# Integrating genome and epigenome data to identify tissue-specific DNA methylation biomarkers for cancer risk

**DOI:** 10.1101/2023.08.09.23293899

**Authors:** Yaohua Yang, Yaxin Chen, Shuai Xu, Xingyi Guo, Guochong Jia, Jie Ping, Xiang Shu, Tianying Zhao, Fangcheng Yuan, Gang Wang, Yufang Xie, Hang Ci, Dan Liu, Fei Ye, Xiao-Ou Shu, Wei Zheng, Li Li, Qiuyin Cai, Jirong Long

## Abstract

The relationship between tissue-specific DNA methylation and cancer risk remains inadequately elucidated. Leveraging the Genotype-Tissue Expression (GTEx) consortium, we developed genetic models to predict DNA methylation at CpG sites (CpGs) across the genome for seven tissues and applied these models to genome-wide association study (GWAS) data of corresponding cancers, namely breast, colorectal, renal cell, lung, ovarian, prostate, and testicular germ cell cancers. At Bonferroni-corrected *P*<0.05, we identified 2,776 CpGs significantly associated with cancer risk, of which 92.7% (2,572) were specific to a particular cancer type. Notably, 57 CpGs within 35 putative novel loci retained significant associations with cancer risk after conditioning on proximal GWAS-identified signals. Further integrative multi-omics analyses revealed 791 CpG-gene-cancer trios, suggesting that DNA methylation at 248 distinct CpGs might influence cancer risk through regulating expression of 145 unique *cis*-genes. These findings substantially advance our understanding of the interplay between genetics, epigenetics, and gene expression in cancer etiology.

---

Genome-wide association studies (GWAS) have identified over 1,000 common variants associated with cancer risk ^1–7^. However, these variants are mainly situated in non-coding regions, posing challenges in identifying target genes and mechanisms ^1–7^. While expression quantitative trait loci (eQTL) studies have uncovered many genes associated with GWAS-identified variants, most of them were limited by the small proportion of gene expression variation captured by individual eQTL variants ^8^. Transcriptome-wide association studies (TWAS) addressed this by incorporating multiple *cis*-variants to predict gene expression, unveiling hundreds of candidate cancer susceptibility genes ^9^. However, over half of GWAS loci lacked TWAS-identified genes, implying the possible existence of additional mechanisms contributing to the genetic susceptibility of cancer risk beyond *cis*-gene expression regulation.

DNA methylation plays crucial roles in regulating gene expression regulation, maintaining genomic stability, and establishing of cell identity ^10^. Aberrant DNA methylation patterns, such as global hypomethylation and gene-specific hypermethylation, are hallmarks of cancers ^11^. In addition to environmental factors, DNA methylation is also shaped by genetics ^12^. In the largest GWAS of blood DNA methylation conducted to date (n=32,851), methylation QTLs (meQTLs) were identified for ∼45.2% (190,102) of CpGs on the Illumina HumanMethylation450 BeadChip^12^. A recent study employed the Illumina MethylationEPIC BeadChip to profile DNA methylation across nine distinct tissue types and discovered meQTLs for 37.9% (286,152) of all investigated CpGs. Of these meQTLs, 37% were detected among all tissues while 5% were specific to a particular tissue type ^13^. In addition, a subset of these meQTLs were found to colocalize with GWAS-identified loci for various traits in biologically relevant tissues ^13^. However, most of these colocalizations did not involve eQTLs, suggesting that genetic effects on trait variation in those loci were more likely to be mediated by DNA methylation rather than gene expression ^13^. Therefore, dissecting tissue-specific genetically determined DNA methylation holds promise in unravelling the genetic susceptibility to complex traits, including cancer susceptibility.

We previously discovered 1,343 CpGs with genetically predicted DNA methylation levels in blood associated with cancer risk ^14–17^. However, the lack of tissue DNA methylation data hindered the evaluation of these findings in cancer-relevant tissues. The present study aimed to identify tissue-specific DNA methylation biomarkers associated with cancer risk and decipher the underlying mechanisms. Leveraging normal tissue DNA methylation data and paired genetic data of cancer-free donors from the Gene-Tissue Expression (GTEx) consortium, we developed statistical models for predicting DNA methylation at CpGs across the genome for seven tissue types. These models were subsequently applied to cancer GWAS data to infer associations between genetically predicted CpG methylation and the risk of breast, colorectal, renal cell, lung, ovarian, prostate, and testicular germ cell cancers, respectively. For identified cancer-associated-CpGs, we employed integrative analyses of DNA methylomic, transcriptomic, genomic, and cancer GWAS data to further explore whether they may affect cancer risk through modulating the expression of nearby genes.

## Results

### Tissue-specific DNA methylation prediction models

The analytical framework of this study is illustrated in **Figure 1**. Processed DNA methylation data, including beta mixture quantile (BMIQ)-normalized β values of 754,054 CpGs across 987 tissue samples from cancer-free GTEx subjects, were obtained from the Gene Expression Omnibus (GEO). After excluding breast tissue samples from males and samples from non-European descendants or without genetic data, 30 breast, 167 colon, 41 kidney, 163 lung, 118 ovary, 96 prostate, 44 testis, 42 whole blood, and 37 muscle tissue samples were retained. For each CpG in each tissue, a single-tissue model was built using elastic net ^18^ and a joint-tissue models was established using multivariate-response penalized regression ^19^ to borrow information from all the other tissues, respectively, and the one with higher prediction performance was kept. Of the 754,054 CpGs investigated, models for 510,444 (67.7%) exhibited reliable prediction performance of R>0.1 and *P*<0.05. Notably, 45% (n=229,907) of these models were highly tissue-specific, found exclusively in one tissue, while only 2% (n=10,145) were ubiquitous across all tissues. Specifically, we built 99,707 models for breast, 202,922 for colon, 138,849 for kidney, 204,339 for lung, 192,188 for ovary, 163,663 for prostate, and 128,843 for testis tissues, respectively.

**Figure 1.**
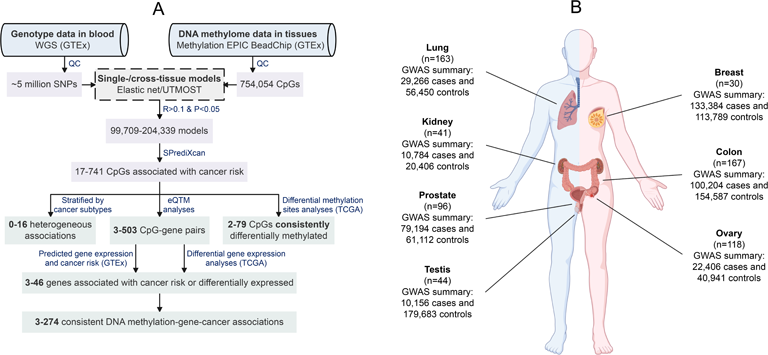
Overall workflow and resources of the present study. **A**, the analysis pipeline. WGS, whole genome sequencing; GTEx, Gene-Tissue Expression; QC, quality control; eQTM, expression quantitative trait methylation; TCGA, The Cancer Genome Atlas. **B,** tissue samples used in DNA methylation prediction model development and cancer GWAS data used in association analyses.

### Association between genetically predicted DNA methylation and cancer risk

For each tissue type, prediction models were applied to GWAS data of the corresponding cancer using SPrediXcan ^20^ to identify CpGs with genetically predicted DNA methylation levels significantly associated with cancer risk at Bonferroni-corrected *P*<0.05. These analyses involved GWAS data of seven cancers, including breast (n=424,601), colorectal (n=254,791), renal cell (n=31,190), lung (n=85,716), ovarian (n=63,347), prostate (n=140,306), and testicular germ cell (n=28,135) cancers. All subjects included in these GWAS were of European ancestry, expect for those in colorectal cancer GWAS, of which 73% and 25% were of European and Asian (27%) ancestry, respectively. In total, 2,776 CpGs were found to be significantly associated with the risk of at least one cancer (**Figure 2**; **Supplementary Tables 1-7**). A total of 95,438 genetic variants were included in prediction models of these 2,776 CpGs, with a median of 29 variants per model (interquartile range [IQR]: 17-46). Remarkably, 92.7% (2,572) of these 2,776 CpGs showed associations that were exclusively to a specific cancer type, including 371 (92.3% of 402) for breast cancer, 715 (95.6% of 748) for colorectal cancer, 17 (100%) for renal cell cancer, 386 (87.7% of 440) for lung cancer, 226 (89.3% of 253) for ovarian cancer, 721 (93.5% of 771) for prostate cancer, and 136 (93.8% of 145) for testicular germ cell cancer (**Figure S1)**. Among these 2,776 CpGs, 2,572 are localized within 315 of the 690 (45.6%) caner susceptibility loci identified by previous GWAS ^1–7^, while the remaining 204 CpGs at 42 loci are at least one mega base (Mb) away from any GWAS-identified cancer risk variants (**Figure 2**).

**Figure 2.**
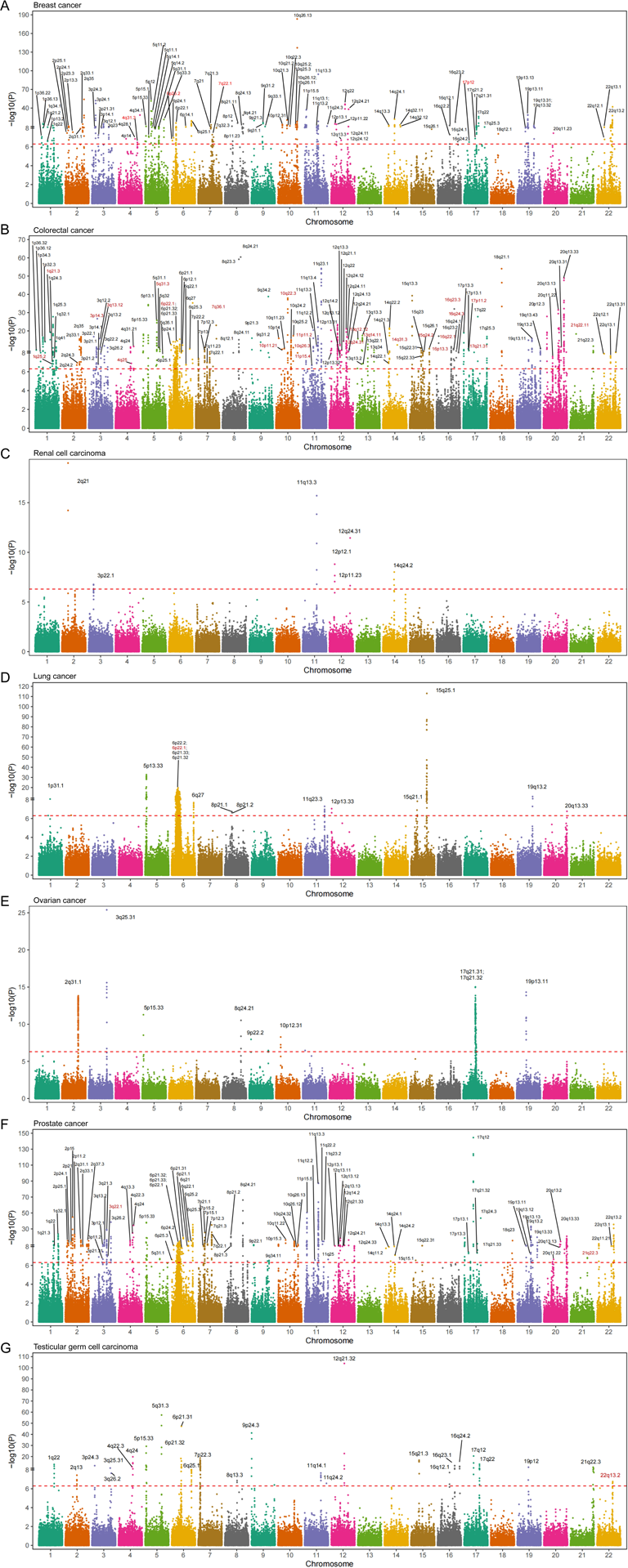
Manhattan plots showing associations between genetically predicted DNA methylation at CpGs and cancer risk. **A**, breast cancer. **B.** colorectal cancer. **C,** renal cell cancer. **D,** lung cancer. **E,** ovarian cancer. **F,** prostate cancer. **G.** testicular germ cell cancer. The dashed red line in each plot denotes a threshold of Bonferroni-corrected *P*<0.05. Cytoband information instead of CpGs were annotated because of the large number of cancer-associated-CpGs. Potential novel loci are highlighted in red.

Upon adjusting for all independent GWAS signals in the nearest known loci, 130 of the 2,776 CpGs sustained a significant association with cancer risk at the same Bonferroni-corrected thresholds (**Supplementary Tables 1-7**). Among these 130 CpGs, 57 CpGs at 35 loci are >1Mb away from any GWAS-reported variants, including 15 CpGs at four loci for breast cancer, 35 CpGs at 27 loci for colorectal cancer, one CpG at one locus for lung cancer, three CpGs at two loci for prostate cancer, and three CpGs at one locus for testicular germ cell cancer (**Table 1; Supplementary Tables 1-7**), suggesting that these loci might be potential novel risk loci for cancers. The remaining 73 CpGs reside in 18 known risk loci, including eight CpGs at five loci for breast cancer, 34 CpGs at seven loci for colorectal cancer, and 31 CpGs at six loci for prostate cancer (**Table 2; Supplementary Tables 1-7**).

**Table 1.** DNA methylation marks associated with cancer risk identified in genomic regions not yet reported for cancer risk.

| CpG | Cytoband | Closest gene | Classification | Z score <sup>a</sup> | OR (95% CI) <sup>a</sup> | *P* <sup>a</sup> | R <sup>b</sup> | Nearest risk variants | Distance (Mb) | *P* <sup>a</sup> adjusted for risk variants |
| --- | --- | --- | --- | --- | --- | --- | --- | --- | --- | --- |
| Breast cancer |  |  |  |  |  |  |  |  |  |  |
| cg26218269 | 4q31.3 | *MAB21L2; LRBA* | 5'URT | 5.34 | 1.11 (1.07-1.16) | 9.23×10<sup>-8</sup> | 0.51 | rs6828523 | 24.3 | 9.24×10<sup>-8</sup> |
| cg03611078 | 6p25.2 | *C6orf201* | 5'UTR | 5.19 | 1.12 (1.08-1.18) | 2.15×10<sup>-7</sup> | 0.45 | rs11242675 | 2.8 | 4.65×10<sup>-7</sup> |
| cg03637179 | 7q22.1 | *TAF6* | 5'UTR | -5.45 | 0.80 (0.74-0.87) | 5.14×10<sup>-8</sup> | 0.36 | rs79518236 | 1.7 | 5.36×10<sup>-8</sup> |
| cg22652561 | 17p12 | *MAP2K4* | TSS200 | 5.12 | 1.07 (1.04-1.10) | 3.09×10<sup>-7</sup> | 0.40 | rs78378222 | 4.4 | 1.78×10<sup>-7</sup> |
| Colorectal cancer |  |  |  |  |  |  |  |  |  |  |
| cg18755616 | 1q21.3 | *KCNN3* | Intronic | 9.15 | 1.31 (1.24-1.39) | 5.94×10<sup>-20</sup> | 0.62 | rs5028523 | 18.1 | 6.75×10<sup>-20</sup> |
| cg15503006 | 1q25.2 | *FAM20B* | TSS200 | -5.37 | 0.78 (0.71-0.85) | 8.07×10<sup>-8</sup> | 0.18 | rs10911251 | 4.1 | 3.36×10<sup>-8</sup> |
| cg06965496 | 3p14.3 | *FLNB* | Intronic | 10.84 | 1.38 (1.30-1.46) | 2.14×10<sup>-27</sup> | 0.23 | rs9831861 | 5.0 | 8.52×10<sup>-28</sup> |
| cg24683684 | 3q13.12 | *BBX* | Intronic | -6.80 | 0.87 (0.84-0.91) | 1.03×10<sup>-11</sup> | 0.23 | rs12635946 | 5.6 | 2.70×10<sup>-14</sup> |
| cg10945313 | 4q25 | *ANK2* | 5'URT | 5.28 | 1.13 (1.08-1.18) | 1.31×10<sup>-7</sup> | 0.16 | rs2388976 | 1.8 | 1.21×10<sup>-11</sup> |
| cg03172688 | 5q31.3 | *PCDH1* | Intronic | -6.78 | 0.87 (0.84-0.91) | 1.22×10<sup>-11</sup> | 0.30 | rs647161 | 6.8 | 6.58×10<sup>-20</sup> |
| cg24248978 | 6p22.1 | *ZNF165* | 5'URT | -7.76 | 0.86 (0.83-0.90) | 8.77×10<sup>-15</sup> | 0.16 | rs1476570 | 1.8 | 2.19×10<sup>-14</sup> |
| cg25824330 | 6p21.1 | *MRPS18A* | Intronic | 5.74 | 1.31 (1.19-1.43) | 9.42×10<sup>-9</sup> | 0.16 | rs57939401 | 1.9 | 2.70×10<sup>-8</sup> |
| cg06766817 | 6q25.3 | *SYNJ2* | Intronic | -6.51 | 0.73 (0.66-0.80) | 7.69×10<sup>-11</sup> | 0.21 | rs151127921 | 24.5 | 9.71×10<sup>-11</sup> |
| cg04095069 | 7q36.1 | *NUB1* | Intronic | -10.05 | 0.85 (0.82-0.88) | 8.80×10<sup>-24</sup> | 0.16 | rs73161913 | 20.5 | 7.61×10<sup>-25</sup> |
| cg05877768 | 10p11.21 | *PARD3* | Intronic | 6.44 | 1.25 (1.17-1.34) | 1.21×10<sup>-10</sup> | 0.17 | rs1773860 | 5.1 | 7.56×10<sup>-15</sup> |
| cg00171421 | 10q22.3 | *KCNMA1* | Exonic | -7.72 | 0.84 (0.80-0.88) | 1.17×10<sup>-14</sup> | 0.18 | rs704017 | 1.4 | 2.50×10<sup>-16</sup> |
| cg12338214 | 10q26.3 | *BNIP3* | Intronic | -5.73 | 0.89 (0.85-0.93) | 1.01×10<sup>-8</sup> | 0.32 | rs11196172 | 19.1 | 1.04×10<sup>-8</sup> |
| cg16984897 | 11p15.4 | *STIM1* | TSS1500 | -5.39 | 0.83 (0.78-0.89) | 7.01×10<sup>-8</sup> | 0.27 | rs4450168 | 6.4 | 1.55×10<sup>-9</sup> |
| cg07438103 | 11p11.2 | *C11orf49* | Intronic | -5.19 | 0.80 (0.74-0.87) | 2.07×10<sup>-7</sup> | 0.27 | rs174537 | 14.4 | 2.08×10<sup>-7</sup> |
| cg05190884 | 12q24.31 | *ZNF664-RFLNA* | Intronic | -6.50 | 0.91 (0.89-0.94) | 7.95×10<sup>-11</sup> | 0.43 | rs73208120 | 6.8 | 7.52×10<sup>-10</sup> |
| cg24776343 | 13q12.12 | *TNFRSF19* | Intronic | 8.22 | 1.28 (1.21-1.36) | 2.02×10<sup>-16</sup> | 0.19 | rs116964464 | 3.4 | 8.93×10<sup>-15</sup> |
| cg24467989 | 13q14.11 | *ELF1* | Intronic | -5.27 | 0.67 (0.58-0.78) | 1.35×10<sup>-7</sup> | 0.18 | rs7333607 | 4.1 | 7.08×10<sup>-9</sup> |
| cg13259103 | 14q31.3 | *GALC* | Intronic | -7.23 | 0.80 (0.75-0.85) | 4.76×10<sup>-13</sup> | 0.18 | rs61975764 | 4.6 | 1.45×10<sup>-13</sup> |
| cg24797896 | 15q24.2 | *C15orf39* | Intronic | -6.06 | 0.91 (0.89-0.94) | 1.36×10<sup>-9</sup> | 0.34 | rs8031386 | 3.0 | 5.41×10<sup>-9</sup> |
| cg27634195 | 16p13.3 | *UNKL* | Intronic | -8.60 | 0.88 (0.85-0.90) | 7.82×10<sup>-18</sup> | 0.26 | rs9929218 | 67.4 | 3.48×10<sup>-26</sup> |
| cg09232225 | 16q22.1 | *CES4A* | TSS200 | -5.24 | 0.95 (0.93-0.97) | 1.63×10<sup>-7</sup> | 0.42 | rs9929218 | 1.8 | 7.05×10<sup>-6</sup> |
| cg04290162 | 16q23.3 | *MPHOSPH6* | TSS1500 | -11.92 | 0.78 (0.75-0.81) | 9.26×10<sup>-33</sup> | 0.16 | rs9930005 | 2.2 | 2.86×10<sup>-41</sup> |
| cg04996089 | 16q24.3 | *PABPNIL; CBFA2T3* | Intergenic | -5.78 | 0.89 (0.86-0.93) | 7.68×10<sup>-9</sup> | 0.39 | rs2696839 | 2.6 | 3.95×10<sup>-8</sup> |
| cg13876325 | 17p11.2 | *PIGL* | Intronic | -6.19 | 0.37 (0.27-0.51) | 5.98×10<sup>-10</sup> | 0.20 | rs1078643 | 5.5 | 1.08×10<sup>-8</sup> |
| cg15210992 | 17q21.31 | *AOC3* | TSS1500 | -5.83 | 0.95 (0.93-0.96) | 5.42×10<sup>-9</sup> | 0.27 | rs983318 | 29.4 | 6.78×10<sup>-8</sup> |
| cg11650372 | 21q22.11 | *ITSN1* | Intronic | 8.92 | 1.22 (1.16-1.27) | 4.49×10<sup>-19</sup> | 0.18 | rs9983528 | 12.6 | 2.19×10<sup>-20</sup> |
| Lung cancer |  |  |  |  |  |  |  |  |  |  |
| cg23119604 | 6p22.1 | *HLA-L* | Exonic | 5.59 | 1.11 (1.07-1.16) | 2.28×10<sup>-8</sup> | 0.49 | rs9267123 | 1.2 | 1.53×10<sup>-7</sup> |
| Prostate cancer |  |  |  |  |  |  |  |  |  |  |
| cg21411781 | 3q22.1 | *CPNE4; ACPP* | Intergenic | -5.31 | 0.89 (0.85-0.93) | 1.11×10<sup>-7</sup> | 0.20 | rs35006112 | 3.8 | 3.00×10<sup>-10</sup> |
| cg10650821 | 6p21.33 | *TNF* | Exonic | 6.98 | 1.65(1.43-1.90) | 2.96×10<sup>-12</sup> | 0.21 | rs9275160 | 1.1 | 2.61×10<sup>-8</sup> |
| Testicular germ cell cancer |  |  |  |  |  |  |  |  |  |  |
| cg18042004 | 22q13.2 | *POLR3H* | TSS200 | -5.15 | 0.60(0.49-0.73) | 2.65×10<sup>-7</sup> | 0.381 | rs739525 | 20.6 | 2.68×10<sup>-7</sup> |

OR, odds ratio per standard deviation (SD) increase in genetically predicted DNA methylation level; CI, confidence interval; Mb, megabase; TSS, transcription start site; UTR, untranslated region.
<sup>a</sup> Association Z scores, ORs, 95% CIs, and *P* values were estimated using SPrediXcan. All statistical tests were two-sided.
<sup>b</sup> Coefficients of correlation between predicted and measured DNA methylation levels.

**Table 2.** DNA methylation marks associated with cancer risk identified in genomic regions within 1Mb of known cancer risk variants but representing independent association signals.

| CpG | Cytoband | Closest gene | Classification | Z score <sup>a</sup> | OR (95% CI) <sup>a</sup> | *P* <sup>a</sup> | R <sup>b</sup> | Closest risk variants | Distance (Mb) | *P* <sup>a</sup> adjusted for risk variants |
| --- | --- | --- | --- | --- | --- | --- | --- | --- | --- | --- |
| **Breast cancer** |  |  |  |  |  |  |  |  |  |  |
| cg23178958 | 2p13.3 | ANTXR1 | Intronic | -5.56 | 0.92 (0.89-0.94) | 2.77×10<sup>-8</sup> | 0.38 | rs4602255 | 0.005 | 1.08×10<sup>-7</sup> |
| cg20242213 | 5p12 | MRPS30-DT | Intronic | 7.97 | 1.07 (1.05-1.09) | 1.64×10<sup>-15</sup> | 0.49 | rs10941679 | 0.068 | 8.74×10<sup>-8</sup> |
| cg01444058 | 6q25.1 | *LOC102723831; AP12* | Intergenic | -7.35 | 0.87 (0.83-0.90) | 2.04×10<sup>-13</sup> | 0.55 | rs3757322 | 0.383 | 1.31×10<sup>-10</sup> |
| cg08353955 | 11q13.3 | LOC102724265;<br>LINC01488 | Intergenic | -20.70 | 0.66 (0.64-0.69) | 3.48×10<sup>-95</sup> | 0.64 | rs554219 | 0.042 | 3.82×10<sup>-11</sup> |
| cg02633117 | 19p13.11 | MYO9B | Intronic | -5.04 | 0.90 (0.87-0.94) | 4.64×10<sup>-7</sup> | 0.44 | rs67397200 | 0.214 | 4.15×10<sup>-8</sup> |
| **Colorectal cancer** |  |  |  |  |  |  |  |  |  |  |
| cg10474949 | 6p22.1 | ZFP57; HLA-F | Intergenic | 7.10 | 1.72 (1.48-2.00) | 1.25×10<sup>-12</sup> | 0.26 | rs1476570 | 0.132 | 1.51×10<sup>-9</sup> |
| cg09037630 | 6p21.33 | LOC100287329 | Exonic | -6.98 | 0.71 (0.65-0.78) | 3.01×10<sup>-12</sup> | 0.24 | rs3830041 | 0.663 | 9.46×10<sup>-12</sup> |
| cg04105091 | 6p21.32 | LOC100507547 | Exonic | -6.95 | 0.86 (0.82-0.89) | 3.63×10<sup>-12</sup> | 0.21 | rs3830041 | 0.070 | 1.20×10<sup>-7</sup> |
| cg12444411 | 7p22.2 | GNA12 | 5'UTR | 5.49 | 1.07 (1.05-1.10) | 3.94×10<sup>-8</sup> | 0.58 | rs1182197 | 0.061 | 6.66×10<sup>-58</sup> |
| cg14334258 | 9q31.2 | *KLF4; ACTL7B* | Intergenic | -7.69 | 0.91 (0.89-0.94) | 1.51×10<sup>-14</sup> | 0.31 | rs10978941 | 0.222 | 2.10×10<sup>-11</sup> |
| cg14472025 | 9q34.2 | GBGT1 | Intronic | 13.15 | 1.60 (1.49-1.71) | 1.59×10<sup>-39</sup> | 0.19 | rs7038489a | 0.643 | 6.95×10<sup>-40</sup> |
| cg05475172 | 22q13.31 | WNT7B | Intronic | -5.85 | 0.94 (0.93-0.96) | 5.05×10<sup>-9</sup> | 0.65 | rs736037 | 0.646 | 1.08×10<sup>-8</sup> |
| **Prostate cancer** |  |  |  |  |  |  |  |  |  |  |
| cg11886554 | 3q26.2 | SKIL | Intronic | 5.12 | 1.10 (1.06-1.13) | 3.01×10<sup>-7</sup> | 0.67 | rs78416326 | 0.002 | 2.96×10<sup>-8</sup> |
| cg26668675 | 6p21.33 | *PSORS1C3* | Intronic | 7.28 | 1.28 (1.19-1.36) | 3.37×10<sup>-13</sup> | 0.41 | rs62407547 | 0.932 | 1.69×10<sup>-9</sup> |
| cg06131755 | 6q25.3 | *SOD2-OT1* | Exonic | -5.82 | 0.90(0.87-0.93) | 5.89×10<sup>-9</sup> | 0.50 | rs963800 | 0.032 | 9.36×10<sup>-8</sup> |
| cg20253542 | 7p15.2 | *EVX1; HIBADH* | Intergenic | -9.14 | 0.85 (0.82-0.88) | 6.40×10<sup>-20</sup> | 0.54 | rs6956484 | 0.004 | 2.54×10<sup>-7</sup> |
| cg04726730 | 11p15.5 | *TNNI2; LSP1* | Intergenic | 7.54 | 1.89 (1.60-2.24) | 4.53×10<sup>-14</sup> | 0.26 | rs1881502 | 0.361 | 1.35×10<sup>-14</sup> |
| cg16166568 | 11q13.3 | *LOC338694; MYEOV* | Intergenic | -19.88 | 0.79 (0.77-0.80) | 6.39×10<sup>-88</sup> | 0.77 | rs11825796 | 0.006 | 4.62×10<sup>-16</sup> |

OR, odds ratio per standard deviation (SD) increase in genetically predicted DNA methylation level; CI, confidence interval; Mb, megabase; UTR, untranslated region.
<sup>a</sup> Association Z scores, ORs, 95% CIs, and *P* values were estimated using SPrediXcan. All statistical tests were two-sided.
<sup>b</sup> Correlation coefficients of predicted and measured DNA methylation levels.

We compared the capability of approach we employed here with the TWAS approach in delineating associations at known cancer susceptibility loci identified by previous GWAS. Results of gene- and splicing-based TWAS were either acquired from recent studies ^3,21^ or generated by SPrediXcan analyses using GTEx (v8)-based prediction models and cancer GWAS data. Despite the on average ∼46% (9% to 85%) smaller sample size in prediction model development of our study, we uncovered significant associations in more known loci than TWAS (315 vs. 302), particularly for renal cell cancer (six vs. one) and prostate cancer (81 vs. 65).

Noteworthy, in 27.9% of (88 out of 315) known loci which have CpGs significantly associated with cancer risk, TWAS was unable to identify any significant associations. These results emphasize the pronounced effectiveness of our approach in detecting association signals within GWAS-identified loci compared to TWAS.

The 2,776 cancer-associated-CpGs we identified showed significant enrichments in various genomic regulatory regions in relevant tissues and cell lines at a false discovery rate (FDR)<0.05 (**Supplementary Figure 2**). For instance, colorectal-cancer-associated CpGs were significantly enriched at active genomic regions that were featured by DNase I hypersensitive sites (DHS), chromatin states with enhancer and promoter signatures, and histone marks including H3K4me1, H3K4me3 and H3K36me3 in intestine tissues/cells. In contrast, lung-cancer-associated CpGs exhibited significant enrichments in inactive genomic regions that were characterized by Polycomb-repressed chromatin and the H3K27me3 mark in lung tissues/cells. For cancers with GWAS data stratified by histological types available, most CpGs showing significant associations with overall cancer risk were consistently found to be associated with the risk of most cancer subtypes. Nevertheless, significant cross-subtype heterogeneity was observed for associations of 16, 3, and 8 CpGs with the risk of breast, lung, and ovarian cancers, respectively, at the Bonferroni-corrected thresholds utilized in main analyses (**Supplementary Tables 1, 4, 5**).

Except for ovarian cancer and testicular germ cell cancer, CpGs associated with any of the other five cancers were investigated for their differential methylation between tumor and adjacent normal tissues using data from The Cancer Genome Atlas (TCGA). Among these 2,378 CpGs, data on 1,198 (∼50%) CpGs were available in TCGA, which is consistent with the coverage difference between the DNA methylome profiling arrays used by the present study (Illumina MethylaitonEPIC array) and TCGA (Humanmehylation450 BeadChip). Of these 1,198 CpGs, 300 (∼25%) showed differential DNA methylation at *P*<0.05 with directions of effects consistent with those of CpG-cancer associations. This included 63 out of 188 CpGs for breast cancer, 88 out of 370 CpGs for colorectal cancer, two out of six CpGs for renal cell cancer, 68 out of 317 CpGs for lung cancer, and 79 out of 316 CpGs for prostate cancer (**Supplementary Tables 8-12**).

### DNA methylation influencing cancer risk through modulating *cis*-gene expression

To search for potential target genes of cancer-associated CpGs, we performed expression quantitative trait methylation (eQTM) analyses using GTEx data of the corresponding tissue. At FDR<0.05, we identified 1,242 CpG-gene association pairs, including 245 (55 CpGs and 50 genes) in breast, 72 (49 CpGs and 58 genes) in colon, three (three CpGs and three genes) in kidney, 358 (87 CpGs and 51 genes) in lung, 503 (85 CpGs and 36 genes) in ovary, 48 (34 CpGs and 45 genes) in prostate, and 13 (10 CpGs and 12 genes) in testis. Genes involved in these CpG-gene pairs were then assessed for their impacts on the proliferation of relevant cancer cells using Clustered Regularly Interspaced Short Palindromic Repeat (CRISPR)-Cas9 essentiality screening data from DepMap. Out of 117 genes with data available in DepMap, 19 demonstrated essential roles in cell proliferation at a commonly used threshold of median CERES Score <-0.5 ^22^, including *ELL*, *ADSL*, and *KANSL1* in breast cancer cells, *ZNF574* in colorectal cancer cells, *CCND1* for renal cell cancer cells, *DHX16*, *HYOU1*, *ABCF1*, *PPP1R10*, and *DDX39B* for lung cancer cells, *NSF* and *KANSL1* for ovarian cancer cells, *LSM2*, *CCT4*, *CTDP1*, *CPSF3*, *MRPL45*, *FDPS*, *PRRC2A*, and *NOLC1* in prostate cancer cells (**Supplementary Figure 3**).

We further investigated target genes of cancer-associated CpGs for their predicted expression in association with cancer risk using GTEx and cancer GWAS data, as well as for their differential expression between tumor and adjacent normal tissues using TCGA data. At FDR<0.05, an average of 88.2% (80.6% to 100.0%) of these genes showed a significant association or differential expression. By integrating findings from associations between CpGs and cancers, between CpGs and genes, and between genes and cancers, we revealed 791 CpG-gene-cancer trios. Within each trio, the relationships of CpG-cancer, CpG-gene, and gene-cancer showed consistent directions (**Table 3**; **Supplementary Tables 13-19**). Involved in these 791 trios were 248 unique CpGs, 145 distinct *cis*-genes of these CpGs, and seven cancers. Such trios provided evidence supporting the potential mechanism that these 248 CpG might influence cancer risk through regulating the expression of these 145 genes. For example, as shown in **Table 3 and Figure 3**, genetically predicted DNA methylation at cg22872885 was associated with decreased breast cancer risk, which was consistent with the lower methylation of this CpG in breast cancer tissues than in adjacent normal tissues. This association may be explained by the negative association between DNA methylation at cg22872885 and expression of the *ZMIZ1* gene, and the positive association between genetically predicted expression of *ZMIZ1* and breast cancer risk. Consistently, the expression of *ZMIZ1* was significantly higher in breast cancer tissues than in adjacent normal tissues.

**Figure 3.**
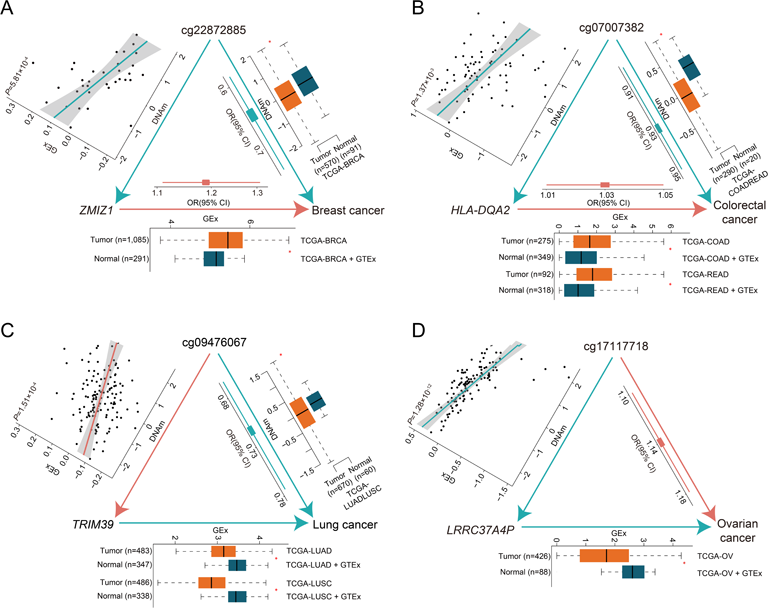
Examples of CpG-gene-cancer trios suggest DNA methylation affecting cancer risk by modulating nearby gene expression. Red arrows, lines, and blocks denote positive associations, while green ones denote negative associations. In boxplots, red and green boxes represent data of tumor and adjacent normal tissues, respectively. DNAm, DNA methylation; GEx, gene expression; OR, odds ratio; CI, confidence interval; TCGA, The Cancer Genome Atlas; BRCA, breast invasive carcinoma; COAD, colon adenocarcinoma; READ, rectum adenocarcinoma; LUAD, lung adenocarcinoma, LUSC, lung squamous cell carcinoma; OV, ovarian serous cystadenocarcinoma. **A**, DNA methylation at cg22872885 may decrease breast cancer risk by suppressing the expression of *ZMIZ1*. **B**, DNA methylation at cg07007382 may decrease colorectal cancer risk by suppressing the expression of *HLA-DQA2*. **C**, DNA methylation at cg09476067 may decrease lung cancer risk by promoting the expression of *TRIM39*. **D**, DNA methylation at cg17117718 may increase ovarian cancer risk by suppressing the expression of *LRCC37A4P*.

**Table 3.**
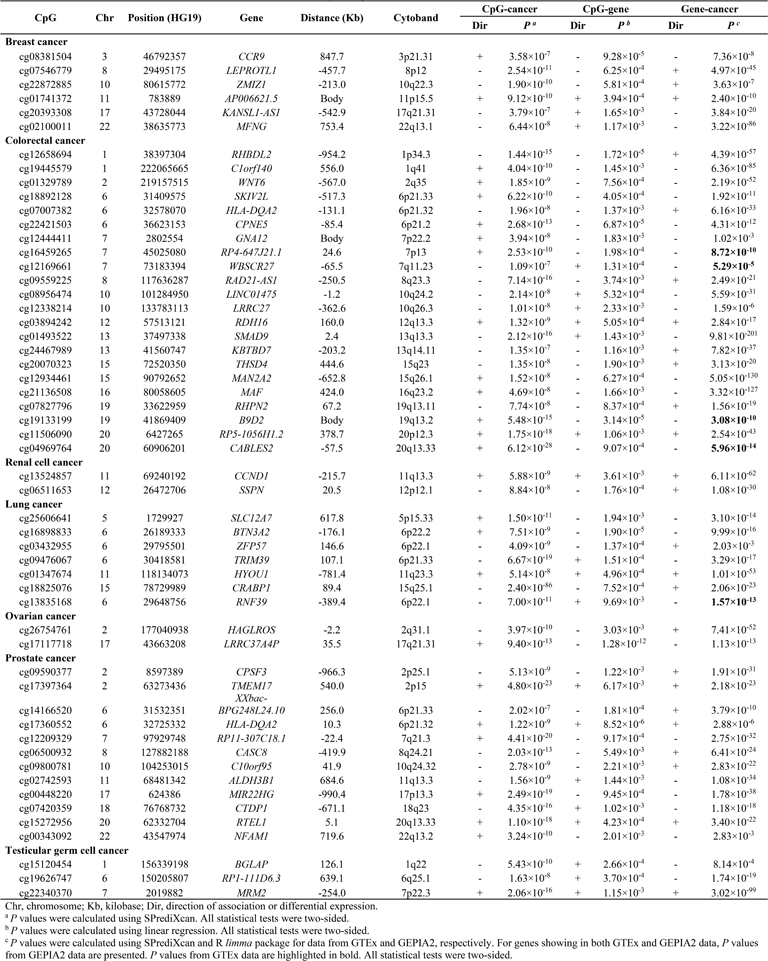
Consistent directions of associations across DNA methylation, gene expression, and cancer risk.

## Discussion

In this comprehensive investigation of tissue-specific DNA methylation and cancer risk using genetic instruments, we identified 2,776 cancer-associated CpGs, with more than 92% being specific to a particular cancer type. Integrative analyses of multi-omics data unveiled 791 CpG-gene-cancer trios indicating that unique 248 CpGs may be associated with cancer risk via modulating the expression of 145 distinct *cis*-genes. These findings strengthen our understanding of the intricate interplays among genetics, epigenetics, and gene expression in cancer etiology.

Compared to a previous study that identified meQTLs for 286,152 CpGs across nine tissues ^13^, our study established prediction models for ∼1.8 times more CpGs (n=510,444) spanning seven tissues. In addition, diverging from their observation that 5% of meQTLs were specific to a particular tissue and 37% were shared across all tissues, prediction models built in the present study showcased a distinctive pattern, with almost 45% of models exclusive to a specific tissue type and only 2% being detected among all tissues. These results indicate the increased sensitivity of our prediction models in capturing tissue-specific genetic determinants of DNA methylation. Notably, more than 92% of cancer-associated CpGs identified by our study were specific to a particular cancer, underlining the tissue-specific epigenetic mechanisms driving carcinogenesis. Nonetheless, it is worth noting that the relatively modest sample size of tissue samples used for model construction and the varied samples sizes of cancer GWAS might also contributed to the tissue-specificity of models and cancer-associated-CpGs.

Our preceding studies using blood-based models identified multiple CpGs associated with the risk of four cancers ^14–17^. In the present study based on tissue-specific models, we identified a comparable number of CpGs for breast, lung, and prostate cancer, but nearly three times more CpGs for ovarian cancer, than our previous studies. Nearly 80% of CpG-cancer associations identified for these four cancers in the current study were not found in our previous studies. This disparity might be attributed to the tissue-specific nature of these CpGs or their absence in the Illumina 450K array, which was used in our previous studies.

A recent study investigated genetically predicted colorectal tissue DNA methylation and colorectal cancer risk ^3^. Our study of colorectal cancer utilized the same GWAS dataset, yet with profound improvements in prediction model development. First, we exclusively employed data of transverse colon tissues from cancer-free individuals, while the recent study involved various colorectal tissue types, including those adjacent to tumors of colorectal cancer patients. In addition, our study benefited from the inclusion of data from eight other distinct tissue types, enabling the development of colon-specific models while leveraging information from other tissues. In contrast, the recent study was confined to building single-tissue models. Finally, the current study, even with a more stringent threshold to select models (R>0.10 and *P*<0.05), established models for almost 6.7 times more CpGs (202,922 vs. 30,385) than the recent study. As a result, our association analyses identified nearly 1.5 times as many colorectal-cancer-associated CpGs (748 vs. 501) and replicated almost 20% of the CpGs reported by the recent study ^3^.

For ∼12% of cancer-associated CpGs, we pinpointed potential target genes in corresponding tissues. This observation is in alignment with previous findings that a large proportion of meQTL-GWAS colocalizations lack eQTL involvement ^13^. These results suggest the potential presence of mechanisms that are alternative to gene expression regulation underlying most variant-cancer associations. Noteworthy, for 19 of these genes, we found evidence from CRISPR-Cas9 screening data supporting their essential roles in the proliferation of corresponding cancer cells, many of which have been implicated in cell proliferation, such as *ADSL* in breast cancer ^23^ and *FDPS* in prostate cancer ^24^. Further, 145 of these target genes were involved in 791 CpG-gene-cancer trios, implying the impacts of 248 CpG on cancer risk by regulating expression of these genes. Among them, nine genes were essential for cancer cell proliferation, including *KANSL1* for breast cancer, *CCND1* for renal cell cancer, *DHX16*, *HYOU1* and *ABCF1* for lung cancer, *NSF* for ovarian cancer, and *CCT4*, *CTDP1*, and *CPSF3* for prostate cancer. Altogether, these findings demonstrated the capability of our study to identify functional genes that might be involved in putative genetic variants-DNA methylation-gene expression-cancer pathways.

Our study has notable strengths. First, we utilized Illumina MethylationEPIC BeadChip DNA methylation data from normal tissue samples of cancer-free individuals, enabling unbiased estimation of genetic determinants of DNA methylation at ∼750,000 CpGs. In addition, both single- and joint-tissue prediction models were developed, considering both tissue-specific and shared genetic determinants to improve prediction accuracy. Moreover, despite a smaller sample size involved in model development, our study successfully identified significant associations in a greater number of known GWAS loci compared to gene- and splicing-based TWAS. Further, the replication of a substantial proportion of associations using external data of tumor and adjacent normal tissue samples strengthened the validity of our findings. Finally, the discovery of CpG-gene-cancer trios provided mechanistic insights into the critical roles of epigenetics in the genetic etiology of cancer.

Several limitations should be noted. First, the sample size for prediction model development, despite being one of the largest for many tissue types, remained relatively small compared to the extensive number of examined CpGs. Enlarging the sample sizes, particularly for tissues with limited samples, would enhance model precision and possibly unveil additional significant associations. Second, the GWAS data of colorectal cancer included individuals of both European and East Asian ancestries. Genetic differences between populations might affect the performance of certain models, particularly in regions with ancestral variations, which might consequently impact the detection of significant associations ^4^. Finally, the differential methylation analyses using TCGA data were limited by the small sample size and the potential differences in DNA methylation profiles between normal tissues adjacent to tumors and those obtained from cancer-free subjects. Future studies employing normal tissue samples from cancer-free individuals, coupled with functional experiments, are needed to further corroborate our findings.

In summary, we identified more than 2,500 CpGs showing tissue-specific associations with cancer risk, nearly 250 of which may influence cancer risk by regulating neighbor gene expression. Our findings emphasize the effectiveness of multi-omics integration in cancer biomarker discovery and enhance our comprehension of the critical role of genetics and epigenetics in cancer etiology.

### Online Methods Data Acquisition

Whole-genome sequencing (WGS) data of blood samples and Illumina MethylationEPIC BeadChip DNA methylation data of normal tissue samples from GTEx (v8) were used as references to build DNA methylation prediction models ^8,13^. Genotype and phenotype data were downloaded from the database of Genotype and Phenotype (dbGaP) (phs000424.v8.p2).

Normalized DNA methylation data of nine tissue types, including breast, colon, kidney, lung, ovary, prostate, testis, whole blood, and muscle, was obtained from GEO (GSE213478). Detailed information on sample preparation, sequencing, and data processing were described elsewhere ^8,13^. Briefly, WGS libraries built from blood DNA samples from 838 donors were sequenced on the Illumina HiSeq X or Hiseq 2000 platform at the Broad Institute with a median coverage of ∼32X. Genotype data of European ancestry were extracted and non-palindromic variants with missing data <5%, minor allele frequency (MAF) >5%, and Hardy-Weinberg equilibrium (HWE) *P*>10^-4^ were retained for subsequent analyses. Epigenome-wide DNA methylation profiling was performed using the Illumina MethylationEPIC BeadChip based on 1,000 tissue DNA samples across nine unique tissue types obtained from 424 subjects. The R package *ChAMP* (v.2.8.6) ^25^ was utilized to process raw data to exclude low-quality samples and CpGs and estimate DNA methylation β values ^13^. After background correction using the single sample normal-exponential out-of-band (ssnoob) method implemented in the R package *minfi* (v.1.36.0) ^26^, β values were normalized using the BMIQ method ^13^. Finally, DNA methylation data of 754,119 CpGs among 738 samples and genotype data of ∼5.1 (IQR: 4.6-5.6) variants from 317 subjects were included in prediction model development.

Summary statistics of GWAS data for breast, colorectal, renal cell, lung, ovarian, prostate, and testicular germ cell cancers were acquired from different sources ^1–7^. Except for colorectal cancer, data of which were from a meta-analysis of GWAS among European and Asian descendants, data for all the other cancers were from GWAS among European descendants. Breast cancer data came from a meta-analysis of the Breast Cancer Association Consortium and UK Biobank, including 133,511 cases and 291,090 controls ^21^. Colorectal cancer data was accessed from GWAS catalog (GCST90129505), including 100,204 CRC cases and 154,587 controls, comprising 78,473 cases and 107,143 controls of European ancestry from Genetics and Epidemiology of Colorectal Cancer Consortium (GECCO), the Colorectal Cancer Transdisciplinary Study (CORECT) and the Colon Cancer Family Registry (CCFR), and 21,731 and 47,444 of Asian Ancestry from the Asia Colorectal Cancer Consortium (ACCC) ^3^. Renal cell cancer data was retrieved from dbGaP (phs001736.v2.p1), including 10,784 cases and 20,406 controls from six datasets, two from International Agency for Research on Cancer (IARC), two from National Cancer Institute (NCI), one from the University of Texas MD Anderson Cancer, and one from the Institute of Cancer Research, UK ^7^. Lung cancer data was downloaded from GWAS catalog (GCST004746), including 29,266 cases and 56,450 controls from the Transdisciplinary Research of Cancer in Lung of the International Lung Cancer Consortium (TRICL-ILCCO) and the Lung Cancer Cohort Consortium (LC3) ^1^. Ovarian cancer data was obtained from the Ovarian Cancer Association Consortium, including 22,406 cases and 40,941 controls ^5^. Prostate cancer data was accessed from the Prostate Cancer Association Group to Investigate Cancer Associated Alterations in the Genome (PRACTICAL) consortium, including 79,194 cases and 61,112 controls ^2^. Testicular germ cell cancer data was retrieved from dbGaP (phs001349), including 10,156 cases and 17,979 from The Testicular Cancer Consortium ^6^.

### DNA Methylation Prediction Model Development

For each tissue, BMIQ-normalized DNA methylation β values were inverse-normalized within each CpG and regressed on covariates to get residuals. These covariates included top five genetic principle components (PCs), Probabilistic Estimation of Expression Residuals (PEER) ^27^ factors (n=5 for breast, kidney, testis, muscle, and blood; n=20 for colon, lung, ovary, and prostate), sex (only for colon, kidney, lung, muscle, blood), and indicators for WGS sequencing platform (HiSeq X or HiSeq 2000) and library construction protocol indicator (PCR based or PCR-free). For each CpG, a single-tissue prediction model was built using genetic variants within its 500Kb flanking region to predict its inverse-normalized methylation residuals by fitting an elastic net model (α=0.5) ^18^. In addition, a cross-tissue prediction model borrowing information from DNA methylation data of all the other eight tissues from the same set of donors was established using the multivariate-response penalized regression method implemented in Unified Test for MOlecular SignaTures (UTMOST) ^19^. For both strategies, five-fold cross-validation was performed to evaluate prediction performance and satisfactory models were identified at R>0.1, 10% correlation between predicted and measured DNA methylation levels, and *P*<0.05 ^19^. CpGs with at least one model meeting these criteria were considered in downstream association analyses. If both models are qualified, the model with higher R value was used in downstream association analyses.

### Association Analyses between Genetically Predicted DNA Methylation and Cancer Risk

SPrediXcan ^20^ was utilized to assess associations between genetically predicted DNA methylation level at CpGs and cancer risk. The association Z score was calculated following the below formula, in which *W*_SNP–m_ represents the weight of variant *S* on DNA methylation levels at CpG *m*, 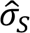 and 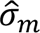 represents estimated variances of variant *S* and CpG *m*, and 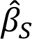 and (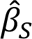) represents effect size and standard error of the association between variant *S* and cancer risk, respectively.

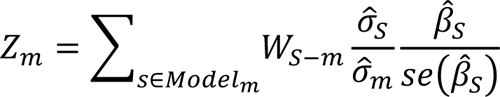

Association analyses and Bonferroni-correction were conducted for each cancer type separately and significant associations were identified at Bonferroni-corrected *P*<0.05, corresponding to 5.01×10^-7^ (0.05/99,707) for breast cancer, 2.46×10^-7^ (0.05/202,922) for colorectal cancer, 3.60×10^-7^ (0.05/138,849) for renal cell cancer, 2.45×10^-7^ (0.05/204,339) for lung cancer, 2.60×10^-7^ (0.05/192,188) for ovarian cancer, 3.05×10^-7^ (0.05/163,663) for prostate cancer, and 3.88×10^-7^ (0.05/128,843) for testicular germ cell cancer.

For each CpG-cancer association, we evaluated its independence from its nearest GWAS risk signals. Briefly, for each CpG, we identified variants that were independently associated with cancer risk at *P*<5×10^-8^ in its nearest GWAS-reported cancer susceptibility locus through a stepwise model selection procedure implemented in GCTA-COJO (v1.91.3 beta) ^28^. Then for each variant included in the prediction model of this CpG, its association with cancer risk conditioning on all variants identified in the first step was evaluated using GCTA-COJO. Finally, SPrediXcan analysis was conducted using the summary statistics generated in the second step and the Bonferroni-corrected thresholds used in main analyses were applied to determine significance.

For CpGs significantly associated with cancer risk, we first carried out eFORGE (v2.0) analyses with default settings to evaluate their overlap with DHS, chromatin states, and histone marks, respectively, in relevant tissues and cell lines using data from the Roadmap Epigenomics Project^29^. Then for those associated with breast, renal cell, lung, and ovarian cancer, stratification analyses by cancer subtypes were performed and for each cancer, the heterogeneity in associations between subtypes was examined via Cochrane’s Q test and significant heterogeneity was detected at the Bonferroni-corrected thresholds used in main analyses. Further, except for ovarian and testicular germ cell cancer, CpGs associated with other cancer were interrogated for their differential methylation between tumor and adjacent normal tissues using Illumina HumanMethylation450 BeadChip DNA methylation data from TCGA. DNA methylation β values of 485,577 CpGs and patient information were obtained from the National Cancer Institute (NCI) Genomic Data Commons Data Portal. For each cancer, data from subtypes were combined to improve statistical power and subjects of non-European ancestry and CpGs with any missing data were excluded. Then, β values were quantile-normalized within-sample and then inverse-normalized within-CpG. For each CpG, differential methylation analysis was performed by fitting a linear mixed-effects model implemented in the R package *nlme* (v 3.1.140) with tissue type (tumor/adjacent normal) modeled as a random effect, adjusting for age, sample type indicator (FFPE or not), and top three DNA methylation PCs ^30^. For analyses of colorectal, renal cell, and lung cancers, sex and cancer subtype were additionally adjusted. For lung cancer, smoking status (current/former/never) and pack-year of smoking were further adjusted. Numbers of tumor and adjacent normal tissue samples included in the final analyses were 791 vs. 97 for breast, 404 vs. 45 for colorectal, 597 vs. 205 for renal cell, 839 vs. 74 for lung, and 502 vs. 50 for prostate cancer. CpGs showing a differential expression at *P*<0.05 with directions of effect sizes consistent with Z scores of their associations with cancer risk in the main analyses were considered validated.

### Identifying CpG-gene-cancer Trios

For each cancer-associated CpG, eQTM analyses were conducted to search for potential target genes in its 500Kb flanking region using data of corresponding tissue. DNA methylation data involved in prediction model development in the present study and gene expression data downloaded from GTExPortal were utilized for these analyses. Subjects of non-European descent were also involved to improve statistical power. In total, 34 breast, 75 colon, 131 lung, 112 ovary, 44 prostate, and 25 testis tissue samples with both DNA methylation and gene expression data available were involved in this analysis. For each tissue type, genes with ≥6 read and >0.1 Transcript Per Million (TPM) were retained and TPM values were quantile-normalized within samples and then inverse-normalized within genes. Then for each CpG-gene pair, a linear regression model was fitted with inverse-normalized DNA methylation values as the exposure and inverse-normalized gene expression values as the outcome. Five DNA methylation PEERs and five gene expression PEERs were additionally adjusted. Finally, FDR correction was applied to the nominal *P* values and significant associations were identified at FDR<0.05. Such analyses could not be performed for kidney tissue due to the extremely small sample size (n=5). To address this, results of CpG-gene associations based on data of 414 normal non-neoplastic kidney tissue samples were accessed from a previous study ^31^.

For genes significantly associated with cancer-associated CpGs, we first evaluated their effects on essentiality for proliferation of corresponding cancer cells using CRISPR-Cas9 screening data. CERES values of these genes in cells relevant to breast (n=48), colon and rectum (n=57), kidney (n=32), lung (n=114), ovary (n=57), and prostate (n=10) were obtained from the DepMap Public 23Q2 release. For a particular cancer, genes with a median CERES value <-0.5 across all cells were considered essential for proliferation ^22^. Next, these genes were investigated for their genetically predicted expression in association with cancer risk. Single- and cross-tissue gene expression prediction models developed using GTEx (v8) data via elastic net and Joint-Tissue Imputation (JTI) approaches were acquired from PredictDB ^32^ and Zenodo ^33^, respectively. For each of the genes that had at least one model with R^2^>0.01 and *P*<0.05, only the model with higher R^2^ value was used in association analyses with cancer risk using SPrediXcan ^20^. Further, we examined the differential expression of these genes between tumor and normal tissues.

Results from the Gene Expression Profiling Interactive Analysis (GEPIA2) web server ^34^ were used. Data curation and analyses by the GEPIA2 team are described in detail elsewhere ^34^. Briefly, for each of 33 cancer types, raw RNA-seq data from TCGA and GTEx were processed using a uniform pipeline and differential expression analyses between TCGA tumor tissues and TCGA adjacent normal tissues plus GTEx normal tissues were conducted using the R *limma* package ^34^. For both SPrediXcan and differential expression analyses, FDR-correction was applied for each cancer separately and FDR<0.05 was used to determine significance.

Finally, to identify CpG-gene-cancer trios supporting DNA methylation of cancer-associated CpGs influencing cancer risk by modulating neighboring gene expression, we integrated results from CpG-cancer associations, CpG-gene associations, and gene-cancer relationships to evaluate the consistency of association directions. For gene-cancer relationships, we counted genes either consistently associated with cancer risk or consistently differentially expressed between tumor and normal tissues.

## Data availability

Genotype, DNA methylation, and gene expression data of GTEx participants were obtained from dbGaP (phs000424.v8.p2), GEO (GSE213478), and GTExPortal (https://www.gtexportal.org/home/), respectively. DNA methylation data of TCGA participants was acquired from NCI Genomic Data Commons Data Portal (https://portal.gdc.cancer.gov/). GTEx v8-based gene expression and splicing prediction models were downloaded from PredictDB Data Repository (https://predictdb.org/). Differential gene expression data was accessed from GEPIA2 (http://gepia2.cancer-pku.cn/#index).

## Code availability

All codes that could be used to replicate our findings, along with all DNA methylation prediction models developed in this study are available at Zenodo (https://zenodo.org/deposit/8226213).

## Acknowledgments

This analysis includes data generated by the GTEx consortium and the TCGA National Cancer Institute Program. These data were obtained from dbGaP (phs000424.v8.p2), GEO (GSE213478), and NCI Genomic Data Commons Data Portal, respectively. Y.Y. is partially supported by the NCI grant R00CA248822. This work was also supported in part by NCI grants R01CA249863 (MPIs: C.Q. and L.J.) and R01CA247987 (MPIs: L.J. and Y.F.). The funders had no role in study design, data collection and analysis, decision to publish, or preparation of the manuscript. Data analyses were conducted on the Rivanna High-Performance Computing (PHC) system at the University of Virginia and the Advanced Computing Center for Research and Education (ACCRE) HPC system at the Vanderbilt University.

## Contributions

Y.Y., L.J., and C.Q. conceived of and directed the study. Y.Y. and C.Y. performed all bioinformatics and statistical analyses, prepared tables, figures, and supplements, and wrote the manuscript, under the supervision of L.J., and C.Q. L.J., C.Q., Z.W., S.XO., Y.F., and L.L. provided valuable advice on analytical methods, results interpretation, and manuscript writing, and critically revised the original version of manuscript. X.S., W.G., X.Y, C.H., L.D. intently checked analyses pipelines and codes and finalized tables, figures, and supplements. All authors contributed essential feedback throughout the study, and reviewed and approved the final manuscript.

## References

1. Byun, J. et al. Cross-ancestry genome-wide meta-analysis of 61,047 cases and 947,237 controls identifies new susceptibility loci contributing to lung cancer. Nature genetics 54, 1167–1177 (2022).

2. Conti, D.V. et al. Trans-ancestry genome-wide association meta-analysis of prostate cancer identifies new susceptibility loci and informs genetic risk prediction. Nature genetics 53, 65–75 (2021).

3. Fernandez-Rozadilla, C. et al. Deciphering colorectal cancer genetics through multi-omic analysis of 100,204 cases and 154,587 controls of European and east Asian ancestries. Nature genetics 55, 89–99 (2023).

4. Jia, G. et al. Genome-and transcriptome-wide association studies of 386,000 Asian and European-ancestry women provide new insights into breast cancer genetics. The American Journal of Human Genetics 109, 2185–2195 (2022).

5. Phelan, C.M. et al. Identification of 12 new susceptibility loci for different histotypes of epithelial ovarian cancer. Nature genetics 49, 680–691 (2017).

6. Pluta, J. et al. Identification of 22 susceptibility loci associated with testicular germ cell tumors. Nature communications 12, 4487 (2021).

7. Scelo, G. et al. Genome-wide association study identifies multiple risk loci for renal cell carcinoma. Nature communications 8, 15724 (2017).

8. Consortium, G. The GTEx Consortium atlas of genetic regulatory effects across human tissues. Science 369, 1318–1330 (2020).

9. Lu, M. et al. TWAS atlas: a curated knowledgebase of transcriptome-wide association studies. Nucleic Acids Research 51, D1179–D1187 (2023).

10. Greenberg, M.V. & Bourc’his, D. The diverse roles of DNA methylation in mammalian development and disease. Nature reviews Molecular cell biology 20, 590–607 (2019).

11. Nishiyama, A. & Nakanishi, M. Navigating the DNA methylation landscape of cancer. Trends in Genetics 37, 1012–1027 (2021).

12. Min, J.L. et al. Genomic and phenotypic insights from an atlas of genetic effects on DNA methylation. Nature genetics 53, 1311–1321 (2021).

13. Oliva, M. et al. DNA methylation QTL mapping across diverse human tissues provides molecular links between genetic variation and complex traits. Nature genetics 55, 112–122 (2023).

14. Wu, L. et al. An integrative multi-omics analysis to identify candidate DNA methylation biomarkers related to prostate cancer risk. Nature communications 11, 3905 (2020).

15. Yang, Y. et al. Genetic Data from Nearly 63,000 Women of European Descent Predicts DNA Methylation Biomarkers and Epithelial Ovarian Cancer RiskDNA Methylation Biomarkers and Epithelial Ovarian Cancer. Cancer research 79, 505–517 (2019).

16. Yang, Y. et al. Genetically predicted levels of DNA methylation biomarkers and breast cancer risk: data from 228 951 women of European descent. JNCI: Journal of the National Cancer Institute 112, 295–304 (2020).

17. Zhu, J. et al. Integrating genome and methylome data to identify candidate DNA methylation biomarkers for pancreatic cancer risk. *Cancer Epidemiology*, Biomarkers & Prevention 30, 2079–2087 (2021).

18. Gamazon, E.R. et al. A gene-based association method for mapping traits using reference transcriptome data. Nature genetics 47, 1091–1098 (2015).

19. Hu, Y. et al. A statistical framework for cross-tissue transcriptome-wide association analysis. Nature genetics 51, 568–576 (2019).

20. Barbeira, A.N. et al. Exploring the phenotypic consequences of tissue specific gene expression variation inferred from GWAS summary statistics. Nature communications 9, 1825 (2018).

21. Gao, G. et al. A joint transcriptome-wide association study across multiple tissues identifies candidate breast cancer susceptibility genes. The American Journal of Human Genetics 110, 950–962 (2023).

22. Meyers, R.M. et al. Computational correction of copy number effect improves specificity of CRISPR–Cas9 essentiality screens in cancer cells. Nature genetics 49, 1779–1784 (2017).

23. Zurlo, G. et al. Prolyl hydroxylase substrate adenylosuccinate lyase is an oncogenic driver in triple negative breast cancer. Nature Communications 10, 5177 (2019).

24. Seshacharyulu, P. et al. FDPS cooperates with PTEN loss to promote prostate cancer progression through modulation of small GTPases/AKT axis. Oncogene 38, 5265–5280 (2019).

25. Tian, Y. et al. ChAMP: updated methylation analysis pipeline for Illumina BeadChips. Bioinformatics 33, 3982–3984 (2017).

26. Fortin, J.-P., Triche Jr, T.J. & Hansen, K.D. Preprocessing, normalization and integration of the Illumina HumanMethylationEPIC array with minfi. Bioinformatics 33, 558–560 (2017).

27. Stegle, O., Parts, L., Piipari, M., Winn, J. & Durbin, R. Using probabilistic estimation of expression residuals (PEER) to obtain increased power and interpretability of gene expression analyses. Nature protocols 7, 500–507 (2012).

28. Yang, J. et al. Conditional and joint multiple-SNP analysis of GWAS summary statistics identifies additional variants influencing complex traits. Nature genetics 44, 369–375 (2012).

29. Breeze, C.E. et al. eFORGE v2. 0: updated analysis of cell type-specific signal in epigenomic data. Bioinformatics 35, 4767–4769 (2019).

30. Zhou, H.J., Li, L., Li, Y., Li, W. & Li, J.J. PCA outperforms popular hidden variable inference methods for molecular QTL mapping. Genome biology 23, 1–17 (2022).

31. Liu, H. et al. Epigenomic and transcriptomic analyses define core cell types, genes and targetable mechanisms for kidney disease. Nature Genetics 54, 950–962 (2022).

32. Barbeira, A.N. et al. Exploiting the GTEx resources to decipher the mechanisms at GWAS loci. Genome biology 22, 1–24 (2021).

33. Zhou, D. et al. A unified framework for joint-tissue transcriptome-wide association and Mendelian randomization analysis. Nature genetics 52, 1239–1246 (2020).

34. Tang, Z., Kang, B., Li, C., Chen, T. & Zhang, Z. GEPIA2: an enhanced web server for large-scale expression profiling and interactive analysis. Nucleic acids research 47, W556–W560 (2019).

