## Supplementary Figures 1-4 for "Integrating genome and epigenome data to identify tissue-specific DNA methylation biomarkers for cancer risk"

**Table of Contents**

**Supplementary Figure Legends**

**Supplementary Figure 1. Tissue-specificity of cancer-associated CpGs.**

**Supplementary Figure 2. Cell type-specific enrichment with functional genomic regions of cancer-associated CpGs.**

**Supplementary Figure 3. Essential roles in cell proliferation of genes associated with cancer-associated CpGs.**

**Supplementary Figure 4. Examples of CpG-gene-cancer trios suggesting DNA methylation at CpGs influencing cancer risk by regulating nearby gene expression.**

#### Supplementary Figure Legends

**Supplementary Figure 1. Tissue-specificity of cancer-associated CpGs.** For each cancer, significant associations were identified at Bonferroni-corrected $P<0.05$. CpGs that were exclusively associated with a particular cancer are defined as tissue specific.

**Supplementary Figure 2. Cell type-specific enrichment with functional genomic regions of cancer susceptibility CpGs.** Analyses were conducted and plots were generated by eFORGE (v2.0). For each cancer, CpGs significantly associated with cancer risk at Bonferroni-corrected $P<0.05$ were tested for enrichments in DNase I hypersensitive sites (DHS), 15 chromatin states, and five histone marks. In all plots, hollow circles, except for those blue ones, denote significant enrichments at false-discovery rate (FDR)<0.05 while all solid circles denote significant enrichment at FDR<0.01.

**Supplementary Figure 3. Essential roles in cell proliferation of genes associated with cancer-associated CpGs.** Boxplots show effects of genes on cell proliferation using experimental data from CRISPR screens (see **Methods**). The dashed red line in each plot denotes the significant threshold of median CERES values<-0.5. Genes shown in CpG-gene-cancer trios revealed by downstream analyses (see **Methods**) are highlighted in red.

**Supplementary Figure 4. Examples of CpG-gene-cancer trios suggesting DNA methylation influencing cancer risk by regulating nearby gene expression.** Red arrows, lines, and blocks indicate positive associations, while green ones indicate negative associations. In boxplots, red and green boxes represent data of tumor and adjacent normal tissues, respectively. DNAm, DNA methylation; GEx, gene expression; OR, odds ratio; CI, confidence interval; TCGA, The Cancer Genome Atlas; PRAD, prostate adenocarcinoma; TGCT, testicular germ cell tumors; KIRC, kidney renal clear cell carcinoma; KIRP, kidney renal papillary cell carcinoma. **A**, DNA methylation at cg17397364 may increase prostate cancer risk by promoting the expression of *TMEM17*. **B**, DNA methylation at cg22340370 may increase testicular germ cell cancer risk by suppressing the expression of *MRM2*. **C**, DNA methylation at cg06511653 may decrease renal cell cancer risk by suppressing the expression of *SSPN*.

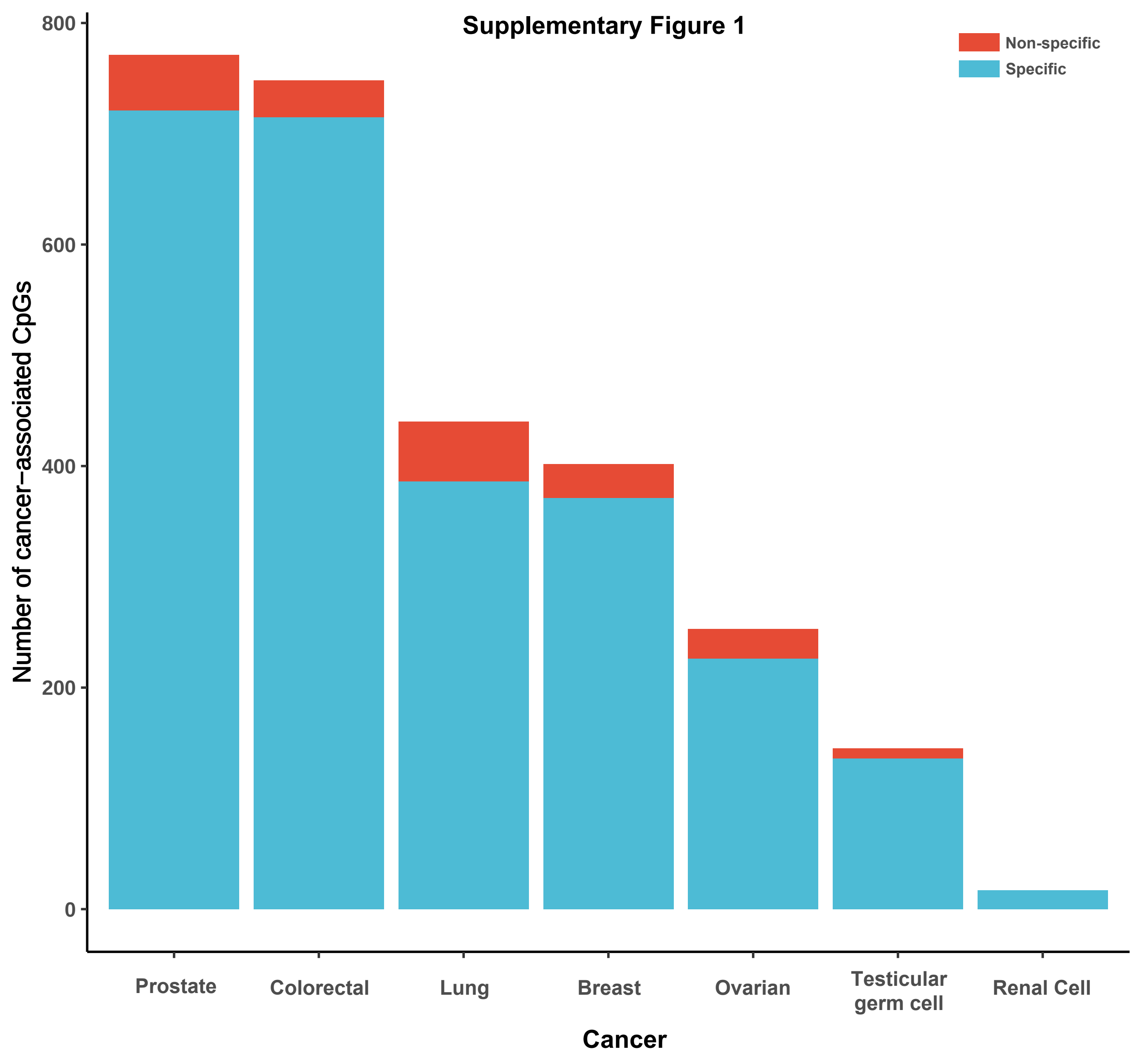
Supplementary Figure 1
Non-specific
Specific
Number of cancer-associated CpGs
800
600
400
200
0
Prostate
Colorectal
Lung
Breast
Ovarian
Testicular germ cell
Renal Cell
Cancer

Supplementary Figure 2

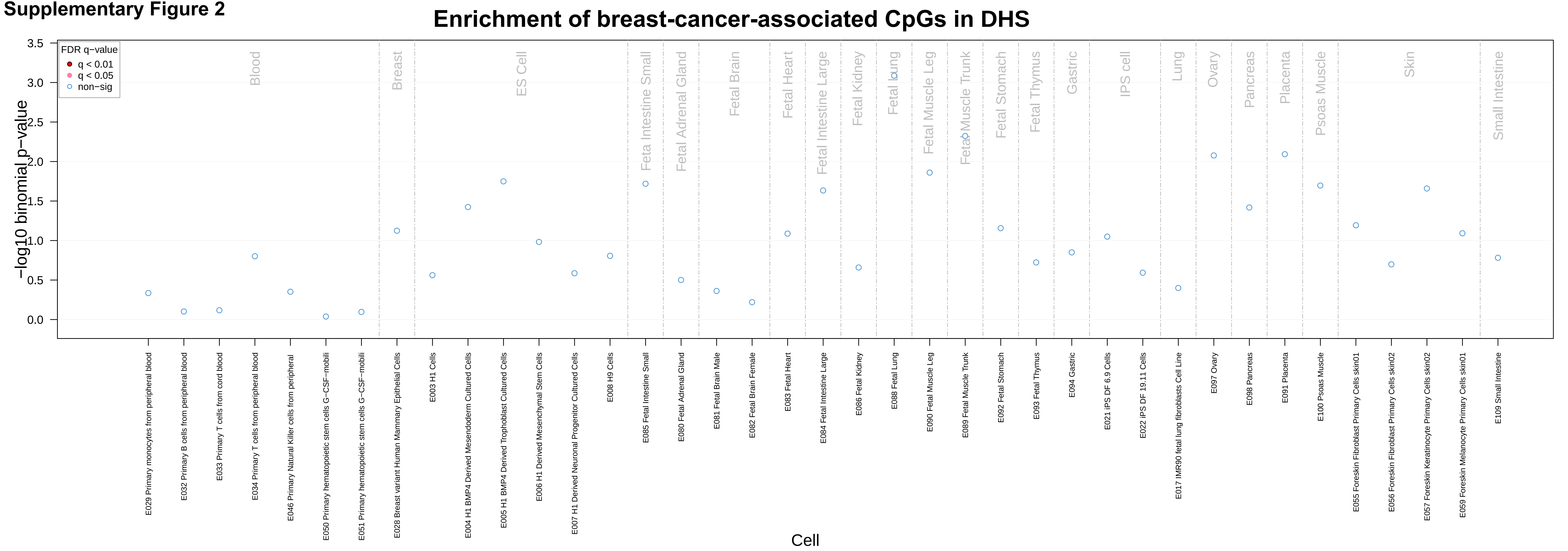

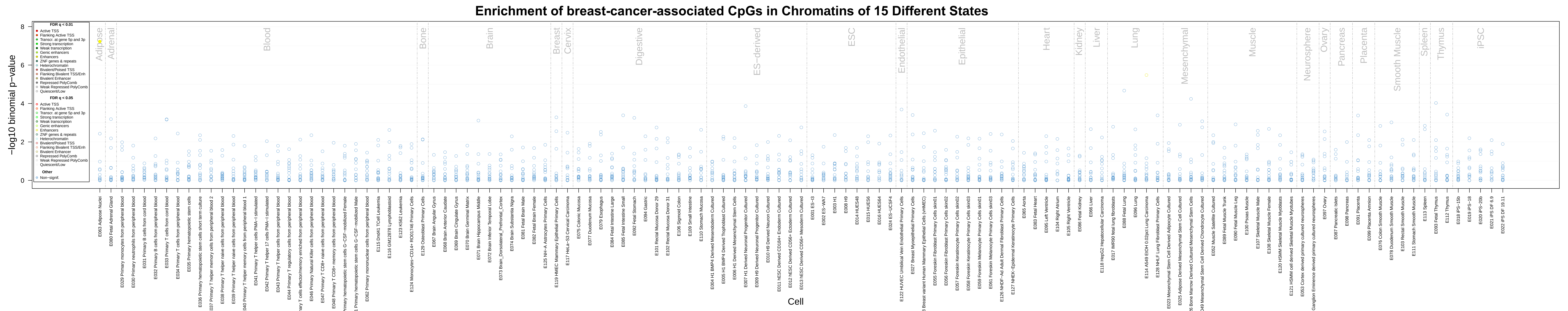

Enrichment of breast-cancer-associated CpGs in Chromatins of 15 Different States
−log10 binomial p−value
Cell
FDR q < 0.01
FDR q < 0.05
Other
Non−signif.
Adipose
Adrenal
Blood
Bone
Brain
Breast
Cervix
Digestive
ES−deriv
ESC
Endothelial
Epithelial
Heart
Kidney
Liver
Lung
Mesenchymal
Muscle
Neurosphere
Ovary
Pancreas
Placenta
Smooth Muscle
Spleen
Thymus
iPSC

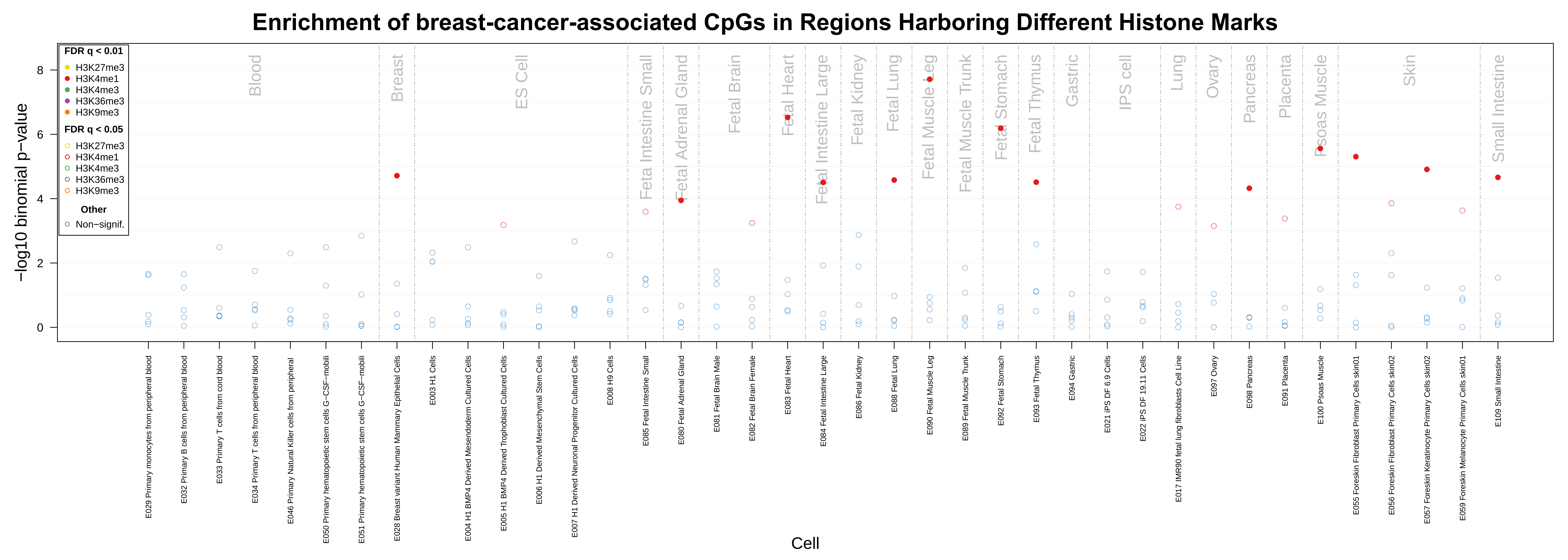
Enrichment of breast-cancer-associated CpGs in Regions Harboring Different Histone Marks
FDR q < 0.01
H3K27me3
H3K4me1
H3K4me3
H3K36me3
H3K9me3
FDR q < 0.05
H3K27me3
H3K4me1
H3K4me3
H3K36me3
H3K9me3
Other
Non−signif.
−log10 binomial p−value
0
2
4
6
8
Blood
Breast
ES Cell
Feta Intestine Small
Fetal Adrenal Gland
Fetal Brain
Fetal Heart
Fetal Intestine Large
Fetal Kidney
Fetal Lung
Fetal Muscle Leg
Fetal Muscle Trunk
Fetal Stomach
Fetal Thymus
Gastric
IPS cell
Lung
Ovary
Pancreas
Placenta
Psoas Muscle
Skin
Small Intestine
E029 Primary monocytes from peripheral blood
E032 Primary B cells from peripheral blood
E033 Primary T cells from cord blood
E034 Primary T cells from peripheral blood
E046 Primary Natural Killer cells from peripheral
E050 Primary hematopoietic stem cells G−CSF−mobili
E051 Primary hematopoietic stem cells G−CSF−mobili
E028 Breast variant Human Mammary Epithelial Cells
E003 H1 Cells
E004 H1 BMP4 Derived Mesendoderm Cultured Cells
E005 H1 BMP4 Derived Trophoblast Cultured Cells
E006 H1 Derived Mesenchymal Stem Cells
E007 H1 Derived Neuronal Progenitor Cultured Cells
E008 H9 Cells
E085 Fetal Intestine Small
E080 Fetal Adrenal Gland
E081 Fetal Brain Male
E082 Fetal Brain Female
E083 Fetal Heart
E084 Fetal Intestine Large
E086 Fetal Kidney
E088 Fetal Lung
E090 Fetal Muscle Leg
E089 Fetal Muscle Trunk
E092 Fetal Stomach
E093 Fetal Thymus
E094 Gastric
E021 iPS DF 6.9 Cells
E022 iPS DF 19.11 Cells
E017 IMR90 fetal lung fibroblasts Cell Line
E097 Ovary
E098 Pancreas
E091 Placenta
E100 Psoas Muscle
E055 Foreskin Fibroblast Primary Cells skin01
E056 Foreskin Fibroblast Primary Cells skin02
E057 Foreskin Keratinocyte Primary Cells skin02
E059 Foreskin Melanocyte Primary Cells skin01
E109 Small Intestine
Cell

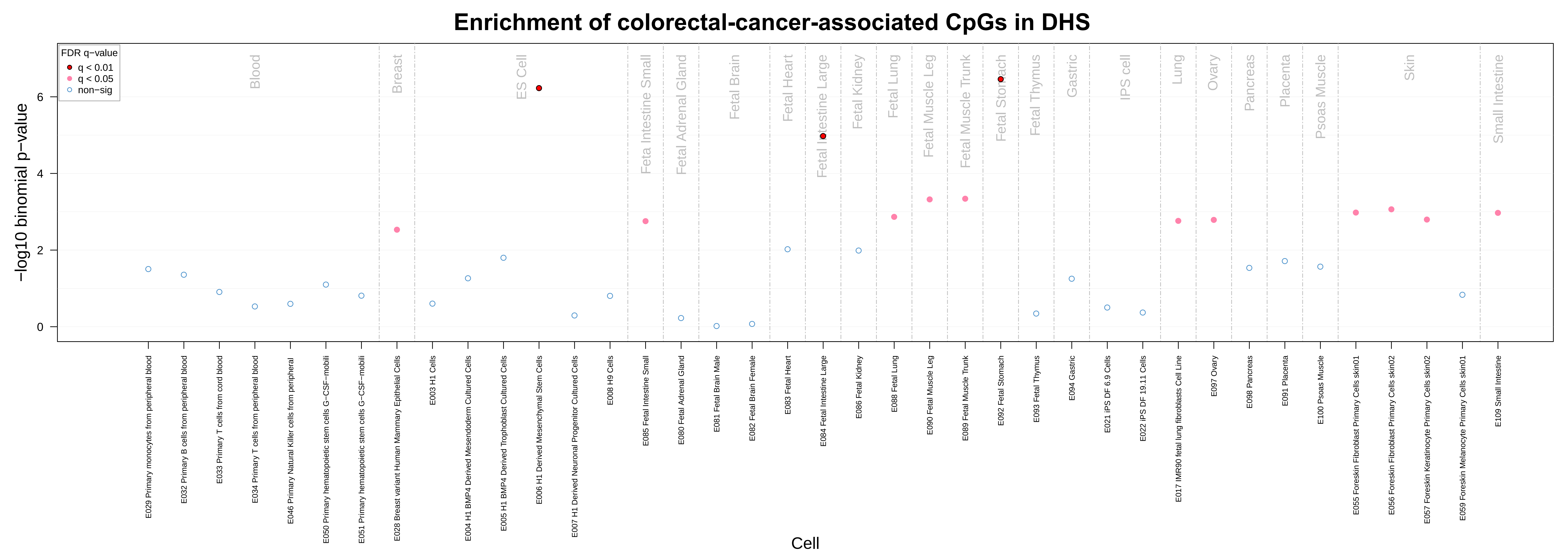

Enrichment of colorectal-cancer-associated CpGs in DHS
FDR q−value
q < 0.01
q < 0.05
non−sig
−log10 binomial p−value
Cell
Blood
Breast
ES Cell
Feta Intestine Small
Fetal Adrenal Gland
Fetal Brain
Fetal Heart
Fetal Intestine Large
Fetal Kidney
Fetal Lung
Fetal Muscle Leg
Fetal Muscle Trunk
Fetal Stomach
Fetal Thymus
Gastric
IPS cell
Lung
Ovary
Pancreas
Placenta
Psoas Muscle
Skin
Small Intestine
E029 Primary monocytes from peripheral blood
E032 Primary B cells from peripheral blood
E033 Primary T cells from cord blood
E034 Primary T cells from peripheral blood
E046 Primary Natural Killer cells from peripheral
E050 Primary hematopoietic stem cells G−CSF−mobili
E051 Primary hematopoietic stem cells G−CSF−mobili
E028 Breast variant Human Mammary Epithelial Cells
E003 H1 Cells
E004 H1 BMP4 Derived Mesendoderm Cultured Cells
E005 H1 BMP4 Derived Trophoblast Cultured Cells
E006 H1 Derived Mesenchymal Stem Cells
E007 H1 Derived Neuronal Progenitor Cultured Cells
E008 H9 Cells
E085 Fetal Intestine Small
E080 Fetal Adrenal Gland
E081 Fetal Brain Male
E082 Fetal Brain Female
E083 Fetal Heart
E084 Fetal Intestine Large
E086 Fetal Kidney
E088 Fetal Lung
E090 Fetal Muscle Leg
E089 Fetal Muscle Trunk
E092 Fetal Stomach
E093 Fetal Thymus
E094 Gastric
E021 iPS DF 6.9 Cells
E022 iPS DF 19.11 Cells
E017 IMR90 fetal lung fibroblasts Cell Line
E097 Ovary
E098 Pancreas
E091 Placenta
E100 Psoas Muscle
E055 Foreskin Fibroblast Primary Cells skin01
E056 Foreskin Fibroblast Primary Cells skin02
E057 Foreskin Keratinocyte Primary Cells skin02
E059 Foreskin Melanocyte Primary Cells skin01
E109 Small Intestine

**Enrichment of colorectal-cancer-associated CpGs in Chromatins of 15 Different States**

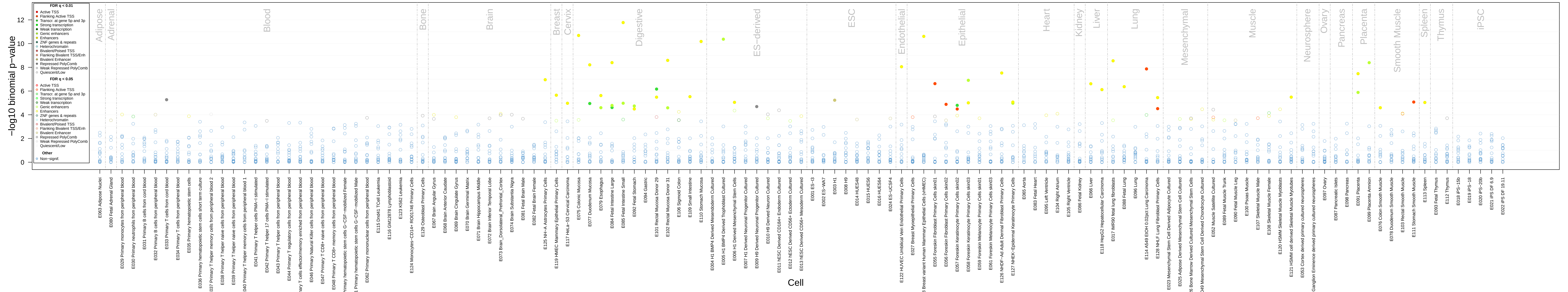

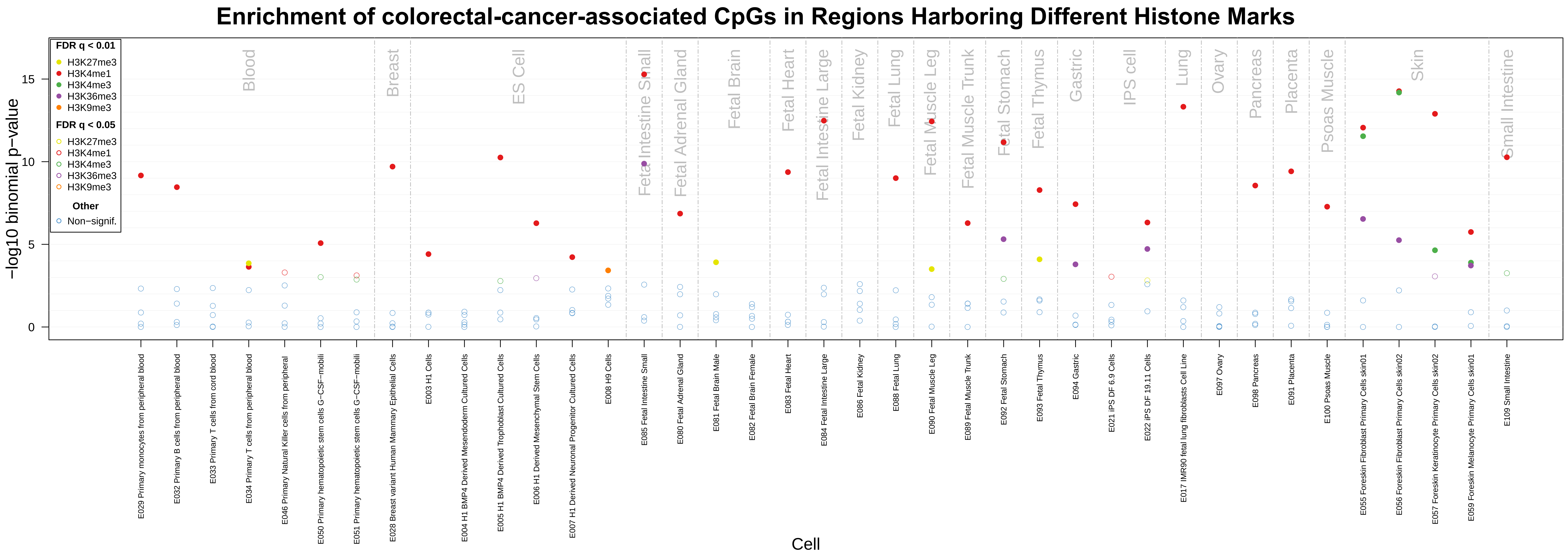

Enrichment of colorectal-cancer-associated CpGs in Regions Harboring Different Histone Marks
FDR q < 0.01
H3K27me3
H3K4me1
H3K4me3
H3K36me3
H3K9me3
FDR q < 0.05
H3K27me3
H3K4me1
H3K4me3
H3K36me3
H3K9me3
Other
Non−signif.
−log10 binomial p−value
0
5
10
15
Blood
Breast
ES Cell
Fetal Intestine Small
Fetal Adrenal Gland
Fetal Brain
Fetal Heart
Fetal Intestine Large
Fetal Kidney
Fetal Lung
Fetal Muscle Leg
Fetal Muscle Trunk
Fetal Stomach
Fetal Thymus
Gastric
IPS cell
Lung
Ovary
Pancreas
Placenta
Psoas Muscle
Skin
Small Intestine
E029 Primary monocytes from peripheral blood
E032 Primary B cells from peripheral blood
E033 Primary T cells from cord blood
E034 Primary T cells from peripheral blood
E046 Primary Natural Killer cells from peripheral
E050 Primary hematopoietic stem cells G−CSF−mobili
E051 Primary hematopoietic stem cells G−CSF−mobili
E028 Breast variant Human Mammary Epithelial Cells
E003 H1 Cells
E004 H1 BMP4 Derived Mesendoderm Cultured Cells
E005 H1 BMP4 Derived Trophoblast Cultured Cells
E006 H1 Derived Mesenchymal Stem Cells
E007 H1 Derived Neuronal Progenitor Cultured Cells
E008 H9 Cells
E085 Fetal Intestine Small
E080 Fetal Adrenal Gland
E081 Fetal Brain Male
E082 Fetal Brain Female
E083 Fetal Heart
E084 Fetal Intestine Large
E086 Fetal Kidney
E088 Fetal Lung
E090 Fetal Muscle Leg
E089 Fetal Muscle Trunk
E092 Fetal Stomach
E093 Fetal Thymus
E094 Gastric
E021 iPS DF 6.9 Cells
E022 iPS DF 19.11 Cells
E017 IMR90 fetal lung fibroblasts Cell Line
E097 Ovary
E098 Pancreas
E091 Placenta
E100 Psoas Muscle
E055 Foreskin Fibroblast Primary Cells skin01
E056 Foreskin Fibroblast Primary Cells skin02
E057 Foreskin Keratinocyte Primary Cells skin02
E059 Foreskin Melanocyte Primary Cells skin01
E109 Small Intestine
Cell

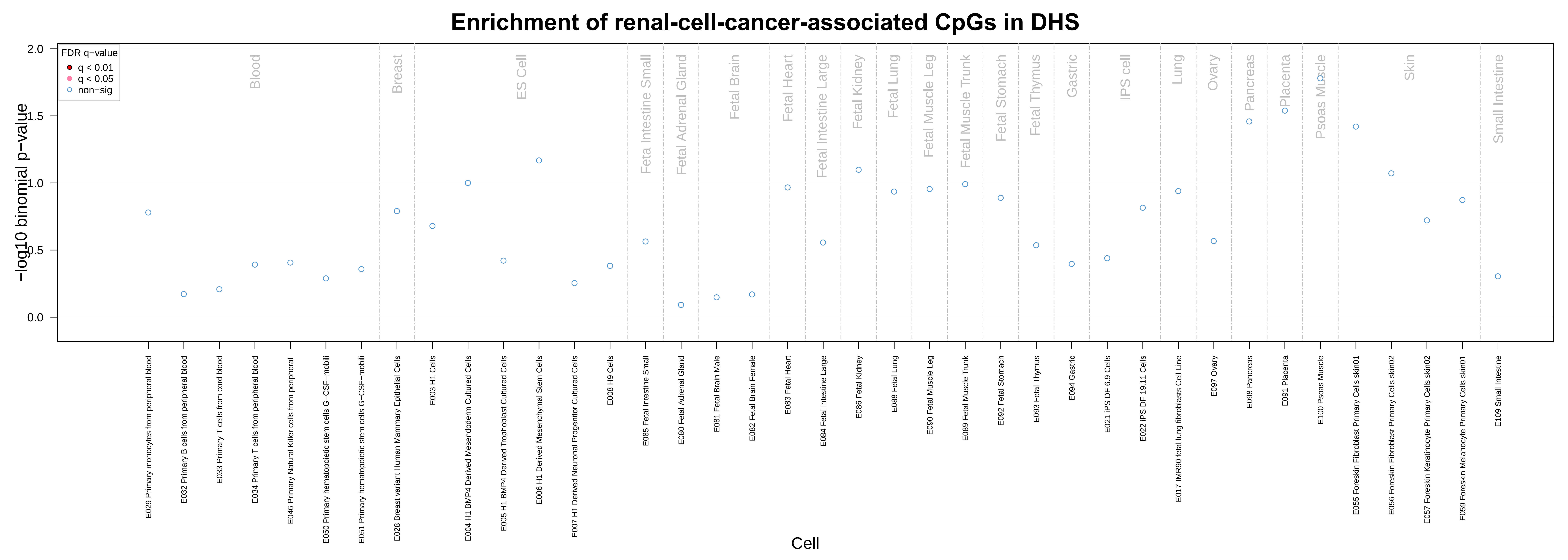
Enrichment of renal-cell-cancer-associated CpGs in DHS
FDR q−value
q < 0.01
q < 0.05
non−sig
−log10 binomial p−value
0.0
0.5
1.0
1.5
2.0
Blood
Breast
ES Cell
Feta Intestine Small
Fetal Adrenal Gland
Fetal Brain
Fetal Heart
Fetal Intestine Large
Fetal Kidney
Fetal Lung
Fetal Muscle Leg
Fetal Muscle Trunk
Fetal Stomach
Fetal Thymus
Gastric
IPS cell
Lung
Ovary
Pancreas
Placenta
Psoas Muscle
Skin
Small Intestine
E029 Primary monocytes from peripheral blood
E032 Primary B cells from peripheral blood
E033 Primary T cells from cord blood
E034 Primary T cells from peripheral blood
E046 Primary Natural Killer cells from peripheral
E050 Primary hematopoietic stem cells G−CSF−mobili
E051 Primary hematopoietic stem cells G−CSF−mobili
E028 Breast variant Human Mammary Epithelial Cells
E003 H1 Cells
E004 H1 BMP4 Derived Mesendoderm Cultured Cells
E005 H1 BMP4 Derived Trophoblast Cultured Cells
E006 H1 Derived Mesenchymal Stem Cells
E007 H1 Derived Neuronal Progenitor Cultured Cells
E008 H9 Cells
E085 Fetal Intestine Small
E080 Fetal Adrenal Gland
E081 Fetal Brain Male
E082 Fetal Brain Female
E083 Fetal Heart
E084 Fetal Intestine Large
E086 Fetal Kidney
E088 Fetal Lung
E090 Fetal Muscle Leg
E089 Fetal Muscle Trunk
E092 Fetal Stomach
E093 Fetal Thymus
E094 Gastric
E021 iPS DF 6.9 Cells
E022 iPS DF 19.11 Cells
E017 IMR90 fetal lung fibroblasts Cell Line
E097 Ovary
E098 Pancreas
E091 Placenta
E100 Psoas Muscle
E055 Foreskin Fibroblast Primary Cells skin01
E056 Foreskin Fibroblast Primary Cells skin02
E057 Foreskin Keratinocyte Primary Cells skin02
E059 Foreskin Melanocyte Primary Cells skin01
E109 Small Intestine
Cell

#### Enrichment of renal-cell-cancer-associated CpGs in Chromatins of 15 Different States

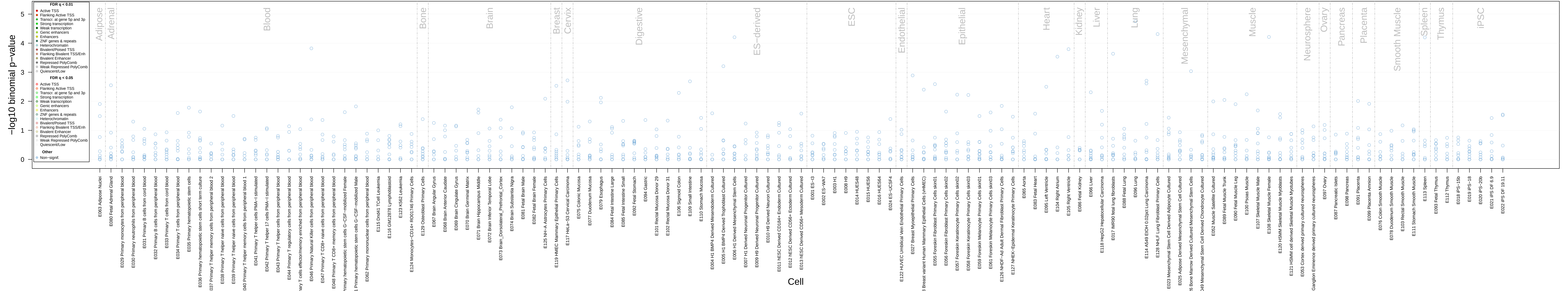

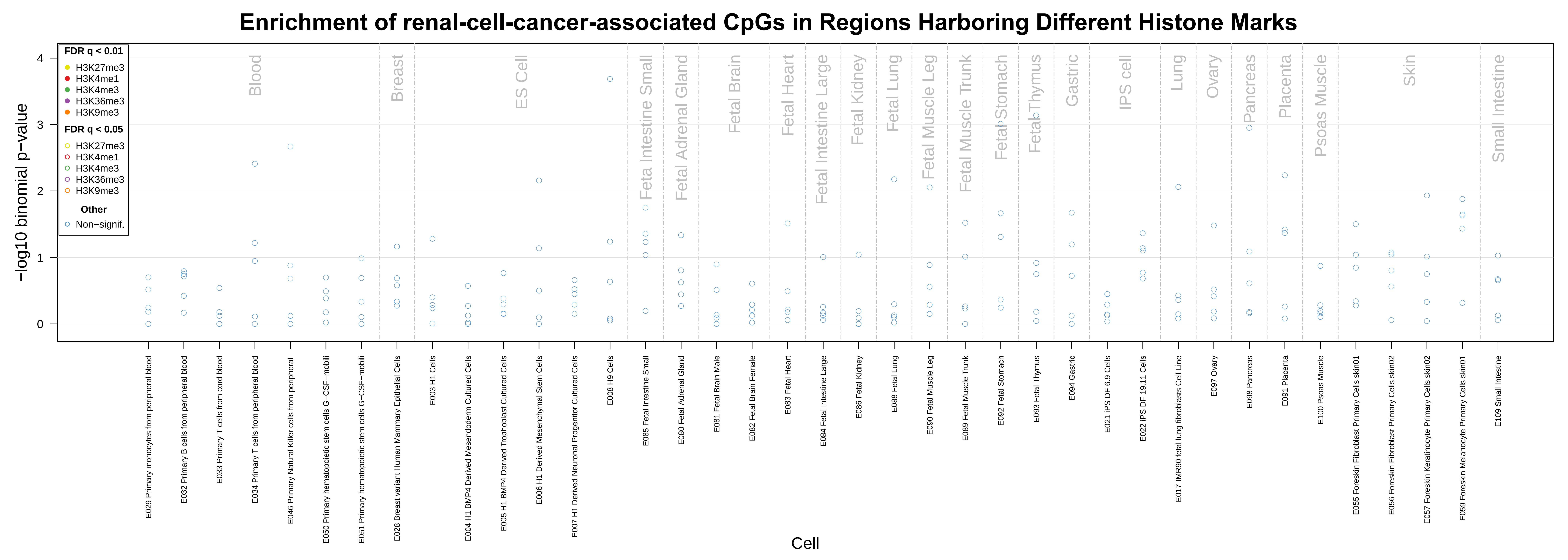

Enrichment of renal-cell-cancer-associated CpGs in Regions Harboring Different Histone Marks
−log10 binomial p−value
Cell
FDR q < 0.01
H3K27me3
H3K4me1
H3K4me3
H3K36me3
H3K9me3
FDR q < 0.05
H3K27me3
H3K4me1
H3K4me3
H3K36me3
H3K9me3
Other
Non−signif.
Blood
Breast
ES Cell
Feta Intestine Small
Fetal Adrenal Gland
Fetal Brain
Fetal Heart
Fetal Intestine Large
Fetal Kidney
Fetal Lung
Fetal Muscle Leg
Fetal Muscle Trunk
Fetal Stomach
Fetal Thymus
Gastric
IPS cell
Lung
Ovary
Pancreas
Placenta
Psoas Muscle
Skin
Small Intestine
E029 Primary monocytes from peripheral blood
E032 Primary B cells from peripheral blood
E033 Primary T cells from cord blood
E034 Primary T cells from peripheral blood
E046 Primary Natural Killer cells from peripheral
E050 Primary hematopoietic stem cells G−CSF−mobili
E051 Primary hematopoietic stem cells G−CSF−mobili
E028 Breast variant Human Mammary Epithelial Cells
E003 H1 Cells
E004 H1 BMP4 Derived Mesendoderm Cultured Cells
E005 H1 BMP4 Derived Trophoblast Cultured Cells
E006 H1 Derived Mesenchymal Stem Cells
E007 H1 Derived Neuronal Progenitor Cultured Cells
E008 H9 Cells
E085 Fetal Intestine Small
E080 Fetal Adrenal Gland
E081 Fetal Brain Male
E082 Fetal Brain Female
E083 Fetal Heart
E084 Fetal Intestine Large
E086 Fetal Kidney
E088 Fetal Lung
E090 Fetal Muscle Leg
E089 Fetal Muscle Trunk
E092 Fetal Stomach
E093 Fetal Thymus
E094 Gastric
E021 iPS DF 6.9 Cells
E022 iPS DF 19.11 Cells
E017 IMR90 fetal lung fibroblasts Cell Line
E097 Ovary
E098 Pancreas
E091 Placenta
E100 Psoas Muscle
E055 Foreskin Fibroblast Primary Cells skin01
E056 Foreskin Fibroblast Primary Cells skin02
E057 Foreskin Keratinocyte Primary Cells skin02
E059 Foreskin Melanocyte Primary Cells skin01
E109 Small Intestine

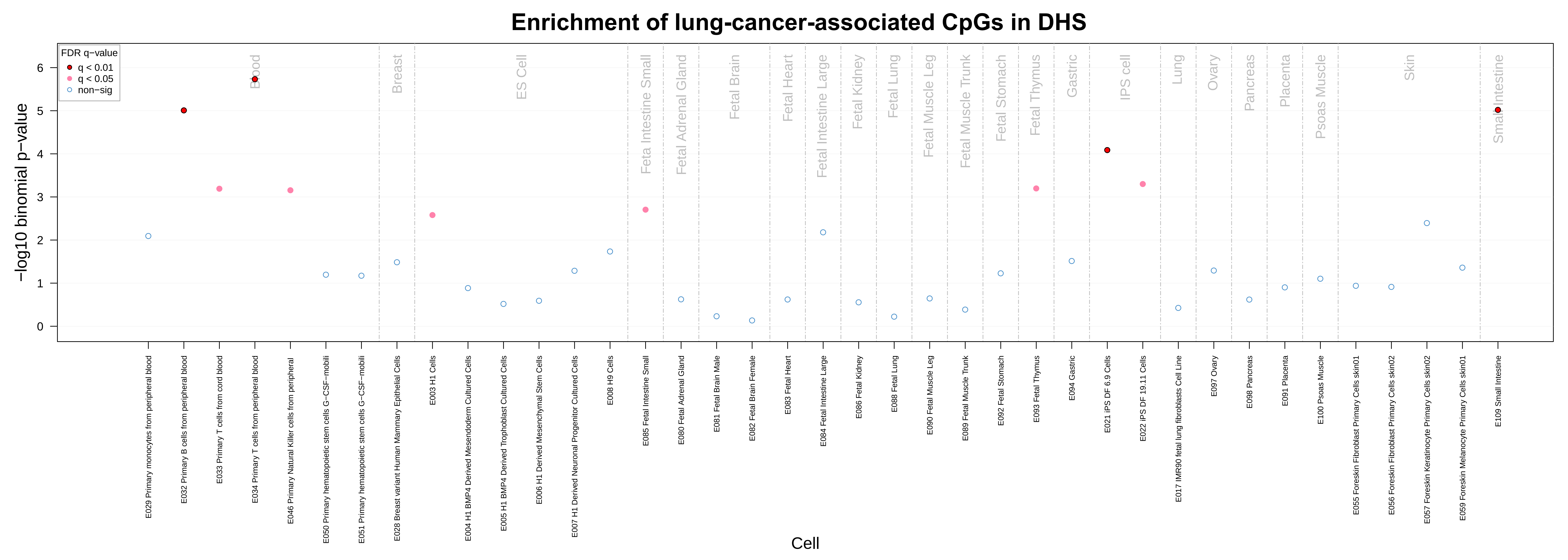

Enrichment of lung-cancer-associated CpGs in DHS
FDR q−value
q < 0.01
q < 0.05
non−sig
−log10 binomial p−value
0
1
2
3
4
5
6
Blood
Breast
ES Cell
Feta Intestine Small
Fetal Adrenal Gland
Fetal Brain
Fetal Heart
Fetal Intestine Large
Fetal Kidney
Fetal Lung
Fetal Muscle Leg
Fetal Muscle Trunk
Fetal Stomach
Fetal Thymus
Gastric
IPS cell
Lung
Ovary
Pancreas
Placenta
Psoas Muscle
Skin
Small Intestine
E029 Primary monocytes from peripheral blood
E032 Primary B cells from peripheral blood
E033 Primary T cells from cord blood
E034 Primary T cells from peripheral blood
E046 Primary Natural Killer cells from peripheral
E050 Primary hematopoietic stem cells G−CSF−mobili
E051 Primary hematopoietic stem cells G−CSF−mobili
E028 Breast variant Human Mammary Epithelial Cells
E003 H1 Cells
E004 H1 BMP4 Derived Mesendoderm Cultured Cells
E005 H1 BMP4 Derived Trophoblast Cultured Cells
E006 H1 Derived Mesenchymal Stem Cells
E007 H1 Derived Neuronal Progenitor Cultured Cells
E008 H9 Cells
E085 Fetal Intestine Small
E080 Fetal Adrenal Gland
E081 Fetal Brain Male
E082 Fetal Brain Female
E083 Fetal Heart
E084 Fetal Intestine Large
E086 Fetal Kidney
E088 Fetal Lung
E090 Fetal Muscle Leg
E089 Fetal Muscle Trunk
E092 Fetal Stomach
E093 Fetal Thymus
E094 Gastric
E021 iPS DF 6.9 Cells
E022 iPS DF 19.11 Cells
E017 IMR90 fetal lung fibroblasts Cell Line
E097 Ovary
E098 Pancreas
E091 Placenta
E100 Psoas Muscle
E055 Foreskin Fibroblast Primary Cells skin01
E056 Foreskin Fibroblast Primary Cells skin02
E057 Foreskin Keratinocyte Primary Cells skin02
E059 Foreskin Melanocyte Primary Cells skin01
E109 Small Intestine
Cell

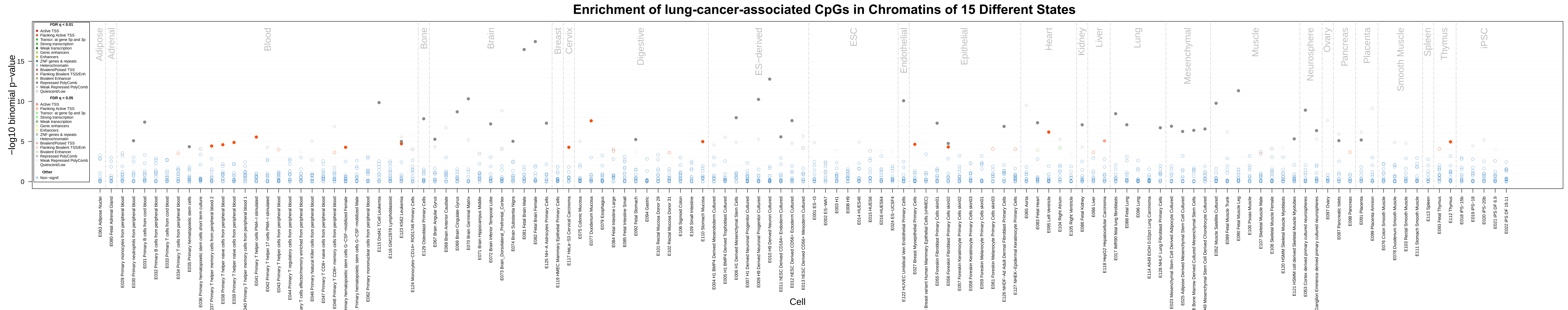
Enrichment of lung-cancer-associated CpGs in Chromatins of 15 Different States
−log10 binomial p−value
Cell
FDR q < 0.01
Active TSS
Flanking Active TSS
Transcr. at gene 5p and 3p
Strong transcription
Weak transcription
Genic enhancers
Enhancers
ZNF genes & repeats
Heterochromatin
Bivalent/Poised TSS
Flanking Bivalent TSS/Enh
Bivalent Enhancer
Repressed PolyComb
Weak Repressed PolyComb
Quiescent/Low
FDR q < 0.05
Other
Non−signif.
Adipose
Adrenal
Blood
Bone
Brain
Breast
Cervix
Digestive
ES−deriv
ESC
Endothelial
Epithelial
Heart
Kidney
Liver
Lung
Mesenchymal
Muscle
Neurosphere
Ovary
Pancreas
Placenta
Smooth Muscle
Spleen
Thymus
iPSC

#### Enrichment of lung-cancer-associated CpGs in Regions Harboring Different Histone Marks

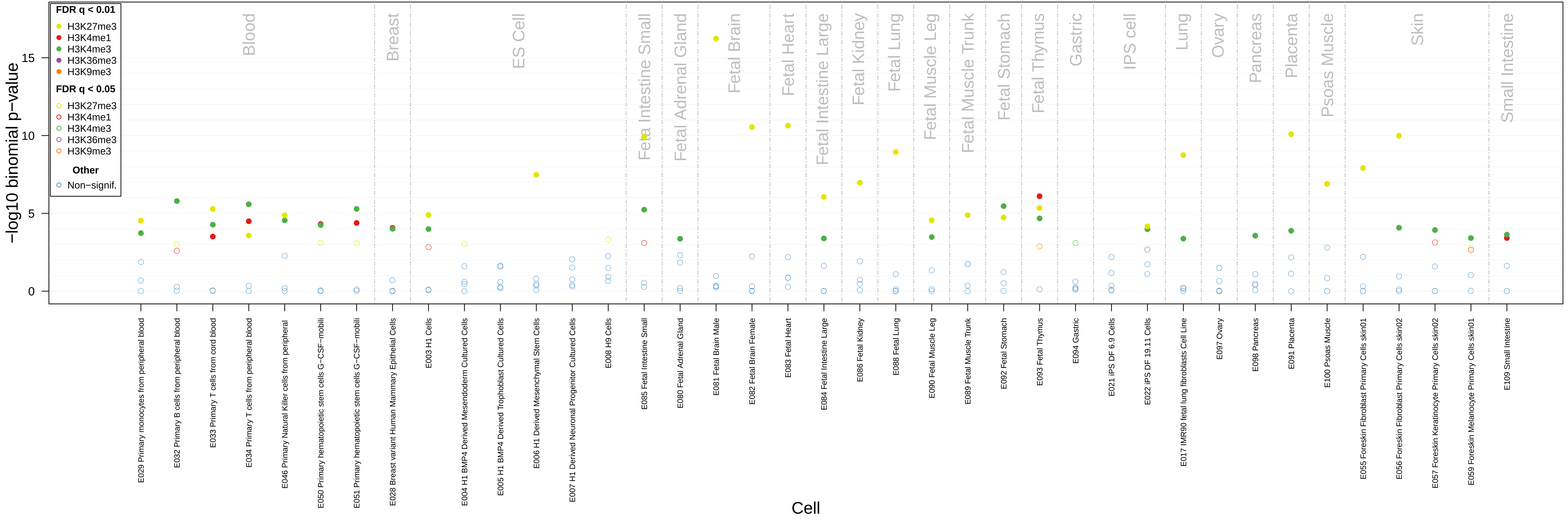

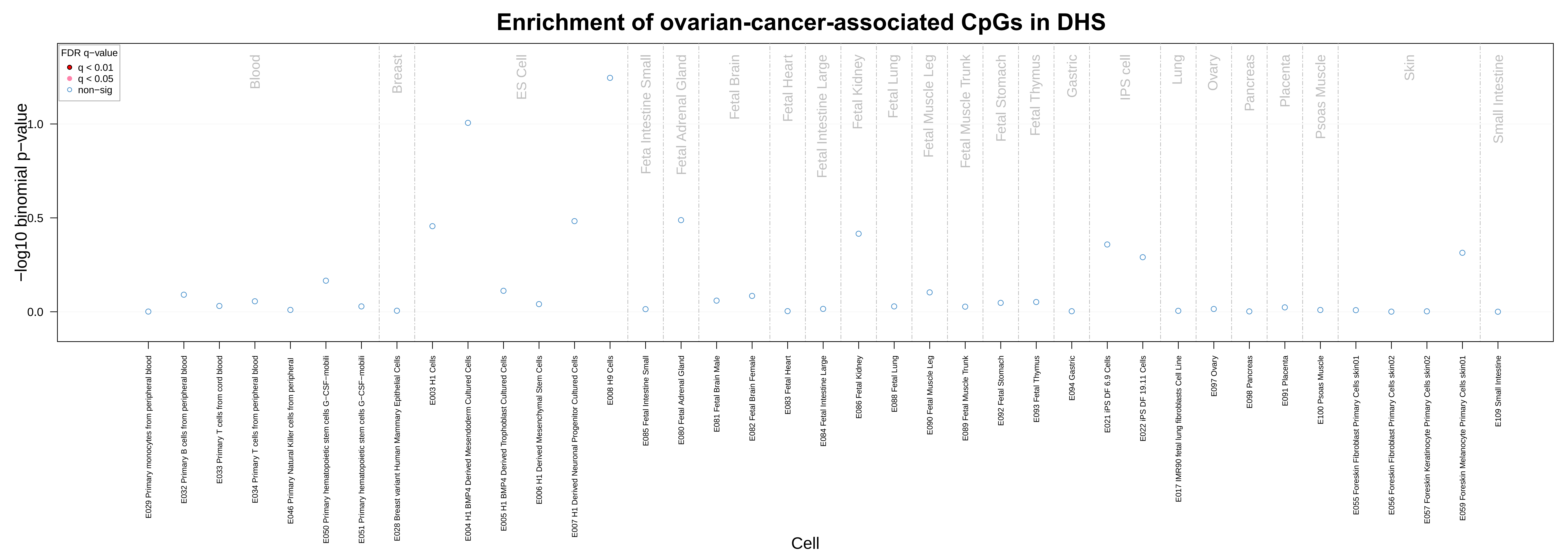
Enrichment of ovarian-cancer-associated CpGs in DHS
FDR q−value
q < 0.01
q < 0.05
non−sig
−log10 binomial p−value
0.0
0.5
1.0
Blood
Breast
ES Cell
Feta Intestine Small
Fetal Adrenal Gland
Fetal Brain
Fetal Heart
Fetal Intestine Large
Fetal Kidney
Fetal Lung
Fetal Muscle Leg
Fetal Muscle Trunk
Fetal Stomach
Fetal Thymus
Gastric
IPS cell
Lung
Ovary
Pancreas
Placenta
Psoas Muscle
Skin
Small Intestine
E029 Primary monocytes from peripheral blood
E032 Primary B cells from peripheral blood
E033 Primary T cells from cord blood
E034 Primary T cells from peripheral blood
E046 Primary Natural Killer cells from peripheral
E050 Primary hematopoietic stem cells G−CSF−mobili
E051 Primary hematopoietic stem cells G−CSF−mobili
E028 Breast variant Human Mammary Epithelial Cells
E003 H1 Cells
E004 H1 BMP4 Derived Mesendoderm Cultured Cells
E005 H1 BMP4 Derived Trophoblast Cultured Cells
E006 H1 Derived Mesenchymal Stem Cells
E007 H1 Derived Neuronal Progenitor Cultured Cells
E008 H9 Cells
E085 Fetal Intestine Small
E080 Fetal Adrenal Gland
E081 Fetal Brain Male
E082 Fetal Brain Female
E083 Fetal Heart
E084 Fetal Intestine Large
E086 Fetal Kidney
E088 Fetal Lung
E090 Fetal Muscle Leg
E089 Fetal Muscle Trunk
E092 Fetal Stomach
E093 Fetal Thymus
E094 Gastric
E021 iPS DF 6.9 Cells
E022 iPS DF 19.11 Cells
E017 IMR90 fetal lung fibroblasts Cell Line
E097 Ovary
E098 Pancreas
E091 Placenta
E100 Psoas Muscle
E055 Foreskin Fibroblast Primary Cells skin01
E056 Foreskin Fibroblast Primary Cells skin02
E057 Foreskin Keratinocyte Primary Cells skin02
E059 Foreskin Melanocyte Primary Cells skin01
E109 Small Intestine
Cell

### Enrichment of ovarian-cancer-associated CpGs in Chromatins of 15 Different States

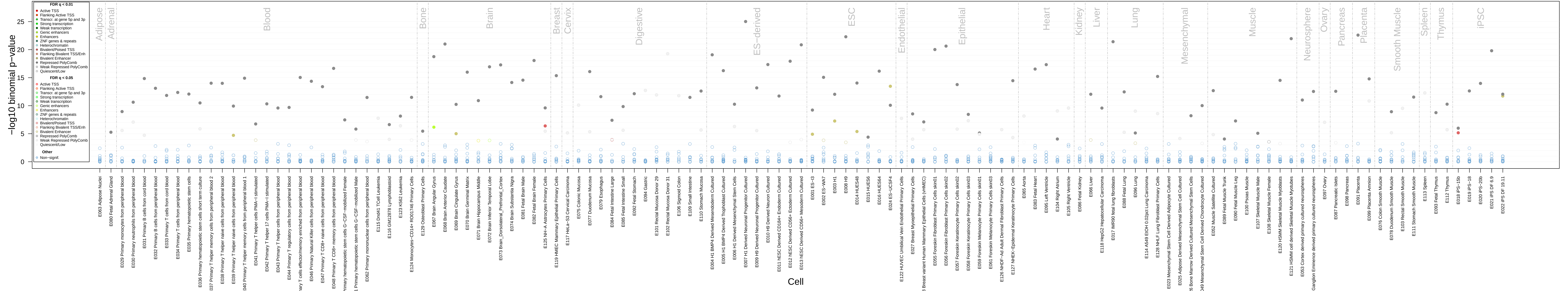

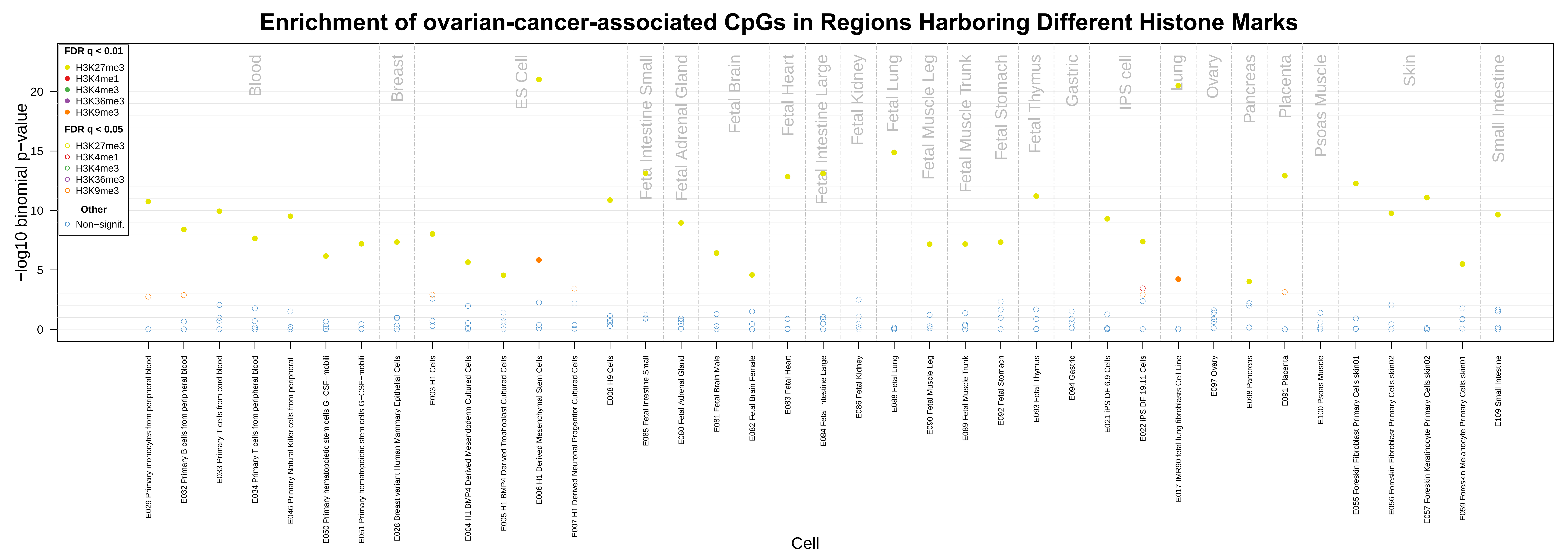

Enrichment of ovarian-cancer-associated CpGs in Regions Harboring Different Histone Marks
FDR q < 0.01
H3K27me3
H3K4me1
H3K4me3
H3K36me3
H3K9me3
FDR q < 0.05
H3K27me3
H3K4me1
H3K4me3
H3K36me3
H3K9me3
Other
Non−signif.
−log10 binomial p−value
0
5
10
15
20
Blood
Breast
ES Cell
Fetal Intestine Small
Fetal Adrenal Gland
Fetal Brain
Fetal Heart
Fetal Intestine Large
Fetal Kidney
Fetal Lung
Fetal Muscle Leg
Fetal Muscle Trunk
Fetal Stomach
Fetal Thymus
Gastric
IPS cell
Lung
Ovary
Pancreas
Placenta
Psoas Muscle
Skin
Small Intestine
E029 Primary monocytes from peripheral blood
E032 Primary B cells from peripheral blood
E033 Primary T cells from cord blood
E034 Primary T cells from peripheral blood
E046 Primary Natural Killer cells from peripheral
E050 Primary hematopoietic stem cells G−CSF−mobili
E051 Primary hematopoietic stem cells G−CSF−mobili
E028 Breast variant Human Mammary Epithelial Cells
E003 H1 Cells
E004 H1 BMP4 Derived Mesendoderm Cultured Cells
E005 H1 BMP4 Derived Trophoblast Cultured Cells
E006 H1 Derived Mesenchymal Stem Cells
E007 H1 Derived Neuronal Progenitor Cultured Cells
E008 H9 Cells
E085 Fetal Intestine Small
E080 Fetal Adrenal Gland
E081 Fetal Brain Male
E082 Fetal Brain Female
E083 Fetal Heart
E084 Fetal Intestine Large
E086 Fetal Kidney
E088 Fetal Lung
E090 Fetal Muscle Leg
E089 Fetal Muscle Trunk
E092 Fetal Stomach
E093 Fetal Thymus
E094 Gastric
E021 iPS DF 6.9 Cells
E022 iPS DF 19.11 Cells
E017 IMR90 fetal lung fibroblasts Cell Line
E097 Ovary
E098 Pancreas
E091 Placenta
E100 Psoas Muscle
E055 Foreskin Fibroblast Primary Cells skin01
E056 Foreskin Fibroblast Primary Cells skin02
E057 Foreskin Keratinocyte Primary Cells skin02
E059 Foreskin Melanocyte Primary Cells skin01
E109 Small Intestine
Cell

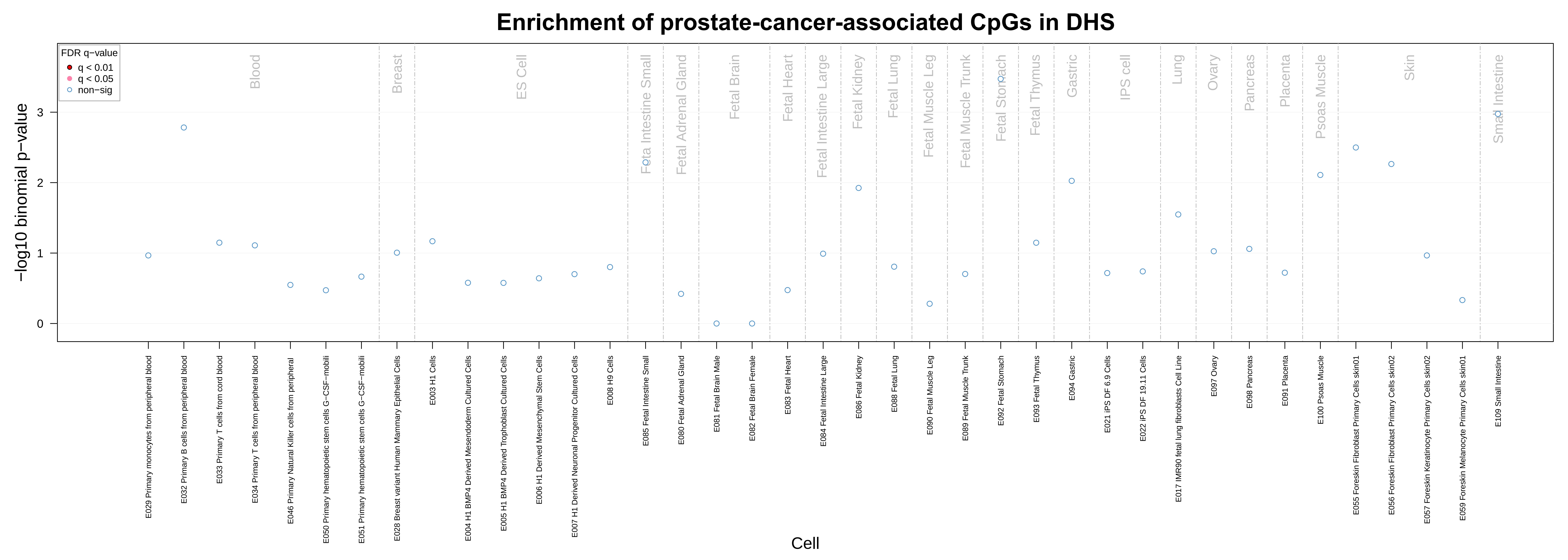

Enrichment of prostate-cancer-associated CpGs in DHS
FDR q-value
q < 0.01
q < 0.05
non−sig
−log10 binomial p−value
Cell
Blood
Breast
ES Cell
Feta Intestine Small
Fetal Adrenal Gland
Fetal Brain
Fetal Heart
Fetal Intestine Large
Fetal Kidney
Fetal Lung
Fetal Muscle Leg
Fetal Muscle Trunk
Fetal Stomach
Fetal Thymus
Gastric
IPS cell
Lung
Ovary
Pancreas
Placenta
Psoas Muscle
Skin
Small Intestine
E029 Primary monocytes from peripheral blood
E032 Primary B cells from peripheral blood
E033 Primary T cells from cord blood
E034 Primary T cells from peripheral blood
E046 Primary Natural Killer cells from peripheral
E050 Primary hematopoietic stem cells G−CSF−mobili
E051 Primary hematopoietic stem cells G−CSF−mobili
E028 Breast variant Human Mammary Epithelial Cells
E003 H1 Cells
E004 H1 BMP4 Derived Mesendoderm Cultured Cells
E005 H1 BMP4 Derived Trophoblast Cultured Cells
E006 H1 Derived Mesenchymal Stem Cells
E007 H1 Derived Neuronal Progenitor Cultured Cells
E008 H9 Cells
E085 Fetal Intestine Small
E080 Fetal Adrenal Gland
E081 Fetal Brain Male
E082 Fetal Brain Female
E083 Fetal Heart
E084 Fetal Intestine Large
E086 Fetal Kidney
E088 Fetal Lung
E090 Fetal Muscle Leg
E089 Fetal Muscle Trunk
E092 Fetal Stomach
E093 Fetal Thymus
E094 Gastric
E021 iPS DF 6.9 Cells
E022 iPS DF 19.11 Cells
E017 IMR90 fetal lung fibroblasts Cell Line
E097 Ovary
E098 Pancreas
E091 Placenta
E100 Psoas Muscle
E055 Foreskin Fibroblast Primary Cells skin01
E056 Foreskin Fibroblast Primary Cells skin02
E057 Foreskin Keratinocyte Primary Cells skin02
E059 Foreskin Melanocyte Primary Cells skin01
E109 Small Intestine
0
1
2
3

#### Enrichment of prostate-cancer-associated CpGs in Chromatins of 15 Different States

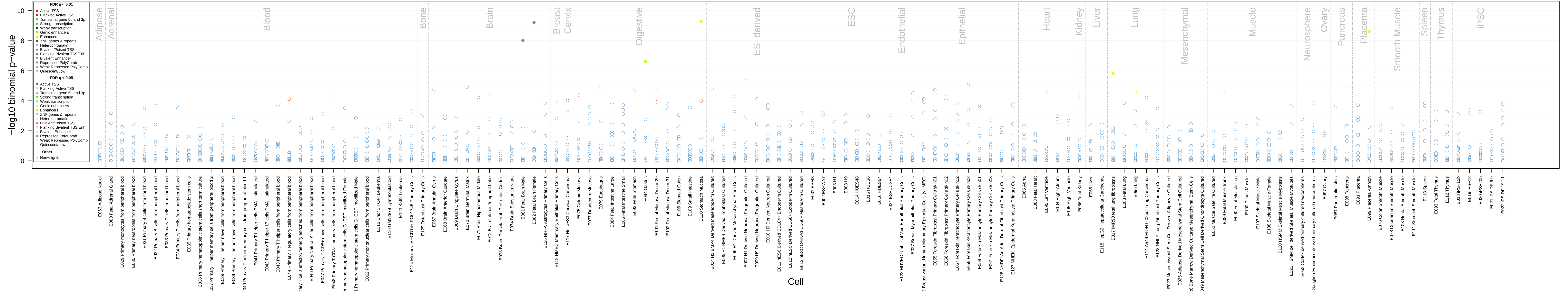

**Enrichment of prostate-cancer-associated CpGs in Regions Harboring Different Histone Marks**

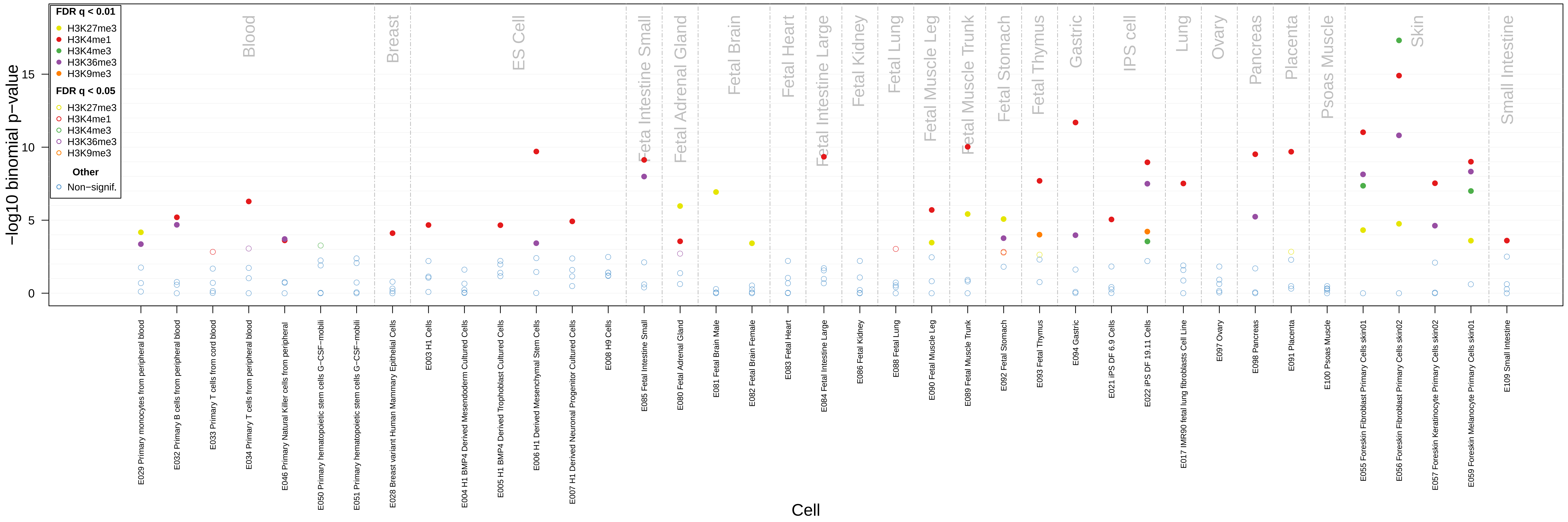

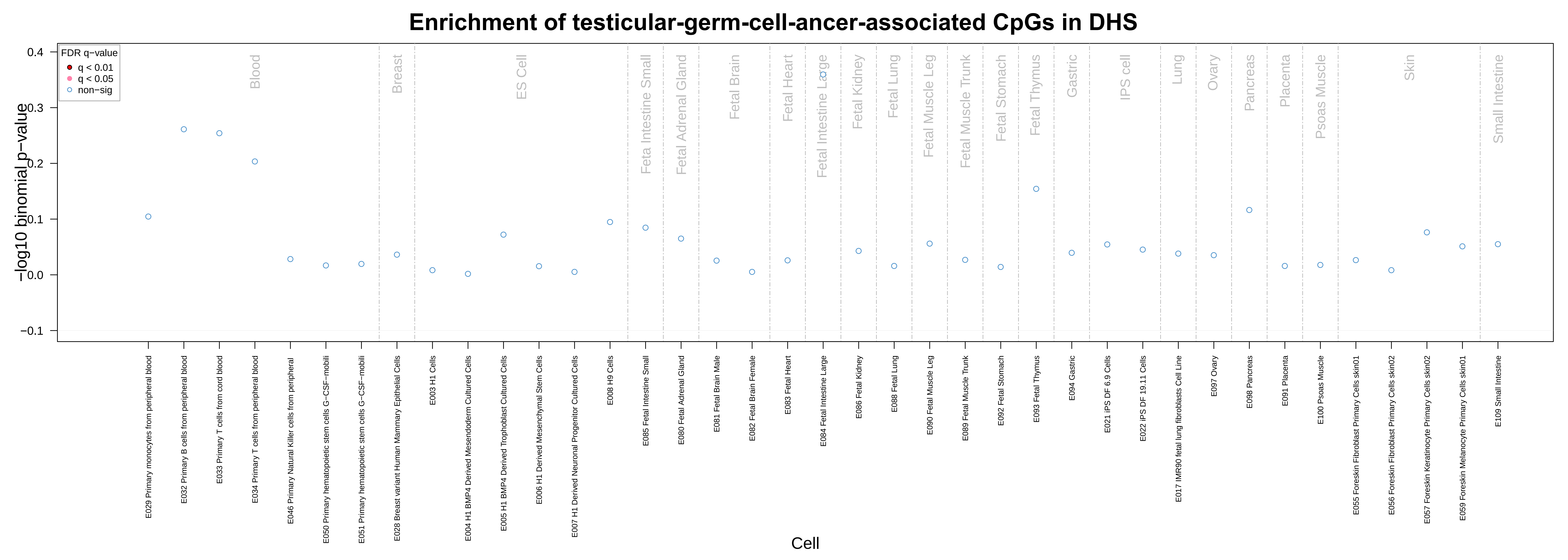

Enrichment of testicular-germ-cell-ancer-associated CpGs in DHS
FDR q−value
q < 0.01
q < 0.05
non−sig
−log10 binomial p−value
0.4
0.3
0.2
0.1
0.0
−0.1
Blood
Breast
ES Cell
Feta Intestine Small
Fetal Adrenal Gland
Fetal Brain
Fetal Heart
Fetal Intestine Large
Fetal Kidney
Fetal Lung
Fetal Muscle Leg
Fetal Muscle Trunk
Fetal Stomach
Fetal Thymus
Gastric
IPS cell
Lung
Ovary
Pancreas
Placenta
Psoas Muscle
Skin
Small Intestine
E029 Primary monocytes from peripheral blood
E032 Primary B cells from peripheral blood
E033 Primary T cells from cord blood
E034 Primary T cells from peripheral blood
E046 Primary Natural Killer cells from peripheral
E050 Primary hematopoietic stem cells G−CSF−mobili
E051 Primary hematopoietic stem cells G−CSF−mobili
E028 Breast variant Human Mammary Epithelial Cells
E003 H1 Cells
E004 H1 BMP4 Derived Mesendoderm Cultured Cells
E005 H1 BMP4 Derived Trophoblast Cultured Cells
E006 H1 Derived Mesenchymal Stem Cells
E007 H1 Derived Neuronal Progenitor Cultured Cells
E008 H9 Cells
E085 Fetal Intestine Small
E080 Fetal Adrenal Gland
E081 Fetal Brain Male
E082 Fetal Brain Female
E083 Fetal Heart
E084 Fetal Intestine Large
E086 Fetal Kidney
E088 Fetal Lung
E090 Fetal Muscle Leg
E089 Fetal Muscle Trunk
E092 Fetal Stomach
E093 Fetal Thymus
E094 Gastric
E021 iPS DF 6.9 Cells
E022 iPS DF 19.11 Cells
E017 IMR90 fetal lung fibroblasts Cell Line
E097 Ovary
E098 Pancreas
E091 Placenta
E100 Psoas Muscle
E055 Foreskin Fibroblast Primary Cells skin01
E056 Foreskin Fibroblast Primary Cells skin02
E057 Foreskin Keratinocyte Primary Cells skin02
E059 Foreskin Melanocyte Primary Cells skin01
E109 Small Intestine
Cell

Enrichment of testicular-germ-cell-ancer-associated CpGs in Chromatins of 15 Different States

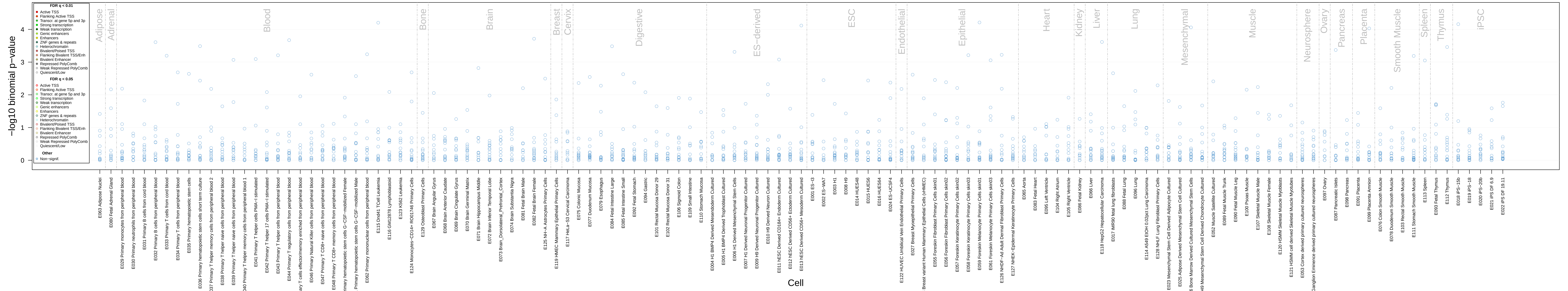

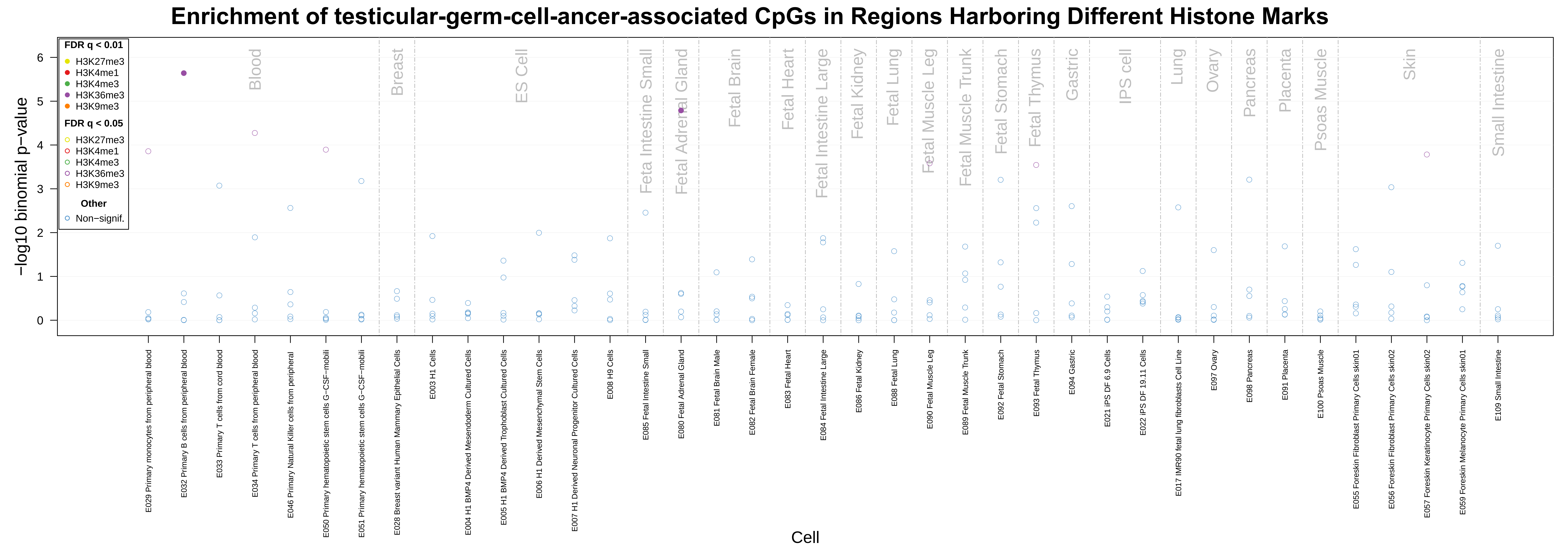

Enrichment of testicular-germ-cell-ancer-associated CpGs in Regions Harboring Different Histone Marks
FDR q < 0.01
H3K27me3
H3K4me1
H3K4me3
H3K36me3
H3K9me3
FDR q < 0.05
H3K27me3
H3K4me1
H3K4me3
H3K36me3
H3K9me3
Other
Non−signif.
−log10 binomial p−value
Blood
Breast
ES Cell
Feta Intestine Small
Fetal Adrenal Gland
Fetal Brain
Fetal Heart
Fetal Intestine Large
Fetal Kidney
Fetal Lung
Fetal Muscle Leg
Fetal Muscle Trunk
Fetal Stomach
Fetal Thymus
Gastric
IPS cell
Lung
Ovary
Pancreas
Placenta
Psoas Muscle
Skin
Small Intestine
E029 Primary monocytes from peripheral blood
E032 Primary B cells from peripheral blood
E033 Primary T cells from cord blood
E034 Primary T cells from peripheral blood
E046 Primary Natural Killer cells from peripheral
E050 Primary hematopoietic stem cells G−CSF−mobili
E051 Primary hematopoietic stem cells G−CSF−mobili
E028 Breast variant Human Mammary Epithelial Cells
E003 H1 Cells
E004 H1 BMP4 Derived Mesendoderm Cultured Cells
E005 H1 BMP4 Derived Trophoblast Cultured Cells
E006 H1 Derived Mesenchymal Stem Cells
E007 H1 Derived Neuronal Progenitor Cultured Cells
E008 H9 Cells
E085 Fetal Intestine Small
E080 Fetal Adrenal Gland
E081 Fetal Brain Male
E082 Fetal Brain Female
E083 Fetal Heart
E084 Fetal Intestine Large
E086 Fetal Kidney
E088 Fetal Lung
E090 Fetal Muscle Leg
E089 Fetal Muscle Trunk
E092 Fetal Stomach
E093 Fetal Thymus
E094 Gastric
E021 iPS DF 6.9 Cells
E022 iPS DF 19.11 Cells
E017 IMR90 fetal lung fibroblasts Cell Line
E097 Ovary
E098 Pancreas
E091 Placenta
E100 Psoas Muscle
E055 Foreskin Fibroblast Primary Cells skin01
E056 Foreskin Fibroblast Primary Cells skin02
E057 Foreskin Keratinocyte Primary Cells skin02
E059 Foreskin Melanocyte Primary Cells skin01
E109 Small Intestine
Cell

**Supplementary Figure 3**

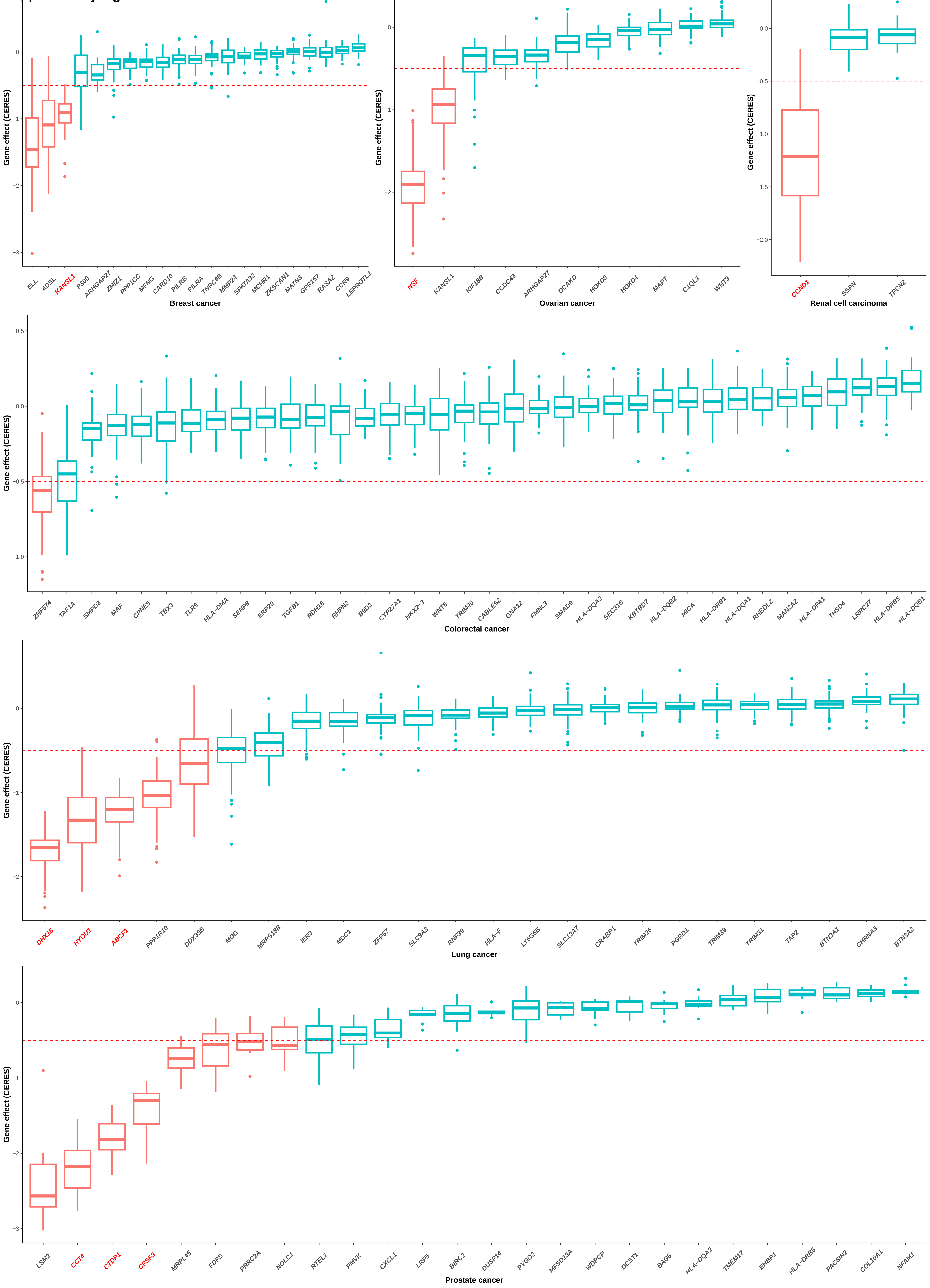

#### Supplementary Figure 4

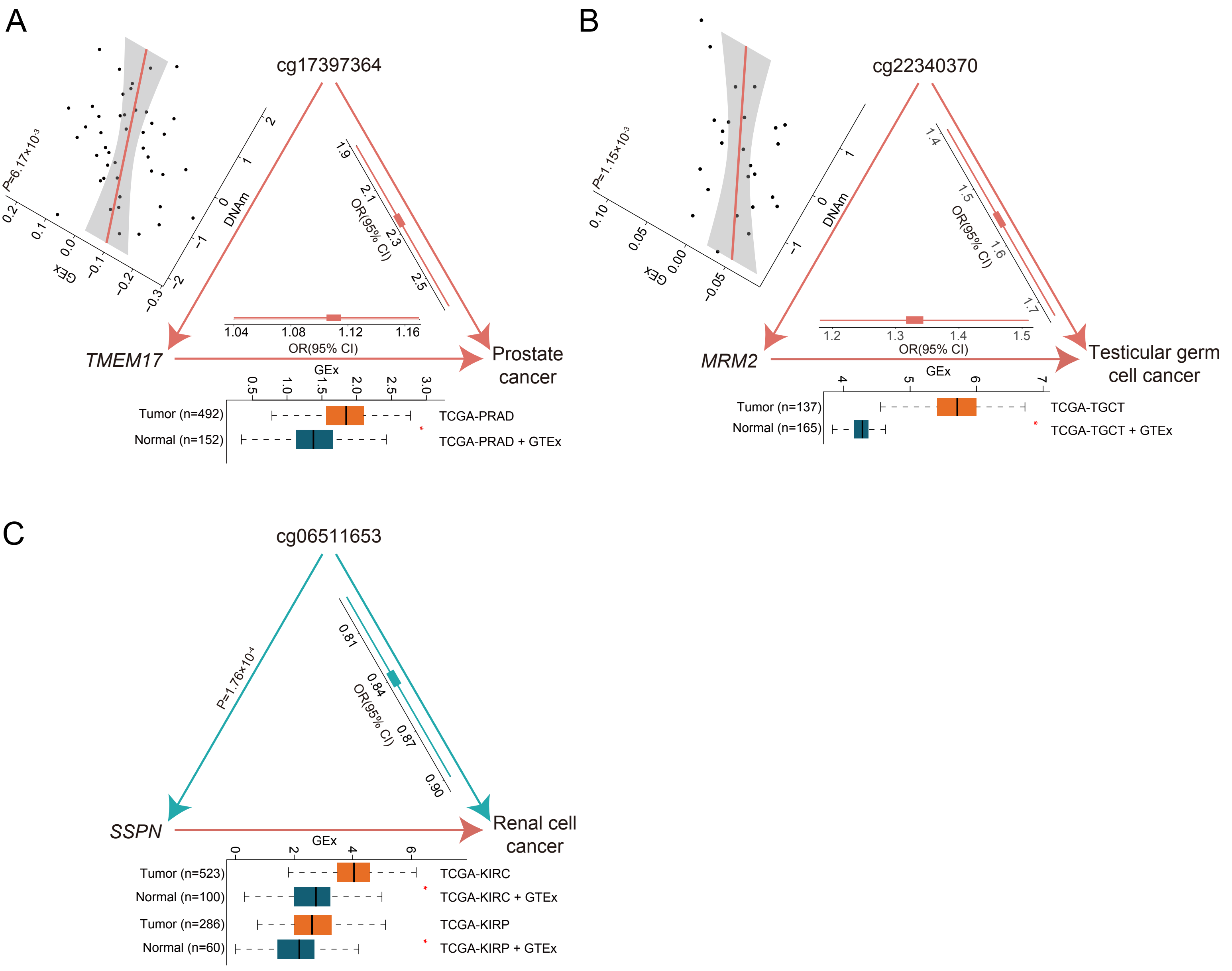
