## Supplementary Tables 1-19 for "Integrating genome and epigenome data to identify tissue-specific DNA methylation biomarkers for cancer risk"

**Supplementary Table 1. Associations with risks of breast cancer subtypes for CpGs showing a significant association with overall breast cancer risk**

| CpG | Chr | Pos | Cytoband | Z score [a] | *P* [a] | R [b] | Closest gene | Classificatio | Luminal A-like | | Luminal B-like | | Luminal B/HER2- | | HER2-enriched-like | | Triple-negative | | $I^2$ | Cochran's *P* |
|---|---|---|---|---|---|---|---|---|---|---|---|---|---|---|---|---|---|---|---|---|
| | | | | | | | | | Z score [a] | *P* [a] | Z score [a] | *P* [a] | Z score [a] | *P* [a] | Z score [a] | *P* [a] | Z score [a] | *P* [a] | | |
| **Associations showing a significant heterogeneity across subtypes (Cochran's *P*<5.01E-07[0.05/99,709; Bonferroni correction])** | | | | | | | | | | | | | | | | | | | | |
| cg0504021 | 1 | 18,807,594 | 1p36.13 | 8.04 | 8.64E-16 | 0.52 | *KLHDC7A* | Exonic | 7.29 | 3.03E-13 | 4.59 | 4.46E-06 | 4.47 | 7.84E-06 | 1.23 | 0.22 | 0.01 | 0.99 | 95.8 | 8.66E-20 |
| cg1267497 | 2 | 217,930,512 | 2q35 | 15.58 | 9.65E-55 | 0.39 | *LINC01921* ; *DIRC3* -AS | Intergenic | 15.35 | 3.37E-53 | 2.83 | 4.71E-03 | 4.06 | 4.84E-05 | 4.03 | 5.60E-05 | 0.05 | 0.96 | 92.3 | 1.59E-10 |
| cg1820458 | 2 | 217,939,092 | 2q35 | 11.15 | 7.31E-29 | 0.38 | *LINC01921* ; *DIRC3-AS* | Intergenic | 11.83 | 2.75E-32 | 3.62 | 2.90E-04 | 2.44 | 0.01 | 2.06 | 0.04 | -0.13 | 0.89 | 93.7 | 4.77E-13 |
| cg0668245 | 2 | 218,231,928 | 2q35 | 10.38 | 3.15E-25 | 0.54 | *DIRC3* | Intronic | 10.04 | 1.01E-23 | 2.58 | 9.97E-03 | 2.86 | 4.30E-03 | 2.25 | 0.02 | 0.29 | 0.77 | 95.5 | 1.53E-18 |
| cg1456128 | 4 | 175,839,468 | 4q34.1 | -11.29 | 1.44E-29 | 0.38 | *ADAM29* | TSS200 | -10.09 | 6.30E-24 | -4.30 | 1.72E-05 | -4.39 | 1.13E-05 | 0.75 | 0.45 | 1.32 | 0.19 | 91.4 | 1.93E-09 |
| cg1465492 | 5 | 44,388,688 | 5p12 | -12.54 | 4.28E-36 | 0.37 | *FGF10* | Exonic | -12.91 | 3.89E-38 | -0.80 | 0.42 | -3.55 | 3.80E-04 | 0.07 | 0.94 | -1.94 | 0.05 | 92.5 | 1.12E-08 |
| cg0897681 | 5 | 44,389,406 | 5p12 | -15.21 | 3.21E-52 | 0.41 | *FGF10-AS1* | Intronic | -16.40 | 1.91E-60 | -3.22 | 1.28E-03 | -5.21 | 1.84E-07 | 0.03 | 0.97 | -1.48 | 0.14 | 93.1 | 8.54E-12 |
| cg0541780 | 10 | 21,807,024 | 10p12.31 | -6.06 | 1.40E-09 | 0.47 | *SKIDA1* | 5'UTR | -7.02 | 2.25E-12 | -2.80 | 5.07E-03 | -2.59 | 9.48E-03 | -0.99 | 0.32 | 2.98 | 2.91E-03 | 88.6 | 4.47E-07 |
| cg2519579 | 10 | 21,807,252 | 10p12.31 | -7.60 | 2.90E-14 | 0.38 | *SKIDA1* | 5'UTR | -8.46 | 2.65E-17 | -2.68 | 7.43E-03 | -2.94 | 3.33E-03 | 0.02 | 0.98 | 2.56 | 0.01 | 90 | 4.30E-08 |
| cg0713523 | 10 | 21,808,126 | 10p12.31 | -7.55 | 4.51E-14 | 0.42 | *SKIDA1* | 5'UTR | -8.65 | 4.97E-18 | -3.23 | 1.24E-03 | -3.50 | 4.63E-04 | -1.07 | 0.28 | 3.74 | 1.87E-04 | 92.6 | 4.40E-11 |
| cg0389159 | 10 | 21,814,313 | 10p12.31 | 7.56 | 4.14E-14 | 0.46 | *SKIDA1* | Intronic | 8.37 | 5.53E-17 | 3.08 | 2.05E-03 | 2.75 | 6.04E-03 | 0.59 | 0.55 | -2.70 | 6.88E-03 | 90.1 | 3.68E-08 |
| cg2219986 | 10 | 22,233,858 | 10p12.31 | -7.09 | 1.32E-12 | 0.44 | *DNAJC1* | Intronic | -7.86 | 3.77E-15 | -3.26 | 1.11E-03 | -3.09 | 2.01E-03 | -0.83 | 0.40 | 3.64 | 2.76E-04 | 91.7 | 8.30E-10 |
| cg1504910 | 10 | 123,353,889 | 10q26.13 | -28.93 | 5.31E-184 | 0.37 | *FGFR2* | Intronic | -25.83 | 4.14E-147 | -9.14 | 6.07E-20 | -12.93 | 3.09E-38 | -1.10 | 0.27 | 0.96 | 0.34 | 97.9 | 5.97E-41 |
| cg2703014 | 10 | 123,688,359 | 10q26.13 | -24.98 | 1.13E-137 | 0.43 | *ATE1-AS1* | Intronic | -22.52 | 2.66E-112 | -8.19 | 2.58E-16 | -11.18 | 5.36E-29 | -0.87 | 0.39 | -0.44 | 0.66 | 97.1 | 1.34E-28 |
| g08353955 | 11 | 69,289,746 | 11q13.3 | -20.70 | 3.48E-95 | 0.64 | *LOC102724265* ; *LINC01488* | Intergenic | -22.13 | 1.65E-108 | -3.62 | 2.92E-04 | -5.76 | 8.46E-09 | -0.96 | 0.34 | 0.58 | 0.56 | 97.2 | 1.93E-29 |
| cg1794110 | 19 | 17,407,198 | 19p13.11 | -6.38 | 1.75E-10 | 0.41 | *ABHD8* | Intronic | 0.55 | 0.58 | -2.88 | 3.93E-03 | -2.44 | 0.01 | 0.23 | 0.82 | -11.02 | 3.10E-28 | 96.1 | 3.20E-21 |
| **Associations showing a nominal significant heterogeneity across subtypes (5.01E-07<Cochran's *P*<0.05)** | | | | | | | | | | | | | | | | | | | | |
| cg0441623 | 1 | 10,051,507 | 1p36.22 | 5.29 | 1.23E-07 | 0.37 | *NMNAT1* ; *RBP7* | Intergenic | -0.02 | 0.98 | 3.42 | 6.28E-04 | 3.81 | 1.41E-04 | 3.48 | 4.97E-04 | 2.47 | 0.01 | 75.4 | 2.71E-03 |
| cg0738416 | 1 | 10,488,281 | 1p36.22 | -5.08 | 3.85E-07 | 0.64 | *PGD* ; *CENPS* | Intergenic | 0.11 | 0.92 | -3.36 | 7.68E-04 | -3.65 | 2.59E-04 | -3.56 | 3.75E-04 | -2.37 | 0.02 | 83.7 | 6.12E-05 |
| cg2712137 | 1 | 11,022,902 | 1p36.22 | -5.35 | 8.56E-08 | 0.47 | *C1orf127* | Intronic | -0.33 | 0.74 | -2.65 | 8.04E-03 | -3.66 | 2.54E-04 | -3.47 | 5.29E-04 | -2.27 | 0.02 | 76.1 | 2.16E-03 |
| cg1300721 | 1 | 18,807,272 | 1p36.13 | 7.88 | 3.16E-15 | 0.66 | *KLHDC7A* | TSS200 | 7.15 | 8.41E-13 | 4.57 | 4.77E-06 | 4.65 | 3.36E-06 | 1.42 | 0.15 | -0.08 | 0.93 | 77.1 | 1.55E-03 |
| cg2020020 | 1 | 18,807,388 | 1p36.13 | 7.65 | 2.03E-14 | 0.68 | *KLHDC7A* | TSS200 | 7.21 | 5.50E-13 | 4.52 | 6.30E-06 | 3.96 | 7.38E-05 | 1.14 | 0.26 | -0.22 | 0.83 | 75.8 | 2.37E-03 |
| cg1435655 | 1 | 18,808,102 | 1p36.13 | 5.97 | 2.37E-09 | 0.52 | *KLHDC7A* | Exonic | 6.08 | 1.24E-09 | 3.41 | 6.48E-04 | 1.68 | 0.09 | 0.31 | 0.75 | -0.44 | 0.66 | 69.7 | 0.01 |
| cg1589609 | 1 | 45,966,115 | 1p34.1 | 5.15 | 2.55E-07 | 0.44 | *MMACHC* | Intronic | 4.30 | 1.73E-05 | 2.70 | 6.92E-03 | 1.45 | 0.15 | 2.00 | 0.05 | -1.08 | 0.28 | 65.1 | 0.02 |
| cg0953942 | 1 | 46,180,057 | 1p34.1 | 6.25 | 4.20E-10 | 0.40 | *IPP* | Exonic | 4.82 | 1.46E-06 | 2.45 | 0.01 | 3.59 | 3.36E-04 | 2.51 | 0.01 | -1.05 | 0.29 | 73.1 | 4.94E-03 |
| cg0884463 | 1 | 46,487,578 | 1p34.1 | -5.17 | 2.35E-07 | 0.37 | *MAST2* | Intronic | -3.86 | 1.13E-04 | -1.55 | 0.12 | -3.57 | 3.62E-04 | -2.26 | 0.02 | 0.47 | 0.64 | 59.1 | 0.04 |
| cg1715791 | 1 | 150,222,676 | 1q21.2 | -7.07 | 1.57E-12 | 0.48 | *ANP32E* ; *CA14* | Intergenic | -6.97 | 3.15E-12 | -2.67 | 7.67E-03 | -0.26 | 0.80 | -0.91 | 0.36 | -2.02 | 0.04 | 59.6 | 0.04 |
| cg2098011 | 1 | 155,108,432 | 1q22 | -5.07 | 3.99E-07 | 0.39 | *SLC50A1* | Exonic | -5.29 | 1.20E-07 | -1.94 | 0.05 | -1.76 | 0.08 | -0.61 | 0.54 | -0.47 | 0.64 | 60 | 0.04 |
| cg1052881 | 1 | 155,699,019 | 1q22 | 5.63 | 1.80E-08 | 0.42 | *DAP3* | Exonic | 5.19 | 2.14E-07 | 0.87 | 0.39 | 1.58 | 0.11 | 1.60 | 0.11 | -0.83 | 0.41 | 62.5 | 0.03 |
| cg2109390 | 2 | 25,097,146 | 2p23.3 | 6.22 | 5.08E-10 | 0.39 | *ADCY3* | Intronic | 3.39 | 6.91E-04 | 2.99 | 2.80E-03 | 0.02 | 0.99 | 3.28 | 1.02E-03 | 2.54 | 0.01 | 61.5 | 0.03 |
| cg0458662 | 2 | 25,135,609 | 2p23.3 | -6.71 | 1.93E-11 | 0.66 | *ADCY3* | Intronic | -3.65 | 2.61E-04 | -3.09 | 1.98E-03 | -0.24 | 0.81 | -3.30 | 9.57E-04 | -3.13 | 1.74E-03 | 62 | 0.03 |
| cg0847131 | 2 | 25,141,735 | 2p23.3 | -6.45 | 1.10E-10 | 0.59 | *ADCY3* | Exonic | -3.33 | 8.75E-04 | -2.69 | 7.10E-03 | -0.21 | 0.83 | -3.42 | 6.15E-04 | -2.76 | 5.82E-03 | 60.9 | 0.04 |
| cg0895250 | 2 | 201,450,610 | 2q33.1 | 5.56 | 2.65E-08 | 0.61 | *AOX1* | TSS200 | 2.60 | 9.33E-03 | 2.43 | 0.01 | -0.16 | 0.87 | -0.43 | 0.67 | 4.91 | 8.89E-07 | 74.8 | 3.23E-03 |
| cg1964138 | 2 | 201,694,128 | 2q33.1 | -6.21 | 5.33E-10 | 0.76 | *BZW1* ; *CLK1* | Intergenic | -2.47 | 0.01 | -2.43 | 0.02 | 0.67 | 0.50 | 0.73 | 0.47 | -4.63 | 3.63E-06 | 78.1 | 1.10E-03 |
| cg0079495 | 2 | 201,694,177 | 2q33.1 | -6.33 | 2.47E-10 | 0.75 | *BZW1* ; *CLK1* | Intergenic | -2.52 | 0.01 | -2.40 | 0.02 | 0.53 | 0.60 | 0.86 | 0.39 | -4.82 | 1.41E-06 | 79.3 | 6.83E-04 |
| cg2764676 | 2 | 217,957,999 | 2q35 | -11.18 | 5.11E-29 | 0.41 | *LINC01921* ; *DIRC3-AS* | Intergenic | -10.66 | 1.55E-26 | -4.82 | 1.41E-06 | -2.88 | 4.01E-03 | -3.18 | 1.49E-03 | -1.25 | 0.21 | 78.7 | 8.59E-04 |
| cg0321311 | 3 | 26,691,758 | 3p24.1 | 10.20 | 2.00E-24 | 0.47 | *LRRC3B* | Intronic | 8.71 | 3.04E-18 | 2.46 | 0.01 | 3.92 | 8.69E-05 | 0.40 | 0.69 | 1.21 | 0.22 | 68.6 | 0.01 |
| cg0311695 | 3 | 26,736,240 | 3p24.1 | -6.54 | 6.05E-11 | 0.65 | *LRRC3B* | 5'UTR | -6.25 | 4.15E-10 | -1.87 | 0.06 | -3.68 | 2.38E-04 | -0.50 | 0.62 | -0.14 | 0.89 | 76.7 | 1.80E-03 |
| cg0362634 | 3 | 27,368,232 | 3p24.1 | 9.43 | 4.07E-21 | 0.48 | *NEK10* | Intronic | 7.55 | 4.46E-14 | 1.16 | 0.24 | 4.33 | 1.47E-05 | -0.31 | 0.76 | 2.00 | 0.05 | 69.2 | 0.01 |
| cg0399592 | 3 | 27,408,898 | 3p24.1 | -5.93 | 3.07E-09 | 0.46 | *NEK10* | Intronic | -4.81 | 1.53E-06 | -1.15 | 0.25 | -1.15 | 0.25 | -0.38 | 0.70 | -2.66 | 7.89E-03 | 77 | 1.63E-03 |
| cg2103239 | 3 | 27,528,777 | 3p24.1 | 15.29 | 9.35E-53 | 0.45 | *SLC4A7* ; *EOMES* | Intergenic | 12.88 | 5.78E-38 | 4.51 | 6.39E-06 | 5.57 | 2.61E-08 | 0.27 | 0.79 | 3.20 | 1.38E-03 | 84.5 | 3.53E-05 |
| cg0247634 | 3 | 27,670,457 | 3p24.1 | -11.20 | 4.04E-29 | 0.52 | *SLC4A7* ; *EOMES* | Intergenic | -9.91 | 3.60E-23 | -4.03 | 5.63E-05 | -2.94 | 3.30E-03 | -2.11 | 0.04 | -1.58 | 0.11 | 68.2 | 0.01 |
| cg1455753 | 3 | 27,763,581 | 3p24.1 | -14.59 | 3.47E-48 | 0.39 | *EOMES* | Exonic | -13.49 | 1.68E-41 | -3.05 | 2.29E-03 | -5.76 | 8.50E-09 | -0.97 | 0.33 | -2.75 | 5.99E-03 | 83.8 | 5.80E-05 |
| cg2340373 | 3 | 99,877,771 | 3q12.1 | -7.91 | 2.47E-15 | 0.49 | *CMSS1* | Intronic | -6.90 | 5.32E-12 | -2.38 | 0.02 | -2.05 | 0.04 | -0.04 | 0.97 | -0.97 | 0.33 | 61.8 | 0.03 |
| cg0815851 | 3 | 140,866,589 | 3q23 | -6.47 | 9.80E-11 | 0.63 | *SPSB4* | 3'UTR | -5.65 | 1.59E-08 | -3.29 | 9.95E-04 | -4.47 | 7.90E-06 | -2.07 | 0.04 | -0.44 | 0.66 | 61.2 | 0.04 |
| cg0701068 | 5 | 1,217,576 | 5p15.33 | -5.39 | 6.90E-08 | 0.36 | *SLC6A19* | Intronic | -2.96 | 3.10E-03 | -0.68 | 0.50 | -1.54 | 0.12 | -0.38 | 0.70 | -5.03 | 4.86E-07 | 66.8 | 0.02 |
| cg0062441 | 5 | 1,282,357 | 5p15.33 | -5.14 | 2.70E-07 | 0.38 | *TERT* | Intronic | -3.06 | 2.21E-03 | -1.50 | 0.13 | -1.58 | 0.11 | -1.32 | 0.19 | -7.95 | 1.81E-15 | 88.2 | 8.06E-07 |
| cg1533996 | 5 | 45,693,264 | 5p12 | -7.35 | 2.02E-13 | 0.57 | *HCN1* | Intronic | -8.07 | 7.15E-16 | -1.43 | 0.15 | -1.48 | 0.14 | -0.71 | 0.48 | -0.88 | 0.38 | 78.8 | 8.26E-04 |
| cg0225824 | 5 | 55,777,117 | 5q11.2 | 9.96 | 2.22E-23 | 0.38 | *LINC01948* | Intronic | 10.57 | 4.29E-26 | 3.11 | 1.87E-03 | 4.04 | 5.30E-05 | 0.91 | 0.37 | 0.67 | 0.51 | 84.1 | 4.77E-05 |
| cg2278814 | 5 | 56,033,159 | 5q11.2 | -8.17 | 3.07E-16 | 0.39 | *C5orf67* ; *LOC105378979* | Intergenic | -6.71 | 1.92E-11 | -5.49 | 4.04E-08 | -4.00 | 6.33E-05 | 0.55 | 0.58 | -2.19 | 0.03 | 69.6 | 0.01 |
| cg0897173 | 5 | 56,110,438 | 5q11.2 | -12.48 | 9.85E-36 | 0.38 | *MAP3K1* | TSS1500 | -12.06 | 1.77E-33 | -4.39 | 1.12E-05 | -4.58 | 4.70E-06 | -1.15 | 0.25 | -1.23 | 0.22 | 83 | 9.96E-05 |
| cg1780928 | 5 | 56,205,270 | 5q11.2 | -5.59 | 2.23E-08 | 0.41 | *SETD9* | 5'UTR | -5.87 | 4.32E-09 | -4.34 | 1.45E-05 | -0.72 | 0.47 | -0.84 | 0.40 | 0.01 | 1.00 | 75.9 | 2.34E-03 |
| cg0380077 | 5 | 131,743,782 | 5q31.1 | -5.19 | 2.09E-07 | 0.41 | *SLC22A5* ; *C5orf56* | Intergenic | -2.50 | 0.01 | -3.73 | 1.92E-04 | -1.32 | 0.19 | -0.86 | 0.39 | 0.46 | 0.64 | 59.6 | 0.04 |
| cg0156820 | 5 | 132,722,571 | 5q31.1 | -5.06 | 4.14E-07 | 0.54 | *FSTL4* | Intronic | -4.97 | 6.86E-07 | -1.48 | 0.14 | -0.92 | 0.36 | 0.31 | 0.76 | 0.74 | 0.46 | 64.9 | 0.02 |
| g14306534 | 6 | 151,712,952 | 6q25.1 | 8.12 | 4.72E-16 | 0.41 | *ZBTB2* | TSS200 | 3.26 | 1.10E-03 | 3.26 | 1.10E-03 | 3.83 | 1.26E-04 | 2.71 | 6.65E-03 | 5.28 | 1.29E-07 | 71.1 | 7.81E-03 |
| cg1322983 | 6 | 152,010,481 | 6q25.1 | -9.21 | 3.21E-20 | 0.37 | *CCDC170* ; *ESR1* | Intergenic | -4.59 | 4.54E-06 | -3.84 | 1.21E-04 | -3.33 | 8.77E-04 | -2.58 | 9.83E-03 | -6.24 | 4.39E-10 | 69.8 | 0.01 |
| cg1770137 | 7 | 94,285,343 | 7q21.3 | 7.45 | 9.46E-14 | 0.45 | *SGCE* | Exonic | 4.92 | 8.80E-07 | 3.03 | 2.44E-03 | 3.95 | 7.88E-05 | 0.69 | 0.49 | -0.45 | 0.65 | 64.9 | 0.02 |
| cg2215054 | 7 | 102,059,183 | 7q22.1 | -5.34 | 9.30E-08 | 0.45 | *LOC100630923* | Intronic | -4.16 | 3.15E-05 | 0.84 | 0.40 | -1.32 | 0.19 | -2.87 | 4.17E-03 | -0.54 | 0.59 | 63.6 | 0.03 |
| cg0203670 | 8 | 29,529,211 | 8p12 | -9.22 | 2.84E-20 | 0.72 | *DUSP4* ; *LINC00589* | Intergenic | -8.15 | 3.60E-16 | -1.76 | 0.08 | -2.32 | 0.02 | 1.13 | 0.26 | -5.45 | 5.07E-08 | 78.9 | 7.88E-04 |
| cg1860488 | 8 | 124,408,541 | 8q24.13 | 6.03 | 1.67E-09 | 0.53 | *ATAD2* | Exonic | 5.32 | 1.02E-07 | 0.33 | 0.74 | 1.58 | 0.11 | -0.38 | 0.70 | 4.20 | 2.71E-05 | 60.2 | 0.04 |
| cg1425099 | 9 | 106,856,677 | 9q31.1 | 5.36 | 8.28E-08 | 0.49 | *SMC2* | 5'UTR | 1.98 | 0.05 | 4.82 | 1.42E-06 | 3.08 | 2.06E-03 | 0.86 | 0.39 | 2.59 | 9.47E-03 | 72.6 | 5.55E-03 |
| cg1114825 | 9 | 106,856,714 | 9q31.1 | -5.14 | 2.73E-07 | 0.57 | *SMC2* | 5'UTR | -1.71 | 0.09 | -4.66 | 3.16E-06 | -2.96 | 3.11E-03 | -0.84 | 0.40 | -2.75 | 6.02E-03 | 72.8 | 5.30E-03 |
| cg1292731 | 9 | 106,856,733 | 9q31.1 | 5.12 | 3.07E-07 | 0.53 | *SMC2* | 5'UTR | 1.59 | 0.11 | 4.68 | 2.94E-06 | 3.02 | 2.56E-03 | 0.82 | 0.41 | 2.84 | 4.45E-03 | 74.5 | 3.51E-03 |
| cg2452217 | 9 | 106,856,962 | 9q31.1 | -5.08 | 3.79E-07 | 0.59 | *SMC2* | 5'UTR | -1.54 | 0.12 | -4.68 | 2.81E-06 | -3.00 | 2.74E-03 | -0.87 | 0.39 | -2.85 | 4.31E-03 | 74.8 | 3.18E-03 |
| cg2074527 | 9 | 110,309,675 | 9q31.2 | 8.04 | 9.14E-16 | 0.47 | *KLF4* ; *ACTL7B* | Intergenic | 5.26 | 1.45E-07 | 3.75 | 1.76E-04 | 2.64 | 8.22E-03 | 1.66 | 0.10 | -0.51 | 0.61 | 63.9 | 0.03 |
| cg0927936 | 9 | 110,822,615 | 9q31.2 | -8.49 | 2.04E-17 | 0.45 | *KLF4* ; *ACTL7B* | Intergenic | -7.86 | 3.92E-15 | -2.03 | 0.04 | -4.37 | 1.24E-05 | -2.10 | 0.04 | -0.74 | 0.46 | 61.7 | 0.03 |
| cg0334010 | 10 | 22,028,443 | 10p12.31 | -5.53 | 3.17E-08 | 0.46 | *MLLT10* | Intronic | -5.52 | 3.37E-08 | -2.14 | 0.03 | -2.82 | 4.74E-03 | -0.18 | 0.86 | 1.77 | 0.08 | 85.2 | 1.46E-04 |
| cg0960330 | 10 | 114,781,546 | 10q25.2 | -7.64 | 2.21E-14 | 0.65 | *TCF7L2* | Intronic | -6.17 | 6.71E-10 | -0.65 | 0.51 | -0.56 | 0.58 | 0.31 | 0.76 | -5.86 | 4.73E-09 | 81.8 | 2.00E-04 |
| cg1042096 | 10 | 114,812,813 | 10q25.2 | 7.31 | 2.61E-13 | 0.47 | *TCF7L2* | Intronic | 6.15 | 7.74E-10 | 0.68 | 0.49 | 0.60 | 0.55 | -0.19 | 0.85 | 5.93 | 2.96E-09 | 81.7 | 2.19E-04 |
| cg0807041 | 10 | 115,201,116 | 10q25.3 | -5.25 | 1.50E-07 | 0.68 | *TCF7L2* ; *HABP2* | Intergenic | -6.29 | 3.25E-10 | 0.60 | 0.55 | -3.01 | 2.58E-03 | -1.27 | 0.21 | -2.32 | 0.02 | 59.2 | 0.04 |
| cg2545938 | 10 | 122,948,144 | 10q26.12 | 8.75 | 2.18E-18 | 0.48 | *WDR11* ; *FGFR2* | Intergenic | 7.62 | 2.46E-14 | 3.68 | 2.34E-04 | 4.30 | 1.71E-05 | 0.68 | 0.50 | 0.32 | 0.75 | 86 | 9.15E-06 |
| cg2263303 | 10 | 123,355,576 | 10q26.13 | -7.97 | 1.54E-15 | 0.42 | *FGFR2* | Intronic | -6.13 | 8.61E-10 | -2.36 | 0.02 | -3.53 | 4.17E-04 | 0.33 | 0.74 | 0.26 | 0.79 | 68 | 0.01 |
| cg1771077 | 10 | 123,356,331 | 10q26.13 | -9.00 | 2.18E-19 | 0.59 | *FGFR2* | Intronic | -8.53 | 1.53E-17 | -2.49 | 0.01 | -4.33 | 1.51E-05 | 0.59 | 0.55 | -1.43 | 0.15 | 72 | 6.37E-03 |
| cg0329383 | 10 | 123,506,700 | 10q26.13 | -5.87 | 4.23E-09 | 0.41 | *ATE1* | Intronic | -5.11 | 3.16E-07 | -2.34 | 0.02 | -1.90 | 0.06 | 1.79 | 0.07 | 0.09 | 0.93 | 69.7 | 0.01 |
| cg2369125 | 10 | 123,688,108 | 10q26.13 | -6.71 | 2.01E-11 | 0.42 | *ATE1-AS1* | Exonic | -7.46 | 8.63E-14 | -1.48 | 0.14 | -2.48 | 0.01 | -0.33 | 0.74 | 0.68 | 0.50 | 77.2 | 1.53E-03 |
| cg0744448 | 10 | 123,688,279 | 10q26.13 | -7.32 | 2.48E-13 | 0.56 | *ATE1-AS1* | Intronic | -7.11 | 1.13E-12 | -1.64 | 0.10 | -3.37 | 7.60E-04 | 0.20 | 0.84 | -0.24 | 0.81 | 78.9 | 8.02E-04 |
| cg0673379 | 10 | 123,922,970 | 10q26.13 | -6.31 | 2.72E-10 | 0.42 | *TACC2* | Intronic | -4.98 | 6.25E-07 | -1.62 | 0.10 | -1.97 | 0.05 | 0.41 | 0.68 | 0.44 | 0.66 | 59.4 | 0.04 |
| cg1142066 | 11 | 1,750,353 | 11p15.5 | 7.00 | 2.55E-12 | 0.39 | *KRTAP5* -6; *IFITM10* | Intergenic | 5.29 | 1.26E-07 | 4.98 | 6.28E-07 | 1.60 | 0.11 | 0.31 | 0.76 | 1.30 | 0.19 | 67.9 | 0.01 |
| cg1336751 | 11 | 1,942,466 | 11p15.5 | 12.86 | 7.10E-38 | 0.45 | *TNNT3* | Intronic | 9.74 | 1.97E-22 | 6.15 | 7.65E-10 | 2.87 | 4.13E-03 | 3.03 | 2.46E-03 | 2.70 | 6.89E-03 | 68.2 | 0.01 |
| cg0302289 | 11 | 1,947,791 | 11p15.5 | 10.81 | 3.03E-27 | 0.47 | *TNNT3* | Intronic | 8.90 | 5.53E-19 | 5.73 | 9.81E-09 | 2.16 | 0.03 | 3.11 | 1.87E-03 | 2.22 | 0.03 | 71.6 | 6.96E-03 |
| cg0833795 | 11 | 1,948,933 | 11p15.5 | 6.61 | 3.77E-11 | 0.45 | *TNNT3* | Intronic | 6.15 | 7.89E-10 | 4.32 | 1.53E-05 | 0.69 | 0.49 | 0.77 | 0.44 | 2.07 | 0.04 | 60.5 | 0.04 |
| cg1803250 | 11 | 1,949,113 | 11p15.5 | 6.68 | 2.34E-11 | 0.46 | *TNNT3* | Intronic | 6.18 | 6.37E-10 | 4.32 | 1.56E-05 | 0.75 | 0.45 | 0.75 | 0.45 | 2.07 | 0.04 | 59.6 | 0.04 |
| cg2388213 | 11 | 68,779,826 | 11q13.3 | -5.14 | 2.69E-07 | 0.46 | *MRGPRF-AS1* | Exonic | -4.76 | 1.91E-06 | -1.06 | 0.29 | -2.33 | 0.02 | 2.44 | 0.01 | -1.40 | 0.16 | 72.4 | 5.83E-03 |
| cg2347854 | 11 | 69,259,265 | 11q13.3 | -7.27 | 3.55E-13 | 0.60 | *LOC102724265* ; *LINC01488* | Intergenic | -6.15 | 7.84E-10 | -1.48 | 0.14 | -3.35 | 8.14E-04 | 1.11 | 0.27 | -1.07 | 0.28 | 66 | 0.02 |
| cg0724166 | 11 | 69,259,336 | 11q13.3 | -6.56 | 5.53E-11 | 0.46 | *LOC102724265* ; *LINC01488* | Intergenic | -5.40 | 6.83E-08 | -1.45 | 0.15 | -3.24 | 1.20E-03 | 1.41 | 0.16 | -1.14 | 0.26 | 64 | 0.03 |
| cg0096666 | 11 | 69,259,631 | 11q13.3 | -6.72 | 1.83E-11 | 0.44 | *LOC102724265* ; *LINC01488* | Intergenic | -5.70 | 1.21E-08 | -1.56 | 0.12 | -3.36 | 7.66E-04 | 1.66 | 0.10 | -1.30 | 0.19 | 69.6 | 0.01 |
| cg0706804 | 11 | 69,312,202 | 11q13.3 | -9.01 | 1.97E-19 | 0.53 | *LINC01488* ; *CCND1* | Intergenic | -8.91 | 5.28E-19 | 0.21 | 0.83 | -2.39 | 0.02 | -1.33 | 0.18 | 0.59 | 0.56 | 87 | 3.39E-06 |
| cg0853536 | 12 | 96,177,524 | 12q22 | 14.26 | 3.70E-46 | 0.66 | *NTN4* | Intronic | 11.44 | 2.51E-30 | 4.75 | 2.07E-06 | 4.07 | 4.63E-05 | 0.70 | 0.49 | 5.41 | 6.20E-08 | 58.2 | 0.05 |
| cg0721176 | 12 | 115,102,290 | 12q24.21 | 6.55 | 5.95E-11 | 0.51 | *TBX5-AS1* ; *TBX3* | Intergenic | 5.62 | 1.94E-08 | 2.03 | 0.04 | 3.65 | 2.66E-04 | -0.24 | 0.81 | 0.31 | 0.76 | 58.4 | 0.05 |
| cg1092444 | 12 | 115,825,501 | 12q24.21 | -13.03 | 7.74E-39 | 0.57 | *TBX3* ; *MED13L* | Intergenic | -12.57 | 3.14E-36 | -4.33 | 1.51E-05 | -3.59 | 3.34E-04 | -3.19 | 1.42E-03 | -0.81 | 0.42 | 86.2 | 7.88E-06 |
| cg2611987 | 14 | 37,530,475 | 14q13.3 | 6.79 | 1.14E-11 | 0.37 | *SLC25A21* | Intronic | 7.37 | 1.73E-13 | 1.45 | 0.15 | 2.15 | 0.03 | 0.84 | 0.40 | 0.60 | 0.55 | 63.8 | 0.03 |
| cg0310853 | 14 | 68,746,051 | 14q24.1 | -9.84 | 7.35E-23 | 0.37 | *RAD51B* | Intronic | -8.90 | 5.62E-19 | -2.19 | 0.03 | -2.48 | 0.01 | -0.50 | 0.62 | -2.72 | 6.50E-03 | 58.4 | 0.05 |
| cg1051190 | 14 | 91,842,949 | 14q32.11 | 7.41 | 1.30E-13 | 0.42 | *CCDC88C* | Intronic | 5.66 | 1.49E-08 | 4.48 | 7.58E-06 | 2.81 | 5.03E-03 | 1.61 | 0.11 | -0.17 | 0.86 | 69.3 | 0.01 |
| cg0106691 | 15 | 91,478,537 | 15q26.1 | -5.68 | 1.34E-08 | 0.45 | *UNC45A* | 5'UTR | -3.43 | 6.13E-04 | -3.24 | 1.18E-03 | -5.20 | 2.03E-07 | -2.21 | 0.03 | -2.47 | 0.01 | 62.3 | 0.03 |
| g22652561 | 17 | 11,924,127 | 17p12 | 5.12 | 3.09E-07 | 0.40 | *MAP2K4* | TSS200 | 4.11 | 4.00E-05 | 0.35 | 0.72 | 2.32 | 0.02 | 1.08 | 0.28 | -1.07 | 0.28 | 58.7 | 0.05 |
| cg0164072 | 17 | 44,029,468 | 17q21.31 | 6.49 | 8.41E-11 | 0.51 | *MAPT* | Intronic | 4.22 | 2.48E-05 | 2.45 | 0.01 | 0.17 | 0.87 | 2.27 | 0.02 | -0.10 | 0.92 | 63.5 | 0.03 |
| cg2459989 | 17 | 44,270,211 | 17q21.31 | 5.54 | 3.11E-08 | 0.37 | *KANSL1* | 5'UTR | 3.72 | 1.98E-04 | 2.09 | 0.04 | 0.33 | 0.74 | 2.05 | 0.04 | -1.06 | 0.29 | 62.8 | 0.03 |
| cg2499200 | 17 | 44,270,238 | 17q21.31 | 6.04 | 1.51E-09 | 0.49 | *KANSL1* | 5'UTR | 4.52 | 6.22E-06 | 2.34 | 0.02 | 0.73 | 0.47 | 2.16 | 0.03 | -0.68 | 0.50 | 62.3 | 0.03 |

| | | | | | | | | | | | | | | | | | | | | |
|---|---|---|---|---|---|---|---|---|---|---|---|---|---|---|---|---|---|---|---|---|
| cg05358814 | 17 | 53,046,663 | 17q22 | -8.22 | 2.05E-16 | 0.63 | STXBP4 | Intronic | -8.27 | 1.35E-16 | -1.34 | 0.18 | -3.04 | 2.35E-03 | -2.17 | 0.03 | -0.47 | 0.64 | 69.7 | 0.01 |
| cg23202870 | 17 | 53,245,218 | 17q22 | 10.78 | 4.37E-27 | 0.49 | STXBP4 | 3'UTR | 10.66 | 1.58E-26 | 3.98 | 6.79E-05 | 3.57 | 3.59E-04 | 1.33 | 0.19 | 0.04 | 0.97 | 79.8 | 5.36E-04 |
| cg14861570 | 17 | 53,499,954 | 17q22 | -10.21 | 1.88E-24 | 0.58 | MMD | TSS1500 | -10.17 | 2.58E-24 | -3.58 | 3.39E-04 | -3.18 | 1.45E-03 | -1.34 | 0.18 | 0.03 | 0.98 | 87.6 | 1.77E-06 |
| cg17996099 | 17 | 53,588,349 | 17q22 | 6.88 | 6.11E-12 | 0.40 | MMD ; TMEM100 | Intergenic | 6.90 | 5.24E-12 | 3.59 | 3.36E-04 | 2.99 | 2.76E-03 | -0.41 | 0.68 | 1.14 | 0.26 | 68.8 | 0.01 |
| cg08479950 | 19 | 13,960,019 | 19p13.13 | -5.82 | 5.93E-09 | 0.43 | LOC284454 ; MIR181C | Intergenic | -3.88 | 1.06E-04 | -1.74 | 0.08 | -2.33 | 0.02 | -1.22 | 0.22 | 1.37 | 0.17 | 61 | 0.04 |
| cg04304033 | 19 | 18,547,070 | 19p13.11 | -9.89 | 4.39E-23 | 0.37 | ISYNA1 | Intronic | -9.35 | 8.90E-21 | -3.09 | 1.97E-03 | -1.85 | 0.06 | -1.76 | 0.08 | -2.31 | 0.02 | 63.1 | 0.03 |
| cg22640344 | 19 | 18,548,211 | 19p13.11 | -11.29 | 1.41E-29 | 0.51 | ISYNA1 | Intronic | -10.23 | 1.49E-24 | -3.22 | 1.27E-03 | -1.80 | 0.07 | -2.18 | 0.03 | -2.96 | 3.04E-03 | 68.4 | 0.01 |
| cg04845985 | 19 | 18,548,229 | 19p13.11 | -11.30 | 1.27E-29 | 0.65 | ISYNA1 | Intronic | -10.29 | 8.11E-25 | -3.22 | 1.29E-03 | -1.80 | 0.07 | -2.08 | 0.04 | -2.97 | 2.93E-03 | 69 | 0.01 |
| cg11102782 | 19 | 18,549,136 | 19p13.11 | 8.29 | 1.09E-16 | 0.43 | ISYNA1 | TSS200 | 8.00 | 1.25E-15 | 2.57 | 0.01 | 0.55 | 0.59 | 1.12 | 0.26 | 2.59 | 9.54E-03 | 69 | 0.01 |
| cg27342294 | 20 | 34,570,157 | 20q11.23 | 5.51 | 3.55E-08 | 0.38 | CNBD2 | Intronic | 2.93 | 3.44E-03 | 2.02 | 0.04 | 4.98 | 6.43E-07 | -0.58 | 0.56 | 1.25 | 0.21 | 77.3 | 1.48E-03 |
| cg00711252 | 22 | 29,108,527 | 22q12.1 | 6.19 | 5.92E-10 | 0.40 | CHEK2 | Intronic | 3.77 | 1.63E-04 | 3.96 | 7.63E-05 | 3.92 | 8.78E-05 | 0.99 | 0.32 | 0.30 | 0.77 | 60.6 | 0.04 |
| cg16381371 | 22 | 29,552,438 | 22q12.1 | -5.24 | 1.64E-07 | 0.46 | KREMEN1 | Intronic | -3.10 | 1.96E-03 | -3.74 | 1.80E-04 | -3.06 | 2.19E-03 | -2.50 | 0.01 | 1.05 | 0.30 | 75.2 | 2.87E-03 |
| cg13868216 | 22 | 38,484,884 | 22q13.1 | 5.81 | 6.40E-09 | 0.37 | BAIAP2L2 | Exonic | 5.10 | 3.47E-07 | 1.66 | 0.10 | 2.66 | 7.92E-03 | -0.08 | 0.94 | -0.97 | 0.33 | 67.1 | 0.02 |
| cg20207973 | 22 | 38,506,712 | 22q13.1 | -7.24 | 4.57E-13 | 0.43 | BAIAP2L2 ; PLA2G6 | TSS200 | -6.34 | 2.28E-10 | -1.89 | 0.06 | -3.21 | 1.33E-03 | -0.80 | 0.42 | 0.46 | 0.65 | 67.4 | 0.02 |
| cg09247692 | 22 | 38,506,781 | 22q13.1 | -6.15 | 7.89E-10 | 0.39 | BAIAP2L2 ; PLA2G6 | TSS200 | -5.50 | 3.90E-08 | -1.70 | 0.09 | -3.13 | 1.77E-03 | -0.77 | 0.44 | 0.57 | 0.57 | 62.4 | 0.03 |
| cg24671288 | 22 | 38,534,055 | 22q13.1 | 5.17 | 2.38E-07 | 0.71 | PLA2G6 | Intronic | 5.48 | **4.36E-08** | 1.11 | 0.27 | 2.25 | 0.02 | 0.93 | 0.35 | -0.87 | 0.39 | 63.1 | 0.03 |
| cg13427897 | 22 | 38,568,891 | 22q13.1 | 6.50 | 8.14E-11 | 0.40 | PLA2G6 | Intronic | 5.36 | **8.40E-08** | 1.73 | 0.08 | 2.63 | 8.59E-03 | 1.42 | 0.15 | -1.01 | 0.31 | 65.9 | 0.02 |
| cg16924930 | 22 | 38,570,825 | 22q13.1 | -6.66 | 2.73E-11 | 0.50 | PLA2G6 | Intronic | -5.62 | **1.91E-08** | -1.67 | 0.09 | -2.69 | 7.20E-03 | -1.44 | 0.15 | 0.99 | 0.32 | 67.8 | 0.01 |
| cg11131752 | 22 | 38,576,506 | 22q13.1 | 7.31 | 2.76E-13 | 0.56 | PLA2G6 | Intronic | 6.37 | **1.94E-10** | 1.87 | 0.06 | 2.81 | 4.99E-03 | 1.29 | 0.20 | -0.81 | 0.42 | 70.7 | 8.46E-03 |
| cg03162506 | 22 | 38,580,953 | 22q13.1 | -7.37 | 1.69E-13 | 0.54 | PLA2G6 ; MAFF | Intergenic | -6.30 | **2.98E-10** | -2.01 | 0.04 | -3.11 | 1.86E-03 | -1.39 | 0.16 | 0.39 | 0.70 | 63.4 | 0.03 |
| cg25457927 | 22 | 38,595,422 | 22q13.1 | 6.98 | 3.02E-12 | 0.60 | PLA2G6 ; MAFF | Intergenic | 5.66 | **1.52E-08** | 1.80 | 0.07 | 3.26 | 1.13E-03 | 1.22 | 0.22 | -0.35 | 0.73 | 58 | 0.05 |
| cg09984479 | 22 | 38,635,620 | 22q13.1 | -6.56 | 5.50E-11 | 0.49 | TMEM184B | Intronic | -5.69 | **1.28E-08** | -1.59 | 0.11 | -4.34 | 1.42E-05 | -0.75 | 0.45 | 0.98 | 0.33 | 76.5 | 1.89E-03 |
| cg02100011 | 22 | 38,635,773 | 22q13.1 | -5.62 | 1.97E-08 | 0.40 | TMEM184B | Intronic | -4.71 | 2.49E-06 | -1.22 | 0.22 | -4.48 | 7.43E-06 | -0.36 | 0.72 | 0.87 | 0.39 | 75 | 3.05E-03 |
| cg20648350 | 22 | 40,730,728 | 22q13.1 | -7.39 | 1.46E-13 | 0.69 | TNRC6B | 3'UTR | -6.20 | **5.49E-10** | -2.17 | 0.03 | -1.00 | 0.32 | -1.03 | 0.30 | -3.87 | 1.07E-04 | 61.5 | 0.03 |
| cg06594008 | 22 | 41,033,637 | 22q13.2 | -9.62 | 6.55E-22 | 0.40 | MRTFA | TSS1500 | -7.27 | **3.48E-13** | -2.97 | 2.94E-03 | -0.38 | 0.70 | -1.00 | 0.32 | -3.82 | 1.34E-04 | 87.3 | 2.46E-06 |
| cg06264060 | 22 | 41,075,007 | 22q13.2 | -9.30 | 1.36E-20 | 0.46 | MCHR1 | TSS200 | -6.97 | **3.27E-12** | -3.67 | 2.44E-04 | -0.86 | 0.39 | -1.69 | 0.09 | -5.08 | **3.82E-07** | 58.2 | 0.05 |
| cg14218819 | 22 | 41,075,206 | 22q13.2 | -8.64 | 5.45E-18 | 0.41 | MCHR1 | 5'UTR | -6.38 | **1.73E-10** | -3.14 | 1.69E-03 | -0.15 | 0.88 | -2.35 | 0.02 | -4.42 | 9.81E-06 | 79.5 | 6.29E-04 |
| cg04023129 | 22 | 41,460,038 | 22q13.2 | -13.71 | 9.37E-43 | 0.40 | RBX1 ; SNORD140 | Intergenic | -10.33 | **5.40E-25** | -4.23 | 2.32E-05 | -1.87 | 0.06 | -2.27 | 0.02 | -6.35 | **2.16E-10** | 69.4 | 0.01 |
| **Associations not showing significant heterogeneity across subtypes (Cochran's *P*>0.05)** | | | | | | | | | | | | | | | | | | | | |
| cg09123533 | 1 | 114,302,919 | 1p13.2 | 5.19 | 2.10E-07 | 0.39 | PHTF1 | TSS1500 | 4.90 | 9.40E-07 | 1.48 | 0.14 | 1.56 | 0.12 | -0.69 | 0.49 | 1.89 | 0.06 | 18.3 | 0.30 |
| cg01331754 | 1 | 149,909,124 | 1q21.2 | 5.86 | 4.55E-09 | 0.44 | MTMR11 ; OTUD7B | TSS1500 | 5.75 | **8.84E-09** | 1.94 | 0.05 | 0.36 | 0.72 | 0.48 | 0.63 | 2.71 | 6.76E-03 | 42.8 | 0.14 |
| cg15328025 | 1 | 155,107,693 | 1q22 | -5.28 | 1.30E-07 | 0.42 | SLC50A1 ; EFNA1 | TSS200 | -4.85 | 1.21E-06 | -2.15 | 0.03 | -2.14 | 0.03 | -0.59 | 0.55 | -0.54 | 0.59 | 2.4 | 0.39 |
| cg24589936 | 1 | 155,180,595 | 1q22 | -5.19 | 2.16E-07 | 0.64 | MTX1 | Intronic | -4.99 | 5.96E-07 | -1.81 | 0.07 | -1.69 | 0.09 | -1.84 | 0.07 | 0.31 | 0.76 | 47.8 | 0.10 |
| cg17623587 | 1 | 155,231,541 | 1q22 | 5.05 | 4.42E-07 | 0.47 | SCAMP3 | Intronic | 4.41 | 1.02E-05 | 1.44 | 0.15 | 1.75 | 0.08 | 1.23 | 0.22 | 0.04 | 0.97 | 0 | 0.53 |
| cg22041635 | 1 | 155,247,476 | 1q22 | 5.88 | 3.98E-09 | 0.37 | HCN3 | Exonic | 5.19 | **2.16E-07** | 1.29 | 0.20 | 2.14 | 0.03 | 1.47 | 0.14 | -0.52 | 0.60 | 52.5 | 0.08 |
| cg06053528 | 1 | 156,047,061 | 1q22 | 5.04 | 4.65E-07 | 0.41 | MEX3A | Exonic | 5.10 | **3.35E-07** | 1.02 | 0.31 | 1.72 | 0.08 | 0.37 | 0.71 | -0.43 | 0.67 | 49.3 | 0.10 |
| cg14124917 | 2 | 10,090,974 | 2p25.1 | -5.52 | 3.43E-08 | 0.36 | GRHL1 | TSS1500 | -4.31 | 1.62E-05 | -3.07 | 2.12E-03 | -3.02 | 2.54E-03 | -0.20 | 0.84 | -1.03 | 0.30 | 18.4 | 0.30 |
| cg20393901 | 2 | 19,320,934 | 2p24.1 | 5.63 | 1.80E-08 | 0.59 | LINC01376 ; MIR4757 | Intergenic | 2.42 | 0.02 | 0.59 | 0.55 | 2.99 | 2.78E-03 | 1.36 | 0.18 | 4.19 | 2.81E-05 | 55.6 | 0.06 |
| cg15020926 | 2 | 25,097,091 | 2p23.3 | 6.23 | 4.58E-10 | 0.54 | ADCY3 | Intronic | 3.45 | 5.55E-04 | 2.86 | 4.21E-03 | 0.12 | 0.90 | 3.11 | 1.90E-03 | 2.52 | 0.01 | 54.6 | 0.07 |
| cg27147322 | 2 | 25,136,820 | 2p23.3 | -6.32 | 2.66E-10 | 0.53 | ADCY3 | Intronic | -3.44 | 5.90E-04 | -2.80 | 5.06E-03 | -0.03 | 0.97 | -3.17 | 1.51E-03 | -2.67 | 7.68E-03 | 55.4 | 0.06 |
| cg19602045 | 2 | 25,138,079 | 2p23.3 | -6.63 | 3.37E-11 | 0.65 | ADCY3 | Intronic | -3.64 | 2.75E-04 | -2.71 | 6.74E-03 | -0.47 | 0.64 | -3.47 | 5.19E-04 | -2.80 | 5.06E-03 | 56.4 | 0.06 |
| cg20518994 | 2 | 25,141,532 | 2p23.3 | -6.52 | 6.98E-11 | 0.71 | ADCY3 | Exonic | -3.64 | 2.76E-04 | -2.82 | 4.79E-03 | -0.59 | 0.55 | -2.99 | 2.75E-03 | -3.10 | 1.91E-03 | 46.4 | 0.11 |
| cg15423357 | 2 | 25,149,977 | 2p23.3 | -6.06 | 1.40E-09 | 0.38 | ADCY3 ; DNAJC27 | Intergenic | -3.25 | 1.17E-03 | -2.59 | 9.51E-03 | -0.24 | 0.81 | -2.61 | 9.01E-03 | -3.09 | 1.97E-03 | 43.8 | 0.13 |
| cg01884057 | 2 | 25,150,051 | 2p23.3 | -6.42 | 1.39E-10 | 0.52 | ADCY3 ; DNAJC27 | Intergenic | -3.67 | 2.39E-04 | -2.80 | 5.16E-03 | -0.65 | 0.51 | -2.88 | 4.04E-03 | -2.88 | 3.95E-03 | 37.6 | 0.17 |
| cg11218383 | 2 | 25,233,015 | 2p23.3 | 6.02 | 1.79E-09 | 0.58 | DNAJC27-AS1 | Intronic | 3.53 | 4.09E-04 | 2.67 | 7.49E-03 | 1.38 | 0.17 | 1.89 | 0.06 | 3.59 | 3.31E-04 | 6.3 | 0.37 |
| cg20303441 | 2 | 25,496,390 | 2p23.3 | -5.22 | 1.83E-07 | 0.43 | DNMT3A | Intronic | -2.86 | 4.28E-03 | -2.02 | 0.04 | -1.26 | 0.21 | -1.77 | 0.08 | -2.08 | 0.04 | 0 | 0.85 |
| g23178958 | 2 | 69,387,295 | 2p13.3 | -5.56 | 2.77E-08 | 0.38 | ANTXR1 | Intronic | -3.32 | 9.02E-04 | -1.92 | 0.06 | -2.90 | 3.76E-03 | -2.75 | 6.00E-03 | -2.17 | 0.03 | 0 | 0.50 |
| cg08796252 | 2 | 172,378,641 | 2q31.1 | 5.71 | 1.16E-08 | 0.70 | CYBRD1 | TSS200 | 5.02 | 5.26E-07 | 1.80 | 0.07 | 3.19 | 1.44E-03 | -1.18 | 0.24 | 0.94 | 0.35 | 57 | 0.05 |
| cg22391094 | 2 | 201,398,514 | 2q33.1 | -5.63 | 1.76E-08 | 0.38 | SGO2 | Intronic | -3.53 | 4.18E-04 | -1.71 | 0.09 | -2.01 | 0.04 | 0.77 | 0.44 | -4.09 | 4.39E-05 | 48.1 | 0.10 |
| cg21017626 | 2 | 201,715,305 | 2q33.1 | 5.60 | 2.13E-08 | 0.42 | BZW1 ; CLK1 | Intergenic | 3.89 | 1.02E-04 | 1.04 | 0.30 | 2.16 | 0.03 | 0.28 | 0.78 | 2.40 | 0.02 | 0 | 0.89 |
| cg03277598 | 2 | 202,004,869 | 2q33.1 | -7.79 | 6.77E-15 | 0.48 | CFLAR | Intronic | -3.24 | 1.20E-03 | -3.03 | 2.44E-03 | -1.85 | 0.06 | -0.40 | 0.69 | -4.38 | 1.16E-05 | 47.9 | 0.10 |
| cg19448993 | 2 | 202,097,129 | 2q33.1 | 5.56 | 2.63E-08 | 0.41 | CASP10 ; CASP8 | Intergenic | 1.41 | 0.16 | 2.00 | 0.05 | 1.65 | 0.10 | 0.06 | 0.95 | 3.13 | 1.73E-03 | 30.4 | 0.22 |
| cg19494818 | 2 | 202,561,266 | 2q33.1 | 5.84 | 5.20E-09 | 0.50 | MPP4 | Intronic | 2.96 | 3.04E-03 | 2.26 | 0.02 | 1.36 | 0.17 | 0.33 | 0.75 | 2.82 | 4.84E-03 | 0 | 0.64 |
| cg21884962 | 2 | 218,225,246 | 2q35 | -5.39 | 7.21E-08 | 0.45 | DIRC3 | Intronic | -5.71 | **1.14E-08** | -1.43 | 0.15 | -1.61 | 0.11 | -0.72 | 0.47 | -0.54 | 0.59 | 48.5 | 0.10 |
| cg07152837 | 2 | 218,438,884 | 2q35 | -5.99 | 2.08E-09 | 0.42 | DIRC3 | Intronic | -5.24 | **1.57E-07** | -3.41 | 6.41E-04 | -1.05 | 0.29 | -2.00 | 0.05 | -1.73 | 0.08 | 9.2 | 0.35 |
| cg09509293 | 3 | 16,783,065 | 3p24.3 | -5.88 | 4.09E-09 | 0.60 | DAZL ; PLCL2 | Intergenic | -4.08 | 4.58E-05 | -1.07 | 0.29 | -1.24 | 0.22 | -1.62 | 0.11 | -2.13 | 0.03 | 0 | 0.86 |
| cg26263234 | 3 | 26,666,060 | 3p24.1 | 6.22 | 4.85E-10 | 0.49 | LRRC3B | Intronic | 4.70 | 2.65E-06 | 0.46 | 0.65 | 1.65 | 0.10 | -0.69 | 0.49 | 0.94 | 0.35 | 40.9 | 0.15 |
| cg22415969 | 3 | 27,411,065 | 3p24.1 | -5.29 | 1.24E-07 | 0.43 | NEK10 | TSS200 | -5.90 | **3.62E-09** | -1.57 | 0.12 | -1.05 | 0.29 | -1.82 | 0.07 | -1.54 | 0.12 | 19.3 | 0.29 |
| cg09563400 | 3 | 27,509,190 | 3p24.1 | -5.51 | 3.65E-08 | 0.50 | SLC4A7 | Intronic | -5.89 | **3.88E-09** | -1.51 | 0.13 | -2.18 | 0.03 | -0.88 | 0.38 | -0.31 | 0.76 | 55.3 | 0.06 |
| cg20739013 | 3 | 27,761,677 | 3p24.1 | -6.51 | 7.54E-11 | 0.51 | EOMES | Exonic | -5.07 | **4.03E-07** | -1.93 | 0.05 | -1.85 | 0.06 | -1.31 | 0.19 | -1.12 | 0.26 | 0 | 0.74 |
| cg22309949 | 3 | 46,792,192 | 3p21.31 | 5.32 | 1.06E-07 | 0.68 | PRSS45P ; PRSS43P | Intergenic | 2.84 | 4.57E-03 | 2.81 | 4.92E-03 | 3.80 | 1.47E-04 | 0.59 | 0.56 | 3.44 | 5.90E-04 | 46.1 | 0.12 |
| cg23231268 | 3 | 46,792,462 | 3p21.31 | 5.43 | 5.70E-08 | 0.44 | PRSS45P ; PRSS43P | Intergenic | 2.86 | 4.24E-03 | 2.70 | 6.85E-03 | 4.14 | 3.40E-05 | 0.90 | 0.37 | 3.19 | 1.41E-03 | 46.4 | 0.11 |
| cg13182339 | 3 | 64,058,252 | 3p14.1 | -5.81 | 6.36E-09 | 0.41 | PRICKLE2-AS1 | Intronic | -3.95 | 7.89E-05 | -2.47 | 0.01 | -1.14 | 0.25 | -1.04 | 0.30 | -1.67 | 0.10 | 0 | 0.84 |
| cg16276021 | 3 | 99,015,077 | 3q12.1 | -6.59 | 4.37E-11 | 0.50 | DCBLD2 ; COL8A1 | Intergenic | -4.44 | 9.02E-06 | -2.65 | 8.12E-03 | -2.09 | 0.04 | -0.65 | 0.51 | -1.19 | 0.23 | 0 | 0.72 |
| cg03021388 | 3 | 99,565,499 | 3q12.1 | 7.69 | 1.52E-14 | 0.46 | CMSS1 ; FILIP1L | Intronic | 4.96 | 6.94E-07 | 2.23 | 0.03 | 3.41 | 6.53E-04 | 0.74 | 0.46 | 0.82 | 0.41 | 0.5 | 0.40 |
| cg21419939 | 3 | 99,598,918 | 3q12.1 | 6.10 | 1.07E-09 | 0.47 | CMSS1 ; FILIP1L | Intronic | 5.83 | **5.59E-09** | 0.89 | 0.37 | 2.21 | 0.03 | -0.84 | 0.40 | 1.23 | 0.22 | 55.9 | 0.06 |
| cg16808912 | 3 | 99,603,495 | 3q12.1 | 6.55 | 5.75E-11 | 0.49 | CMSS1 ; FILIP1L | Intronic | 5.15 | **2.66E-07** | 1.20 | 0.23 | 1.38 | 0.17 | 0.25 | 0.80 | 0.72 | 0.47 | 17.6 | 0.30 |
| cg05878887 | 3 | 99,620,389 | 3q12.1 | 5.99 | 2.15E-09 | 0.53 | CMSS1 ; FILIP1L | Intronic | 4.99 | 6.13E-07 | 2.20 | 0.03 | 1.72 | 0.09 | 0.40 | 0.69 | 0.85 | 0.40 | 10.8 | 0.34 |
| cg17983017 | 3 | 99,649,860 | 3q12.1 | -6.92 | 4.52E-12 | 0.49 | FILIP1L | Exonic | -4.25 | 2.10E-05 | -1.76 | 0.08 | -3.27 | 1.08E-03 | -0.12 | 0.91 | -1.17 | 0.24 | 0 | 0.50 |
| cg26581146 | 3 | 99,654,279 | 3q12.1 | 6.31 | 2.81E-10 | 0.39 | CMSS1 ; FILIP1L | Intronic | 3.47 | 5.17E-04 | 1.94 | 0.05 | 3.40 | 6.73E-04 | 0.60 | 0.55 | 0.65 | 0.52 | 1.5 | 0.40 |
| cg17560007 | 3 | 99,881,060 | 3q12.1 | -5.49 | 4.06E-08 | 0.44 | CMSS1 | Intronic | -3.09 | 2.01E-03 | -1.38 | 0.17 | -2.28 | 0.02 | 0.42 | 0.68 | -0.25 | 0.81 | 0 | 0.43 |
| cg04730047 | 3 | 99,979,355 | 3q12.1 | 6.42 | 1.37E-10 | 0.51 | TBC1D23 | TSS1500 | 3.53 | 4.16E-04 | 1.00 | 0.32 | 2.02 | 0.04 | 0.55 | 0.58 | 0.39 | 0.70 | 0 | 0.71 |
| cg19019371 | 3 | 140,770,984 | 3q23 | -5.44 | 5.32E-08 | 0.38 | SPSB4 | TSS1500 | -4.27 | 1.99E-05 | -1.83 | 0.07 | -3.04 | 2.39E-03 | -0.99 | 0.32 | -2.25 | 0.02 | 0 | 0.95 |
| cg00563531 | 3 | 140,834,923 | 3q23 | -6.12 | 9.16E-10 | 0.53 | SPSB4 | Intronic | -3.58 | 3.40E-04 | -3.15 | 1.64E-03 | -3.65 | 2.67E-04 | -1.17 | 0.24 | -0.90 | 0.37 | 0 | 0.44 |
| cg18030972 | 3 | 140,993,197 | 3q23 | -5.25 | 1.49E-07 | 0.41 | PXYLP1 | Intronic | -3.00 | 2.67E-03 | -2.08 | 0.04 | -3.61 | 3.09E-04 | -1.20 | 0.23 | -2.83 | 4.61E-03 | 23.6 | 0.26 |
| cg10066494 | 3 | 141,150,514 | 3q23 | -5.33 | 9.64E-08 | 0.53 | ZBTB38 | Intronic | -4.48 | 7.62E-06 | -1.69 | 0.09 | -3.44 | 5.75E-04 | -2.58 | 9.79E-03 | 0.27 | 0.79 | 0 | 0.41 |
| cg20054786 | 4 | 38,799,406 | 4p14 | 5.27 | 1.39E-07 | 0.42 | TLR1 | Exonic | 3.24 | 1.19E-03 | 1.48 | 0.14 | 1.06 | 0.29 | -0.03 | 0.97 | 0.14 | 0.89 | 0 | 0.54 |
| cg12444746 | 4 | 126,572,264 | 4q28.1 | 5.26 | 1.43E-07 | 0.47 | MIR2054 ; INTU | Intergenic | 2.93 | 3.38E-03 | 1.50 | 0.13 | 1.35 | 0.18 | 1.71 | 0.09 | 1.86 | 0.06 | 0 | 0.94 |
| g13529678 | 4 | 151,181,171 | 4q31.3 | -5.10 | 3.38E-07 | 0.67 | DCLK2 ; LRBA | Intergenic | -3.84 | 1.22E-04 | -0.69 | 0.49 | -1.49 | 0.14 | -2.29 | 0.02 | -3.04 | 2.33E-03 | 0 | 0.45 |
| g26218269 | 4 | 151,503,227 | 4q31.3 | 5.09 | 3.57E-07 | 0.51 | LRBA | Intronic | 4.16 | 3.19E-05 | 0.62 | 0.54 | 0.95 | 0.34 | 2.37 | 0.02 | 3.80 | 1.44E-04 | 42.2 | 0.14 |
| cg21623028 | 5 | 420,443 | 5p15.33 | -5.16 | 2.53E-07 | 0.45 | AHRR | Intronic | -2.75 | 5.99E-03 | -1.91 | 0.06 | -2.51 | 0.01 | -0.76 | 0.45 | -1.96 | 0.05 | 0 | 0.88 |
| cg27311041 | 5 | 1,300,340 | 5p15.33 | -6.67 | 2.64E-11 | 0.47 | TERT ; MIR4457 | Intergenic | -5.06 | **4.10E-07** | -2.04 | 0.04 | -3.24 | 1.21E-03 | -1.91 | 0.06 | -2.84 | 4.49E-03 | 0 | 0.98 |
| cg06782035 | 5 | 16,179,135 | 5p15.1 | -5.38 | 7.27E-08 | 0.45 | MARCHF11 | Intronic | -4.31 | 1.64E-05 | -2.39 | 0.02 | -1.65 | 0.10 | -1.28 | 0.20 | -2.54 | 0.01 | 0 | 0.95 |
| cg06760839 | 5 | 16,742,666 | 5p15.1 | -5.34 | 9.25E-08 | 0.57 | MYO10 | Intronic | -4.41 | 1.04E-05 | -2.34 | 0.02 | -1.02 | 0.31 | -1.46 | 0.14 | -2.15 | 0.03 | 0 | 0.70 |
| g20242213 | 5 | 44,774,282 | 5p12 | 7.97 | 1.64E-15 | 0.49 | MRPS30-DT | Intronic | 5.75 | **8.70E-09** | 2.06 | 0.04 | 3.05 | 2.25E-03 | 1.65 | 0.10 | 2.98 | 2.86E-03 | 0 | 0.99 |
| cg11557886 | 5 | 44,811,891 | 5p12 | -8.84 | 9.48E-19 | 0.49 | MRPS30 | Intronic | -8.59 | **8.40E-18** | -1.45 | 0.15 | -2.96 | 3.12E-03 | -1.06 | 0.29 | -2.86 | 4.25E-03 | 38.9 | 0.16 |
| cg14979674 | 5 | 45,260,623 | 5p12 | 5.40 | 6.83E-08 | 0.42 | HCN1 | 3'UTR | 4.09 | 4.30E-05 | 3.78 | 1.59E-04 | 1.26 | 0.21 | 1.22 | 0.22 | 2.25 | 0.02 | 5.4 | 0.38 |
| cg10471743 | 5 | 49,739,158 | 5q11.1 | 5.07 | 3.88E-07 | 0.43 | EMB ; PARP8 | Intergenic | 2.16 | 0.03 | 1.80 | 0.07 | 3.49 | 4.91E-04 | 1.72 | 0.09 | 3.00 | 2.74E-03 | 36.1 | 0.18 |
| cg16524385 | 5 | 50,103,490 | 5q11.1 | 5.49 | 4.12E-08 | 0.49 | PARP8 | Intronic | 3.28 | 1.03E-03 | 1.01 | 0.31 | 3.27 | 1.09E-03 | 1.69 | 0.09 | 2.11 | 0.03 | 0 | 0.62 |
| cg06009016 | 5 | 56,135,074 | 5q11.2 | 6.21 | 5.18E-10 | 0.57 | MAP3K1 | Intronic | 6.91 | **4.87E-12** | 2.16 | 0.03 | 1.33 | 0.18 | 1.24 | 0.21 | 0.85 | 0.40 | 53.5 | 0.07 |
| cg22565248 | 5 | 81,051,199 | 5q14.1 | -6.11 | 1.01E-09 | 0.45 | SSBP2 ; ATG10 | Intergenic | -5.75 | **9.07E-09** | -1.27 | 0.20 | -2.42 | 0.02 | -0.64 | 0.52 | -0.48 | 0.63 | 31 | 0.21 |
| cg15871215 | 5 | 81,402,204 | 5q14.2 | 5.73 | 1.02E-08 | 0.50 | ATG10 | Intronic | 3.67 | 2.43E-04 | 1.38 | 0.17 | 2.22 | 0.03 | -0.96 | 0.34 | 2.37 | 0.02 | 14.3 | 0.32 |
| cg18035979 | 5 | 81,575,199 | 5q14.2 | 5.29 | 1.22E-07 | 0.40 | ATP6AP1L | 5'UTR | 4.63 | 3.74E-06 | 0.20 | 0.84 | 2.11 | 0.03 | -0.02 | 0.98 | 0.68 | 0.50 | 30.4 | 0.23 |
| cg00402173 | 5 | 81,709,024 | 5q14.2 | -5.05 | 4.37E-07 | 0.58 | ATP6AP1L ; MIR3977 | Intergenic | -3.66 | 2.51E-04 | -0.97 | 0.33 | -1.88 | 0.06 | -1.10 | 0.27 | -1.11 | 0.27 | 0 | 0.94 |
| cg16465129 | 5 | 131,408,149 | 5q31.1 | 5.13 | 2.95E-07 | 0.52 | IL3 ; CSF2 | Intergenic | 3.90 | 9.77E-05 | 2.69 | 7.12E-03 | 1.98 | 0.05 | -0.02 | 0.99 | 0.48 | 0.63 | 31 | 0.22 |
| cg24502014 | 5 | 131,563,623 | 5q31.1 | -5.04 | 4.59E-07 | 0.37 | P4HA2 | TSS200 | -2.60 | 9.19E-03 | -2.97 | 2.95E-03 | -2.08 | 0.04 | 0.12 | 0.90 | -0.81 | 0.42 | 25.3 | 0.26 |
| cg11033092 | 5 | 131,564,679 | 5q31.1 | 6.17 | 6.99E-10 | 0.55 | P4HA2 ; PDLIM4 | Intergenic | 4.31 | 1.61E-05 | 2.92 | 3.46E-03 | 2.96 | 3.04E-03 | -0.75 | 0.45 | 0.76 | 0.44 | 48.9 | 0.10 |
| cg25550088 | 5 | 131,573,526 | 5q31.1 | -5.03 | 4.96E-07 | 0.50 | P4HA2 ; PDLIM4 | Intergenic | -3.73 | 1.93E-04 | -2.47 | 0.01 | -2.35 | 0.02 | 0.62 | 0.54 | -0.60 | 0.55 | 26.4 | 0.25 |
| cg12564285 | 5 | 131,593,104 | 5q31.1 | 5.89 | 3.97E-09 | 0.53 | PDLIM4 | TSS1500 | 4.52 | 6.09E-06 | 2.74 | 6.13E-03 | 2.70 | 6.91E-03 | -1.09 | 0.28 | 1.49 | 0.14 | 44.9 | 0.12 |
| cg04518342 | 5 | 131,593,106 | 5q31.1 | 6.13 | 8.70E-10 | 0.59 | PDLIM4 | TSS1500 | 4.56 | 5.16E-06 | 2.91 | 3.61E-03 | 2.77 | 5.61E-03 | -0.96 | 0.34 | 1.67 | 0.10 | 41.8 | 0.14 |
| cg18301423 | 5 | 131,593,218 | 5q31.1 | 5.84 | 5.26E-09 | 0.50 | PDLIM4 | TSS200 | 4.43 | 9.47E-06 | 2.93 | 3.41E-03 | 2.57 | 0.01 | -0.94 | 0.35 | 1.54 | 0.12 | 40.6 | 0.15 |
| cg18758796 | 5 | 131,593,413 | 5q31.1 | 6.42 | 1.38E-10 | 0.54 | PDLIM4 | 5'UTR | 4.79 | 1.70E-06 | 3.09 | 2.00E-03 | 2.84 | 4.53E-03 | -0.83 | 0.41 | 1.54 | 0.12 | 43.3 | 0.13 |
| cg26229274 | 5 | 131,629,510 | 5q31.1 | -6.04 | 1.50E-09 | 0.38 | SLC22A4 | TSS1500 | -4.23 | 2.31E-05 | -3.54 | 3.98E-04 | -2.22 | 0.03 | -0.41 | 0.68 | -0.24 | 0.81 | 47.4 | 0.11 |
| cg13654253 | 5 | 131,629,910 | 5q31.1 | -5.07 | 3.92E-07 | 0.45 | SLC22A4 | TSS1500 | -3.90 | 9.60E-05 | -2.83 | 4.66E-03 | -1.59 | 0.11 | 0.73 | 0.47 | -0.85 | 0.40 | 33.7 | 0.20 |
| cg17111895 | 5 | 131,630,387 | 5q31.1 | -5.31 | 1.09E-07 | 0.39 | SLC22A4 | Exonic | -3.33 | 8.56E-04 | -3.19 | 1.45E-03 | -2.61 | 9.17E-03 | -1.71 | 0.09 | -0.21 | 0.83 | 35.3 | 0.19 |
| cg05231888 | 5 | 131,630,754 | 5q31.1 | -5.70 | 1.23E-08 | 0.48 | SLC22A4 | Intronic | -3.18 | 1.49E-03 | -3.06 | 2.22E-03 | -1.83 | 0.07 | -0.13 | 0.90 | 0.24 | 0.81 | 41.4 | 0.15 |
| cg14622915 | 5 | 131,657,711 | 5q31.1 | 5.15 | 2.59E-07 | 0.50 | MIR3936HG | Intronic | 2.50 | 0.01 | 3.33 | 8.58E-04 | 1.45 | 0.15 | -0.02 | 0.98 | -0.40 | 0.69 | 58.5 | 0.07 |

| | | | | | | | | | | | | | | | | | | | | |
|---|---|---|---|---|---|---|---|---|---|---|---|---|---|---|---|---|---|---|---|---|
| cg1765083 | 5 | 131,679,878 | 5q31.1 | -5.20 | 1.98E-07 | 0.41 | *MIR3936HG* | Intronic | -2.87 | 4.13E-03 | -3.24 | 1.20E-03 | -2.13 | 0.03 | -0.82 | 0.41 | 0.71 | 0.48 | 54.6 | 0.07 |
| cg0753894 | 5 | 131,705,188 | 5q31.1 | 5.99 | 2.07E-09 | 0.50 | *MIR3936HG* | Intronic | 4.14 | 3.41E-05 | 2.84 | 4.57E-03 | 2.22 | 0.03 | -0.22 | 0.82 | 0.25 | 0.80 | 36.8 | 0.18 |
| cg0651653 | 5 | 131,732,471 | 5q31.1 | 6.51 | 7.43E-11 | 0.39 | *SLC22A5* ; *C5orf56* | Intergenic | 3.70 | 2.19E-04 | 3.64 | 2.77E-04 | 2.51 | 0.01 | 0.50 | 0.62 | 0.04 | 0.97 | 51.2 | 0.08 |
| cg2037302 | 5 | 131,746,552 | 5q31.1 | 6.01 | 1.88E-09 | 0.53 | *C5orf56* | TSS200 | 3.82 | 1.35E-04 | 3.31 | 9.38E-04 | 2.32 | 0.02 | 0.00 | 1.00 | 0.54 | 0.59 | 36.8 | 0.19 |
| cg2113840 | 5 | 131,827,807 | 5q31.1 | -5.08 | 3.69E-07 | 0.47 | *IRF1* ; *IL5* | Intergenic | -2.94 | 3.30E-03 | -3.70 | 2.19E-04 | -1.71 | 0.09 | -1.39 | 0.17 | 0.33 | 0.74 | 57.1 | 0.05 |
| cg1168837 | 5 | 132,082,633 | 5q31.1 | -5.39 | 7.03E-08 | 0.46 | *CCNI2* | TSS1500 | -4.79 | 1.67E-06 | -2.13 | 0.03 | -1.25 | 0.21 | 0.37 | 0.71 | -0.06 | 0.95 | 39.1 | 0.16 |
| cg2007568 | 5 | 132,083,910 | 5q31.1 | -5.36 | 8.11E-08 | 0.40 | *CCNI2* | Intronic | -4.39 | 1.12E-05 | -1.78 | 0.08 | -1.28 | 0.20 | 0.42 | 0.67 | 0.06 | 0.95 | 38.4 | 0.17 |
| cg1038608 | 5 | 158,313,279 | 5q33.3 | 9.71 | 2.60E-22 | 0.45 | *EBF1* | Intronic | 6.05 | **1.43E-09** | 2.54 | 0.01 | 3.21 | 1.34E-03 | 2.34 | 0.02 | 3.27 | 1.09E-03 | 0 | 0.99 |
| cg0781072 | 5 | 158,524,649 | 5q33.3 | -5.76 | 8.28E-09 | 0.62 | *EBF1* | 5'UTR | -4.56 | 5.13E-06 | -3.51 | 4.53E-04 | -1.99 | 0.05 | 0.23 | 0.82 | -3.09 | 1.98E-03 | 22.5 | 0.27 |
| cg1596356 | 5 | 158,528,040 | 5q33.3 | -5.34 | 9.32E-08 | 0.51 | *LINC02202* | Intronic | -3.60 | 3.21E-04 | -2.77 | 5.59E-03 | -1.71 | 0.09 | -0.50 | 0.62 | -2.51 | 0.01 | 0 | 0.74 |
| cg1005443 | 5 | 158,637,265 | 5q33.3 | -5.61 | 2.01E-08 | 0.52 | *RNF145* | TSS1500 | -2.71 | 6.75E-03 | -0.66 | 0.51 | -1.98 | 0.05 | -3.28 | 1.05E-03 | -1.91 | 0.06 | 35.5 | 0.18 |
| g03611078 | 6 | 4,087,591 | 6p25.2 | 5.19 | 2.15E-07 | 0.45 | *C6orf201* | 5'UTR | 3.20 | 1.39E-03 | -0.58 | 0.56 | 1.34 | 0.18 | 0.41 | 0.68 | 2.74 | 6.13E-03 | 37.1 | 0.17 |
| cg0114221 | 6 | 13,282,393 | 6p24.1 | -5.31 | 1.08E-07 | 0.50 | *LOC100130357* | Intronic | -5.58 | **2.46E-08** | -1.60 | 0.11 | -2.13 | 0.03 | -0.58 | 0.56 | -0.68 | 0.50 | 0 | 0.44 |
| cg1148686 | 6 | 27,436,870 | 6p22.1 | -5.06 | 4.11E-07 | 0.45 | *ZNF184* | Intronic | -3.79 | 1.51E-04 | -0.03 | 0.98 | -0.61 | 0.54 | -1.19 | 0.23 | -0.68 | 0.50 | 1.7 | 0.40 |
| cg1087752 | 6 | 28,480,871 | 6p22.1 | 6.11 | 1.03E-09 | 0.41 | *GPX6* | Intronic | 3.52 | 4.34E-04 | 1.19 | 0.23 | -0.11 | 0.91 | 1.37 | 0.17 | 2.70 | 6.93E-03 | 5.2 | 0.38 |
| cg2614567 | 6 | 28,554,497 | 6p22.1 | -5.83 | 5.49E-09 | 0.52 | *ZBED9* | 5'UTR | -3.67 | 2.42E-04 | -1.64 | 0.10 | -1.35 | 0.18 | -1.22 | 0.22 | -1.36 | 0.17 | 0 | 0.87 |
| cg1436885 | 6 | 28,732,188 | 6p22.1 | -5.77 | 7.95E-09 | 0.50 | *LINC00533* ; *LINC0162* | Intergenic | -2.70 | 6.92E-03 | 0.10 | 0.92 | -0.20 | 0.84 | -0.49 | 0.62 | -3.57 | 3.53E-04 | 51.2 | 0.08 |
| cg2232825 | 6 | 28,757,381 | 6p22.1 | -8.30 | 1.05E-16 | 0.41 | *LINC00533* ; *LINC0162* | Intergenic | -4.73 | 2.22E-06 | -1.12 | 0.26 | -0.63 | 0.53 | -1.40 | 0.16 | -3.37 | 7.53E-04 | 4.1 | 0.38 |
| cg1402840 | 6 | 28,793,656 | 6p22.1 | -5.08 | 3.69E-07 | 0.44 | *LINC00533* ; *LINC0162* | Intergenic | -3.47 | 5.12E-04 | -1.46 | 0.14 | -0.30 | 0.77 | -1.93 | 0.05 | -2.44 | 0.01 | 0 | 0.54 |
| cg0246597 | 6 | 28,872,035 | 6p22.1 | -5.47 | 4.38E-08 | 0.49 | *TRIM27* | Exonic | -4.04 | 5.30E-05 | -1.01 | 0.31 | 0.85 | 0.39 | -0.14 | 0.89 | -1.36 | 0.17 | 46.8 | 0.11 |
| cg2284208 | 6 | 28,880,239 | 6p22.1 | -5.82 | 6.00E-09 | 0.60 | *TRIM27* | Intronic | -3.23 | 1.25E-03 | -1.08 | 0.28 | 0.51 | 0.61 | -0.83 | 0.40 | -2.73 | 6.29E-03 | 48.4 | 0.10 |
| cg0060996 | 6 | 28,911,199 | 6p22.1 | -5.26 | 1.43E-07 | 0.46 | *LINC01556* | TSS1500 | -3.27 | 1.08E-03 | -0.36 | 0.72 | 0.70 | 0.48 | -0.77 | 0.44 | -3.03 | 2.42E-03 | 50.6 | 0.09 |
| cg2052766 | 6 | 28,953,824 | 6p22.1 | -5.41 | 6.43E-08 | 0.45 | *HCG15* | Exonic | -3.58 | 3.39E-04 | -1.96 | 0.05 | 0.79 | 0.43 | -1.03 | 0.30 | -1.36 | 0.17 | 44 | 0.13 |
| cg1938057 | 6 | 28,968,973 | 6p22.1 | -6.54 | 5.97E-11 | 0.47 | *ZNF311* | Intronic | -3.83 | 1.26E-04 | -1.22 | 0.22 | -0.02 | 0.98 | -1.21 | 0.23 | -1.99 | 0.05 | 8.1 | 0.36 |
| cg2487696 | 6 | 28,971,902 | 6p22.1 | 5.45 | 5.08E-08 | 0.58 | *ZNF311* | Intronic | 2.80 | 5.19E-03 | 0.38 | 0.70 | -0.36 | 0.72 | 0.38 | 0.70 | 3.69 | 2.28E-04 | 55.1 | 0.06 |
| cg2704575 | 6 | 28,971,967 | 6p22.1 | 5.65 | 1.63E-08 | 0.75 | *ZNF311* | Intronic | 3.21 | 1.34E-03 | 0.50 | 0.62 | -0.47 | 0.64 | 0.43 | 0.66 | 3.48 | 5.04E-04 | 53.4 | 0.07 |
| cg2671247 | 6 | 28,984,445 | 6p22.1 | -5.25 | 1.51E-07 | 0.48 | *ZNF311* ; *LOC100129636* | Intergenic | -2.97 | 2.97E-03 | -0.65 | 0.52 | 0.39 | 0.70 | -0.54 | 0.59 | -2.49 | 0.01 | 16.6 | 0.31 |
| cg2086225 | 6 | 29,054,523 | 6p22.1 | 5.91 | 3.34E-09 | 0.45 | *OR2B3* | Exonic | 2.10 | 0.04 | 0.24 | 0.81 | 1.14 | 0.25 | 1.33 | 0.18 | 4.19 | 2.74E-05 | 51.3 | 0.08 |
| cg0506815 | 6 | 29,408,587 | 6p22.1 | 5.20 | 2.00E-07 | 0.43 | *OR10C1* | Exonic | 2.98 | 2.84E-03 | 0.55 | 0.58 | -0.35 | 0.73 | 0.08 | 0.94 | 2.98 | 2.89E-03 | 38.5 | 0.16 |
| cg0102479 | 6 | 29,524,067 | 6p22.1 | 5.88 | 4.16E-09 | 0.59 | *UBD* | Exonic | 3.59 | 3.26E-04 | 1.07 | 0.29 | 1.58 | 0.11 | 0.88 | 0.38 | 2.57 | 0.01 | 0 | 0.92 |
| cg2358568 | 6 | 82,161,324 | 6q14.1 | -6.35 | 2.13E-10 | 0.53 | *BCKDHB* ; *TENT5A* | Intergenic | -5.50 | **3.83E-08** | -2.54 | 0.01 | -3.09 | 1.99E-03 | -1.90 | 0.06 | -2.89 | 3.85E-03 | 0 | 1.00 |
| g01444058 | 6 | 151,558,900 | 6q25.1 | -7.35 | 2.04E-13 | 0.55 | *LOC102723831* ; *AKAP12* | Intergenic | -3.79 | 1.48E-04 | -3.02 | 2.51E-03 | -3.66 | 2.53E-04 | -1.67 | 0.09 | -4.85 | 1.23E-06 | 49.2 | 0.10 |
| cg0858631 | 6 | 151,560,577 | 6q25.1 | -7.74 | 9.62E-15 | 0.51 | *AKAP12* | TSS1500 | -4.04 | 5.25E-05 | -2.80 | 5.12E-03 | -3.53 | 4.10E-04 | -1.58 | 0.12 | -5.10 | **3.37E-07** | 48.3 | 0.10 |
| cg2407533 | 6 | 151,712,594 | 6q25.1 | -5.39 | 7.11E-08 | 0.45 | *ZBTB2* | 5'UTR | -2.56 | 0.01 | -1.46 | 0.15 | -2.02 | 0.04 | 0.39 | 0.70 | -2.78 | 5.49E-03 | 0 | 0.45 |
| cg2073069 | 6 | 151,803,839 | 6q25.1 | 6.95 | 3.75E-12 | 0.44 | *ARMT1* ; *CCDC170* | Intergenic | 2.35 | 0.02 | 1.99 | 0.05 | 1.51 | 0.13 | 2.26 | 0.02 | 3.42 | 6.22E-04 | 30.2 | 0.22 |
| g06611115 | 6 | 152,011,200 | 6q25.1 | -8.95 | 3.60E-19 | 0.44 | *ESR1* | TSS1500 | -4.12 | 3.83E-05 | -3.28 | 1.04E-03 | -3.36 | 7.76E-04 | -2.43 | 0.02 | -5.00 | 5.79E-07 | 49.6 | 0.09 |
| cg2211717 | 7 | 91,764,530 | 7q21.2 | 5.72 | 1.07E-08 | 0.44 | *CYP51A1-AS1* | Intronic | 3.53 | 4.12E-04 | 3.08 | 2.07E-03 | 1.67 | 0.10 | 1.13 | 0.26 | 1.22 | 0.22 | 0 | 0.62 |
| cg0371477 | 7 | 91,764,589 | 7q21.2 | 5.70 | 1.19E-08 | 0.41 | *CYP51A1-AS1* | Intronic | 3.51 | 4.55E-04 | 3.03 | 2.46E-03 | 1.65 | 0.10 | 1.36 | 0.17 | 1.25 | 0.21 | 0 | 0.65 |
| cg0168965 | 7 | 91,764,605 | 7q21.2 | 6.41 | 1.44E-10 | 0.37 | *CYP51A1-AS1* | Intronic | 3.95 | 7.69E-05 | 3.09 | 1.98E-03 | 2.08 | 0.04 | 1.55 | 0.12 | 1.30 | 0.19 | 0 | 0.67 |
| cg2628356 | 7 | 94,284,274 | 7q21.3 | 6.29 | 3.20E-10 | 0.40 | *SGCE* | Intronic | 4.71 | 2.42E-06 | 2.94 | 3.23E-03 | 3.20 | 1.39E-03 | 0.91 | 0.36 | -0.36 | 0.72 | 54.8 | 0.06 |
| g03637179 | 7 | 99,716,580 | 7q22.1 | -5.45 | 5.14E-08 | 0.36 | *TAF6* | 5'UTR | -2.78 | 5.47E-03 | -2.29 | 0.02 | -2.29 | 0.02 | -1.21 | 0.22 | -1.63 | 0.10 | 0 | 0.88 |
| g20272595 | 7 | 99,717,190 | 7q22.1 | -5.23 | 1.66E-07 | 0.44 | *TAF6* | Intronic | -2.77 | 5.64E-03 | -2.49 | 0.01 | -1.39 | 0.17 | -0.81 | 0.42 | -1.80 | 0.07 | 0 | 0.85 |
| g20773800 | 7 | 99,934,006 | 7q22.1 | 5.17 | 2.28E-07 | 0.77 | *STAG3L5P* ; *STAG3L5P-PVRIG2P-PILRB* | Intronic | 2.95 | 3.21E-03 | 2.17 | 0.03 | 1.92 | 0.05 | 0.89 | 0.37 | 1.62 | 0.10 | 0 | 0.96 |
| g24531009 | 7 | 99,987,853 | 7q22.1 | 5.18 | 2.18E-07 | 0.42 | *PILRA* | Intronic | 3.01 | 2.61E-03 | 2.56 | 0.01 | 1.61 | 0.11 | 0.65 | 0.51 | 1.66 | 0.10 | 0 | 0.85 |
| g14587961 | 7 | 99,991,523 | 7q22.1 | 5.19 | 2.12E-07 | 0.72 | *PILRA* | Intronic | 3.01 | 2.57E-03 | 2.63 | 8.44E-03 | 1.44 | 0.15 | 0.62 | 0.53 | 1.66 | 0.10 | 0 | 0.81 |
| g13153964 | 7 | 100,026,424 | 7q22.1 | 5.13 | 2.83E-07 | 0.74 | *MEPCE* | 5'UTR | 2.92 | 3.47E-03 | 2.65 | 8.06E-03 | 1.69 | 0.09 | 0.45 | 0.66 | 1.87 | 0.06 | 0 | 0.77 |
| g23667466 | 7 | 100,026,434 | 7q22.1 | 5.20 | 1.97E-07 | 0.73 | *MEPCE* | 5'UTR | 2.97 | 3.03E-03 | 2.60 | 9.34E-03 | 1.74 | 0.08 | 0.54 | 0.59 | 1.83 | 0.07 | 0 | 0.82 |
| g21679810 | 7 | 100,027,352 | 7q22.1 | 5.16 | 2.48E-07 | 0.70 | *MEPCE* | Intronic | 2.95 | 3.16E-03 | 2.69 | 7.23E-03 | 1.61 | 0.11 | 0.49 | 0.62 | 1.82 | 0.07 | 0 | 0.77 |
| g07160278 | 7 | 100,027,354 | 7q22.1 | 5.15 | 2.63E-07 | 0.76 | *MEPCE* | Intronic | 2.93 | 3.36E-03 | 2.67 | 7.65E-03 | 1.67 | 0.10 | 0.44 | 0.66 | 1.87 | 0.06 | 0 | 0.77 |
| g20601288 | 7 | 100,031,831 | 7q22.1 | 5.13 | 2.88E-07 | 0.72 | *MEPCE* | Downstream | 2.91 | 3.66E-03 | 2.65 | 8.03E-03 | 1.74 | 0.08 | 0.47 | 0.64 | 1.84 | 0.07 | 0 | 0.78 |
| g15448098 | 7 | 100,083,084 | 7q22.1 | -5.20 | 1.98E-07 | 0.69 | *NYAP1* | Intronic | -2.96 | 3.06E-03 | -2.44 | 0.01 | -1.86 | 0.06 | -0.72 | 0.47 | -1.70 | 0.09 | 0 | 0.90 |
| cg0232753 | 7 | 100,091,786 | 7q22.1 | 5.13 | 2.94E-07 | 0.41 | *NYAP1* | 3'UTR | 3.21 | 1.32E-03 | 2.18 | 0.03 | 0.95 | 0.34 | 0.45 | 0.65 | 1.91 | 0.06 | 0 | 0.85 |
| cg0542330 | 7 | 102,500,111 | 7q22.1 | -5.14 | 2.80E-07 | 0.45 | *FBXL13* | Intronic | -4.32 | 1.53E-05 | -1.29 | 0.20 | -0.46 | 0.64 | -2.47 | 0.01 | -2.32 | 0.02 | 9 | 0.36 |
| cg0413240 | 7 | 130,626,376 | 7q32.3 | -5.79 | 6.98E-09 | 0.51 | *LINC-PINT* | Downstream | -4.99 | 5.97E-07 | -0.39 | 0.70 | -0.23 | 0.82 | -0.25 | 0.80 | -2.95 | 3.16E-03 | 51.9 | 0.08 |
| cg0754677 | 8 | 29,495,175 | 8p12 | -6.75 | 1.46E-11 | 0.52 | *DUSP4* ; *LINC00589* | Intergenic | -5.71 | **1.11E-08** | -1.75 | 0.08 | -2.04 | 0.04 | 0.57 | 0.57 | -3.36 | 7.76E-04 | 35.8 | 0.18 |
| cg2150481 | 8 | 37,285,286 | 8p11.23 | -5.15 | 2.55E-07 | 0.44 | *LINC01605* | Intronic | -2.84 | 4.49E-03 | -2.22 | 0.03 | 0.38 | 0.71 | -0.32 | 0.75 | -2.88 | 4.02E-03 | 47.4 | 0.11 |
| cg1732808 | 8 | 75,916,668 | 8q21.11 | 8.26 | 1.46E-16 | 0.36 | *CRISPLD1* | Intronic | 5.49 | **3.91E-08** | 3.45 | 5.61E-04 | 4.15 | 3.32E-05 | 1.84 | 0.07 | 2.27 | 0.02 | 0 | 0.63 |
| cg1548873 | 8 | 124,194,891 | 8q24.13 | -5.70 | 1.21E-08 | 0.46 | *FAM83A* | 5'UTR | -4.83 | 1.39E-06 | -1.35 | 0.18 | -0.67 | 0.50 | -0.20 | 0.84 | -4.06 | 4.86E-05 | 49.8 | 0.09 |
| cg0729768 | 8 | 124,254,762 | 8q24.13 | -6.33 | 2.48E-10 | 0.38 | *ZHX1-C8orf76* | Intronic | -5.52 | **3.31E-08** | -1.38 | 0.17 | -0.77 | 0.44 | 0.00 | 1.00 | -4.23 | 2.30E-05 | 47.4 | 0.11 |
| cg1573082 | 8 | 124,838,057 | 8q24.13 | -5.57 | 2.54E-08 | 0.40 | *FAM91A1* ; *FER1L6* | Intergenic | -5.01 | 5.52E-07 | -0.72 | 0.47 | -1.38 | 0.17 | -0.22 | 0.83 | -1.93 | 0.05 | 0 | 0.45 |
| cg2489515 | 8 | 127,837,199 | 8q24.21 | 5.56 | 2.68E-08 | 0.46 | *LRATD2* ; *PCAT1* | Intergenic | 4.51 | 6.34E-06 | 2.05 | 0.04 | 2.62 | 8.87E-03 | 1.85 | 0.06 | 0.82 | 0.41 | 0 | 0.66 |
| cg0192617 | 8 | 128,671,375 | 8q24.21 | 5.50 | 3.89E-08 | 0.53 | *CASC8* ; *CASC11* | Intergenic | 4.64 | 3.46E-06 | -0.07 | 0.95 | 2.90 | 3.77E-03 | 0.75 | 0.45 | 1.49 | 0.14 | 0 | 0.76 |
| cg1532684 | 8 | 128,850,899 | 8q24.21 | -11.27 | 1.85E-29 | 0.38 | *PVT1* | Intronic | -8.94 | **3.95E-19** | -3.44 | 5.84E-04 | -4.04 | 5.36E-05 | -2.62 | 8.80E-03 | -1.82 | 0.07 | 35.8 | 0.18 |
| cg0095631 | 8 | 128,944,009 | 8q24.21 | -5.76 | 8.48E-09 | 0.54 | *PVT1* | Intronic | -4.69 | 2.78E-06 | -1.92 | 0.05 | -0.71 | 0.48 | -0.32 | 0.75 | 0.66 | 0.51 | 56.8 | 0.06 |
| cg0500452 | 8 | 129,163,376 | 8q24.21 | -7.89 | 3.01E-15 | 0.42 | *MIR1208* | Downstream | -6.25 | **4.18E-10** | -2.29 | 0.02 | -2.42 | 0.02 | -0.08 | 0.94 | -1.02 | 0.31 | 24.6 | 0.26 |
| cg2757473 | 8 | 129,206,020 | 8q24.21 | 5.80 | 6.52E-09 | 0.43 | *MIR1208* ; *LINC00824* | Intergenic | 4.71 | 2.53E-06 | 1.95 | 0.05 | 0.72 | 0.47 | 0.35 | 0.72 | -0.69 | 0.49 | 57.5 | 0.05 |
| cg1598832 | 9 | 22,098,908 | 9p21.3 | -9.35 | 9.11E-21 | 0.38 | *CDKN2B-AS1* | Intronic | -5.67 | **1.39E-08** | -3.01 | 2.63E-03 | -3.21 | 1.31E-03 | -3.00 | 2.69E-03 | -2.01 | 0.04 | 0 | 0.58 |
| cg0734316 | 9 | 119,448,853 | 9q33.1 | -6.58 | 4.71E-11 | 0.39 | *ASTN2* | Intronic | -4.04 | 5.28E-05 | -1.22 | 0.22 | -1.91 | 0.06 | -0.36 | 0.72 | -0.56 | 0.57 | 0 | 0.55 |
| cg2283227 | 10 | 63,805,067 | 10q21.2 | 7.29 | 3.08E-13 | 0.68 | *ARID5B* | Intronic | 4.77 | 1.80E-06 | 3.22 | 1.30E-03 | 2.36 | 0.02 | 0.87 | 0.38 | 1.12 | 0.26 | 0 | 0.42 |
| cg1255299 | 10 | 64,374,314 | 10q21.2 | 5.71 | 1.12E-08 | 0.61 | *ZNF365* | Intronic | 3.42 | 6.27E-04 | 3.25 | 1.15E-03 | 2.46 | 0.01 | 1.10 | 0.27 | 2.33 | 0.02 | 0 | 0.67 |
| cg2756776 | 10 | 64,573,184 | 10q21.3 | 9.52 | 1.74E-21 | 0.41 | *EGR2* | Exonic | 7.36 | **1.84E-13** | 3.28 | 1.04E-03 | 2.56 | 0.01 | 2.00 | 0.05 | 0.01 | 0.99 | 0 | 0.75 |
| cg1443560 | 10 | 64,578,186 | 10q21.3 | 6.95 | 3.62E-12 | 0.54 | *EGR2* | Intronic | 5.24 | **1.60E-07** | 3.59 | 3.37E-04 | 2.72 | 6.51E-03 | 1.60 | 0.11 | 1.35 | 0.18 | 0 | 0.61 |
| cg2287288 | 10 | 80,615,772 | 10q22.3 | -6.92 | 4.61E-12 | 0.55 | *LINC00595* ; *ZMIZ1-AS* | Intergenic | -5.83 | **5.51E-09** | -0.53 | 0.60 | -2.72 | 6.48E-03 | -1.19 | 0.23 | -2.88 | 3.93E-03 | 0 | 0.47 |
| cg2533157 | 10 | 114,779,655 | 10q25.2 | -6.26 | 3.82E-10 | 0.44 | *TCF7L2* | Intronic | -4.26 | 2.01E-05 | -0.90 | 0.37 | -1.03 | 0.30 | 0.80 | 0.42 | -3.82 | 1.35E-04 | 56.4 | 0.06 |
| cg1434146 | 10 | 121,201,554 | 10q26.11 | 5.67 | 1.45E-08 | 0.55 | *GRK5* | Exonic | 3.65 | 2.57E-04 | 1.62 | 0.11 | 2.11 | 0.03 | 1.34 | 0.18 | 0.98 | 0.33 | 0 | 0.93 |
| cg1707586 | 10 | 121,287,467 | 10q26.11 | 5.75 | 9.07E-09 | 0.42 | *RGS10* | Intronic | 3.55 | 3.84E-04 | 1.61 | 0.11 | 1.10 | 0.27 | 1.67 | 0.09 | 0.92 | 0.36 | 0 | 0.82 |
| cg2083566 | 10 | 123,688,330 | 10q26.13 | -5.24 | 1.63E-07 | 0.41 | *ATE1-AS1* | Intronic | -5.18 | **2.19E-07** | -1.84 | 0.07 | -2.25 | 0.02 | 0.34 | 0.73 | -1.35 | 0.18 | 0 | 0.49 |
| cg0604649 | 11 | 320,940 | 11p15.5 | 5.97 | 2.44E-09 | 0.42 | *IFITM3* | TSS200 | 2.43 | 0.01 | 2.02 | 0.04 | 1.52 | 0.13 | 2.75 | 6.02E-03 | 2.90 | 3.68E-03 | 26 | 0.25 |
| cg0132241 | 11 | 781,977 | 11p15.5 | 6.42 | 1.40E-10 | 0.45 | *LOC171391* | Intronic | 4.93 | 8.13E-07 | 1.83 | 0.07 | 1.72 | 0.09 | 1.39 | 0.17 | 1.61 | 0.11 | 0 | 0.89 |
| cg0174137 | 11 | 783,889 | 11p15.5 | 6.61 | 3.91E-11 | 0.52 | *LOC171391* | Exonic | 4.97 | 6.56E-07 | 1.90 | 0.06 | 1.77 | 0.08 | 1.49 | 0.14 | 1.69 | 0.09 | 0 | 0.90 |
| cg2162096 | 11 | 802,895 | 11p15.5 | 6.70 | 2.05E-11 | 0.47 | *PIDD1* | Intronic | 4.81 | 1.50E-06 | 2.40 | 0.02 | 1.54 | 0.12 | 2.01 | 0.04 | 1.77 | 0.08 | 0 | 0.80 |
| cg1051479 | 11 | 802,938 | 11p15.5 | 6.02 | 1.70E-09 | 0.40 | *PIDD1* | Intronic | 4.24 | 2.23E-05 | 1.94 | 0.05 | 1.51 | 0.13 | 2.04 | 0.04 | 1.56 | 0.12 | 0 | 0.86 |
| cg1762889 | 11 | 804,452 | 11p15.5 | -7.53 | 5.26E-14 | 0.42 | *PIDD1* | 5'UTR | -4.82 | 1.40E-06 | -2.78 | 5.46E-03 | -1.50 | 0.13 | -2.34 | 0.02 | -2.34 | 0.02 | 0 | 0.69 |
| cg0552240 | 11 | 1,502,226 | 11p15.5 | -10.49 | 9.67E-26 | 0.39 | *MOB2* | Intronic | -8.21 | **2.12E-16** | -4.41 | 1.05E-05 | -3.21 | 1.35E-03 | -1.72 | 0.09 | -2.98 | 2.87E-03 | 0 | 0.53 |
| cg1305821 | 11 | 1,947,949 | 11p15.5 | 6.06 | 1.33E-09 | 0.49 | *TNNT3* | Intronic | 4.43 | 9.45E-06 | 2.53 | 0.01 | 1.71 | 0.09 | 0.59 | 0.55 | 0.95 | 0.34 | 0 | 0.65 |
| cg0558660 | 11 | 1,965,447 | 11p15.5 | -7.27 | 3.72E-13 | 0.36 | *TNNT3* ; *MRPL23* | Intergenic | -5.49 | **3.94E-08** | -3.53 | 4.10E-04 | -2.73 | 6.32E-03 | -0.78 | 0.43 | -1.71 | 0.09 | 0 | 0.59 |
| cg0649896 | 11 | 1,981,264 | 11p15.5 | -6.99 | 2.72E-12 | 0.58 | *MRPL23* ; *MRPL23-AS1* | Intergenic | -5.49 | **4.04E-08** | -3.52 | 4.26E-04 | -2.43 | 0.02 | -0.96 | 0.34 | -1.50 | 0.13 | 0 | 0.49 |
| cg0589471 | 11 | 1,989,899 | 11p15.5 | 7.38 | 1.56E-13 | 0.40 | *MRPL23* ; *MRPL23-AS1* | Intergenic | 5.97 | **2.32E-09** | 2.65 | 8.03E-03 | 2.77 | 5.60E-03 | 0.86 | 0.39 | 1.97 | 0.05 | 0 | 0.75 |
| cg2465985 | 11 | 65,222,491 | 11q13.1 | 6.08 | 1.22E-09 | 0.39 | *LOC101927789* | Upstream | 4.82 | 1.41E-06 | 2.34 | 0.02 | 2.56 | 0.01 | 1.73 | 0.08 | 0.22 | 0.83 | 12.2 | 0.34 |
| cg0653892 | 11 | 65,579,765 | 11q13.1 | 5.74 | 9.65E-09 | 0.67 | *OVOL1* ; *SNX32* | Intergenic | 5.27 | **1.37E-07** | 1.82 | 0.07 | 3.02 | 2.52E-03 | 0.76 | 0.45 | 0.15 | 0.88 | 35.5 | 0.18 |
| cg0405510 | 11 | 65,626,734 | 11q13.1 | 6.74 | 1.59E-11 | 0.48 | *CFL1* ; *MUS81* | Intergenic | 5.98 | **2.22E-09** | 2.31 | 0.02 | 1.59 | 0.11 | 2.89 | 3.84E-03 | 0.61 | 0.54 | 48.6 | 0.10 |
| cg0057633 | 11 | 65,640,516 | 11q13.1 | -5.13 | 2.90E-07 | 0.48 | *EFEMP2* | TSS200 | -4.56 | 5.03E-06 | -1.84 | 0.07 | -2.77 | 5.63E-03 | -0.77 | 0.44 | -0.03 | 0.98 | 30 | 0.22 |
| cg1979280 | 11 | 65,647,270 | 11q13.1 | 5.64 | 1.66E-08 | 0.47 | *CTSW* | TSS200 | 5.07 | **3.97E-07** | 1.69 | 0.09 | 2.23 | 0.03 | 1.86 | 0.06 | 0.68 | 0.50 | 0 | 0.51 |
| cg2234552 | 11 | 65,667,998 | 11q13.1 | 5.75 | 8.69E-09 | 0.40 | *FOSL1* | TSS200 | 4.98 | 6.48E-07 | 2.16 | 0.03 | 2.65 | 8.09E-03 | 1.30 | 0.19 | 0.23 | 0.82 | 13.6 | 0.33 |
| cg0222732 | 11 | 66,008,686 | 11q13.2 | 5.51 | 3.55E-08 | 0.37 | *PACS1* | Intronic | 4.49 | 7.01E-06 | 2.23 | 0.03 | 2.66 | 7.88E-03 | 1.04 | 0.30 | 0.03 | 0.97 | 0 | 0.67 |
| cg1089297 | 11 | 66,028,483 | 11q13.2 | 5.61 | 2.05E-08 | 0.47 | *KLC2* | Intronic | 4.57 | 4.85E-06 | 0.73 | 0.47 | 2.12 | 0.03 | 2.24 | 0.03 | 0.49 | 0.62 | 15 | 0.32 |
| cg0919004 | 11 | 66,118,069 | 11q13.2 | 5.22 | 1.76E-07 | 0.37 | *LOC102724064* | Intronic | 4.50 | 6.93E-06 | 2.67 | 7.49E-03 | 2.60 | 9.23E-03 | 1.05 | 0.29 | 1.64 | 0.10 | 0 | 0.90 |
| cg0966396 | 11 | 68,939,830 | 11q13.3 | -5.72 | 1.08E-08 | 0.43 | *LOC338694* | TSS1500 | -4.54 | 5.75E-06 | -0.69 | 0.49 | -1.46 | 0.14 | -1.92 | 0.06 | -1.19 | 0.23 | 0 | 0.85 |
| cg1505949 | 11 | 68,980,857 | 11q13.3 | 5.48 | 4.26E-08 | 0.47 | *LOC338694* ; *MYEOV* | Intergenic | 4.67 | 3.04E-06 | 0.67 | 0.50 | 1.27 | 0.20 | 2.46 | 0.01 | 0.35 | 0.73 | 38.2 | 0.17 |
| cg0710441 | 11 | 69,259,247 | 11q13.3 | -5.68 | 1.32E-08 | 0.55 | *LOC102724265* ; *LINC01488* | Intergenic | -4.38 | 1.17E-05 | -1.27 | 0.21 | -2.96 | 3.10E-03 | 1.25 | 0.21 | -1.13 | 0.26 | 50.3 | 0.09 |
| cg0186509 | 11 | 129,763,954 | 11q24.3 | -6.79 | 1.09E-11 | 0.51 | *NFRKB* | Intronic | -4.35 | 1.35E-05 | -2.31 | 0.02 | -1.75 | 0.08 | -2.74 | 6.12E-03 | -2.99 | 2.82E-03 | 0 | 0.58 |

| | | | | | | | | | | | | | | | | | | | | |
|---|---|---|---|---|---|---|---|---|---|---|---|---|---|---|---|---|---|---|---|---|
| cg1336853 | 12 | 14,414,242 | 12p13.1 | -7.87 | 3.49E-15 | 0.55 | *GRIN2B* ; *ATF7IP* | Intergenic | -5.53 | **3.20E-08** | -2.29 | 0.02 | -2.21 | 0.03 | -2.42 | 0.02 | -2.24 | 0.02 | 0 | 0.87 |
| cg1421098 | 12 | 28,115,804 | 12p11.22 | -5.67 | 1.45E-08 | 0.45 | *PTHLH* | 3'UTR | -4.40 | 1.06E-05 | -1.91 | 0.06 | -1.04 | 0.30 | -0.15 | 0.88 | -4.73 | 2.20E-06 | 56.8 | 0.05 |
| cg1544639 | 12 | 28,501,308 | 12p11.22 | 5.13 | 2.94E-07 | 0.40 | *CCDC91* | Intronic | 3.80 | 1.46E-04 | 2.05 | 0.04 | 1.00 | 0.32 | 0.50 | 0.61 | 3.22 | 1.29E-03 | 0 | 0.54 |
| cg0157945 | 12 | 28,651,427 | 12p11.22 | 7.71 | 1.26E-14 | 0.44 | *CCDC91* | Intronic | 6.72 | **1.78E-11** | 2.31 | 0.02 | 2.53 | 0.01 | 1.37 | 0.17 | 4.21 | 2.56E-05 | 0 | 0.69 |
| cg2548664 | 12 | 57,472,858 | 12q13.3 | -5.20 | 1.99E-07 | 0.50 | *NEMP1* | TSS1500 | -4.11 | 3.88E-05 | -0.14 | 0.89 | -1.50 | 0.13 | -0.35 | 0.73 | -3.75 | 1.77E-04 | 29.5 | 0.23 |
| cg1356003 | 12 | 96,099,778 | 12q22 | -13.14 | 1.91E-39 | 0.39 | *NTN4* | Intronic | -10.49 | **9.33E-26** | -4.14 | 3.47E-05 | -4.10 | 4.20E-05 | -1.06 | 0.29 | -4.75 | 2.08E-06 | 22.2 | 0.27 |
| cg2574182 | 12 | 96,180,994 | 12q22 | 14.41 | 4.46E-47 | 0.68 | *NTN4* | Exonic | 11.43 | **2.93E-30** | 4.97 | 6.86E-07 | 4.30 | 1.75E-05 | 0.95 | 0.34 | 5.60 | **2.18E-08** | 49 | 0.10 |
| cg1928483 | 12 | 96,186,464 | 12q22 | 14.27 | 3.20E-46 | 0.54 | *NTN4* ; *LOC105369921* | Intergenic | 11.47 | **1.92E-30** | 4.91 | 9.03E-07 | 4.18 | 2.94E-05 | 0.89 | 0.37 | 5.25 | **1.55E-07** | 51 | 0.09 |
| cg1756448 | 12 | 111,396,889 | 12q24.11 | -5.49 | 4.13E-08 | 0.47 | *LOC105369980* ; *CUX2* | Intergenic | -5.05 | **4.44E-07** | -0.80 | 0.42 | -2.15 | 0.03 | -1.96 | 0.05 | -2.02 | 0.04 | 0 | 0.68 |
| cg0793219 | 12 | 112,008,034 | 12q24.12 | -5.49 | 3.93E-08 | 0.55 | *ATXN2* | Intronic | -4.44 | 9.00E-06 | -1.13 | 0.26 | -1.85 | 0.06 | -1.78 | 0.07 | -1.69 | 0.09 | 0 | 0.88 |
| cg2274428 | 12 | 112,200,054 | 12q24.12 | -5.23 | 1.66E-07 | 0.44 | *ACAD10* ; *ALDH2* | Intergenic | -4.62 | 3.76E-06 | -1.08 | 0.28 | -2.32 | 0.02 | -1.40 | 0.16 | -1.25 | 0.21 | 0 | 0.80 |
| cg2662053 | 12 | 115,102,726 | 12q24.21 | 6.55 | 5.75E-11 | 0.44 | *TBX5-AS1* ; *TBX3* | Intergenic | 5.31 | **1.09E-07** | 2.35 | 0.02 | 3.57 | 3.52E-04 | 0.21 | 0.84 | 0.44 | 0.66 | 45.3 | 0.12 |
| cg2664891 | 14 | 37,666,927 | 14q13.3 | -7.18 | 6.91E-13 | 0.37 | *MIPOL1* | TSS200 | -6.37 | **1.92E-10** | -1.98 | 0.05 | -2.08 | 0.04 | 0.11 | 0.91 | -1.47 | 0.14 | 36.9 | 0.18 |
| cg1775402 | 14 | 68,410,569 | 14q24.1 | 5.50 | 3.72E-08 | 0.40 | *RAD51B* | Intronic | 3.77 | 1.65E-04 | 3.00 | 2.72E-03 | 1.22 | 0.22 | 1.57 | 0.12 | 1.75 | 0.08 | 0 | 0.66 |
| cg0841518 | 14 | 68,578,658 | 14q24.1 | -5.20 | 1.96E-07 | 0.43 | *RAD51B* | Intronic | -4.31 | 1.63E-05 | -2.55 | 0.01 | -0.88 | 0.38 | -1.66 | 0.10 | -0.86 | 0.39 | 0 | 0.44 |
| cg0238769 | 14 | 68,887,349 | 14q24.1 | 7.04 | 1.87E-12 | 0.60 | *RAD51B* | Intronic | 5.27 | **1.38E-07** | 3.16 | 1.56E-03 | 2.14 | 0.03 | 1.68 | 0.09 | 1.82 | 0.07 | 0 | 0.77 |
| cg1076621 | 14 | 68,915,105 | 14q24.1 | 7.32 | 2.45E-13 | 0.64 | *RAD51B* | Intronic | 5.45 | **5.00E-08** | 3.20 | 1.39E-03 | 2.33 | 0.02 | 1.67 | 0.10 | 2.15 | 0.03 | 0 | 0.86 |
| cg1690355 | 14 | 68,993,143 | 14q24.1 | 7.41 | 1.25E-13 | 0.39 | *RAD51B* | Intronic | 5.53 | **3.23E-08** | 3.31 | 9.20E-04 | 1.73 | 0.08 | 2.36 | 0.02 | 1.60 | 0.11 | 0 | 0.41 |
| cg2093113 | 14 | 69,045,498 | 14q24.1 | -6.49 | 8.68E-11 | 0.37 | *RAD51B* | Intronic | -6.34 | **2.27E-10** | -1.25 | 0.21 | -1.03 | 0.30 | 0.08 | 0.94 | -1.92 | 0.06 | 53 | 0.07 |
| cg1322556 | 14 | 93,106,414 | 14q32.12 | 6.49 | 8.81E-11 | 0.47 | *RIN3* | Intronic | 3.73 | 1.93E-04 | 1.84 | 0.07 | 2.75 | 5.95E-03 | 2.07 | 0.04 | 2.32 | 0.02 | 0 | 0.89 |
| cg0194254 | 16 | 52,504,616 | 16q12.1 | 7.15 | 8.90E-13 | 0.47 | *TOX3* | Intronic | 4.46 | 8.32E-06 | 1.08 | 0.28 | 1.75 | 0.08 | 0.64 | 0.52 | 1.36 | 0.17 | 0 | 0.67 |
| cg0307326 | 16 | 54,008,588 | 16q12.2 | -7.19 | 6.51E-13 | 0.39 | *FTO* | Intronic | -3.87 | 1.10E-04 | -2.95 | 3.20E-03 | -2.83 | 4.59E-03 | -2.51 | 0.01 | -4.36 | 1.29E-05 | 33.4 | 0.20 |
| cg1581711 | 16 | 54,097,364 | 16q12.2 | 5.45 | 5.07E-08 | 0.44 | *FTO* | Intronic | 2.92 | 3.48E-03 | 2.62 | 8.81E-03 | 1.77 | 0.08 | 2.40 | 0.02 | 3.81 | 1.40E-04 | 45.7 | 0.12 |
| cg1482249 | 16 | 80,662,527 | 16q23.2 | 8.69 | 3.75E-18 | 0.48 | *CDYL2* | Intronic | 7.79 | **6.63E-15** | 3.22 | 1.27E-03 | 4.53 | 5.86E-06 | 0.60 | 0.55 | 2.45 | 0.01 | 27.6 | 0.24 |
| cg0705664 | 16 | 86,546,785 | 16q24.1 | 5.14 | 2.76E-07 | 0.49 | *FOXF1* | 3'UTR | 3.32 | 8.94E-04 | 2.86 | 4.18E-03 | 1.69 | 0.09 | 1.47 | 0.14 | 1.27 | 0.21 | 0 | 0.71 |
| cg0544196 | 16 | 87,505,566 | 16q24.2 | -5.46 | 4.80E-08 | 0.38 | *ZCCHC14* | Intronic | -4.28 | 1.86E-05 | -3.23 | 1.23E-03 | -1.11 | 0.27 | -1.06 | 0.29 | -2.25 | 0.02 | 0 | 0.48 |
| cg1486682 | 17 | 40,853,095 | 17q21.2 | -5.28 | 1.32E-07 | 0.41 | *EZH1* | 3'UTR | -3.24 | 1.18E-03 | -0.27 | 0.78 | -0.09 | 0.93 | -3.44 | 5.85E-04 | -2.54 | 0.01 | 56.4 | 0.06 |
| cg2447740 | 17 | 43,484,506 | 17q21.31 | 6.28 | 3.35E-10 | 0.58 | *ARHGAP27* | Intronic | 4.29 | 1.79E-05 | 2.55 | 0.01 | 0.74 | 0.46 | 2.42 | 0.02 | 0.00 | 1.00 | 27 | 0.25 |
| cg0220072 | 17 | 43,489,199 | 17q21.31 | 6.24 | 4.36E-10 | 0.53 | *ARHGAP27* | Intronic | 3.88 | 1.04E-04 | 2.21 | 0.03 | 0.54 | 0.59 | 1.58 | 0.11 | 0.65 | 0.52 | 0 | 0.41 |
| cg0706757 | 17 | 43,506,829 | 17q21.31 | 6.25 | 4.18E-10 | 0.38 | *ARHGAP27* | 3'UTR | 4.27 | 1.93E-05 | 2.13 | 0.03 | 0.33 | 0.74 | 2.14 | 0.03 | -0.23 | 0.82 | 57.1 | 0.05 |
| cg0171896 | 17 | 43,516,991 | 17q21.31 | 6.15 | 7.58E-10 | 0.63 | *PLEKHM1* | Exonic | 4.25 | 2.16E-05 | 2.26 | 0.02 | 1.17 | 0.24 | 2.09 | 0.04 | -0.02 | 0.99 | 0 | 0.57 |
| cg0091697 | 17 | 43,580,461 | 17q21.31 | 6.61 | 3.77E-11 | 0.51 | *ARL17B* | Upstream | 4.27 | 1.99E-05 | 2.65 | 7.98E-03 | 0.24 | 0.81 | 2.07 | 0.04 | 0.59 | 0.56 | 42.1 | 0.14 |
| cg2243321 | 17 | 43,662,623 | 17q21.31 | -6.96 | 3.44E-12 | 0.60 | *ARL17A* ; *MAPK8IP1P2* | Intergenic | -4.29 | 1.79E-05 | -3.03 | 2.41E-03 | -0.28 | 0.78 | -2.06 | 0.04 | -0.81 | 0.42 | 44.7 | 0.12 |
| cg0134121 | 17 | 43,662,625 | 17q21.31 | -6.99 | 2.79E-12 | 0.64 | *ARL17A* ; *MAPK8IP1P2* | Intergenic | -4.31 | 1.61E-05 | -3.07 | 2.13E-03 | -0.28 | 0.78 | -2.04 | 0.04 | -0.83 | 0.41 | 45.2 | 0.12 |
| cg0188239 | 17 | 43,717,810 | 17q21.31 | -6.87 | 6.50E-12 | 0.46 | *LINC02210* | Exonic | -4.11 | 3.95E-05 | -3.12 | 1.79E-03 | -0.19 | 0.85 | -2.14 | 0.03 | -0.80 | 0.42 | 48.1 | 0.10 |
| cg2039330 | 17 | 43,728,044 | 17q21.31 | -5.80 | 6.50E-09 | 0.72 | *LINC02210-CRHR1* | Intronic | -3.87 | 1.10E-04 | -2.85 | 4.38E-03 | -0.37 | 0.71 | -1.95 | 0.05 | 0.14 | 0.89 | 39.3 | 0.16 |
| cg0653739 | 17 | 43,799,898 | 17q21.31 | -6.65 | 2.84E-11 | 0.53 | *LINC02210-CRHR1* | Intronic | -4.15 | 3.34E-05 | -2.76 | 5.75E-03 | -0.48 | 0.63 | -2.09 | 0.04 | -0.26 | 0.80 | 40.7 | 0.15 |
| cg0323827 | 17 | 43,825,672 | 17q21.31 | -6.89 | 5.58E-12 | 0.65 | *LINC02210-CRHR1* | Intronic | -4.43 | 9.22E-06 | -2.77 | 5.59E-03 | -0.37 | 0.71 | -2.26 | 0.02 | -0.21 | 0.83 | 50.5 | 0.09 |
| cg1622835 | 17 | 43,848,958 | 17q21.31 | 6.91 | 4.85E-12 | 0.52 | *LINC02210-CRHR1* | Intronic | 4.23 | 2.33E-05 | 2.86 | 4.29E-03 | 0.23 | 0.82 | 2.00 | 0.05 | 0.70 | 0.48 | 42.2 | 0.14 |
| cg1429779 | 17 | 43,867,801 | 17q21.31 | -6.25 | 4.13E-10 | 0.42 | *CRHR1* ; *LINC02210-CRHR1* | Intronic | -4.21 | 2.53E-05 | -3.12 | 1.81E-03 | -0.64 | 0.52 | -1.55 | 0.12 | -0.81 | 0.42 | 24.5 | 0.26 |
| cg1584480 | 17 | 43,916,233 | 17q21.31 | 6.50 | 7.91E-11 | 0.49 | *LINC02210-CRHR1* ; *MAPT-AS1* | Intergenic | 4.07 | 4.65E-05 | 2.60 | 9.27E-03 | 1.07 | 0.29 | 2.40 | 0.02 | 1.10 | 0.27 | 2.8 | 0.39 |
| cg1583477 | 17 | 43,919,576 | 17q21.31 | 5.65 | 1.57E-08 | 0.39 | *LINC02210-CRHR1* ; *MAPT-AS1* | Intergenic | 3.33 | 8.70E-04 | 2.62 | 8.78E-03 | 0.16 | 0.88 | 1.74 | 0.08 | 1.13 | 0.26 | 3.9 | 0.38 |
| cg2320227 | 17 | 43,971,911 | 17q21.31 | -6.94 | 4.05E-12 | 0.62 | *MAPT-AS1* | Intronic | -4.36 | 1.32E-05 | -2.88 | 3.99E-03 | -0.29 | 0.77 | -2.11 | 0.03 | -0.76 | 0.44 | 43.7 | 0.13 |
| cg2009941 | 17 | 43,971,919 | 17q21.31 | -6.94 | 3.88E-12 | 0.44 | *MAPT-AS1* | Intronic | -4.31 | 1.67E-05 | -3.02 | 2.56E-03 | -0.28 | 0.78 | -2.08 | 0.04 | -0.83 | 0.40 | 44.2 | 0.13 |
| cg1148926 | 17 | 43,973,426 | 17q21.31 | -6.98 | 3.02E-12 | 0.46 | *MAPT-IT1* | Exonic | -4.35 | 1.35E-05 | -3.03 | 2.48E-03 | -0.31 | 0.76 | -2.15 | 0.03 | -0.71 | 0.48 | 47 | 0.11 |
| cg1022460 | 17 | 43,975,063 | 17q21.31 | -6.33 | 2.38E-10 | 0.46 | *MAPT-IT1* | Exonic | -4.20 | 2.62E-05 | -2.40 | 0.02 | -0.10 | 0.92 | -1.88 | 0.06 | -0.42 | 0.67 | 36.6 | 0.18 |
| cg2159949 | 17 | 43,998,761 | 17q21.31 | -5.71 | 1.13E-08 | 0.39 | *MAPT* | Intronic | -4.08 | 4.46E-05 | -2.67 | 7.60E-03 | -1.55 | 0.12 | -1.71 | 0.09 | -1.30 | 0.19 | 0 | 0.74 |
| cg2392623 | 17 | 44,024,256 | 17q21.31 | 6.61 | 3.72E-11 | 0.47 | *MAPT* | Intronic | 4.22 | 2.40E-05 | 2.81 | 4.97E-03 | 0.25 | 0.80 | 2.24 | 0.03 | 0.33 | 0.74 | 50.4 | 0.09 |
| cg1858691 | 17 | 44,040,821 | 17q21.31 | 6.83 | 8.77E-12 | 0.59 | *MAPT* | Intronic | 4.29 | 1.80E-05 | 2.96 | 3.11E-03 | 0.25 | 0.80 | 2.10 | 0.04 | 0.68 | 0.49 | 46.4 | 0.11 |
| cg0193406 | 17 | 44,064,242 | 17q21.31 | -6.30 | 2.99E-10 | 0.42 | *MAPT* | Intronic | -4.19 | 2.79E-05 | -2.53 | 0.01 | -0.25 | 0.80 | -2.09 | 0.04 | -0.12 | 0.91 | 43.4 | 0.13 |
| cg0572148 | 17 | 44,071,124 | 17q21.31 | -6.97 | 3.25E-12 | 0.48 | *MAPT* | Intronic | -4.43 | 9.31E-06 | -2.86 | 4.19E-03 | -0.66 | 0.51 | -1.95 | 0.05 | -0.57 | 0.57 | 35.8 | 0.18 |
| cg1272797 | 17 | 44,075,500 | 17q21.31 | 6.00 | 1.95E-09 | 0.38 | *MAPT* | Intronic | 3.41 | 6.48E-04 | 2.50 | 0.01 | 0.34 | 0.73 | 1.03 | 0.30 | 1.08 | 0.28 | 0 | 0.64 |
| cg1568285 | 17 | 44,076,429 | 17q21.31 | 5.47 | 4.40E-08 | 0.47 | *MAPT* | Intronic | 2.61 | 9.08E-03 | 3.35 | 8.13E-04 | -0.14 | 0.89 | 1.86 | 0.06 | 0.03 | 0.98 | 55.7 | 0.06 |
| cg0831866 | 17 | 44,122,580 | 17q21.31 | -6.53 | 6.37E-11 | 0.56 | *KANSL1* | Intronic | -4.24 | 2.28E-05 | -2.66 | 7.77E-03 | -0.40 | 0.69 | -2.02 | 0.04 | -0.65 | 0.52 | 34 | 0.19 |
| cg0247899 | 17 | 44,180,961 | 17q21.31 | 6.56 | 5.22E-11 | 0.50 | *KANSL1* | Intronic | 3.99 | 6.48E-05 | 2.86 | 4.23E-03 | 0.10 | 0.92 | 2.38 | 0.02 | 0.28 | 0.78 | 53.1 | 0.07 |
| cg0190611 | 17 | 44,208,035 | 17q21.31 | 6.57 | 5.10E-11 | 0.61 | *KANSL1* | Intronic | 3.77 | 1.60E-04 | 2.93 | 3.36E-03 | 0.01 | 0.99 | 2.30 | 0.02 | 0.60 | 0.55 | 56.7 | 0.06 |
| cg1906479 | 17 | 44,257,101 | 17q21.31 | 6.31 | 2.80E-10 | 0.52 | *KANSL1* | Intronic | 4.31 | 1.63E-05 | 2.24 | 0.02 | 0.41 | 0.68 | 2.03 | 0.04 | 0.57 | 0.57 | 30.6 | 0.22 |
| cg0232203 | 17 | 44,269,535 | 17q21.31 | 6.43 | 1.27E-10 | 0.36 | *KANSL1* | Intronic | 4.39 | 1.16E-05 | 3.29 | 1.00E-03 | 1.02 | 0.31 | 2.20 | 0.03 | 0.89 | 0.37 | 32.6 | 0.20 |
| cg1613203 | 17 | 44,292,685 | 17q21.31 | 6.75 | 1.49E-11 | 0.47 | *KANSL1* | Intronic | 4.35 | 1.36E-05 | 2.93 | 3.36E-03 | 0.40 | 0.69 | 1.95 | 0.05 | 0.76 | 0.45 | 39.3 | 0.16 |
| cg1791178 | 17 | 44,343,683 | 17q21.31 | 6.08 | 1.23E-09 | 0.43 | *KANSL1* ; *ARL17B* | Intergenic | 3.96 | 7.58E-05 | 2.28 | 0.02 | 0.99 | 0.32 | 2.59 | 9.56E-03 | 0.58 | 0.56 | 28 | 0.23 |
| cg0515980 | 17 | 44,343,776 | 17q21.31 | 6.88 | 5.98E-12 | 0.64 | *KANSL1* ; *ARL17B* | Intergenic | 4.31 | 1.66E-05 | 3.03 | 2.42E-03 | 0.37 | 0.71 | 2.01 | 0.04 | 0.72 | 0.47 | 43.8 | 0.13 |
| cg0498637 | 17 | 77,768,687 | 17q25.3 | -6.30 | 2.89E-10 | 0.48 | *CBX8* | Exonic | -3.74 | 1.86E-04 | -3.74 | 1.86E-04 | -1.44 | 0.15 | -2.02 | 0.04 | -3.63 | 2.79E-04 | 37.3 | 0.17 |
| cg1087541 | 18 | 25,707,409 | 18q12.1 | 5.48 | 4.19E-08 | 0.52 | *CDH2* | Intronic | 2.50 | 0.01 | 2.38 | 0.02 | 1.44 | 0.15 | 2.28 | 0.02 | 3.63 | 2.78E-04 | 31.4 | 0.21 |
| cg1252744 | 19 | 13,908,134 | 19p13.13 | 5.58 | 2.36E-08 | 0.42 | *ZSWIM4* | Intronic | 2.97 | 3.01E-03 | 0.98 | 0.32 | 2.38 | 0.02 | 1.35 | 0.18 | -0.07 | 0.94 | 0 | 0.80 |
| g02633117 | 19 | 17,187,589 | 19p13.11 | -5.04 | 4.64E-07 | 0.44 | *MYO9B* | Intronic | -2.77 | 5.68E-03 | -1.49 | 0.14 | -2.20 | 0.03 | -2.16 | 0.03 | -0.71 | 0.47 | 0 | 0.62 |
| g11385249 | 19 | 17,215,602 | 19p13.11 | -5.06 | 4.11E-07 | 0.40 | *MYO9B* | Intronic | -2.57 | 0.01 | -2.96 | 3.09E-03 | -2.20 | 0.03 | -3.22 | 1.27E-03 | -0.69 | 0.49 | 55 | 0.06 |
| cg0952966 | 19 | 18,531,111 | 19p13.11 | -9.43 | 4.01E-21 | 0.47 | *SSBP4* | Intronic | -8.26 | **1.42E-16** | -3.08 | 2.08E-03 | -1.62 | 0.11 | -1.69 | 0.09 | -2.57 | 0.01 | 44.2 | 0.13 |
| cg1836244 | 19 | 18,544,419 | 19p13.11 | -6.24 | 4.50E-10 | 0.40 | *SSBP4* | Exonic | -4.61 | 4.06E-06 | -0.78 | 0.43 | -0.96 | 0.34 | -1.89 | 0.06 | -0.87 | 0.38 | 6.6 | 0.37 |
| cg0106597 | 19 | 18,549,689 | 19p13.11 | 7.25 | 4.02E-13 | 0.60 | *ISYNA1* | TSS1500 | 7.45 | **9.62E-14** | 2.23 | 0.03 | 1.69 | 0.09 | 0.93 | 0.35 | 2.20 | 0.03 | 34.6 | 0.19 |
| cg1079298 | 19 | 18,575,406 | 19p13.11 | -7.17 | 7.45E-13 | 0.54 | *ELL* | Intronic | -7.44 | **1.01E-13** | -1.71 | 0.09 | -1.16 | 0.24 | -1.43 | 0.15 | -1.85 | 0.06 | 53.8 | 0.07 |
| cg0403941 | 19 | 18,717,779 | 19p13.11 | -7.57 | 3.85E-14 | 0.37 | *CRLF1* | TSS200 | -6.52 | **6.89E-11** | -1.58 | 0.12 | -1.37 | 0.17 | -1.33 | 0.18 | -2.71 | 6.76E-03 | 6.5 | 0.37 |
| cg0238927 | 19 | 18,717,781 | 19p13.11 | -7.36 | 1.90E-13 | 0.37 | *CRLF1* | TSS200 | -6.76 | **1.43E-11** | -1.68 | 0.09 | -1.56 | 0.12 | -1.49 | 0.14 | -2.18 | 0.03 | 10.6 | 0.35 |
| cg2601685 | 19 | 19,028,003 | 19p13.11 | -5.71 | 1.14E-08 | 0.36 | *COPE* | Intronic | -4.94 | 7.85E-07 | -2.07 | 0.04 | -0.60 | 0.55 | 0.47 | 0.64 | -2.61 | 8.98E-03 | 31.8 | 0.21 |
| cg0288745 | 19 | 19,495,540 | 19p13.11 | -5.07 | 4.05E-07 | 0.42 | *MAU2* ; *GATAD2A* | Intergenic | -2.17 | 0.03 | -1.21 | 0.23 | -3.45 | 5.61E-04 | -0.28 | 0.78 | -1.51 | 0.13 | 11.8 | 0.34 |
| cg0370901 | 19 | 19,516,395 | 19p13.11 | 5.03 | 4.91E-07 | 0.57 | *GATAD2A* | Intronic | 3.81 | 1.41E-04 | 1.04 | 0.30 | 2.26 | 0.02 | 2.27 | 0.02 | 1.80 | 0.07 | 0 | 0.80 |
| cg0385407 | 19 | 19,626,428 | 19p13.11 | -5.82 | 5.97E-09 | 0.40 | *NDUFA13* ; *TSSK6* | 5'UTR | -2.75 | 6.04E-03 | -1.66 | 0.10 | -3.27 | 1.09E-03 | -0.76 | 0.44 | -1.98 | 0.05 | 0 | 0.45 |
| cg2673261 | 19 | 19,648,335 | 19p13.11 | 6.23 | 4.72E-10 | 0.39 | *YJEFN3* | 3'UTR | 3.44 | 5.78E-04 | 1.54 | 0.12 | 3.41 | 6.59E-04 | 1.45 | 0.15 | 1.82 | 0.07 | 0 | 0.71 |
| cg2637317 | 19 | 44,306,615 | 19q13.31 | -7.00 | 2.52E-12 | 0.47 | *LYPD5* | Intronic | -5.22 | **1.76E-07** | -2.14 | 0.03 | -2.47 | 0.01 | -1.98 | 0.05 | -4.34 | 1.44E-05 | 0 | 0.62 |
| cg1199392 | 19 | 44,307,056 | 19q13.31 | -6.29 | 3.24E-10 | 0.47 | *LYPD5* | Intronic | -4.77 | 1.89E-06 | -1.69 | 0.09 | -2.20 | 0.03 | -1.66 | 0.10 | -4.04 | 5.44E-05 | 0 | 0.62 |
| cg0130257 | 19 | 44,325,595 | 19q13.31 | 6.80 | 1.05E-11 | 0.52 | *LYPD5* | TSS1500 | 3.83 | 1.30E-04 | 3.59 | 3.31E-04 | 1.44 | 0.15 | 2.70 | 6.98E-03 | 3.04 | 2.38E-03 | 38.2 | 0.17 |
| cg0863329 | 19 | 44,405,433 | 19q13.31 | 6.88 | 6.10E-12 | 0.43 | *LOC100505715* | Exonic | 5.25 | **1.51E-07** | 3.08 | 2.04E-03 | 2.89 | 3.83E-03 | 3.25 | 1.16E-03 | 3.95 | 7.95E-05 | 0 | 0.51 |
| cg0551185 | 19 | 44,719,305 | 19q13.31 | -6.10 | 1.09E-09 | 0.51 | *ZNF227* | Intronic | -5.19 | **2.12E-07** | -1.61 | 0.11 | -1.61 | 0.11 | -0.81 | 0.42 | -2.03 | 0.04 | 0 | 0.79 |
| cg2114925 | 19 | 44,861,706 | 19q13.31 | -5.26 | 1.43E-07 | 0.51 | *ZNF112* | Intronic | -2.76 | 5.75E-03 | -3.09 | 1.98E-03 | -0.13 | 0.89 | -0.40 | 0.69 | -2.72 | 6.47E-03 | 51 | 0.09 |
| cg1982230 | 19 | 46,181,546 | 19q13.32 | -5.82 | 6.05E-09 | 0.52 | *GIPR* | Intronic | -4.57 | 4.89E-06 | -1.65 | 0.10 | -3.11 | 1.86E-03 | -1.83 | 0.07 | -1.11 | 0.27 | 0 | 0.67 |
| cg1102947 | 22 | 38,202,626 | 22q13.1 | 5.28 | 1.26E-07 | 0.36 | *H1F0* | 3'UTR | 3.85 | 1.17E-04 | 1.74 | 0.08 | 1.18 | 0.24 | 1.16 | 0.24 | 1.46 | 0.14 | 0 | 0.95 |
| cg1604880 | 22 | 38,203,755 | 22q13.1 | -6.16 | 7.10E-10 | 0.37 | *GCAT* | TSS200 | -5.31 | **1.08E-07** | -2.37 | 0.02 | -2.62 | 8.73E-03 | -1.35 | 0.18 | -0.23 | 0.82 | 0 | 0.60 |
| cg0837321 | 22 | 38,669,477 | 22q13.1 | -5.31 | 1.11E-07 | 0.62 | *TMEM184B* | TSS1500 | -4.98 | 6.50E-07 | -1.72 | 0.09 | -3.39 | 6.97E-04 | -0.79 | 0.43 | -0.32 | 0.75 | 19.8 | 0.29 |
| cg2450816 | 22 | 40,405,832 | 22q13.1 | 5.14 | 2.81E-07 | 0.52 | *FAM83F* | Intronic | 3.71 | 2.10E-04 | 3.05 | 2.27E-03 | 0.61 | 0.54 | 0.45 | 0.65 | 2.99 | 2.77E-03 | 23.3 | 0.27 |
| cg0163753 | 22 | 40,417,285 | 22q13.1 | -7.02 | 2.16E-12 | 0.39 | *FAM83F* | Intronic | -6.61 | **3.84E-11** | -2.46 | 0.01 | -0.67 | 0.50 | -1.75 | 0.08 | -2.87 | 4.07E-03 | 0 | 0.77 |
| cg1771650 | 22 | 40,600,549 | 22q13.1 | 8.17 | 3.21E-16 | 0.45 | *TNRC6B* | Intronic | 6.04 | **1.57E-09** | 2.31 | 0.02 | 1.35 | 0.18 | 1.48 | 0.14 | 4.08 | 4.52E-05 | 0 | 0.54 |
| cg2369896 | 22 | 40,772,232 | 22q13.1 | 7.98 | 1.45E-15 | 0.42 | *ADSL* ; *SGSM3* | Intronic | 6.49 | **8.37E-11** | 2.92 | 3.45E-03 | 0.70 | 0.48 | 2.22 | 0.03 | 3.86 | 1.14E-04 | 42.4 | 0.14 |
| cg0770420 | 22 | 40,805,579 | 22q13.1 | -5.47 | 4.38E-08 | 0.40 | *SGSM3* | Intronic | -4.01 | 5.99E-05 | -1.35 | 0.18 | -0.55 | 0.58 | -2.30 | 0.02 | -2.65 | 7.98E-03 | 26.3 | 0.25 |
| cg2744644 | 22 | 40,859,364 | 22q13.1 | 8.49 | 1.98E-17 | 0.40 | *MRTFA* | 5'UTR | 6.60 | **3.98E-11** | 2.48 | 0.01 | 1.66 | 0.10 | 0.87 | 0.38 | 3.19 | 1.40E-03 | 0 | 0.87 |
| cg2719941 | 22 | 40,901,737 | 22q13.1 | -6.13 | 9.04E-10 | 0.46 | *MRTFA* | Intronic | -4.49 | 7.29E-06 | -1.64 | 0.10 | -1.29 | 0.20 | 0.14 | 0.89 | -2.62 | 8.89E-03 | 0 | 0.83 |
| cg0078244 | 22 | 41,051,038 | 22q13.2 | 11.09 | 1.42E-28 | 0.49 | *MRTFA* ; *MCHR1* | Intergenic | 8.14 | **4.08E-16** | 3.45 | 5.54E-04 | 3.21 | 1.32E-03 | 2.21 | 0.03 | 6.26 | **3.75E-10** | 23.1 | 0.27 |
| cg0090242 | 22 | 41,074,991 | 22q13.2 | -7.94 | 2.04E-15 | 0.38 | *MCHR1* | TSS200 | -5.72 | **1.06E-08** | -3.46 | 5.46E-04 | -1.53 | 0.13 | -1.80 | 0.07 | -4.32 | 1.53E-05 | 10.2 | 0.35 |
| cg2027443 | 22 | 41,075,992 | 22q13.2 | -5.89 | 3.87E-09 | 0.39 | *MCHR1* | Intronic | -4.38 | 1.17E-05 | -2.46 | 0.01 | -0.70 | 0.48 | -0.72 | 0.47 | -4.08 | 4.52E-05 | 48.8 | 0.10 |
| cg0595141 | 22 | 41,310,349 | 22q13.2 | -5.44 | 5.32E-08 | 0.38 | *XPNPEP3* | Intronic | -4.23 | 2.32E-05 | -1.97 | 0.05 | 0.30 | 0.77 | -1.34 | 0.18 | -2.35 | 0.02 | 0 | 0.42 |

OR, odds ratio per standard deviation (SD) increase in genetically predicted DNA methylation level; CI, confidence interval; TSS, transcription start site; UTR, untranslated region.

**Supplementary Table 2. CpGs showing a significant association with colorectal cancer risk**

| CpG | Chr | Pos | Cytoband | Z score [a] | *P* [a] | R [b] | Closest gene | Classification |
|---|---|---|---|---|---|---|---|---|
| cg11332951 | 1 | 2,396,472 | 1p36.32 | -6.31 | 2.85E-10 | 0.27 | *PEX10* ; *PLCH2* | Intergenic |
| cg01981829 | 1 | 22,706,864 | 1p36.12 | 5.36 | 8.32E-08 | 0.45 | *MIR4418* ; *ZBTB40* | Intergenic |
| cg26255052 | 1 | 23,038,321 | 1p36.12 | -5.76 | 8.56E-09 | 0.21 | *EPHB2* | Intronic |
| cg12658694 | 1 | 38,397,304 | 1p34.3 | -7.98 | 1.44E-15 | 0.66 | *INPP5B* | Intronic |
| cg20409009 | 1 | 38,405,906 | 1p34.3 | -7.75 | 9.02E-15 | 0.37 | *INPP5B* | Intronic |
| cg26729965 | 1 | 38,455,861 | 1p34.3 | 8.30 | 1.02E-16 | 0.59 | *SF3A3* | TSS200 |
| cg26398791 | 1 | 38,455,867 | 1p34.3 | 7.37 | 1.76E-13 | 0.24 | *SF3A3* | TSS200 |
| cg18147567 | 1 | 38,457,103 | 1p34.3 | 8.35 | 6.88E-17 | 0.20 | *SF3A3* ; *FHL3* | Intergenic |
| cg05202204 | 1 | 38,460,950 | 1p34.3 | -8.42 | 3.62E-17 | 0.64 | *SF3A3* ; *FHL3* | Intergenic |
| cg09847153 | 1 | 38,461,540 | 1p34.3 | -8.21 | 2.20E-16 | 0.59 | *FHL3* | Downstream |
| cg17077180 | 1 | 38,461,687 | 1p34.3 | -8.20 | 2.35E-16 | 0.57 | *FHL3* | Downstream |
| cg04673462 | 1 | 38,461,896 | 1p34.3 | -8.39 | 4.84E-17 | 0.57 | *FHL3* | Downstream |
| cg26528311 | 1 | 38,462,546 | 1p34.3 | -7.67 | 1.71E-14 | 0.33 | *FHL3* | 3'UTR |
| cg16585132 | 1 | 38,464,943 | 1p34.3 | -6.05 | 1.49E-09 | 0.50 | *FHL3* | Exonic |
| cg10610196 | 1 | 38,465,296 | 1p34.3 | -6.91 | 4.97E-12 | 0.32 | *FHL3* | Intronic |
| cg24519393 | 1 | 38,470,540 | 1p34.3 | -8.45 | 2.89E-17 | 0.21 | *FHL3* | Intronic |
| cg23427534 | 1 | 38,480,016 | 1p34.3 | -6.15 | 7.68E-10 | 0.39 | *UTP11* | Intronic |
| cg02584939 | 1 | 55,248,099 | 1p32.3 | 7.76 | 8.29E-15 | 0.28 | *TTC22* | Exonic |
| cg14542186 | 1 | 55,257,914 | 1p32.3 | -5.75 | 9.09E-09 | 0.31 | *TTC22* | Intronic |
| cg18755616 [c] | 1 | 154,733,492 | 1q21.3 | 9.15 | 5.94E-20 | 0.62 | *KCNN3* | Intronic |
| cg07506908 [c] | 1 | 154,733,494 | 1q21.3 | 8.08 | 6.42E-16 | 0.57 | *KCNN3* | Intronic |
| cg20675262 | 1 | 172,874,951 | 1q24.3 | 5.26 | 1.44E-07 | 0.49 | *FASLG* ; *TNFSF18* | Intergenic |
| cg15503006 [c] | 1 | 178,994,942 | 1q25.2 | -5.37 | 8.07E-08 | 0.18 | *FAM20B* | 5'UTR |
| cg10931420 | 1 | 182,558,407 | 1q25.3 | 5.40 | 6.78E-08 | 0.18 | *RNASEL* | TSS200 |
| cg04472447 | 1 | 182,931,960 | 1q25.3 | -11.07 | 1.66E-28 | 0.27 | *SHCBP1L* ; *LAMC1* | TSS1500 |
| cg21523751 | 1 | 182,988,639 | 1q25.3 | -9.90 | 4.05E-23 | 0.22 | *SHCBP1L* ; *LAMC1* | TSS200 |
| cg00767544 | 1 | 183,013,562 | 1q25.3 | 8.76 | 1.89E-18 | 0.32 | *LAMC1* | TSS200 |
| cg11532216 | 1 | 183,063,517 | 1q25.3 | 10.15 | 3.17E-24 | 0.81 | *LAMC1* | TSS200 |
| cg07752015 | 1 | 183,079,647 | 1q25.3 | -9.84 | 7.87E-23 | 0.60 | *LAMC1* | TSS200 |
| cg17644690 | 1 | 183,093,894 | 1q25.3 | 8.68 | 3.98E-18 | 0.53 | *LAMC1* | Exonic |
| cg16418754 | 1 | 183,093,901 | 1q25.3 | -10.04 | 1.01E-23 | 0.30 | *LAMC1* | Exonic |
| cg07627133 | 1 | 183,094,584 | 1q25.3 | 9.78 | 1.43E-22 | 0.26 | *LAMC1* | Exonic |
| cg17352178 | 1 | 183,096,966 | 1q25.3 | 8.94 | 3.73E-19 | 0.37 | *LAMC1* | Intronic |
| cg07820189 | 1 | 183,149,515 | 1q25.3 | 5.85 | 5.03E-09 | 0.21 | *LAMC1* ; *LAMC2* | Intergenic |
| cg00208830 | 1 | 183,154,778 | 1q25.3 | 5.72 | 1.06E-08 | 0.36 | *LAMC2* | TSS1500 |
| cg12552165 | 1 | 183,154,981 | 1q25.3 | 5.39 | 6.96E-08 | 0.59 | *LAMC2* | TSS200 |
| cg05366923 | 1 | 183,155,056 | 1q25.3 | 5.71 | 1.11E-08 | 0.43 | *LAMC2* | TSS200 |
| cg01949993 | 1 | 183,155,116 | 1q25.3 | 5.98 | 2.22E-09 | 0.67 | *LAMC2* | TSS200 |
| cg05929019 | 1 | 183,155,154 | 1q25.3 | 6.00 | 2.01E-09 | 0.73 | *LAMC2* | TSS200 |
| cg21378578 | 1 | 183,155,235 | 1q25.3 | 5.83 | 5.68E-09 | 0.60 | *LAMC2* | 5'UTR |
| cg04218899 | 1 | 183,155,366 | 1q25.3 | 5.95 | 2.66E-09 | 0.66 | *LAMC2* | 5'UTR |
| cg06211724 | 1 | 183,155,437 | 1q25.3 | 5.90 | 3.69E-09 | 0.55 | *LAMC2* | 5'UTR |
| cg23696949 | 1 | 183,155,644 | 1q25.3 | 5.74 | 9.49E-09 | 0.54 | *LAMC2* | Intronic |
| cg02767663 | 1 | 183,167,718 | 1q25.3 | 5.85 | 5.00E-09 | 0.46 | *LAMC2* | Intronic |
| cg10581625 | 1 | 183,208,626 | 1q25.3 | 5.39 | 7.23E-08 | 0.16 | *LAMC2* | Exonic |
| cg19755016 | 1 | 183,261,947 | 1q25.3 | -6.70 | 2.10E-11 | 0.18 | *NMNAT2* | Exonic |
| cg06934615 | 1 | 183,441,736 | 1q25.3 | 5.25 | 1.48E-07 | 0.18 | *SMG7* | 5'UTR |
| cg25008509 | 1 | 183,562,693 | 1q25.3 | -5.65 | 1.64E-08 | 0.46 | *NCF2* ; *ARPC5* | Intergenic |
| cg04041654 | 1 | 183,579,470 | 1q25.3 | 5.64 | 1.74E-08 | 0.29 | *NCF2* ; *ARPC5* | Intergenic |
| cg26915799 | 1 | 201,852,272 | 1q32.1 | -5.54 | 3.02E-08 | 0.17 | *IPO9* | 3'UTR |
| cg10766380 | 1 | 221,928,176 | 1q41 | 5.31 | 1.07E-07 | 0.17 | *DUSP10* ; *LINC01655* | Intergenic |
| cg23805594 | 1 | 221,933,302 | 1q41 | -5.39 | 7.06E-08 | 0.34 | *DUSP10* ; *LINC01655* | Intergenic |
| cg14231155 | 1 | 221,939,069 | 1q41 | 5.42 | 6.12E-08 | 0.18 | *DUSP10* ; *LINC01655* | Intergenic |
| cg08747931 | 1 | 222,014,008 | 1q41 | 6.16 | 7.25E-10 | 0.37 | *LINC01655* | Exonic |
| cg17131975 | 1 | 222,039,219 | 1q41 | 6.86 | 6.74E-12 | 0.52 | *LINC01655* ; *LINC02257* | Intergenic |
| cg19445579 | 1 | 222,065,665 | 1q41 | 6.25 | 4.04E-10 | 0.52 | *LINC01655* ; *LINC02257* | Intergenic |
| cg13871948 | 1 | 222,071,062 | 1q41 | -6.68 | 2.42E-11 | 0.52 | *LINC01655* ; *LINC02257* | Intergenic |

| | | | | | | | | |
|---|---|---|---|---|---|---|---|---|
| cg14124465 | 1 | 222,082,182 | 1q41 | 6.10 | 1.07E-09 | 0.27 | *LINC01655* ; *LINC02257* | Intergenic |
| cg08564019 | 1 | 222,164,794 | 1q41 | 5.25 | 1.52E-07 | 0.35 | *LINC02474* ; *HHIPL2* | Intergenic |
| cg23713008 | 1 | 222,224,071 | 1q41 | 7.83 | 4.85E-15 | 0.27 | *LINC02474* ; *HHIPL2* | Intergenic |
| cg24722070 | 1 | 222,227,490 | 1q41 | 6.00 | 1.95E-09 | 0.22 | *LINC02474* ; *HHIPL2* | Intergenic |
| cg07827139 | 2 | 159,992,206 | 2q24.2 | -5.20 | 1.99E-07 | 0.19 | *TANC1* | Intronic |
| cg10150255 | 2 | 169,025,365 | 2q24.3 | -5.30 | 1.16E-07 | 0.51 | *STK39* | Intronic |
| cg05392844 | 2 | 199,639,143 | 2q33.1 | 5.34 | 9.27E-08 | 0.20 | *LINC01923* ; *SATB2* | Intergenic |
| cg04112147 | 2 | 199,778,914 | 2q33.1 | -8.01 | 1.16E-15 | 0.63 | *LINC01923* ; *SATB2* | Intergenic |
| cg20230808 | 2 | 200,055,484 | 2q33.1 | 8.19 | 2.64E-16 | 0.24 | *LINC01923* ; *SATB2* | Intergenic |
| cg09024069 | 2 | 200,126,093 | 2q33.1 | 6.96 | 3.47E-12 | 0.83 | *LINC01923* ; *SATB2* | Intergenic |
| cg22973789 | 2 | 200,321,011 | 2q33.1 | 6.38 | 1.79E-10 | 0.17 | *SATB2* | Intronic |
| cg26101402 | 2 | 200,323,147 | 2q33.1 | -5.26 | 1.42E-07 | 0.17 | *SATB2* | Intronic |
| cg02329011 | 2 | 200,330,901 | 2q33.1 | 5.21 | 1.89E-07 | 0.23 | *SATB2* | Intronic |
| cg16518266 | 2 | 218,621,552 | 2q35 | -6.14 | 8.03E-10 | 0.19 | *DIRC3* | TSS1500 |
| cg20131290 | 2 | 218,990,364 | 2q35 | -5.34 | 9.08E-08 | 0.52 | *CXCR2* | TSS200 |
| cg10591797 | 2 | 218,990,598 | 2q35 | -5.63 | 1.75E-08 | 0.26 | *CXCR2* | Intronic |
| cg17536959 | 2 | 219,021,022 | 2q35 | 5.82 | 5.86E-09 | 0.26 | *CXCR2* ; *CXCR1* | Intergenic |
| cg22635982 | 2 | 219,080,665 | 2q35 | 6.43 | 1.32E-10 | 0.37 | *CXCR1* ; *ARPC2* | Intergenic |
| cg00012203 | 2 | 219,082,015 | 2q35 | 6.75 | 1.48E-11 | 0.80 | *ARPC2* | Intronic |
| cg04731861 | 2 | 219,085,781 | 2q35 | -6.79 | 1.14E-11 | 0.69 | *ARPC2* | Intronic |
| cg17651887 | 2 | 219,117,224 | 2q35 | 8.21 | 2.13E-16 | 0.17 | *ARPC2* | Intronic |
| cg19710355 | 2 | 219,124,882 | 2q35 | 8.53 | 1.46E-17 | 0.47 | *GPBAR1* | 5'UTR |
| cg20019365 | 2 | 219,134,978 | 2q35 | 8.17 | 2.97E-16 | 0.82 | *AAMP* ; *PNKD* | TSS200 |
| cg08494069 | 2 | 219,144,417 | 2q35 | 8.00 | 1.20E-15 | 0.34 | *PNKD* ; *TMBIM1* | Intronic |
| cg09474840 | 2 | 219,146,931 | 2q35 | -7.89 | 2.94E-15 | 0.29 | *PNKD* ; *TMBIM1* | Intronic |
| cg13835894 | 2 | 219,148,914 | 2q35 | 5.80 | 6.52E-09 | 0.49 | *PNKD* ; *TMBIM1* | Intronic |
| cg01329789 | 2 | 219,157,515 | 2q35 | 6.01 | 1.85E-09 | 0.18 | *PNKD* | Intronic |
| cg02383163 | 2 | 219,157,675 | 2q35 | 7.97 | 1.64E-15 | 0.16 | *PNKD* | Intronic |
| cg03371758 | 2 | 219,181,427 | 2q35 | 5.71 | 1.14E-08 | 0.25 | *PNKD* | Intronic |
| cg05991184 | 2 | 219,186,017 | 2q35 | -7.72 | 1.17E-14 | 0.46 | *PNKD* | Intronic |
| cg25618457 | 2 | 219,187,295 | 2q35 | -8.37 | 5.65E-17 | 0.29 | *PNKD* | Intronic |
| cg04184094 | 2 | 219,187,299 | 2q35 | -8.43 | 3.58E-17 | 0.48 | *PNKD* | Intronic |
| cg15100426 | 2 | 219,187,432 | 2q35 | -8.11 | 5.15E-16 | 0.19 | *PNKD* | Intronic |
| cg04018708 | 2 | 219,188,804 | 2q35 | 8.38 | 5.50E-17 | 0.31 | *PNKD* | Intronic |
| cg04050850 | 2 | 219,536,800 | 2q35 | -6.54 | 6.35E-11 | 0.22 | *RNF25* | 5'UTR |
| cg03364247 | 3 | 40,570,062 | 3p22.1 | -8.53 | 1.47E-17 | 0.20 | *ZNF621* | Intronic |
| cg11530593 | 3 | 52,022,173 | 3p21.2 | -5.56 | 2.76E-08 | 0.17 | *ABHD14A-ACY1* ; *ACY1* | Intronic |
| cg00690270 | 3 | 52,324,148 | 3p21.1 | -5.25 | 1.55E-07 | 0.59 | *GLYCTK* | Intronic |
| cg19999141 | 3 | 52,424,765 | 3p21.1 | -6.12 | 9.43E-10 | 0.21 | *DNAH1* | Intronic |
| cg21742876 | 3 | 52,817,404 | 3p21.1 | -5.65 | 1.57E-08 | 0.52 | *ITIH1* | Intronic |
| cg06204229 | 3 | 52,865,917 | 3p21.1 | -6.84 | 7.67E-12 | 0.69 | *ITIH4* ; *MUSTN1* | Intergenic |
| cg03278365 | 3 | 52,877,203 | 3p21.1 | -5.30 | 1.17E-07 | 0.23 | *STIMATE* ; *STIMATE-MUSTN1* | Intronic |
| cg25058201 | 3 | 52,877,271 | 3p21.1 | 5.57 | 2.52E-08 | 0.68 | *STIMATE* ; *STIMATE-MUSTN1* | Intronic |
| cg13643186 | 3 | 52,877,402 | 3p21.1 | -5.67 | 1.41E-08 | 0.32 | *STIMATE* ; *STIMATE-MUSTN1* | Intronic |
| cg25367572 | 3 | 52,926,875 | 3p21.1 | 6.48 | 8.95E-11 | 0.24 | *STIMATE* ; *STIMATE-MUSTN1* | Intronic |
| cg04865290 | 3 | 52,927,548 | 3p21.1 | 6.64 | 3.04E-11 | 0.60 | *STIMATE* ; *STIMATE-MUSTN1* | Intronic |
| cg20625060 | 3 | 52,927,957 | 3p21.1 | 5.54 | 3.06E-08 | 0.47 | *STIMATE* ; *STIMATE-MUSTN1* | Intronic |
| cg11613427 | 3 | 52,932,208 | 3p21.1 | 5.81 | 6.27E-09 | 0.27 | *TMEM110* ; *TMEM110-MUSTN1* | TSS1500 |
| cg09977701 | 3 | 53,060,719 | 3p21.1 | -6.59 | 4.48E-11 | 0.23 | *SFMBT1* | Intronic |
| cg11375938 | 3 | 53,085,910 | 3p21.1 | 5.76 | 8.26E-09 | 0.24 | *SFMBT1* ; *RFT1* | Intergenic |
| cg26965574 [c] | 3 | 57,963,665 | 3p14.3 | 6.51 | 7.75E-11 | 0.17 | *SLMAP* ; *FLNB* | Intergenic |
| cg06965496 [c] | 3 | 58,063,095 | 3p14.3 | 10.84 | 2.14E-27 | 0.23 | *FLNB* | Intronic |
| cg00225902 | 3 | 66,444,122 | 3p14.1 | -5.24 | 1.60E-07 | 0.30 | *LRIG1* | Intronic |
| cg16813459 | 3 | 100,430,811 | 3q12.2 | -6.49 | 8.59E-11 | 0.23 | *TFG* | Intronic |
| cg06607226 | 3 | 100,617,729 | 3q12.2 | 8.25 | 1.58E-16 | 0.33 | *ABI3BP* | Exonic |
| cg24683684 [c] | 3 | 107,318,514 | 3q13.12 | -6.80 | 1.03E-11 | 0.23 | *BBX* | Intronic |
| cg23646888 | 3 | 112,929,672 | 3q13.2 | -5.69 | 1.28E-08 | 0.20 | *BOC* | TSS1500 |
| cg20083609 | 3 | 112,980,454 | 3q13.2 | -5.50 | 3.75E-08 | 0.29 | *BOC* | TSS1500 |

| | | | | | | | | |
|---|---|---|---|---|---|---|---|---|
| cg07708788 | 3 | 133,748,311 | 3q22.2 | 7.06 | 1.62E-12 | 0.24 | *SLCO2A1* | Intronic |
| cg04531097 | 3 | 133,931,867 | 3q22.2 | -5.59 | 2.22E-08 | 0.23 | *RYK* | Intronic |
| cg07283011 | 3 | 133,970,158 | 3q22.2 | -5.44 | 5.20E-08 | 0.28 | *RYK* | TSS1500 |
| cg00169633 | 3 | 134,093,551 | 3q22.2 | -5.32 | 1.06E-07 | 0.25 | *AMOTL2* | TSS1500 |
| cg14222479 | 3 | 169,487,675 | 3q26.2 | -5.92 | 3.30E-09 | 0.26 | *ACTRT3* | 5'UTR |
| cg21687591 | 3 | 169,492,002 | 3q26.2 | 6.73 | 1.67E-11 | 0.28 | *MYNN* | Intronic |
| cg26469215 | 4 | 105,888,253 | 4q24 | 6.00 | 1.95E-09 | 0.40 | *CXXC4-AS1* ; *TET2* | Intergenic |
| cg01769750 | 4 | 105,888,571 | 4q24 | 6.86 | 6.69E-12 | 0.26 | *CXXC4-AS1* ; *TET2* | Intergenic |
| cg05865911 | 4 | 106,066,844 | 4q24 | 5.77 | 7.94E-09 | 0.25 | *TET2* | TSS1500 |
| cg17862558 | 4 | 106,137,048 | 4q24 | -6.69 | 2.28E-11 | 0.39 | *TET2-AS1* | Intronic |
| cg10945313 [c] | 4 | 113,739,291 | 4q25 | 5.28 | 1.31E-07 | 0.16 | *ANK2* | 5'UTR |
| cg11195926 | 4 | 145,621,327 | 4q31.21 | -6.96 | 3.43E-12 | 0.58 | *HHIP* | Intronic |
| cg11697194 | 5 | 964,196 | 5p15.33 | -5.79 | 7.23E-09 | 0.23 | *TRIP13* ; *LOC100506688* | Intergenic |
| cg01101522 | 5 | 1,037,853 | 5p15.33 | 7.07 | 1.53E-12 | 0.16 | *NKD2* | Intronic |
| cg27010096 | 5 | 1,060,613 | 5p15.33 | 5.79 | 7.25E-09 | 0.56 | *SLC12A7* | Intronic |
| cg07384136 | 5 | 1,115,158 | 5p15.33 | -6.07 | 1.29E-09 | 0.19 | *SLC12A7* ; *CTD-3080P12.3* | Intergenic |
| cg06593603 | 5 | 1,218,042 | 5p15.33 | 7.79 | 6.91E-15 | 0.43 | *SLC6A19* | Intronic |
| cg02327812 | 5 | 1,218,507 | 5p15.33 | -8.87 | 7.31E-19 | 0.24 | *SLC6A19* | Intronic |
| cg14323943 | 5 | 1,226,480 | 5p15.33 | 8.28 | 1.26E-16 | 0.48 | *SLC6A18* | Intronic |
| cg11908978 | 5 | 1,236,791 | 5p15.33 | 8.72 | 2.77E-18 | 0.31 | *SLC6A18* | Intronic |
| cg21817287 | 5 | 40,066,956 | 5p13.1 | 7.94 | 1.96E-15 | 0.26 | *LINC00603* ; *PTGER4* | Intergenic |
| cg06784095 | 5 | 40,151,155 | 5p13.1 | -5.51 | 3.69E-08 | 0.78 | *LINC00603* ; *PTGER4* | Intergenic |
| cg21879236 | 5 | 40,227,327 | 5p13.1 | 5.16 | 2.43E-07 | 0.21 | *LINC00603* ; *PTGER4* | Intergenic |
| cg22637518 | 5 | 40,260,640 | 5p13.1 | -8.75 | 2.23E-18 | 0.36 | *LINC00603* ; *PTGER4* | Intergenic |
| cg09756010 | 5 | 40,260,734 | 5p13.1 | -7.50 | 6.60E-14 | 0.54 | *LINC00603* ; *PTGER4* | Intergenic |
| cg00928088 | 5 | 40,268,273 | 5p13.1 | 8.39 | 4.68E-17 | 0.29 | *LINC00603* ; *PTGER4* | Intergenic |
| cg17138835 | 5 | 40,269,596 | 5p13.1 | -12.27 | 1.37E-34 | 0.78 | *LINC00603* ; *PTGER4* | Intergenic |
| cg04287854 | 5 | 40,276,980 | 5p13.1 | -12.35 | 4.71E-35 | 0.70 | *LINC00603* ; *PTGER4* | Intergenic |
| cg08192208 | 5 | 133,970,810 | 5q31.1 | -5.22 | 1.83E-07 | 0.25 | *SAR1B* ; *SEC24A* | Intergenic |
| cg10740944 | 5 | 134,367,807 | 5q31.1 | -7.52 | 5.52E-14 | 0.19 | *PITX1* | Intronic |
| cg26452341 | 5 | 134,446,260 | 5q31.1 | 8.44 | 3.17E-17 | 0.70 | *C5orf66* | Intronic |
| cg00070172 | 5 | 134,451,867 | 5q31.1 | -6.08 | 1.21E-09 | 0.19 | *C5orf66* | Intronic |
| cg26493306 | 5 | 134,463,458 | 5q31.1 | 9.18 | 4.30E-20 | 0.38 | *C5orf66* | Intronic |
| cg05835599 | 5 | 134,469,813 | 5q31.1 | 10.66 | 1.63E-26 | 0.24 | *C5orf66* | Intronic |
| cg15128679 | 5 | 134,476,728 | 5q31.1 | 8.02 | 1.06E-15 | 0.26 | *C5orf66* | Intronic |
| cg15732431 | 5 | 134,478,061 | 5q31.1 | 10.37 | 3.42E-25 | 0.51 | *C5orf66* | Intronic |
| cg06148154 | 5 | 134,510,531 | 5q31.1 | -8.37 | 5.76E-17 | 0.81 | *C5orf66* | Intronic |
| cg21147203 | 5 | 134,529,659 | 5q31.1 | 7.17 | 7.78E-13 | 0.67 | *C5orf66* | Intronic |
| cg07474586 | 5 | 134,530,497 | 5q31.1 | 7.08 | 1.49E-12 | 0.20 | *C5orf66* | Intronic |
| cg14671011 | 5 | 134,544,423 | 5q31.1 | 8.37 | 5.54E-17 | 0.34 | *C5orf66* | Intronic |
| cg22034072 | 5 | 134,551,974 | 5q31.1 | -6.28 | 3.48E-10 | 0.46 | *C5orf66* | Intronic |
| cg16603653 | 5 | 134,566,929 | 5q31.1 | -5.41 | 6.13E-08 | 0.22 | *C5orf66* | Intronic |
| cg01146539 | 5 | 134,688,623 | 5q31.1 | 6.40 | 1.54E-10 | 0.16 | *H2AFY* | Intronic |
| cg03172688 [c] | 5 | 141,254,129 | 5q31.3 | -6.78 | 1.22E-11 | 0.30 | *PCDH1* | Intronic |
| cg09661503 | 5 | 149,568,693 | 5q32 | 6.03 | 1.62E-09 | 0.56 | *SLC6A7* | TSS1500 |
| cg21212995 | 5 | 149,570,189 | 5q32 | -6.41 | 1.49E-10 | 0.21 | *SLC6A7* | Intronic |
| cg10782034 | 5 | 172,323,758 | 5q35.1 | -5.55 | 2.83E-08 | 0.44 | *ERGIC1* | Intronic |
| cg13582457 | 5 | 172,324,773 | 5q35.1 | -5.57 | 2.50E-08 | 0.79 | *ERGIC1* | Intronic |
| cg27518156 | 6 | 6,751,984 | 6p25.1 | -5.19 | 2.11E-07 | 0.42 | *LY86* ; *RREB1* | Intergenic |
| cg17617182 | 6 | 12,279,872 | 6p24.1 | -6.29 | 3.20E-10 | 0.22 | *HIVEP1* ; *EDN1* | Intergenic |
| cg27565517 | 6 | 12,292,676 | 6p24.1 | -7.29 | 3.21E-13 | 0.82 | *EDN1* | Exonic |
| cg24248978 [c] | 6 | 28,049,235 | 6p22.1 | -7.76 | 8.77E-15 | 0.16 | *ZNF165* | 5'UTR |
| cg14173258 [d] | 6 | 28,832,474 | 6p22.1 | 5.61 | 2.01E-08 | 0.16 | *LINC01623* ; *HCG14* | Intergenic |
| cg26481402 | 6 | 28,853,050 | 6p22.1 | -5.17 | 2.35E-07 | 0.57 | *LINC01623* ; *HCG14* | Intergenic |
| cg15490715 | 6 | 29,521,568 | 6p22.1 | -5.23 | 1.73E-07 | 0.20 | *LINC01015* ; *UBD* | Intergenic |
| cg10474949 [d] | 6 | 29,678,319 | 6p22.1 | 7.10 | 1.25E-12 | 0.26 | *ZFP57* ; *HLA-F* | Intergenic |
| cg11441173 [d] | 6 | 29,707,887 | 6p22.1 | 5.44 | 5.38E-08 | 0.42 | *HLA-F-AS1* | Intronic |
| cg17105013 [d] | 6 | 29,716,712 | 6p22.1 | -6.73 | 1.67E-11 | 0.19 | *HLA-F-AS1* | Exonic |
| cg03835151 | 6 | 29,727,844 | 6p22.1 | 5.26 | 1.44E-07 | 0.53 | *IFITM4P* ; *HCG4* | Intergenic |

| | | | | | | | | |
|---|---|---|---|---|---|---|---|---|
| cg03432955 | 6 | 29,795,501 | 6p22.1 | -5.46 | 4.85E-08 | 0.83 | *HLA-G* | Intronic |
| cg00126638 | 6 | 29,795,583 | 6p22.1 | -5.34 | 9.19E-08 | 0.81 | *HLA-G* | 5'UTR |
| cg03521696 | 6 | 29,795,595 | 6p22.1 | -5.27 | 1.38E-07 | 0.76 | *HLA-G* | 5'UTR |
| cg21529533 | 6 | 29,796,015 | 6p22.1 | -5.22 | 1.75E-07 | 0.69 | *HLA-G* | Exonic |
| cg17102747 | 6 | 30,076,980 | 6p22.1 | -5.18 | 2.23E-07 | 0.17 | *TRIM31-AS1* | Intronic |
| cg03368643 [d] | 6 | 30,181,600 | 6p22.1 | 5.26 | 1.44E-07 | 0.26 | *TRIM26* | TSS1500 |
| cg23071690 [d] | 6 | 30,232,015 | 6p22.1 | 5.54 | 3.07E-08 | 0.20 | *HLA-L* | Exonic |
| cg20595636 | 6 | 30,279,111 | 6p22.1 | -5.96 | 2.47E-09 | 0.26 | *HCG17* ; *HCG18* | Intronic |
| cg23778943 | 6 | 30,421,189 | 6p21.33 | -5.75 | 9.19E-09 | 0.34 | *RPP21* ; *HLA-E* | Intergenic |
| cg27375351 | 6 | 30,452,529 | 6p21.33 | -5.16 | 2.43E-07 | 0.17 | *RPP21; HLA-E* | Intergenic |
| cg21758773 | 6 | 30,458,730 | 6p21.33 | -5.49 | 4.01E-08 | 0.27 | *HLA-E* | Intronic |
| cg17615629 | 6 | 30,459,867 | 6p21.33 | -5.32 | 1.04E-07 | 0.27 | *HLA-E* | Intronic |
| cg22410026 | 6 | 30,483,923 | 6p21.33 | 5.74 | 9.64E-09 | 0.20 | *LINC02569* | TSS1500 |
| cg06251929 | 6 | 30,539,459 | 6p21.33 | 5.25 | 1.50E-07 | 0.39 | *ABCF1* | Intronic |
| cg05177685 | 6 | 30,558,470 | 6p21.33 | 5.30 | 1.19E-07 | 0.45 | *ABCF1* | Exonic |
| cg16212881 | 6 | 30,640,519 | 6p21.33 | 7.74 | 9.62E-15 | 0.17 | *DHX16* | Exonic |
| cg13357602 | 6 | 30,655,720 | 6p21.33 | -5.45 | 5.04E-08 | 0.17 | *PPP1R18* | TSS1500 |
| cg06596877 | 6 | 30,659,513 | 6p21.33 | 5.45 | 5.03E-08 | 0.38 | *NRM* | TSS1500 |
| cg05505711 [d] | 6 | 30,714,519 | 6p21.33 | 5.40 | 6.80E-08 | 0.71 | *IER3* ; *HCG20* | Intergenic |
| cg18730733 | 6 | 30,816,447 | 6p21.33 | 5.24 | 1.63E-07 | 0.39 | *LINC02570* | TSS1500 |
| cg24875484 | 6 | 30,910,583 | 6p21.33 | -6.03 | 1.61E-09 | 0.31 | *MUCL3* | Intronic |
| cg06855286 | 6 | 31,010,597 | 6p21.33 | -6.05 | 1.43E-09 | 0.58 | *MUC22* ; *HCG22* | Intergenic |
| cg16398511 | 6 | 31,010,701 | 6p21.33 | -5.81 | 6.17E-09 | 0.65 | *MUC22* ; *HCG22* | Intergenic |
| cg18595780 | 6 | 31,010,742 | 6p21.33 | -6.65 | 2.95E-11 | 0.69 | *MUC22* ; *HCG22* | Intergenic |
| cg14259707 | 6 | 31,021,957 | 6p21.33 | -6.57 | 5.12E-11 | 0.18 | *HCG22* | 5'UTR |
| cg07717512 | 6 | 31,026,287 | 6p21.33 | -5.89 | 3.85E-09 | 0.18 | *HCG22* | Intronic |
| cg02451928 | 6 | 31,125,948 | 6p21.33 | 5.55 | 2.92E-08 | 0.43 | *CCHCR1* | 5'UTR |
| cg00160818 [d] | 6 | 31,126,356 | 6p21.33 | 5.54 | 2.96E-08 | 0.19 | *CCHCR1* ; *TCF19* | TSS1500 |
| cg15948871 [d] | 6 | 31,139,620 | 6p21.33 | 5.47 | 4.58E-08 | 0.42 | *POU5F1* ; *PSORS1C3* | Intergenic |
| cg17931227 | 6 | 31,148,370 | 6p21.33 | -5.24 | 1.64E-07 | 0.54 | *PSORS1C3* | Intronic |
| cg09179646 | 6 | 31,148,383 | 6p21.33 | -5.31 | 1.08E-07 | 0.55 | *PSORS1C3* | Intronic |
| cg22291762 | 6 | 31,148,404 | 6p21.33 | -5.41 | 6.28E-08 | 0.55 | *PSORS1C3* | Intronic |
| cg24427850 | 6 | 31,148,409 | 6p21.33 | -5.20 | 1.98E-07 | 0.53 | *PSORS1C3* | Intronic |
| cg26668675 | 6 | 31,148,463 | 6p21.33 | -5.33 | 9.65E-08 | 0.53 | *PSORS1C3* | Intronic |
| cg03078486 | 6 | 31,148,474 | 6p21.33 | -5.25 | 1.49E-07 | 0.54 | *PSORS1C3* | Intronic |
| cg26818629 | 6 | 31,148,516 | 6p21.33 | -5.47 | 4.47E-08 | 0.46 | *PSORS1C3* | Intronic |
| cg14036627 | 6 | 31,148,657 | 6p21.33 | -5.19 | 2.06E-07 | 0.52 | *PSORS1C3* | Intronic |
| cg23457357 | 6 | 31,148,748 | 6p21.33 | -5.75 | 9.07E-09 | 0.51 | *PSORS1C3* | Intronic |
| cg03921216 | 6 | 31,166,799 | 6p21.33 | -5.54 | 2.99E-08 | 0.41 | *HCG27* | Intronic |
| cg02773045 [d] | 6 | 31,168,568 | 6p21.33 | 6.51 | 7.47E-11 | 0.19 | *HCG27* | Intronic |
| cg10367909 | 6 | 31,367,038 | 6p21.33 | 5.36 | 8.26E-08 | 0.52 | *MICA-AS1* | Intronic |
| cg21001008 | 6 | 31,368,503 | 6p21.33 | 5.21 | 1.94E-07 | 0.29 | *MICA* | 5'UTR |
| cg18892128 | 6 | 31,409,575 | 6p21.33 | 6.18 | 6.22E-10 | 0.80 | *LINC01149* | Exonic |
| cg05054864 [d] | 6 | 31,458,496 | 6p21.33 | -6.93 | 4.24E-12 | 0.49 | *MICB-DT* | Intronic |
| cg14646094 | 6 | 31,496,752 | 6p21.33 | 5.35 | 9.00E-08 | 0.34 | *MCCD1* | 5'UTR |
| cg20248972 [d] | 6 | 31,515,526 | 6p21.33 | 5.54 | 3.10E-08 | 0.16 | *NFKBIL1* | Exonic |
| cg09037630 [d] | 6 | 31,528,239 | 6p21.33 | -6.98 | 3.01E-12 | 0.24 | *LOC100287329* | Exonic |
| cg00983060 | 6 | 31,531,190 | 6p21.33 | 5.81 | 6.38E-09 | 0.40 | *LOC100287329* | Intronic |
| cg09736959 | 6 | 31,540,114 | 6p21.33 | 5.68 | 1.35E-08 | 0.78 | *LOC100287329* | Intronic |
| cg24216966 | 6 | 31,540,121 | 6p21.33 | 5.66 | 1.53E-08 | 0.73 | *LOC100287329* | Intronic |
| cg11586857 | 6 | 31,540,136 | 6p21.33 | 5.74 | 9.32E-09 | 0.65 | *LOC100287329* | Intronic |
| cg17709873 | 6 | 31,540,456 | 6p21.33 | 5.58 | 2.43E-08 | 0.65 | *LOC100287329* | Exonic |
| cg21222743 [d] | 6 | 31,543,545 | 6p21.33 | -5.52 | 3.35E-08 | 0.21 | *TNF* | Exonic |
| cg08610982 [d] | 6 | 31,560,871 | 6p21.33 | -5.55 | 2.80E-08 | 0.29 | *NCR3* | TSS200 |
| cg00124375 | 6 | 31,698,218 | 6p21.33 | 5.35 | 8.63E-08 | 0.30 | *DDAH2* | TSS1500 |
| cg17983217 | 6 | 31,698,223 | 6p21.33 | 5.58 | 2.36E-08 | 0.32 | *DDAH2* | TSS1500 |
| cg00686197 [d] | 6 | 31,733,619 | 6p21.33 | 5.78 | 7.61E-09 | 0.35 | *VWA7* | Intronic |
| cg02002586 | 6 | 31,733,894 | 6p21.33 | 5.39 | 7.12E-08 | 0.30 | *VWA7* | Intronic |

| | | | | | | | | |
|---|---|---|---|---|---|---|---|---|
| cg12988118 [d] | 6 | 31,744,524 | 6p21.33 | 5.83 | 5.56E-09 | 0.30 | *VWA7* | Exonic |
| cg27143082 [d] | 6 | 31,744,628 | 6p21.33 | 5.98 | 2.25E-09 | 0.25 | *VWA7* | Intronic |
| cg24065328 [d] | 6 | 31,744,735 | 6p21.33 | 5.56 | 2.67E-08 | 0.28 | *VWA7* | Intronic |
| cg19868593 | 6 | 31,744,816 | 6p21.33 | 5.84 | 5.33E-09 | 0.31 | *VWA7* | Intronic |
| cg24508713 | 6 | 31,870,783 | 6p21.33 | 5.27 | 1.34E-07 | 0.22 | *C2* | Intronic |
| cg12460721 [d] | 6 | 31,922,556 | 6p21.33 | 6.00 | 1.97E-09 | 0.19 | *NELFE* | Exonic |
| cg14218053 | 6 | 32,049,376 | 6p21.33 | 5.69 | 1.28E-08 | 0.28 | *TNXB* | Exonic |
| cg16496219 | 6 | 32,085,017 | 6p21.33 | 6.94 | 3.86E-12 | 0.28 | *ATF6B* | Intronic |
| cg02429905 | 6 | 32,119,944 | 6p21.32 | 5.91 | 3.43E-09 | 0.20 | *PRRT1* | 5'UTR |
| cg04749507 | 6 | 32,120,203 | 6p21.32 | 5.20 | 1.97E-07 | 0.23 | *PRRT1* | Intronic |
| cg12738718 | 6 | 32,121,204 | 6p21.32 | -5.63 | 1.84E-08 | 0.23 | *LOC100507547* | Exonic |
| cg05133205 | 6 | 32,121,249 | 6p21.32 | -5.47 | 4.42E-08 | 0.25 | *LOC100507547* | Exonic |
| cg04105091 [d] | 6 | 32,121,355 | 6p21.32 | -6.95 | 3.63E-12 | 0.21 | *LOC100507547* | Exonic |
| cg12585943 | 6 | 32,121,368 | 6p21.32 | -5.20 | 2.01E-07 | 0.20 | *LOC100507547* | Exonic |
| cg00086577 | 6 | 32,122,894 | 6p21.32 | 5.55 | 2.94E-08 | 0.24 | *PPT2* | Exonic |
| cg06902929 | 6 | 32,123,258 | 6p21.32 | 7.89 | 3.11E-15 | 0.20 | *PPT2-EGFL8* | Intronic |
| cg11122280 | 6 | 32,123,479 | 6p21.32 | -5.47 | 4.58E-08 | 0.24 | *PPT2* | Exonic |
| cg15829057 | 6 | 32,129,834 | 6p21.32 | -6.92 | 4.58E-12 | 0.16 | *PPT2-EGFL8* | Intronic |
| cg02681370 | 6 | 32,156,482 | 6p21.32 | 6.09 | 1.10E-09 | 0.33 | *PBX2* | Intronic |
| cg07324546 | 6 | 32,261,558 | 6p21.32 | -5.26 | 1.46E-07 | 0.17 | *TSBP1* | Exonic |
| cg11539695 | 6 | 32,287,314 | 6p21.32 | 6.22 | 4.98E-10 | 0.27 | *TSBP1-AS1* | Intronic |
| cg06545464 | 6 | 32,299,649 | 6p21.32 | -6.10 | 1.07E-09 | 0.17 | *TSBP1-AS1* | Intronic |
| cg09817599 | 6 | 32,326,581 | 6p21.32 | 6.08 | 1.21E-09 | 0.21 | *TSBP1-AS1* | Intronic |
| cg22863148 | 6 | 32,339,554 | 6p21.32 | -5.58 | 2.41E-08 | 0.20 | *TSBP1-AS1* | Intronic |
| cg04797846 | 6 | 32,362,707 | 6p21.32 | -6.90 | 5.14E-12 | 0.35 | *BTNL2* | Exonic |
| cg01626341 | 6 | 32,440,987 | 6p21.32 | -5.48 | 4.34E-08 | 0.66 | *HLA-DRA* ; *HLA-DRB5* | Intergenic |
| cg00440797 | 6 | 32,493,873 | 6p21.32 | -6.56 | 5.28E-11 | 0.76 | *HLA-DRB5* | Intronic |
| cg14645244 | 6 | 32,552,205 | 6p21.32 | 6.71 | 1.94E-11 | 0.71 | *HLA-DRB1* | Intronic |
| cg07007382 | 6 | 32,578,070 | 6p21.32 | -5.62 | 1.96E-08 | 0.78 | *HLA-DRB1* ; *HLA-DQA1* | Intergenic |
| cg08831072 | 6 | 32,685,319 | 6p21.32 | -5.23 | 1.73E-07 | 0.21 | *HLA-DQB1* ; *HLA-DQA2* | Intergenic |
| cg03638120 | 6 | 32,729,720 | 6p21.32 | -5.70 | 1.19E-08 | 0.39 | *HLA-DQB2* | Intronic |
| cg22421503 | 6 | 36,623,153 | 6p21.2 | 7.31 | 2.68E-13 | 0.69 | *MIR3925* ; *PANDAR* | Intergenic |
| cg13378192 | 6 | 36,625,207 | 6p21.2 | 6.00 | 1.92E-09 | 0.68 | *MIR3925* ; *PANDAR* | Intergenic |
| cg15106294 | 6 | 36,626,044 | 6p21.2 | 6.35 | 2.22E-10 | 0.53 | *MIR3925* ; *PANDAR* | Intergenic |
| cg20731623 | 6 | 36,632,462 | 6p21.2 | 5.53 | 3.17E-08 | 0.53 | *MIR3925* ; *PANDAR* | Intergenic |
| cg02730678 | 6 | 36,641,554 | 6p21.2 | -8.16 | 3.34E-16 | 0.30 | *PANDAR* | Exonic |
| cg11920449 | 6 | 36,645,608 | 6p21.2 | -7.26 | 4.00E-13 | 0.24 | *DINOL* | Exonic |
| cg24425727 | 6 | 36,645,648 | 6p21.2 | -7.00 | 2.54E-12 | 0.53 | *CDKN1A* | Exonic |
| cg15474579 | 6 | 36,645,812 | 6p21.2 | -6.65 | 2.83E-11 | 0.38 | *DINOL* | Exonic |
| cg03714916 | 6 | 36,645,886 | 6p21.2 | -6.65 | 2.87E-11 | 0.24 | *DINOL* | Exonic |
| cg25824330 [c] | 6 | 43,655,393 | 6p21.1 | 5.74 | 9.42E-09 | 0.16 | *MRPS18A* | Intronic |
| cg14471687 | 6 | 55,569,418 | 6p12.1 | 6.11 | 9.71E-10 | 0.22 | *HMGCLL1* ; *BMP5* | Intergenic |
| cg01890197 | 6 | 55,886,930 | 6p12.1 | -7.20 | 5.82E-13 | 0.18 | *BMP5* ; *COL21A1* | Intergenic |
| cg11412288 | 6 | 117,802,725 | 6q22.1 | 5.22 | 1.78E-07 | 0.27 | *ROS1* ; *DCBLD1* | Intergenic |
| cg02575483 | 6 | 117,826,706 | 6q22.1 | -5.21 | 1.88E-07 | 0.26 | *DCBLD1* | Intronic |
| cg06766817 [c] | 6 | 158,515,036 | 6q25.3 | -6.51 | 7.69E-11 | 0.21 | *SYNJ2* | Intronic |
| cg21782315 | 6 | 170,531,847 | 6q27 | -5.45 | 5.02E-08 | 0.20 | *LOC102724511* ; *LOC154449* | Intergenic |
| cg03481035 | 6 | 170,786,302 | 6q27 | -5.28 | 1.28E-07 | 0.21 | *FAM120B* ; *PSMB1* | Intergenic |
| cg20556304 | 6 | 170,819,931 | 6q27 | 12.53 | 5.06E-36 | 0.35 | *FAM120B* ; *PSMB1* | Intergenic |
| cg21143896 [d] | 7 | 2,802,374 | 7p22.2 | 5.44 | 5.33E-08 | 0.55 | *GNA12* | Exonic |
| cg12444411 [d] | 7 | 2,802,554 | 7p22.2 | 5.49 | 3.94E-08 | 0.58 | *GNA12* | 5'UTR |
| cg23285459 [d] | 7 | 2,802,560 | 7p22.2 | 5.35 | 8.57E-08 | 0.45 | *GNA12* | 5'UTR |
| cg00947599 [d] | 7 | 2,802,697 | 7p22.2 | 5.27 | 1.39E-07 | 0.60 | *GNA12* | 5'UTR |
| cg19524238 [d] | 7 | 2,802,976 | 7p22.2 | 5.19 | 2.10E-07 | 0.54 | *AMZ1* ; *GNA12* | Intronic |
| cg23333125 [d] | 7 | 2,803,065 | 7p22.2 | 5.23 | 1.73E-07 | 0.43 | *AMZ1* ; *GNA12* | Intronic |
| cg03185022 [d] | 7 | 2,884,158 | 7p22.2 | -5.26 | 1.43E-07 | 0.35 | *GNA12* | TSS200 |
| cg06152383 [d] | 7 | 2,884,448 | 7p22.2 | -5.24 | 1.60E-07 | 0.33 | *GNA12* | TSS1500 |
| cg12487466 | 7 | 45,001,963 | 7p13 | -6.00 | 1.94E-09 | 0.29 | *MYO1G* | Downstream |

| | | | | | | | | |
|---|---|---|---|---|---|---|---|---|
| cg16459265 | 7 | 45,025,080 | 7p13 | 6.33 | 2.53E-10 | 0.16 | *SNORA9* | Exonic |
| cg26433975 | 7 | 45,066,858 | 7p13 | -5.99 | 2.04E-09 | 0.19 | *CCM2* | Intronic |
| cg18205668 | 7 | 45,066,895 | 7p13 | -5.61 | 2.08E-08 | 0.38 | *CCM2* | Intronic |
| cg23991989 | 7 | 45,067,055 | 7p13 | -5.53 | 3.24E-08 | 0.37 | *CCM2* | Intronic |
| cg06872437 | 7 | 45,107,913 | 7p13 | 5.59 | 2.21E-08 | 0.48 | *CCM2* | Intronic |
| cg12908802 | 7 | 45,129,214 | 7p13 | -5.32 | 1.01E-07 | 0.27 | *NACAD* | TSS1500 |
| cg04785587 | 7 | 45,154,359 | 7p13 | -6.28 | 3.28E-10 | 0.66 | *TBRG4* ; *RAMP3* | Intergenic |
| cg08684900 | 7 | 45,164,170 | 7p13 | 5.99 | 2.14E-09 | 0.24 | *TBRG4* ; *RAMP3* | Intergenic |
| cg23378318 | 7 | 45,178,951 | 7p13 | 5.39 | 6.88E-08 | 0.27 | *TBRG4* ; *RAMP3* | Intergenic |
| cg01610333 | 7 | 45,184,614 | 7p13 | 5.51 | 3.62E-08 | 0.27 | *TBRG4* ; *RAMP3* | Intergenic |
| cg27622369 | 7 | 46,926,447 | 7p12.3 | 5.38 | 7.50E-08 | 0.69 | *LOC730338* ; *TNS3* | Intergenic |
| cg09905635 | 7 | 47,525,243 | 7p12.3 | 6.75 | 1.50E-11 | 0.27 | *TNS3* | Intronic |
| cg12169661 | 7 | 73,183,394 | 7q11.23 | -5.31 | 1.09E-07 | 0.66 | *CLDN3* | 3'UTR |
| cg00686915 | 7 | 99,076,604 | 7q22.1 | -5.53 | 3.24E-08 | 0.23 | *ZNF789* | Intronic |
| cg02004150 | 7 | 99,474,922 | 7q22.1 | 5.91 | 3.38E-09 | 0.37 | *OR2AE1* | TSS1500 |
| cg12773992 | 7 | 99,496,588 | 7q22.1 | -5.68 | 1.34E-08 | 0.18 | *TRIM4* | Intronic |
| cg07902050 | 7 | 99,542,108 | 7q22.1 | 5.37 | 7.78E-08 | 0.19 | *GJC3* ; *AZGP1* | Intergenic |
| cg22792024 | 7 | 99,566,441 | 7q22.1 | 5.45 | 4.91E-08 | 0.78 | *AZGP1* | Intronic |
| cg00641178 | 7 | 99,621,385 | 7q22.1 | -5.30 | 1.15E-07 | 0.30 | *ZKSCAN1* | Exonic |
| cg13621337 | 7 | 99,662,524 | 7q22.1 | 5.32 | 1.03E-07 | 0.18 | *ZNF3* | Exonic |
| cg01758041 | 7 | 99,686,291 | 7q22.1 | 5.44 | 5.31E-08 | 0.26 | *COPS6* | TSS1500 |
| cg18090197 | 7 | 99,769,602 | 7q22.1 | 5.69 | 1.27E-08 | 0.40 | *GPC2* | Intronic |
| cg04095069 [c] | 7 | 151,064,240 | 7q36.1 | -10.05 | 8.80E-24 | 0.16 | *NUB1* | Intronic |
| cg14873699 [c] | 7 | 151,469,323 | 7q36.1 | 8.23 | 1.88E-16 | 0.19 | *PRKAG2* | Intronic |
| cg06362710 | 8 | 59,769,256 | 8q12.1 | 5.74 | 9.58E-09 | 0.28 | *TOX* | Intronic |
| cg06945559 | 8 | 117,564,306 | 8q23.3 | 7.41 | 1.31E-13 | 0.18 | *LINC00536* ; *EIF3H* | Intergenic |
| cg27409870 | 8 | 117,635,401 | 8q23.3 | -16.33 | 6.09E-60 | 0.18 | *LINC00536* ; *EIF3H* | Intergenic |
| cg09559225 | 8 | 117,636,287 | 8q23.3 | -8.07 | 7.14E-16 | 0.35 | *LINC00536* ; *EIF3H* | Intergenic |
| cg24004040 | 8 | 117,650,383 | 8q23.3 | -5.90 | 3.53E-09 | 0.19 | *LINC00536* ; *EIF3H* | Intergenic |
| cg00284516 | 8 | 117,778,669 | 8q24.11 | -6.81 | 9.79E-12 | 0.75 | *UTP23* | TSS200 |
| cg13578020 | 8 | 117,887,218 | 8q24.11 | -6.12 | 9.18E-10 | 0.17 | *RAD21-AS1* | Exonic |
| cg00080252 | 8 | 117,948,764 | 8q24.11 | -5.81 | 6.18E-09 | 0.19 | *RAD21-AS1* ; *AARD* | Intergenic |
| cg20597722 | 8 | 128,421,921 | 8q24.21 | 16.50 | 3.65E-61 | 0.20 | *CASC8* | Intronic |
| cg10520134 | 9 | 22,052,525 | 9p21.3 | 6.37 | 1.87E-10 | 0.20 | *CDKN2B-AS1* | Intronic |
| cg10734259 | 9 | 22,103,886 | 9p21.3 | -8.31 | 9.45E-17 | 0.37 | *CDKN2B-AS1* | Intronic |
| cg14334258 [d] | 9 | 110,595,657 | 9q31.2 | -7.69 | 1.51E-14 | 0.31 | *KLF4* ; *ACTL7B* | Intergenic |
| cg09279362 | 9 | 110,822,615 | 9q31.2 | -6.19 | 5.99E-10 | 0.33 | *KLF4* ; *ACTL7B* | Intergenic |
| cg14472025 [d] | 9 | 136,039,125 | 9q34.2 | 13.15 | 1.59E-39 | 0.19 | *GBGT1* | Intronic |
| cg09513259 [d] | 9 | 136,039,474 | 9q34.2 | -5.73 | 1.00E-08 | 0.20 | *GBGT1* | TSS200 |
| cg14050782 | 9 | 136,932,722 | 9q34.2 | 5.54 | 2.95E-08 | 0.21 | *BRD3* | Intronic |
| cg00303989 | 10 | 8,302,769 | 10p14 | 8.28 | 1.20E-16 | 0.25 | *LINC00708* | Exonic |
| cg05877768 [c] | 10 | 34,427,076 | 10p11.21 | 6.44 | 1.21E-10 | 0.17 | *PARD3* | Intronic |
| cg03817621 | 10 | 52,645,399 | 10q11.23 | 6.55 | 5.69E-11 | 0.35 | *A1CF* | 5'UTR |
| cg24411946 | 10 | 52,645,408 | 10q11.23 | 6.58 | 4.83E-11 | 0.34 | *A1CF* | 5'UTR |
| cg00171421 [c] | 10 | 79,396,127 | 10q22.3 | -7.72 | 1.17E-14 | 0.18 | *KCNMA1* | Exonic |
| cg13198551 | 10 | 80,565,202 | 10q22.3 | 5.70 | 1.22E-08 | 0.15 | *LINC00595* ; *ZMIZ1-AS1* | Intergenic |
| cg04708661 | 10 | 80,862,455 | 10q22.3 | 8.77 | 1.79E-18 | 0.25 | *ZMIZ1* | Intronic |
| cg23346408 | 10 | 81,001,957 | 10q22.3 | 11.91 | 1.09E-32 | 0.55 | *ZMIZ1* | Intronic |
| cg17853216 | 10 | 81,002,480 | 10q22.3 | 12.86 | 7.68E-38 | 0.59 | *ZMIZ1* | Intronic |
| cg17705334 | 10 | 81,002,615 | 10q22.3 | 13.00 | 1.24E-38 | 0.59 | *ZMIZ1* | Intronic |
| cg02283485 | 10 | 81,003,020 | 10q22.3 | 11.51 | 1.22E-30 | 0.18 | *ZMIZ1* | Intronic |
| cg17100355 | 10 | 81,043,875 | 10q22.3 | 5.41 | 6.19E-08 | 0.36 | *ZMIZ1* | Intronic |
| cg04609265 | 10 | 81,045,085 | 10q22.3 | -5.51 | 3.66E-08 | 0.32 | *ZMIZ1* | Intronic |
| cg13574913 | 10 | 81,045,166 | 10q22.3 | -5.31 | 1.12E-07 | 0.17 | *ZMIZ1* | Intronic |
| cg13115165 | 10 | 81,045,939 | 10q22.3 | -5.78 | 7.49E-09 | 0.38 | *ZMIZ1* | Intronic |
| cg00674004 | 10 | 101,280,750 | 10q24.2 | 5.50 | 3.81E-08 | 0.17 | *GOT1* ; *LINC01475* | Intergenic |
| cg19960420 | 10 | 101,282,185 | 10q24.2 | 5.48 | 4.29E-08 | 0.68 | *GOT1* ; *LINC01475* | Intergenic |
| cg01987516 | 10 | 101,282,726 | 10q24.2 | -5.95 | 2.63E-09 | 0.45 | *GOT1* ; *LINC01475* | Intergenic |
| cg23904955 | 10 | 101,282,759 | 10q24.2 | -5.91 | 3.46E-09 | 0.58 | *GOT1* ; *LINC01475* | Intergenic |

| | | | | | | | | |
|---|---|---|---|---|---|---|---|---|
| cg17888390 | 10 | 101,282,816 | 10q24.2 | -5.70 | 1.23E-08 | 0.44 | *GOT1* ; *LINC01475* | Intergenic |
| cg07044859 | 10 | 101,282,883 | 10q24.2 | -5.66 | 1.52E-08 | 0.51 | *GOT1* ; *LINC01475* | Intergenic |
| cg14748795 | 10 | 101,283,090 | 10q24.2 | -5.80 | 6.47E-09 | 0.56 | *GOT1* ; *LINC01475* | Intergenic |
| cg11081894 | 10 | 101,283,847 | 10q24.2 | -5.66 | 1.54E-08 | 0.33 | *GOT1* ; *LINC01475* | Intergenic |
| cg11232245 | 10 | 101,284,535 | 10q24.2 | -6.42 | 1.39E-10 | 0.27 | *GOT1* ; *LINC01475* | Intergenic |
| cg08956474 | 10 | 101,284,950 | 10q24.2 | -5.60 | 2.14E-08 | 0.32 | *GOT1* ; *LINC01475* | Intergenic |
| cg04972745 | 10 | 101,287,846 | 10q24.2 | -5.64 | 1.73E-08 | 0.24 | *LINC01475* | Exonic |
| cg25214158 | 10 | 101,289,600 | 10q24.2 | -6.05 | 1.45E-09 | 0.20 | *LINC01475* | Intronic |
| cg10377903 | 10 | 101,290,699 | 10q24.2 | -7.64 | 2.15E-14 | 0.19 | *LINC01475* | Intronic |
| cg02030034 | 10 | 101,292,403 | 10q24.2 | -5.73 | 9.96E-09 | 0.22 | *NKX2-3* | TSS1500 |
| cg09788492 | 10 | 101,292,477 | 10q24.2 | 5.47 | 4.52E-08 | 0.33 | *NKX2-3* | TSS1500 |
| cg14506196 | 10 | 101,295,400 | 10q24.2 | 6.78 | 1.22E-11 | 0.17 | *NKX2-3* | Exonic |
| cg03711485 | 10 | 101,296,036 | 10q24.2 | 5.25 | 1.56E-07 | 0.33 | *NKX2-3* | 3'UTR |
| cg19129200 | 10 | 101,355,830 | 10q24.2 | -6.99 | 2.75E-12 | 0.29 | *NKX2-3* ; *SLC25A28* | Intergenic |
| cg15561668 | 10 | 101,390,456 | 10q24.2 | -8.05 | 8.27E-16 | 0.63 | *SLC25A28* ; *ENTPD7* | Intergenic |
| cg02917360 | 10 | 101,445,268 | 10q24.2 | 7.96 | 1.69E-15 | 0.21 | *ENTPD7* | Intronic |
| cg17885402 | 10 | 101,492,224 | 10q24.2 | 7.90 | 2.87E-15 | 0.46 | *COX15* | 5'UTR |
| cg04965811 | 10 | 101,492,459 | 10q24.2 | 7.16 | 7.91E-13 | 0.16 | *CUTC* | Intronic |
| cg00573079 | 10 | 114,292,138 | 10q25.2 | 5.38 | 7.58E-08 | 0.21 | *VTI1A* | Intronic |
| cg18883674 | 10 | 114,729,144 | 10q25.2 | -6.75 | 1.48E-11 | 0.18 | *TCF7L2* | Intronic |
| cg12454663 | 10 | 114,805,006 | 10q25.2 | -8.66 | 4.66E-18 | 0.16 | *TCF7L2* | Intronic |
| cg12338214 [c] | 10 | 133,783,113 | 10q26.3 | -5.73 | 1.01E-08 | 0.32 | *BNIP3* | Intronic |
| cg16984897 [c] | 11 | 3,875,715 | 11p15.4 | -5.39 | 7.01E-08 | 0.27 | *RHOG* ; *STIM1* | Intergenic |
| cg22319263 [c] | 11 | 3,875,774 | 11p15.4 | -5.32 | 1.01E-07 | 0.22 | *RHOG* ; *STIM1* | Intergenic |
| cg15489575 [c] | 11 | 3,875,823 | 11p15.4 | -5.24 | 1.61E-07 | 0.28 | *RHOG* ; *STIM1* | Intergenic |
| cg07438103 [c] | 11 | 47,175,143 | 11p11.2 | -5.19 | 2.07E-07 | 0.27 | *C11orf49* | Intronic |
| cg23917844 | 11 | 61,314,901 | 11q12.2 | 6.05 | 1.46E-09 | 0.19 | *SYT7* | Intronic |
| cg05094984 | 11 | 61,544,890 | 11q12.2 | 6.93 | 4.17E-12 | 0.20 | *MYRF* | Exonic |
| cg07689907 | 11 | 61,582,574 | 11q12.2 | -9.44 | 3.85E-21 | 0.25 | *FADS1* | Intronic |
| cg14725641 | 11 | 61,582,763 | 11q12.2 | 8.30 | 1.08E-16 | 0.33 | *FADS1* | Intronic |
| cg08093537 | 11 | 61,595,225 | 11q12.2 | 6.09 | 1.15E-09 | 0.53 | *FADS2* | Intronic |
| cg01400685 | 11 | 61,598,025 | 11q12.2 | 6.11 | 9.77E-10 | 0.25 | *FADS2* | Intronic |
| cg22796604 | 11 | 61,602,232 | 11q12.2 | 6.70 | 2.03E-11 | 0.22 | *FADS2* | Intronic |
| cg00264176 | 11 | 61,612,775 | 11q12.2 | -8.31 | 9.95E-17 | 0.47 | *FADS2* | Intronic |
| cg02297460 | 11 | 69,920,437 | 11q13.3 | 5.45 | 5.00E-08 | 0.20 | *LINC02584* | Intronic |
| cg24846725 | 11 | 69,921,715 | 11q13.3 | 7.15 | 8.68E-13 | 0.26 | *LINC02584* | TSS1500 |
| cg26705266 | 11 | 69,924,339 | 11q13.3 | 5.28 | 1.30E-07 | 0.36 | *ANO1* | TSS200 |
| cg15682194 | 11 | 69,925,136 | 11q13.3 | 5.31 | 1.11E-07 | 0.40 | *ANO1* | Intronic |
| cg25018928 | 11 | 69,943,421 | 11q13.3 | 7.04 | 1.92E-12 | 0.52 | *ANO1* | Intronic |
| cg18944952 | 11 | 74,108,494 | 11q13.4 | 7.19 | 6.27E-13 | 0.16 | *PGM2L1* | Intronic |
| cg07431994 | 11 | 74,218,301 | 11q13.4 | -10.51 | 7.58E-26 | 0.31 | *LOC100287896* ; *POLD3* | Intergenic |
| cg22403983 | 11 | 74,295,300 | 11q13.4 | 7.55 | 4.26E-14 | 0.22 | *LOC100287896* ; *POLD3* | Intergenic |
| cg11913491 | 11 | 74,303,901 | 11q13.4 | 6.46 | 1.08E-10 | 0.34 | *POLD3* | 5'UTR |
| cg12406507 | 11 | 74,348,674 | 11q13.4 | -9.82 | 9.43E-23 | 0.21 | *POLD3* | Intronic |
| cg14195722 | 11 | 74,413,754 | 11q13.4 | -8.10 | 5.69E-16 | 0.41 | *CHRDL2* | Intronic |
| cg11102819 | 11 | 74,421,983 | 11q13.4 | -5.60 | 2.09E-08 | 0.38 | *CHRDL2* | Exonic |
| cg00376252 | 11 | 74,427,166 | 11q13.4 | -8.31 | 9.92E-17 | 0.34 | *CHRDL2* | Intronic |
| cg16180424 | 11 | 74,428,603 | 11q13.4 | -5.41 | 6.43E-08 | 0.48 | *CHRDL2* | Intronic |
| cg08536044 | 11 | 74,431,442 | 11q13.4 | -8.63 | 5.90E-18 | 0.40 | *CHRDL2* | Intronic |
| cg12233692 | 11 | 74,431,850 | 11q13.4 | -7.67 | 1.68E-14 | 0.43 | *CHRDL2* | Intronic |
| cg08700454 | 11 | 74,432,028 | 11q13.4 | -7.00 | 2.62E-12 | 0.33 | *CHRDL2* | Intronic |
| cg16209924 | 11 | 74,437,016 | 11q13.4 | -6.33 | 2.39E-10 | 0.24 | *CHRDL2* | Intronic |
| cg22902532 | 11 | 74,440,516 | 11q13.4 | -6.17 | 6.90E-10 | 0.21 | *CHRDL2* | Intronic |
| cg22255888 | 11 | 110,754,112 | 11q23.1 | -5.33 | 9.78E-08 | 0.32 | *ARHGAP20* ; *LINC02550* | Intergenic |
| cg05820396 | 11 | 111,055,240 | 11q23.1 | -12.36 | 4.47E-35 | 0.30 | *LINC02550* ; *C11orf53* | Intergenic |
| cg14675241 | 11 | 111,154,375 | 11q23.1 | 7.44 | 1.00E-13 | 0.20 | *C11orf53* | Intronic |
| cg03078767 | 11 | 111,154,742 | 11q23.1 | 15.61 | 6.63E-55 | 0.47 | *C11orf53* | Intronic |
| cg23091777 | 11 | 111,154,754 | 11q23.1 | 15.24 | 1.85E-52 | 0.41 | *C11orf53* | Intronic |
| cg21127079 | 11 | 111,154,826 | 11q23.1 | 14.15 | 1.85E-45 | 0.39 | *C11orf53* | Exonic |

| | | | | | | | | |
|---|---|---|---|---|---|---|---|---|
| cg24861920 | 11 | 111,169,126 | 11q23.1 | 15.22 | 2.71E-52 | 0.28 | *COLCA1* | 5'UTR |
| cg20221957 | 11 | 111,169,182 | 11q23.1 | 15.55 | 1.50E-54 | 0.54 | *COLCA1* | 5'UTR |
| cg21250150 | 11 | 111,169,251 | 11q23.1 | 14.44 | 2.79E-47 | 0.31 | *COLCA1* | 5'UTR |
| cg20465794 | 11 | 111,169,943 | 11q23.1 | 15.18 | 4.84E-52 | 0.22 | *COLCA1* | 5'UTR |
| cg18547942 | 11 | 111,170,390 | 11q23.1 | 12.70 | 6.20E-37 | 0.21 | *COLCA1* ; *COLCA2* | 5'UTR |
| cg13202534 | 11 | 111,170,911 | 11q23.1 | 14.71 | 5.76E-49 | 0.37 | *COLCA1* | 5'UTR |
| cg09538401 | 11 | 111,171,330 | 11q23.1 | 9.62 | 6.83E-22 | 0.40 | *COLCA2* | Exonic |
| cg13833445 | 11 | 111,173,264 | 11q23.1 | 5.60 | 2.12E-08 | 0.26 | *COLCA1* ; *COLCA2* | Intronic |
| cg08329092 | 11 | 111,203,997 | 11q23.1 | -6.57 | 4.87E-11 | 0.37 | *COLCA2* ; *MIR4491* | Intergenic |
| cg19126910 | 11 | 111,249,659 | 11q23.1 | 5.80 | 6.80E-09 | 0.59 | *POU2AF1* | Intronic |
| cg12832280 | 11 | 111,307,431 | 11q23.1 | 6.52 | 7.02E-11 | 0.51 | *LOC100132078* ; *BTG4* | Intergenic |
| cg24290496 | 11 | 111,313,817 | 11q23.1 | -12.58 | 2.65E-36 | 0.21 | *LOC100132078* ; *BTG4* | Intergenic |
| cg15943751 | 11 | 111,319,463 | 11q23.1 | -5.45 | 5.00E-08 | 0.45 | *LOC100132078* ; *BTG4* | Intergenic |
| cg02532096 | 11 | 111,411,289 | 11q23.1 | 7.40 | 1.40E-13 | 0.17 | *LAYN* | 5'UTR |
| cg07896967 | 11 | 111,502,580 | 11q23.1 | -5.40 | 6.64E-08 | 0.18 | *SIK2* | Intronic |
| cg17762753 | 12 | 4,374,220 | 12p13.32 | -6.18 | 6.23E-10 | 0.20 | *CCND2-AS1* | Intronic |
| cg25519457 | 12 | 4,387,827 | 12p13.32 | -5.71 | 1.12E-08 | 0.26 | *CCND2* | Intronic |
| cg12594237 | 12 | 4,399,955 | 12p13.32 | -6.21 | 5.42E-10 | 0.41 | *CCND2* | Intronic |
| cg00417288 | 12 | 4,399,998 | 12p13.32 | -9.43 | 4.01E-21 | 0.36 | *CCND2* | Intronic |
| cg15247141 | 12 | 6,387,977 | 12p13.31 | 7.81 | 5.77E-15 | 0.17 | *CD9* ; *PLEKHG6* | Intergenic |
| cg25273936 | 12 | 6,421,474 | 12p13.31 | 6.28 | 3.33E-10 | 0.65 | *PLEKHG6* | Exonic |
| cg03560395 | 12 | 6,427,068 | 12p13.31 | 6.57 | 4.91E-11 | 0.70 | *PLEKHG6* | Exonic |
| cg05863443 | 12 | 50,419,463 | 12q13.12 | -5.70 | 1.19E-08 | 0.17 | *RACGAP1* | TSS200 |
| cg08189863 | 12 | 50,468,624 | 12q13.12 | 5.20 | 1.99E-07 | 0.57 | *ASIC1* | Intronic |
| cg01043616 | 12 | 50,475,035 | 12q13.12 | -5.54 | 3.02E-08 | 0.33 | *ASIC1* | Intronic |
| cg16615454 | 12 | 50,475,167 | 12q13.12 | -6.12 | 9.49E-10 | 0.21 | *ASIC1* | Intronic |
| cg02553054 | 12 | 50,478,480 | 12q13.12 | 6.56 | 5.23E-11 | 0.28 | *SMARCD1* | TSS1500 |
| cg27207430 | 12 | 50,478,730 | 12q13.12 | -5.71 | 1.12E-08 | 0.18 | *SMARCD1* | TSS1500 |
| cg25437271 | 12 | 50,494,820 | 12q13.12 | -7.13 | 1.03E-12 | 0.17 | *SMARCD1* | Downstream |
| cg15405652 | 12 | 50,529,977 | 12q13.12 | -7.23 | 4.67E-13 | 0.75 | *CERS5* | Intronic |
| cg21422335 | 12 | 50,612,068 | 12q13.12 | 7.19 | 6.35E-13 | 0.24 | *LIMA1* | Intronic |
| cg24245830 | 12 | 50,616,100 | 12q13.12 | 9.58 | 9.37E-22 | 0.43 | *LIMA1* | Exonic |
| cg08247202 | 12 | 50,616,162 | 12q13.12 | 6.87 | 6.45E-12 | 0.46 | *LIMA1* | Exonic |
| cg11635421 | 12 | 50,616,262 | 12q13.12 | 9.42 | 4.40E-21 | 0.44 | *LIMA1* | Exonic |
| cg01577165 | 12 | 50,616,434 | 12q13.12 | 10.01 | 1.36E-23 | 0.40 | *LIMA1* | 5'UTR |
| cg23497020 | 12 | 50,616,562 | 12q13.12 | 9.78 | 1.38E-22 | 0.25 | *LIMA1* | Intronic |
| cg12892471 | 12 | 50,616,779 | 12q13.12 | 8.87 | 7.43E-19 | 0.44 | *LIMA1* | Intronic |
| cg15819890 | 12 | 50,678,329 | 12q13.12 | -5.98 | 2.22E-09 | 0.51 | *LIMA1* ; *FAM186A* | Intergenic |
| cg26865274 | 12 | 50,697,206 | 12q13.12 | 5.63 | 1.76E-08 | 0.22 | *LIMA1* ; *FAM186A* | Intergenic |
| cg14249095 | 12 | 50,745,607 | 12q13.12 | -8.52 | 1.56E-17 | 0.19 | *FAM186A* | Exonic |
| cg26960637 | 12 | 50,784,193 | 12q13.12 | -7.66 | 1.87E-14 | 0.26 | *FAM186A* | Intronic |
| cg27563558 | 12 | 50,895,943 | 12q13.12 | 6.51 | 7.71E-11 | 0.32 | *LARP4* ; *DIP2B* | Intergenic |
| cg03907575 | 12 | 50,898,411 | 12q13.12 | -9.02 | 1.82E-19 | 0.53 | *DIP2B* | TSS1500 |
| cg20014596 | 12 | 50,898,483 | 12q13.12 | -7.96 | 1.78E-15 | 0.43 | *DIP2B* | TSS1500 |
| cg00665405 | 12 | 50,899,548 | 12q13.12 | 8.37 | 5.97E-17 | 0.18 | *DIP2B* | Intronic |
| cg18504661 | 12 | 50,931,745 | 12q13.12 | 6.54 | 6.36E-11 | 0.56 | *DIP2B* | Intronic |
| cg11534400 | 12 | 50,943,912 | 12q13.12 | -6.37 | 1.93E-10 | 0.31 | *DIP2B* | Intronic |
| cg01485708 | 12 | 50,968,242 | 12q13.12 | -7.33 | 2.25E-13 | 0.27 | *DIP2B* | Intronic |
| cg04217475 | 12 | 51,007,498 | 12q13.12 | 7.96 | 1.72E-15 | 0.24 | *DIP2B* | Intronic |
| cg14953804 | 12 | 51,021,072 | 12q13.12 | -5.48 | 4.24E-08 | 0.18 | *DIP2B* | Intronic |
| cg11471172 | 12 | 51,128,843 | 12q13.12 | 8.10 | 5.53E-16 | 0.26 | *DIP2B* | Intronic |
| cg15168922 | 12 | 51,128,872 | 12q13.12 | 8.04 | 8.87E-16 | 0.25 | *DIP2B* | Exonic |
| cg23040489 | 12 | 51,146,086 | 12q13.12 | -10.14 | 3.73E-24 | 0.19 | *DIP2B* ; *ATF1* | Intergenic |
| cg12399160 | 12 | 51,179,369 | 12q13.12 | -5.45 | 5.11E-08 | 0.16 | *ATF1* | Intronic |
| cg08740063 | 12 | 51,180,192 | 12q13.12 | -7.28 | 3.27E-13 | 0.39 | *ATF1* | Intronic |
| cg14405934 | 12 | 51,316,220 | 12q13.12 | 8.43 | 3.51E-17 | 0.24 | *TMPRSS12* ; *METTL7A* | Intergenic |
| cg02101662 | 12 | 51,316,730 | 12q13.12 | 6.36 | 1.96E-10 | 0.16 | *TMPRSS12* ; *METTL7A* | Intergenic |
| cg04477962 | 12 | 51,317,375 | 12q13.12 | 7.91 | 2.55E-15 | 0.34 | *TMPRSS12* ; *METTL7A* | Intergenic |
| cg10487714 | 12 | 51,322,158 | 12q13.12 | 6.32 | 2.68E-10 | 0.26 | *METTL7A* | Intronic |

| | | | | | | | | |
|---|---|---|---|---|---|---|---|---|
| cg22366350 | 12 | 57,082,604 | 12q13.3 | 5.75 | 8.95E-09 | 0.27 | *PTGES3* | TSS1500 |
| cg16632593 | 12 | 57,444,752 | 12q13.3 | 5.54 | 2.94E-08 | 0.17 | *MYO1A* | 5'UTR |
| cg07134316 | 12 | 57,488,112 | 12q13.3 | 6.19 | 5.94E-10 | 0.41 | *NAB2* | Intronic |
| cg07926491 | 12 | 57,489,971 | 12q13.3 | -7.33 | 2.38E-13 | 0.39 | *STAT6* | 3'UTR |
| cg16896579 | 12 | 57,496,251 | 12q13.3 | -5.55 | 2.84E-08 | 0.32 | *STAT6* | Exonic |
| cg13685273 | 12 | 57,512,715 | 12q13.3 | 5.71 | 1.16E-08 | 0.40 | *STAT6* ; *LRP1* | Intergenic |
| cg03894242 | 12 | 57,513,121 | 12q13.3 | 6.06 | 1.32E-09 | 0.38 | *STAT6* ; *LRP1* | Intergenic |
| cg10233454 | 12 | 57,529,389 | 12q13.3 | 7.12 | 1.06E-12 | 0.38 | *LRP1* | Intronic |
| cg01883322 | 12 | 64,233,376 | 12q14.2 | -5.17 | 2.33E-07 | 0.16 | *MIR10527* ; *SRGAP1* | Intergenic |
| cg11362183 | 12 | 64,403,751 | 12q14.2 | -5.34 | 9.32E-08 | 0.30 | *SRGAP1* | Intronic |
| cg14731391 | 12 | 71,523,979 | 12q21.1 | -5.34 | 9.18E-08 | 0.16 | *TSPAN8* | Intronic |
| cg25115154 | 12 | 71,533,408 | 12q21.1 | 5.64 | 1.70E-08 | 0.30 | *TSPAN8* | Intronic |
| cg19045468 | 12 | 71,533,592 | 12q21.1 | 6.27 | 3.57E-10 | 0.40 | *TSPAN8* | Exonic |
| cg19013339 | 12 | 96,052,514 | 12q22 | 7.83 | 4.70E-15 | 0.30 | *PGAM1P5* | Intronic |
| cg09763086 | 12 | 111,358,601 | 12q24.11 | 6.94 | 3.87E-12 | 0.16 | *MYL2* | TSS200 |
| cg07060816 | 12 | 111,395,124 | 12q24.11 | 6.56 | 5.25E-11 | 0.25 | *LOC105369980* | Intronic |
| cg20940052 | 12 | 111,396,409 | 12q24.11 | 7.38 | 1.57E-13 | 0.58 | *LOC105369980* | TSS1500 |
| cg19351627 | 12 | 111,396,962 | 12q24.11 | 7.53 | 5.12E-14 | 0.32 | *LOC105369980* ; *CUX2* | Intergenic |
| cg19702880 | 12 | 111,403,583 | 12q24.11 | 5.69 | 1.24E-08 | 0.28 | *LOC105369980* ; *CUX2* | Intergenic |
| cg18964988 | 12 | 111,470,767 | 12q24.11 | 6.65 | 2.93E-11 | 0.15 | *LOC105369980* ; *CUX2* | Intergenic |
| cg16235197 | 12 | 111,758,873 | 12q24.12 | 5.30 | 1.13E-07 | 0.17 | *CUX2* | Intronic |
| cg08894980 | 12 | 111,768,251 | 12q24.12 | -7.06 | 1.65E-12 | 0.38 | *CUX2* | Intronic |
| cg03354235 | 12 | 111,832,504 | 12q24.12 | -9.19 | 4.06E-20 | 0.19 | *PHETA1* ; *SH2B3* | Intergenic |
| cg20759222 | 12 | 111,834,955 | 12q24.12 | -5.17 | 2.36E-07 | 0.17 | *PHETA1* ; *SH2B3* | Intergenic |
| cg07932199 | 12 | 112,008,034 | 12q24.12 | 5.26 | 1.45E-07 | 0.26 | *ATXN2* | Intronic |
| cg00418629 | 12 | 112,062,661 | 12q24.12 | -5.55 | 2.79E-08 | 0.17 | *ATXN2-AS* ; *BRAP* | Intergenic |
| cg09553684 | 12 | 112,191,404 | 12q24.12 | 9.60 | 8.30E-22 | 0.27 | *ACAD10* | Intronic |
| cg19186356 | 12 | 112,204,327 | 12q24.12 | -6.87 | 6.45E-12 | 0.26 | *ALDH2* | TSS1500 |
| cg20637653 | 12 | 112,450,938 | 12q24.13 | 5.80 | 6.54E-09 | 0.19 | *TMEM116* | 5'UTR |
| cg10101109 | 12 | 112,455,255 | 12q24.13 | 7.88 | 3.32E-15 | 0.20 | *ERP29* | Intronic |
| cg12171396 | 12 | 112,563,139 | 12q24.13 | 5.30 | 1.16E-07 | 0.15 | *TRAFD1* | TSS1500 |
| cg12198432 | 12 | 115,104,183 | 12q24.21 | 6.72 | 1.83E-11 | 0.37 | *TBX5-AS1* ; *TBX3* | Intergenic |
| cg16406892 | 12 | 115,118,777 | 12q24.21 | -6.22 | 4.85E-10 | 0.43 | *TBX3* | Exonic |
| cg13829104 | 12 | 115,118,806 | 12q24.21 | -5.85 | 4.90E-09 | 0.27 | *TBX3* | Exonic |
| cg02694170 | 12 | 115,888,757 | 12q24.21 | -8.51 | 1.75E-17 | 0.41 | *TBX3* ; *MED13L* | Intergenic |
| cg24172291 | 12 | 115,889,509 | 12q24.21 | -7.21 | 5.70E-13 | 0.30 | *TBX3* ; *MED13L* | Intergenic |
| cg00387115 | 12 | 115,889,598 | 12q24.21 | -8.44 | 3.22E-17 | 0.47 | *TBX3* ; *MED13L* | Intergenic |
| cg09605392 | 12 | 115,906,012 | 12q24.21 | 8.54 | 1.38E-17 | 0.31 | *TBX3* ; *MED13L* | Intergenic |
| cg04020319 [c] | 12 | 121,445,882 | 12q24.31 | 5.94 | 2.84E-09 | 0.54 | *C12orf43* | Intronic |
| cg05190884 [c] | 12 | 124,528,472 | 12q24.31 | -6.50 | 7.95E-11 | 0.43 | *ZNF664-RFLNA* | Intronic |
| cg24776343 [c] | 13 | 24,189,786 | 13q12.12 | 8.22 | 2.02E-16 | 0.19 | *TNFRSF19* | Intronic |
| cg22259344 | 13 | 34,068,089 | 13q13.2 | 6.31 | 2.74E-10 | 0.34 | *STARD13* | Intronic |
| cg08051523 | 13 | 34,083,938 | 13q13.2 | 5.25 | 1.53E-07 | 0.47 | *STARD13* | Intronic |
| cg02985381 | 13 | 37,494,744 | 13q13.3 | 6.12 | 9.57E-10 | 0.47 | *SMAD9* | TSS1500 |
| cg01034396 | 13 | 37,494,802 | 13q13.3 | 8.37 | 5.66E-17 | 0.66 | *SMAD9* | TSS1500 |
| cg08643012 | 13 | 37,494,946 | 13q13.3 | 7.86 | 3.77E-15 | 0.69 | *SMAD9* | TSS1500 |
| cg03940408 | 13 | 37,494,994 | 13q13.3 | 8.70 | 3.27E-18 | 0.46 | *SMAD9* | TSS1500 |
| cg25917725 | 13 | 37,495,343 | 13q13.3 | 8.04 | 8.70E-16 | 0.47 | *SMAD9* | TSS1500 |
| cg27598979 | 13 | 37,495,632 | 13q13.3 | 7.87 | 3.61E-15 | 0.51 | *SMAD9* ; *ALG5* | Intergenic |
| cg01493522 | 13 | 37,497,338 | 13q13.3 | -8.22 | 2.12E-16 | 0.39 | *SMAD9* ; *ALG5* | Intergenic |
| cg24467989 [c] | 13 | 41,560,747 | 13q14.11 | -5.27 | 1.35E-07 | 0.18 | *ELF1* | Intronic |
| cg20314763 | 13 | 73,628,948 | 13q22.1 | -5.35 | 8.86E-08 | 0.31 | *KLF5* | TSS200 |
| cg09667226 | 13 | 73,632,762 | 13q22.1 | 6.79 | 1.12E-11 | 0.17 | *KLF5* | Intronic |
| cg07749664 | 13 | 73,634,670 | 13q22.1 | -5.50 | 3.80E-08 | 0.20 | *KLF5* | Intronic |
| cg22065124 | 13 | 73,649,955 | 13q22.1 | -7.35 | 1.94E-13 | 0.34 | *KLF5* | Exonic |
| cg11664362 | 13 | 73,675,471 | 13q22.1 | -6.39 | 1.68E-10 | 0.38 | *KLF5* ; *LINC00392* | Intergenic |
| cg08556772 | 13 | 111,077,142 | 13q34 | 6.30 | 3.03E-10 | 0.43 | *COL4A2* | Exonic |
| cg04300287 | 13 | 111,077,156 | 13q34 | 5.82 | 6.06E-09 | 0.44 | *COL4A2* | Exonic |
| cg13153710 | 13 | 111,077,257 | 13q34 | 5.35 | 8.83E-08 | 0.21 | *COL4A2* | Intronic |

| | | | | | | | | |
|---|---|---|---|---|---|---|---|---|
| cg16284305 | 14 | 50,698,462 | 14q21.3 | 5.69 | 1.31E-08 | 0.15 | *SOS2* | TSS1500 |
| cg13046688 | 14 | 51,340,344 | 14q22.1 | -5.35 | 8.81E-08 | 0.31 | *ABHD12B* | Intronic |
| cg21052932 | 14 | 51,342,320 | 14q22.1 | -5.28 | 1.28E-07 | 0.40 | *ABHD12B* | Intronic |
| cg16720578 | 14 | 54,410,717 | 14q22.2 | 6.04 | 1.54E-09 | 0.34 | *LOC101927620* ; *MIR5580* | Intergenic |
| cg01964170 | 14 | 54,414,764 | 14q22.2 | 5.50 | 3.88E-08 | 0.18 | *MIR5580* | Downstream |
| cg05928290 | 14 | 54,418,728 | 14q22.2 | -9.56 | 1.15E-21 | 0.26 | *BMP4* | Exonic |
| cg05923197 | 14 | 54,418,804 | 14q22.2 | -9.05 | 1.42E-19 | 0.19 | *BMP4* | Exonic |
| cg09367901 | 14 | 54,418,851 | 14q22.2 | -8.69 | 3.58E-18 | 0.19 | *BMP4* | Exonic |
| cg07685795 | 14 | 54,418,880 | 14q22.2 | -9.68 | 3.61E-22 | 0.19 | *BMP4* | Exonic |
| cg16190896 | 14 | 54,423,436 | 14q22.2 | 5.81 | 6.10E-09 | 0.18 | *BMP4* | 5'UTR |
| cg12128696 | 14 | 54,429,756 | 14q22.2 | 5.46 | 4.80E-08 | 0.31 | *BMP4* ; *CDKN3* | Intergenic |
| cg18844229 | 14 | 54,434,563 | 14q22.2 | -6.07 | 1.28E-09 | 0.35 | *BMP4* ; *CDKN3* | Intergenic |
| cg13259103 [c] | 14 | 88,456,132 | 14q31.3 | -7.23 | 4.76E-13 | 0.18 | *GALC* | Intronic |
| cg05737860 | 15 | 32,969,475 | 15q13.3 | -8.68 | 3.93E-18 | 0.29 | *ARHGAP11A-SCG5* ; *SCG5* | Intronic |
| cg01722420 | 15 | 32,993,763 | 15q13.3 | 5.22 | 1.76E-07 | 0.34 | *ARHGAP11A-SCG5* ; *GREM1* | Intergenic |
| cg04870120 | 15 | 32,999,748 | 15q13.3 | 12.76 | 2.77E-37 | 0.32 | *ARHGAP11A-SCG5* ; *GREM1* | Intergenic |
| cg07578199 | 15 | 33,001,552 | 15q13.3 | -13.24 | 5.32E-40 | 0.75 | *ARHGAP11A-SCG5* ; *GREM1* | Intergenic |
| cg09442789 | 15 | 33,009,115 | 15q13.3 | 9.86 | 6.30E-23 | 0.25 | *ARHGAP11A-SCG5* ; *GREM1* | Intergenic |
| cg09656020 | 15 | 33,009,409 | 15q13.3 | 9.94 | 2.75E-23 | 0.21 | *GREM1* | TSS1500 |
| cg11193064 | 15 | 67,008,444 | 15q22.31 | -6.60 | 4.08E-11 | 0.76 | *SMAD6* | Intronic |
| cg19684296 | 15 | 67,008,514 | 15q22.31 | -5.38 | 7.35E-08 | 0.43 | *SMAD6* | Intronic |
| cg06846320 | 15 | 67,401,751 | 15q22.33 | -5.58 | 2.36E-08 | 0.21 | *SMAD3* | Intronic |
| cg24579218 | 15 | 68,104,479 | 15q23 | 5.55 | 2.78E-08 | 0.64 | *MAP2K5* ; *SKOR1* | Intergenic |
| cg27178290 | 15 | 68,110,793 | 15q23 | -5.75 | 9.08E-09 | 0.61 | *MAP2K5* ; *SKOR1* | Intergenic |
| cg02764055 | 15 | 68,110,824 | 15q23 | -6.11 | 9.98E-10 | 0.39 | *MAP2K5* ; *SKOR1* | Intergenic |
| cg04618848 | 15 | 68,111,941 | 15q23 | -5.22 | 1.76E-07 | 0.16 | *MAP2K5* ; *SKOR1* | Intergenic |
| cg09469610 | 15 | 68,113,110 | 15q23 | -6.19 | 5.95E-10 | 0.19 | *MAP2K5* ; *SKOR1* | Intergenic |
| cg18916403 | 15 | 68,115,877 | 15q23 | -5.23 | 1.66E-07 | 0.24 | *MAP2K5* ; *SKOR1* | Intergenic |
| cg07010088 | 15 | 68,116,505 | 15q23 | -5.59 | 2.29E-08 | 0.22 | *MAP2K5* ; *SKOR1* | Intergenic |
| cg23074691 | 15 | 72,183,545 | 15q23 | 5.40 | 6.69E-08 | 0.17 | *MYO9A* | Intronic |
| cg21214455 | 15 | 72,412,075 | 15q23 | -5.70 | 1.23E-08 | 0.51 | *SENP8* | Intronic |
| cg16672083 | 15 | 72,433,130 | 15q23 | -5.62 | 1.94E-08 | 0.69 | *SENP8* | 3'UTR |
| cg25472872 | 15 | 72,491,290 | 15q23 | 5.70 | 1.20E-08 | 0.34 | *GRAMD2* | TSS1500 |
| cg20070323 | 15 | 72,520,350 | 15q23 | -5.68 | 1.35E-08 | 0.21 | *PKM* | Intronic |
| cg23314488 | 15 | 72,522,205 | 15q23 | -5.61 | 1.97E-08 | 0.27 | *PKM* | 5'UTR |
| cg12670525 | 15 | 72,563,412 | 15q23 | 5.82 | 5.72E-09 | 0.19 | *PARP6* | 5'UTR |
| cg10040866 | 15 | 72,690,640 | 15q23 | -5.73 | 9.96E-09 | 0.21 | *TMEM202* | TSS200 |
| cg24797896 [c] | 15 | 75,496,647 | 15q24.2 | -6.06 | 1.36E-09 | 0.34 | *C15orf39* | Intronic |
| cg22293481 | 15 | 90,745,248 | 15q26.1 | 6.55 | 5.92E-11 | 0.18 | *SEMA4B* | Intronic |
| cg00425431 | 15 | 90,792,223 | 15q26.1 | 5.75 | 9.10E-09 | 0.73 | *TTLL13P* | TSS1500 |
| cg20027684 | 15 | 90,792,300 | 15q26.1 | 5.75 | 8.95E-09 | 0.24 | *TTLL13P* | TSS1500 |
| cg12934461 | 15 | 90,792,652 | 15q26.1 | 5.66 | 1.52E-08 | 0.46 | *TTLL13P* | TSS1500 |
| cg25423822 | 15 | 90,792,717 | 15q26.1 | 5.79 | 7.01E-09 | 0.43 | *TTLL13P* | TSS1500 |
| cg05239308 | 15 | 90,793,056 | 15q26.1 | 5.38 | 7.33E-08 | 0.64 | *TTLL13P* | Intronic |
| cg03959527 | 15 | 90,894,952 | 15q26.1 | 5.77 | 7.77E-09 | 0.16 | *ZNF774* | TSS1500 |
| cg09027889 | 15 | 91,039,456 | 15q26.1 | -5.34 | 9.44E-08 | 0.38 | *IQGAP1* | TSS1500 |
| cg15046585 | 15 | 91,072,757 | 15q26.1 | 5.22 | 1.78E-07 | 0.48 | *CRTC3* | TSS1500 |
| cg04099705 [c] | 16 | 1,397,981 | 16p13.3 | -7.53 | 5.06E-14 | 0.16 | *BAIAP3* | Exonic |
| cg27634195 [c] | 16 | 1,440,421 | 16p13.3 | -8.60 | 7.82E-18 | 0.26 | *UNKL* | Intronic |
| cg09232225 | 16 | 67,022,472 | 16q22.1 | -5.24 | 1.63E-07 | 0.42 | *CES4A* | TSS200 |
| cg27235071 | 16 | 68,560,373 | 16q22.1 | 5.19 | 2.12E-07 | 0.38 | *SMPD3* ; *ZFP90* | TSS200 |
| cg03487276 | 16 | 68,563,106 | 16q22.1 | 5.42 | 6.04E-08 | 0.46 | *SMPD3* ; *ZFP90* | Intergenic |
| cg13922712 | 16 | 68,721,436 | 16q22.1 | -5.26 | 1.47E-07 | 0.70 | *CDH3* | Exonic |
| cg01231543 | 16 | 68,741,748 | 16q22.1 | -5.50 | 3.91E-08 | 0.30 | *CDH3* ; *CDH1* | Intergenic |
| cg24766462 | 16 | 68,749,583 | 16q22.1 | -6.24 | 4.43E-10 | 0.40 | *CDH3* ; *CDH1* | Intergenic |
| cg10133464 | 16 | 68,758,578 | 16q22.1 | 5.45 | 4.98E-08 | 0.29 | *CDH3* ; *CDH1* | Intergenic |
| cg27512737 | 16 | 68,804,435 | 16q22.1 | 5.99 | 2.14E-09 | 0.28 | *CDH1* | Intronic |
| cg20543683 | 16 | 68,812,420 | 16q22.1 | -5.79 | 6.87E-09 | 0.16 | *CDH1* | Intronic |
| cg08696401 | 16 | 68,818,222 | 16q22.1 | 6.66 | 2.66E-11 | 0.44 | *CDH1* | Intronic |

| | | | | | | | | |
|---|---|---|---|---|---|---|---|---|
| cg14240326 | 16 | 68,877,911 | 16q22.1 | 6.91 | 4.89E-12 | 0.18 | *TANGO6* | Intronic |
| cg10689889 | 16 | 79,634,732 | 16q23.2 | 6.37 | 1.88E-10 | 0.15 | *MAF* | TSS1500 |
| cg02232320 | 16 | 79,804,683 | 16q23.2 | 6.52 | 7.21E-11 | 0.18 | *LINC01229* | Intronic |
| cg21136508 | 16 | 80,058,605 | 16q23.2 | 5.46 | 4.69E-08 | 0.16 | *LINC01229* ; *LOC102724084* | Intergenic |
| cg24591575 | 16 | 80,094,284 | 16q23.2 | -6.54 | 6.32E-11 | 0.49 | *LINC01229* ; *LOC102724084* | Intergenic |
| cg04290162 [c] | 16 | 82,204,401 | 16q23.3 | -11.92 | 9.26E-33 | 0.16 | *MPHOSPH6* | TSS1500 |
| cg03105604 [c] | 16 | 82,675,170 | 16q23.3 | 5.40 | 6.64E-08 | 0.23 | *CDH13* | TSS1500 |
| cg18238236 | 16 | 86,320,980 | 16q24.1 | 5.53 | 3.29E-08 | 0.22 | *LINC02135* | Intronic |
| cg05831517 | 16 | 86,337,750 | 16q24.1 | -6.11 | 9.86E-10 | 0.16 | *LINC02135* ; *LINC00917* | Intergenic |
| cg04996089 [c] | 16 | 88,937,216 | 16q24.3 | -5.78 | 7.68E-09 | 0.39 | *PABPN1L* ; *CBFA2T3* | Intergenic |
| cg07716131 [c] | 16 | 88,937,225 | 16q24.3 | -5.86 | 4.74E-09 | 0.36 | *PABPN1L* ; *CBFA2T3* | Intergenic |
| cg00448220 | 17 | 624,386 | 17p13.3 | -5.24 | 1.59E-07 | 0.32 | *VPS53* ; *TLCD3A* | Intergenic |
| cg27345989 | 17 | 790,735 | 17p13.3 | 5.35 | 8.91E-08 | 0.60 | *NXN* | Intronic |
| cg19956147 | 17 | 800,431 | 17p13.3 | 5.69 | 1.26E-08 | 0.38 | *NXN* | Intronic |
| cg08190450 | 17 | 800,717 | 17p13.3 | 6.24 | 4.50E-10 | 0.41 | *NXN* | Intronic |
| cg08104960 | 17 | 807,555 | 17p13.3 | 6.38 | 1.76E-10 | 0.72 | *NXN* | Intronic |
| cg07965995 | 17 | 811,389 | 17p13.3 | -7.59 | 3.26E-14 | 0.81 | *NXN* | Intronic |
| cg08081173 | 17 | 811,505 | 17p13.3 | -6.18 | 6.49E-10 | 0.46 | *NXN* | Intronic |
| cg00383244 | 17 | 812,734 | 17p13.3 | 5.59 | 2.27E-08 | 0.46 | *NXN* | Intronic |
| cg17655357 | 17 | 813,389 | 17p13.3 | 9.07 | 1.18E-19 | 0.55 | *LOC101927727* | Intronic |
| cg07128379 | 17 | 10,533,368 | 17p13.1 | -6.52 | 6.87E-11 | 0.18 | *MYH3* | Intronic |
| cg13876325 [c] | 17 | 16,190,098 | 17p11.2 | -6.19 | 5.98E-10 | 0.20 | *PIGL* | Intronic |
| cg15210992 [c] | 17 | 41,002,900 | 17q21.31 | -5.83 | 5.42E-09 | 0.27 | *AOC3* ; *AOC2* | TSS1500 |
| cg20629788 | 17 | 55,055,383 | 17q22 | 5.37 | 7.82E-08 | 0.35 | *SCPEP1* | TSS200 |
| cg15870043 | 17 | 55,175,165 | 17q22 | 11.08 | 1.59E-28 | 0.22 | *AKAP1* | TSS200 |
| cg10060631 | 17 | 81,024,353 | 17q25.3 | 5.41 | 6.34E-08 | 0.31 | *B3GNTL1* ; *METRNL* | Intergenic |
| cg07005960 | 17 | 81,050,498 | 17q25.3 | -5.30 | 1.16E-07 | 0.33 | *METRNL* | Intronic |
| cg13054523 | 17 | 81,055,722 | 17q25.3 | -5.50 | 3.81E-08 | 0.49 | *METRNL* ; *RPL23AP87* | Intergenic |
| cg15082992 | 17 | 81,057,757 | 17q25.3 | 7.35 | 2.02E-13 | 0.36 | *METRNL* ; *RPL23AP87* | Intergenic |
| cg27176264 | 17 | 81,060,149 | 17q25.3 | 8.29 | 1.10E-16 | 0.83 | *METRNL* ; *RPL23AP87* | Intergenic |
| cg13023205 | 17 | 81,060,243 | 17q25.3 | 8.24 | 1.67E-16 | 0.82 | *METRNL* ; *RPL23AP87* | Intergenic |
| cg03185552 | 17 | 81,060,259 | 17q25.3 | 8.55 | 1.25E-17 | 0.82 | *METRNL* ; *RPL23AP87* | Intergenic |
| cg19341630 | 18 | 46,141,962 | 18q21.1 | 5.97 | 2.42E-09 | 0.36 | *CTIF* | Intronic |
| cg21082132 | 18 | 46,226,956 | 18q21.1 | -15.59 | 8.84E-55 | 0.21 | *CTIF* | Intronic |
| cg02216442 | 18 | 46,381,931 | 18q21.1 | 6.13 | 8.52E-10 | 0.60 | *CTIF* | Intronic |
| cg02994053 | 18 | 46,398,337 | 18q21.1 | -14.42 | 3.92E-47 | 0.18 | *CTIF* ; *SMAD7* | Intergenic |
| cg23229732 | 18 | 46,398,778 | 18q21.1 | -5.18 | 2.21E-07 | 0.40 | *CTIF* ; *SMAD7* | Intergenic |
| cg11909137 | 18 | 46,461,202 | 18q21.1 | -12.17 | 4.24E-34 | 0.20 | *SMAD7* | Intronic |
| cg21984044 | 18 | 46,465,677 | 18q21.1 | 5.91 | 3.52E-09 | 0.23 | *SMAD7* | Intronic |
| cg20800216 | 18 | 46,471,349 | 18q21.1 | -5.79 | 7.00E-09 | 0.19 | *SMAD7* | Intronic |
| cg01898573 | 18 | 46,479,537 | 18q21.1 | -5.59 | 2.25E-08 | 0.35 | *SMAD7* ; *LOC100129878* | Intergenic |
| cg08747744 | 19 | 33,166,019 | 19q13.11 | 8.00 | 1.25E-15 | 0.20 | *ANKRD27* | 5'UTR |
| cg16468363 | 19 | 33,543,282 | 19q13.11 | -6.31 | 2.74E-10 | 0.35 | *RHPN2* | Intronic |
| cg03563238 | 19 | 33,554,763 | 19q13.11 | 5.43 | 5.56E-08 | 0.51 | *RHPN2* | Intronic |
| cg07827796 | 19 | 33,622,959 | 19q13.11 | -5.37 | 7.74E-08 | 0.35 | *WDR88* | TSS200 |
| cg08477640 | 19 | 41,863,820 | 19q13.2 | 8.26 | 1.52E-16 | 0.37 | *B9D2* | TSS200 |
| cg19133199 | 19 | 41,869,409 | 19q13.2 | 7.82 | 5.48E-15 | 0.47 | *B9D2* | Exonic |
| cg02063076 | 19 | 41,881,163 | 19q13.2 | -6.41 | 1.43E-10 | 0.65 | *TMEM91* | Intronic |
| cg19497572 | 19 | 41,882,098 | 19q13.2 | 8.00 | 1.22E-15 | 0.39 | *TMEM91* | Intronic |
| cg07663732 | 19 | 41,882,741 | 19q13.2 | -6.60 | 4.10E-11 | 0.29 | *TMEM91* | 5'UTR |
| cg14132834 | 19 | 41,945,861 | 19q13.2 | -5.20 | 2.01E-07 | 0.48 | *ATP5SL* | TSS1500 |
| cg10384570 | 19 | 41,947,118 | 19q13.2 | 5.94 | 2.85E-09 | 0.18 | *DMAC2* ; *ERICH4* | Intergenic |
| cg25675409 | 19 | 58,893,475 | 19q13.43 | -5.75 | 8.68E-09 | 0.23 | *ZNF837* ; *MIR4754* | Intergenic |
| cg24465855 | 19 | 58,986,510 | 19q13.43 | 5.38 | 7.30E-08 | 0.32 | *ZNF324* | 3'UTR |
| cg10598434 | 19 | 59,028,520 | 19q13.43 | 5.77 | 7.98E-09 | 0.19 | *ZBTB45* | Exonic |
| cg21618549 | 19 | 59,067,270 | 19q13.43 | -5.65 | 1.60E-08 | 0.36 | *UBE2M* | 3'UTR |
| cg04170623 | 19 | 59,071,414 | 19q13.43 | -6.10 | 1.04E-09 | 0.66 | *MZF1-AS1* | Intronic |
| cg11707067 | 19 | 59,073,430 | 19q13.43 | -5.78 | 7.32E-09 | 0.17 | *MZF1-AS1* | Intronic |
| cg26954695 | 19 | 59,085,561 | 19q13.43 | -6.57 | 5.10E-11 | 0.38 | *MZF1-AS1* | Exonic |

| | | | | | | | | |
|---|---|---|---|---|---|---|---|---|
| cg07786254 | 19 | 59,085,608 | 19q13.43 | -5.70 | 1.17E-08 | 0.29 | *MZF1-AS1* | Exonic |
| cg18113259 | 20 | 5,986,702 | 20p12.3 | -6.47 | 1.00E-10 | 0.17 | *CRLS1* | 5'UTR |
| cg18304342 | 20 | 6,362,368 | 20p12.3 | 7.36 | 1.83E-13 | 0.21 | *FERMT1* ; *CASC20* | Intergenic |
| cg11506090 | 20 | 6,427,265 | 20p12.3 | 8.77 | 1.75E-18 | 0.16 | *CASC20* | Intronic |
| cg12917994 | 20 | 6,685,966 | 20p12.3 | 6.76 | 1.40E-11 | 0.26 | *CASC20* ; *LINC01713* | Intergenic |
| cg07617394 | 20 | 6,690,692 | 20p12.3 | 8.28 | 1.19E-16 | 0.28 | *CASC20* ; *LINC01713* | Intergenic |
| cg20416330 | 20 | 6,693,183 | 20p12.3 | -11.10 | 1.20E-28 | 0.31 | *CASC20* ; *LINC01713* | Intergenic |
| cg26536259 | 20 | 6,748,006 | 20p12.3 | 5.37 | 7.88E-08 | 0.20 | *BMP2* | TSS1500 |
| cg01797527 | 20 | 6,749,480 | 20p12.3 | 9.15 | 5.76E-20 | 0.17 | *BMP2* | 5'UTR |
| cg06879302 | 20 | 6,760,769 | 20p12.3 | 6.52 | 7.02E-11 | 0.19 | *BMP2* | 3'UTR |
| cg14334648 | 20 | 7,831,901 | 20p12.3 | 9.04 | 1.58E-19 | 0.24 | *MIR8062* ; *HAO1* | Intergenic |
| cg03552151 | 20 | 33,103,584 | 20q11.22 | -5.29 | 1.25E-07 | 0.26 | *DYNLRB1* | TSS1500 |
| cg00436005 | 20 | 33,115,150 | 20q11.22 | 5.57 | 2.49E-08 | 0.33 | *DYNLRB1* | Intronic |
| cg05054831 | 20 | 33,265,543 | 20q11.22 | -5.50 | 3.72E-08 | 0.31 | *PIGU* | TSS1500 |
| cg16987856 | 20 | 47,201,581 | 20q13.13 | -5.46 | 4.88E-08 | 0.65 | *LINC00494* ; *PREX1* | Intergenic |
| cg09517540 | 20 | 47,231,625 | 20q13.13 | -5.59 | 2.25E-08 | 0.22 | *LINC00494* ; *PREX1* | Intergenic |
| cg03740761 | 20 | 47,237,270 | 20q13.13 | -5.52 | 3.45E-08 | 0.17 | *LINC00494* ; *PREX1* | Intergenic |
| cg12681499 | 20 | 47,250,851 | 20q13.13 | 5.77 | 7.79E-09 | 0.32 | *PREX1* | Intronic |
| cg00481013 | 20 | 47,260,660 | 20q13.13 | -5.78 | 7.41E-09 | 0.22 | *PREX1* | Intronic |
| cg05997891 | 20 | 47,332,991 | 20q13.13 | -7.06 | 1.71E-12 | 0.22 | *PREX1* | Intronic |
| cg06903512 | 20 | 47,340,424 | 20q13.13 | 11.42 | 3.46E-30 | 0.57 | *PREX1* | Intronic |
| cg13900919 | 20 | 47,340,601 | 20q13.13 | 11.81 | 3.49E-32 | 0.53 | *PREX1* | Intronic |
| cg11494288 | 20 | 47,341,147 | 20q13.13 | 9.66 | 4.37E-22 | 0.28 | *PREX1* | Intronic |
| cg13145583 | 20 | 47,345,118 | 20q13.13 | 5.66 | 1.54E-08 | 0.29 | *PREX1* | Intronic |
| cg15849237 | 20 | 47,360,150 | 20q13.13 | -6.16 | 7.47E-10 | 0.58 | *PREX1* | Intronic |
| cg09219302 | 20 | 47,435,660 | 20q13.13 | -7.52 | 5.50E-14 | 0.19 | *PREX1* | Intronic |
| cg08162572 | 20 | 47,663,143 | 20q13.13 | -6.46 | 1.02E-10 | 0.20 | *CSE1L* | Intronic |
| cg07615977 | 20 | 48,770,918 | 20q13.13 | -5.73 | 1.01E-08 | 0.69 | *TMEM189* ; *TMEM189-UBE2V1* | TSS1500 |
| cg06824303 | 20 | 48,780,410 | 20q13.13 | 5.45 | 5.07E-08 | 0.29 | *TMEM189-UBE2V1* ; *LINC01273* | Intergenic |
| cg03319948 | 20 | 48,784,962 | 20q13.13 | -5.81 | 6.19E-09 | 0.22 | *TMEM189-UBE2V1* ; *LINC01273* | Intergenic |
| cg15674748 | 20 | 48,788,359 | 20q13.13 | 5.55 | 2.91E-08 | 0.59 | *LINC01273* | TSS1500 |
| cg05499559 | 20 | 48,789,176 | 20q13.13 | 5.63 | 1.85E-08 | 0.21 | *LINC01273* | Exonic |
| cg05993759 | 20 | 48,964,698 | 20q13.13 | 6.45 | 1.13E-10 | 0.33 | *LINC01271* ; *PTPN1* | Intergenic |
| cg08013764 | 20 | 48,983,213 | 20q13.13 | 6.87 | 6.55E-12 | 0.37 | *LINC01271* ; *PTPN1* | Intergenic |
| cg08438589 | 20 | 48,986,614 | 20q13.13 | -9.12 | 7.61E-20 | 0.41 | *LINC01271* ; *PTPN1* | Intergenic |
| cg01252585 | 20 | 48,999,791 | 20q13.13 | -7.94 | 2.10E-15 | 0.40 | *LINC01271* ; *PTPN1* | Intergenic |
| cg04183873 | 20 | 49,044,372 | 20q13.13 | -6.04 | 1.52E-09 | 0.36 | *LINC01271* ; *PTPN1* | Intergenic |
| cg05098991 | 20 | 49,057,225 | 20q13.13 | -9.72 | 2.39E-22 | 0.27 | *LINC01271* ; *PTPN1* | Intergenic |
| cg27204212 | 20 | 49,062,303 | 20q13.13 | 9.52 | 1.80E-21 | 0.33 | *LINC01271* ; *PTPN1* | Intergenic |
| cg15287917 | 20 | 49,092,675 | 20q13.13 | 5.35 | 8.64E-08 | 0.38 | *LINC01271* ; *PTPN1* | Intergenic |
| cg27085045 | 20 | 49,093,894 | 20q13.13 | 5.39 | 6.95E-08 | 0.46 | *LINC01271* ; *PTPN1* | Intergenic |
| cg25654021 | 20 | 49,261,777 | 20q13.13 | 5.32 | 1.02E-07 | 0.34 | *RIPOR3* | Intronic |
| cg18111764 | 20 | 49,261,887 | 20q13.13 | 5.16 | 2.45E-07 | 0.58 | *RIPOR3* | Intronic |
| cg08013100 | 20 | 49,279,727 | 20q13.13 | -5.21 | 1.84E-07 | 0.35 | *RIPOR3* | Intronic |
| cg15154178 | 20 | 49,289,699 | 20q13.13 | -5.43 | 5.61E-08 | 0.26 | *RIPOR3* | Intronic |
| cg10795973 | 20 | 55,629,748 | 20q13.31 | -5.87 | 4.42E-09 | 0.19 | *TFAP2C* ; *BMP7* | Intergenic |
| cg03140378 | 20 | 55,787,633 | 20q13.31 | -6.13 | 8.68E-10 | 0.28 | *BMP7* | Intronic |
| cg11406886 | 20 | 55,804,869 | 20q13.31 | 6.22 | 5.12E-10 | 0.52 | *BMP7* | Intronic |
| cg13089205 | 20 | 55,809,563 | 20q13.31 | -5.41 | 6.18E-08 | 0.79 | *BMP7* | Intronic |
| cg13236649 | 20 | 55,835,020 | 20q13.31 | -6.90 | 5.38E-12 | 0.66 | *BMP7* | Intronic |
| cg05067690 | 20 | 60,806,710 | 20q13.33 | -5.34 | 9.11E-08 | 0.35 | *LOC105369209* ; *MIR1-1HG-AS1* | Intronic |
| cg08079813 | 20 | 60,812,344 | 20q13.33 | 5.52 | 3.39E-08 | 0.39 | *MIR1-1HG-AS1* | Intronic |
| cg03199926 | 20 | 60,860,733 | 20q13.33 | 5.34 | 9.07E-08 | 0.75 | *MIR1-1HG-AS1* | Intronic |
| cg10213353 | 20 | 60,860,905 | 20q13.33 | 5.34 | 9.34E-08 | 0.81 | *MIR1-1HG-AS1* | Intronic |
| cg10925829 | 20 | 60,860,984 | 20q13.33 | 5.27 | 1.40E-07 | 0.79 | *MIR1-1HG-AS1* | Intronic |

| | | | | | | | | |
|---|---|---|---|---|---|---|---|---|
| cg10764574 | 20 | 60,884,293 | 20q13.33 | -6.57 | 5.11E-11 | 0.29 | *MIR1-1HG-AS1* | Intronic |
| cg16496398 | 20 | 60,884,376 | 20q13.33 | 7.60 | 2.87E-14 | 0.57 | *MIR1-1HG-AS1* | Intronic |
| cg09594475 | 20 | 60,884,601 | 20q13.33 | -6.99 | 2.85E-12 | 0.68 | *MIR1-1HG-AS1* | Intronic |
| cg17213352 | 20 | 60,886,007 | 20q13.33 | 10.95 | 6.31E-28 | 0.24 | *LAMA5* | Exonic |
| cg19674797 | 20 | 60,886,077 | 20q13.33 | 5.43 | 5.53E-08 | 0.27 | *LAMA5* | Exonic |
| cg08597733 | 20 | 60,886,158 | 20q13.33 | 7.34 | 2.10E-13 | 0.28 | *MIR1-1HG-AS1* | Intronic |
| cg12382846 | 20 | 60,892,121 | 20q13.33 | -7.23 | 4.77E-13 | 0.65 | *MIR1-1HG-AS1* | Intronic |
| cg08176005 | 20 | 60,901,339 | 20q13.33 | 9.93 | 3.09E-23 | 0.27 | *MIR1-1HG-AS1* | Intronic |
| cg17889447 | 20 | 60,901,850 | 20q13.33 | -7.02 | 2.22E-12 | 0.44 | *MIR1-1HG-AS1* | Intronic |
| cg22307297 | 20 | 60,903,441 | 20q13.33 | -10.00 | 1.50E-23 | 0.39 | *LAMA5* | Exonic |
| cg15193198 | 20 | 60,906,057 | 20q13.33 | 11.01 | 3.51E-28 | 0.60 | *MIR1-1HG-AS1* | Intronic |
| cg04969764 | 20 | 60,906,201 | 20q13.33 | 10.96 | 6.12E-28 | 0.40 | *MIR1-1HG-AS1* | Intronic |
| cg07447854 | 20 | 60,907,525 | 20q13.33 | 11.27 | 1.77E-29 | 0.22 | *LAMA5* | Exonic |
| cg03055693 | 20 | 60,910,388 | 20q13.33 | 7.62 | 2.47E-14 | 0.18 | *MIR1-1HG-AS1* | Intronic |
| cg23251868 | 20 | 60,911,989 | 20q13.33 | 9.21 | 3.23E-20 | 0.51 | *MIR1-1HG-AS1* | Intronic |
| cg26418193 | 20 | 60,918,171 | 20q13.33 | 14.62 | 2.16E-48 | 0.27 | *MIR1-1HG-AS1* | Intronic |
| cg18668449 | 20 | 60,925,179 | 20q13.33 | 8.71 | 3.12E-18 | 0.40 | *MIR1-1HG-AS1* | Intronic |
| cg01059881 | 20 | 60,927,922 | 20q13.33 | 12.58 | 2.63E-36 | 0.21 | *MIR1-1HG-AS1* | Intronic |
| cg23491107 | 20 | 60,927,975 | 20q13.33 | 10.73 | 7.23E-27 | 0.26 | *MIR1-1HG-AS1* | Intronic |
| cg02024094 | 20 | 60,937,585 | 20q13.33 | 10.25 | 1.14E-24 | 0.52 | *LAMA5* | Exonic |
| cg02605258 | 20 | 60,940,772 | 20q13.33 | 11.34 | 8.16E-30 | 0.36 | *MIR1-1HG-AS1* | Intronic |
| cg24461506 | 20 | 60,948,495 | 20q13.33 | 5.17 | 2.32E-07 | 0.28 | *MIR1-1HG-AS1* | Intronic |
| cg06419409 | 20 | 60,948,961 | 20q13.33 | -7.12 | 1.09E-12 | 0.68 | *MIR1-1HG-AS1* | Intronic |
| cg24112000 | 20 | 60,950,667 | 20q13.33 | 11.16 | 6.23E-29 | 0.81 | *MIR1-1HG-AS1* | Intronic |
| cg16585368 | 20 | 60,956,324 | 20q13.33 | -11.95 | 6.49E-33 | 0.65 | *MIR1-1HG-AS1* | Intronic |
| cg25415966 | 20 | 60,964,137 | 20q13.33 | 6.30 | 3.02E-10 | 0.15 | *MIR1-1HG-AS1* | Intronic |
| cg22601191 | 20 | 60,968,625 | 20q13.33 | -12.77 | 2.40E-37 | 0.57 | *CABLES2* | Exonic |
| cg23531748 | 20 | 60,969,906 | 20q13.33 | -6.49 | 8.72E-11 | 0.17 | *MIR1-1HG-AS1* | Intronic |
| cg10923088 | 20 | 60,976,987 | 20q13.33 | -7.01 | 2.44E-12 | 0.24 | *MIR1-1HG-AS1* | Intronic |
| cg14208357 | 20 | 60,977,008 | 20q13.33 | -14.81 | 1.28E-49 | 0.27 | *MIR1-1HG-AS1* | Intronic |
| cg15674680 | 20 | 60,982,522 | 20q13.33 | -6.00 | 1.98E-09 | 0.50 | *MIR1-1HG-AS1* | Intronic |
| cg22633769 | 20 | 60,982,531 | 20q13.33 | -7.38 | 1.57E-13 | 0.62 | *MIR1-1HG-AS1* | Intronic |
| cg22541963 | 20 | 60,982,533 | 20q13.33 | -7.81 | 5.61E-15 | 0.71 | *MIR1-1HG-AS1* | Intronic |
| cg03924136 | 20 | 62,367,108 | 20q13.33 | 5.87 | 4.38E-09 | 0.27 | *LIME1* | 5'UTR |
| cg11650372 [c] | 21 | 35,171,932 | 21q22.11 | 8.92 | 4.49E-19 | 0.18 | *ITSN1* | Intronic |
| cg15089774 | 21 | 47,739,184 | 21q22.3 | -5.77 | 7.77E-09 | 0.41 | *C21orf58* | Intronic |
| cg00989069 | 21 | 47,777,635 | 21q22.3 | 6.18 | 6.35E-10 | 0.33 | *PCNT* | Intronic |
| cg26884033 | 21 | 47,786,512 | 21q22.3 | 5.93 | 3.11E-09 | 0.54 | *PCNT* | Exonic |
| cg10943305 | 21 | 47,790,232 | 21q22.3 | 7.01 | 2.35E-12 | 0.69 | *PCNT* | Intronic |
| cg00879155 | 21 | 47,829,917 | 21q22.3 | -5.89 | 3.96E-09 | 0.42 | *PCNT* | Intronic |
| cg20080983 | 22 | 29,430,271 | 22q12.1 | 5.69 | 1.25E-08 | 0.38 | *ZNRF3* | Intronic |
| cg12768652 | 22 | 39,619,399 | 22q13.1 | -5.59 | 2.32E-08 | 0.59 | *PDGFB* | 3'UTR |
| cg25676471 | 22 | 39,633,497 | 22q13.1 | -5.33 | 9.81E-08 | 0.78 | *PDGFB* | Intronic |
| cg15641875 | 22 | 39,637,489 | 22q13.1 | 5.29 | 1.22E-07 | 0.17 | *PDGFB* | Intronic |
| cg17070577 | 22 | 39,642,168 | 22q13.1 | -5.60 | 2.20E-08 | 0.27 | *PDGFB* ; *RPL3* | Intergenic |
| cg12059304 | 22 | 39,642,701 | 22q13.1 | -5.52 | 3.43E-08 | 0.47 | *PDGFB* ; *RPL3* | Intergenic |
| cg20539366 [d] | 22 | 46,370,644 | 22q13.31 | -5.84 | 5.18E-09 | 0.48 | *WNT7B* | Intronic |
| cg05475172 [d] | 22 | 46,371,152 | 22q13.31 | -5.85 | 5.05E-09 | 0.65 | *WNT7B* | Intronic |
| cg24036761 | 22 | 46,394,385 | 22q13.31 | -5.33 | 9.79E-08 | 0.42 | *WNT7B* ; *LOC730668* | Intergenic |

OR, odds ratio per standard deviation (SD) increase in genetically predicted DNA methylation level; CI, confidence interval; TSS, transcription start site; UTR, untranslated region.

[a] Association Z scores, ORs, 95% CIs, and *P* values were estimated using SPrediXcan. All statistical tests were two-sided.

[b] Correlation coefficients of predicted and measured DNA methylation levels.

[c] CpGs that are >1Mb away from any GWAS-identified SNPs and remain significantly associated with colorectal cancer risk after conditioning on nearest GWAS signals.

[d] CpGs within 1Mb of GWAS-identified SNPs but remain significantly associated with colorectal cancer risk after conditioning on nearest GWAS signals.

**Supplementary Table 3. Associations with risks of renal cell cancer subtypes for CpGs showing a significant association with overall renal cell cancer risk**

| CpG | Chr | Pos | Cytoband | Z score [a] | *P* [a] | R [b] | Closest gene | Classification | Clear cell RCC | | Papillary RCC | | $I^2$ | Cochran's *P* |
|---|---|---|---|---|---|---|---|---|---|---|---|---|---|---|
| | | | | | | | | | Z score [a] | *P* [a] | Z score [a] | *P* [a] | | |
| **Associations showing a nominal significant heterogeneity across subtypes (3.60E-07 [0.05/138,849; Bonferroni correction]<Cochran's *P*<0.05)** | | | | | | | | | | | | | | |
| cg14358543 | 2 | 46,555,124 | 2p21 | -9.09 | 1.02E-19 | 0.50 | *EPAS1* | Intronic | -7.40 | **1.41E-13** | -0.62 | 0.54 | 75.2 | 0.04 |
| cg27476253 | 11 | 69,066,655 | 11q13.3 | -7.69 | 1.46E-14 | 0.40 | *MYEOV; LOC102724265* | Intergenic | -6.23 | **4.56E-10** | 0.53 | 0.60 | 90.0 | 1.59E-03 |
| cg13524857 | 11 | 69,240,192 | 11q13.3 | 5.82 | 5.88E-09 | 0.41 | *LOC102724265* | upstream | 5.37 | **7.79E-08** | -3.34 | 8.47E-04 | 96.1 | 4.75E-07 |
| cg07156636 | 11 | 69,240,438 | 11q13.3 | 6.77 | 1.26E-11 | 0.61 | *LOC102724265* | 5'UTR | 4.73 | 2.19E-06 | -2.55 | 0.01 | 94.1 | 3.88E-05 |
| cg06597413 | 11 | 69,251,883 | 11q13.3 | -8.22 | 1.99E-16 | 0.33 | *LOC102724265*; *LINC01488* | Intergenic | -5.36 | **8.37E-08** | 1.11 | 0.27 | 88.9 | 2.66E-03 |
| cg09854615 | 11 | 69,259,788 | 11q13.3 | -5.24 | 1.63E-07 | 0.41 | *LOC102724265*; *LINC01488* | Intergenic | -3.34 | 8.43E-04 | 1.69 | 0.09 | 87.1 | 5.30E-03 |
| cg00522213 | 12 | 125,318,539 | 12q24.31 | -6.95 | 3.56E-12 | 0.72 | *SCARB1* | Intronic | -6.11 | **1.02E-09** | 1.99 | 0.05 | 93.7 | 6.61E-05 |
| cg07948866 | 12 | 125,336,990 | 12q24.31 | 5.18 | 2.25E-07 | 0.37 | *SCARB1* | Intronic | 5.09 | 3.65E-07 | -0.58 | 0.56 | 82.3 | 0.02 |
| cg06917998 | 14 | 73,269,031 | 14q24.2 | -5.20 | 2.00E-07 | 0.51 | *DPF3* | Intronic | -4.38 | 1.20E-05 | 0.80 | 0.42 | 81.0 | 0.02 |
| **Associations not showing significant heterogeneity across subtypes (Cochran's *P*>0.05)** | | | | | | | | | | | | | | |
| cg26526276 | 2 | 46,585,929 | 2p21 | -7.80 | 6.00E-15 | 0.33 | *EPAS1* | Intronic | -5.60 | **2.08E-08** | -0.30 | 0.77 | 65.1 | 0.09 |
| cg14818391 | 3 | 40,428,506 | 3p22.1 | -5.23 | 1.69E-07 | 0.47 | *ENTPD3* | TSS1500 | -3.70 | 2.16E-04 | -1.04 | 0.30 | 0.0 | 0.71 |
| cg10755058 | 3 | 40,428,713 | 3p22.1 | -5.20 | 1.94E-07 | 0.53 | *ENTPD3* | 5'UTR | -3.56 | 3.74E-04 | -1.11 | 0.27 | 0.0 | 0.81 |
| cg00048149 | 12 | 26,472,528 | 12p12.1 | -6.04 | 1.56E-09 | 0.63 | *SSPN*; *ITPR2* | Intergenic | -5.45 | **5.17E-08** | -0.78 | 0.43 | 40.7 | 0.19 |
| cg06511653 | 12 | 26,472,706 | 12p12.1 | -5.35 | 8.84E-08 | 0.51 | *SSPN*; *ITPR2* | Intergenic | -4.72 | 2.35E-06 | -1.81 | 0.07 | 0.0 | 0.99 |
| cg23332732 | 12 | 26,986,274 | 12p11.23 | -5.60 | 2.20E-08 | 0.49 | *ITPR2* | TSS200 | -4.35 | 1.39E-05 | -0.90 | 0.37 | 0.0 | 0.49 |
| cg02788686 | 14 | 72,920,506 | 14q24.2 | 5.44 | 5.44E-08 | 0.37 | *RGS6* | Intronic | 5.06 | 4.30E-07 | 0.51 | 0.61 | 28.9 | 0.24 |
| cg26376566 | 14 | 73,603,660 | 14q24.2 | -5.74 | 9.62E-09 | 0.39 | *PSEN1* | Intronic | -5.53 | **3.23E-08** | -0.67 | 0.50 | 47.2 | 0.17 |

OR, odds ratio per standard deviation (SD) increase in genetically predicted DNA methylation level; CI, confidence interval; TSS, transcription start site; UTR, untranslated region.

[a] Association Z scores, ORs, 95% CIs, and P values were estimated using SPrediXcan. *P* values that are less than 3.60E-07 (Bonferroni-corrected threshold for 138,849 tested CpGs) are highlighted in bold. All statistical tests were two-sided.

[b] Correlation coefficients of predicted and measured DNA methylation levels.

**Supplementary Table 4. Associations with risks of lung cancer subtypes for CpGs showing a significant association with overall lung cancer risk**

| CpG | Chr | Pos | Cytoband | Z score [a] | *P* [a] | R [b] | Closest gene | Classification | Adenocarcinoma | | Squamous Cell Carcinoma | | Small Cell Lung Cancer | | $I^2$ | Cochran's *P* |
|---|---|---|---|---|---|---|---|---|---|---|---|---|---|---|---|---|
| | | | | | | | | | Z score [a] | *P* [a] | Z score [a] | *P* [a] | Z score [a] | *P* [a] | | |
| **Associations showing a significant heterogeneity across subtypes (Cochran's *P*<2.45E-07[0.05/204,339; Bonferroni correction])** | | | | | | | | | | | | | | | | |
| cg11908978 | 5 | 1,236,791 | 5p15.33 | -5.83 | 5.41E-09 | 0.19 | *SLC6A18* | Intronic | -8.60 | **7.88E-18** | 0.31 | 0.75 | -0.77 | 0.44 | 94.8 | **4.32E-09** |
| cg25045012 | 5 | 1,277,563 | 5p15.33 | -7.59 | 3.32E-14 | 0.73 | *TERT* | Intronic | -9.51 | **1.82E-21** | -1.04 | 0.30 | -2.15 | 0.03 | 93.5 | **1.88E-07** |
| cg00624418 | 5 | 1,282,357 | 5p15.33 | -6.82 | 9.41E-12 | 0.77 | *TERT* | Intronic | -9.26 | **2.02E-20** | -0.06 | 0.95 | -2.32 | 0.02 | 94.4 | **1.71E-08** |
| **Associations showing a nominal significant heterogeneity across subtypes (2.45E-07<Cochran's *P*<0.05)** | | | | | | | | | | | | | | | | |
| cg25518571 | 5 | 1,286,042 | 5p15.33 | -7.36 | 1.86E-13 | 0.30 | *TERT* | Intronic | -10.32 | **5.82E-25** | -0.03 | 0.98 | -2.26 | 0.02 | 91.1 | 1.31E-05 |
| cg23516848 | 5 | 1,297,490 | 5p15.33 | 5.21 | 1.84E-07 | 0.22 | *TERT*; *MIR4457* | Intergenic | 6.06 | **1.33E-09** | 2.00 | 0.05 | -0.44 | 0.66 | 84.7 | 1.48E-03 |
| cg27491040 | 5 | 1,313,825 | 5p15.33 | 11.24 | 2.73E-29 | 0.25 | *MIR4457*; *CLPTM1L* | Intergenic | 9.19 | **4.11E-20** | 7.95 | **1.83E-15** | 2.90 | 3.79E-03 | 77.7 | 0.01 |
| cg26209169 | 5 | 1,316,264 | 5p15.33 | -11.57 | 5.76E-31 | 0.49 | *MIR4457*; *CLPTM1L* | Intergenic | -9.10 | **8.75E-20** | -8.97 | **2.86E-19** | -2.98 | 2.85E-03 | 70.7 | 0.03 |
| cg07774410 | 5 | 1,323,142 | 5p15.33 | 7.00 | 2.51E-12 | 0.33 | *CLPTM1L* | Intronic | 7.17 | **7.48E-13** | 2.17 | 0.03 | 1.37 | 0.17 | 73.2 | 0.02 |
| cg25077823 | 5 | 1,324,772 | 5p15.33 | -6.39 | 1.62E-10 | 0.35 | *CLPTM1L* | Intronic | -3.51 | 4.56E-04 | -7.11 | **1.16E-12** | -1.56 | 0.12 | 91.4 | 9.08E-06 |
| cg13325231 | 5 | 1,342,170 | 5p15.33 | 11.72 | 9.84E-32 | 0.31 | *CLPTM1L* | Intronic | 9.45 | **3.35E-21** | 9.11 | **7.97E-20** | 3.17 | 1.51E-03 | 75.3 | 0.02 |
| cg07493874 | 5 | 1,342,172 | 5p15.33 | 11.80 | 3.68E-32 | 0.52 | *CLPTM1L* | Intronic | 9.45 | **3.39E-21** | 9.20 | **3.50E-20** | 3.11 | 1.86E-03 | 67.0 | 0.05 |
| cg22739207 | 5 | 1,356,382 | 5p15.33 | -5.34 | 9.18E-08 | 0.17 | *CLPTM1L*; *LINC01511* | Intergenic | -5.22 | **1.79E-07** | -1.58 | 0.11 | 0.73 | 0.47 | 82.9 | 2.88E-03 |
| cg13311315 | 6 | 26,124,257 | 6p22.2 | -6.85 | 7.39E-12 | 0.29 | *HIST1H2AC*; *HIST1H2BC* | TSS200 | -2.16 | 0.03 | -7.00 | **2.63E-12** | -3.89 | 1.01E-04 | 89.6 | 6.73E-05 |
| cg16898833 | 6 | 26,189,333 | 6p22.2 | 5.78 | 7.51E-09 | 0.51 | *HIST1H4D* | TSS200 | 1.14 | 0.25 | 6.40 | **1.57E-10** | 3.01 | 2.58E-03 | 88.8 | 1.34E-04 |
| cg12315302 | 6 | 26,189,340 | 6p22.2 | 5.67 | 1.44E-08 | 0.48 | *HIST1H4D* | TSS200 | 0.98 | 0.33 | 6.23 | **4.76E-10** | 3.58 | 3.38E-04 | 90.8 | 1.99E-05 |
| cg05493094 | 6 | 26,200,045 | 6p22.2 | -5.86 | 4.56E-09 | 0.57 | *HIST1H2BF* | Exonic | -2.55 | 0.01 | -5.67 | **1.47E-08** | -3.25 | 1.14E-03 | 84.2 | 1.76E-03 |
| cg01168740 | 6 | 26,229,103 | 6p22.2 | 5.42 | 5.96E-08 | 0.22 | *HIST1H3E*; *HIST1H1D* | Intergenic | 2.66 | 7.73E-03 | 4.49 | 7.00E-06 | 2.72 | 6.47E-03 | 76.0 | 0.02 |
| cg25783550 | 6 | 27,512,906 | 6p22.1 | -5.84 | 5.20E-09 | 0.22 | *ZNF184*; *LINC01012* | Intergenic | -2.27 | 0.02 | -5.83 | **5.44E-09** | -2.41 | 0.02 | 77.0 | 0.01 |
| cg10046620 | 6 | 27,775,042 | 6p22.1 | 6.28 | 3.41E-10 | 0.22 | *HIST1H2AI*; *HIST1H2BL* | TSS1500 | 1.77 | 0.08 | 6.07 | **1.28E-09** | 3.08 | 2.06E-03 | 83.4 | 2.45E-03 |
| cg02228804 | 6 | 27,775,720 | 6p22.1 | 5.85 | 4.96E-09 | 0.27 | *HIST1H2AI*; *HIST1H2BL* | TSS1500 | 1.61 | 0.11 | 5.18 | **2.20E-07** | 3.19 | 1.44E-03 | 79.7 | 7.25E-03 |
| cg00856041 | 6 | 27,798,455 | 6p22.1 | 5.87 | 4.23E-09 | 0.33 | *HIST1H4K* | Downstream | 1.46 | 0.14 | 6.08 | **1.21E-09** | 2.83 | 4.65E-03 | 84.9 | 1.33E-03 |
| cg22955778 | 6 | 27,798,556 | 6p22.1 | 6.49 | 8.85E-11 | 0.40 | *HIST1H4K* | Downstream | 1.97 | 0.05 | 5.66 | **1.52E-08** | 3.63 | 2.83E-04 | 83.2 | 2.59E-03 |
| cg10856605 | 6 | 28,048,756 | 6p22.1 | 5.57 | 2.55E-08 | 0.18 | *ZNF165* | 5'UTR | 1.50 | 0.13 | 5.67 | **1.40E-08** | 2.56 | 0.01 | 82.8 | 3.03E-03 |
| cg16446308 | 6 | 28,235,899 | 6p22.1 | 5.53 | 3.19E-08 | 0.23 | *ZSCAN26* | Intronic | 1.21 | 0.23 | 5.96 | **2.51E-09** | 2.81 | 4.91E-03 | 85.5 | 1.02E-03 |
| cg08638886 | 6 | 28,704,807 | 6p22.1 | -6.48 | 9.09E-11 | 0.49 | *LINC00533*; *LINC01623* | Intergenic | -2.69 | 7.25E-03 | -5.89 | **3.97E-09** | -3.28 | 1.05E-03 | 72.0 | 0.03 |
| cg19918208 | 6 | 28,828,364 | 6p22.1 | -7.20 | 6.07E-13 | 0.30 | *LINC01623* | Exonic | -2.62 | 8.79E-03 | -6.39 | **1.61E-10** | -3.03 | 2.44E-03 | 74.9 | 0.02 |
| cg12914966 | 6 | 28,830,789 | 6p22.1 | -6.64 | 3.18E-11 | 0.52 | *LINC01623* | Intronic | -2.01 | 0.04 | -6.47 | **9.72E-11** | -3.18 | 1.47E-03 | 83.6 | 2.22E-03 |
| cg16677399 | 6 | 28,830,902 | 6p22.1 | -7.25 | 4.02E-13 | 0.51 | *LINC01623* | Intronic | -2.89 | 3.86E-03 | -6.45 | **1.16E-10** | -3.34 | 8.51E-04 | 76.3 | 0.01 |
| cg00903577 | 6 | 28,831,109 | 6p22.1 | -6.40 | 1.53E-10 | 0.55 | *LINC01623* | Exonic | -2.32 | 0.02 | -5.69 | **1.27E-08** | -2.82 | 4.73E-03 | 68.2 | 0.04 |
| cg13269645 | 6 | 28,864,832 | 6p22.1 | 6.82 | 9.04E-12 | 0.51 | *HCG14* | Intronic | 2.00 | 0.05 | 6.60 | **4.25E-11** | 3.39 | 7.00E-04 | 85.4 | 1.05E-03 |
| cg03270340 | 6 | 28,891,204 | 6p22.1 | -6.77 | 1.32E-11 | 0.49 | *TRIM27* | Exonic | -1.93 | 0.05 | -6.42 | **1.34E-10** | -3.50 | 4.64E-04 | 85.6 | 9.76E-04 |
| cg26531141 | 6 | 28,912,013 | 6p22.1 | -5.52 | 3.33E-08 | 0.27 | *LINC01556* | Exonic | -2.00 | 0.05 | -4.93 | 8.14E-07 | -2.87 | 4.15E-03 | 68.2 | 0.04 |
| cg02405128 | 6 | 29,432,580 | 6p22.1 | 5.86 | 4.70E-09 | 0.16 | *OR2H1* | Downstream | 2.48 | 0.01 | 5.66 | **1.49E-08** | 1.86 | 0.06 | 67.1 | 0.05 |
| cg10420223 | 6 | 29,526,561 | 6p22.1 | -8.06 | 7.65E-16 | 0.51 | *UBD* | Intronic | -2.66 | 7.87E-03 | -7.29 | **3.14E-13** | -3.74 | 1.81E-04 | 86.1 | 7.69E-04 |
| cg25729445 | 6 | 29,595,347 | 6p22.1 | -6.23 | 4.58E-10 | 0.19 | *GABBR1* | Exonic | -2.77 | 5.54E-03 | -6.59 | **4.46E-11** | -2.41 | 0.02 | 79.7 | 7.22E-03 |
| cg11358777 | 6 | 29,638,498 | 6p22.1 | 7.17 | 7.29E-13 | 0.28 | *MOG* | Exonic | 3.17 | 1.54E-03 | 5.57 | **2.60E-08** | 3.83 | 1.26E-04 | 70.6 | 0.03 |
| cg12833048 | 6 | 29,639,582 | 6p22.1 | 6.78 | 1.18E-11 | 0.45 | *MOG* | 3'UTR | 2.95 | 3.16E-03 | 5.39 | **6.99E-08** | 3.75 | 1.76E-04 | 68.2 | 0.04 |
| cg10648573 | 6 | 29,648,348 | 6p22.1 | -7.07 | 1.53E-12 | 0.73 | *ZFP57* | Intronic | -3.34 | 8.25E-04 | -5.57 | **2.52E-08** | -3.94 | 8.20E-05 | 67.1 | 0.05 |
| cg00588198 | 6 | 29,648,452 | 6p22.1 | -6.23 | 4.60E-10 | 0.77 | *ZFP57* | Intronic | -2.68 | 7.29E-03 | -4.98 | 6.48E-07 | -4.03 | 5.66E-05 | 72.4 | 0.03 |
| cg20228636 | 6 | 29,648,525 | 6p22.1 | -6.87 | 6.60E-12 | 0.76 | *ZFP57* | Intronic | -3.14 | 1.71E-03 | -5.41 | **6.25E-08** | -3.98 | 6.76E-05 | 67.8 | 0.04 |
| cg03198009 | 6 | 29,648,604 | 6p22.1 | -6.47 | 9.74E-11 | 0.77 | *ZFP57* | Intronic | -2.77 | 5.64E-03 | -5.15 | 2.62E-07 | -4.00 | 6.45E-05 | 71.5 | 0.03 |
| cg13835168 | 6 | 29,648,756 | 6p22.1 | -6.52 | 7.00E-11 | 0.74 | *ZFP57* | Intronic | -2.98 | 2.91E-03 | -5.24 | **1.62E-07** | -3.83 | 1.27E-04 | 67.2 | 0.05 |
| cg08041448 | 6 | 29,648,901 | 6p22.1 | -5.39 | 7.18E-08 | 0.72 | *ZFP57* | 5'UTR | -2.12 | 0.03 | -4.41 | 1.03E-05 | -3.90 | 9.74E-05 | 73.5 | 0.02 |
| cg25252977 | 6 | 29,696,650 | 6p22.1 | -7.64 | 2.10E-14 | 0.53 | *HLA-F-AS1* | Intronic | -4.16 | 3.17E-05 | -5.50 | **3.80E-08** | -4.58 | 4.72E-06 | 80.2 | 6.40E-03 |
| cg11363230 | 6 | 29,796,351 | 6p22.1 | -6.57 | 4.94E-11 | 0.42 | *HLA-G* | Exonic | -3.12 | 1.81E-03 | -5.90 | **3.58E-09** | -3.11 | 1.90E-03 | 68.8 | 0.04 |
| cg21855392 | 6 | 29,798,387 | 6p22.1 | 5.45 | 5.06E-08 | 0.26 | *HLA-G* | Intronic | 2.08 | 0.04 | 5.00 | 5.69E-07 | 3.55 | 3.91E-04 | 73.8 | 0.02 |
| cg26121931 | 6 | 29,895,204 | 6p22.1 | 6.20 | 5.70E-10 | 0.72 | *HCG4B* | TSS1500 | 2.50 | 0.01 | 5.51 | **3.69E-08** | 3.48 | 4.93E-04 | 79.0 | 8.54E-03 |
| cg27538003 | 6 | 29,945,149 | 6p22.1 | -6.53 | 6.41E-11 | 0.37 | *HCG9* | Intronic | -2.81 | 4.92E-03 | -5.66 | **1.50E-08** | -2.96 | 3.07E-03 | 67.4 | 0.05 |
| cg16690356 | 6 | 29,945,948 | 6p22.1 | -8.75 | 2.18E-18 | 0.59 | *HCG9* | Intronic | -3.92 | 8.86E-05 | -7.30 | **2.78E-13** | -3.39 | 7.09E-04 | 75.5 | 0.02 |
| cg15267112 | 6 | 30,000,566 | 6p22.1 | -6.00 | 1.94E-09 | 0.16 | *ZNRD1ASP* | Intronic | -2.52 | 0.01 | -5.60 | **2.17E-08** | -2.93 | 3.43E-03 | 71.2 | 0.03 |
| cg21852658 | 6 | 30,002,671 | 6p22.1 | 7.92 | 2.47E-15 | 0.22 | *ZNRD1ASP* | Intronic | 3.85 | 1.20E-04 | 6.69 | **2.16E-11** | 3.43 | 6.04E-04 | 69.3 | 0.04 |
| cg22184136 | 6 | 30,038,720 | 6p22.1 | -6.27 | 3.66E-10 | 0.17 | *RNF39* | 3'UTR | -2.86 | 4.26E-03 | -4.99 | 6.02E-07 | -4.19 | 2.83E-05 | 73.0 | 0.02 |
| cg06180152 | 6 | 30,043,492 | 6p22.1 | -8.10 | 5.72E-16 | 0.30 | *RNF39* | Exonic | -4.54 | 5.73E-06 | -6.92 | **4.64E-12** | -1.80 | 0.07 | 77.3 | 0.01 |
| cg03782043 | 6 | 30,044,206 | 6p22.1 | -7.06 | 1.70E-12 | 0.41 | *RNF39* | TSS1500 | -2.81 | 4.99E-03 | -6.61 | **3.84E-11** | -3.26 | 1.10E-03 | 77.4 | 0.01 |
| cg01692379 | 6 | 30,044,259 | 6p22.1 | -6.31 | 2.80E-10 | 0.44 | *RNF39* | TSS1500 | -2.76 | 5.70E-03 | -6.03 | **1.64E-09** | -2.79 | 5.33E-03 | 73.4 | 0.02 |
| cg02868178 | 6 | 30,045,280 | 6p22.1 | -7.12 | 1.07E-12 | 0.47 | *RNF39*; *TRIM31* | Intergenic | -3.31 | 9.24E-04 | -6.51 | **7.69E-11** | -3.29 | 1.00E-03 | 73.8 | 0.02 |
| cg16219552 | 6 | 30,057,025 | 6p22.1 | -7.15 | 8.43E-13 | 0.55 | *RNF39*; *TRIM31* | Intergenic | -3.01 | 2.63E-03 | -6.39 | **1.64E-10** | -3.46 | 5.33E-04 | 77.6 | 0.01 |
| cg03228974 | 6 | 30,067,855 | 6p22.1 | -8.14 | 4.01E-16 | 0.44 | *RNF39*; *TRIM31* | Intergenic | -3.60 | 3.23E-04 | -6.95 | **3.55E-12** | -3.87 | 1.08E-04 | 78.7 | 9.06E-03 |
| cg27041554 | 6 | 30,068,899 | 6p22.1 | 5.27 | 1.34E-07 | 0.54 | *RNF39*; *TRIM31* | Intergenic | 1.82 | 0.07 | 4.98 | 6.43E-07 | 3.41 | 6.42E-04 | 76.1 | 0.02 |
| cg02301724 | 6 | 30,068,934 | 6p22.1 | 5.87 | 4.44E-09 | 0.35 | *RNF39*; *TRIM31* | Intergenic | 2.36 | 0.02 | 5.47 | **4.51E-08** | 3.14 | 1.71E-03 | 75.0 | 0.02 |
| cg22442895 | 6 | 30,069,213 | 6p22.1 | 6.06 | 1.37E-09 | 0.23 | *RNF39*; *TRIM31* | Intergenic | 2.67 | 7.52E-03 | 5.32 | **1.01E-07** | 3.24 | 1.19E-03 | 66.7 | 0.05 |
| cg09791883 | 6 | 30,069,659 | 6p22.1 | 6.84 | 8.04E-12 | 0.30 | *RNF39*; *TRIM31* | Intergenic | 2.55 | 0.01 | 5.87 | **4.47E-09** | 4.10 | 4.06E-05 | 77.4 | 0.01 |
| cg04906656 | 6 | 30,069,681 | 6p22.1 | 6.72 | 1.84E-11 | 0.42 | *TRIM31* | Downstream | 2.97 | 2.96E-03 | 5.30 | **1.14E-07** | 3.70 | 2.13E-04 | 67.1 | 0.05 |
| cg15384453 | 6 | 30,070,082 | 6p22.1 | -5.81 | 6.41E-09 | 0.38 | *TRIM31* | Downstream | -2.27 | 0.02 | -4.75 | 1.99E-06 | -3.50 | 4.60E-04 | 70.0 | 0.04 |
| cg13831136 | 6 | 30,070,342 | 6p22.1 | -6.15 | 7.83E-10 | 0.36 | *TRIM31* | Downstream | -2.69 | 7.07E-03 | -5.01 | 5.32E-07 | -3.58 | 3.42E-04 | 68.5 | 0.04 |
| cg10283825 | 6 | 30,071,268 | 6p22.1 | 6.22 | 5.12E-10 | 0.30 | *TRIM31* | 3'UTR | 2.11 | 0.03 | 5.34 | **9.15E-08** | 3.57 | 3.56E-04 | 78.6 | 9.37E-03 |
| cg22377040 | 6 | 30,071,412 | 6p22.1 | 5.74 | 9.56E-09 | 0.50 | *TRIM31* | Exonic | 1.96 | 0.05 | 4.99 | 6.11E-07 | 3.67 | 2.47E-04 | 75.3 | 0.02 |
| cg01249016 | 6 | 30,072,468 | 6p22.1 | -6.76 | 1.35E-11 | 0.39 | *TRIM31* | Intronic | -2.43 | 0.02 | -5.76 | **8.62E-09** | -4.11 | 3.91E-05 | 80.7 | 5.59E-03 |
| cg17102747 | 6 | 30,076,980 | 6p22.1 | -7.19 | 6.68E-13 | 0.16 | *TRIM31-AS1* | Intronic | -3.11 | 1.85E-03 | -6.18 | **6.36E-10** | -3.42 | 6.26E-04 | 72.4 | 0.03 |
| cg03339321 | 6 | 30,080,642 | 6p22.1 | 5.81 | 6.07E-09 | 0.44 | *TRIM31-AS1* | Intronic | 1.41 | 0.16 | 5.43 | **5.61E-08** | 3.86 | 1.13E-04 | 84.4 | 1.62E-03 |
| cg04763709 | 6 | 30,101,683 | 6p22.1 | 5.38 | 7.40E-08 | 0.30 | *TRIM31-AS1*; *TRIM40* | Intergenic | 1.92 | 0.06 | 4.97 | 6.75E-07 | 3.10 | 1.97E-03 | 72.3 | 0.03 |
| cg12612406 | 6 | 30,103,459 | 6p22.1 | -5.66 | 1.48E-08 | 0.22 | *TRIM40* | TSS1500 | -2.43 | 0.02 | -5.60 | **2.13E-08** | -1.68 | 0.09 | 70.8 | 0.03 |
| cg13280080 | 6 | 30,113,786 | 6p22.1 | -7.33 | 2.28E-13 | 0.27 | *TRIM40* | Exonic | -3.19 | 1.40E-03 | -6.07 | **1.25E-09** | -3.68 | 2.37E-04 | 70.7 | 0.03 |
| cg02772121 | 6 | 30,130,881 | 6p22.1 | -5.74 | 9.63E-09 | 0.43 | *TRIM15* | TSS200 | -2.42 | 0.02 | -5.28 | **1.28E-07** | -2.88 | 4.03E-03 | 67.2 | 0.05 |
| cg06051311 | 6 | 30,131,001 | 6p22.1 | -5.52 | 3.47E-08 | 0.24 | *TRIM15* | 5URT | -1.68 | 0.09 | -5.26 | **1.46E-07** | -3.54 | 4.00E-04 | 81.5 | 4.43E-03 |
| cg21856784 | 6 | 30,131,214 | 6p22.1 | -5.43 | 5.53E-08 | 0.35 | *TRIM15* | 5URT | -0.89 | 0.37 | -6.42 | **1.35E-10** | -4.17 | 3.01E-05 | 91.1 | 1.31E-05 |
| cg00720829 | 6 | 30,131,219 | 6p22.1 | -6.91 | 4.91E-12 | 0.38 | *TRIM15* | 5URT;1stExon | -2.39 | 0.02 | -6.20 | **5.59E-10** | -4.61 | 4.11E-06 | 85.5 | 1.00E-03 |

| | | | | | | | | | | | | | | | | |
|---|---|---|---|---|---|---|---|---|---|---|---|---|---|---|---|---|
| cg13396830 | 6 | 30,181,581 | 6p22.1 | 7.75 | 9.22E-15 | 0.17 | *TRIM26* | TSS1500 | 3.46 | 5.35E-04 | 6.83 | **8.34E-12** | 2.59 | 9.56E-03 | 75.8 | 0.02 |
| cg09262446 | 6 | 30,231,689 | 6p22.1 | -7.37 | 1.68E-13 | 0.53 | *HLA-L* | Exonic | -3.40 | 6.63E-04 | -6.25 | **4.23E-10** | -2.62 | 8.79E-03 | 72.2 | 0.03 |
| cg19869086 | 6 | 30,306,285 | 6p22.1 | -7.76 | 8.38E-15 | 0.35 | *TRIM39* ; *TRIM39-RPP21* | Intronic | -3.51 | 4.56E-04 | -6.64 | **3.15E-11** | -2.50 | 0.01 | 73.1 | 0.02 |
| cg22850066 | 6 | 30,313,042 | 6p22.1 | 5.64 | 1.65E-08 | 0.18 | *RPP21* ; *TRIM39-RPP21* | Intronic | 2.24 | 0.02 | 4.98 | 6.25E-07 | 3.59 | 3.26E-04 | 71.3 | 0.03 |
| cg01316659 | 6 | 30,418,115 | 6p21.33 | -6.21 | 5.20E-10 | 0.34 | *RPP21* ;*HLA-E* | Intergenic | -2.12 | 0.03 | -5.98 | **2.20E-09** | -3.41 | 6.59E-04 | 84.0 | 1.90E-03 |
| cg09476067 | 6 | 30,418,581 | 6p21.33 | -8.88 | 6.67E-19 | 0.28 | *RPP21* ; *HLA-E* | Intergenic | -3.84 | 1.24E-04 | -7.69 | **1.50E-14** | -3.50 | 4.73E-04 | 80.7 | 5.59E-03 |
| cg18940843 | 6 | 30,418,773 | 6p21.33 | -6.58 | 4.73E-11 | 0.38 | *RPP21* ; *HLA-E* | Intergenic | -2.95 | 3.16E-03 | -5.30 | **1.13E-07** | -4.19 | 2.79E-05 | 71.4 | 0.03 |
| cg12078775 | 6 | 30,419,543 | 6p21.33 | -7.16 | 8.25E-13 | 0.30 | *RPP21* ; *HLA-E* | Intergenic | -2.25 | 0.02 | -7.19 | **6.60E-13** | -3.95 | 7.82E-05 | 90.9 | 1.77E-05 |
| cg27572120 | 6 | 30,419,551 | 6p21.33 | -6.50 | 8.30E-11 | 0.38 | *RPP21* ; *HLA-E* | Intergenic | -1.94 | 0.05 | -6.23 | **4.77E-10** | -3.18 | 1.47E-03 | 87.9 | 2.50E-04 |
| cg26570901 | 6 | 30,419,576 | 6p21.33 | -6.79 | 1.14E-11 | 0.30 | *RPP21* ; *HLA-E* | Intergenic | -1.93 | 0.05 | -6.84 | **8.08E-12** | -3.61 | 3.08E-04 | 90.5 | 2.68E-05 |
| cg18048542 | 6 | 30,619,113 | 6p21.33 | -5.96 | 2.57E-09 | 0.18 | *C6orf136* | Exonic | -3.32 | 9.04E-04 | -5.53 | **3.18E-08** | -0.67 | 0.50 | 78.5 | 9.48E-03 |
| cg12247101 | 6 | 30,653,549 | 6p21.33 | -5.53 | 3.24E-08 | 0.33 | *PPP1R18* | Exonic | -2.26 | 0.02 | -5.00 | 5.82E-07 | -2.99 | 2.83E-03 | 69.4 | 0.04 |
| cg05978527 | 6 | 30,656,692 | 6p21.33 | 6.08 | 1.18E-09 | 0.16 | *NRM* | Exonic | 2.56 | 0.01 | 4.78 | 1.74E-06 | 4.18 | 2.96E-05 | 74.8 | 0.02 |
| cg24813995 | 6 | 30,657,882 | 6p21.33 | -5.48 | 4.17E-08 | 0.28 | *NRM* | Exonic | -2.37 | 0.02 | -4.98 | 6.21E-07 | -3.97 | 7.24E-05 | 74.9 | 0.02 |
| cg10573158 | 6 | 30,689,496 | 6p21.33 | 6.71 | 1.91E-11 | 0.19 | *TUBB* | 5'UTR | 2.56 | 0.01 | 6.17 | **6.85E-10** | 4.32 | 1.53E-05 | 82.7 | 3.07E-03 |
| cg23397584 | 6 | 30,726,973 | 6p21.33 | 7.26 | 3.89E-13 | 0.28 | *IER3* ;*HCG20* | Intergenic | 2.84 | 4.46E-03 | 7.03 | **2.11E-12** | 4.80 | 1.57E-06 | 86.4 | 6.55E-04 |
| cg15570745 | 6 | 30,731,791 | 6p21.33 | 7.14 | 9.37E-13 | 0.21 | *IER3* ; *HCG20* | Intergenic | 3.35 | 8.18E-04 | 5.83 | **5.65E-09** | 5.45 | **5.15E-08** | 82.5 | 3.25E-03 |
| cg26146308 | 6 | 30,851,884 | 6p21.33 | 7.63 | 2.30E-14 | 0.35 | *DDR1* | 5'UTR | 4.15 | 3.30E-05 | 6.83 | **8.52E-12** | 2.28 | 0.02 | 67.1 | 0.05 |
| cg07265873 | 6 | 30,851,940 | 6p21.33 | 9.22 | 2.94E-20 | 0.35 | *DDR1* | Intronic | 4.03 | 5.63E-05 | 8.07 | **7.32E-16** | 3.82 | 1.31E-04 | 81.9 | 3.96E-03 |
| cg06879746 | 6 | 30,883,768 | 6p21.33 | 5.66 | 1.56E-08 | 0.16 | *VARS2* | Exonic | 2.09 | 0.04 | 6.07 | **1.26E-09** | 4.22 | 2.40E-05 | 79.1 | 8.42E-03 |
| cg14813923 | 6 | 30,886,665 | 6p21.33 | 8.52 | 1.56E-17 | 0.48 | *VARS2* | Exonic | 4.08 | 4.44E-05 | 7.19 | **6.29E-13** | 5.02 | 5.12E-07 | 82.2 | 3.58E-03 |
| cg15992932 | 6 | 30,899,405 | 6p21.33 | -7.06 | 1.66E-12 | 0.48 | *SFTA2* | Intronic | -3.50 | 4.65E-04 | -6.08 | **1.19E-09** | -4.52 | 6.20E-06 | 75.1 | 0.02 |
| cg13561028 | 6 | 30,899,649 | 6p21.33 | -8.00 | 1.26E-15 | 0.73 | *SFTA2* | Intronic | -3.82 | 1.33E-04 | -7.08 | **1.40E-12** | -4.55 | 5.48E-06 | 77.9 | 0.01 |
| cg15063443 | 6 | 30,900,560 | 6p21.33 | 5.76 | 8.40E-09 | 0.43 | *SFTA2* | TSS1500 | 2.68 | 7.40E-03 | 5.44 | **5.24E-08** | 4.23 | 2.29E-05 | 75.3 | 0.02 |
| cg04559908 | 6 | 30,920,123 | 6p21.33 | -8.13 | 4.12E-16 | 0.71 | *MUCL3* | Exonic | -3.75 | 1.76E-04 | -7.09 | **1.39E-12** | -4.67 | 3.07E-06 | 80.0 | 6.69E-03 |
| cg11485823 | 6 | 30,921,592 | 6p21.33 | 6.62 | 3.69E-11 | 0.44 | *HCG21* | Intronic | 2.77 | 5.59E-03 | 5.89 | **3.96E-09** | 3.37 | 7.40E-04 | 73.5 | 0.02 |
| cg18964706 | 6 | 30,921,630 | 6p21.33 | 5.98 | 2.17E-09 | 0.30 | *HCG21* | Intronic | 2.07 | 0.04 | 5.81 | **6.44E-09** | 3.59 | 3.31E-04 | 78.3 | 1.00E-02 |
| cg12797044 | 6 | 30,922,043 | 6p21.33 | 6.29 | 3.23E-10 | 0.30 | *HCG21* | Intronic | 2.90 | 3.77E-03 | 5.33 | **1.00E-07** | 4.18 | 2.93E-05 | 72.6 | 0.03 |
| cg03513762 | 6 | 30,923,420 | 6p21.33 | 6.49 | 8.55E-11 | 0.17 | *HCG21* | Upstream | 2.35 | 0.02 | 6.74 | **1.54E-11** | 4.21 | 2.58E-05 | 86.6 | 5.78E-04 |
| cg13126982 | 6 | 30,949,988 | 6p21.33 | -7.30 | 2.86E-13 | 0.25 | *HCG21* ; *MUC21* | Intergenic | -4.16 | 3.23E-05 | -7.02 | **2.21E-12** | -2.25 | 0.02 | 85.4 | 1.05E-03 |
| cg10715333 | 6 | 31,018,523 | 6p21.33 | -6.74 | 1.60E-11 | 0.37 | *MUC22* ; *HCG22* | Intergenic | -3.04 | 2.35E-03 | -5.92 | **3.24E-09** | -3.44 | 5.81E-04 | 69.0 | 0.04 |
| cg17038049 | 6 | 31,021,365 | 6p21.33 | 5.37 | 7.99E-08 | 0.45 | *HCG22* | Intronic | 1.27 | 0.20 | 5.66 | **1.51E-08** | 3.43 | 6.05E-04 | 83.7 | 2.15E-03 |
| cg22786748 | 6 | 31,034,042 | 6p21.33 | -7.55 | 4.20E-14 | 0.30 | *HCG22* ; *C6orf15* | Intergenic | -2.45 | 0.01 | -7.75 | **8.87E-15** | -4.00 | 6.40E-05 | 89.4 | 8.21E-05 |
| cg16519217 | 6 | 31,038,713 | 6p21.33 | 6.04 | 1.57E-09 | 0.34 | *HCG22* ; *C6orf15* | Intergenic | 1.98 | 0.05 | 5.88 | **4.10E-09** | 3.98 | 6.90E-05 | 85.0 | 1.25E-03 |
| cg22110654 | 6 | 31,051,644 | 6p21.33 | 8.19 | 2.68E-16 | 0.64 | *HCG22* ; *C6orf15* | Intergenic | 3.70 | 2.12E-04 | 7.69 | **1.42E-14** | 4.65 | 3.24E-06 | 84.6 | 1.49E-03 |
| cg08951273 | 6 | 31,079,152 | 6p21.33 | 6.10 | 1.06E-09 | 0.43 | *C6orf15* | 3'UTR | 2.21 | 0.03 | 5.98 | **2.24E-09** | 1.50 | 0.13 | 70.1 | 0.04 |
| cg08097157 | 6 | 31,080,048 | 6p21.33 | 5.24 | 1.57E-07 | 0.37 | *C6orf15* | Exonic | 1.94 | 0.05 | 5.21 | **1.94E-07** | 1.54 | 0.12 | 73.6 | 0.02 |
| cg17849733 | 6 | 31,082,200 | 6p21.33 | -5.81 | 6.10E-09 | 0.51 | *PSORS1C1* ; *CDSN* | TSS1500 | -2.95 | 3.22E-03 | -5.27 | **1.39E-07** | -1.13 | 0.26 | 67.4 | 0.05 |
| cg05875819 | 6 | 31,090,280 | 6p21.33 | -6.00 | 1.98E-09 | 0.29 | *PSORS1C1* | Intronic | -1.81 | 0.07 | -6.15 | **7.68E-10** | -3.01 | 2.65E-03 | 81.4 | 4.56E-03 |
| cg02053920 | 6 | 31,093,846 | 6p21.33 | -5.45 | 5.08E-08 | 0.32 | *PSORS1C1* | Intronic | -3.39 | 7.12E-04 | -4.76 | 1.93E-06 | -0.38 | 0.70 | 74.3 | 0.02 |
| cg02451928 | 6 | 31,125,948 | 6p21.33 | 5.97 | 2.33E-09 | 0.52 | *CCHCR1* | 5'UTR | 2.79 | 5.29E-03 | 5.42 | **5.87E-08** | 4.56 | 5.14E-06 | 78.3 | 0.01 |
| cg23252259 | 6 | 31,148,612 | 6p21.33 | -5.65 | 1.58E-08 | 0.28 | *PSORS1C3* | Intronic | -2.35 | 0.02 | -5.75 | **9.09E-09** | -4.89 | 1.02E-06 | 83.0 | 2.83E-03 |
| cg02936944 | 6 | 31,168,943 | 6p21.33 | -5.83 | 5.66E-09 | 0.18 | *HCG27* | Intronic | -2.17 | 0.03 | -5.60 | **2.13E-08** | -3.49 | 4.75E-04 | 77.9 | 0.01 |
| cg00888093 | 6 | 31,336,635 | 6p21.33 | -5.33 | 1.01E-07 | 0.56 | *HLA-B* ; *MICA-AS1* | Intergenic | -1.55 | 0.12 | -6.08 | **1.18E-09** | -3.89 | 9.94E-05 | 86.8 | 5.27E-04 |
| cg10367909 | 6 | 31,367,038 | 6p21.33 | 5.17 | 2.38E-07 | 0.45 | *MICA-AS1* | Intronic | 1.21 | 0.22 | 6.74 | **1.62E-11** | 3.61 | 3.02E-04 | 90.3 | 3.50E-05 |
| cg03694466 | 6 | 31,409,757 | 6p21.33 | -5.27 | 1.40E-07 | 0.78 | *LINC01149* | Exonic | -1.30 | 0.19 | -6.44 | **1.23E-10** | -3.55 | 3.85E-04 | 90.8 | 1.86E-05 |
| cg02956255 | 6 | 31,430,641 | 6p21.33 | -5.61 | 2.00E-08 | 0.40 | *HCP5* | TSS1500 | -1.78 | 0.07 | -5.71 | **1.12E-08** | -2.80 | 5.17E-03 | 79.3 | 7.99E-03 |
| cg14646094 | 6 | 31,496,752 | 6p21.33 | 5.17 | 2.32E-07 | 0.47 | *MCCD1* | 5'UTR | 1.29 | 0.20 | 5.36 | **8.29E-08** | 3.87 | 1.10E-04 | 87.4 | 3.52E-04 |
| cg20443451 | 6 | 31,497,233 | 6p21.33 | 5.97 | 2.44E-09 | 0.30 | *MCCD1* | Intronic | 2.11 | 0.03 | 6.02 | **1.78E-09** | 3.31 | 9.47E-04 | 81.8 | 4.05E-03 |
| cg11468089 | 6 | 31,506,280 | 6p21.33 | -5.55 | 2.91E-08 | 0.42 | *ATP6V1G2-DDX39B* | Intronic | -1.37 | 0.17 | -6.04 | **1.56E-09** | -4.13 | 3.69E-05 | 88.8 | 1.28E-04 |
| cg05182583 | 6 | 31,598,464 | 6p21.33 | 5.86 | 4.52E-09 | 0.61 | *PRRC2A* | Exonic | 1.37 | 0.17 | 6.60 | **4.09E-11** | 4.01 | 6.09E-05 | 88.2 | 2.08E-04 |
| cg05053818 | 6 | 31,618,987 | 6p21.33 | -6.58 | 4.70E-11 | 0.34 | *BAG6* | Intronic | -2.78 | 5.50E-03 | -6.18 | **6.59E-10** | -3.66 | 2.56E-04 | 72.9 | 0.02 |
| cg03689048 | 6 | 31,619,544 | 6p21.33 | -6.26 | 3.94E-10 | 0.21 | *BAG6* | 5'UTR | -1.74 | 0.08 | -6.72 | **1.85E-11** | -3.58 | 3.44E-04 | 87.5 | 3.45E-04 |
| cg16587838 | 6 | 31,704,705 | 6p21.33 | 6.67 | 2.55E-11 | 0.27 | *CLIC1* | 5'UTR | 2.90 | 3.77E-03 | 6.54 | **6.35E-11** | 2.50 | 0.01 | 80.8 | 5.50E-03 |
| cg10563645 | 6 | 31,705,988 | 6p21.33 | 7.67 | 1.78E-14 | 0.25 | *CLIC1* | TSS1500 | 2.52 | 0.01 | 7.43 | **1.08E-13** | 4.07 | 4.79E-05 | 85.7 | 9.03E-04 |
| cg21091985 | 6 | 31,706,222 | 6p21.33 | 6.52 | 6.87E-11 | 0.50 | *CLIC1* ; *MSH5* | Intergenic | 2.81 | 4.99E-03 | 5.96 | **2.57E-09** | 3.59 | 3.37E-04 | 74.2 | 0.02 |
| cg23634079 | 6 | 31,712,195 | 6p21.33 | -8.13 | 4.22E-16 | 0.59 | *MSH5-SAPCD1* | Intronic | -3.41 | 6.52E-04 | -7.88 | **3.31E-15** | -3.69 | 2.28E-04 | 85.3 | 1.09E-03 |
| cg00330953 | 6 | 31,733,810 | 6p21.33 | -5.67 | 1.40E-08 | 0.21 | *VWA7* | Exonic | -0.99 | 0.32 | -6.81 | **9.95E-12** | -4.31 | 1.64E-05 | 92.3 | 2.14E-06 |
| cg27482961 | 6 | 31,774,303 | 6p21.33 | 5.49 | 4.08E-08 | 0.22 | *LSM2* | Intronic | 2.41 | 0.02 | 3.13 | 1.77E-03 | 3.86 | 1.16E-04 | 67.6 | 0.05 |
| cg12678191 | 6 | 31,802,415 | 6p21.33 | 5.67 | 1.45E-08 | 0.20 | *SNHG32* ; *SNORD48* | TSS1500 | 1.80 | 0.07 | 4.84 | 1.30E-06 | 3.25 | 1.15E-03 | 74.1 | 0.02 |
| cg26747317 | 6 | 31,829,793 | 6p21.33 | -6.21 | 5.22E-10 | 0.16 | *NEU1* | Exonic | -2.42 | 0.02 | -7.22 | **5.37E-13** | -3.91 | 9.19E-05 | 89.0 | 1.10E-04 |
| cg16505946 | 6 | 31,870,305 | 6p21.33 | 8.29 | 1.18E-16 | 0.47 | *C2* | Intronic | 3.51 | 4.41E-04 | 7.84 | **4.35E-15** | 3.69 | 2.27E-04 | 82.3 | 3.57E-03 |
| cg16868572 | 6 | 31,870,310 | 6p21.33 | 6.88 | 5.97E-12 | 0.17 | *C2* | Intronic | 2.52 | 0.01 | 7.07 | **1.55E-12** | 3.22 | 1.30E-03 | 84.6 | 1.52E-03 |
| cg17591348 | 6 | 31,923,212 | 6p21.33 | 6.05 | 1.46E-09 | 0.20 | *NELFE* | Intronic | 2.58 | 9.82E-03 | 6.56 | **5.55E-11** | 2.28 | 0.02 | 90.9 | 1.66E-05 |
| cg03031281 | 6 | 31,941,422 | 6p21.33 | 6.00 | 1.93E-09 | 0.18 | *STK19* | Intronic | 1.50 | 0.13 | 6.70 | **2.06E-11** | 3.67 | 2.42E-04 | 88.7 | 1.48E-04 |
| cg04771084 | 6 | 31,973,255 | 6p21.33 | -6.70 | 2.13E-11 | 0.46 | *CYP21A1P* | TSS200 | -2.53 | 0.01 | -6.34 | **2.30E-10** | -3.33 | 8.63E-04 | 80.0 | 6.75E-03 |
| cg18790646 | 6 | 32,006,196 | 6p21.33 | -5.82 | 5.86E-09 | 0.28 | *CYP21A2* | 5'UTR | -1.97 | 0.05 | -5.60 | **2.20E-08** | -3.27 | 1.06E-03 | 82.4 | 3.43E-03 |
| cg24055029 | 6 | 32,055,137 | 6p21.33 | -5.83 | 5.69E-09 | 0.25 | *TNXB* | Intronic | -1.47 | 0.14 | -5.53 | **3.28E-08** | -2.47 | 0.01 | 79.9 | 6.91E-03 |
| cg26266427 | 6 | 32,063,838 | 6p21.33 | -5.29 | 1.19E-07 | 0.27 | *TNXB* | Exonic | -1.65 | 0.10 | -5.99 | **2.04E-09** | -1.91 | 0.06 | 85.6 | 9.62E-04 |
| cg01992382 | 6 | 32,064,212 | 6p21.33 | -5.22 | 1.76E-07 | 0.31 | *TNXB* | Exonic | -1.16 | 0.25 | -5.13 | 2.94E-07 | -2.21 | 0.03 | 84.1 | 1.82E-03 |
| cg15745284 | 6 | 32,064,923 | 6p21.33 | -7.88 | 3.38E-15 | 0.25 | *TNXB* | Exonic | -3.26 | 1.12E-03 | -7.64 | **2.18E-14** | -4.36 | 1.28E-05 | 88.2 | 2.06E-04 |
| cg27068095 | 6 | 32,119,016 | 6p21.32 | -5.27 | 1.36E-07 | 0.22 | *PRRT1* | Intronic | -2.48 | 0.01 | -5.15 | 2.55E-07 | -2.51 | 0.01 | 72.7 | 0.03 |
| cg05133205 | 6 | 32,121,249 | 6p21.32 | -6.37 | 1.88E-10 | 0.33 | *LOC100507547* | Exonic | -2.71 | 6.68E-03 | -6.42 | **1.38E-10** | -3.32 | 8.94E-04 | 84.3 | 1.71E-03 |
| cg17329164 | 6 | 32,121,259 | 6p21.32 | -5.74 | 9.33E-09 | 0.16 | *LOC100507547* | Exonic | -1.31 | 0.19 | -6.13 | **8.91E-10** | -3.80 | 1.47E-04 | 88.1 | 2.25E-04 |
| cg04105091 | 6 | 32,121,355 | 6p21.32 | -6.02 | 1.79E-09 | 0.16 | *LOC100507547* | Exonic | -1.84 | 0.07 | -6.34 | **2.36E-10** | -3.77 | 1.61E-04 | 86.6 | 5.65E-04 |
| cg00552704 | 6 | 32,121,420 | 6p21.32 | -5.44 | 5.28E-08 | 0.15 | *LOC100507547* | Exonic | -2.32 | 0.02 | -6.45 | **1.10E-10** | -2.67 | 7.48E-03 | 87.3 | 3.72E-04 |
| cg24009529 | 6 | 32,122,470 | 6p21.32 | 5.50 | 3.72E-08 | 0.59 | *PPT2* | Exonic | 1.58 | 0.11 | 5.13 | 2.90E-07 | 2.87 | 4.16E-03 | 74.9 | 0.02 |
| cg11108115 | 6 | 32,135,186 | 6p21.32 | 7.46 | 8.99E-14 | 0.16 | *EGFL8* | Exonic | 2.97 | 2.97E-03 | 7.36 | **1.84E-13** | 2.88 | 3.91E-03 | 84.9 | 1.34E-03 |
| cg06863878 | 6 | 32,168,732 | 6p21.32 | -6.23 | 4.70E-10 | 0.33 | *NOTCH4* | Exonic | -1.73 | 0.08 | -6.67 | **2.51E-11** | -3.30 | 9.75E-04 | 84.6 | 1.49E-03 |
| cg24446242 | 6 | 32,185,340 | 6p21.32 | 5.54 | 3.09E-08 | 0.34 | *NOTCH4* | Intronic | 1.27 | 0.21 | 6.00 | **2.03E-09** | 3.39 | 6.92E-04 | 88.0 | 2.40E-04 |
| cg18416871 | 6 | 32,185,984 | 6p21.32 | 5.35 | 8.63E-08 | 0.23 | *NOTCH4* | Intronic | 1.50 | 0.13 | 5.19 | **2.07E-07** | 2.49 | 0.01 | 75.8 | 0.02 |
| cg25258233 | 6 | 32,407,570 | 6p21.32 | -5.43 | 5.70E-08 | 0.16 | *HLA-DRA* | TSS200 | -2.08 | 0.04 | -5.60 | **2.09E-08** | -1.68 | 0.09 | 78.5 | 9.48E-03 |
| cg15255946 | 6 | 32,729,563 | 6p21.32 | -5.36 | 8.37E-08 | 0.18 | *HLA-DQB2* | Exonic | -2.00 | 0.05 | -5.75 | **8.82E-09** | -3.40 | 6.65E-04 | 84.7 | 1.47E-03 |
| cg04903089 | 6 | 32,905,190 | 6p21.32 | 5.16 | 2.43E-07 | 0.27 | *HLA-DMB* | Exonic | 1.71 | 0.09 | 4.80 | 1.59E-06 | 1.69 | 0.09 | 67.3 | 0.05 |
| cg22961241 | 6 | 32,917,502 | 6p21.32 | 5.77 | 7.71E-09 | 0.33 | *HLA-DMA* | Exonic | 2.18 | 0.03 | 5.82 | **5.82E-09** | 1.49 | 0.14 | 77.0 | 0.01 |

| | | | | | | | | | | | | | | | | |
|---|---|---|---|---|---|---|---|---|---|---|---|---|---|---|---|---|
| cg03531211 | 6 | 32,920,102 | 6p21.32 | 6.14 | 8.10E-10 | 0.22 | *HLA-DMA* | Intronic | 2.39 | 0.02 | 6.28 | **3.35E-10** | 1.16 | 0.25 | 81.4 | 4.61E-03 |
| cg01347674 | 11 | 118,134,073 | 11q23.3 | 5.45 | 5.14E-08 | 0.40 | *MPZL2* | Intronic | 5.75 | **9.03E-09** | 1.50 | 0.13 | 1.78 | 0.07 | 71.3 | 0.03 |
| cg11869183 | 12 | 1,008,948 | 12p13.33 | -5.35 | 9.00E-08 | 0.43 | *WNK1* | Intronic | -1.26 | 0.21 | -5.86 | **4.66E-09** | -4.12 | 3.77E-05 | 90.8 | 1.92E-05 |
| cg07745104 | 15 | 49,430,666 | 15q21.1 | 5.66 | 1.48E-08 | 0.37 | *COPS2* | Intronic | 6.99 | **2.66E-12** | -0.41 | 0.68 | 1.19 | 0.24 | 92.3 | 2.38E-06 |
| cg13561554 | 15 | 78,795,944 | 15q25.1 | -14.55 | 5.83E-48 | 0.46 | *IREB2* ; *HYKK* | Intergenic | -8.72 | **2.85E-18** | -3.29 | 9.97E-04 | -6.58 | **4.75E-11** | 68.9 | 0.04 |
| cg04495498 | 15 | 79,074,250 | 15q25.1 | -11.63 | 2.80E-31 | 0.33 | *ADAMTS7* | Intronic | -6.93 | **4.26E-12** | -7.22 | **5.09E-13** | -4.17 | 3.10E-05 | 87.4 | 3.54E-04 |
| cg21242079 | 15 | 79,101,063 | 15q25.1 | 6.12 | 9.26E-10 | 0.67 | *ADAMTS7* | Intronic | 4.18 | 2.87E-05 | 4.16 | 3.25E-05 | 2.86 | 4.27E-03 | 86.9 | 4.86E-04 |
| cg03938525 | 15 | 79,262,857 | 15q25.1 | 6.03 | 1.65E-09 | 0.17 | *RASGRF1* | Intronic | 3.54 | 3.96E-04 | 3.46 | 5.34E-04 | 5.26 | **1.47E-07** | 81.6 | 4.35E-03 |
| **Associations not showing significant heterogeneity across subtypes (Cochran's *P* >0.05)** | | | | | | | | | | | | | | | | |
| cg08491437 | 1 | 78,442,533 | 1p31.1 | 5.95 | 2.64E-09 | 0.48 | *FUBP1* | Intronic | 5.38 | **7.47E-08** | 3.50 | 4.57E-04 | 2.47 | 0.01 | 0.0 | 0.89 |
| cg26152188 | 5 | 863,294 | 5p15.33 | -6.28 | 3.42E-10 | 0.18 | *BRD9* | Downstream | -4.66 | 3.10E-06 | -3.31 | 9.48E-04 | -1.64 | 0.10 | 0.0 | 0.95 |
| cg04184427 | 5 | 1,076,892 | 5p15.33 | 9.11 | 8.46E-20 | 0.17 | *SLC12A7* | Exonic | 7.13 | **1.00E-12** | 6.30 | **2.94E-10** | 2.38 | 0.02 | 14.4 | 0.31 |
| cg18836287 | 5 | 1,145,044 | 5p15.33 | -6.46 | 1.07E-10 | 0.24 | *SLC12A7* ; *CTD-3080P12.3* | Intergenic | -6.65 | **2.91E-11** | -2.42 | 0.02 | -3.14 | 1.70E-03 | 64.8 | 0.06 |
| cg17026582 | 5 | 1,298,573 | 5p15.33 | -9.48 | 2.59E-21 | 0.25 | *TERT* ; *MIR4457* | Intergenic | -7.18 | **7.11E-13** | -7.59 | **3.21E-14** | -2.63 | 8.44E-03 | 46.3 | 0.16 |
| cg11624060 | 5 | 1,316,038 | 5p15.33 | -11.54 | 8.48E-31 | 0.25 | *MIR4457* ; *CLPTM1L* | Intergenic | -9.35 | **8.62E-21** | -8.81 | **1.27E-18** | -3.01 | 2.59E-03 | 59.9 | 0.08 |
| cg10441424 | 5 | 1,316,636 | 5p15.33 | -11.38 | 5.26E-30 | 0.30 | *MIR4457* ; *CLPTM1L* | Intergenic | -9.29 | **1.56E-20** | -8.79 | **1.52E-18** | -2.90 | 3.77E-03 | 63.5 | 0.06 |
| cg20007063 | 5 | 1,326,407 | 5p15.33 | -11.13 | 8.91E-29 | 0.31 | *CLPTM1L* | Intronic | -8.66 | **4.80E-18** | -8.65 | **5.18E-18** | -2.81 | 4.88E-03 | 61.4 | 0.07 |
| cg20768760 | 5 | 1,332,573 | 5p15.33 | -11.76 | 6.55E-32 | 0.69 | *CLPTM1L* | Intronic | -9.58 | **9.51E-22** | -8.59 | **8.58E-18** | -3.11 | 1.85E-03 | 54.1 | 0.11 |
| cg21202862 | 5 | 1,337,955 | 5p15.33 | -12.06 | 1.80E-33 | 0.35 | *CLPTM1L* | Intronic | -9.90 | **4.37E-23** | -8.89 | **6.23E-19** | -3.26 | 1.11E-03 | 56.4 | 0.10 |
| cg09508356 | 5 | 1,346,092 | 5p15.33 | -8.10 | 5.71E-16 | 0.34 | *CLPTM1L* | TSS1500 | -6.63 | **3.38E-11** | -5.79 | **7.11E-09** | -1.86 | 0.06 | 28.3 | 0.25 |
| cg21519219 | 5 | 1,723,384 | 5p15.33 | 6.53 | 6.38E-11 | 0.19 | *MIR4277* ; *MRPL36* | Intergenic | 6.18 | **6.27E-10** | 3.73 | 1.89E-04 | 2.18 | 0.03 | 0.0 | 0.38 |
| cg25606641 | 5 | 1,729,927 | 5p15.33 | 6.75 | 1.50E-11 | 0.16 | *MIR4277* ; *MRPL36* | Intergenic | 6.33 | **2.44E-10** | 4.47 | 7.89E-06 | 1.42 | 0.16 | 28.6 | 0.25 |
| cg02110333 | 5 | 1,800,184 | 5p15.33 | 5.39 | 7.07E-08 | 0.19 | *MRPL36* | TSS1500 | 4.41 | 1.06E-05 | 0.88 | 0.38 | 2.49 | 0.01 | 59.5 | 0.08 |
| cg05353884 | 6 | 26,226,203 | 6p22.2 | -6.50 | 8.28E-11 | 0.23 | *HIST1H3E* | Downstream | -3.97 | 7.27E-05 | -5.74 | **9.20E-09** | -3.69 | 2.21E-04 | 55.7 | 0.10 |
| cg17866778 | 6 | 26,233,442 | 6p22.2 | 5.76 | 8.61E-09 | 0.18 | *HIST1H1D* | Downstream | 3.48 | 4.94E-04 | 5.16 | 2.51E-07 | 3.23 | 1.25E-03 | 38.7 | 0.20 |
| cg05868516 | 6 | 26,286,170 | 6p22.2 | -6.36 | 1.96E-10 | 0.42 | *HIST1H4H* | TSS1500 | -3.11 | 1.90E-03 | -5.48 | **4.32E-08** | 1.10 | 0.27 | 66.0 | 0.05 |
| cg13569146 | 6 | 26,330,589 | 6p22.2 | 5.99 | 2.08E-09 | 0.27 | *HIST1H4H* ; *BTN3A2* | Intergenic | 3.32 | 8.85E-04 | 4.56 | 5.08E-06 | NA | NA | 39.6 | 0.20 |
| cg12539606 | 6 | 27,777,856 | 6p22.1 | 5.38 | 7.59E-08 | 0.27 | *HIST1H3H* | Exonic | 1.95 | 0.05 | 4.94 | 7.93E-07 | 2.14 | 0.03 | 65.4 | 0.06 |
| cg01084722 | 6 | 28,483,060 | 6p22.1 | 5.19 | 2.12E-07 | 0.18 | *GPX6* | Intronic | 2.45 | 0.01 | 3.35 | 7.99E-04 | 1.94 | 0.05 | 0.0 | 0.84 |
| cg13866093 | 6 | 28,502,727 | 6p22.1 | 5.19 | 2.15E-07 | 0.36 | *GPX5* | 3'UTR | 3.43 | 6.10E-04 | 3.45 | 5.64E-04 | 2.46 | 0.01 | 0.0 | 0.89 |
| cg15026241 | 6 | 28,784,980 | 6p22.1 | 6.48 | 9.13E-11 | 0.15 | *LINC00533* ; *LINC01623* | Intergenic | 3.24 | 1.20E-03 | 5.51 | **3.63E-08** | 2.62 | 8.80E-03 | 0.0 | 0.77 |
| cg13082535 | 6 | 28,864,756 | 6p22.1 | -5.60 | 2.10E-08 | 0.19 | *HCG14* | Intronic | -3.61 | 3.12E-04 | -5.06 | 4.26E-07 | -2.01 | 0.04 | 25.8 | 0.26 |
| cg23756442 | 6 | 28,892,465 | 6p22.1 | -6.35 | 2.17E-10 | 0.27 | *TRIM27* | TSS1500 | -2.63 | 8.48E-03 | -5.55 | **2.89E-08** | -2.69 | 7.17E-03 | 61.0 | 0.08 |
| cg02057917 | 6 | 29,323,269 | 6p22.1 | -6.25 | 4.03E-10 | 0.16 | *OR5V1* | Exonic | -3.03 | 2.45E-03 | -4.43 | 9.51E-06 | -3.76 | 1.73E-04 | 53.5 | 0.12 |
| cg06308109 | 6 | 29,548,691 | 6p22.1 | 5.39 | 6.93E-08 | 0.26 | *UBD* ; *SNORD32B* | Intergenic | 4.30 | 1.67E-05 | 3.48 | 5.08E-04 | 2.27 | 0.02 | 44.7 | 0.16 |
| cg23742601 | 6 | 29,589,895 | 6p22.1 | 6.02 | 1.70E-09 | 0.27 | *GABBR1* | Exonic | 3.47 | 5.16E-04 | 4.31 | 1.63E-05 | 3.25 | 1.17E-03 | 0.0 | 0.61 |
| cg21100518 | 6 | 29,595,002 | 6p22.1 | -6.06 | 1.35E-09 | 0.21 | *GABBR1* | Intronic | -3.42 | 6.18E-04 | -5.74 | **9.69E-09** | -3.03 | 2.48E-03 | 35.3 | 0.21 |
| cg08862148 | 6 | 29,595,315 | 6p22.1 | -5.55 | 2.87E-08 | 0.20 | *GABBR1* | Exonic | -2.51 | 0.01 | -4.10 | 4.14E-05 | -2.96 | 3.04E-03 | 35.6 | 0.21 |
| cg02014853 | 6 | 29,595,335 | 6p22.1 | -6.63 | 3.30E-11 | 0.20 | *GABBR1* | Exonic | -3.29 | 1.02E-03 | -5.42 | **6.13E-08** | -2.95 | 3.17E-03 | 51.3 | 0.13 |
| cg25880387 | 6 | 29,609,770 | 6p22.1 | 6.79 | 1.14E-11 | 0.25 | *GABBR1* ; *MOG* | Intergenic | 3.24 | 1.21E-03 | 5.62 | **1.86E-08** | 3.19 | 1.43E-03 | 55.3 | 0.11 |
| cg00433866 | 6 | 29,623,646 | 6p22.1 | 5.84 | 5.20E-09 | 0.35 | *GABBR1* ; *MOG* | Intergenic | 3.05 | 2.32E-03 | 4.68 | 2.82E-06 | 2.61 | 9.02E-03 | 24.1 | 0.27 |
| cg13537510 | 6 | 29,623,721 | 6p22.1 | 6.81 | 9.95E-12 | 0.21 | *GABBR1* ; *MOG* | Intergenic | 3.17 | 1.53E-03 | 5.09 | 3.66E-07 | 3.38 | 7.17E-04 | 50.8 | 0.13 |
| cg17977304 | 6 | 29,623,992 | 6p22.1 | 6.10 | 1.05E-09 | 0.31 | *MOG* | TSS1500 | 3.49 | 4.89E-04 | 4.82 | 1.43E-06 | 3.20 | 1.36E-03 | 29.5 | 0.24 |
| cg16118803 | 6 | 29,629,716 | 6p22.1 | 7.02 | 2.28E-12 | 0.62 | *MOG* | Intronic | 3.99 | 6.70E-05 | 4.62 | 3.91E-06 | 3.17 | 1.55E-03 | 0.0 | 0.52 |
| cg05279622 | 6 | 29,629,758 | 6p22.1 | 7.27 | 3.70E-13 | 0.61 | *MOG* | Intronic | 4.08 | 4.59E-05 | 5.11 | 3.24E-07 | 3.76 | 1.72E-04 | 22.2 | 0.28 |
| cg05813221 | 6 | 29,629,988 | 6p22.1 | 7.73 | 1.10E-14 | 0.24 | *MOG* | Intronic | 4.30 | 1.71E-05 | 6.06 | **1.39E-09** | 3.48 | 5.01E-04 | 40.3 | 0.19 |
| cg03147503 | 6 | 29,633,969 | 6p22.1 | 5.94 | 2.87E-09 | 0.51 | *MOG* | Exonic | 3.49 | 4.85E-04 | 3.88 | 1.04E-04 | 2.03 | 0.04 | 0.0 | 0.80 |
| cg24444631 | 6 | 29,636,366 | 6p22.1 | 5.70 | 1.20E-08 | 0.39 | *MOG* | Intronic | 3.05 | 2.30E-03 | 4.24 | 2.27E-05 | 2.10 | 0.04 | 0.0 | 0.48 |
| cg06061002 | 6 | 29,638,918 | 6p22.1 | 6.95 | 3.59E-12 | 0.48 | *MOG* | 3'UTR | 3.39 | 6.88E-04 | 5.47 | **4.51E-08** | 4.09 | 4.36E-05 | 64.9 | 0.06 |
| cg00539542 | 6 | 29,644,544 | 6p22.1 | -7.16 | 8.17E-13 | 0.55 | *ZFP57* | Intronic | -3.71 | 2.10E-04 | -5.07 | 4.00E-07 | -3.98 | 7.03E-05 | 56.3 | 0.10 |
| cg12463578 | 6 | 29,644,756 | 6p22.1 | -7.11 | 1.18E-12 | 0.65 | *ZFP57* | Exonic | -3.45 | 5.51E-04 | -5.21 | **1.89E-07** | -4.00 | 6.40E-05 | 59.9 | 0.08 |
| cg25978138 | 6 | 29,648,161 | 6p22.1 | -7.24 | 4.56E-13 | 0.73 | *ZFP57* | Intronic | -3.75 | 1.76E-04 | -5.34 | **9.17E-08** | -3.90 | 9.50E-05 | 51.9 | 0.13 |
| cg11747594 | 6 | 29,648,225 | 6p22.1 | -6.94 | 3.81E-12 | 0.75 | *ZFP57* | Intronic | -3.35 | 8.14E-04 | -5.38 | **7.55E-08** | -3.90 | 9.77E-05 | 64.3 | 0.06 |
| cg15708526 | 6 | 29,648,271 | 6p22.1 | -6.86 | 6.88E-12 | 0.75 | *ZFP57* | Intronic | -3.46 | 5.43E-04 | -5.25 | **1.50E-07** | -3.65 | 2.58E-04 | 44.2 | 0.17 |
| cg04071440 | 6 | 29,648,275 | 6p22.1 | -7.09 | 1.38E-12 | 0.75 | *ZFP57* | Intronic | -3.58 | 3.46E-04 | -5.34 | **9.43E-08** | -3.90 | 9.58E-05 | 55.7 | 0.10 |
| cg08022281 | 6 | 29,648,345 | 6p22.1 | -7.39 | 1.46E-13 | 0.72 | *ZFP57* | Intronic | -3.77 | 1.62E-04 | -5.39 | **7.22E-08** | -4.04 | 5.27E-05 | 57.7 | 0.09 |
| cg12644888 | 6 | 29,648,360 | 6p22.1 | -6.91 | 4.87E-12 | 0.72 | *ZFP57* | Intronic | -3.39 | 7.06E-04 | -5.28 | **1.27E-07** | -3.95 | 7.94E-05 | 60.2 | 0.08 |
| cg22494932 | 6 | 29,648,379 | 6p22.1 | -6.89 | 5.78E-12 | 0.73 | *ZFP57* | Intronic | -3.27 | 1.08E-03 | -5.39 | **7.12E-08** | -3.78 | 1.58E-04 | 62.9 | 0.07 |
| cg25699073 | 6 | 29,648,381 | 6p22.1 | -7.24 | 4.35E-13 | 0.72 | *ZFP57* | Intronic | -3.97 | 7.14E-05 | -5.04 | 4.56E-07 | -3.89 | 9.92E-05 | 30.1 | 0.24 |
| cg07134666 | 6 | 29,648,400 | 6p22.1 | -6.99 | 2.69E-12 | 0.73 | *ZFP57* | Intronic | -3.31 | 9.27E-04 | -5.51 | **3.66E-08** | -3.87 | 1.09E-04 | 65.1 | 0.06 |
| cg16885113 | 6 | 29,648,507 | 6p22.1 | -6.82 | 9.19E-12 | 0.77 | *ZFP57* | Intronic | -3.15 | 1.61E-03 | -5.42 | **6.01E-08** | -3.89 | 1.00E-04 | 66.3 | 0.05 |
| cg11383134 | 6 | 29,648,590 | 6p22.1 | -6.76 | 1.36E-11 | 0.73 | *ZFP57* | Intronic | -3.25 | 1.14E-03 | -5.27 | **1.38E-07** | -3.80 | 1.44E-04 | 61.4 | 0.08 |
| cg03449857 | 6 | 29,648,623 | 6p22.1 | -6.63 | 3.39E-11 | 0.76 | *ZFP57* | Intronic | -3.28 | 1.02E-03 | -5.20 | **1.96E-07** | -3.80 | 1.45E-04 | 58.9 | 0.09 |
| cg15570656 | 6 | 29,648,628 | 6p22.1 | -6.93 | 4.27E-12 | 0.75 | *ZFP57* | Intronic | -3.36 | 7.91E-04 | -5.33 | **9.60E-08** | -3.84 | 1.25E-04 | 59.8 | 0.08 |
| cg03278703 | 6 | 29,691,107 | 6p22.1 | 6.52 | 7.25E-11 | 0.21 | *HLA-F* | TSS200 | 3.04 | 2.38E-03 | 4.83 | 1.40E-06 | 3.26 | 1.12E-03 | 40.3 | 0.19 |
| cg20927242 | 6 | 29,692,011 | 6p22.1 | -6.62 | 3.71E-11 | 0.23 | *HLA-F* | Exonic | -3.67 | 2.41E-04 | -4.05 | 5.19E-05 | -2.54 | 0.01 | 0.0 | 0.77 |
| cg03536022 | 6 | 29,706,879 | 6p22.1 | -6.50 | 8.21E-11 | 0.51 | *HLA-F-AS1* | Intronic | -3.78 | 1.55E-04 | -4.47 | 7.89E-06 | -3.80 | 1.44E-04 | 22.0 | 0.28 |
| cg11441173 | 6 | 29,707,887 | 6p22.1 | 7.12 | 1.09E-12 | 0.23 | *HLA-F-AS1* | Intronic | 4.45 | 8.61E-06 | 5.17 | **2.30E-07** | 2.98 | 2.86E-03 | 0.0 | 0.45 |
| cg08451469 | 6 | 29,711,588 | 6p22.1 | -6.24 | 4.29E-10 | 0.73 | *HLA-F-AS1* | Intronic | -3.26 | 1.11E-03 | -4.08 | 4.49E-05 | -3.57 | 3.61E-04 | 12.4 | 0.32 |
| cg08560874 | 6 | 29,759,947 | 6p22.1 | 5.63 | 1.80E-08 | 0.72 | *HCG4* ; *HLA-V* | Exonic | 2.30 | 0.02 | 4.29 | 1.78E-05 | 2.94 | 3.26E-03 | 50.0 | 0.14 |
| cg08696107 | 6 | 29,759,949 | 6p22.1 | 5.72 | 1.08E-08 | 0.69 | *HCG4* ; *HLA-V* | Exonic | 2.43 | 0.02 | 4.39 | 1.13E-05 | 2.90 | 3.74E-03 | 47.0 | 0.15 |
| cg13688808 | 6 | 29,760,158 | 6p22.1 | 6.09 | 1.14E-09 | 0.36 | *HCG4* | Exonic | 2.57 | 0.01 | 4.84 | 1.33E-06 | 3.21 | 1.35E-03 | 58.9 | 0.09 |
| cg25107289 | 6 | 29,760,856 | 6p22.1 | 5.30 | 1.13E-07 | 0.27 | *HLA-V* | Intronic | 3.52 | 4.29E-04 | 3.07 | 2.16E-03 | 1.17 | 0.24 | 0.0 | 0.74 |
| cg13143120 | 6 | 29,761,042 | 6p22.1 | -7.62 | 2.48E-14 | 0.19 | *HLA-V* | Intronic | -3.61 | 3.09E-04 | -5.37 | **7.95E-08** | -3.82 | 1.32E-04 | 50.6 | 0.13 |
| cg05973840 | 6 | 29,761,044 | 6p22.1 | -5.64 | 1.74E-08 | 0.43 | *HLA-V* | Intronic | -2.24 | 0.03 | -4.25 | 2.13E-05 | -3.15 | 1.65E-03 | 55.2 | 0.11 |
| cg03432955 | 6 | 29,795,501 | 6p22.1 | -5.88 | 4.09E-09 | 0.82 | *HLA-G* | Intronic | -2.78 | 5.41E-03 | -4.67 | 3.02E-06 | -2.86 | 4.22E-03 | 38.2 | 0.20 |
| cg00126638 | 6 | 29,795,583 | 6p22.1 | -5.50 | 3.83E-08 | 0.81 | *HLA-G* | 5'UTR | -2.55 | 0.01 | -4.38 | 1.20E-05 | -2.81 | 4.93E-03 | 36.1 | 0.21 |
| cg03521696 | 6 | 29,795,595 | 6p22.1 | -5.58 | 2.44E-08 | 0.77 | *HLA-G* | 5'UTR | -2.62 | 8.86E-03 | -4.41 | 1.05E-05 | -2.86 | 4.30E-03 | 38.0 | 0.20 |
| cg03997176 | 6 | 29,796,066 | 6p22.1 | 6.46 | 1.05E-10 | 0.38 | *HLA-G* | Exonic | 3.70 | 2.14E-04 | 4.17 | 3.08E-05 | 2.09 | 0.04 | 0.0 | 0.88 |
| cg19141489 | 6 | 29,796,068 | 6p22.1 | 6.10 | 1.08E-09 | 0.33 | *HLA-G* | Exonic | 3.80 | 1.46E-04 | 3.63 | 2.85E-04 | 1.85 | 0.06 | 0.0 | 0.92 |
| cg11862551 | 6 | 29,817,963 | 6p22.1 | -5.25 | 1.56E-07 | 0.62 | *ZNRD1ASP* ; *HCP5B* | Intergenic | -3.08 | 2.06E-03 | -3.53 | 4.22E-04 | -1.73 | 0.08 | 0.0 | 0.80 |
| cg03863342 | 6 | 29,831,661 | 6p22.1 | -8.01 | 1.13E-15 | 0.69 | *ZNRD1ASP* ; *HCP5B* | Intergenic | -3.83 | 1.26E-04 | -6.01 | **1.82E-09** | -3.91 | 9.33E-05 | 34.0 | 0.22 |
| cg24838316 | 6 | 29,895,260 | 6p22.1 | 6.64 | 3.09E-11 | 0.79 | *HCG4B* | TSS1500 | 3.20 | 1.39E-03 | 5.63 | **1.83E-08** | 2.97 | 3.02E-03 | 38.4 | 0.20 |

| | | | | | | | | | | | | | | | | |
|---|---|---|---|---|---|---|---|---|---|---|---|---|---|---|---|---|
| cg05523662 | 6 | 29,910,101 | 6p22.1 | 5.66 | 1.51E-08 | 0.52 | *HLA-A* | TSS200 | 2.32 | 0.02 | 4.35 | 1.39E-05 | 3.21 | 1.33E-03 | 54.9 | 0.11 |
| cg15319255 | 6 | 29,910,269 | 6p22.1 | -6.29 | 3.27E-10 | 0.68 | *HLA-A* | 5'UTR | -3.81 | 1.38E-04 | -3.42 | 6.18E-04 | -1.94 | 0.05 | 6.8 | 0.34 |
| cg19593490 | 6 | 29,943,188 | 6p22.1 | -6.37 | 1.89E-10 | 0.70 | *HCG9* | Exonic | -3.87 | 1.10E-04 | -4.12 | 3.72E-05 | -1.59 | 0.11 | 31.0 | 0.23 |
| cg00599564 | 6 | 29,943,209 | 6p22.1 | -5.89 | 3.88E-09 | 0.68 | *HCG9* | Exonic | -2.57 | 0.01 | -3.98 | 6.97E-05 | -2.24 | 0.03 | 0.0 | 0.90 |
| cg23244913 | 6 | 29,943,401 | 6p22.1 | -5.19 | 2.07E-07 | 0.71 | *HCG9* | Intronic | -2.88 | 3.94E-03 | -3.01 | 2.64E-03 | -1.69 | 0.09 | 0.0 | 0.98 |
| cg16368146 | 6 | 29,943,425 | 6p22.1 | -5.54 | 3.03E-08 | 0.74 | *HCG9* | Intronic | -2.95 | 3.22E-03 | -3.56 | 3.71E-04 | -1.92 | 0.06 | 0.0 | 0.83 |
| cg17093638 | 6 | 29,944,505 | 6p22.1 | 5.30 | 1.18E-07 | 0.21 | *HCG9* | Intronic | 2.12 | 0.03 | 4.76 | 1.90E-06 | 2.74 | 6.16E-03 | 62.6 | 0.07 |
| cg10432093 | 6 | 29,945,155 | 6p22.1 | -6.17 | 6.95E-10 | 0.49 | *HCG9* | Intronic | -2.94 | 3.27E-03 | -5.12 | 3.02E-07 | -2.21 | 0.03 | 48.0 | 0.15 |
| cg06507043 | 6 | 29,945,728 | 6p22.1 | -7.09 | 1.30E-12 | 0.32 | *HCG9* | Intronic | -3.50 | 4.59E-04 | -5.46 | **4.74E-08** | -3.40 | 6.71E-04 | 31.9 | 0.23 |
| cg04983903 | 6 | 29,974,065 | 6p22.1 | -5.68 | 1.35E-08 | 0.71 | *HLA-J* ; *ZNRD1ASP* | Intronic | -3.07 | 2.13E-03 | -4.06 | 4.89E-05 | -2.42 | 0.02 | 0.0 | 0.54 |
| cg19448822 | 6 | 29,974,773 | 6p22.1 | 5.41 | 6.34E-08 | 0.58 | *HLA-J* | Exonic | 2.78 | 5.44E-03 | 4.06 | 4.91E-05 | 2.59 | 9.60E-03 | 0.0 | 0.56 |
| cg13207333 | 6 | 29,974,798 | 6p22.1 | 6.13 | 9.02E-10 | 0.48 | *HLA-J* | Exonic | 3.02 | 2.51E-03 | 4.63 | 3.61E-06 | 2.47 | 0.01 | 0.0 | 0.48 |
| cg11867546 | 6 | 29,974,810 | 6p22.1 | 8.37 | 5.99E-17 | 0.59 | *HLA-J* | Exonic | 3.79 | 1.49E-04 | 6.75 | **1.46E-11** | 4.22 | 2.46E-05 | 62.8 | 0.07 |
| cg08163199 | 6 | 29,974,858 | 6p22.1 | 5.17 | 2.39E-07 | 0.33 | *HLA-J* ; *ZNRD1ASP* | Intronic | 3.20 | 1.38E-03 | 3.72 | 2.03E-04 | 1.58 | 0.11 | 0.0 | 0.70 |
| cg25318809 | 6 | 29,974,863 | 6p22.1 | 6.33 | 2.52E-10 | 0.32 | *HLA-J* ; *ZNRD1ASP* | Intronic | 2.86 | 4.21E-03 | 5.14 | 2.72E-07 | 2.83 | 4.72E-03 | 40.4 | 0.19 |
| cg14781281 | 6 | 29,974,868 | 6p22.1 | 5.22 | 1.82E-07 | 0.45 | *HLA-J* ; *ZNRD1ASP* | Intronic | 3.02 | 2.49E-03 | 3.50 | 4.67E-04 | 2.16 | 0.03 | 0.0 | 0.86 |
| cg08325845 | 6 | 29,974,886 | 6p22.1 | 6.75 | 1.47E-11 | 0.53 | *HLA-J* ; *ZNRD1ASP* | Intronic | 3.57 | 3.52E-04 | 5.26 | **1.46E-07** | 2.89 | 3.84E-03 | 14.4 | 0.31 |
| cg15726260 | 6 | 29,974,900 | 6p22.1 | 5.94 | 2.80E-09 | 0.34 | *HLA-J* ; *ZNRD1ASP* | Intronic | 3.74 | 1.81E-04 | 4.30 | 1.74E-05 | 2.24 | 0.02 | 0.0 | 0.77 |
| cg12976581 | 6 | 29,975,032 | 6p22.1 | 7.64 | 2.20E-14 | 0.46 | *HLA-J* ; *ZNRD1ASP* | Intronic | 4.27 | 1.97E-05 | 5.88 | **4.22E-09** | 2.92 | 3.46E-03 | 9.1 | 0.33 |
| cg14432143 | 6 | 29,975,061 | 6p22.1 | 6.08 | 1.21E-09 | 0.26 | *HLA-J* ; *ZNRD1ASP* | Intronic | 2.84 | 4.58E-03 | 5.54 | **2.95E-08** | 1.96 | 0.05 | 64.5 | 0.06 |
| cg04211303 | 6 | 29,975,675 | 6p22.1 | -7.53 | 5.07E-14 | 0.33 | *HLA-J* ; *ZNRD1ASP* | Intronic | -3.68 | 2.35E-04 | -5.86 | **4.58E-09** | -3.65 | 2.61E-04 | 54.3 | 0.11 |
| cg05062511 | 6 | 30,028,447 | 6p22.1 | 5.23 | 1.70E-07 | 0.22 | *ZNRD1ASP* | Intronic | 3.26 | 1.11E-03 | 3.59 | 3.37E-04 | 2.52 | 0.01 | 0.0 | 0.77 |
| cg02383089 | 6 | 30,029,002 | 6p22.1 | -7.39 | 1.45E-13 | 0.20 | *ZNRD1* ; *ZNRD1ASP* | TSS200 | -4.21 | 2.55E-05 | -5.56 | **2.71E-08** | -1.67 | 0.09 | 1.4 | 0.36 |
| cg13919341 | 6 | 30,029,929 | 6p22.1 | 6.64 | 3.06E-11 | 0.59 | *ZNRD1* | Intronic | 3.78 | 1.59E-04 | 5.48 | **4.23E-08** | 2.05 | 0.04 | 0.0 | 0.40 |
| cg06155771 | 6 | 30,029,935 | 6p22.1 | 7.38 | 1.63E-13 | 0.61 | *ZNRD1* | Intronic | 4.14 | 3.41E-05 | 5.30 | **1.16E-07** | 2.97 | 3.00E-03 | 0.0 | 0.60 |
| cg07468289 | 6 | 30,030,010 | 6p22.1 | 6.51 | 7.54E-11 | 0.41 | *ZNRD1* | Exonic | 3.52 | 4.36E-04 | 4.47 | 7.77E-06 | 2.58 | 9.86E-03 | 0.0 | 0.82 |
| cg09093485 | 6 | 30,038,882 | 6p22.1 | -5.34 | 9.05E-08 | 0.41 | *RNF39* | 3'UTR | -3.49 | 4.75E-04 | -3.99 | 6.72E-05 | -2.03 | 0.04 | 0.0 | 0.81 |
| cg05827186 | 6 | 30,038,915 | 6p22.1 | -6.10 | 1.06E-09 | 0.26 | *RNF39* | Exonic | -3.24 | 1.19E-03 | -4.58 | 4.68E-06 | -2.38 | 0.02 | 0.0 | 0.48 |
| cg19046933 | 6 | 30,038,922 | 6p22.1 | -7.82 | 5.19E-15 | 0.23 | *RNF39* | Exonic | -4.00 | 6.34E-05 | -6.05 | **1.45E-09** | -3.26 | 1.11E-03 | 30.6 | 0.24 |
| cg03293507 | 6 | 30,038,975 | 6p22.1 | -5.87 | 4.25E-09 | 0.57 | *RNF39* | Exonic | -4.22 | 2.48E-05 | -4.01 | 6.10E-05 | -1.49 | 0.14 | 0.0 | 0.71 |
| cg20249327 | 6 | 30,039,374 | 6p22.1 | -5.77 | 7.91E-09 | 0.55 | *RNF39* | Exonic | -3.95 | 7.93E-05 | -3.78 | 1.59E-04 | -1.08 | 0.28 | 0.0 | 0.44 |
| cg15877520 | 6 | 30,039,376 | 6p22.1 | -5.23 | 1.72E-07 | 0.56 | *RNF39* | Exonic | -3.46 | 5.42E-04 | -3.69 | 2.20E-04 | -1.24 | 0.22 | 0.0 | 0.63 |
| cg13918754 | 6 | 30,039,380 | 6p22.1 | -5.85 | 5.02E-09 | 0.50 | *RNF39* | Exonic | -4.19 | 2.78E-05 | -3.96 | 7.34E-05 | -1.39 | 0.16 | 0.0 | 0.63 |
| cg23752348 | 6 | 30,042,137 | 6p22.1 | 6.04 | 1.56E-09 | 0.29 | *RNF39* | Intronic | 4.03 | 5.60E-05 | 4.12 | 3.75E-05 | 2.00 | 0.04 | 0.0 | 0.70 |
| cg11724134 | 6 | 30,043,727 | 6p22.1 | -7.70 | 1.40E-14 | 0.39 | *RNF39* | TSS200 | -3.82 | 1.33E-04 | -6.48 | **8.88E-11** | -3.44 | 5.79E-04 | 66.0 | 0.05 |
| cg12521832 | 6 | 30,043,737 | 6p22.1 | -7.71 | 1.26E-14 | 0.49 | *RNF39* | TSS200 | -4.24 | 2.27E-05 | -6.20 | **5.50E-10** | -2.96 | 3.12E-03 | 46.6 | 0.15 |
| cg09530011 | 6 | 30,043,818 | 6p22.1 | -7.33 | 2.25E-13 | 0.51 | *RNF39* | TSS200 | -3.47 | 5.14E-04 | -6.21 | **5.46E-10** | -2.31 | 0.02 | 65.1 | 0.06 |
| cg11963660 | 6 | 30,043,820 | 6p22.1 | -7.28 | 3.40E-13 | 0.48 | *RNF39* | TSS200 | -3.63 | 2.81E-04 | -6.11 | **1.00E-09** | -3.30 | 9.76E-04 | 60.1 | 0.08 |
| cg22325294 | 6 | 30,043,829 | 6p22.1 | -5.61 | 2.08E-08 | 0.41 | *RNF39* | TSS1500 | -3.19 | 1.42E-03 | -4.85 | 1.26E-06 | -1.88 | 0.06 | 16.1 | 0.30 |
| cg13169221 | 6 | 30,043,924 | 6p22.1 | -5.28 | 1.26E-07 | 0.48 | *RNF39* | TSS1500 | -3.22 | 1.29E-03 | -3.97 | 7.16E-05 | -1.51 | 0.13 | 0.0 | 0.61 |
| cg15448721 | 6 | 30,045,041 | 6p22.1 | -6.77 | 1.25E-11 | 0.54 | *RNF39* ; *TRIM31* | Intergenic | -3.23 | 1.25E-03 | -5.79 | **6.84E-09** | -2.87 | 4.07E-03 | 58.7 | 0.09 |
| cg21671020 | 6 | 30,068,493 | 6p22.1 | 7.24 | 4.33E-13 | 0.17 | *RNF39* ; *TRIM31* | Intergenic | 4.00 | 6.28E-05 | 5.16 | 2.47E-07 | 2.25 | 0.02 | 0.0 | 0.64 |
| cg23161794 | 6 | 30,068,942 | 6p22.1 | 5.69 | 1.24E-08 | 0.27 | *RNF39; TRIM31* | Intergenic | 3.32 | 9.15E-04 | 3.63 | 2.85E-04 | 3.10 | 1.91E-03 | 0.0 | 0.53 |
| cg04163475 | 6 | 30,069,065 | 6p22.1 | 5.28 | 1.30E-07 | 0.42 | *RNF39* ; *TRIM31* | Intergenic | 2.59 | 9.72E-03 | 4.73 | 2.24E-06 | 2.90 | 3.69E-03 | 51.5 | 0.13 |
| cg19178742 | 6 | 30,070,099 | 6p22.1 | -5.52 | 3.37E-08 | 0.59 | *TRIM31* | Downstream | -2.21 | 0.03 | -4.34 | 1.41E-05 | -3.25 | 1.14E-03 | 59.3 | 0.09 |
| cg15093247 | 6 | 30,071,060 | 6p22.1 | 6.27 | 3.62E-10 | 0.24 | *TRIM31* | 3'UTR | 2.74 | 6.15E-03 | 4.64 | 3.47E-06 | 1.91 | 0.06 | 35.2 | 0.21 |
| cg20797270 | 6 | 30,071,288 | 6p22.1 | 5.49 | 4.05E-08 | 0.45 | *TRIM31* | 3'UTR | 2.36 | 0.02 | 4.67 | 2.97E-06 | 3.45 | 5.56E-04 | 61.8 | 0.07 |
| cg10194160 | 6 | 30,071,387 | 6p22.1 | 6.25 | 4.00E-10 | 0.17 | *TRIM31* | Exonic | 2.31 | 0.02 | 5.04 | 4.66E-07 | 3.19 | 1.42E-03 | 61.2 | 0.08 |
| cg23642250 | 6 | 30,071,394 | 6p22.1 | 5.51 | 3.61E-08 | 0.52 | *TRIM31* | Exonic | 2.24 | 0.03 | 4.73 | 2.21E-06 | 3.31 | 9.45E-04 | 65.1 | 0.06 |
| cg00787996 | 6 | 30,072,616 | 6p22.1 | -6.90 | 5.34E-12 | 0.35 | *TRIM31* | Intronic | -2.71 | 6.77E-03 | -5.61 | **2.02E-08** | -3.30 | 9.56E-04 | 61.5 | 0.07 |
| cg25721950 | 6 | 30,074,078 | 6p22.1 | 7.13 | 1.03E-12 | 0.54 | *TRIM31-AS1* | Intronic | 3.31 | 9.38E-04 | 5.46 | **4.63E-08** | 3.20 | 1.36E-03 | 45.3 | 0.16 |
| cg11229497 | 6 | 30,078,954 | 6p22.1 | 5.70 | 1.16E-08 | 0.72 | *TRIM31-AS1* | Intronic | 3.30 | 9.81E-04 | 3.78 | 1.59E-04 | 2.85 | 4.34E-03 | 0.0 | 0.61 |
| cg13927501 | 6 | 30,079,090 | 6p22.1 | 5.25 | 1.54E-07 | 0.50 | *TRIM31-AS1* | Intronic | 3.49 | 4.77E-04 | 2.82 | 4.78E-03 | 2.49 | 0.01 | 0.0 | 0.88 |
| cg21232488 | 6 | 30,079,203 | 6p22.1 | 5.37 | 7.78E-08 | 0.63 | *TRIM31-AS1* | Intronic | 3.36 | 7.91E-04 | 3.30 | 9.82E-04 | 2.63 | 8.54E-03 | 0.0 | 0.80 |
| cg21809927 | 6 | 30,079,265 | 6p22.1 | 5.52 | 3.38E-08 | 0.61 | *TRIM31-AS1* | Intronic | 3.31 | 9.18E-04 | 3.43 | 6.03E-04 | 2.73 | 6.43E-03 | 0.0 | 0.77 |
| cg20591745 | 6 | 30,079,524 | 6p22.1 | 5.49 | 3.93E-08 | 0.32 | *TRIM31-AS1* | Exonic | 3.56 | 3.70E-04 | 3.43 | 6.12E-04 | 1.53 | 0.13 | 0.0 | 0.58 |
| cg04176169 | 6 | 30,080,264 | 6p22.1 | 8.09 | 6.06E-16 | 0.43 | *TRIM31* | Exonic | 3.60 | 3.22E-04 | 6.26 | **3.87E-10** | 4.05 | 5.22E-05 | 65.6 | 0.05 |
| cg05068206 | 6 | 30,095,195 | 6p22.1 | -5.82 | 5.95E-09 | 0.35 | *TRIM31-AS1* ; *TRIM40* | Intergenic | -3.04 | 2.40E-03 | -4.60 | 4.14E-06 | -1.21 | 0.23 | 34.1 | 0.22 |
| cg11963400 | 6 | 30,112,769 | 6p22.1 | -7.39 | 1.45E-13 | 0.35 | *TRIM40* | Intronic | -3.50 | 4.59E-04 | -5.31 | **1.10E-07** | -3.79 | 1.53E-04 | 50.0 | 0.14 |
| cg14708905 | 6 | 30,113,074 | 6p22.1 | 5.90 | 3.67E-09 | 0.17 | *TRIM40* | Intronic | 2.73 | 6.33E-03 | 3.62 | 2.96E-04 | 3.33 | 8.78E-04 | 5.6 | 0.35 |
| cg08996678 | 6 | 30,113,783 | 6p22.1 | -5.62 | 1.88E-08 | 0.20 | *TRIM40* | Exonic | -2.66 | 7.89E-03 | -4.23 | 2.33E-05 | -3.16 | 1.58E-03 | 42.6 | 0.18 |
| cg12258492 | 6 | 30,114,802 | 6p22.1 | 6.16 | 7.28E-10 | 0.19 | *TRIM40* | Exonic | 2.96 | 3.05E-03 | 4.78 | 1.75E-06 | 3.31 | 9.19E-04 | 42.7 | 0.17 |
| cg00372886 | 6 | 30,126,176 | 6p22.1 | -5.82 | 5.90E-09 | 0.24 | *TRIM10* | Exonic | -3.04 | 2.38E-03 | -3.46 | 5.36E-04 | -2.99 | 2.75E-03 | 0.0 | 0.57 |
| cg04560098 | 6 | 30,127,760 | 6p22.1 | 7.41 | 1.23E-13 | 0.26 | *TRIM10* | Intronic | 4.30 | 1.73E-05 | 5.14 | 2.79E-07 | 1.75 | 0.08 | 0.0 | 0.42 |
| cg27201802 | 6 | 30,138,225 | 6p22.1 | 5.40 | 6.61E-08 | 0.16 | *TRIM15* | Intronic | 2.91 | 3.60E-03 | 3.98 | 6.81E-05 | 3.64 | 2.77E-04 | 49.0 | 0.14 |
| cg16257013 | 6 | 30,167,403 | 6p22.1 | 7.31 | 2.58E-13 | 0.37 | *TRIM26* | Intronic | 3.61 | 3.02E-04 | 6.10 | **1.04E-09** | 2.35 | 0.02 | 60.1 | 0.08 |
| cg18200389 | 6 | 30,170,340 | 6p22.1 | 7.06 | 1.62E-12 | 0.16 | *TRIM26* | Intronic | 3.21 | 1.33E-03 | 5.41 | **6.29E-08** | 3.55 | 3.78E-04 | 62.7 | 0.07 |
| cg03368643 | 6 | 30,181,600 | 6p22.1 | 7.74 | 9.62E-15 | 0.32 | *TRIM26* | TSS1500 | 3.77 | 1.66E-04 | 6.02 | **1.78E-09** | 3.55 | 3.87E-04 | 56.5 | 0.10 |
| cg18241904 | 6 | 30,181,604 | 6p22.1 | 7.76 | 8.65E-15 | 0.19 | *TRIM26* | TSS1500 | 4.64 | 3.47E-06 | 6.56 | **5.53E-11** | 2.96 | 3.10E-03 | 0.0 | 0.37 |
| cg23119604 [c] | 6 | 30,227,854 | 6p22.1 | 5.59 | 2.28E-08 | 0.49 | *HLA-L* | Exonic | 2.15 | 0.03 | 4.89 | 1.00E-06 | 3.16 | 1.56E-03 | 64.3 | 0.06 |
| cg04884612 | 6 | 30,231,639 | 6p22.1 | 7.36 | 1.89E-13 | 0.34 | *HLA-L* | Exonic | 3.59 | 3.35E-04 | 6.20 | **5.75E-10** | 2.48 | 0.01 | 63.2 | 0.07 |
| cg16055914 | 6 | 30,231,887 | 6p22.1 | 5.90 | 3.62E-09 | 0.50 | *HLA-L* | Exonic | 2.99 | 2.80E-03 | 4.31 | 1.67E-05 | 2.52 | 0.01 | 0.0 | 0.44 |
| cg09815377 | 6 | 30,309,236 | 6p22.1 | -5.17 | 2.30E-07 | 0.23 | *TRIM39* ; *TRIM39-RPP21* | Intronic | -2.46 | 0.01 | -4.69 | 2.68E-06 | -2.42 | 0.02 | 57.3 | 0.10 |
| cg21636096 | 6 | 30,313,077 | 6p22.1 | 7.30 | 2.92E-13 | 0.21 | *RPP21* ; *TRIM39-RPP21* | Exonic | 3.32 | 8.92E-04 | 5.42 | **5.80E-08** | 1.94 | 0.05 | 44.9 | 0.16 |
| cg24213773 | 6 | 30,419,201 | 6p21.33 | -6.47 | 9.61E-11 | 0.20 | *RPP21* ; *HLA-E* | Intergenic | -3.83 | 1.29E-04 | -4.86 | 1.18E-06 | -2.50 | 0.01 | 0.0 | 0.53 |
| cg20644330 | 6 | 30,457,133 | 6p21.33 | 5.29 | 1.26E-07 | 0.18 | *HLA-E* | TSS200 | 2.19 | 0.03 | 4.57 | 4.86E-06 | 2.84 | 4.50E-03 | 62.3 | 0.07 |
| cg13092958 | 6 | 30,524,521 | 6p21.33 | 5.99 | 2.08E-09 | 0.17 | *GNL1* ; *PRR3* | 5'UTR | 3.89 | 1.02E-04 | 4.43 | 9.47E-06 | 1.06 | 0.29 | 11.8 | 0.32 |
| cg17762168 | 6 | 30,585,409 | 6p21.33 | 5.46 | 4.72E-08 | 0.15 | *MRPS18B* ; *PPP1R10* | TSS1500 | 3.18 | 1.47E-03 | 3.18 | 1.49E-03 | 1.95 | 0.05 | 0.0 | 1.00 |
| cg04686398 | 6 | 30,683,190 | 6p21.33 | -6.14 | 8.12E-10 | 0.25 | *MDC1* | Intronic | -3.33 | 8.79E-04 | -4.53 | 5.83E-06 | -3.64 | 2.75E-04 | 33.7 | 0.22 |
| cg23897356 | 6 | 30,688,013 | 6p21.33 | 5.88 | 4.18E-09 | 0.16 | *TUBB* | TSS1500 | 4.50 | 6.84E-06 | 4.17 | 3.05E-05 | 1.24 | 0.22 | 0.0 | 0.49 |

| | | | | | | | | | | | | | | | | |
|---|---|---|---|---|---|---|---|---|---|---|---|---|---|---|---|---|
| cg01554625 | 6 | 30,688,579 | 6p21.33 | -6.63 | 3.39E-11 | 0.16 | *TUBB* | Intronic | -3.10 | 1.92E-03 | -5.82 | **5.83E-09** | -2.50 | 0.01 | 62.7 | 0.07 |
| cg20802826 | 6 | 30,782,080 | 6p21.33 | -7.90 | 2.90E-15 | 0.63 | *LINC00243* | Exonic | -4.13 | 3.71E-05 | -6.54 | **6.05E-11** | -3.36 | 7.86E-04 | 36.6 | 0.21 |
| cg19579005 | 6 | 30,782,272 | 6p21.33 | -8.66 | 4.69E-18 | 0.47 | *LINC00243* | Exonic | -4.48 | 7.41E-06 | -7.58 | **3.52E-14** | -3.48 | 5.06E-04 | 43.1 | 0.17 |
| cg12457901 | 6 | 30,882,671 | 6p21.33 | 5.24 | 1.58E-07 | 0.81 | *VARS2* | Exonic | 2.90 | 3.69E-03 | 3.12 | 1.81E-03 | 3.22 | 1.29E-03 | 0.0 | 0.38 |
| cg05680196 | 6 | 30,884,915 | 6p21.33 | 6.50 | 8.14E-11 | 0.38 | *VARS2* | Exonic | 3.57 | 3.59E-04 | 5.82 | **6.03E-09** | 2.82 | 4.86E-03 | 41.3 | 0.18 |
| cg27185298 | 6 | 30,885,205 | 6p21.33 | 5.33 | 9.62E-08 | 0.37 | *VARS2* | Intronic | 2.69 | 7.10E-03 | 4.29 | 1.75E-05 | 3.53 | 4.17E-04 | 23.7 | 0.27 |
| cg26672776 | 6 | 30,899,568 | 6p21.33 | -5.89 | 3.83E-09 | 0.75 | *SFTA2* | Exonic | -2.96 | 3.11E-03 | -4.88 | 1.06E-06 | -3.85 | 1.16E-04 | 63.9 | 0.06 |
| cg11554728 | 6 | 30,921,757 | 6p21.33 | 6.10 | 1.08E-09 | 0.34 | *HCG21* | Intronic | 3.29 | 9.97E-04 | 4.63 | 3.70E-06 | 4.28 | 1.84E-05 | 60.8 | 0.08 |
| cg03059420 | 6 | 30,923,241 | 6p21.33 | 5.75 | 9.09E-09 | 0.54 | *HCG21* | Upstream | 2.67 | 7.64E-03 | 4.31 | 1.63E-05 | 3.80 | 1.42E-04 | 60.9 | 0.08 |
| cg00474080 | 6 | 30,923,243 | 6p21.33 | 5.37 | 8.07E-08 | 0.25 | *HCG21* | Upstream | 2.59 | 9.53E-03 | 3.80 | 1.45E-04 | 3.91 | 9.14E-05 | 61.7 | 0.07 |
| cg00334245 | 6 | 30,923,446 | 6p21.33 | -5.27 | 1.39E-07 | 0.36 | *HCG21* | Upstream | -2.27 | 0.02 | -4.57 | 4.95E-06 | -2.40 | 0.02 | 61.8 | 0.07 |
| cg15878568 | 6 | 30,923,865 | 6p21.33 | 5.24 | 1.62E-07 | 0.71 | *HCG21* ; *MUC21* | Intergenic | 2.43 | 0.02 | 3.85 | 1.19E-04 | 3.78 | 1.54E-04 | 59.8 | 0.08 |
| cg00357273 | 6 | 31,138,611 | 6p21.33 | 5.41 | 6.24E-08 | 0.52 | *POU5F1* | TSS200 | 3.54 | 3.96E-04 | 3.76 | 1.71E-04 | 2.28 | 0.02 | 0.0 | 0.93 |
| cg11805138 | 6 | 31,148,332 | 6p21.33 | -6.00 | 1.98E-09 | 0.51 | *PSORS1C3* | Intronic | -3.31 | 9.29E-04 | -4.97 | 6.67E-07 | -3.71 | 2.07E-04 | 47.4 | 0.15 |
| cg17931227 | 6 | 31,148,370 | 6p21.33 | -5.82 | 6.03E-09 | 0.49 | *PSORS1C3* | Intronic | -3.47 | 5.13E-04 | -4.46 | 8.35E-06 | -3.52 | 4.29E-04 | 12.6 | 0.32 |
| cg09179646 | 6 | 31,148,383 | 6p21.33 | -5.72 | 1.09E-08 | 0.52 | *PSORS1C3* | Intronic | -3.42 | 6.35E-04 | -4.33 | 1.48E-05 | -3.44 | 5.81E-04 | 3.5 | 0.35 |
| cg22291762 | 6 | 31,148,404 | 6p21.33 | -5.99 | 2.06E-09 | 0.50 | *PSORS1C3* | Intronic | -3.57 | 3.53E-04 | -4.81 | 1.50E-06 | -3.45 | 5.69E-04 | 22.2 | 0.28 |
| cg24427850 | 6 | 31,148,409 | 6p21.33 | -5.76 | 8.22E-09 | 0.51 | *PSORS1C3* | Intronic | -3.44 | 5.83E-04 | -4.58 | 4.66E-06 | -3.39 | 6.98E-04 | 16.6 | 0.30 |
| cg26668675 | 6 | 31,148,463 | 6p21.33 | -6.05 | 1.41E-09 | 0.54 | *PSORS1C3* | Intronic | -3.55 | 3.87E-04 | -4.82 | 1.44E-06 | -3.57 | 3.53E-04 | 26.8 | 0.26 |
| cg03078486 | 6 | 31,148,474 | 6p21.33 | -5.86 | 4.52E-09 | 0.52 | *PSORS1C3* | Intronic | -3.51 | 4.52E-04 | -4.64 | 3.55E-06 | -3.41 | 6.50E-04 | 13.3 | 0.32 |
| cg22701603 | 6 | 31,148,483 | 6p21.33 | -5.86 | 4.54E-09 | 0.51 | *PSORS1C3* | Intronic | -3.70 | 2.16E-04 | -4.38 | 1.21E-05 | -3.31 | 9.45E-04 | 0.0 | 0.49 |
| cg26818629 | 6 | 31,148,516 | 6p21.33 | -5.83 | 5.66E-09 | 0.50 | *PSORS1C3* | Intronic | -3.66 | 2.52E-04 | -4.43 | 9.37E-06 | -3.18 | 1.50E-03 | 0.0 | 0.47 |
| cg27547543 | 6 | 31,148,524 | 6p21.33 | -5.84 | 5.09E-09 | 0.52 | *PSORS1C3* | Intronic | -3.29 | 9.96E-04 | -5.25 | **1.48E-07** | -3.78 | 1.57E-04 | 57.6 | 0.09 |
| cg09357589 | 6 | 31,148,552 | 6p21.33 | -6.08 | 1.19E-09 | 0.52 | *PSORS1C3* | Intronic | -3.61 | 3.05E-04 | -4.67 | 3.01E-06 | -3.70 | 2.14E-04 | 23.2 | 0.27 |
| cg14036627 | 6 | 31,148,657 | 6p21.33 | -5.52 | 3.34E-08 | 0.50 | *PSORS1C3* | Intronic | -3.23 | 1.23E-03 | -4.39 | 1.12E-05 | -3.46 | 5.31E-04 | 27.4 | 0.25 |
| cg11811828 | 6 | 31,148,666 | 6p21.33 | -6.23 | 4.81E-10 | 0.51 | *PSORS1C3* | Intronic | -3.40 | 6.79E-04 | -5.06 | 4.26E-07 | -3.53 | 4.22E-04 | 44.2 | 0.17 |
| cg23457357 | 6 | 31,148,748 | 6p21.33 | -6.39 | 1.66E-10 | 0.48 | *PSORS1C3* | Intronic | -3.62 | 2.94E-04 | -5.30 | **1.17E-07** | -3.62 | 2.98E-04 | 43.7 | 0.17 |
| cg03081173 | 6 | 31,166,502 | 6p21.33 | -5.37 | 7.88E-08 | 0.68 | *HCG27* | Intronic | -2.84 | 4.53E-03 | -3.82 | 1.34E-04 | -2.43 | 0.01 | 0.0 | 0.52 |
| cg27536700 | 6 | 31,166,759 | 6p21.33 | -5.64 | 1.70E-08 | 0.65 | *HCG27* | Intronic | -2.96 | 3.05E-03 | -4.94 | 7.76E-07 | -2.39 | 0.02 | 39.3 | 0.19 |
| cg03921216 | 6 | 31,166,799 | 6p21.33 | -5.54 | 3.00E-08 | 0.27 | *HCG27* | Intronic | -3.20 | 1.38E-03 | -4.41 | 1.01E-05 | -1.99 | 0.05 | 0.0 | 0.50 |
| cg10473907 | 6 | 31,167,664 | 6p21.33 | 5.21 | 1.93E-07 | 0.50 | *HCG27* | Intronic | 3.99 | 6.71E-05 | 3.48 | 5.08E-04 | 1.26 | 0.21 | 0.0 | 0.45 |
| cg00218406 | 6 | 31,431,407 | 6p21.33 | -5.47 | 4.45E-08 | 0.16 | *HCP5* | Exonic | -3.51 | 4.46E-04 | -4.44 | 9.12E-06 | -3.19 | 1.43E-03 | 0.0 | 0.42 |
| cg03880509 | 6 | 31,548,599 | 6p21.33 | 5.84 | 5.07E-09 | 0.16 | *LTB* | Exonic | 2.36 | 0.02 | 5.44 | **5.36E-08** | 2.47 | 0.01 | 55.0 | 0.11 |
| cg14661811 | 6 | 31,619,002 | 6p21.33 | -5.46 | 4.77E-08 | 0.31 | *BAG6* | Intronic | -3.38 | 7.24E-04 | -3.77 | 1.62E-04 | -1.24 | 0.21 | 0.0 | 0.74 |
| cg14986464 | 6 | 31,669,902 | 6p21.33 | 5.20 | 1.96E-07 | 0.18 | *ABHD16A* | Exonic | 2.27 | 0.02 | 5.27 | **1.35E-07** | 2.45 | 0.01 | 64.4 | 0.06 |
| cg21769117 | 6 | 31,705,273 | 6p21.33 | 5.88 | 4.18E-09 | 0.26 | *CLIC1* | TSS1500 | 3.18 | 1.46E-03 | 4.95 | 7.47E-07 | 2.41 | 0.02 | 0.0 | 0.48 |
| cg03619586 | 6 | 31,705,409 | 6p21.33 | 8.23 | 1.80E-16 | 0.27 | *CLIC1* | TSS1500 | 4.21 | 2.58E-05 | 6.77 | **1.33E-11** | 3.61 | 3.01E-04 | 63.4 | 0.07 |
| cg16143804 | 6 | 31,706,344 | 6p21.33 | 7.61 | 2.68E-14 | 0.25 | *CLIC1* ; *MSH5* | Intergenic | 4.23 | 2.37E-05 | 6.72 | **1.77E-11** | 2.33 | 0.02 | 64.7 | 0.06 |
| cg01787834 | 6 | 31,762,723 | 6p21.33 | 5.28 | 1.28E-07 | 0.19 | *VARS* | Exonic | 3.48 | 5.06E-04 | 3.58 | 3.39E-04 | 1.65 | 0.10 | 0.0 | 0.86 |
| cg09085842 | 6 | 31,782,961 | 6p21.33 | -5.30 | 1.18E-07 | 0.17 | *HSPA1L* | 5'UTR | -2.48 | 0.01 | -5.18 | **2.23E-07** | -2.38 | 0.02 | 65.8 | 0.05 |
| cg24707219 | 6 | 31,847,009 | 6p21.33 | -5.44 | 5.40E-08 | 0.46 | *SLC44A4* ; *EHMT2* | TSS200 | -4.29 | 1.83E-05 | -2.56 | 0.01 | -0.62 | 0.53 | 20.6 | 0.28 |
| cg13197078 | 6 | 31,963,919 | 6p21.33 | 6.19 | 6.07E-10 | 0.28 | *C4A* ; *C4B* | Intronic | 4.03 | 5.59E-05 | 4.86 | 1.19E-06 | 1.63 | 0.10 | 0.0 | 0.43 |
| cg22575347 | 6 | 31,973,180 | 6p21.33 | 5.60 | 2.18E-08 | 0.38 | *CYP21A1P* | TSS1500 | 3.88 | 1.06E-04 | 3.30 | 9.53E-04 | 0.53 | 0.60 | 0.0 | 0.50 |
| cg11825829 | 6 | 31,997,979 | 6p21.33 | 6.80 | 1.02E-11 | 0.33 | *C4B*; *C4B_2* | Intronic | 4.24 | 2.28E-05 | 5.75 | **9.12E-09** | 2.08 | 0.04 | 6.8 | 0.34 |
| cg20914572 | 6 | 32,119,874 | 6p21.32 | -5.75 | 9.02E-09 | 0.48 | *PRRT1* | Exonic | -3.75 | 1.74E-04 | -3.52 | 4.25E-04 | -1.29 | 0.20 | 0.0 | 0.66 |
| cg11122280 | 6 | 32,123,479 | 6p21.32 | -5.32 | 1.06E-07 | 0.35 | *PPT2* | Exonic | -2.18 | 0.03 | -4.66 | 3.10E-06 | -2.53 | 0.01 | 58.4 | 0.09 |
| cg16809040 | 6 | 32,189,610 | 6p21.32 | 6.70 | 2.09E-11 | 0.19 | *NOTCH4* | Intronic | 3.20 | 1.37E-03 | 5.39 | **6.86E-08** | 3.16 | 1.56E-03 | 4.9 | 0.35 |
| cg01738650 | 6 | 32,223,076 | 6p21.32 | 5.98 | 2.30E-09 | 0.41 | *TSBP1-AS1* | Exonic | 4.44 | 9.16E-06 | 2.86 | 4.25E-03 | 1.00 | 0.32 | 0.0 | 0.39 |
| cg18060330 | 6 | 32,376,066 | 6p21.32 | 5.43 | 5.63E-08 | 0.70 | *TSBP1-AS1* | TSS1500 | 2.35 | 0.02 | 4.94 | 7.72E-07 | 2.05 | 0.04 | 46.6 | 0.15 |
| cg08831072 | 6 | 32,685,319 | 6p21.32 | -6.00 | 2.00E-09 | 0.43 | *HLA-DQB1* ; *HLA-DQA2* | Intergenic | -3.27 | 1.07E-03 | -4.61 | 4.03E-06 | -1.62 | 0.11 | 0.0 | 0.58 |
| cg10123514 | 6 | 32,904,061 | 6p21.32 | 5.50 | 3.77E-08 | 0.30 | *HLA-DMB* | Intronic | 2.83 | 4.72E-03 | 4.35 | 1.38E-05 | 2.65 | 8.09E-03 | 29.8 | 0.24 |
| cg23945333 | 6 | 33,134,078 | 6p21.32 | -5.32 | 1.06E-07 | 0.19 | *COL11A2* | Intronic | -2.64 | 8.30E-03 | -4.92 | 8.52E-07 | -1.76 | 0.08 | 44.0 | 0.17 |
| cg05966420 | 6 | 166,970,431 | 6q27 | -5.30 | 1.14E-07 | 0.27 | *RPS6KA2* | Intronic | -4.08 | 4.44E-05 | -3.17 | 1.52E-03 | -2.78 | 5.39E-03 | 0.0 | 0.76 |
| cg04901951 | 6 | 167,189,174 | 6q27 | 5.49 | 4.05E-08 | 0.22 | *RPS6KA2* | Intronic | 4.12 | 3.83E-05 | 3.48 | 4.93E-04 | 3.02 | 2.50E-03 | 0.0 | 0.76 |
| cg24885369 | 6 | 167,364,028 | 6q27 | 5.56 | 2.76E-08 | 0.24 | *RNASET2* | Intronic | 4.30 | 1.68E-05 | 3.31 | 9.28E-04 | 3.13 | 1.73E-03 | 0.0 | 0.46 |
| cg03583463 | 6 | 167,372,938 | 6q27 | -5.61 | 2.03E-08 | 0.57 | *RNASET2* ; *MIR3939* | Intergenic | -4.22 | 2.45E-05 | -3.27 | 1.08E-03 | -2.93 | 3.35E-03 | 0.0 | 0.79 |
| cg20406979 | 6 | 167,373,233 | 6q27 | -5.48 | 4.36E-08 | 0.62 | *RNASET2* ; *MIR3939* | Intergenic | -4.28 | 1.87E-05 | -3.30 | 9.75E-04 | -2.88 | 4.03E-03 | 0.0 | 0.80 |
| cg18837922 | 6 | 167,391,203 | 6q27 | 5.40 | 6.73E-08 | 0.83 | *RNASET2* ; *MIR3939* | Intergenic | 4.14 | 3.52E-05 | 3.06 | 2.21E-03 | 3.01 | 2.58E-03 | 0.0 | 0.66 |
| cg11749617 | 6 | 167,522,162 | 6q27 | -5.41 | 6.18E-08 | 0.21 | *FGFR1OP* ; *CCR6* | Intergenic | -4.30 | 1.74E-05 | -2.55 | 0.01 | -2.65 | 8.14E-03 | 0.0 | 0.59 |
| cg02953306 | 8 | 27,336,626 | 8p21.2 | 5.16 | 2.43E-07 | 0.47 | *CHRNA2* | 5'UTR | 3.21 | 1.31E-03 | 3.68 | 2.35E-04 | 1.85 | 0.06 | 0.0 | 0.78 |
| cg08029165 | 8 | 27,405,666 | 8p21.1 | 5.21 | 1.92E-07 | 0.46 | *EPHX2* ; *CLU* | Intergenic | 3.67 | 2.43E-04 | 4.28 | 1.88E-05 | 2.59 | 9.61E-03 | 0.0 | 0.65 |
| cg26487031 | 11 | 118,134,801 | 11q23.3 | 5.20 | 2.01E-07 | 0.52 | *MPZL2* | Intronic | 5.25 | **1.53E-07** | 1.63 | 0.10 | 1.78 | 0.07 | 59.3 | 0.09 |
| cg18399703 | 11 | 118,134,846 | 11q23.3 | 5.25 | 1.55E-07 | 0.41 | *MPZL2* | Exonic | 5.28 | **1.27E-07** | 1.84 | 0.07 | 1.60 | 0.11 | 55.4 | 0.11 |
| cg07315693 | 11 | 118,135,203 | 11q23.3 | 5.31 | 1.07E-07 | 0.21 | *MPZL2* | 5'UTR | 5.09 | 3.61E-07 | 1.76 | 0.08 | 1.53 | 0.13 | 52.0 | 0.12 |
| cg11072540 | 15 | 47,566,294 | 15q21.1 | -5.28 | 1.28E-07 | 0.36 | *SEMA6D* | Intronic | -2.94 | 3.27E-03 | -2.69 | 7.23E-03 | -3.59 | 3.28E-04 | 35.3 | 0.21 |
| cg21715896 | 15 | 78,439,388 | 15q25.1 | -7.45 | 9.21E-14 | 0.22 | *CIB2* ; *IDH3A* | Intergenic | -5.41 | **6.38E-08** | -2.91 | 3.57E-03 | -4.10 | 4.13E-05 | 52.7 | 0.12 |
| cg15991190 | 15 | 78,495,720 | 15q25.1 | -11.73 | 8.72E-32 | 0.21 | *ACSBG1* | Intronic | -7.57 | **3.73E-14** | -8.85 | **8.94E-19** | -5.39 | **6.89E-08** | 47.8 | 0.15 |
| cg17086672 | 15 | 78,506,328 | 15q25.1 | -5.66 | 1.52E-08 | 0.27 | *ACSBG1* | Intronic | -3.32 | 8.98E-04 | -4.60 | 4.17E-06 | -2.45 | 0.01 | 8.5 | 0.34 |
| cg01786994 | 15 | 78,556,945 | 15q25.1 | -9.12 | 7.31E-20 | 0.16 | *DNAJA4* | Intronic | -5.57 | **2.57E-08** | -4.84 | 1.30E-06 | -4.01 | 5.98E-05 | 0.0 | 0.64 |
| cg25291474 | 15 | 78,609,546 | 15q25.1 | -6.00 | 1.98E-09 | 0.27 | *WDR61* ; *CRABP1* | Intergenic | -2.39 | 0.02 | -4.10 | 4.14E-05 | -0.25 | 0.80 | 40.4 | 0.19 |
| cg15933925 | 15 | 78,726,587 | 15q25.1 | 6.11 | 1.01E-09 | 0.52 | *CRABP1* ; *IREB2* | Intergenic | 4.43 | 9.21E-06 | 3.04 | 2.38E-03 | 2.22 | 0.03 | 0.0 | 0.84 |
| cg04214706 | 15 | 78,729,669 | 15q25.1 | -18.67 | 8.17E-78 | 0.73 | *IREB2* | TSS1500 | -12.35 | **5.00E-35** | -11.39 | **4.71E-30** | -7.89 | **2.93E-15** | 0.0 | 0.58 |
| cg18825076 | 15 | 78,729,989 | 15q25.1 | -19.69 | 2.40E-86 | 0.80 | *IREB2* | 5'UTR | -12.76 | **2.72E-37** | -12.30 | **9.41E-35** | -8.46 | **2.66E-17** | 7.8 | 0.34 |
| cg08092050 | 15 | 78,734,858 | 15q25.1 | 19.28 | 7.62E-83 | 0.61 | *IREB2* | Intronic | 12.48 | **9.68E-36** | 11.93 | **8.71E-33** | 8.24 | **1.74E-16** | 0.0 | 0.39 |
| cg25138288 | 15 | 78,741,649 | 15q25.1 | -9.90 | 4.23E-23 | 0.28 | *IREB2* | Intronic | -7.06 | **1.66E-12** | -5.59 | **2.22E-08** | -4.15 | 3.36E-05 | 0.0 | 0.85 |
| cg08426200 | 15 | 78,802,280 | 15q25.1 | 13.24 | 5.41E-40 | 0.20 | *HYKK* | Intronic | 9.07 | **1.20E-19** | 8.87 | **7.48E-19** | 5.03 | 5.02E-07 | 0.0 | 0.89 |
| cg06917634 | 15 | 78,832,804 | 15q25.1 | -7.77 | 8.08E-15 | 0.78 | *PSMA4* | 5'UTR | -5.82 | **5.77E-09** | -3.45 | 5.57E-04 | -2.95 | 3.16E-03 | 0.0 | 0.49 |
| cg21291873 | 15 | 78,832,830 | 15q25.1 | -5.72 | 1.06E-08 | 0.38 | *PSMA4* | 5'UTR | -3.11 | 1.87E-03 | -3.82 | 1.34E-04 | -3.48 | 5.04E-04 | 42.5 | 0.18 |
| cg13782957 | 15 | 78,832,906 | 15q25.1 | -8.42 | 3.61E-17 | 0.20 | *PSMA4* | Intronic | -4.60 | 4.30E-06 | -4.76 | 1.98E-06 | -4.17 | 3.07E-05 | 15.2 | 0.31 |
| cg04140906 | 15 | 78,833,505 | 15q25.1 | -11.08 | 1.59E-28 | 0.48 | *PSMA4* | Intronic | -6.81 | **9.70E-12** | -7.97 | **1.62E-15** | -5.08 | 3.78E-07 | 33.6 | 0.22 |
| cg22563815 | 15 | 78,856,949 | 15q25.1 | -7.63 | 2.28E-14 | 0.72 | *CHRNA5* | TSS1500 | -5.82 | **5.88E-09** | -3.37 | 7.55E-04 | -2.88 | 4.03E-03 | 0.0 | 0.46 |
| cg17108064 | 15 | 78,857,060 | 15q25.1 | -8.44 | 3.28E-17 | 0.50 | *CHRNA5* | TSS1500 | -6.30 | **2.93E-10** | -3.88 | 1.03E-04 | -3.10 | 1.94E-03 | 0.0 | 0.50 |
| cg19696491 | 15 | 78,857,124 | 15q25.1 | -7.61 | 2.76E-14 | 0.41 | *CHRNA5* | TSS1500 | -5.72 | **1.06E-08** | -3.41 | 6.40E-04 | -2.81 | 5.00E-03 | 0.0 | 0.52 |
| cg22498099 | 15 | 78,857,571 | 15q25.1 | -7.94 | 1.95E-15 | 0.19 | *CHRNA5* | TSS1500 | -5.82 | **5.98E-09** | -3.81 | 1.38E-04 | -2.89 | 3.81E-03 | 0.0 | 0.67 |
| cg21642963 | 15 | 78,858,002 | 15q25.1 | 18.66 | 1.12E-77 | 0.46 | *CHRNA5* | 5'UTR | 12.68 | **7.96E-37** | 11.21 | **3.46E-29** | 7.88 | **3.15E-15** | 0.0 | 0.71 |
| cg16751781 | 15 | 78,858,589 | 15q25.1 | -6.05 | 1.44E-09 | 0.38 | *CHRNA5* | Intronic | -4.40 | 1.06E-05 | -2.92 | 3.49E-03 | -1.24 | 0.22 | 0.0 | 0.63 |
| cg24031919 | 15 | 78,884,874 | 15q25.1 | -13.73 | 6.83E-43 | 0.32 | *CHRNA5* | Intronic | -9.58 | **1.00E-21** | -7.80 | **6.14E-15** | -5.24 | **1.57E-07** | 0.0 | 0.95 |

| | | | | | | | | | | | | | | | | |
|---|---|---|---|---|---|---|---|---|---|---|---|---|---|---|---|---|
| cg01718004 | 15 | 78,894,581 | 15q25.1 | 11.66 | 2.08E-31 | 0.22 | *CHRNA3* | Exonic | 6.94 | **3.90E-12** | 7.54 | **4.56E-14** | 5.22 | **1.80E-07** | 27.1 | 0.25 |
| cg08862537 | 15 | 78,911,620 | 15q25.1 | -6.25 | 4.18E-10 | 0.52 | *CHRNA3* | Intronic | -4.86 | 1.15E-06 | -2.64 | 8.39E-03 | -1.87 | 0.06 | 0.0 | 0.46 |
| cg07973591 | 15 | 78,911,824 | 15q25.1 | -7.45 | 9.51E-14 | 0.65 | *CHRNA3* | Intronic | -5.71 | **1.16E-08** | -3.37 | 7.58E-04 | -2.46 | 0.01 | 0.0 | 0.49 |
| cg23880581 | 15 | 78,912,387 | 15q25.1 | -6.76 | 1.39E-11 | 0.46 | *CHRNA3* | Intronic | -5.09 | 3.67E-07 | -3.11 | 1.85E-03 | -2.10 | 0.04 | 0.0 | 0.60 |
| cg04882995 | 15 | 78,912,658 | 15q25.1 | 22.67 | 8.23E-114 | 0.44 | *CHRNA3* | Intronic | 15.11 | **1.45E-51** | 14.17 | **1.35E-45** | 9.42 | **4.42E-21** | 0.0 | 0.71 |
| cg25739259 | 15 | 78,913,363 | 15q25.1 | -19.87 | 7.21E-88 | 0.37 | *CHRNA3* | 5URT | -13.71 | **8.74E-43** | -11.71 | **1.18E-31** | -8.34 | **7.22E-17** | 0.0 | 0.72 |
| cg19993258 | 15 | 78,914,264 | 15q25.1 | -13.60 | 3.94E-42 | 0.26 | *CHRNA3* | TSS1500 | -9.70 | **2.98E-22** | -7.60 | **2.88E-14** | -5.30 | **1.19E-07** | 0.0 | 0.85 |
| cg14364242 | 15 | 78,914,367 | 15q25.1 | -7.97 | 1.62E-15 | 0.45 | *CHRNA3* ; *CHRNB4* | Intergenic | -5.74 | **9.70E-09** | -4.10 | 4.11E-05 | -2.82 | 4.75E-03 | 0.0 | 0.81 |
| cg09252970 | 15 | 78,916,632 | 15q25.1 | 8.12 | 4.80E-16 | 0.45 | *CHRNB4* | 3'UTR | 6.27 | **3.50E-10** | 4.09 | 4.28E-05 | 3.08 | 2.07E-03 | 0.0 | 0.56 |
| cg08321268 | 15 | 78,917,214 | 15q25.1 | -5.38 | 7.50E-08 | 0.16 | *CHRNB4* | 3'UTR | -3.56 | 3.77E-04 | -4.58 | 4.60E-06 | -1.77 | 0.08 | 7.8 | 0.34 |
| cg01971720 | 15 | 78,921,123 | 15q25.1 | -7.85 | 4.14E-15 | 0.53 | *CHRNB4* | Intronic | -6.02 | **1.72E-09** | -3.58 | 3.49E-04 | -2.56 | 0.01 | 0.0 | 0.49 |
| cg11412248 | 15 | 78,930,962 | 15q25.1 | -10.00 | 1.53E-23 | 0.35 | *CHRNB4* | Intronic | -5.82 | **5.74E-09** | -7.51 | **6.09E-14** | -3.95 | 7.97E-05 | 65.0 | 0.06 |
| cg13379055 | 15 | 78,942,528 | 15q25.1 | -5.71 | 1.12E-08 | 0.37 | *CHRNB4* ; *LOC646938* | Intergenic | -4.33 | 1.51E-05 | -3.29 | 1.01E-03 | -1.67 | 0.10 | 0.0 | 0.96 |
| cg23236406 | 15 | 78,952,937 | 15q25.1 | -7.41 | 1.28E-13 | 0.16 | *CHRNB4* ; *LOC646938* | Intergenic | -4.96 | 7.10E-07 | -1.56 | 0.12 | -2.93 | 3.37E-03 | 0.0 | 0.73 |
| cg00061860 | 15 | 78,953,028 | 15q25.1 | -7.20 | 5.89E-13 | 0.34 | *CHRNB4* ; *LOC646938* | Intergenic | -4.68 | 2.91E-06 | -0.39 | 0.70 | -2.88 | 3.94E-03 | 0.0 | 0.95 |
| cg01653922 | 15 | 78,965,749 | 15q25.1 | -13.15 | 1.75E-39 | 0.21 | *CHRNB4* ; *LOC646938* | Intergenic | -9.02 | **1.97E-19** | -7.44 | **1.03E-13** | -4.83 | 1.36E-06 | 0.0 | 0.96 |
| cg00727663 | 15 | 78,968,879 | 15q25.1 | -12.30 | 9.00E-35 | 0.26 | *CHRNB4* ; *LOC646938* | Intergenic | -8.14 | **3.88E-16** | -5.55 | **2.84E-08** | -4.21 | 2.50E-05 | 0.0 | 0.52 |
| cg08860612 | 15 | 79,084,223 | 15q25.1 | -10.61 | 2.75E-26 | 0.83 | *ADAMTS7* | Intronic | -7.15 | **8.61E-13** | -6.26 | **3.82E-10** | -3.93 | 8.33E-05 | 0.0 | 0.64 |
| cg22753661 | 15 | 79,092,743 | 15q25.1 | 5.90 | 3.67E-09 | 0.56 | *ADAMTS7* | Exonic | 3.85 | 1.18E-04 | 3.74 | 1.84E-04 | 1.45 | 0.15 | 0.0 | 0.60 |
| cg12645284 | 15 | 79,092,878 | 15q25.1 | 5.98 | 2.26E-09 | 0.53 | *ADAMTS7* | Exonic | 3.86 | 1.14E-04 | 3.70 | 2.12E-04 | 1.52 | 0.13 | 0.0 | 0.79 |
| cg00746121 | 15 | 79,120,042 | 15q25.1 | -8.08 | 6.49E-16 | 0.45 | *ADAMTS7* ; *MORF4L1* | Intergenic | -6.25 | **4.14E-10** | -5.00 | 5.64E-07 | -2.28 | 0.02 | 0.0 | 0.52 |
| cg18432286 | 15 | 79,120,806 | 15q25.1 | -7.32 | 2.43E-13 | 0.18 | *ADAMTS7* ; *MORF4L1* | Intergenic | -5.46 | **4.72E-08** | -4.39 | 1.13E-05 | -2.85 | 4.38E-03 | 0.0 | 0.90 |
| cg05250119 | 15 | 79,123,384 | 15q25.1 | 7.48 | 7.62E-14 | 0.37 | *ADAMTS7* ; *MORF4L1* | Intergenic | 5.13 | 2.92E-07 | 4.95 | 7.24E-07 | 1.83 | 0.07 | 0.0 | 0.44 |
| cg00540400 | 15 | 79,124,168 | 15q25.1 | -8.16 | 3.41E-16 | 0.75 | *ADAMTS7* ; *MORF4L1* | Intergenic | -6.50 | **8.11E-11** | -5.09 | 3.63E-07 | -2.18 | 0.03 | 0.0 | 0.48 |
| cg15822222 | 15 | 79,164,806 | 15q25.1 | -5.59 | 2.30E-08 | 0.62 | *MORF4L1* | TSS1500 | -4.31 | 1.64E-05 | -2.06 | 0.04 | -3.60 | 3.13E-04 | 0.0 | 0.60 |
| cg19318224 | 19 | 41,197,264 | 19q13.2 | 5.44 | 5.23E-08 | 0.71 | *NUMBL* ; *COQ8B* | TSS1500 | 4.53 | 5.93E-06 | 3.45 | 5.62E-04 | 3.04 | 2.34E-03 | 0.0 | 0.72 |
| cg12345610 | 19 | 41,340,569 | 19q13.2 | -6.84 | 8.16E-12 | 0.70 | *CYP2T1P* ; *CYP2A6* | Intergenic | -4.99 | 5.97E-07 | -5.61 | **2.00E-08** | -2.10 | 0.04 | 0.0 | 0.37 |
| cg24067755 | 19 | 41,870,099 | 19q13.2 | 6.09 | 1.16E-09 | 0.19 | *TMEM91* | Intronic | 4.21 | 2.52E-05 | 3.81 | 1.38E-04 | 1.85 | 0.06 | 30.3 | 0.24 |
| cg23159704 | 20 | 62,570,397 | 20q13.33 | 5.23 | 1.72E-07 | 0.48 | *UCKL1* | Downstream | 5.37 | **8.00E-08** | 2.18 | 0.03 | 2.11 | 0.03 | 49.7 | 0.14 |

OR, odds ratio per standard deviation (SD) increase in genetically predicted DNA methylation level; CI, confidence interval; TSS, transcription start site; UTR, untranslated region.

[a] Association Z scores, ORs, 95% CIs, and P values were estimated using SPrediXcan. *P* values that are less than 2.45E-07 (Bonferroni-corrected threshold for 204,339 tested CpGs) are highlighted in bold. All statistical tests were two-sided.

[b] Correlation coefficients of predicted and measured DNA methylation levels.

[c] CpGs that are >1Mb away from any GWAS-identified SNPs and remain significantly associated with lung cancer risk after conditioning on nearest GWAS signals.

**Supplementary Table 5. Associations with risks of ovarian cancer subtypes for CpGs showing a significant association with overall ovarian cancer risk**

| CpG | Chr | Pos | Cytoband | Z score [a] | *P* [a] | R [b] | *Closest gene* | Classification | Clear Cell Ovarian Carcinoma | | Endometrioid Ovarian Carcinoma | | Mucinous Carcinoma | | Serous Ovrian Carcinoma | | $I^2$ | Cochran's *P* |
|---|---|---|---|---|---|---|---|---|---|---|---|---|---|---|---|---|---|---|
| | | | | | | | | | Z score [a] | *P* [a] | Z score [a] | *P* [a] | Z score [a] | *P* [a] | Z score [a] | *P* [a] | | |
| **Associations showing a significant heterogeneity across subtypes (Cochran's *P* <2.60E-07[0.05/192,188; Bonferroni correction])** | | | | | | | | | | | | | | | | | | |
| cg23901857 | 2 | 176,993,221 | 2q31.1 | -7.37 | 1.67E-13 | 0.25 | *HOXD9* ; *HOXD8* | Intergenic | 1.46 | 0.14 | -3.07 | 2.13E-03 | -7.71 | **1.22E-14** | -7.46 | **8.69E-14** | 91.1 | **2.26E-07** |
| cg15617032 | 17 | 44,020,778 | 17q21.31 | 8.04 | 9.31E-16 | 0.56 | *MAPT* | Intronic | 0.34 | 0.74 | 3.13 | 1.73E-03 | 2.41 | 0.02 | 7.33 | **2.32E-13** | 92.9 | **3.25E-09** |
| cg24781419 | 19 | 17,371,555 | 19p13.11 | 6.13 | 9.02E-10 | 0.46 | *USHBP1* | Intronic | -0.96 | 0.33 | -0.20 | 0.85 | -0.85 | 0.39 | 7.57 | **3.65E-14** | 91.1 | **2.17E-07** |
| cg26667659 | 19 | 17,377,598 | 19p13.11 | 6.74 | 1.59E-11 | 0.24 | *BABAM1* | TSS1500 | -0.71 | 0.48 | -1.00 | 0.32 | -1.20 | 0.23 | 8.44 | **3.12E-17** | 93.2 | **1.60E-09** |
| cg20372155 | 19 | 17,377,600 | 19p13.11 | 6.63 | 3.33E-11 | 0.44 | *BABAM1* | TSS1500 | -0.81 | 0.42 | -1.01 | 0.31 | -1.29 | 0.20 | 8.38 | **5.51E-17** | 93.2 | **1.31E-09** |
| cg03745160 | 19 | 17,393,902 | 19p13.11 | 7.70 | 1.37E-14 | 0.33 | *ANKLE1* | Intronic | 1.57 | 0.12 | -0.73 | 0.47 | -0.78 | 0.43 | 9.32 | **1.12E-20** | 92.7 | **5.66E-09** |
| cg12247222 | 19 | 17,403,313 | 19p13.11 | 7.83 | 4.82E-15 | 0.22 | *ABHD8* | UTR3 | 0.07 | 0.94 | -1.31 | 0.19 | -0.98 | 0.33 | 9.59 | **9.15E-22** | 94.2 | **4.17E-11** |
| cg06952291 | 19 | 17,414,123 | 19p13.11 | -5.69 | 1.24E-08 | 0.27 | *ABHD8* | UTR5 | 1.29 | 0.20 | 1.34 | 0.18 | 1.01 | 0.31 | -7.39 | **1.47E-13** | 92.4 | **1.50E-08** |
| **Associations showing a nominal significant heterogeneity across subtypes (2.60E-07<Cochran's *P* <0.05)** | | | | | | | | | | | | | | | | | | |
| cg11597131 | 2 | 176,988,939 | 2q31.1 | -6.25 | 4.06E-10 | 0.40 | *HOXD9* | UTR3 | 0.40 | 0.69 | -2.95 | 3.15E-03 | -6.25 | **4.17E-10** | -6.29 | **3.12E-10** | 83.2 | 4.61E-04 |
| cg09173768 | 2 | 176,989,349 | 2q31.1 | -7.09 | 1.36E-12 | 0.22 | *HOXD9* | UTR3 | 1.24 | 0.21 | -3.33 | 8.77E-04 | -7.15 | **8.39E-13** | -7.36 | **1.90E-13** | 89.0 | 5.25E-06 |
| cg19373813 | 2 | 176,989,586 | 2q31.1 | -6.43 | 1.30E-10 | 0.43 | *HOXD9* | UTR3 | 1.18 | 0.24 | -3.06 | 2.22E-03 | -6.69 | **2.16E-11** | -6.72 | **1.77E-11** | 87.4 | 2.75E-05 |
| cg19412900 | 2 | 176,990,595 | 2q31.1 | -7.60 | 3.03E-14 | 0.32 | *HOXD9* | Downstream | 1.03 | 0.30 | -3.97 | 7.31E-05 | -7.63 | **2.36E-14** | -7.68 | **1.64E-14** | 89.4 | 2.98E-06 |
| cg00035316 | 2 | 176,993,017 | 2q31.1 | -7.01 | 2.31E-12 | 0.30 | *HOXD9* ; *HOXD8* | Intergenic | 0.76 | 0.45 | -3.52 | 4.39E-04 | -6.82 | **8.88E-12** | -7.12 | **1.10E-12** | 85.8 | 1.03E-04 |
| cg18448949 | 2 | 176,993,089 | 2q31.1 | -5.96 | 2.47E-09 | 0.24 | *HOXD9* ; *HOXD8* | Intergenic | 1.13 | 0.26 | -2.50 | 0.01 | -6.19 | **5.99E-10** | -6.28 | **3.34E-10** | 85.6 | 1.15E-04 |
| cg03321133 | 2 | 176,993,115 | 2q31.1 | -6.62 | 3.64E-11 | 0.36 | *HOXD9* ; *HOXD8* | Intergenic | 0.65 | 0.52 | -2.89 | 3.85E-03 | -6.98 | **2.93E-12** | -6.63 | **3.25E-11** | 86.0 | 8.86E-05 |
| cg01293179 | 2 | 176,996,285 | 2q31.1 | -6.57 | 4.91E-11 | 0.43 | *HOXD8* | Exonic | 0.43 | 0.67 | -3.39 | 6.96E-04 | -6.65 | **2.83E-11** | -6.49 | **8.71E-11** | 81.9 | 8.63E-04 |
| cg07783843 | 2 | 176,997,311 | 2q31.1 | -7.54 | 4.74E-14 | 0.44 | *HOXD8* | UTR3 | 1.02 | 0.31 | -4.01 | 6.10E-05 | -7.56 | **3.97E-14** | -7.55 | **4.25E-14** | 89.2 | 4.02E-06 |
| cg21932416 | 2 | 176,997,707 | 2q31.1 | -6.19 | 6.12E-10 | 0.44 | *HOXD8* | Downstream | 1.21 | 0.23 | -2.91 | 3.59E-03 | -6.40 | **1.59E-10** | -6.18 | **6.53E-10** | 86.8 | 4.43E-05 |
| cg06011461 | 2 | 177,002,813 | 2q31.1 | -7.55 | 4.27E-14 | 0.54 | *LOC401021* | Intronic | 1.01 | 0.31 | -3.94 | 8.25E-05 | -7.50 | **6.55E-14** | -7.65 | **2.07E-14** | 89.0 | 4.96E-06 |
| cg12721835 | 2 | 177,002,820 | 2q31.1 | -7.55 | 4.28E-14 | 0.45 | *LOC401021* | Intronic | 0.96 | 0.34 | -3.91 | 9.15E-05 | -7.53 | **5.18E-14** | -7.66 | **1.93E-14** | 88.9 | 5.53E-06 |
| cg10503083 | 2 | 177,003,280 | 2q31.1 | -7.15 | 8.97E-13 | 0.30 | *LOC401021* | Intronic | 1.30 | 0.19 | -3.97 | 7.16E-05 | -7.12 | **1.11E-12** | -7.19 | **6.42E-13** | 89.3 | 3.44E-06 |
| cg08182501 | 2 | 177,003,406 | 2q31.1 | -7.47 | 7.94E-14 | 0.52 | *LOC401021* | Intronic | 0.99 | 0.32 | -3.97 | 7.06E-05 | -7.52 | **5.32E-14** | -7.49 | **6.66E-14** | 89.0 | 4.92E-06 |
| cg06896987 | 2 | 177,003,735 | 2q31.1 | -5.78 | 7.37E-09 | 0.56 | *LOC401021* | Exonic | 0.18 | 0.86 | -2.45 | 0.01 | -6.32 | **2.62E-10** | -5.67 | **1.44E-08** | 82.5 | 6.46E-04 |
| cg11248896 | 2 | 177,003,747 | 2q31.1 | -5.98 | 2.23E-09 | 0.52 | *LOC401021* | Exonic | 0.40 | 0.69 | -2.61 | 9.13E-03 | -6.58 | **4.66E-11** | -5.89 | **3.79E-09** | 84.5 | 2.33E-04 |
| cg11375458 | 2 | 177,003,996 | 2q31.1 | -5.79 | 6.91E-09 | 0.46 | *LOC401021* | Exonic | 0.43 | 0.67 | -2.54 | 0.01 | -5.90 | **3.63E-09** | -5.67 | **1.42E-08** | 80.5 | 1.53E-03 |
| cg18516557 | 2 | 177,004,015 | 2q31.1 | -6.22 | 4.88E-10 | 0.54 | *LOC401021* | Exonic | 0.75 | 0.46 | -3.04 | 2.37E-03 | -6.48 | **9.26E-11** | -6.18 | **6.47E-10** | 85.1 | 1.57E-04 |
| cg21818470 | 2 | 177,004,975 | 2q31.1 | -7.13 | 1.02E-12 | 0.25 | *LOC401021* | Exonic | 0.44 | 0.66 | -3.67 | 2.44E-04 | -7.13 | **9.88E-13** | -7.00 | **2.58E-12** | 85.9 | 9.14E-05 |
| cg01473155 | 2 | 177,010,984 | 2q31.1 | -7.32 | 2.50E-13 | 0.41 | *LOC401021* ; *MIR10B* | Intergenic | 1.06 | 0.29 | -3.76 | 1.69E-04 | -7.48 | **7.47E-14** | -7.31 | **2.64E-13** | 89.3 | 3.73E-06 |
| cg14298457 | 2 | 177,012,117 | 2q31.1 | -7.46 | 8.43E-14 | 0.40 | *LOC401021* ; *MIR10B* | Intergenic | 1.02 | 0.31 | -4.05 | 5.17E-05 | -7.54 | **4.88E-14** | -7.47 | **7.79E-14** | 89.2 | 4.25E-06 |
| cg22344631 | 2 | 177,012,133 | 2q31.1 | -6.54 | 6.19E-11 | 0.44 | *LOC401021* ; *MIR10B* | Intergenic | 0.81 | 0.42 | -2.76 | 5.74E-03 | -6.22 | **4.86E-10** | -6.68 | **2.36E-11** | 83.9 | 3.24E-04 |
| cg04194947 | 2 | 177,012,191 | 2q31.1 | -7.21 | 5.49E-13 | 0.37 | *LOC401021* ; *MIR10B* | Intergenic | 0.99 | 0.32 | -3.79 | 1.51E-04 | -7.47 | **8.14E-14** | -7.23 | **4.80E-13** | 89.0 | 5.36E-06 |
| cg15990972 | 2 | 177,012,372 | 2q31.1 | -7.22 | 5.02E-13 | 0.35 | *LOC401021* ; *MIR10B* | Intergenic | 0.94 | 0.35 | -3.58 | 3.48E-04 | -7.42 | **1.14E-13** | -7.38 | **1.57E-13** | 88.9 | 6.06E-06 |
| cg02857760 | 2 | 177,012,562 | 2q31.1 | -7.06 | 1.64E-12 | 0.57 | *LOC401021* ; *MIR10B* | Intergenic | 1.23 | 0.22 | -3.87 | 1.11E-04 | -7.19 | **6.42E-13** | -7.21 | **5.44E-13** | 89.0 | 5.11E-06 |
| cg17818471 | 2 | 177,012,640 | 2q31.1 | -7.55 | 4.50E-14 | 0.53 | *LOC401021* ; *MIR10B* | Intergenic | 1.03 | 0.30 | -4.04 | 5.41E-05 | -7.70 | **1.40E-14** | -7.56 | **4.02E-14** | 89.6 | 2.34E-06 |
| cg01043845 | 2 | 177,013,071 | 2q31.1 | -7.59 | 3.28E-14 | 0.46 | *LOC401021* ; *MIR10B* | Intergenic | 1.03 | 0.30 | -4.01 | 5.95E-05 | -7.69 | **1.52E-14** | -7.63 | **2.40E-14** | 89.6 | 2.47E-06 |
| cg24000528 | 2 | 177,013,630 | 2q31.1 | -7.40 | 1.37E-13 | 0.42 | *LOC401021* ; *MIR10B* | Intergenic | 1.00 | 0.32 | -3.78 | 1.56E-04 | -7.13 | **1.04E-12** | -7.52 | **5.43E-14** | 87.8 | 1.82E-05 |
| cg23460578 | 2 | 177,014,009 | 2q31.1 | -5.84 | 5.30E-09 | 0.31 | *LOC401021* ; *MIR10B* | Intergenic | 0.94 | 0.35 | -2.73 | 6.30E-03 | -5.21 | **1.84E-07** | -6.02 | **1.79E-09** | 79.5 | 2.16E-03 |
| cg19519747 | 2 | 177,014,555 | 2q31.1 | -6.88 | 6.14E-12 | 0.44 | *MIR10B* | TSS1500 | 1.26 | 0.21 | -3.37 | 7.52E-04 | -6.80 | **1.03E-11** | -7.14 | **9.47E-13** | 87.3 | 2.94E-05 |
| cg07011913 | 2 | 177,014,626 | 2q31.1 | -7.26 | 4.00E-13 | 0.54 | *MIR10B* | TSS1500 | 1.11 | 0.27 | -3.55 | 3.90E-04 | -7.37 | **1.67E-13** | -7.44 | **9.99E-14** | 89.1 | 4.34E-06 |
| cg12127282 | 2 | 177,014,685 | 2q31.1 | -5.51 | 3.56E-08 | 0.43 | *MIR10B* ; *HOXD4* | TSS1500 | 1.28 | 0.20 | -2.44 | 0.01 | -4.90 | 9.62E-07 | -5.79 | **7.09E-09** | 80.2 | 1.72E-03 |
| cg08717880 | 2 | 177,014,849 | 2q31.1 | -7.19 | 6.65E-13 | 0.40 | *MIR10B* | TSS200 | 0.92 | 0.36 | -3.65 | 2.62E-04 | -6.50 | **7.81E-11** | -7.20 | **6.15E-13** | 85.4 | 1.32E-04 |
| cg27160395 | 2 | 177,014,949 | 2q31.1 | -7.60 | 2.88E-14 | 0.44 | *MIR10B* | TSS200 | 1.04 | 0.30 | -3.98 | 6.90E-05 | -7.67 | **1.76E-14** | -7.67 | **1.71E-14** | 89.6 | 2.55E-06 |
| cg17104824 | 2 | 177,014,959 | 2q31.1 | -5.92 | 3.20E-09 | 0.44 | *MIR10B* | TSS200 | 1.57 | 0.12 | -3.16 | 1.59E-03 | -6.48 | **9.31E-11** | -6.10 | **1.07E-09** | 88.5 | 8.83E-06 |
| cg00014998 | 2 | 177,014,962 | 2q31.1 | -7.37 | 1.65E-13 | 0.49 | *MIR10B* | TSS200 | 1.01 | 0.31 | -3.56 | 3.73E-04 | -7.20 | **6.11E-13** | -7.56 | **4.11E-14** | 88.1 | 1.33E-05 |
| cg25942990 | 2 | 177,014,996 | 2q31.1 | -7.31 | 2.76E-13 | 0.57 | *MIR10B* | TSS200 | 0.99 | 0.32 | -3.85 | 1.17E-04 | -7.06 | **1.65E-12** | -7.30 | **2.81E-13** | 87.7 | 2.06E-05 |
| cg01152019 | 2 | 177,015,044 | 2q31.1 | -7.65 | 1.98E-14 | 0.55 | *MIR10B* | Exonic | 0.89 | 0.37 | -4.00 | 6.39E-05 | -7.37 | **1.73E-13** | -7.72 | **1.15E-14** | 88.2 | 1.21E-05 |
| cg14399060 | 2 | 177,015,070 | 2q31.1 | -7.60 | 3.01E-14 | 0.53 | *MIR10B* | Exonic | 1.08 | 0.28 | -3.87 | 1.08E-04 | -7.52 | **5.51E-14** | -7.72 | **1.18E-14** | 89.3 | 3.63E-06 |
| cg00767581 | 2 | 177,015,125 | 2q31.1 | -7.52 | 5.69E-14 | 0.62 | *MIR10B* | Exonic | 1.02 | 0.31 | -3.92 | 8.92E-05 | -7.45 | **9.21E-14** | -7.63 | **2.32E-14** | 88.9 | 5.53E-06 |
| cg19478682 | 2 | 177,015,812 | 2q31.1 | -7.23 | 4.66E-13 | 0.52 | *HOXD4* | TSS1500 | 0.87 | 0.39 | -3.99 | 6.60E-05 | -7.26 | **4.02E-13** | -7.25 | **4.31E-13** | 87.8 | 1.80E-05 |
| cg20739994 | 2 | 177,015,844 | 2q31.1 | -7.33 | 2.32E-13 | 0.43 | *HOXD4* | TSS1500 | 0.84 | 0.40 | -4.06 | 4.91E-05 | -6.94 | **3.93E-12** | -7.22 | **5.19E-13** | 87.1 | 3.51E-05 |
| cg21011616 | 2 | 177,015,992 | 2q31.1 | -7.50 | 6.41E-14 | 0.59 | *HOXD4* | TSS200 | 1.08 | 0.28 | -3.85 | 1.18E-04 | -7.57 | **3.78E-14** | -7.61 | **2.73E-14** | 89.4 | 2.97E-06 |
| cg09583532 | 2 | 177,015,999 | 2q31.1 | -7.56 | 3.94E-14 | 0.53 | *HOXD4* | TSS200 | 1.00 | 0.32 | -4.02 | 5.76E-05 | -7.69 | **1.47E-14** | -7.56 | **3.88E-14** | 89.5 | 2.86E-06 |
| cg04843067 | 2 | 177,016,037 | 2q31.1 | -6.96 | 3.42E-12 | 0.56 | *HOXD4* | TSS200 | 0.85 | 0.39 | -3.50 | 4.61E-04 | -7.16 | **8.03E-13** | -7.03 | **2.04E-12** | 87.7 | 2.17E-05 |
| cg12539415 | 2 | 177,016,167 | 2q31.1 | -7.51 | 5.69E-14 | 0.64 | *HOXD4* | UTR5 | 1.08 | 0.28 | -3.97 | 7.14E-05 | -7.73 | **1.09E-14** | -7.55 | **4.24E-14** | 89.8 | 1.72E-06 |
| cg18750960 | 2 | 177,016,417 | 2q31.1 | -7.55 | 4.46E-14 | 0.57 | *HOXD4* | Exonic | 1.01 | 0.31 | -3.98 | 6.78E-05 | -7.64 | **2.19E-14** | -7.57 | **3.64E-14** | 89.4 | 3.00E-06 |
| cg19509393 | 2 | 177,016,490 | 2q31.1 | -7.57 | 3.70E-14 | 0.61 | *HOXD4* | Exonic | 1.04 | 0.30 | -4.00 | 6.25E-05 | -7.67 | **1.69E-14** | -7.62 | **2.59E-14** | 89.6 | 2.53E-06 |
| cg25951210 | 2 | 177,016,619 | 2q31.1 | -7.55 | 4.30E-14 | 0.67 | *HOXD4* | Exonic | 1.01 | 0.31 | -3.95 | 7.78E-05 | -7.52 | **5.58E-14** | -7.64 | **2.13E-14** | 89.1 | 4.71E-06 |
| cg10803577 | 2 | 177,016,837 | 2q31.1 | -7.62 | 2.53E-14 | 0.66 | *HOXD4* | Intronic | 1.03 | 0.31 | -3.89 | 1.02E-04 | -7.67 | **1.70E-14** | -7.70 | **1.33E-14** | 89.5 | 2.83E-06 |
| cg09981233 | 2 | 177,016,899 | 2q31.1 | -7.58 | 3.42E-14 | 0.65 | *HOXD4* | Intronic | 1.03 | 0.30 | -4.00 | 6.30E-05 | -7.68 | **1.63E-14** | -7.63 | **2.35E-14** | 89.6 | 2.50E-06 |
| cg26654148 | 2 | 177,016,978 | 2q31.1 | -7.62 | 2.44E-14 | 0.60 | *HOXD4* | Intronic | 1.01 | 0.31 | -3.89 | 1.01E-04 | -7.62 | **2.45E-14** | -7.71 | **1.25E-14** | 89.4 | 3.32E-06 |
| cg25304107 | 2 | 177,017,331 | 2q31.1 | -7.42 | 1.15E-13 | 0.65 | *HOXD4* | Intronic | 1.17 | 0.24 | -3.59 | 3.25E-04 | -7.44 | **9.75E-14** | -7.59 | **3.28E-14** | 89.1 | 4.70E-06 |
| cg16218721 | 2 | 177,017,352 | 2q31.1 | -7.60 | 3.05E-14 | 0.49 | *HOXD4* | Exonic | 1.03 | 0.30 | -3.97 | 7.11E-05 | -7.64 | **2.23E-14** | -7.67 | **1.70E-14** | 89.5 | 2.89E-06 |
| cg26590744 | 2 | 177,017,389 | 2q31.1 | -7.09 | 1.32E-12 | 0.62 | *HOXD4* | Exonic | 1.16 | 0.25 | -3.30 | 9.75E-04 | -6.85 | **7.62E-12** | -7.35 | **1.94E-13** | 87.4 | 2.81E-05 |
| cg01414185 | 2 | 177,017,449 | 2q31.1 | -7.57 | 3.62E-14 | 0.62 | *HOXD4* | Exonic | 1.03 | 0.30 | -4.01 | 6.11E-05 | -7.68 | **1.59E-14** | -7.62 | **2.62E-14** | 89.6 | 2.47E-06 |
| cg14900124 | 2 | 177,017,630 | 2q31.1 | -7.32 | 2.41E-13 | 0.45 | *HOXD4* | Exonic | 0.89 | 0.38 | -3.67 | 2.45E-04 | -7.36 | **1.89E-13** | -7.30 | **2.93E-13** | 88.3 | 1.08E-05 |
| cg09701582 | 2 | 177,017,797 | 2q31.1 | -7.60 | 2.90E-14 | 0.46 | *HOXD4* | UTR3 | 1.03 | 0.30 | -3.98 | 6.97E-05 | -7.64 | **2.22E-14** | -7.68 | **1.62E-14** | 89.5 | 2.92E-06 |
| cg14940165 | 2 | 177,021,702 | 2q31.1 | -6.87 | 6.40E-12 | 0.36 | *HOXD3* | Upstream | 1.24 | 0.22 | -3.45 | 5.52E-04 | -6.86 | **7.07E-12** | -7.01 | **2.43E-12** | 88.4 | 1.05E-05 |
| cg26091021 | 2 | 177,021,826 | 2q31.1 | -7.33 | 2.36E-13 | 0.49 | *HOXD3* | Upstream | 1.00 | 0.32 | -3.76 | 1.70E-04 | -7.19 | **6.40E-13** | -7.41 | **1.25E-13** | 88.4 | 1.01E-05 |
| cg25708328 | 2 | 177,022,056 | 2q31.1 | -7.04 | 1.96E-12 | 0.48 | *HOXD3* | UTR5 | 0.99 | 0.32 | -3.78 | 1.59E-04 | -6.37 | **1.88E-10** | -7.33 | **2.38E-13** | 85.2 | 1.51E-04 |
| cg25137403 | 2 | 177,022,172 | 2q31.1 | -7.53 | 5.11E-14 | 0.43 | *HOXD3* | UTR5 | 1.04 | 0.30 | -4.00 | 6.28E-05 | -7.66 | **1.82E-14** | -7.54 | **4.82E-14** | 89.6 | 2.57E-06 |
| cg14454907 | 2 | 177,022,285 | 2q31.1 | -7.10 | 1.28E-12 | 0.51 | *HOXD3* | Intronic | 1.16 | 0.25 | -3.69 | 2.29E-04 | -7.06 | **1.70E-12** | -7.23 | **4.84E-13** | 88.4 | 1.02E-05 |
| cg04201253 | 2 | 177,022,517 | 2q31.1 | -7.14 | 9.20E-13 | 0.51 | *HOXD3* | Intronic | 1.19 | 0.24 | -3.74 | 1.82E-04 | -7.16 | **8.35E-13** | -7.24 | **4.39E-13** | 88.7 | 7.37E-06 |
| cg05525812 | 2 | 177,022,646 | 2q31.1 | -6.91 | 4.86E-12 | 0.55 | *HOXD3* | Intronic | 1.29 | 0.20 | -3.54 | 4.06E-04 | -6.89 | **5.42E-12** | -7.09 | **1.33E-12** | 87.7 | 2.00E-05 |
| cg26821745 | 2 | 177,023,183 | 2q31.1 | -7.22 | 5.02E-13 | 0.50 | *HOXD3* | Intronic | 1.20 | 0.23 | -3.28 | 1.03E-03 | -7.20 | **5.90E-13** | -7.50 | **6.15E-14** | 89.1 | 4.41E-06 |
| cg18944451 | 2 | 177,023,716 | 2q31.1 | -6.75 | 1.48E-11 | 0.53 | *HOXD3* | Intronic | 1.10 | 0.27 | -3.28 | 1.04E-03 | -6.56 | **5.44E-11** | -7.06 | **1.66E-12** | 85.7 | 1.04E-04 |
| cg25916282 | 2 | 177,023,760 | 2q31.1 | -7.58 | 3.44E-14 | 0.42 | *HOXD3* | Intronic | 1.03 | 0.30 | -4.00 | 6.47E-05 | -7.66 | **1.85E-14** | -7.64 | **2.25E-14** | 89.5 | 2.65E-06 |
| cg00223681 | 2 | 177,024,117 | 2q31.1 | -6.67 | 2.59E-11 | 0.44 | *HOXD3* | Intronic | 1.26 | 0.21 | -3.64 | 2.71E-04 | -6.51 | **7.33E-11** | -6.74 | **1.58E-11** | 86.7 | 4.99E-05 |
| cg05342515 | 2 | 177,024,172 | 2q31.1 | -7.58 | 3.53E-14 | 0.24 | *HOXD3* | Intronic | 0.97 | 0.33 | -3.74 | 1.86E-04 | -7.30 | **2.93E-13** | -7.69 | **1.42E-14** | 88.4 | 9.68E-06 |
| cg23676302 | 2 | 177,025,859 | 2q31.1 | -5.86 | 4.53E-09 | 0.46 | *HOXD3* | Intronic | 1.10 | 0.27 | -3.33 | 8.73E-04 | -4.75 | 2.07E-06 | -5.94 | **2.83E-09** | 74.5 | 8.25E-03 |
| cg07930539 | 2 | 177,025,975 | 2q31.1 | -5.46 | 4.67E-08 | 0.29 | *HOXD3* | Intronic | 1.36 | 0.17 | -3.05 | 2.26E-03 | -5.10 | 3.38E-07 | -5.72 | **1.09E-08** | 82.1 | 7.93E-04 |
| cg01871127 | 2 | 177,026,301 | 2q31.1 | -5.65 | 1.57E-08 | 0.45 | *HOXD3* | Intronic | 2.21 | 0.03 | -3.53 | 4.12E-04 | -6.52 | **6.90E-11** | -5.87 | **4.40E-09** | 90.4 | 7.25E-07 |
| cg12938020 | 2 | 177,027,075 | 2q31.1 | -6.31 | 2.84E-10 | 0.39 | *HOXD3* | Intronic | 0.52 | 0.61 | -3.43 | 6.06E-04 | -5.18 | **2.20E-07** | -6.45 | **1.15E-10** | 73.9 | 9.33E-03 |
| cg02773086 | 2 | 177,027,440 | 2q31.1 | -6.60 | 4.18E-11 | 0.31 | *HOXD3* | Intronic | 1.09 | 0.28 | -3.52 | 4.38E-04 | -5.96 | **2.59E-09** | -6.82 | **9.00E-12** | 84.1 | 2.93E-04 |
| cg01014615 | 2 | 177,027,621 | 2q31.1 | -6.93 | 4.24E-12 | 0.25 | *HOXD3* | Intronic | 1.13 | 0.26 | -3.89 | 9.95E-05 | -6.42 | **1.38E-10** | -6.98 | **2.93E-12** | 86.3 | 6.99E-05 |
| cg13316854 | 2 | 177,027,856 | 2q31.1 | -5.37 | 7.96E-08 | 0.23 | *HOXD3* | Intronic | 0.62 | 0.54 | -2.07 | 0.04 | -4.36 | 1.29E-05 | -5.65 | **1.60E-08** | 68.0 | 0.02 |
| cg06145336 | 2 | 177,027,931 | 2q31.1 | -6.54 | 6.05E-11 | 0.48 | *HOXD3* | Intronic | 1.42 | 0.16 | -3.06 | 2.24E-03 | -6.19 | **6.21E-10** | -7.06 | **1.65E-12** | 86.5 | 5.79E-05 |
| cg25938806 | 2 | 177,028,162 | 2q31.1 | -6.84 | 8.04E-12 | 0.50 | *HOXD3* | Intronic | 1.15 | 0.25 | -3.49 | 4.74E-04 | -6.50 | **8.23E-11** | -7.03 | **2.12E-12** | 86.7 | 4.93E-05 |
| cg01128482 | 2 | 177,028,621 | 2q31.1 | -6.62 | 3.60E-11 | 0.27 | *HOXD3* | Intronic | 0.98 | 0.32 | -3.18 | 1.47E-03 | -7.11 | **1.18E-12** | -6.65 | **2.98E-11** | 88.3 | 1.11E-05 |
| cg02408153 | 2 | 177,028,655 | 2q31.1 | -7.57 | 3.72E-14 | 0.34 | *HOXD3* | Intronic | 1.05 | 0.29 | -4.00 | 6.21E-05 | -7.73 | **1.04E-14** | -7.60 | **3.07E-14** | 89.8 | 1.92E-06 |
| cg24541426 | 2 | 177,029,459 | 2q31.1 | -7.20 | 5.94E-13 | 0.61 | *HOXD3* | Intronic | 1.12 | 0.26 | -3.86 | 1.14E-04 | -7.27 | **3.51E-13** | -7.24 | **4.44E-13** | 88.8 | 6.47E-06 |
| cg15812753 | 2 | 177,029,813 | 2q31.1 | -6.67 | 2.63E-11 | 0.47 | *HOXD3* | Intronic | 1.35 | 0.18 | -3.69 | 2.20E-04 | -7.24 | **4.37E-13** | -6.71 | **1.98E-11** | 89.7 | 2.09E-06 |
| cg04316624 | 2 | 177,036,809 | 2q31.1 | -5.36 | 8.25E-08 | 0.21 | *HOXD3* | Exonic | 1.75 | 0.08 | -2.92 | 3.54E-03 | -6.03 | **1.67E-09** | -5.55 | **2.87E-08** | 88.0 | 1.48E-05 |
| cg00278547 | 2 | 177,037,212 | 2q31.1 | -7.16 | 7.98E-13 | 0.39 | *HOXD3* | UTR3 | 1.32 | 0.19 | -3.82 | 1.32E-04 | -7.56 | **4.13E-14** | -7.26 | **3.85E-13** | 90.3 | 8.69E-07 |
| cg10159630 | 2 | 177,037,364 | 2q31.1 | -7.47 | 8.03E-14 | 0.45 | *HOXD3* | UTR3 | 1.16 | 0.25 | -3.93 | 8.63E-05 | -7.61 | **2.71E-14** | -7.56 | **4.07E-14** | 89.9 | 1.68E-06 |

| | | | | | | | | | | | | | | | | | | |
|---|---|---|---|---|---|---|---|---|---|---|---|---|---|---|---|---|---|---|
| cg13053653 | 2 | 177,037,631 | 2q31.1 | -7.60 | 3.03E-14 | 0.49 | *HOXD3* | UTR3 | 1.04 | 0.30 | -3.97 | 7.17E-05 | -7.66 | **1.90E-14** | -7.67 | **1.73E-14** | 89.5 | 2.62E-06 |
| cg21902187 | 2 | 177,037,710 | 2q31.1 | -6.51 | 7.28E-11 | 0.41 | *HOXD3* | UTR3 | 1.33 | 0.18 | -3.24 | 1.18E-03 | -6.51 | **7.57E-11** | -6.76 | **1.38E-11** | 87.1 | 3.53E-05 |
| cg22934308 | 2 | 177,038,617 | 2q31.1 | -7.54 | 4.61E-14 | 0.75 | *HAGLR* | Exonic | 0.99 | 0.32 | -3.97 | 7.18E-05 | -7.52 | **5.58E-14** | -7.60 | **2.88E-14** | 89.1 | 4.59E-06 |
| cg10477905 | 2 | 177,039,295 | 2q31.1 | -7.42 | 1.19E-13 | 0.79 | *HAGLR* | Exonic | 1.12 | 0.26 | -3.91 | 9.14E-05 | -7.56 | **4.04E-14** | -7.50 | **6.19E-14** | 89.6 | 2.43E-06 |
| cg13768137 | 2 | 177,039,426 | 2q31.1 | -7.15 | 8.64E-13 | 0.54 | *HAGLR* | Exonic | 1.11 | 0.27 | -3.80 | 1.44E-04 | -6.99 | **2.72E-12** | -7.36 | **1.82E-13** | 87.9 | 1.76E-05 |
| cg23322546 | 2 | 177,040,094 | 2q31.1 | -7.59 | 3.29E-14 | 0.72 | *HAGLR* | Exonic | 0.60 | 0.55 | -3.80 | 1.46E-04 | -7.16 | **7.88E-13** | -7.55 | **4.33E-14** | 86.6 | 5.57E-05 |
| cg10086659 | 2 | 177,040,226 | 2q31.1 | -7.34 | 2.09E-13 | 0.46 | *HAGLR* | Exonic | 1.21 | 0.23 | -3.61 | 3.06E-04 | -7.11 | **1.16E-12** | -7.64 | **2.22E-14** | 88.3 | 1.10E-05 |
| cg26754761 | 2 | 177,040,938 | 2q31.1 | -6.26 | 3.97E-10 | 0.47 | *HAGLR* | Exonic | 1.36 | 0.17 | -3.50 | 4.69E-04 | -6.82 | **9.02E-12** | -6.28 | **3.38E-10** | 88.7 | 7.31E-06 |
| cg23139698 | 2 | 177,041,515 | 2q31.1 | -7.32 | 2.40E-13 | 0.57 | *HAGLR* | Intronic | 1.01 | 0.31 | -3.43 | 5.99E-04 | -6.97 | **3.19E-12** | -7.46 | **8.97E-14** | 87.2 | 3.29E-05 |
| cg03401519 | 2 | 177,042,493 | 2q31.1 | -6.37 | 1.87E-10 | 0.59 | *HAGLR* | Exonic | 1.46 | 0.14 | -3.40 | 6.74E-04 | -7.00 | **2.59E-12** | -6.52 | **7.08E-11** | 89.5 | 2.58E-06 |
| cg13092806 | 2 | 177,043,255 | 2q31.1 | -7.13 | 1.00E-12 | 0.42 | *HAGLROS* | Exonic | 0.67 | 0.50 | -3.96 | 7.52E-05 | -7.27 | **3.58E-13** | -6.90 | **5.15E-12** | 87.4 | 2.68E-05 |
| cg04248916 | 2 | 177,052,295 | 2q31.1 | -7.47 | 7.82E-14 | 0.63 | *HAGLR* | Intronic | 0.96 | 0.34 | -3.92 | 8.78E-05 | -7.46 | **8.85E-14** | -7.57 | **3.86E-14** | 88.8 | 6.63E-06 |
| cg11418595 | 2 | 177,052,486 | 2q31.1 | -7.69 | 1.49E-14 | 0.75 | *HAGLR* | Intronic | 0.68 | 0.50 | -3.91 | 9.30E-05 | -7.30 | **2.96E-13** | -7.75 | **8.97E-15** | 87.2 | 3.29E-05 |
| cg07797397 | 2 | 177,052,527 | 2q31.1 | -7.48 | 7.20E-14 | 0.66 | *HAGLR* | Intronic | 1.00 | 0.32 | -3.93 | 8.40E-05 | -7.41 | **1.29E-13** | -7.62 | **2.50E-14** | 88.7 | 6.96E-06 |
| cg03553278 | 2 | 177,055,168 | 2q31.1 | 6.45 | 1.13E-10 | 0.39 | *HOXD1* | UTR3 | -1.31 | 0.19 | 3.83 | 1.29E-04 | 6.41 | **1.46E-10** | 6.31 | **2.73E-10** | 87.4 | 2.73E-05 |
| cg13295262 | 2 | 177,355,072 | 2q31.1 | -5.96 | 2.54E-09 | 0.41 | *MTX2* ; *MIR1246* | Intergenic | -0.81 | 0.42 | -3.57 | 3.62E-04 | -6.83 | **8.23E-12** | -5.08 | 3.80E-07 | 76.8 | 4.76E-03 |
| cg26791400 | 2 | 177,502,087 | 2q31.1 | -5.43 | 5.59E-08 | 0.25 | *LINC01116* | Intronic | 0.20 | 0.84 | -3.41 | 6.45E-04 | -5.52 | **3.46E-08** | -4.66 | 3.19E-06 | 72.8 | 0.01 |
| cg15616007 | 3 | 156,365,686 | 3q25.31 | -7.62 | 2.50E-14 | 0.48 | *SSR3* ; *TIPARP-AS1* | Intergenic | 0.10 | 0.92 | -1.79 | 0.07 | -1.11 | 0.27 | -8.37 | **5.93E-17** | 87.6 | 2.28E-05 |
| cg11886469 | 3 | 156,391,394 | 3q25.31 | 7.94 | 2.01E-15 | 0.32 | *TIPARP-AS1* | Exonic | -0.63 | 0.53 | 2.48 | 0.01 | 0.16 | 0.87 | 8.42 | **3.71E-17** | 90.2 | 1.05E-06 |
| cg09684262 | 3 | 156,391,685 | 3q25.31 | 10.57 | 4.05E-26 | 0.32 | *TIPARP-AS1* | Exonic | 1.39 | 0.16 | 2.85 | 4.42E-03 | 0.53 | 0.60 | 11.66 | **1.98E-31** | 88.8 | 6.85E-06 |
| cg24262469 | 3 | 156,391,694 | 3q25.31 | 7.80 | 6.05E-15 | 0.48 | *TIPARP-AS1* | Exonic | 0.46 | 0.65 | 1.92 | 0.05 | 0.79 | 0.43 | 8.77 | **1.71E-18** | 80.6 | 1.46E-03 |
| cg09015090 | 3 | 156,391,735 | 3q25.31 | 8.19 | 2.56E-16 | 0.21 | *TIPARP-AS1* | Exonic | 1.84 | 0.07 | 3.38 | 7.19E-04 | 1.03 | 0.30 | 8.39 | **4.78E-17** | 62.2 | 0.05 |
| cg01265971 | 3 | 156,398,928 | 3q25.31 | -8.04 | 8.85E-16 | 0.38 | *TIPARP* | Intronic | -0.73 | 0.47 | -1.59 | 0.11 | -0.69 | 0.49 | -8.84 | **9.29E-19** | 64.0 | 0.04 |
| cg26111729 | 3 | 156,878,163 | 3q25.31 | 6.55 | 5.76E-11 | 0.19 | *CCNL1* | UTR5 | 1.82 | 0.07 | 1.45 | 0.15 | -0.27 | 0.79 | 7.07 | **1.56E-12** | 71.4 | 0.01 |
| cg00624418 | 5 | 1,282,357 | 5p15.33 | -6.90 | 5.26E-12 | 0.65 | *TERT* | Intronic | 0.03 | 0.97 | -2.03 | 0.04 | -0.16 | 0.88 | -7.54 | **4.55E-14** | 81.5 | 1.03E-03 |
| cg25518571 | 5 | 1,286,042 | 5p15.33 | -5.93 | 2.99E-09 | 0.22 | *TERT* | Intronic | -0.07 | 0.94 | -2.59 | 9.52E-03 | -0.20 | 0.84 | -6.48 | **9.43E-11** | 79.1 | 2.50E-03 |
| cg07518058 | 8 | 129,199,528 | 8q24.21 | -5.88 | 4.17E-09 | 0.27 | *MIR1208* ; *LINC00824* | Intergenic | 2.03 | 0.04 | -1.45 | 0.15 | -1.51 | 0.13 | -7.10 | **1.22E-12** | 87.0 | 3.75E-05 |
| cg10462685 | 8 | 129,440,930 | 8q24.21 | 5.21 | 1.89E-07 | 0.19 | *LINC00824* | Intronic | -1.86 | 0.06 | 0.46 | 0.64 | 0.20 | 0.84 | 6.62 | **3.60E-11** | 87.8 | 1.80E-05 |
| cg12903012 | 8 | 129,569,577 | 8q24.21 | 6.65 | 3.02E-11 | 0.26 | *LINC00824* | Intronic | -2.70 | 6.91E-03 | 1.68 | 0.09 | 1.69 | 0.09 | 7.80 | **6.29E-15** | 90.7 | 4.58E-07 |
| cg18956029 | 9 | 16,870,888 | 9p22.2 | 5.71 | 1.10E-08 | 0.52 | *BNC2* | TSS200 | 1.59 | 0.11 | 0.64 | 0.52 | -1.14 | 0.26 | 6.69 | **2.16E-11** | 85.5 | 1.25E-04 |
| cg14260695 | 17 | 43,506,184 | 17q21.31 | -5.56 | 2.74E-08 | 0.22 | *ARHGAP27* | Intronic | -1.24 | 0.21 | -2.12 | 0.03 | 0.11 | 0.91 | -5.69 | **1.26E-08** | 73.4 | 0.01 |
| cg07368061 | 17 | 44,090,862 | 17q21.31 | 5.80 | 6.47E-09 | 0.44 | *MAPT* | Intronic | 0.31 | 0.76 | 2.49 | 0.01 | 1.17 | 0.24 | 5.22 | **1.82E-07** | 67.2 | 0.03 |
| cg03496533 | 17 | 46,035,183 | 17q21.32 | -6.04 | 1.56E-09 | 0.31 | *PRR15L* | UTR5 | -2.21 | 0.03 | -3.41 | 6.55E-04 | 0.56 | 0.57 | -5.94 | **2.85E-09** | 75.9 | 6.04E-03 |
| cg15738800 | 17 | 46,035,272 | 17q21.32 | -5.23 | 1.67E-07 | 0.34 | *PRR15L* | TSS200 | -0.86 | 0.39 | -3.83 | 1.30E-04 | -0.54 | 0.59 | -4.69 | 2.69E-06 | 71.0 | 0.02 |
| cg09607679 | 17 | 46,035,297 | 17q21.32 | -6.74 | 1.57E-11 | 0.49 | *PRR15L* | TSS200 | -1.72 | 0.09 | -4.06 | 4.96E-05 | -0.24 | 0.81 | -6.61 | **3.74E-11** | 78.6 | 2.89E-03 |
| cg24247040 | 17 | 46,231,565 | 17q21.32 | -5.47 | 4.54E-08 | 0.30 | *SKAP1* | Intronic | -1.25 | 0.21 | -1.27 | 0.20 | 0.11 | 0.91 | -5.74 | **9.66E-09** | 65.4 | 0.03 |
| cg06186330 | 17 | 46,522,513 | 17q21.32 | 6.20 | 5.51E-10 | 0.26 | *LOC101927166* | Exonic | 1.01 | 0.31 | 1.54 | 0.12 | -0.24 | 0.81 | 6.62 | **3.61E-11** | 76.1 | 5.74E-03 |
| cg06249421 | 17 | 46,567,806 | 17q21.32 | -5.42 | 5.87E-08 | 0.44 | *LOC101927166* ; *HOXB1* | Intergenic | -0.58 | 0.56 | -1.14 | 0.26 | 0.36 | 0.72 | -5.78 | **7.63E-09** | 71.8 | 0.01 |
| cg20401567 | 17 | 46,619,555 | 17q21.32 | -5.31 | 1.07E-07 | 0.42 | *HOXB2* | Downstream | -0.10 | 0.92 | -0.93 | 0.35 | -0.18 | 0.86 | -5.68 | **1.36E-08** | 68.1 | 0.02 |
| cg19047660 | 17 | 46,619,923 | 17q21.32 | -5.27 | 1.33E-07 | 0.32 | *HOXB2* | Downstream | -0.42 | 0.67 | -1.07 | 0.29 | -0.31 | 0.76 | -5.62 | **1.88E-08** | 65.5 | 0.03 |
| cg02494572 | 17 | 46,624,268 | 17q21.32 | -5.23 | 1.68E-07 | 0.26 | *HOXB-AS1* | Intronic | -0.54 | 0.59 | -0.96 | 0.34 | 0.29 | 0.77 | -5.59 | **2.22E-08** | 69.6 | 0.02 |
| cg15649236 | 17 | 46,657,504 | 17q21.32 | 5.26 | 1.44E-07 | 0.22 | *HOXB3* | Intronic | 1.34 | 0.18 | 3.69 | 2.20E-04 | 0.18 | 0.86 | 5.28 | **1.29E-07** | 64.7 | 0.04 |
| cg07898252 | 19 | 17,458,126 | 19p13.11 | -6.78 | 1.23E-11 | 0.56 | *GTPBP3* ; *PLVAP* | Intergenic | -0.33 | 0.74 | 0.24 | 0.81 | -0.13 | 0.90 | -7.96 | **1.76E-15** | 89.3 | 3.39E-06 |
| cg04749549 | 19 | 17,459,798 | 19p13.11 | -5.99 | 2.07E-09 | 0.46 | *GTPBP3* ; *PLVAP* | Intergenic | 0.43 | 0.67 | 1.10 | 0.27 | 0.41 | 0.68 | -7.52 | **5.28E-14** | 90.7 | 4.51E-07 |
| **Associations not showing significant heterogeneity across subtypes (Cochran's *P*>0.05)** | | | | | | | | | | | | | | | | | | |
| cg18702197 | 2 | 177,027,641 | 2q31.1 | -5.76 | 8.24E-09 | 0.30 | *HOXD3* | Intronic | 0.12 | 0.90 | -2.45 | 0.01 | -4.64 | 3.51E-06 | -6.11 | **9.86E-10** | 40.2 | 0.17 |
| cg07608961 | 3 | 156,511,429 | 3q25.31 | 5.21 | 1.89E-07 | 0.20 | *LINC00886* | Intronic | 2.27 | 0.02 | 1.90 | 0.06 | -0.34 | 0.73 | 5.64 | **1.70E-08** | 59.1 | 0.06 |
| cg06760536 | 10 | 21,785,735 | 10p12.31 | -5.43 | 5.77E-08 | 0.21 | *MIR1915HG* | Exonic | -2.64 | 8.40E-03 | -1.80 | 0.07 | -0.64 | 0.52 | -5.08 | 3.68E-07 | 32.7 | 0.22 |
| cg25349828 | 10 | 21,809,392 | 10p12.31 | 5.84 | 5.27E-09 | 0.45 | *SKIDA1* | Intronic | 2.60 | 9.40E-03 | 1.86 | 0.06 | 0.61 | 0.54 | 5.54 | **3.09E-08** | 39.1 | 0.18 |
| cg05913198 | 10 | 21,822,977 | 10p12.31 | -5.28 | 1.29E-07 | 0.19 | *MLLT10* | Intronic | -2.36 | 0.02 | -1.86 | 0.06 | -0.45 | 0.65 | -4.93 | 8.34E-07 | 26.3 | 0.25 |
| cg22199864 | 10 | 22,233,858 | 10p12.31 | -5.27 | 1.36E-07 | 0.55 | *DNAJC1* | Intronic | -2.71 | 6.72E-03 | -1.85 | 0.06 | -0.55 | 0.58 | -4.88 | 1.08E-06 | 29.7 | 0.23 |
| cg19943578 | 17 | 43,483,527 | 17q21.31 | -6.26 | 3.76E-10 | 0.19 | *ARHGAP27* | Intronic | -0.48 | 0.63 | -2.47 | 0.01 | -1.39 | 0.16 | -5.74 | **9.30E-09** | 22.4 | 0.28 |
| cg24477401 | 17 | 43,484,506 | 17q21.31 | -7.37 | 1.77E-13 | 0.62 | *ARHGAP27* | Intronic | -1.56 | 0.12 | -3.05 | 2.31E-03 | -0.98 | 0.33 | -6.95 | **3.64E-12** | 44.8 | 0.14 |
| cg05314706 | 17 | 43,508,250 | 17q21.31 | -7.47 | 8.03E-14 | 0.30 | *ARHGAP27* | Intronic | -1.32 | 0.19 | -2.85 | 4.40E-03 | -1.03 | 0.30 | -7.02 | **2.23E-12** | 49.1 | 0.12 |
| cg04487659 | 17 | 43,510,575 | 17q21.31 | 5.88 | 4.21E-09 | 0.34 | *ARHGAP27* | TSS1500 | 0.63 | 0.53 | 2.03 | 0.04 | 0.66 | 0.51 | 5.48 | **4.19E-08** | 35.3 | 0.20 |
| cg00916973 | 17 | 43,580,461 | 17q21.31 | -6.75 | 1.52E-11 | 0.64 | *ARL17B* | Upstream | -0.44 | 0.66 | -2.80 | 5.15E-03 | -1.22 | 0.22 | -6.23 | **4.56E-10** | 43.7 | 0.15 |
| cg22433210 | 17 | 43,662,623 | 17q21.31 | 7.10 | 1.28E-12 | 0.74 | *ARL17A* ; *MAPK8IP1P2* | Intergenic | 0.60 | 0.55 | 3.14 | 1.67E-03 | 1.22 | 0.22 | 6.47 | **9.53E-11** | 45.6 | 0.14 |
| cg01341218 | 17 | 43,662,625 | 17q21.31 | 7.15 | 8.75E-13 | 0.64 | *ARL17A* ; *MAPK8IP1P2* | Intergenic | 0.62 | 0.53 | 3.21 | 1.33E-03 | 1.25 | 0.21 | 6.51 | **7.43E-11** | 45.3 | 0.14 |
| cg12297030 | 17 | 43,662,878 | 17q21.31 | 6.91 | 4.79E-12 | 0.46 | *ARL17A* ; *MAPK8IP1P2* | Intergenic | 0.52 | 0.60 | 2.96 | 3.05E-03 | 1.20 | 0.23 | 6.36 | **2.02E-10** | 45.1 | 0.14 |
| cg17117718 | 17 | 43,663,208 | 17q21.31 | 7.14 | 9.40E-13 | 0.76 | *ARL17A* ; *MAPK8IP1P2* | Intergenic | 0.59 | 0.55 | 3.17 | 1.55E-03 | 1.27 | 0.20 | 6.51 | **7.29E-11** | 45.5 | 0.14 |
| cg05727186 | 17 | 43,697,356 | 17q21.31 | -6.15 | 7.60E-10 | 0.26 | *MGC57346* | TSS1500 | -0.84 | 0.40 | -3.07 | 2.12E-03 | -1.36 | 0.17 | -5.55 | **2.91E-08** | 0.0 | 0.41 |
| cg18815117 | 17 | 43,698,604 | 17q21.31 | -7.11 | 1.13E-12 | 0.75 | *LINC02210* | Intronic | -0.71 | 0.48 | -3.16 | 1.58E-03 | -1.29 | 0.20 | -6.47 | **1.01E-10** | 40.5 | 0.17 |
| cg01882395 | 17 | 43,717,810 | 17q21.31 | 7.35 | 1.93E-13 | 0.70 | *LINC02210* | Exonic | 0.55 | 0.58 | 3.01 | 2.60E-03 | 1.37 | 0.17 | 6.71 | **2.01E-11** | 47.2 | 0.13 |
| cg18753072 | 17 | 43,719,194 | 17q21.31 | 7.33 | 2.37E-13 | 0.35 | *LINC02210* | Exonic | 0.79 | 0.43 | 3.13 | 1.72E-03 | 1.26 | 0.21 | 6.71 | **1.93E-11** | 45.0 | 0.14 |
| cg20059597 | 17 | 43,726,219 | 17q21.31 | 6.98 | 2.99E-12 | 0.58 | *LINC02210* | Intronic | 0.56 | 0.58 | 2.92 | 3.45E-03 | 1.23 | 0.22 | 6.46 | **1.05E-10** | 45.5 | 0.14 |
| cg20393308 | 17 | 43,728,044 | 17q21.31 | 7.12 | 1.12E-12 | 0.74 | *LINC02210* | Intronic | 0.59 | 0.55 | 3.15 | 1.61E-03 | 1.22 | 0.22 | 6.51 | **7.43E-11** | 46.6 | 0.13 |
| cg16050799 | 17 | 43,734,332 | 17q21.31 | 7.06 | 1.63E-12 | 0.32 | *LINC02210* | Intronic | 0.67 | 0.50 | 3.07 | 2.14E-03 | 1.04 | 0.30 | 6.52 | **7.06E-11** | 49.3 | 0.12 |
| cg08276280 | 17 | 43,743,868 | 17q21.31 | 6.63 | 3.37E-11 | 0.63 | *LINC02210* | Intronic | 0.54 | 0.59 | 2.49 | 0.01 | 1.19 | 0.23 | 6.29 | **3.26E-10** | 43.4 | 0.15 |
| cg19447040 | 17 | 43,750,272 | 17q21.31 | 5.81 | 6.42E-09 | 0.34 | *LINC02210* | Intronic | 0.62 | 0.53 | 1.90 | 0.06 | 0.49 | 0.62 | 5.31 | **1.07E-07** | 36.8 | 0.19 |
| cg01448078 | 17 | 43,752,782 | 17q21.31 | 6.96 | 3.31E-12 | 0.56 | *LINC02210* | Intronic | 0.64 | 0.52 | 3.04 | 2.36E-03 | 1.41 | 0.16 | 6.17 | **7.02E-10** | 19.3 | 0.29 |
| cg06475362 | 17 | 43,763,639 | 17q21.31 | 6.92 | 4.39E-12 | 0.54 | *LINC02210* | Intronic | 1.10 | 0.27 | 2.98 | 2.88E-03 | 1.21 | 0.23 | 6.28 | **3.41E-10** | 0.0 | 0.42 |
| cg07817266 | 17 | 43,765,890 | 17q21.31 | 7.01 | 2.46E-12 | 0.73 | *LINC02210* | Intronic | 0.74 | 0.46 | 2.95 | 3.20E-03 | 1.15 | 0.25 | 6.39 | **1.61E-10** | 41.3 | 0.16 |
| cg07508782 | 17 | 43,783,158 | 17q21.31 | 5.72 | 1.07E-08 | 0.32 | *LINC02210* | Intronic | 0.80 | 0.42 | 1.70 | 0.09 | 0.61 | 0.54 | 5.63 | **1.85E-08** | 43.1 | 0.15 |
| cg03820725 | 17 | 43,787,766 | 17q21.31 | 6.52 | 6.96E-11 | 0.28 | *LINC02210* | Intronic | 0.42 | 0.68 | 1.99 | 0.05 | 0.90 | 0.37 | 6.17 | **6.84E-10** | 53.2 | 0.09 |
| cg13111061 | 17 | 43,797,230 | 17q21.31 | 5.69 | 1.30E-08 | 0.44 | *LINC02210* | Intronic | 0.08 | 0.93 | 2.10 | 0.04 | 1.20 | 0.23 | 5.13 | 2.91E-07 | 26.8 | 0.25 |
| cg08590638 | 17 | 43,798,045 | 17q21.31 | 6.65 | 2.93E-11 | 0.63 | *LINC02210* | Intronic | 0.35 | 0.72 | 2.72 | 6.51E-03 | 1.15 | 0.25 | 6.29 | **3.12E-10** | 49.5 | 0.11 |
| cg06537391 | 17 | 43,799,898 | 17q21.31 | 7.27 | 3.68E-13 | 0.70 | *LINC02210* | Intronic | 0.76 | 0.44 | 3.28 | 1.04E-03 | 1.54 | 0.12 | 6.56 | **5.34E-11** | 33.9 | 0.21 |
| cg03238273 | 17 | 43,825,672 | 17q21.31 | 6.98 | 2.93E-12 | 0.56 | *LINC02210* | Intronic | 0.71 | 0.47 | 3.02 | 2.56E-03 | 1.12 | 0.26 | 6.42 | **1.40E-10** | 44.1 | 0.15 |
| cg10556198 | 17 | 43,826,307 | 17q21.31 | 6.93 | 4.25E-12 | 0.67 | *LINC02210* | Intronic | 0.56 | 0.58 | 2.91 | 3.59E-03 | 1.15 | 0.25 | 6.35 | **2.18E-10** | 45.1 | 0.14 |
| cg24394631 | 17 | 43,863,000 | 17q21.31 | -6.11 | 9.84E-10 | 0.28 | *CRHR1* | Intronic | -0.96 | 0.33 | -3.21 | 1.33E-03 | -0.96 | 0.34 | -5.35 | **8.68E-08** | 17.4 | 0.30 |
| cg11760414 | 17 | 43,863,398 | 17q21.31 | 6.63 | 3.44E-11 | 0.39 | *CRHR1* | Intronic | 0.80 | 0.43 | 3.27 | 1.09E-03 | 0.90 | 0.37 | 5.99 | **2.05E-09** | 35.4 | 0.20 |
| cg23420656 | 17 | 43,865,031 | 17q21.31 | 8.01 | 1.11E-15 | 0.27 | *CRHR1* | Intronic | 0.93 | 0.35 | 3.50 | 4.67E-04 | 1.46 | 0.14 | 7.30 | **2.77E-13** | 45.0 | 0.14 |
| cg03066966 | 17 | 43,902,069 | 17q21.31 | 6.13 | 8.63E-10 | 0.34 | *CRHR1* | Intronic | 0.13 | 0.90 | 1.66 | 0.10 | 0.87 | 0.38 | 6.05 | **1.49E-09** | 57.3 | 0.07 |
| cg00025823 | 17 | 43,909,151 | 17q21.31 | 7.30 | 2.96E-13 | 0.25 | *CRHR1* | Intronic | 0.31 | 0.76 | 2.81 | 4.94E-03 | 1.12 | 0.26 | 6.67 | **2.59E-11** | 55.9 | 0.08 |
| cg26656751 | 17 | 43,910,226 | 17q21.31 | 5.87 | 4.36E-09 | 0.52 | CRHR1 | Intronic | 0.46 | 0.64 | 2.00 | 0.05 | 1.30 | 0.19 | 5.53 | **3.25E-08** | 21.3 | 0.28 |
| cg15844800 | 17 | 43,916,233 | 17q21.31 | 5.82 | 5.83E-09 | 0.36 | *CRHR1* ; *MAPT-AS1* | Intergenic | 0.15 | 0.88 | 2.21 | 0.03 | 1.23 | 0.22 | 5.37 | **7.85E-08** | 26.9 | 0.25 |
| cg15295732 | 17 | 43,942,128 | 17q21.31 | -6.59 | 4.53E-11 | 0.53 | *MAPT-AS1* | Intronic | -0.31 | 0.76 | -2.71 | 6.69E-03 | -1.21 | 0.23 | -6.20 | **5.78E-10** | 46.9 | 0.13 |
| cg13115898 | 17 | 43,951,350 | 17q21.31 | -6.93 | 4.20E-12 | 0.47 | *MAPT-AS1* | Intronic | -0.69 | 0.49 | -3.25 | 1.15E-03 | -1.14 | 0.26 | -6.25 | **3.99E-10** | 42.1 | 0.16 |
| cg23202543 | 17 | 43,971,022 | 17q21.31 | -7.61 | 2.65E-14 | 0.20 | *MAPT-AS1* | Intronic | -0.83 | 0.41 | -3.22 | 1.30E-03 | -1.48 | 0.14 | -6.97 | **3.17E-12** | 57.0 | 0.07 |
| cg17473914 | 17 | 43,971,149 | 17q21.31 | 6.46 | 1.03E-10 | 0.67 | *MAPT-AS1* | Intronic | 0.44 | 0.66 | 2.69 | 7.19E-03 | 0.90 | 0.37 | 6.02 | **1.77E-09** | 31.5 | 0.22 |
| cg05301556 | 17 | 43,971,177 | 17q21.31 | -7.23 | 4.84E-13 | 0.38 | *MAPT-AS1* | Intronic | -0.63 | 0.53 | -3.13 | 1.72E-03 | -1.22 | 0.22 | -6.60 | **4.12E-11** | 47.3 | 0.13 |
| cg16520312 | 17 | 43,971,471 | 17q21.31 | -7.14 | 9.16E-13 | 0.55 | *MAPT-AS1* | Intronic | -0.64 | 0.52 | -3.11 | 1.87E-03 | -1.26 | 0.21 | -6.56 | **5.33E-11** | 45.5 | 0.14 |
| cg13773723 | 17 | 43,971,894 | 17q21.31 | 5.51 | 3.59E-08 | 0.42 | *MAPT-AS1* | Intronic | 0.68 | 0.50 | 1.45 | 0.15 | 0.47 | 0.64 | 5.46 | **4.78E-08** | 46.1 | 0.13 |
| cg07163735 | 17 | 43,971,906 | 17q21.31 | 6.71 | 1.89E-11 | 0.34 | *MAPT-AS1* | Intronic | 0.81 | 0.42 | 2.93 | 3.42E-03 | 1.14 | 0.26 | 6.15 | **7.83E-10** | 32.6 | 0.22 |
| cg23202277 | 17 | 43,971,911 | 17q21.31 | 6.93 | 4.10E-12 | 0.75 | *MAPT-AS1* | Intronic | 0.53 | 0.60 | 3.05 | 2.27E-03 | 1.17 | 0.24 | 6.34 | **2.33E-10** | 45.6 | 0.14 |
| cg20099416 | 17 | 43,971,919 | 17q21.31 | 6.77 | 1.26E-11 | 0.74 | *MAPT-AS1* | Intronic | 0.98 | 0.33 | 2.54 | 0.01 | 0.88 | 0.38 | 6.19 | **6.03E-10** | 36.7 | 0.19 |
| cg00891649 | 17 | 43,972,573 | 17q21.31 | -7.30 | 2.90E-13 | 0.41 | *MAPT-AS1* | Intronic | -0.57 | 0.57 | -2.96 | 3.11E-03 | -1.36 | 0.17 | -6.69 | **2.19E-11** | 48.1 | 0.12 |
| cg11489262 | 17 | 43,973,426 | 17q21.31 | 7.13 | 1.04E-12 | 0.52 | *MAPT-IT1* | Exonic | 0.67 | 0.50 | 3.05 | 2.27E-03 | 1.21 | 0.22 | 6.54 | **6.01E-11** | 45.5 | 0.14 |
| cg10780632 | 17 | 43,973,522 | 17q21.31 | 5.67 | 1.40E-08 | 0.83 | *MAPT-IT1* | Exonic | 0.25 | 0.80 | 1.76 | 0.08 | 1.70 | 0.09 | 5.41 | **6.20E-08** | 37.2 | 0.19 |
| cg14202850 | 17 | 43,974,869 | 17q21.31 | -6.66 | 2.76E-11 | 0.26 | *MAPT-IT1* | Exonic | -0.27 | 0.79 | -2.57 | 0.01 | -1.37 | 0.17 | -6.30 | **3.05E-10** | 46.7 | 0.13 |
| cg11909912 | 17 | 43,974,919 | 17q21.31 | -7.18 | 6.75E-13 | 0.38 | *MAPT-IT1* | Exonic | -0.71 | 0.48 | -2.98 | 2.88E-03 | -1.09 | 0.28 | -6.61 | **3.87E-11** | 48.9 | 0.12 |

| | | | | | | | | | | | | | | | | | | |
|---|---|---|---|---|---|---|---|---|---|---|---|---|---|---|---|---|---|---|
| cg01630519 | 17 | 43,992,416 | 17q21.31 | 7.22 | 5.11E-13 | 0.57 | *MAPT* | Intronic | 0.73 | 0.47 | 3.13 | 1.75E-03 | 1.24 | 0.21 | 6.59 | **4.41E-11** | 44.5 | 0.14 |
| cg13972556 | 17 | 43,993,497 | 17q21.31 | -6.11 | 1.01E-09 | 0.46 | *MAPT* | Intronic | -0.14 | 0.89 | -1.86 | 0.06 | -0.64 | 0.52 | -6.02 | **1.70E-09** | 58.1 | 0.07 |
| cg15739383 | 17 | 43,993,684 | 17q21.31 | -6.72 | 1.85E-11 | 0.59 | *MAPT* | Intronic | -0.23 | 0.82 | -2.53 | 0.01 | -1.35 | 0.18 | -6.42 | **1.39E-10** | 48.9 | 0.12 |
| cg27477690 | 17 | 43,996,840 | 17q21.31 | 7.03 | 2.05E-12 | 0.57 | *MAPT* | Intronic | 0.82 | 0.41 | 3.18 | 1.47E-03 | 1.33 | 0.18 | 6.33 | **2.38E-10** | 31.7 | 0.22 |
| cg15476265 | 17 | 44,012,006 | 17q21.31 | -6.89 | 5.42E-12 | 0.30 | *MAPT* | Intronic | -0.44 | 0.66 | -2.62 | 8.68E-03 | -1.12 | 0.26 | -6.44 | **1.19E-10** | 52.0 | 0.10 |
| cg04226788 | 17 | 44,014,390 | 17q21.31 | 7.15 | 8.69E-13 | 0.27 | *MAPT* | Intronic | 0.56 | 0.57 | 3.15 | 1.61E-03 | 1.25 | 0.21 | 6.52 | **6.81E-11** | 47.0 | 0.13 |
| cg20750326 | 17 | 44,019,832 | 17q21.31 | -6.66 | 2.82E-11 | 0.29 | *MAPT* | Intronic | -0.39 | 0.70 | -2.68 | 7.46E-03 | -0.60 | 0.55 | -6.32 | **2.70E-10** | 60.4 | 0.06 |
| cg16573346 | 17 | 44,020,278 | 17q21.31 | 6.85 | 7.30E-12 | 0.52 | *MAPT* | Intronic | 1.22 | 0.22 | 3.08 | 2.06E-03 | 1.32 | 0.19 | 6.12 | **9.13E-10** | 7.4 | 0.36 |
| cg23926235 | 17 | 44,024,256 | 17q21.31 | -7.15 | 8.65E-13 | 0.73 | *MAPT* | Intronic | -0.69 | 0.49 | -3.07 | 2.14E-03 | -1.20 | 0.23 | -6.54 | **6.08E-11** | 45.4 | 0.14 |
| cg05772917 | 17 | 44,027,251 | 17q21.31 | 6.93 | 4.14E-12 | 0.60 | *MAPT* | Intronic | 0.41 | 0.68 | 2.98 | 2.90E-03 | 1.21 | 0.23 | 6.34 | **2.33E-10** | 47.2 | 0.13 |
| cg01640727 | 17 | 44,029,468 | 17q21.31 | -7.30 | 2.81E-13 | 0.45 | *MAPT* | Intronic | -0.48 | 0.63 | -2.75 | 5.96E-03 | -1.24 | 0.21 | -6.69 | **2.29E-11** | 52.1 | 0.10 |
| cg00112026 | 17 | 44,033,451 | 17q21.31 | 6.57 | 4.90E-11 | 0.38 | *MAPT* | Intronic | 0.23 | 0.82 | 2.54 | 0.01 | 0.94 | 0.35 | 6.15 | **7.74E-10** | 53.4 | 0.09 |
| cg23044891 | 17 | 44,038,886 | 17q21.31 | 6.98 | 2.92E-12 | 0.18 | *MAPT* | Intronic | 1.08 | 0.28 | 3.34 | 8.38E-04 | 1.12 | 0.26 | 6.30 | **2.98E-10** | 34.2 | 0.21 |
| cg02159926 | 17 | 44,039,472 | 17q21.31 | 6.78 | 1.20E-11 | 0.30 | *MAPT* | Intronic | 0.67 | 0.50 | 2.73 | 6.38E-03 | 1.13 | 0.26 | 6.35 | **2.17E-10** | 42.5 | 0.16 |
| cg22635938 | 17 | 44,039,549 | 17q21.31 | -7.70 | 1.36E-14 | 0.35 | *MAPT* | Intronic | -0.97 | 0.33 | -3.08 | 2.05E-03 | -1.75 | 0.08 | -7.11 | **1.20E-12** | 36.5 | 0.19 |
| cg18586914 | 17 | 44,040,821 | 17q21.31 | -6.90 | 5.10E-12 | 0.57 | *MAPT* | Intronic | -0.70 | 0.49 | -2.89 | 3.91E-03 | -1.18 | 0.24 | -6.34 | **2.31E-10** | 40.6 | 0.17 |
| cg26868406 | 17 | 44,049,253 | 17q21.31 | 7.48 | 7.21E-14 | 0.35 | *MAPT* | Exonic | 1.29 | 0.20 | 3.26 | 1.11E-03 | 1.25 | 0.21 | 6.44 | **1.20E-10** | 23.6 | 0.27 |
| cg01934064 | 17 | 44,064,242 | 17q21.31 | 6.91 | 4.92E-12 | 0.66 | *MAPT* | Intronic | 0.67 | 0.51 | 2.82 | 4.88E-03 | 1.05 | 0.29 | 6.42 | **1.40E-10** | 47.1 | 0.13 |
| cg05721485 | 17 | 44,071,124 | 17q21.31 | 7.20 | 5.94E-13 | 0.61 | *MAPT* | Intronic | 0.71 | 0.48 | 3.16 | 1.60E-03 | 1.31 | 0.19 | 6.59 | **4.50E-11** | 42.9 | 0.15 |
| cg25885883 | 17 | 44,075,753 | 17q21.31 | -5.92 | 3.22E-09 | 0.22 | *MAPT* | Intronic | -0.73 | 0.47 | -2.57 | 0.01 | -1.14 | 0.26 | -5.09 | 3.59E-07 | 0.0 | 0.52 |
| cg15682851 | 17 | 44,076,429 | 17q21.31 | -6.95 | 3.60E-12 | 0.27 | *MAPT* | Intronic | -1.07 | 0.28 | -3.42 | 6.23E-04 | -1.13 | 0.26 | -6.05 | **1.48E-09** | 27.4 | 0.25 |
| cg07936825 | 17 | 44,081,997 | 17q21.31 | 6.23 | 4.79E-10 | 0.47 | *MAPT* | Intronic | 0.64 | 0.52 | 2.34 | 0.02 | 0.89 | 0.38 | 6.01 | **1.89E-09** | 43.0 | 0.15 |
| cg00792758 | 17 | 44,087,219 | 17q21.31 | -7.33 | 2.36E-13 | 0.66 | *MAPT* | Intronic | -0.83 | 0.41 | -3.01 | 2.60E-03 | -1.04 | 0.30 | -6.79 | **1.09E-11** | 47.3 | 0.13 |
| cg15269754 | 17 | 44,088,248 | 17q21.31 | 7.15 | 8.67E-13 | 0.50 | *MAPT* | Intronic | 0.83 | 0.41 | 2.87 | 4.09E-03 | 1.39 | 0.17 | 6.05 | **1.43E-09** | 13.2 | 0.33 |
| cg14888901 | 17 | 44,091,062 | 17q21.31 | 6.78 | 1.24E-11 | 0.69 | *MAPT* | Intronic | 0.63 | 0.53 | 2.80 | 5.18E-03 | 1.12 | 0.26 | 6.33 | **2.50E-10** | 43.5 | 0.15 |
| cg24787400 | 17 | 44,097,757 | 17q21.31 | 6.78 | 1.19E-11 | 0.23 | *MAPT* | Intronic | 1.11 | 0.27 | 3.52 | 4.32E-04 | 2.97 | 2.99E-03 | 5.77 | **8.01E-09** | 0.0 | 0.59 |
| cg05533539 | 17 | 44,104,521 | 17q21.31 | 6.84 | 8.15E-12 | 0.44 | *MAPT* | UTR3 | 0.74 | 0.46 | 2.61 | 8.93E-03 | 1.29 | 0.20 | 6.17 | **6.78E-10** | 26.6 | 0.25 |
| cg07298766 | 17 | 44,111,831 | 17q21.31 | -5.99 | 2.13E-09 | 0.25 | *KANSL1* | Intronic | -0.47 | 0.64 | -2.30 | 0.02 | -0.61 | 0.54 | -5.71 | **1.10E-08** | 47.3 | 0.13 |
| cg07694853 | 17 | 44,116,674 | 17q21.31 | -5.76 | 8.46E-09 | 0.20 | *KANSL1* | Intronic | -0.77 | 0.44 | -1.90 | 0.06 | -0.62 | 0.53 | -4.86 | 1.19E-06 | 9.1 | 0.35 |
| cg08318660 | 17 | 44,122,580 | 17q21.31 | 6.36 | 2.06E-10 | 0.56 | *KANSL1* | Intronic | 0.43 | 0.67 | 2.59 | 9.68E-03 | 0.96 | 0.34 | 5.92 | **3.30E-09** | 42.1 | 0.16 |
| cg22192454 | 17 | 44,135,007 | 17q21.31 | -6.17 | 6.96E-10 | 0.64 | *KANSL1* | Intronic | -0.33 | 0.74 | -2.04 | 0.04 | -1.32 | 0.19 | -5.66 | **1.49E-08** | 19.9 | 0.29 |
| cg26340828 | 17 | 44,140,703 | 17q21.31 | 5.23 | 1.70E-07 | 0.29 | *KANSL1* | Intronic | 0.67 | 0.50 | 1.50 | 0.13 | 0.86 | 0.39 | 4.46 | 8.25E-06 | 0.0 | 0.50 |
| cg14642709 | 17 | 44,199,852 | 17q21.31 | 6.26 | 3.81E-10 | 0.35 | *KANSL1* | Intronic | 1.15 | 0.25 | 3.44 | 5.88E-04 | 1.14 | 0.25 | 5.44 | **5.32E-08** | 7.7 | 0.35 |
| cg01906119 | 17 | 44,208,035 | 17q21.31 | -5.45 | 5.00E-08 | 0.42 | *KANSL1* | Intronic | -0.87 | 0.38 | -2.76 | 5.73E-03 | -1.01 | 0.31 | -4.45 | 8.61E-06 | 0.0 | 0.58 |
| cg06721910 | 17 | 44,223,602 | 17q21.31 | -6.32 | 2.57E-10 | 0.54 | *KANSL1* | Intronic | -0.86 | 0.39 | -2.39 | 0.02 | -1.59 | 0.11 | -5.93 | **3.07E-09** | 0.0 | 0.42 |
| cg00512837 | 17 | 44,237,286 | 17q21.31 | 7.01 | 2.45E-12 | 0.32 | *KANSL1* | Intronic | 1.14 | 0.25 | 3.09 | 1.99E-03 | 1.04 | 0.30 | 6.21 | **5.16E-10** | 31.5 | 0.22 |
| cg21214508 | 17 | 44,248,233 | 17q21.31 | -7.55 | 4.29E-14 | 0.45 | *KANSL1* | Exonic | -1.47 | 0.14 | -3.34 | 8.41E-04 | -1.69 | 0.09 | -6.87 | **6.46E-12** | 0.0 | 0.59 |
| cg14374015 | 17 | 44,249,569 | 17q21.31 | 5.91 | 3.49E-09 | 0.30 | *KANSL1* | UTR5 | 0.66 | 0.51 | 2.51 | 0.01 | 1.58 | 0.11 | 5.23 | **1.70E-07** | 0.0 | 0.51 |
| cg25464663 | 17 | 44,254,545 | 17q21.31 | -5.81 | 6.29E-09 | 0.46 | *KANSL1* | Intronic | -0.57 | 0.57 | -2.33 | 0.02 | -1.02 | 0.31 | -5.92 | **3.19E-09** | 38.4 | 0.18 |
| cg10191506 | 17 | 44,260,227 | 17q21.31 | -6.81 | 9.99E-12 | 0.40 | *KANSL1* | Intronic | -0.78 | 0.44 | -3.12 | 1.83E-03 | -1.16 | 0.25 | -6.26 | **3.89E-10** | 38.0 | 0.18 |
| cg17003784 | 17 | 44,268,391 | 17q21.31 | 7.10 | 1.28E-12 | 0.25 | *KANSL1* | Intronic | 0.35 | 0.72 | 2.85 | 4.36E-03 | 1.15 | 0.25 | 6.50 | **7.81E-11** | 52.8 | 0.10 |
| cg24599892 | 17 | 44,270,211 | 17q21.31 | -7.02 | 2.28E-12 | 0.23 | *KANSL1* | UTR5 | -1.28 | 0.20 | -3.20 | 1.37E-03 | -1.70 | 0.09 | -6.28 | **3.34E-10** | 0.0 | 0.46 |
| cg24992007 | 17 | 44,270,238 | 17q21.31 | -6.24 | 4.49E-10 | 0.55 | *KANSL1* | UTR5 | -0.53 | 0.60 | -2.85 | 4.40E-03 | -1.09 | 0.27 | -5.94 | **2.86E-09** | 37.5 | 0.19 |
| cg04739613 | 17 | 44,270,960 | 17q21.31 | 6.51 | 7.64E-11 | 0.36 | *KANSL1-AS1* | Exonic | 0.31 | 0.76 | 2.49 | 0.01 | 1.21 | 0.23 | 5.78 | **7.33E-09** | 34.3 | 0.21 |
| cg09250749 | 17 | 44,271,316 | 17q21.31 | 7.32 | 2.45E-13 | 0.34 | *KANSL1-AS1* | Intronic | 0.68 | 0.49 | 3.85 | 1.17E-04 | 2.20 | 0.03 | 6.79 | **1.12E-11** | 43.1 | 0.15 |
| cg17642057 | 17 | 44,274,734 | 17q21.31 | -5.59 | 2.24E-08 | 0.33 | *KANSL1* | Intronic | -0.50 | 0.62 | -2.50 | 0.01 | -0.98 | 0.33 | -5.47 | **4.41E-08** | 26.3 | 0.25 |
| cg15790431 | 17 | 44,292,828 | 17q21.31 | -6.90 | 5.23E-12 | 0.31 | *KANSL1* | Intronic | -0.71 | 0.48 | -2.97 | 2.99E-03 | -1.20 | 0.23 | -6.46 | **1.07E-10** | 42.6 | 0.16 |
| cg08670715 | 17 | 44,341,754 | 17q21.31 | -6.51 | 7.37E-11 | 0.32 | *KANSL1* ; *ARL17B* | Intergenic | -0.34 | 0.73 | -2.60 | 9.31E-03 | -1.45 | 0.15 | -6.12 | **9.53E-10** | 47.3 | 0.13 |
| cg17911788 | 17 | 44,343,683 | 17q21.31 | -5.85 | 4.82E-09 | 0.69 | *KANSL1* ; *ARL17B* | Intergenic | -0.52 | 0.60 | -2.18 | 0.03 | -1.35 | 0.18 | -5.70 | **1.19E-08** | 20.3 | 0.29 |
| cg05159804 | 17 | 44,343,776 | 17q21.31 | -7.13 | 1.04E-12 | 0.20 | *KANSL1* ; *ARL17B* | Intergenic | -0.58 | 0.56 | -3.18 | 1.45E-03 | -1.25 | 0.21 | -6.53 | **6.38E-11** | 47.0 | 0.13 |
| cg17736285 | 17 | 44,347,643 | 17q21.31 | 7.36 | 1.87E-13 | 0.37 | *KANSL1* ; *ARL17B* | Intergenic | 0.56 | 0.58 | 3.07 | 2.14E-03 | 1.37 | 0.17 | 6.56 | **5.51E-11** | 43.6 | 0.15 |
| cg13438899 | 17 | 44,368,573 | 17q21.31 | 6.21 | 5.45E-10 | 0.21 | *ARL17B* | Intronic | 0.50 | 0.61 | 2.26 | 0.02 | 1.54 | 0.12 | 5.92 | **3.21E-09** | 23.2 | 0.27 |
| cg00140914 | 17 | 44,859,215 | 17q21.31 | 5.64 | 1.74E-08 | 0.39 | *WNT3* | Intronic | 0.85 | 0.40 | 1.99 | 0.05 | 1.26 | 0.21 | 5.52 | **3.44E-08** | 0.0 | 0.39 |
| cg10161440 | 17 | 44,860,024 | 17q21.31 | 6.31 | 2.86E-10 | 0.53 | *WNT3* | Intronic | 0.57 | 0.57 | 2.93 | 3.36E-03 | 1.09 | 0.28 | 5.87 | **4.34E-09** | 38.3 | 0.18 |
| cg20626547 | 17 | 44,863,294 | 17q21.31 | 6.65 | 2.87E-11 | 0.18 | *WNT3* | Intronic | 0.70 | 0.48 | 2.74 | 6.06E-03 | 1.19 | 0.23 | 6.35 | **2.17E-10** | 41.0 | 0.17 |
| cg20859882 | 17 | 46,093,755 | 17q21.32 | 6.16 | 7.12E-10 | 0.27 | *CDK5RAP3* ; *COPZ2* | Intergenic | 1.88 | 0.06 | 1.94 | 0.05 | 0.84 | 0.40 | 6.20 | **5.67E-10** | 37.2 | 0.19 |
| cg23959187 | 17 | 46,102,881 | 17q21.32 | -5.37 | 7.83E-08 | 0.41 | *COPZ2* | Downstream | -1.42 | 0.15 | -2.72 | 6.57E-03 | -0.96 | 0.34 | -5.16 | **2.48E-07** | 0.0 | 0.52 |
| cg24027378 | 17 | 46,103,806 | 17q21.32 | 6.22 | 4.93E-10 | 0.18 | *COPZ2* | Exonic | 1.86 | 0.06 | 2.09 | 0.04 | -0.17 | 0.87 | 6.69 | **2.21E-11** | 60.5 | 0.06 |
| cg11314488 | 17 | 46,491,307 | 17q21.32 | -6.94 | 3.90E-12 | 0.21 | *SKAP1* | Intronic | -1.74 | 0.08 | -2.20 | 0.03 | -0.76 | 0.45 | -7.30 | **2.85E-13** | 61.2 | 0.05 |

OR, odds ratio per standard deviation (SD) increase in genetically predicted DNA methylation level; CI, confidence interval; TSS, transcription start site; UTR, untranslated region.

[a] Association Z scores, ORs, 95% CIs, and P values were estimated using SPrediXcan. P values that are less than 2.60E-07 (Bonferroni-corrected threshold for 192,188 tested CpGs) are highlighted in bold. All statistical tests were two-sided.[b] Correlation coefficients of predicted and measured DNA methylation levels.

**Supplementary Table 6. CpGs showing a significant association with prostate cancer risk**

| CpG | Chr | Pos | Cytoband | Z score [a] | *P* [a] | R [b] | Closest gene | Classification |
|---|---|---|---|---|---|---|---|---|
| cg09229181 | 1 | 150,521,241 | 1q21.3 | -5.33 | 9.75E-08 | 0.39 | *ADAMTSL4* | TSS1500 |
| cg07561848 | 1 | 150,551,931 | 1q21.3 | 5.21 | 1.89E-07 | 0.28 | *MCL1* | Exonic |
| cg07382149 | 1 | 150,601,807 | 1q21.3 | -7.85 | 4.05E-15 | 0.29 | *ENSA* | Intronic |
| cg08202226 | 1 | 153,896,590 | 1q21.3 | -5.67 | 1.41E-08 | 0.52 | *GATAD2B* ; *DENND4B* | Intergenic |
| cg16262392 | 1 | 153,932,122 | 1q21.3 | 5.23 | 1.74E-07 | 0.27 | *SLC39A1* | 3'UTR |
| cg04419630 | 1 | 153,932,221 | 1q21.3 | 6.44 | 1.16E-10 | 0.47 | *SLC39A1* | 3'UTR |
| cg03655142 | 1 | 153,950,843 | 1q21.3 | 5.66 | 1.53E-08 | 0.40 | *JTB* | TSS1500 |
| cg06221963 | 1 | 154,839,813 | 1q21.3 | -6.29 | 3.09E-10 | 0.79 | *KCNN3* | Intronic |
| cg09359103 | 1 | 154,839,909 | 1q21.3 | -6.30 | 3.06E-10 | 0.80 | *KCNN3* | Intronic |
| cg16680214 | 1 | 154,839,983 | 1q21.3 | -6.30 | 2.96E-10 | 0.72 | *KCNN3* | Intronic |
| cg03003668 | 1 | 154,840,889 | 1q21.3 | -6.13 | 8.72E-10 | 0.49 | *KCNN3* | Intronic |
| cg20543366 | 1 | 154,841,288 | 1q21.3 | -6.32 | 2.56E-10 | 0.28 | *KCNN3* | Intronic |
| cg24488589 | 1 | 154,850,415 | 1q21.3 | 6.55 | 5.72E-11 | 0.65 | *KCNN3* ; *PMVK* | Intergenic |
| cg24250549 | 1 | 154,909,240 | 1q21.3 | 6.26 | 3.79E-10 | 0.78 | *PMVK* | Intronic |
| cg03351412 | 1 | 154,909,251 | 1q21.3 | 6.21 | 5.46E-10 | 0.53 | *PMVK* | Intronic |
| cg14706141 | 1 | 155,030,538 | 1q22 | -6.02 | 1.71E-09 | 0.46 | *ADAM15* | Exonic |
| cg22531371 | 1 | 155,162,756 | 1q22 | 5.54 | 3.01E-08 | 0.28 | *MUC1* | TSS200 |
| cg17852453 | 1 | 204,426,281 | 1q32.1 | -5.60 | 2.18E-08 | 0.25 | *PIK3C2B* | Intronic |
| cg21195376 | 1 | 204,449,827 | 1q32.1 | -8.55 | 1.23E-17 | 0.33 | *PIK3C2B* | Intronic |
| cg01064725 | 1 | 204,461,714 | 1q32.1 | -10.16 | 3.03E-24 | 0.33 | *PIK3C2B* ; *MDM4* | Intergenic |
| cg20240347 | 1 | 204,465,584 | 1q32.1 | -9.44 | 3.68E-21 | 0.31 | *PIK3C2B* ; *MDM4* | Intergenic |
| cg10661893 | 1 | 204,482,706 | 1q32.1 | -8.91 | 4.92E-19 | 0.30 | *PIK3C2B* ; *MDM4* | Intergenic |
| cg15097530 | 1 | 204,486,394 | 1q32.1 | 9.42 | 4.57E-21 | 0.30 | *MDM4* | Intronic |
| cg07972458 | 1 | 204,535,000 | 1q32.1 | 9.71 | 2.70E-22 | 0.43 | *MDM4* ; *LRRN2* | Intergenic |
| cg21805731 | 1 | 204,560,985 | 1q32.1 | 6.21 | 5.18E-10 | 0.28 | *MDM4* ; *LRRN2* | Intergenic |
| cg20451944 | 1 | 204,606,773 | 1q32.1 | -6.96 | 3.52E-12 | 0.26 | *LRRN2* | Intronic |
| cg26778733 | 1 | 205,646,435 | 1q32.1 | -5.42 | 6.12E-08 | 0.21 | *SLC45A3* | Intronic |
| cg11143987 | 1 | 205,671,920 | 1q32.1 | -5.88 | 4.17E-09 | 0.40 | *SLC45A3* ; *NUCKS1* | Intergenic |
| cg16031515 | 1 | 205,743,344 | 1q32.1 | 5.88 | 4.16E-09 | 0.47 | *RAB29* | Intronic |
| cg26418147 | 1 | 205,743,515 | 1q32.1 | 5.30 | 1.14E-07 | 0.65 | *RAB29* | Intronic |
| cg13453750 | 1 | 205,783,389 | 1q32.1 | 5.28 | 1.28E-07 | 0.73 | *SLC41A1* ; *PM20D1* | Intergenic |
| cg08720477 | 1 | 205,799,480 | 1q32.1 | -5.51 | 3.68E-08 | 0.48 | *PM20D1* | Exonic |
| cg08750521 | 1 | 205,799,640 | 1q32.1 | -5.71 | 1.13E-08 | 0.31 | *PM20D1* | Intronic |
| cg06815965 | 1 | 205,818,668 | 1q32.1 | -5.22 | 1.79E-07 | 0.45 | *PM20D1* | Intronic |
| cg26354017 | 1 | 205,819,088 | 1q32.1 | -5.25 | 1.51E-07 | 0.78 | *PM20D1* | Exonic |
| cg14159672 | 1 | 205,819,179 | 1q32.1 | -5.23 | 1.72E-07 | 0.77 | *PM20D1* | Exonic |
| cg14893161 | 1 | 205,819,251 | 1q32.1 | -5.22 | 1.80E-07 | 0.78 | *PM20D1* | 5'UTR |
| cg07533224 | 1 | 205,819,345 | 1q32.1 | -5.16 | 2.52E-07 | 0.77 | *PM20D1* | TSS200 |
| cg05841700 | 1 | 205,819,383 | 1q32.1 | -5.21 | 1.84E-07 | 0.76 | *PM20D1* | TSS200 |
| cg11965913 | 1 | 205,819,406 | 1q32.1 | -5.17 | 2.32E-07 | 0.79 | *PM20D1* | TSS200 |
| cg07167872 | 1 | 205,819,463 | 1q32.1 | -5.20 | 1.94E-07 | 0.77 | *PM20D1* | TSS200 |
| cg24503407 | 1 | 205,819,492 | 1q32.1 | -5.22 | 1.76E-07 | 0.77 | *PM20D1* | TSS1500 |
| cg16334093 | 1 | 205,819,600 | 1q32.1 | -5.17 | 2.38E-07 | 0.80 | *PM20D1* | TSS1500 |
| cg07157834 | 1 | 205,819,609 | 1q32.1 | -5.24 | 1.64E-07 | 0.75 | *PM20D1* | TSS1500 |
| cg00947999 | 1 | 205,868,664 | 1q32.1 | -5.79 | 7.14E-09 | 0.46 | *LOC284581; SLC26A9* | Intergenic |
| cg22862005 | 1 | 205,884,321 | 1q32.1 | 5.56 | 2.63E-08 | 0.28 | *SLC26A9* | 3'UTR |
| cg21828629 | 1 | 205,913,422 | 1q32.1 | -5.96 | 2.51E-09 | 0.29 | *SLC26A9* | TSS1500 |
| cg12588880 | 2 | 8,597,159 | 2p25.1 | -5.85 | 4.96E-09 | 0.54 | *LINC00299* ; *LINC01814* | Intergenic |
| cg05204532 | 2 | 8,597,177 | 2p25.1 | -5.98 | 2.26E-09 | 0.50 | *LINC00299* ; *LINC01814* | Intergenic |
| cg10105712 | 2 | 8,597,188 | 2p25.1 | -5.71 | 1.10E-08 | 0.40 | *LINC00299* ; *LINC01814* | Intergenic |
| cg09590377 | 2 | 8,597,389 | 2p25.1 | -5.84 | 5.13E-09 | 0.38 | *LINC00299* ; *LINC01814* | Intergenic |
| cg10453602 | 2 | 8,597,867 | 2p25.1 | 5.30 | 1.13E-07 | 0.38 | *LINC00299* ; *LINC01814* | Intergenic |
| cg21917740 | 2 | 8,597,923 | 2p25.1 | -5.48 | 4.16E-08 | 0.34 | *LINC00299* ; *LINC01814* | Intergenic |
| cg14124917 | 2 | 10,090,974 | 2p25.1 | -5.27 | 1.39E-07 | 0.37 | *GRHL1* | TSS1500 |
| cg00105475 | 2 | 10,696,890 | 2p25.1 | -7.37 | 1.65E-13 | 0.54 | *ODC1-DT* ; *NOL10* | Intergenic |

| | | | | | | | | |
|---|---|---|---|---|---|---|---|---|
| cg20870561 | 2 | 10,728,572 | 2p25.1 | -5.34 | 9.36E-08 | 0.27 | *NOL10* | Intronic |
| cg02310191 | 2 | 10,731,329 | 2p25.1 | -8.08 | 6.73E-16 | 0.36 | *NOL10* | Intronic |
| cg01299579 | 2 | 10,830,716 | 2p25.1 | 6.82 | 9.26E-12 | 0.52 | *RN7SL832P* | Exonic |
| cg23893833 | 2 | 10,857,880 | 2p25.1 | 5.55 | 2.87E-08 | 0.36 | *RN7SL832P* ; *ATP6V1C2* | Intergenic |
| cg14780466 | 2 | 20,870,812 | 2p24.1 | -6.21 | 5.35E-10 | 0.44 | *GDF7* | Exonic |
| cg00283986 | 2 | 20,875,062 | 2p24.1 | 7.00 | 2.49E-12 | 0.38 | *GDF7* | 3'UTR |
| cg04689829 | 2 | 43,390,215 | 2p21 | 5.13 | 2.90E-07 | 0.24 | *LINC02580* ; *ZFP36L2* | Intergenic |
| cg09483650 | 2 | 43,634,819 | 2p21 | -6.82 | 8.97E-12 | 0.26 | *THADA* | Intronic |
| cg21555271 | 2 | 43,826,133 | 2p21 | -5.23 | 1.67E-07 | 0.24 | *THADA* ; *PLEKHH2* | Intergenic |
| cg15461431 | 2 | 43,836,757 | 2p21 | 7.59 | 3.12E-14 | 0.32 | *THADA* ; *PLEKHH2* | Intergenic |
| cg20563666 | 2 | 43,903,842 | 2p21 | -5.66 | 1.48E-08 | 0.24 | *PLEKHH2* | Intronic |
| cg27658811 | 2 | 62,494,078 | 2p15 | -6.22 | 5.10E-10 | 0.28 | *B3GNT2* ; *TMEM17* | Intergenic |
| cg09927842 | 2 | 62,684,417 | 2p15 | -8.31 | 9.61E-17 | 0.32 | *B3GNT2* ; *TMEM17* | Intergenic |
| cg15284384 | 2 | 62,685,556 | 2p15 | 5.38 | 7.48E-08 | 0.59 | *B3GNT2* ; *TMEM17* | Intergenic |
| cg14210094 | 2 | 62,888,566 | 2p15 | -5.31 | 1.07E-07 | 0.26 | *TMEM17* ; *EHBP1* | Intergenic |
| cg19352433 | 2 | 62,889,918 | 2p15 | 9.88 | 5.31E-23 | 0.35 | *TMEM17* ; *EHBP1* | Intergenic |
| cg13883027 | 2 | 62,892,060 | 2p15 | -8.22 | 2.12E-16 | 0.65 | *TMEM17* ; *EHBP1* | Intergenic |
| cg00883311 | 2 | 62,932,096 | 2p15 | 7.30 | 2.86E-13 | 0.31 | *EHBP1* | Intronic |
| cg27099154 | 2 | 62,987,767 | 2p15 | 8.93 | 4.10E-19 | 0.45 | *EHBP1* | Intronic |
| cg21115199 | 2 | 63,086,668 | 2p15 | -7.12 | 1.11E-12 | 0.68 | *EHBP1* | Intronic |
| cg27648677 | 2 | 63,086,804 | 2p15 | -8.60 | 7.64E-18 | 0.50 | *EHBP1* | Intronic |
| cg19825503 | 2 | 63,155,443 | 2p15 | 5.65 | 1.58E-08 | 0.31 | *EHBP1* | Intronic |
| cg17397364 | 2 | 63,273,436 | 2p15 | 9.89 | 4.80E-23 | 0.24 | *LOC100132215* | Intronic |
| cg15853117 | 2 | 63,276,966 | 2p15 | -6.23 | 4.72E-10 | 0.26 | *OTX1* | TSS1500 |
| cg17519650 | 2 | 63,277,830 | 2p15 | 14.17 | 1.42E-45 | 0.59 | *OTX1* | Intronic |
| cg27364741 | 2 | 63,281,069 | 2p15 | 11.42 | 3.21E-30 | 0.22 | *OTX1* | Intronic |
| cg14297410 | 2 | 63,351,486 | 2p15 | -6.12 | 9.64E-10 | 0.23 | *WDPCP* | Intronic |
| cg16932552 | 2 | 63,815,797 | 2p15 | -8.83 | 1.08E-18 | 0.23 | *MDH1* ; *WDPCP* | TSS1500 |
| cg25043210 | 2 | 63,872,394 | 2p15 | 6.32 | 2.69E-10 | 0.55 | *MDH1* ; *UGP2* | Intergenic |
| cg10172864 | 2 | 64,069,452 | 2p15 | -5.40 | 6.81E-08 | 0.30 | *UGP2* | Intronic |
| cg24018736 | 2 | 85,294,691 | 2p11.2 | -7.83 | 4.92E-15 | 0.28 | *LINC01964* | Upstream |
| cg24504864 | 2 | 85,665,051 | 2p11.2 | 5.76 | 8.34E-09 | 0.28 | *SH2D6* | Downstream |
| cg24330906 | 2 | 85,765,176 | 2p11.2 | -6.24 | 4.49E-10 | 0.46 | *PARTICL* | Exonic |
| cg17127132 | 2 | 85,788,382 | 2p11.2 | -8.64 | 5.68E-18 | 0.48 | *GGCX* | Intronic |
| cg23752985 | 2 | 85,803,571 | 2p11.2 | -9.56 | 1.16E-21 | 0.44 | *GGCX* ; *VAMP8* | Intergenic |
| cg02493740 | 2 | 85,810,744 | 2p11.2 | 10.06 | 8.59E-24 | 0.76 | *VAMP5* | TSS1500 |
| cg23902361 | 2 | 85,810,844 | 2p11.2 | 9.37 | 7.47E-21 | 0.47 | *VAMP5* | TSS1500 |
| cg25178838 | 2 | 85,818,634 | 2p11.2 | -5.67 | 1.43E-08 | 0.55 | *VAMP5* | Intronic |
| cg21117768 | 2 | 85,829,779 | 2p11.2 | -6.58 | 4.63E-11 | 0.21 | *TMEM150A* | 5'UTR |
| cg24081470 | 2 | 85,890,816 | 2p11.2 | 5.88 | 4.16E-09 | 0.32 | *SFTPB* | Exonic |
| cg10919674 | 2 | 85,976,079 | 2p11.2 | 5.85 | 4.99E-09 | 0.27 | *GNLY* ; *ATOH8* | Intergenic |
| cg05731808 | 2 | 173,293,392 | 2q31.1 | -5.80 | 6.67E-09 | 0.35 | *ITGA6* | Intronic |
| cg21907768 | 2 | 173,306,941 | 2q31.1 | -7.72 | 1.15E-14 | 0.21 | *ITGA6* | Intronic |
| cg07466807 | 2 | 173,326,947 | 2q31.1 | 6.31 | 2.83E-10 | 0.33 | *ITGA6* | Intronic |
| cg10165864 | 2 | 173,419,899 | 2q31.1 | -8.05 | 8.37E-16 | 0.27 | *PDK1* | TSS1500 |
| cg01937628 | 2 | 173,419,909 | 2q31.1 | -6.44 | 1.18E-10 | 0.41 | *PDK1* | TSS1500 |
| cg07833011 | 2 | 173,419,951 | 2q31.1 | -7.39 | 1.46E-13 | 0.46 | *PDK1* | TSS1500 |
| cg17679246 | 2 | 173,420,265 | 2q31.1 | -8.26 | 1.49E-16 | 0.47 | *PDK1* | TSS1500 |
| cg25617952 | 2 | 173,421,844 | 2q31.1 | -7.98 | 1.52E-15 | 0.27 | *PDK1* | Intronic |
| cg11269324 | 2 | 173,608,957 | 2q31.1 | 5.41 | 6.24E-08 | 0.26 | *RAPGEF4* | Intronic |
| cg12465398 | 2 | 201,753,874 | 2q33.1 | 6.38 | 1.81E-10 | 0.22 | *NIF3L1* ; *PPIL3* | TSS1500 |
| cg16040230 | 2 | 202,022,618 | 2q33.1 | -5.16 | 2.52E-07 | 0.59 | *CFLAR* | Intronic |
| cg17475949 | 2 | 202,050,400 | 2q33.1 | -5.86 | 4.65E-09 | 0.26 | *CASP10* | Intronic |
| cg00169121 | 2 | 238,379,873 | 2q37.3 | -6.13 | 8.80E-10 | 0.51 | *COL6A3* ; *MLPH* | Intergenic |
| cg14458575 | 2 | 238,380,390 | 2q37.3 | 6.55 | 5.93E-11 | 0.68 | *COL6A3* ; *MLPH* | Intergenic |
| cg03190769 | 2 | 238,382,720 | 2q37.3 | -6.18 | 6.21E-10 | 0.24 | *COL6A3* ; *MLPH* | Intergenic |
| cg16989719 | 2 | 238,392,110 | 2q37.3 | -6.13 | 8.83E-10 | 0.41 | *COL6A3* ; *MLPH* | Intergenic |
| cg08992911 | 2 | 238,395,768 | 2q37.3 | 6.63 | 3.31E-11 | 0.56 | *MLPH* | Intronic |

| | | | | | | | | |
|---|---|---|---|---|---|---|---|---|
| cg16426459 | 2 | 238,396,155 | 2q37.3 | 5.75 | 8.76E-09 | 0.31 | *MLPH* | Intronic |
| cg06639874 | 2 | 238,417,703 | 2q37.3 | -6.24 | 4.46E-10 | 0.27 | *MLPH* | Intronic |
| cg24827071 | 2 | 238,446,774 | 2q37.3 | -6.70 | 2.05E-11 | 0.28 | *MLPH* | Intronic |
| cg09067559 | 2 | 238,447,184 | 2q37.3 | 5.51 | 3.66E-08 | 0.32 | *MLPH* | Intronic |
| cg16487647 | 2 | 242,089,276 | 2q37.3 | 5.66 | 1.55E-08 | 0.21 | *PPP1R7* | 5'UTR |
| cg13886799 | 2 | 242,127,948 | 2q37.3 | 5.77 | 7.71E-09 | 0.40 | *ANO7* | 5'UTR |
| cg01752462 | 2 | 242,156,822 | 2q37.3 | -6.07 | 1.32E-09 | 0.27 | *ANO7* | Intronic |
| cg09235936 | 2 | 242,170,211 | 2q37.3 | -6.80 | 1.02E-11 | 0.21 | *HDLBP* | Exonic |
| cg21155796 | 2 | 242,212,141 | 2q37.3 | 8.06 | 7.53E-16 | 0.32 | *HDLBP* | Intronic |
| cg02590516 | 2 | 242,254,326 | 2q37.3 | 6.54 | 6.12E-11 | 0.28 | *HDLBP* | Intronic |
| cg17284139 | 2 | 242,444,691 | 2q37.3 | 5.38 | 7.48E-08 | 0.33 | *STK25* | Intronic |
| cg13220917 | 2 | 242,480,249 | 2q37.3 | 5.21 | 1.89E-07 | 0.37 | *STK25* ; *BOK-AS1* | Intergenic |
| cg23840860 | 2 | 242,572,608 | 2q37.3 | -6.14 | 8.45E-10 | 0.27 | *THAP4* | Exonic |
| cg02636222 | 3 | 48,956,935 | 3p21.31 | -5.28 | 1.29E-07 | 0.27 | *ARIH2* | Intronic |
| cg17991812 | 3 | 86,823,205 | 3p12.1 | -5.88 | 4.14E-09 | 0.37 | *LINC02070* ; *VGLL3* | Intergenic |
| cg04477660 | 3 | 87,137,900 | 3p12.1 | 6.60 | 4.11E-11 | 0.38 | *LINC00506* | TSS1500 |
| cg01807408 | 3 | 87,137,933 | 3p12.1 | 6.64 | 3.03E-11 | 0.42 | *LINC00506* | TSS1500 |
| cg23835812 | 3 | 87,276,149 | 3p11.2 | 5.18 | 2.24E-07 | 0.21 | *CHMP2B* | TSS1500 |
| cg14650344 | 3 | 87,276,155 | 3p11.2 | 5.51 | 3.65E-08 | 0.33 | *CHMP2B* ; *MIR4795* | TSS1500 |
| cg17693183 | 3 | 112,951,443 | 3q13.2 | -6.58 | 4.73E-11 | 0.55 | *BOC* | Intronic |
| cg20083609 | 3 | 112,980,454 | 3q13.2 | 8.29 | 1.13E-16 | 0.45 | *BOC* | Intronic |
| cg11950460 | 3 | 113,046,629 | 3q13.2 | 6.30 | 2.90E-10 | 0.30 | *CFAP44* | Exonic |
| cg15044041 | 3 | 113,160,313 | 3q13.2 | 5.91 | 3.43E-09 | 0.23 | *CFAP44* | 5'UTR |
| cg02743293 | 3 | 113,250,921 | 3q13.2 | 7.37 | 1.76E-13 | 0.44 | *SIDT1* | TSS1500 |
| cg12596171 | 3 | 113,251,061 | 3q13.2 | 7.31 | 2.64E-13 | 0.62 | *SIDT1* | TSS200 |
| cg11695283 | 3 | 113,251,181 | 3q13.2 | 7.32 | 2.51E-13 | 0.54 | *SIDT1* | TSS200 |
| cg11138929 | 3 | 113,251,189 | 3q13.2 | 7.26 | 3.96E-13 | 0.53 | *SIDT1* | TSS200 |
| cg00726615 | 3 | 113,252,640 | 3q13.2 | 7.64 | 2.19E-14 | 0.31 | *SIDT1* | Intronic |
| cg01693830 | 3 | 127,790,462 | 3q21.3 | -6.26 | 3.81E-10 | 0.45 | *SEC61A1* | 3'UTR |
| cg06048819 | 3 | 127,794,219 | 3q21.3 | -5.49 | 4.06E-08 | 0.39 | *RUVBL1* | Intronic |
| cg10056460 | 3 | 127,794,257 | 3q21.3 | -6.26 | 3.84E-10 | 0.27 | *RUVBL1* | Intronic |
| cg13719885 | 3 | 127,795,394 | 3q21.3 | -5.36 | 8.50E-08 | 0.57 | *RUVBL1-AS1* | Intronic |
| cg02510924 | 3 | 127,802,658 | 3q21.3 | 6.38 | 1.83E-10 | 0.31 | *RUVBL1* | Intronic |
| cg03115093 | 3 | 127,861,051 | 3q21.3 | -8.40 | 4.31E-17 | 0.48 | *RUVBL1* | Intronic |
| cg27019594 | 3 | 127,892,866 | 3q21.3 | 7.80 | 6.23E-15 | 0.54 | *EEFSEC* | Intronic |
| cg09718996 | 3 | 127,895,237 | 3q21.3 | 11.38 | 5.00E-30 | 0.55 | *EEFSEC* | Intronic |
| cg14972564 | 3 | 127,935,096 | 3q21.3 | -8.04 | 8.90E-16 | 0.29 | *EEFSEC* | Intronic |
| cg00200020 | 3 | 127,937,549 | 3q21.3 | -5.39 | 7.06E-08 | 0.32 | *EEFSEC* | Intronic |
| cg00340326 | 3 | 127,988,088 | 3q21.3 | -7.82 | 5.25E-15 | 0.65 | *EEFSEC* | Intronic |
| cg15808905 | 3 | 128,038,392 | 3q21.3 | -11.21 | 3.60E-29 | 0.31 | *EEFSEC* | Intronic |
| cg21710443 | 3 | 128,055,577 | 3q21.3 | -7.01 | 2.35E-12 | 0.60 | *EEFSEC* | Intronic |
| cg14208839 | 3 | 128,080,980 | 3q21.3 | 6.20 | 5.65E-10 | 0.33 | *EEFSEC* | Intronic |
| cg15846910 | 3 | 128,081,006 | 3q21.3 | 6.88 | 5.92E-12 | 0.31 | *EEFSEC* | Intronic |
| cg17068417 | 3 | 128,081,087 | 3q21.3 | 6.43 | 1.25E-10 | 0.28 | *EEFSEC* | Intronic |
| cg08443130 | 3 | 128,117,563 | 3q21.3 | 5.86 | 4.74E-09 | 0.34 | *EEFSEC* | Intronic |
| cg05487005 | 3 | 128,148,610 | 3q21.3 | 5.68 | 1.38E-08 | 0.30 | *EEFSEC* ; *DNAJB8* | Intergenic |
| cg02836487 | 3 | 128,206,457 | 3q21.3 | -6.50 | 7.82E-11 | 0.38 | *GATA2* | Intronic |
| cg14795409 | 3 | 128,267,611 | 3q21.3 | -5.16 | 2.48E-07 | 0.24 | *LOC90246* ; *LINC01565* | Intergenic |
| cg21411781 [c] | 3 | 131,984,709 | 3q22.1 | -5.31 | 1.11E-07 | 0.20 | *CPNE4* ; *ACPP* | Intergenic |
| cg14222479 | 3 | 169,487,675 | 3q26.2 | -5.16 | 2.52E-07 | 0.29 | *ACTRT3* | Upstream |
| cg17747518 | 3 | 169,725,031 | 3q26.2 | -5.14 | 2.78E-07 | 0.59 | *SEC62* ; *GPR160* | Intergenic |
| cg25314364 | 3 | 169,899,582 | 3q26.2 | 5.66 | 1.52E-08 | 0.23 | *PHC3* | TSS200 |
| cg04067573 | 3 | 169,899,625 | 3q26.2 | -9.40 | 5.44E-21 | 0.42 | *PHC3* | TSS200 |
| cg11886554 [d] | 3 | 170,076,028 | 3q26.2 | 5.12 | 3.01E-07 | 0.67 | *SKIL* | Intronic |
| cg21678033 | 3 | 170,127,982 | 3q26.2 | 13.10 | 3.29E-39 | 0.22 | *SKIL* ; *CLDN11* | Intergenic |
| cg00894757 | 3 | 170,137,240 | 3q26.2 | 5.48 | 4.24E-08 | 0.31 | *CLDN11* | Intronic |
| cg15776647 | 3 | 170,159,062 | 3q26.2 | -5.17 | 2.37E-07 | 0.20 | *CLDN11* ; *SLC7A14* | Intergenic |

| | | | | | | | | |
|---|---|---|---|---|---|---|---|---|
| cg21743924 | 3 | 170,470,815 | 3q26.2 | -7.29 | 3.00E-13 | 0.21 | *SLC7A14-AS1* | Intronic |
| cg09591065 | 3 | 170,483,432 | 3q26.2 | -8.82 | 1.11E-18 | 0.32 | *SLC7A14-AS1* ; *RPL22L1* | Intergenic |
| cg17549886 | 4 | 73,891,141 | 4q13.3 | 6.19 | 5.88E-10 | 0.33 | *ADAMTS3* ; *COX18* | Intergenic |
| cg21511758 | 4 | 74,129,609 | 4q13.3 | -8.07 | 7.23E-16 | 0.20 | *ANKRD17* ; *ALB* | Intergenic |
| cg15567016 | 4 | 74,174,284 | 4q13.3 | 5.18 | 2.27E-07 | 0.44 | *ANKRD17* ; *ALB* | Intergenic |
| cg10270595 | 4 | 74,472,965 | 4q13.3 | -7.58 | 3.58E-14 | 0.42 | *RASSF6* | Intronic |
| cg12566645 | 4 | 74,483,141 | 4q13.3 | -6.53 | 6.47E-11 | 0.52 | *RASSF6* | Intronic |
| cg20368506 | 4 | 74,486,546 | 4q13.3 | -5.50 | 3.79E-08 | 0.20 | *RASSF6* | TSS200 |
| cg20644745 | 4 | 74,519,591 | 4q13.3 | -6.87 | 6.35E-12 | 0.38 | *RASSF6* ; *CXCL8* | Intergenic |
| cg11021082 | 4 | 95,130,006 | 4q22.3 | -6.52 | 6.84E-11 | 0.25 | *SMARCAD1* | Intronic |
| cg05904325 | 4 | 95,441,526 | 4q22.3 | 7.39 | 1.49E-13 | 0.37 | *PDLIM5* | Intronic |
| cg26612230 | 4 | 95,508,526 | 4q22.3 | -8.19 | 2.63E-16 | 0.34 | *PDLIM5* | 3'UTR |
| cg00096603 | 4 | 95,572,122 | 4q22.3 | 8.39 | 4.74E-17 | 0.23 | *PDLIM5* | Intronic |
| cg07677047 | 4 | 95,587,592 | 4q22.3 | 10.46 | 1.26E-25 | 0.46 | *PDLIM5* | 3'UTR |
| cg22449691 | 4 | 106,062,757 | 4q24 | 5.69 | 1.27E-08 | 0.20 | *CXXC4-AS1* ; *TET2* | Intergenic |
| cg17862558 | 4 | 106,137,048 | 4q24 | 12.38 | 3.40E-35 | 0.33 | *TET2-AS1* | Intronic |
| cg13529269 | 4 | 106,395,383 | 4q24 | 9.21 | 3.34E-20 | 0.35 | *PPA2* | TSS200 |
| cg12993807 | 5 | 1,075,045 | 5p15.33 | -5.20 | 1.99E-07 | 0.31 | *SLC12A7* | Intronic |
| cg11628781 | 5 | 1,089,021 | 5p15.33 | 7.55 | 4.28E-14 | 0.32 | *SLC12A7* | Intronic |
| cg23337501 | 5 | 1,225,328 | 5p15.33 | 7.26 | 3.87E-13 | 0.22 | *SLC6A18* | TSS200 |
| cg05024157 | 5 | 1,238,988 | 5p15.33 | 5.14 | 2.76E-07 | 0.45 | *SLC6A18* | Intronic |
| cg00624418 | 5 | 1,282,357 | 5p15.33 | 12.95 | 2.50E-38 | 0.67 | *TERT* | Intronic |
| cg03935379 | 5 | 1,282,703 | 5p15.33 | 13.04 | 7.52E-39 | 0.63 | *TERT* | Exonic |
| cg06531176 | 5 | 1,282,723 | 5p15.33 | 9.06 | 1.36E-19 | 0.47 | *TERT* | Exonic |
| cg05599246 | 5 | 1,343,592 | 5p15.33 | -5.35 | 8.98E-08 | 0.33 | *CLPTM1L* | Intronic |
| cg17629929 | 5 | 1,875,548 | 5p15.33 | -5.15 | 2.66E-07 | 0.21 | *LINC02116* ; *IRX4* | Intergenic |
| cg07961015 | 5 | 1,876,215 | 5p15.33 | -5.63 | 1.82E-08 | 0.26 | *LINC02116* ; *IRX4* | Intergenic |
| cg14451382 | 5 | 1,876,397 | 5p15.33 | -5.37 | 8.02E-08 | 0.30 | *LINC02116* ; *IRX4* | Intergenic |
| cg21534994 | 5 | 1,881,655 | 5p15.33 | -5.30 | 1.16E-07 | 0.24 | *IRX4* | Intronic |
| cg06333078 | 5 | 1,897,253 | 5p15.33 | -8.97 | 2.92E-19 | 0.41 | *CTD-2194D22.4* | Intronic |
| cg11538020 | 5 | 2,208,611 | 5p15.33 | -6.33 | 2.38E-10 | 0.26 | *CTD-2194D22.4* ; *LOC100506858* | Intergenic |
| cg00922210 | 5 | 133,880,083 | 5q31.1 | -5.17 | 2.37E-07 | 0.32 | *JADE2* | Intronic |
| cg14758967 | 5 | 133,884,935 | 5q31.1 | 5.20 | 1.98E-07 | 0.34 | *JADE2* | Intronic |
| cg03652456 | 6 | 1,670,293 | 6p25.3 | 5.16 | 2.47E-07 | 0.66 | *GMDS* | Intronic |
| cg07019595 | 6 | 11,139,578 | 6p24.2 | -5.59 | 2.21E-08 | 0.29 | *SMIM13* | Downstream |
| cg14563924 | 6 | 11,219,554 | 6p24.2 | -6.20 | 5.58E-10 | 0.32 | *NEDD9* | Intronic |
| cg12061917 | 6 | 29,601,556 | 6p22.1 | -5.97 | 2.37E-09 | 0.26 | *GABBR1* | TSS1500 |
| cg20891813 | 6 | 29,691,735 | 6p22.1 | 5.36 | 8.32E-08 | 0.50 | *HLA-F* | Intronic |
| cg17221604 | 6 | 29,720,989 | 6p22.1 | 5.27 | 1.36E-07 | 0.23 | *IFITM4P* ; *HCG4* | Intergenic |
| cg19141489 | 6 | 29,796,068 | 6p22.1 | -5.21 | 1.90E-07 | 0.27 | *HLA-G* | Exonic |
| cg10181330 | 6 | 29,818,243 | 6p22.1 | 5.25 | 1.53E-07 | 0.60 | *ZNRD1ASP* ; *HCP5B* | Intergenic |
| cg10821226 | 6 | 29,818,277 | 6p22.1 | 5.30 | 1.16E-07 | 0.59 | *ZNRD1ASP* ; *HCP5B* | Intergenic |
| cg18516125 | 6 | 29,924,207 | 6p22.1 | -5.52 | 3.44E-08 | 0.48 | *HLA-A* ; *HCG9* | Intergenic |
| cg16690356 | 6 | 29,945,948 | 6p22.1 | 5.16 | 2.52E-07 | 0.34 | *HCG9* | Intronic |
| cg19872019 | 6 | 29,980,960 | 6p22.1 | -5.54 | 3.10E-08 | 0.24 | *ZNRD1ASP* | Exonic |
| cg13044444 | 6 | 30,029,721 | 6p22.1 | -5.51 | 3.50E-08 | 0.28 | *ZNRD1* | Exonic |
| cg13919341 | 6 | 30,029,929 | 6p22.1 | -5.99 | 2.06E-09 | 0.27 | *ZNRD1* | Intronic |
| cg06155771 | 6 | 30,029,935 | 6p22.1 | -5.98 | 2.29E-09 | 0.34 | *ZNRD1* | Intronic |
| cg07468289 | 6 | 30,030,010 | 6p22.1 | -5.90 | 3.73E-09 | 0.33 | *ZNRD1* | Exonic |
| cg11963660 | 6 | 30,043,820 | 6p22.1 | 5.34 | 9.16E-08 | 0.27 | *RNF39* | TSS200 |
| cg24276787 | 6 | 30,045,068 | 6p22.1 | 5.87 | 4.38E-09 | 0.33 | *RNF39* ; *TRIM31* | Intergenic |
| cg11229497 | 6 | 30,078,954 | 6p22.1 | -5.46 | 4.89E-08 | 0.48 | *TRIM31-AS1* | Intronic |
| cg13927501 | 6 | 30,079,090 | 6p22.1 | -5.24 | 1.59E-07 | 0.40 | *TRIM31-AS1* | Intronic |
| cg27313566 | 6 | 30,079,155 | 6p22.1 | -5.19 | 2.08E-07 | 0.50 | *TRIM31-AS1* | Intronic |
| cg06893296 | 6 | 30,095,136 | 6p22.1 | -6.47 | 9.60E-11 | 0.22 | *TRIM31-AS1* ; *TRIM40* | Intergenic |
| cg12758147 | 6 | 30,103,696 | 6p22.1 | 6.14 | 8.15E-10 | 0.54 | *TRIM40* | TSS1500 |
| cg13044052 | 6 | 30,103,699 | 6p22.1 | 6.35 | 2.12E-10 | 0.58 | *TRIM40* | TSS1500 |

| | | | | | | | | |
|---|---|---|---|---|---|---|---|---|
| cg25540824 | 6 | 30,139,686 | 6p22.1 | -6.55 | 5.70E-11 | 0.64 | *TRIM15* | Exonic |
| cg12423902 | 6 | 30,159,309 | 6p22.1 | -5.58 | 2.37E-08 | 0.29 | *TRIM26* | Intronic |
| cg27136023 | 6 | 30,174,651 | 6p22.1 | -6.05 | 1.46E-09 | 0.39 | *TRIM26* | Intronic |
| cg00206463 | 6 | 30,179,435 | 6p22.1 | 6.58 | 4.58E-11 | 0.54 | *TRIM26* | Intronic |
| cg11409139 | 6 | 30,292,481 | 6p22.1 | 5.67 | 1.43E-08 | 0.58 | *HCG17* ; *HCG18* | Intronic |
| cg06063598 | 6 | 30,314,177 | 6p22.1 | -6.55 | 5.75E-11 | 0.44 | *RPP21* ; *TRIM39-RPP21* | Intronic |
| cg18804722 | 6 | 30,431,770 | 6p21.33 | -5.14 | 2.80E-07 | 0.28 | *RPP21* ; *HLA-E* | Intergenic |
| cg13518670 | 6 | 30,433,838 | 6p21.33 | -5.19 | 2.16E-07 | 0.25 | *RPP21* ; *HLA-E* | Intergenic |
| cg24766816 [d] | 6 | 30,568,535 | 6p21.33 | 6.81 | 9.64E-12 | 0.40 | *PPP1R10* | 3'UTR |
| cg23624891 | 6 | 30,570,123 | 6p21.33 | -6.86 | 7.12E-12 | 0.22 | *PPP1R10* | Exonic |
| cg01245721 | 6 | 30,594,037 | 6p21.33 | 5.20 | 1.98E-07 | 0.21 | *MRPS18B* | 3'UTR |
| cg07816366 | 6 | 30,594,749 | 6p21.33 | 5.35 | 8.93E-08 | 0.20 | *ATAT1* | Exonic |
| cg24851684 [d] | 6 | 30,624,160 | 6p21.33 | -6.02 | 1.70E-09 | 0.23 | *DHX16* | Intronic |
| cg12897786 | 6 | 30,640,295 | 6p21.33 | 7.24 | 4.57E-13 | 0.32 | *DHX16* | Intronic |
| cg13357602 | 6 | 30,655,720 | 6p21.33 | 6.37 | 1.89E-10 | 0.25 | *PPP1R18* ; *NRM* | TSS1500 |
| cg09703123 | 6 | 30,658,281 | 6p21.33 | 5.77 | 8.11E-09 | 0.49 | *NRM* | Intronic |
| cg19179185 | 6 | 30,731,397 | 6p21.33 | 6.25 | 4.13E-10 | 0.38 | *IER3* ; *HCG20* | Intergenic |
| cg20802826 [d] | 6 | 30,782,080 | 6p21.33 | 6.62 | 3.55E-11 | 0.34 | *LINC00243* | Exonic |
| cg18730733 | 6 | 30,816,447 | 6p21.33 | 5.74 | 9.40E-09 | 0.31 | *LINC02570* | Upstream |
| cg16213014 | 6 | 30,869,755 | 6p21.33 | -5.58 | 2.36E-08 | 0.21 | *DDR1* ; *GTF2H4* | Intergenic |
| cg15848685 | 6 | 30,882,641 | 6p21.33 | -5.19 | 2.05E-07 | 0.56 | *VARS2* | Exonic |
| cg12457901 | 6 | 30,882,671 | 6p21.33 | -5.26 | 1.43E-07 | 0.73 | *VARS2* | Exonic |
| cg13561028 [d] | 6 | 30,899,649 | 6p21.33 | 5.94 | 2.87E-09 | 0.62 | *SFTA2* | Intronic |
| cg12797044 | 6 | 30,922,043 | 6p21.33 | -5.53 | 3.20E-08 | 0.41 | *HCG21* | Intronic |
| cg03714531 | 6 | 30,923,590 | 6p21.33 | -5.42 | 5.89E-08 | 0.51 | *HCG21* | Upstream |
| cg09657173 | 6 | 30,923,593 | 6p21.33 | -5.19 | 2.14E-07 | 0.34 | *HCG21* | Upstream |
| cg16398511 | 6 | 31,010,701 | 6p21.33 | 5.23 | 1.71E-07 | 0.24 | *MUC22* ; *HCG22* | Intergenic |
| cg15583958 [d] | 6 | 31,055,396 | 6p21.33 | -5.37 | 7.85E-08 | 0.57 | *HCG22* ; *C6orf15* | Intergenic |
| cg03043553 [d] | 6 | 31,072,036 | 6p21.33 | -5.26 | 1.46E-07 | 0.50 | *HCG22* ; *C6orf15* | Intergenic |
| cg17323974 | 6 | 31,079,110 | 6p21.33 | -5.26 | 1.42E-07 | 0.37 | *C6orf15* | 3'UTR |
| cg08951273 | 6 | 31,079,152 | 6p21.33 | -5.87 | 4.34E-09 | 0.59 | *C6orf15* | 3'UTR |
| cg10185489 | 6 | 31,079,555 | 6p21.33 | -5.71 | 1.10E-08 | 0.28 | *C6orf15* | Exonic |
| cg08097157 [d] | 6 | 31,080,048 | 6p21.33 | -6.44 | 1.17E-10 | 0.37 | *C6orf15* | Exonic |
| cg17849733 [d] | 6 | 31,082,200 | 6p21.33 | 5.30 | 1.14E-07 | 0.71 | *PSORS1C1* | TSS1500 |
| cg04095776 [d] | 6 | 31,106,941 | 6p21.33 | 6.95 | 3.66E-12 | 0.42 | *PSORS1C1* | Intronic |
| cg12044337 | 6 | 31,107,195 | 6p21.33 | -5.38 | 7.35E-08 | 0.66 | *PSORS1C1* | Intronic |
| cg07613268 [d] | 6 | 31,110,473 | 6p21.33 | 7.06 | 1.69E-12 | 0.28 | *CCHCR1* | Exonic |
| cg02247838 | 6 | 31,110,639 | 6p21.33 | 6.12 | 9.49E-10 | 0.44 | *CCHCR1* | Intronic |
| cg02451928 | 6 | 31,125,948 | 6p21.33 | -6.11 | 1.02E-09 | 0.49 | *CCHCR1* | 5'UTR |
| cg04072928 [d] | 6 | 31,125,950 | 6p21.33 | -6.85 | 7.47E-12 | 0.51 | *CCHCR1* | 5'UTR |
| cg09045681 | 6 | 31,127,178 | 6p21.33 | -5.29 | 1.22E-07 | 0.32 | *TCF19* | 5'UTR |
| cg16626479 | 6 | 31,138,420 | 6p21.33 | -5.69 | 1.25E-08 | 0.44 | *POU5F1* | 5'UTR |
| cg03465464 [d] | 6 | 31,141,763 | 6p21.33 | -6.00 | 1.98E-09 | 0.29 | *PSORS1C3* | Exonic |
| cg04035638 [d] | 6 | 31,141,768 | 6p21.33 | -5.77 | 8.07E-09 | 0.21 | *PSORS1C3* | Exonic |
| cg02077524 | 6 | 31,145,335 | 6p21.33 | -5.41 | 6.32E-08 | 0.55 | *PSORS1C3* | Intronic |
| cg11805138 | 6 | 31,148,332 | 6p21.33 | 5.38 | 7.28E-08 | 0.38 | *PSORS1C3* | Intronic |
| cg17931227 | 6 | 31,148,370 | 6p21.33 | 6.38 | 1.77E-10 | 0.48 | *PSORS1C3* | Intronic |
| cg09179646 | 6 | 31,148,383 | 6p21.33 | 6.24 | 4.42E-10 | 0.47 | *PSORS1C3* | Intronic |
| cg22291762 [d] | 6 | 31,148,404 | 6p21.33 | 6.79 | 1.08E-11 | 0.46 | *PSORS1C3* | Intronic |
| cg24427850 [d] | 6 | 31,148,409 | 6p21.33 | 6.60 | 4.03E-11 | 0.44 | *PSORS1C3* | Intronic |
| cg26668675 [d] | 6 | 31,148,463 | 6p21.33 | 7.28 | 3.37E-13 | 0.41 | *PSORS1C3* | Intronic |
| cg03078486 [d] | 6 | 31,148,474 | 6p21.33 | 7.00 | 2.51E-12 | 0.38 | *PSORS1C3* | Intronic |
| cg22701603 [d] | 6 | 31,148,483 | 6p21.33 | 6.77 | 1.26E-11 | 0.47 | *PSORS1C3* | Intronic |
| cg26818629 | 6 | 31,148,516 | 6p21.33 | 6.20 | 5.49E-10 | 0.27 | *PSORS1C3* | Intronic |
| cg27547543 [d] | 6 | 31,148,524 | 6p21.33 | 6.35 | 2.15E-10 | 0.35 | *PSORS1C3* | Intronic |

| | | | | | | | | |
|---|---|---|---|---|---|---|---|---|
| cg09357589 | 6 | 31,148,552 | 6p21.33 | 6.05 | 1.42E-09 | 0.42 | *PSORS1C3* | Intronic |
| cg14036627 [d] | 6 | 31,148,657 | 6p21.33 | 6.55 | 5.90E-11 | 0.38 | *PSORS1C3* | Intronic |
| cg11811828 | 6 | 31,148,666 | 6p21.33 | 6.29 | 3.21E-10 | 0.39 | *PSORS1C3* | Intronic |
| cg23457357 [d] | 6 | 31,148,748 | 6p21.33 | 7.25 | 4.20E-13 | 0.41 | *PSORS1C3* | Intronic |
| cg05891181 | 6 | 31,165,581 | 6p21.33 | -5.46 | 4.74E-08 | 0.29 | *HCG27* | Exonic |
| cg18412521 | 6 | 31,220,536 | 6p21.33 | -5.24 | 1.63E-07 | 0.52 | *HCG27* ; *HLA-C* | Intergenic |
| cg06659144 | 6 | 31,239,939 | 6p21.33 | -5.59 | 2.22E-08 | 0.34 | *HLA-C* | TSS200 |
| cg17476405 | 6 | 31,371,614 | 6p21.33 | -5.15 | 2.57E-07 | 0.57 | *MICA* | Intronic |
| cg02956255 | 6 | 31,430,641 | 6p21.33 | 5.30 | 1.16E-07 | 0.31 | *HCP5* | TSS1500 |
| cg04664856 | 6 | 31,442,092 | 6p21.33 | -5.84 | 5.26E-09 | 0.44 | *HCG26* ; *MICB-DT* | Intergenic |
| cg10942339 | 6 | 31,474,043 | 6p21.33 | -6.25 | 4.11E-10 | 0.32 | *MICB* | Exonic |
| cg11112941 [c] | 6 | 31,491,116 | 6p21.33 | -6.66 | 2.76E-11 | 0.22 | *MICB* ; *MCCD1* | Intergenic |
| cg14646094 | 6 | 31,496,752 | 6p21.33 | -5.82 | 5.80E-09 | 0.46 | *MCCD1* | 5'UTR |
| cg16268563 | 6 | 31,513,680 | 6p21.33 | -5.31 | 1.09E-07 | 0.20 | *ATP6V1G2-DDX39B* | Intronic |
| cg19464633 | 6 | 31,515,291 | 6p21.33 | 5.63 | 1.80E-08 | 0.21 | *NFKBIL1* | Intronic |
| cg14166520 | 6 | 31,532,351 | 6p21.33 | -5.20 | 2.02E-07 | 0.28 | *LOC100287329* | Intronic |
| cg10650821 [c] | 6 | 31,543,686 | 6p21.33 | 6.98 | 2.96E-12 | 0.21 | *TNF* | Exonic |
| cg10373114 | 6 | 31,593,415 | 6p21.33 | 5.71 | 1.10E-08 | 0.23 | *PRRC2A* | Intronic |
| cg05182583 | 6 | 31,598,464 | 6p21.33 | -6.66 | 2.74E-11 | 0.28 | *PRRC2A* | Exonic |
| cg17006042 | 6 | 31,620,002 | 6p21.33 | -6.17 | 6.70E-10 | 0.29 | *BAG6* | Intronic |
| cg18753296 | 6 | 31,630,923 | 6p21.33 | -5.48 | 4.34E-08 | 0.43 | *GPANK1* | Intronic |
| cg11522612 | 6 | 31,687,637 | 6p21.33 | -5.16 | 2.47E-07 | 0.31 | *LY6G6C* | Intronic |
| cg19251326 | 6 | 31,688,080 | 6p21.33 | -6.84 | 8.17E-12 | 0.35 | *LY6G6C* | Intronic |
| cg09906448 | 6 | 31,688,160 | 6p21.33 | -5.94 | 2.88E-09 | 0.41 | *LY6G6C* | Intronic |
| cg20794828 | 6 | 31,688,229 | 6p21.33 | -7.45 | 9.24E-14 | 0.57 | *LY6G6C* | Intronic |
| cg22727233 | 6 | 31,688,428 | 6p21.33 | -5.84 | 5.28E-09 | 0.30 | *LY6G6C* | Intronic |
| cg16668548 | 6 | 31,697,545 | 6p21.33 | -7.26 | 3.94E-13 | 0.39 | *DDAH2* | Intronic |
| cg00110675 | 6 | 31,698,047 | 6p21.33 | -6.48 | 9.33E-11 | 0.71 | *DDAH2* ; *CLIC1* | TSS1500 |
| cg10563645 | 6 | 31,705,988 | 6p21.33 | -5.41 | 6.19E-08 | 0.27 | *CLIC1* | TSS1500 |
| cg18424091 | 6 | 31,781,814 | 6p21.33 | 6.39 | 1.70E-10 | 0.31 | *HSPA1L* | Intronic |
| cg09673200 | 6 | 31,803,103 | 6p21.33 | -5.95 | 2.72E-09 | 0.58 | *SNORD48* | Exonic |
| cg15480114 | 6 | 31,857,040 | 6p21.33 | 5.51 | 3.60E-08 | 0.25 | *EHMT2* | Exonic |
| cg22706070 | 6 | 31,857,895 | 6p21.33 | 5.34 | 9.53E-08 | 0.21 | *EHMT2* | Intronic |
| cg07723114 | 6 | 31,864,706 | 6p21.33 | -6.27 | 3.60E-10 | 0.23 | *EHMT2* | Exonic |
| cg04744301 | 6 | 31,936,154 | 6p21.33 | 5.17 | 2.34E-07 | 0.41 | *SKIV2L* | Exonic |
| cg03239580 | 6 | 31,973,051 | 6p21.33 | 6.31 | 2.84E-10 | 0.35 | *CYP21A1P* | TSS1500 |
| cg02432444 | 6 | 32,016,214 | 6p21.33 | 5.36 | 8.12E-08 | 0.21 | *TNXB* | Exonic |
| cg19964491 | 6 | 32,016,236 | 6p21.33 | 5.39 | 7.17E-08 | 0.33 | *TNXB* | Exonic |
| cg11493661 | 6 | 32,016,239 | 6p21.33 | 7.24 | 4.48E-13 | 0.31 | *TNXB* | Exonic |
| cg09474017 | 6 | 32,016,368 | 6p21.33 | 5.29 | 1.24E-07 | 0.42 | *TNXB* | Exonic |
| cg05598103 | 6 | 32,022,929 | 6p21.33 | 5.27 | 1.35E-07 | 0.66 | *TNXB* | Intronic |
| cg11005651 | 6 | 32,065,890 | 6p21.33 | 7.92 | 2.43E-15 | 0.22 | *TNXB* | Exonic |
| cg02390267 | 6 | 32,096,854 | 6p21.33 | 5.49 | 4.02E-08 | 0.31 | *FKBPL* | Exonic |
| cg07305776 | 6 | 32,097,549 | 6p21.33 | 5.89 | 3.82E-09 | 0.29 | *FKBPL* | Exonic |
| cg17229678 | 6 | 32,121,555 | 6p21.32 | 5.13 | 2.84E-07 | 0.41 | *LOC100507547* | Exonic |
| cg27134827 | 6 | 32,121,566 | 6p21.32 | 5.27 | 1.37E-07 | 0.32 | *LOC100507547* | Exonic |
| cg06024540 | 6 | 32,135,396 | 6p21.32 | -5.18 | 2.25E-07 | 0.24 | *EGFL8* | Exonic |
| cg03280235 | 6 | 32,158,953 | 6p21.32 | 5.26 | 1.40E-07 | 0.51 | *GPSM3* | 3'UTR |
| cg06194738 | 6 | 32,159,607 | 6p21.32 | 6.22 | 4.89E-10 | 0.23 | *GPSM3* | Exonic |
| cg06863878 | 6 | 32,168,732 | 6p21.32 | 6.30 | 2.97E-10 | 0.21 | *NOTCH4* | Exonic |
| cg20671150 | 6 | 32,172,151 | 6p21.32 | -7.25 | 4.12E-13 | 0.25 | *NOTCH4* | Exonic |
| cg27322595 | 6 | 32,181,814 | 6p21.32 | 5.59 | 2.29E-08 | 0.28 | *NOTCH4* | Intronic |
| cg14700707 | 6 | 32,191,840 | 6p21.32 | 6.18 | 6.53E-10 | 0.20 | *NOTCH4* | 5'UTR |
| cg07822788 | 6 | 32,192,350 | 6p21.32 | -5.61 | 2.00E-08 | 0.25 | *NOTCH4* | TSS1500 |
| cg27353721 | 6 | 32,202,748 | 6p21.32 | 5.51 | 3.50E-08 | 0.35 | *NOTCH4* ; *TSBP1-AS1* | Intergenic |
| cg08376583 | 6 | 32,222,492 | 6p21.32 | 6.41 | 1.42E-10 | 0.40 | *TSBP1-AS1* | Upstream |
| cg12898793 | 6 | 32,288,224 | 6p21.32 | -5.18 | 2.27E-07 | 0.56 | *TSBP1-AS1* | Intronic |

| | | | | | | | | |
|---|---|---|---|---|---|---|---|---|
| cg17039645 | 6 | 32,294,503 | 6p21.32 | -5.15 | 2.58E-07 | 0.35 | *TSBP1-AS1* | Intronic |
| cg02957632 | 6 | 32,308,995 | 6p21.32 | -5.53 | 3.26E-08 | 0.56 | *TSBP1-AS1* | Intronic |
| cg05198093 | 6 | 32,315,772 | 6p21.32 | -5.13 | 2.91E-07 | 0.42 | *TSBP1-AS1* | Intronic |
| cg04452581 | 6 | 32,397,683 | 6p21.32 | -6.37 | 1.89E-10 | 0.54 | *TSBP1-AS1* ; *HLA-DRA* | Intergenic |
| cg25982308 | 6 | 32,708,469 | 6p21.32 | 5.44 | 5.35E-08 | 0.57 | *HLA-DQA2* | TSS1500 |
| cg17360552 | 6 | 32,725,332 | 6p21.32 | 6.08 | 1.22E-09 | 0.72 | *HLA-DQB2* | Intronic |
| cg12296550 | 6 | 32,728,862 | 6p21.32 | -6.52 | 6.86E-11 | 0.72 | *HLA-DQB2* | Intronic |
| cg07180897 | 6 | 32,729,130 | 6p21.32 | 6.30 | 2.98E-10 | 0.55 | *HLA-DQB2* | Intronic |
| cg19887824 | 6 | 32,797,476 | 6p21.32 | 6.02 | 1.73E-09 | 0.38 | *TAP2* | Intronic |
| cg25744682 | 6 | 32,797,578 | 6p21.32 | 5.98 | 2.27E-09 | 0.66 | *TAP2* | Intronic |
| cg27176262 | 6 | 32,797,830 | 6p21.32 | -5.51 | 3.56E-08 | 0.24 | *TAP2* | Exonic |
| cg23750151 | 6 | 32,808,504 | 6p21.32 | -5.20 | 2.02E-07 | 0.28 | *PSMB8* | 3'UTR |
| cg11453837 | 6 | 32,810,551 | 6p21.32 | -5.39 | 7.16E-08 | 0.35 | *PSMB8* | Exonic |
| cg17214455 | 6 | 32,977,138 | 6p21.32 | -6.36 | 2.01E-10 | 0.42 | *HLA-DOA* | Intronic |
| cg26168907 | 6 | 32,977,223 | 6p21.32 | -5.68 | 1.34E-08 | 0.31 | *HLA-DOA* | Intronic |
| cg17313945 | 6 | 32,977,983 | 6p21.32 | 5.15 | 2.55E-07 | 0.28 | *HLA-DOA* | TSS1500 |
| cg16335136 | 6 | 34,551,373 | 6p21.31 | -5.40 | 6.69E-08 | 0.56 | *SPDEF* ; *ILRUN* | Intergenic |
| cg27456627 | 6 | 34,708,295 | 6p21.31 | -5.47 | 4.42E-08 | 0.47 | *LOC101929243* ; *SNRPC* | Intergenic |
| cg27374423 | 6 | 34,759,890 | 6p21.31 | 5.51 | 3.59E-08 | 0.51 | *UHRF1BP1* | 5'UTR |
| cg07306190 | 6 | 34,760,872 | 6p21.31 | -5.66 | 1.52E-08 | 0.25 | *UHRF1BP1* | Intronic |
| cg04617914 | 6 | 41,512,853 | 6p21.1 | 9.13 | 6.94E-20 | 0.47 | *FOXP4-AS1* | Intronic |
| cg10571987 | 6 | 41,528,395 | 6p21.1 | 8.78 | 1.57E-18 | 0.36 | *FOXP4* | Intronic |
| cg01916088 | 6 | 41,528,531 | 6p21.1 | 8.79 | 1.45E-18 | 0.37 | *FOXP4* | Intronic |
| cg18912520 | 6 | 41,528,623 | 6p21.1 | 5.43 | 5.67E-08 | 0.36 | *FOXP4* | Intronic |
| cg05315195 | 6 | 109,294,784 | 6q21 | -5.94 | 2.92E-09 | 0.32 | *ARMC2* | 3'UTR |
| cg01742428 | 6 | 116,782,500 | 6q22.1 | 6.17 | 6.87E-10 | 0.24 | *CALHM6* | TSS200 |
| cg00953399 | 6 | 116,867,006 | 6q22.1 | 5.56 | 2.66E-08 | 0.23 | *CALHM4* | Intronic |
| cg16540098 | 6 | 117,197,653 | 6q22.1 | 5.93 | 2.94E-09 | 0.22 | *RFX6* | TSS1500 |
| cg17235822 | 6 | 117,263,227 | 6q22.1 | 7.86 | 3.71E-15 | 0.26 | *RFX6* ; *VGLL2* | Intergenic |
| cg15031989 | 6 | 153,379,631 | 6q25.2 | 6.17 | 7.01E-10 | 0.34 | *RGS17* | Intronic |
| cg17707550 | 6 | 153,380,415 | 6q25.2 | 8.01 | 1.19E-15 | 0.24 | *RGS17* | Intronic |
| cg25339164 | 6 | 153,381,289 | 6q25.2 | -8.01 | 1.11E-15 | 0.34 | *RGS17* | Intronic |
| cg13184051 | 6 | 160,135,586 | 6q25.3 | 5.24 | 1.57E-07 | 0.25 | *SOD2* | Intronic |
| cg19811651 | 6 | 160,147,226 | 6q25.3 | 6.17 | 7.00E-10 | 0.39 | *SOD2* | Intronic |
| cg20372956 | 6 | 160,148,392 | 6q25.3 | -7.12 | 1.11E-12 | 0.34 | *SOD2* ; *WTAP* | Intronic |
| cg15298323 [d] | 6 | 160,182,438 | 6q25.3 | -5.52 | 3.48E-08 | 0.40 | *SOD2-OT1* | Exonic |
| cg06131755 [d] | 6 | 160,182,447 | 6q25.3 | -5.82 | 5.89E-09 | 0.50 | *SOD2-OT1* | Exonic |
| cg10268548 | 6 | 160,184,041 | 6q25.3 | -6.41 | 1.47E-10 | 0.26 | *ACAT2* | Exonic |
| cg24280851 | 6 | 160,201,987 | 6q25.3 | -5.93 | 3.11E-09 | 0.22 | *TCP1* | Exonic |
| cg04794901 | 6 | 160,209,903 | 6q25.3 | -6.47 | 9.94E-11 | 0.33 | *TCP1* | Intronic |
| cg01145583 | 6 | 160,211,477 | 6q25.3 | -6.15 | 7.52E-10 | 0.25 | *MRPL18* ; *TCP1* | TSS1500 |
| cg10210715 | 6 | 160,221,411 | 6q25.3 | 6.12 | 9.52E-10 | 0.51 | *PNLDC1* | Exonic |
| cg27247736 | 6 | 160,241,105 | 6q25.3 | 6.21 | 5.20E-10 | 0.46 | *PNLDC1* | Intronic |
| cg19598255 | 6 | 160,250,987 | 6q25.3 | 6.30 | 2.90E-10 | 0.51 | *PNLDC1* ; *MAS1* | Intergenic |
| cg15320711 | 6 | 160,320,026 | 6q25.3 | 5.13 | 2.88E-07 | 0.33 | *MAS1* | Intronic |
| cg24462934 | 6 | 160,326,705 | 6q25.3 | -8.70 | 3.35E-18 | 0.22 | *MAS1* | Intronic |
| cg16678626 | 6 | 160,513,496 | 6q25.3 | -8.80 | 1.43E-18 | 0.21 | *IGF2R* | Intronic |
| cg04530265 | 6 | 160,642,786 | 6q25.3 | 12.54 | 4.74E-36 | 0.34 | *SLC22A2* | Intronic |
| cg21323120 | 6 | 160,661,063 | 6q25.3 | 8.78 | 1.57E-18 | 0.26 | *SLC22A2* | Intronic |
| cg02666489 | 6 | 160,680,115 | 6q25.3 | -7.36 | 1.91E-13 | 0.26 | *SLC22A2* | TSS200 |
| cg24883237 | 6 | 160,699,650 | 6q25.3 | 5.75 | 9.18E-09 | 0.51 | *SLC22A2* ; *SLC22A3* | Intergenic |
| cg07520919 | 6 | 160,721,435 | 6q25.3 | -11.24 | 2.48E-29 | 0.21 | *SLC22A2* ; *SLC22A3* | Intergenic |
| cg19898437 | 6 | 160,758,934 | 6q25.3 | 10.49 | 9.63E-26 | 0.24 | *SLC22A2* ; *SLC22A3* | Intergenic |
| cg25867286 | 6 | 160,759,551 | 6q25.3 | 6.81 | 9.87E-12 | 0.32 | *SLC22A2* ; *SLC22A3* | Intergenic |
| cg25313204 | 6 | 160,768,801 | 6q25.3 | 11.94 | 6.93E-33 | 0.41 | *SLC22A3* | TSS1500 |
| cg07349212 | 6 | 160,770,346 | 6q25.3 | 11.45 | 2.40E-30 | 0.35 | *SLC22A3* | Intronic |
| cg01739295 | 6 | 160,773,026 | 6q25.3 | 10.87 | 1.68E-27 | 0.21 | *SLC22A3* | Intronic |

| | | | | | | | | |
|---|---|---|---|---|---|---|---|---|
| cg15292008 | 6 | 160,928,551 | 6q25.3 | -8.61 | 7.40E-18 | 0.28 | *LPAL2* | Intronic |
| cg15398520 | 6 | 160,932,709 | 6q25.3 | -9.83 | 8.19E-23 | 0.23 | *LPAL2* | TSS1500 |
| cg14031365 | 7 | 20,816,695 | 7p21.1 | 6.31 | 2.78E-10 | 0.20 | *ABCB5* ; *SP8* | Intergenic |
| cg27191852 | 7 | 20,827,982 | 7p21.1 | 5.74 | 9.40E-09 | 0.25 | *SP8; RPL23P8* | Intergenic |
| cg24203469 | 7 | 20,874,871 | 7p21.1 | -5.35 | 8.67E-08 | 0.39 | *LINC01162* | TSS200 |
| cg00188704 | 7 | 27,140,797 | 7p15.2 | -5.63 | 1.78E-08 | 0.42 | *HOXA2* | Exonic |
| cg00921266 | 7 | 27,153,663 | 7p15.2 | -7.12 | 1.09E-12 | 0.35 | *HOXA3* | Intronic |
| cg13694927 | 7 | 27,184,712 | 7p15.2 | 8.03 | 9.54E-16 | 0.21 | *HOXA-AS3* | Intronic |
| cg10474350 | 7 | 27,196,555 | 7p15.2 | 6.90 | 5.17E-12 | 0.24 | *HOXA7* | TSS1500 |
| cg22260008 | 7 | 27,207,318 | 7p15.2 | 6.07 | 1.28E-09 | 0.21 | *HOXA10-HOXA9* | Intronic |
| cg14849571 | 7 | 27,464,261 | 7p15.2 | 6.86 | 6.84E-12 | 0.42 | *EVX1* ; *HIBADH* | Intergenic |
| cg20253542 [d] | 7 | 27,561,082 | 7p15.2 | -9.14 | 6.40E-20 | 0.54 | *EVX1* ; *HIBADH* | Intergenic |
| cg07838458 | 7 | 27,650,858 | 7p15.2 | 5.33 | 9.59E-08 | 0.26 | *HIBADH* | Intronic |
| cg05299362 | 7 | 27,688,612 | 7p15.2 | -5.42 | 5.88E-08 | 0.23 | *HIBADH* | Intronic |
| cg14334382 | 7 | 27,703,802 | 7p15.2 | 5.60 | 2.14E-08 | 0.40 | *HIBADH* ; *TAX1BP1* | Intergenic |
| cg01662334 | 7 | 27,704,065 | 7p15.2 | 6.82 | 9.20E-12 | 0.30 | *HIBADH* ; *TAX1BP1* | Intergenic |
| cg26225794 | 7 | 27,717,723 | 7p15.2 | 5.18 | 2.21E-07 | 0.34 | *HIBADH* ; *TAX1BP1* | Intergenic |
| cg18600035 | 7 | 27,779,299 | 7p15.2 | 6.83 | 8.25E-12 | 0.25 | *TAX1BP1* | Intronic |
| cg06314335 | 7 | 27,862,072 | 7p15.2 | 10.59 | 3.26E-26 | 0.62 | *TAX1BP1* | Intronic |
| cg07314168 | 7 | 27,866,203 | 7p15.2 | 8.28 | 1.28E-16 | 0.63 | *TAX1BP1* | Intronic |
| cg21726132 | 7 | 27,936,643 | 7p15.2 | -5.75 | 9.13E-09 | 0.74 | *JAZF1* | Intronic |
| cg06642895 | 7 | 27,952,113 | 7p15.2 | -8.23 | 1.90E-16 | 0.23 | *JAZF1* | Intronic |
| cg22714138 | 7 | 27,968,663 | 7p15.2 | -5.66 | 1.56E-08 | 0.35 | *JAZF1* | Intronic |
| cg04084447 | 7 | 28,394,254 | 7p15.1 | 7.19 | 6.29E-13 | 0.29 | *CREB5* | Intronic |
| cg04778099 | 7 | 47,445,517 | 7p12.3 | 6.76 | 1.42E-11 | 0.66 | *TNS3* | Intronic |
| cg23694490 | 7 | 47,445,681 | 7p12.3 | 6.79 | 1.12E-11 | 0.69 | *TNS3* | Intronic |
| cg08414403 | 7 | 47,446,450 | 7p12.3 | 6.50 | 8.19E-11 | 0.40 | *TNS3* | Intronic |
| cg05700881 | 7 | 47,474,420 | 7p12.3 | 5.22 | 1.77E-07 | 0.30 | *TNS3* | Intronic |
| cg09696706 | 7 | 47,479,511 | 7p12.3 | -6.38 | 1.80E-10 | 0.42 | *TNS3* | Intronic |
| cg18676790 | 7 | 97,680,731 | 7q21.3 | -8.47 | 2.41E-17 | 0.28 | *OCM2* ; *LMTK2* | Intergenic |
| cg13397024 | 7 | 97,797,637 | 7q21.3 | 8.51 | 1.75E-17 | 0.33 | *LMTK2* | Intronic |
| cg14507403 | 7 | 97,798,005 | 7q21.3 | 6.21 | 5.22E-10 | 0.68 | *LMTK2* | Intronic |
| cg06991560 | 7 | 97,803,978 | 7q21.3 | -10.05 | 9.33E-24 | 0.30 | *LMTK2* | Intronic |
| cg22295745 | 7 | 97,807,643 | 7q21.3 | -5.22 | 1.79E-07 | 0.41 | *LMTK2* | Intronic |
| cg24562669 | 7 | 97,807,699 | 7q21.3 | -12.49 | 8.80E-36 | 0.28 | *LMTK2* | Intronic |
| cg21770322 | 7 | 97,807,741 | 7q21.3 | -12.46 | 1.21E-35 | 0.22 | *LMTK2* | Intronic |
| cg02328330 | 7 | 97,815,869 | 7q21.3 | 7.51 | 5.82E-14 | 0.22 | *LMTK2* | Intronic |
| cg08169830 | 7 | 97,822,374 | 7q21.3 | 5.15 | 2.54E-07 | 0.30 | *LMTK2* | Exonic |
| cg20320158 | 7 | 97,847,054 | 7q21.3 | 5.62 | 1.90E-08 | 0.25 | *TECPR1* | Exonic |
| cg08052726 | 7 | 97,852,433 | 7q21.3 | 8.34 | 7.32E-17 | 0.29 | *TECPR1* | Exonic |
| cg18724396 | 7 | 97,870,994 | 7q21.3 | -9.55 | 1.28E-21 | 0.30 | *TECPR1* | Intronic |
| cg23184226 | 7 | 97,880,563 | 7q21.3 | -8.14 | 4.01E-16 | 0.21 | *TECPR1* | Intronic |
| cg02985445 | 7 | 97,908,505 | 7q21.3 | 8.48 | 2.20E-17 | 0.39 | *TECPR1* ; *BRI3* | Intergenic |
| cg01877450 | 7 | 97,915,802 | 7q21.3 | 8.12 | 4.50E-16 | 0.45 | *BRI3* | Intronic |
| cg00277769 | 7 | 97,922,759 | 7q21.3 | 9.19 | 3.97E-20 | 0.37 | *BAIAP2L1* | 3'UTR |
| cg17341174 | 7 | 97,923,834 | 7q21.3 | -7.09 | 1.32E-12 | 0.30 | *BAIAP2L1* | Intronic |
| cg12366800 | 7 | 97,929,490 | 7q21.3 | -6.64 | 3.19E-11 | 0.35 | *BAIAP2L1* | Intronic |
| cg12209329 | 7 | 97,929,748 | 7q21.3 | 9.18 | 4.41E-20 | 0.49 | *BAIAP2L1* | Intronic |
| cg17403231 | 7 | 97,930,635 | 7q21.3 | -8.13 | 4.29E-16 | 0.56 | *BAIAP2L1* | Intronic |
| cg23043230 | 7 | 97,935,666 | 7q21.3 | 10.66 | 1.50E-26 | 0.42 | *BAIAP2L1* | Intronic |
| cg03626541 | 7 | 97,982,115 | 7q21.3 | -7.25 | 4.12E-13 | 0.61 | *BAIAP2L1* | Intronic |
| cg00213668 | 7 | 98,028,207 | 7q22.1 | -6.68 | 2.46E-11 | 0.22 | *BAIAP2L1* | Intronic |
| cg08684580 | 7 | 98,029,266 | 7q22.1 | 7.65 | 1.94E-14 | 0.32 | *BAIAP2L1* | Intronic |
| cg15827681 | 7 | 98,029,274 | 7q22.1 | 9.87 | 5.68E-23 | 0.24 | *BAIAP2L1* | Intronic |
| cg22708902 | 8 | 23,100,981 | 8p21.3 | 7.12 | 1.08E-12 | 0.36 | *CHMP7* | TSS200 |
| cg08026493 | 8 | 23,430,824 | 8p21.2 | -5.90 | 3.55E-09 | 0.44 | *SLC25A37* | 3'UTR |
| cg20845639 | 8 | 23,430,886 | 8p21.2 | -6.68 | 2.46E-11 | 0.36 | *SLC25A37* | 3'UTR |

| | | | | | | | | |
|---|---|---|---|---|---|---|---|---|
| cg08349112 | 8 | 23,430,909 | 8p21.2 | -6.50 | 8.05E-11 | 0.26 | *SLC25A37* | 3'UTR |
| cg27392395 | 8 | 23,431,014 | 8p21.2 | -7.12 | 1.06E-12 | 0.33 | *SLC25A37* | 3'UTR |
| cg17635193 | 8 | 23,447,031 | 8p21.2 | 6.94 | 4.04E-12 | 0.21 | *SLC25A37* ; *NKX3-1* | Intergenic |
| cg02821261 | 8 | 23,458,808 | 8p21.2 | -9.18 | 4.35E-20 | 0.29 | *SLC25A37* ; *NKX3-1* | Intergenic |
| cg21481775 | 8 | 23,539,627 | 8p21.2 | -6.05 | 1.44E-09 | 0.25 | *NKX3-1* | Intronic |
| cg18502021 | 8 | 23,540,946 | 8p21.2 | -9.45 | 3.42E-21 | 0.28 | *NKX3-1* | TSS1500 |
| cg25820799 | 8 | 127,548,957 | 8q24.21 | -7.61 | 2.73E-14 | 0.24 | *LOC101927657; LRATD2* | Intergenic |
| cg06500932 | 8 | 127,882,188 | 8q24.21 | -7.35 | 2.03E-13 | 0.45 | *LRATD2* ; *PCAT1* | Intergenic |
| cg01725759 | 8 | 127,916,305 | 8q24.21 | -7.35 | 1.96E-13 | 0.34 | *LRATD2* ; *PCAT1* | Intergenic |
| cg01495299 | 8 | 128,003,923 | 8q24.21 | 9.72 | 2.43E-22 | 0.23 | *LRATD2* ; *PCAT1* | Intergenic |
| cg16113271 | 8 | 128,025,393 | 8q24.21 | 12.81 | 1.38E-37 | 0.52 | *PCAT1* | TSS200 |
| cg05928532 | 8 | 128,243,796 | 8q24.21 | 6.42 | 1.33E-10 | 0.35 | *CCAT1* ; *CASC21* | Intergenic |
| cg20001106 | 8 | 128,344,353 | 8q24.21 | -7.47 | 7.79E-14 | 0.46 | *CASC21* ; *CASC8* | Intronic |
| cg14208978 | 8 | 128,351,421 | 8q24.21 | -5.85 | 4.94E-09 | 0.33 | *CASC21* ; *CASC8* | Intronic |
| cg02871568 | 8 | 128,382,999 | 8q24.21 | -15.40 | 1.76E-53 | 0.26 | *CASC21* ; *CASC8* | Intronic |
| cg04657445 | 8 | 128,390,627 | 8q24.21 | -17.29 | 5.97E-67 | 0.33 | *CASC21* ; *CASC8* | Intronic |
| cg08250469 | 8 | 128,434,474 | 8q24.21 | 8.14 | 3.96E-16 | 0.30 | *CASC8* | Intronic |
| cg17465588 | 8 | 128,453,500 | 8q24.21 | -8.72 | 2.90E-18 | 0.30 | *CASC8* | Intronic |
| cg11218883 | 8 | 128,576,880 | 8q24.21 | 5.33 | 1.01E-07 | 0.27 | *CASC8* ; *CASC11* | Intergenic |
| cg25610826 | 8 | 128,586,421 | 8q24.21 | 5.73 | 9.76E-09 | 0.34 | *CASC8* ; *CASC11* | Intergenic |
| cg03272903 | 8 | 128,683,224 | 8q24.21 | 12.95 | 2.44E-38 | 0.23 | *CASC8* ; *CASC11* | Intergenic |
| cg08725914 | 8 | 128,849,627 | 8q24.21 | -6.70 | 2.12E-11 | 0.35 | *PVT1* | Intronic |
| cg05744690 | 9 | 19,033,256 | 9p22.1 | -6.21 | 5.22E-10 | 0.35 | *SAXO1* | Intronic |
| cg17351239 | 9 | 19,033,306 | 9p22.1 | -6.20 | 5.63E-10 | 0.45 | *SAXO1* | Intronic |
| cg03156866 | 9 | 19,033,365 | 9p22.1 | 6.50 | 8.15E-11 | 0.42 | *SAXO1* | Intronic |
| cg10236024 | 9 | 19,033,634 | 9p22.1 | -6.48 | 9.32E-11 | 0.33 | *SAXO1* | Intronic |
| cg17509612 | 9 | 132,598,774 | 9q34.11 | 5.12 | 3.03E-07 | 0.33 | *USP20* | Intronic |
| cg02700194 | 10 | 834,378 | 10p15.3 | -6.33 | 2.47E-10 | 0.37 | *DIP2C* ; *LARP4B* | Intergenic |
| cg10017260 | 10 | 834,428 | 10p15.3 | -6.70 | 2.02E-11 | 0.43 | *DIP2C* ; *LARP4B* | Intergenic |
| cg09361094 | 10 | 834,503 | 10p15.3 | -5.89 | 3.83E-09 | 0.49 | *DIP2C* ; *LARP4B* | Intergenic |
| cg20503657 | 10 | 835,505 | 10p15.3 | 6.16 | 7.38E-10 | 0.70 | *DIP2C* ; *LARP4B* | Intergenic |
| cg02228420 | 10 | 840,201 | 10p15.3 | -6.21 | 5.42E-10 | 0.48 | *DIP2C* ; *LARP4B* | Intergenic |
| cg13163782 | 10 | 856,428 | 10p15.3 | -6.59 | 4.30E-11 | 0.30 | *LARP4B* | 3'UTR |
| cg20035724 | 10 | 856,514 | 10p15.3 | -6.55 | 5.74E-11 | 0.24 | *LARP4B* | 3'UTR |
| cg06043865 | 10 | 47,045,162 | 10q11.22 | -8.31 | 9.65E-17 | 0.22 | *GPRIN2* ; *NPY4R2* | Intergenic |
| cg09839204 | 10 | 47,049,879 | 10q11.22 | -7.17 | 7.32E-13 | 0.21 | *GPRIN2* ; *NPY4R2* | Intergenic |
| cg10897344 | 10 | 104,227,113 | 10q24.32 | 7.42 | 1.15E-13 | 0.28 | *MFSD13A* | Intronic |
| cg15685307 | 10 | 104,228,081 | 10q24.32 | 6.76 | 1.34E-11 | 0.51 | *MFSD13A* | Intronic |
| cg14976746 | 10 | 104,228,125 | 10q24.32 | 5.79 | 7.06E-09 | 0.32 | *MFSD13A* | Intronic |
| cg19045729 | 10 | 104,252,630 | 10q24.32 | 6.25 | 4.14E-10 | 0.41 | *ACTR1A* | Intronic |
| cg09800781 | 10 | 104,253,015 | 10q24.32 | -5.94 | 2.78E-09 | 0.24 | *ACTR1A* | Intronic |
| cg21184170 | 10 | 104,299,959 | 10q24.32 | -5.59 | 2.28E-08 | 0.43 | *SUFU* | Intronic |
| cg01357317 | 10 | 104,373,849 | 10q24.32 | 6.91 | 4.83E-12 | 0.24 | *SUFU* | Intronic |
| cg08991610 | 10 | 104,398,766 | 10q24.32 | -7.86 | 3.94E-15 | 0.21 | *SUFU* ; *TRIM8* | Intergenic |
| cg07119830 | 10 | 104,412,306 | 10q24.32 | -6.97 | 3.15E-12 | 0.31 | *TRIM8* | Intronic |
| cg05805172 | 10 | 104,417,083 | 10q24.32 | -6.39 | 1.64E-10 | 0.25 | *TRIM8* | Exonic |
| cg24435571 | 10 | 104,436,140 | 10q24.32 | -7.31 | 2.65E-13 | 0.24 | *ARL3* | 3'UTR |
| cg07544899 | 10 | 104,479,373 | 10q24.32 | -7.82 | 5.14E-15 | 0.26 | *SFXN2* | Intronic |
| cg06169961 | 10 | 104,502,638 | 10q24.32 | -6.11 | 9.67E-10 | 0.31 | *SFXN2* | 3'UTR |
| cg10951224 | 10 | 104,502,824 | 10q24.32 | -6.41 | 1.50E-10 | 0.42 | *SFXN2* | 3'UTR |
| cg13349042 | 10 | 104,503,101 | 10q24.32 | -6.04 | 1.50E-09 | 0.59 | *SFXN2* | 3'UTR |
| cg13253055 | 10 | 104,574,946 | 10q24.32 | 6.11 | 1.01E-09 | 0.26 | *WBP1L* | 3'UTR |
| cg07809032 | 10 | 104,592,955 | 10q24.32 | 5.80 | 6.62E-09 | 0.38 | *CYP17A1* | Exonic |
| cg06098344 | 10 | 123,048,167 | 10q26.12 | 5.73 | 1.03E-08 | 0.28 | *WDR11* ; *FGFR2* | Intergenic |
| cg21341012 | 10 | 126,378,832 | 10q26.13 | -6.69 | 2.23E-11 | 0.24 | *FAM53B* | Intronic |
| cg09559798 | 10 | 126,688,792 | 10q26.13 | -5.38 | 7.39E-08 | 0.44 | *CTBP2* | Intronic |
| cg19148024 | 10 | 126,695,914 | 10q26.13 | -5.18 | 2.23E-07 | 0.57 | *CTBP2* | Intronic |
| cg02921122 | 10 | 126,712,074 | 10q26.13 | -6.04 | 1.50E-09 | 0.26 | *CTBP2* | Intronic |

| | | | | | | | | |
|---|---|---|---|---|---|---|---|---|
| cg07048415 | 10 | 126,714,013 | 10q26.13 | 6.24 | 4.48E-10 | 0.23 | *CTBP2* | Intronic |
| cg12446642 | 10 | 126,722,163 | 10q26.13 | -5.30 | 1.14E-07 | 0.39 | *CTBP2* | Intronic |
| cg14250138 [d] | 11 | 1,785,804 | 11p15.5 | 6.21 | 5.27E-10 | 0.26 | *CTSD* | TSS1500 |
| cg04047579 [d] | 11 | 1,834,295 | 11p15.5 | -5.98 | 2.29E-09 | 0.22 | *CTSD* ; *SYT8* | Intergenic |
| cg04726730 [d] | 11 | 1,868,700 | 11p15.5 | 7.54 | 4.53E-14 | 0.26 | *TNNI2* ; *LSP1* | Intergenic |
| cg02246725 | 11 | 2,014,127 | 11p15.5 | -7.57 | 3.83E-14 | 0.38 | *LINC01219* | Intronic |
| cg10337079 | 11 | 2,158,697 | 11p15.5 | 5.74 | 9.68E-09 | 0.47 | *IGF2* | Intronic |
| cg09606495 | 11 | 2,181,269 | 11p15.5 | 6.33 | 2.52E-10 | 0.39 | *INS* ; *INS-IGF2* | Intronic |
| cg02749887 | 11 | 2,182,540 | 11p15.5 | -8.07 | 7.10E-16 | 0.35 | *INS* ; *INS-IGF2* | TSS200 |
| cg21574853 | 11 | 2,182,573 | 11p15.5 | -6.77 | 1.25E-11 | 0.28 | *INS* ; *INS-IGF2* | TSS200 |
| cg03366382 | 11 | 2,182,644 | 11p15.5 | -5.62 | 1.89E-08 | 0.47 | *INS* ; *INS-IGF2* | TSS1500 |
| cg25601886 | 11 | 2,183,420 | 11p15.5 | -5.35 | 8.92E-08 | 0.28 | *INS* ; *INS-IGF2* | TSS1500 |
| cg09864961 | 11 | 2,183,434 | 11p15.5 | -5.40 | 6.81E-08 | 0.36 | *INS* ; *INS-IGF2* | TSS1500 |
| cg05141787 | 11 | 2,193,284 | 11p15.5 | -6.73 | 1.72E-11 | 0.26 | *TH* | TSS1500 |
| cg26253759 | 11 | 2,193,501 | 11p15.5 | -7.76 | 8.22E-15 | 0.22 | *MIR4686* ; *TH* | TSS1500 |
| cg00782718 | 11 | 2,197,180 | 11p15.5 | 6.43 | 1.28E-10 | 0.67 | *MIR4686* ; *ASCL2* | Intergenic |
| cg18847491 | 11 | 2,198,579 | 11p15.5 | 5.64 | 1.66E-08 | 0.52 | *MIR4686* ; *ASCL2* | Intergenic |
| cg05224479 | 11 | 2,208,400 | 11p15.5 | 11.57 | 5.61E-31 | 0.43 | *MIR4686* ; *ASCL2* | Intergenic |
| cg10838469 | 11 | 2,211,741 | 11p15.5 | -12.01 | 3.15E-33 | 0.24 | *MIR4686* ; *ASCL2* | Intergenic |
| cg03628333 | 11 | 2,214,677 | 11p15.5 | -13.09 | 3.74E-39 | 0.60 | *MIR4686* ; *ASCL2* | Intergenic |
| cg15177669 | 11 | 2,233,123 | 11p15.5 | -12.88 | 5.48E-38 | 0.35 | *MIR4686* ; *ASCL2* | Intergenic |
| cg10658853 | 11 | 2,233,512 | 11p15.5 | -10.57 | 3.97E-26 | 0.41 | *MIR4686* ; *ASCL2* | Intergenic |
| cg25575145 | 11 | 2,233,644 | 11p15.5 | 13.90 | 6.10E-44 | 0.33 | *MIR4686* ; *ASCL2* | Intergenic |
| cg25635251 | 11 | 2,234,043 | 11p15.5 | -15.05 | 3.37E-51 | 0.70 | *MIR4686* ; *ASCL2* | Intergenic |
| cg12428727 | 11 | 2,234,665 | 11p15.5 | -12.04 | 2.30E-33 | 0.41 | *MIR4686* ; *ASCL2* | Intergenic |
| cg25080747 | 11 | 2,289,117 | 11p15.5 | -5.40 | 6.67E-08 | 0.28 | *ASCL2* | Downstream |
| cg09261816 | 11 | 61,530,655 | 11q12.2 | 5.14 | 2.77E-07 | 0.35 | *MYRF* | Intronic |
| cg02742593 | 11 | 68,481,342 | 11q13.3 | -6.04 | 1.56E-09 | 0.23 | *TESMIN* | Intronic |
| cg06292310 | 11 | 68,581,324 | 11q13.3 | -5.22 | 1.78E-07 | 0.23 | *CPT1A* | Intronic |
| cg10257063 | 11 | 68,587,159 | 11q13.3 | -8.28 | 1.23E-16 | 0.27 | *CPT1A* | Intronic |
| cg02786073 | 11 | 68,625,231 | 11q13.3 | 13.58 | 5.60E-42 | 0.26 | *CPT1A* ; *MRPL21* | Intergenic |
| cg10606903 | 11 | 68,664,848 | 11q13.3 | -10.41 | 2.35E-25 | 0.27 | *MRPL21* | Intronic |
| cg20156080 | 11 | 68,703,600 | 11q13.3 | -6.45 | 1.12E-10 | 0.23 | *IGHMBP2* | Intronic |
| cg16219649 | 11 | 68,714,011 | 11q13.3 | 6.84 | 7.78E-12 | 0.32 | *IGHMBP2* ; *MRGPRD* | Intergenic |
| cg27214458 | 11 | 68,781,032 | 11q13.3 | -8.33 | 8.12E-17 | 0.25 | *MRGPRF-AS1* | Intronic |
| cg22986999 | 11 | 68,781,976 | 11q13.3 | 7.64 | 2.16E-14 | 0.24 | *MRGPRF-AS1* | Exonic |
| cg02455391 | 11 | 68,786,679 | 11q13.3 | -9.83 | 8.04E-23 | 0.38 | *MRGPRF-AS1* | Downstream |
| cg03494790 | 11 | 68,811,758 | 11q13.3 | 8.81 | 1.27E-18 | 0.31 | *MRGPRF-AS1* ; *TPCN2* | Intergenic |
| cg05951984 | 11 | 68,823,811 | 11q13.3 | -6.82 | 9.27E-12 | 0.31 | *TPCN2* | Intronic |
| cg11648404 | 11 | 68,830,440 | 11q13.3 | -12.13 | 7.67E-34 | 0.31 | *TPCN2* | Exonic |
| cg01385198 | 11 | 68,856,225 | 11q13.3 | 5.40 | 6.66E-08 | 0.28 | *TPCN2* | 3'UTR |
| cg16709353 | 11 | 68,856,817 | 11q13.3 | -6.18 | 6.56E-10 | 0.27 | *TPCN2* | 3'UTR |
| cg12152352 | 11 | 68,856,906 | 11q13.3 | -11.34 | 8.18E-30 | 0.29 | *TPCN2* | 3'UTR |
| cg13834366 | 11 | 68,857,566 | 11q13.3 | 6.19 | 5.90E-10 | 0.26 | *TPCN2* | 3'UTR |
| cg03098712 | 11 | 68,860,771 | 11q13.3 | -8.38 | 5.50E-17 | 0.30 | *TPCN2* ; *LOC338694* | Intergenic |
| cg14286243 | 11 | 68,867,220 | 11q13.3 | 6.00 | 2.03E-09 | 0.56 | *TPCN2* ; *LOC338694* | Intergenic |
| cg16499607 | 11 | 68,883,356 | 11q13.3 | 15.78 | 4.11E-56 | 0.53 | *TPCN2* ; *LOC338694* | Intergenic |
| cg26456404 | 11 | 68,883,434 | 11q13.3 | 15.04 | 4.22E-51 | 0.55 | *TPCN2* ; *LOC338694* | Intergenic |
| cg15175377 | 11 | 68,920,434 | 11q13.3 | 11.53 | 8.80E-31 | 0.27 | *LOC338694* | Intronic |
| cg19820355 | 11 | 68,921,264 | 11q13.3 | 10.22 | 1.57E-24 | 0.30 | *LOC338694* | Intronic |
| cg17075483 | 11 | 68,923,332 | 11q13.3 | 10.08 | 6.79E-24 | 0.28 | *LOC338694* | Intronic |
| cg25179853 | 11 | 68,924,577 | 11q13.3 | 13.28 | 3.01E-40 | 0.38 | *LOC338694* | Intronic |
| cg23740940 | 11 | 68,924,746 | 11q13.3 | 11.85 | 2.03E-32 | 0.43 | *LOC338694* | Intronic |
| cg07882059 | 11 | 68,924,751 | 11q13.3 | 13.73 | 6.65E-43 | 0.43 | *LOC338694* | Intronic |
| cg03469862 | 11 | 68,924,853 | 11q13.3 | 12.35 | 4.73E-35 | 0.38 | *LOC338694* | Intronic |
| cg06185577 | 11 | 68,960,238 | 11q13.3 | 15.23 | 2.15E-52 | 0.47 | *LOC338694* ; *MYEOV* | Intergenic |
| cg25475764 | 11 | 68,963,457 | 11q13.3 | -13.35 | 1.17E-40 | 0.46 | *LOC338694* ; *MYEOV* | Intergenic |

| | | | | | | | | |
|---|---|---|---|---|---|---|---|---|
| cg06798934 | 11 | 68,963,737 | 11q13.3 | -12.67 | 8.16E-37 | 0.36 | *LOC338694* ; *MYEOV* | Intergenic |
| cg26172195 | 11 | 68,972,651 | 11q13.3 | -7.51 | 5.97E-14 | 0.33 | *LOC338694* ; *MYEOV* | Intergenic |
| cg15059493 [d] | 11 | 68,980,857 | 11q13.3 | 16.93 | 2.76E-64 | 0.64 | *LOC338694* ; *MYEOV* | Intergenic |
| cg27158675 | 11 | 68,983,797 | 11q13.3 | -14.43 | 3.54E-47 | 0.23 | *LOC338694* ; *MYEOV* | Intergenic |
| cg16166568 [d] | 11 | 68,986,598 | 11q13.3 | -19.88 | 6.39E-88 | 0.77 | *LOC338694* ; *MYEOV* | Intergenic |
| cg18226337 | 11 | 69,002,666 | 11q13.3 | 14.69 | 6.94E-49 | 0.26 | *LOC338694* ; *MYEOV* | Intergenic |
| cg14485103 [d] | 11 | 69,017,579 | 11q13.3 | -5.62 | 1.93E-08 | 0.33 | *LOC338694* ; *MYEOV* | Intergenic |
| cg13888763 | 11 | 69,023,300 | 11q13.3 | 15.06 | 2.80E-51 | 0.57 | *LOC338694* ; *MYEOV* | Intergenic |
| cg07444963 | 11 | 69,025,650 | 11q13.3 | -14.11 | 3.29E-45 | 0.27 | *LOC338694* ; *MYEOV* | Intergenic |
| cg22779330 | 11 | 69,061,911 | 11q13.3 | -11.97 | 5.41E-33 | 0.41 | *MYEOV* | 5'UTR |
| cg17418276 | 11 | 69,065,442 | 11q13.3 | -8.24 | 1.69E-16 | 0.36 | *MYEOV* | Downstream |
| cg01597130 | 11 | 69,068,016 | 11q13.3 | -13.06 | 5.35E-39 | 0.25 | *MYEOV* ; *LOC102724265* | Intergenic |
| cg02721252 | 11 | 69,151,016 | 11q13.3 | -5.91 | 3.38E-09 | 0.21 | *MYEOV* ; *LOC102724265* | Intergenic |
| cg24568082 | 11 | 69,151,092 | 11q13.3 | -8.28 | 1.25E-16 | 0.33 | *MYEOV* ; *LOC102724265* | Intergenic |
| cg10691700 | 11 | 69,228,187 | 11q13.3 | -5.32 | 1.03E-07 | 0.37 | *MYEOV* ; *LOC102724265* | Intergenic |
| cg06921552 | 11 | 69,258,203 | 11q13.3 | 9.79 | 1.20E-22 | 0.41 | *LOC102724265* ; *LINC01488* | Intergenic |
| cg22381128 | 11 | 69,275,403 | 11q13.3 | -5.13 | 2.86E-07 | 0.31 | *LOC102724265* ; *LINC01488* | Intergenic |
| cg22653345 | 11 | 69,307,400 | 11q13.3 | -8.48 | 2.24E-17 | 0.27 | *LINC01488* | Exonic |
| cg16131807 | 11 | 69,387,941 | 11q13.3 | 5.78 | 7.31E-09 | 0.20 | *LINC01488* ; *CCND1* | Intergenic |
| cg14405341 | 11 | 69,440,295 | 11q13.3 | -6.53 | 6.58E-11 | 0.41 | *LINC01488* ; *CCND1* | Intergenic |
| cg10533055 | 11 | 69,482,953 | 11q13.3 | 6.42 | 1.33E-10 | 0.26 | *LTO1* | Intronic |
| cg25511807 | 11 | 102,401,438 | 11q22.2 | -7.86 | 3.90E-15 | 0.36 | *MMP7* | 5'UTR |
| cg17707274 | 11 | 102,401,523 | 11q22.2 | -6.74 | 1.55E-11 | 0.31 | *MMP7* | TSS200 |
| cg21960184 | 11 | 113,804,386 | 11q23.2 | -7.86 | 3.85E-15 | 0.21 | *HTR3B* | Intronic |
| cg06542609 | 11 | 134,280,166 | 11q25 | -5.17 | 2.36E-07 | 0.40 | *B3GAT1* | Intronic |
| cg19252671 | 12 | 12,871,110 | 12p13.1 | -5.99 | 2.13E-09 | 0.37 | *CDKN1B* | Exonic |
| cg16604812 | 12 | 48,419,819 | 12q13.11 | 6.05 | 1.41E-09 | 0.20 | *COL2A1* ; *SENP1* | Intergenic |
| cg13709639 | 12 | 49,526,040 | 12q13.12 | 5.23 | 1.67E-07 | 0.41 | *TUBA1B* | TSS1500 |
| cg04797936 | 12 | 49,659,154 | 12q13.12 | -6.29 | 3.12E-10 | 0.26 | *TUBA1C* | Intronic |
| cg11878254 | 12 | 49,731,885 | 12q13.12 | 5.43 | 5.57E-08 | 0.34 | *C1QL4* | TSS1500 |
| cg02259081 | 12 | 53,268,433 | 12q13.13 | 5.56 | 2.67E-08 | 0.51 | *KRT78* ; *KRT8* | Intergenic |
| cg06553033 | 12 | 53,270,239 | 12q13.13 | 5.80 | 6.78E-09 | 0.48 | *KRT78* ; *KRT8* | Intergenic |
| cg14581129 | 12 | 53,358,946 | 12q13.13 | -6.27 | 3.54E-10 | 0.69 | *KRT78* ; *KRT8* | Intergenic |
| cg05393297 | 12 | 53,359,155 | 12q13.13 | -6.00 | 1.98E-09 | 0.58 | *KRT78* ; *KRT8* | Intergenic |
| cg25025879 | 12 | 53,359,317 | 12q13.13 | -5.76 | 8.20E-09 | 0.48 | *KRT78* ; *KRT8* | Intergenic |
| cg16329197 | 12 | 53,359,506 | 12q13.13 | -6.11 | 1.02E-09 | 0.60 | *KRT78* ; *KRT8* | Intergenic |
| cg04396185 | 12 | 53,574,334 | 12q13.13 | -8.12 | 4.59E-16 | 0.20 | *CSAD* | Intronic |
| cg20163807 | 12 | 53,773,828 | 12q13.13 | -8.86 | 8.16E-19 | 0.34 | *SP1* | TSS200 |
| cg16098340 | 12 | 65,014,995 | 12q14.2 | -5.78 | 7.36E-09 | 0.22 | *RASSF3* | Intronic |
| cg16907855 | 12 | 89,932,866 | 12q21.33 | 5.14 | 2.70E-07 | 0.28 | *POC1B-AS1* | Intronic |
| cg04602591 | 12 | 90,147,542 | 12q21.33 | -6.24 | 4.43E-10 | 0.21 | *ATP2B1-AS1* ; *LINC02399* | Intergenic |
| cg10233830 | 12 | 114,701,413 | 12q24.21 | 6.23 | 4.74E-10 | 0.25 | *LINC02459* ; *TBX5* | Intergenic |
| cg25399352 | 12 | 114,837,572 | 12q24.21 | -8.32 | 8.60E-17 | 0.28 | *TBX5* | Intronic |
| cg25822783 | 12 | 133,050,826 | 12q24.33 | -5.64 | 1.67E-08 | 0.57 | *LOC101928416* ; *FBRSL1* | Intergenic |
| cg07099606 | 12 | 133,051,136 | 12q24.33 | -5.67 | 1.40E-08 | 0.66 | *LOC101928416* ; *FBRSL1* | Intergenic |
| cg11548404 | 12 | 133,076,622 | 12q24.33 | 5.76 | 8.47E-09 | 0.43 | *FBRSL1* | Intronic |
| cg07775147 | 12 | 133,101,495 | 12q24.33 | 5.74 | 9.23E-09 | 0.24 | *FBRSL1* | Intronic |
| cg14078579 | 12 | 133,120,548 | 12q24.33 | -5.93 | 3.00E-09 | 0.62 | *FBRSL1* | Intronic |
| cg05053403 | 12 | 133,120,676 | 12q24.33 | -5.98 | 2.18E-09 | 0.44 | *FBRSL1* | Intronic |
| cg16756508 | 12 | 133,132,798 | 12q24.33 | -5.56 | 2.68E-08 | 0.33 | *FBRSL1* | Intronic |
| cg20454518 | 12 | 133,135,463 | 12q24.33 | -5.14 | 2.81E-07 | 0.37 | *FBRSL1* | Intronic |
| cg01231485 | 12 | 133,140,633 | 12q24.33 | -5.31 | 1.12E-07 | 0.42 | *FBRSL1* | Intronic |
| cg22325186 | 12 | 133,158,791 | 12q24.33 | 5.12 | 3.05E-07 | 0.40 | *FBRSL1* | Exonic |
| cg18366651 | 14 | 23,305,239 | 14q11.2 | 5.65 | 1.56E-08 | 0.76 | *MMP14* ; *MRPL52* | TSS1500 |
| cg00828721 | 14 | 37,121,260 | 14q13.3 | -5.71 | 1.11E-08 | 0.26 | *NKX2-8* ; *PAX9* | Intergenic |

| | | | | | | | | |
|---|---|---|---|---|---|---|---|---|
| cg13075942 | 14 | 69,035,884 | 14q24.1 | -5.75 | 9.18E-09 | 0.26 | *RAD51B* | Intronic |
| cg25522223 | 14 | 70,937,737 | 14q24.2 | -5.36 | 8.33E-08 | 0.37 | *ADAM20P1* | Exonic |
| cg01516775 | 15 | 41,245,254 | 15q15.1 | 5.13 | 2.96E-07 | 0.23 | *CHAC1* | TSS1500 |
| cg12542836 | 15 | 66,600,178 | 15q22.31 | 6.09 | 1.15E-09 | 0.38 | *DIS3L* | Intronic |
| cg00597982 | 15 | 66,773,999 | 15q22.31 | -6.10 | 1.08E-09 | 0.29 | *MAP2K1* | Intronic |
| cg25045219 | 17 | 263,096 | 17p13.3 | -5.78 | 7.52E-09 | 0.22 | *C17orf97* | Exonic |
| cg13576175 | 17 | 504,991 | 17p13.3 | 6.28 | 3.47E-10 | 0.32 | *VPS53* | Intronic |
| cg12281099 | 17 | 599,829 | 17p13.3 | -6.22 | 4.86E-10 | 0.25 | *VPS53* | Intronic |
| cg21512564 | 17 | 613,778 | 17p13.3 | 5.29 | 1.23E-07 | 0.24 | *VPS53* | Exonic |
| cg12384639 | 17 | 618,140 | 17p13.3 | -6.77 | 1.31E-11 | 0.70 | *VPS53* | TSS200 |
| cg08881796 | 17 | 618,545 | 17p13.3 | -7.24 | 4.65E-13 | 0.35 | *VPS53* | TSS1500 |
| cg06500079 | 17 | 619,033 | 17p13.3 | 5.15 | 2.64E-07 | 0.43 | *VPS53* | TSS1500 |
| cg00448220 | 17 | 624,386 | 17p13.3 | 8.99 | 2.49E-19 | 0.28 | *VPS53* ; *TLCD3A* | Intergenic |
| cg18595005 | 17 | 783,036 | 17p13.3 | 7.55 | 4.45E-14 | 0.30 | *NXN* | Intronic |
| cg23087944 | 17 | 7,757,148 | 17p13.1 | -5.49 | 4.06E-08 | 0.54 | *KDM6B* | 3'UTR |
| cg24664346 | 17 | 7,825,949 | 17p13.1 | 5.43 | 5.53E-08 | 0.35 | *KCNAB3* | 3'UTR |
| cg16672659 | 17 | 36,098,223 | 17q12 | -25.67 | 2.82E-145 | 0.66 | *HNF1B* | Intronic |
| cg02335804 | 17 | 36,105,239 | 17q12 | -15.80 | 3.36E-56 | 0.38 | *HNF1B* | TSS200 |
| cg12788467 | 17 | 36,105,335 | 17q12 | -16.24 | 2.72E-59 | 0.32 | *HNF1B* | TSS1500 |
| cg19378036 | 17 | 36,105,364 | 17q12 | -20.03 | 3.17E-89 | 0.35 | *HNF1B* | TSS1500 |
| cg14487292 | 17 | 36,105,517 | 17q12 | -20.18 | 1.54E-90 | 0.31 | *HNF1B* | TSS1500 |
| cg04198914 | 17 | 36,106,025 | 17q12 | -23.76 | 8.56E-125 | 0.43 | *HNF1B* | TSS1500 |
| cg04105270 | 17 | 36,106,301 | 17q12 | -23.57 | 7.30E-123 | 0.62 | *HNF1B* ; *YWHAEP7* | Intergenic |
| cg01529253 | 17 | 36,125,168 | 17q12 | -16.51 | 3.06E-61 | 0.34 | *HNF1B* ; *YWHAEP7* | Intergenic |
| cg17252620 | 17 | 36,125,170 | 17q12 | -13.23 | 5.61E-40 | 0.34 | *HNF1B* ; *YWHAEP7* | Intergenic |
| cg26998125 | 17 | 36,131,474 | 17q12 | -6.65 | 3.02E-11 | 0.78 | *HNF1B* ; *YWHAEP7* | Intergenic |
| cg18418570 | 17 | 36,176,065 | 17q12 | -5.90 | 3.57E-09 | 0.37 | *HNF1B* ; *YWHAEP7* | Intergenic |
| cg21453579 | 17 | 36,404,819 | 17q12 | 5.82 | 6.04E-09 | 0.35 | *LOC440434* | Intronic |
| cg00651005 | 17 | 36,499,258 | 17q12 | 6.73 | 1.66E-11 | 0.33 | *GPR179* | Exonic |
| cg03709850 | 17 | 46,970,000 | 17q21.32 | 5.46 | 4.80E-08 | 0.22 | *LOC105371814* | Exonic |
| cg08112188 | 17 | 47,440,006 | 17q21.33 | 6.95 | 3.75E-12 | 0.55 | *LOC102724596* | Intronic |
| cg05386508 | 17 | 47,841,644 | 17q21.33 | -5.58 | 2.43E-08 | 0.28 | *FAM117A* | TSS200 |
| cg01701526 | 17 | 69,159,818 | 17q24.3 | -13.95 | 3.03E-44 | 0.33 | *CASC17* | Intronic |
| cg26143874 | 18 | 76,363,295 | 18q23 | -5.44 | 5.24E-08 | 0.27 | *LINC01029* ; *SALL3* | Intergenic |
| cg14191434 | 18 | 76,725,033 | 18q23 | -5.19 | 2.14E-07 | 0.70 | *LINC01029* ; *SALL3* | Intergenic |
| cg07420359 | 18 | 76,768,732 | 18q23 | -8.13 | 4.35E-16 | 0.50 | *SALL3* ; *ATP9B* | Intergenic |
| cg11385249 | 19 | 17,215,602 | 19p13.11 | -5.26 | 1.47E-07 | 0.23 | *MYO9B* | Intronic |
| cg19418318 | 19 | 17,219,073 | 19p13.11 | -6.17 | 6.83E-10 | 0.31 | *MYO9B* | Intronic |
| cg08925954 | 19 | 17,219,214 | 19p13.11 | -5.96 | 2.53E-09 | 0.32 | *MYO9B* | Intronic |
| cg23790272 | 19 | 37,960,331 | 19q13.12 | -5.41 | 6.24E-08 | 0.21 | *ZNF570* | 5'UTR |
| cg03611452 | 19 | 38,183,253 | 19q13.12 | 5.17 | 2.31E-07 | 0.46 | *ZNF781* | TSS200 |
| cg21650740 | 19 | 38,457,168 | 19q13.13 | 5.47 | 4.38E-08 | 0.34 | *SIPA1L3* | Intronic |
| cg16217840 | 19 | 38,574,952 | 19q13.13 | -8.19 | 2.52E-16 | 0.23 | *SIPA1L3* | Intronic |
| cg01723844 | 19 | 38,715,357 | 19q13.2 | 6.29 | 3.14E-10 | 0.29 | *DPF1* | Intronic |
| cg27110831 | 19 | 38,715,363 | 19q13.2 | 6.07 | 1.24E-09 | 0.35 | *DPF1* | Intronic |
| cg19395017 | 19 | 38,715,384 | 19q13.2 | 6.40 | 1.52E-10 | 0.48 | *DPF1* | Intronic |
| cg05234415 | 19 | 38,743,744 | 19q13.2 | -11.40 | 3.98E-30 | 0.38 | *PPP1R14A* | Intronic |
| cg05778847 | 19 | 38,746,538 | 19q13.2 | 11.86 | 1.85E-32 | 0.42 | *PPP1R14A* | Intronic |
| cg08196968 | 19 | 38,746,749 | 19q13.2 | 11.30 | 1.26E-29 | 0.21 | *PPP1R14A* | Intronic |
| cg16881183 | 19 | 38,747,088 | 19q13.2 | 12.14 | 6.87E-34 | 0.68 | *PPP1R14A* | 5'UTR |
| cg14131526 | 19 | 38,747,285 | 19q13.2 | 12.14 | 6.49E-34 | 0.71 | *PPP1R14A* | TSS200 |
| cg17252697 | 19 | 38,754,707 | 19q13.2 | -9.59 | 8.98E-22 | 0.33 | *SPINT2* | TSS1500 |
| cg27176870 | 19 | 38,754,930 | 19q13.2 | -8.21 | 2.18E-16 | 0.32 | *SPINT2* | TSS200 |
| cg15470207 | 19 | 38,760,771 | 19q13.2 | 6.65 | 2.99E-11 | 0.38 | *SPINT2* | Intronic |
| cg06538039 | 19 | 38,761,503 | 19q13.2 | 6.14 | 8.21E-10 | 0.57 | *SPINT2* | Intronic |
| cg17840275 | 19 | 38,791,235 | 19q13.2 | 5.88 | 4.17E-09 | 0.29 | *SPINT2* ; *YIF1B* | Intergenic |
| cg24900663 | 19 | 38,810,166 | 19q13.2 | -5.21 | 1.89E-07 | 0.43 | *KCNK6* | TSS1500 |
| cg07786049 | 19 | 38,842,639 | 19q13.2 | -9.18 | 4.30E-20 | 0.23 | *CATSPERG* | Intronic |

| | | | | | | | | |
|---|---|---|---|---|---|---|---|---|
| cg05640294 | 19 | 38,976,794 | 19q13.2 | -7.47 | 7.78E-14 | 0.36 | *RYR1* | Exonic |
| cg21380145 | 19 | 42,069,883 | 19q13.2 | 6.56 | 5.43E-11 | 0.45 | *CEACAM21* | Intronic |
| cg21459123 | 19 | 42,070,397 | 19q13.2 | 6.50 | 7.88E-11 | 0.45 | *CEACAM21* | Intronic |
| cg22963705 | 19 | 42,070,671 | 19q13.2 | 6.29 | 3.10E-10 | 0.21 | *CEACAM21* | Intronic |
| cg23241878 | 19 | 42,082,350 | 19q13.2 | -6.43 | 1.28E-10 | 0.25 | *CEACAM21* | Intronic |
| cg00921609 | 19 | 42,101,873 | 19q13.2 | -6.46 | 1.03E-10 | 0.25 | *CEACAM21* ; *CEACAM4* | Intergenic |
| cg16255631 | 19 | 50,938,708 | 19q13.33 | 9.24 | 2.53E-20 | 0.30 | *MYBPC2* | Intronic |
| cg06521942 | 19 | 50,996,684 | 19q13.33 | 5.63 | 1.84E-08 | 0.26 | *EMC10* ; *JOSD2* | Intergenic |
| cg07370298 | 19 | 51,022,602 | 19q13.33 | -5.27 | 1.36E-07 | 0.26 | *LRRC4B* | Exonic |
| cg19017177 | 19 | 51,141,227 | 19q13.33 | 5.37 | 7.67E-08 | 0.28 | *SYT3* | 5'UTR |
| cg04257123 | 19 | 51,341,745 | 19q13.33 | -6.13 | 8.76E-10 | 0.21 | *LOC105372441* | Intronic |
| cg16742703 | 19 | 51,357,857 | 19q13.33 | 5.53 | 3.13E-08 | 0.51 | *KLK3* | TSS1500 |
| cg15057670 | 19 | 51,359,872 | 19q13.33 | 5.61 | 2.03E-08 | 0.27 | *KLK3* | Intronic |
| cg13261133 | 19 | 51,373,741 | 19q13.33 | -7.42 | 1.14E-13 | 0.32 | *KLK3* ; *KLK2* | Intergenic |
| cg09618904 | 20 | 34,026,478 | 20q11.22 | 5.17 | 2.30E-07 | 0.56 | *GDF5* | Intronic |
| cg10909788 | 20 | 49,537,272 | 20q13.13 | -6.71 | 1.95E-11 | 0.23 | *ADNP* | Intronic |
| cg08479785 | 20 | 49,734,831 | 20q13.13 | 5.74 | 9.39E-09 | 0.24 | *KCNG1* ; *NFATC2* | Intergenic |
| cg05160573 | 20 | 52,384,833 | 20q13.2 | 6.59 | 4.33E-11 | 0.25 | *LOC105372672* ; *SUMO1P1* | Intergenic |
| cg23033986 | 20 | 52,469,879 | 20q13.2 | -7.81 | 5.73E-15 | 0.57 | *LOC105372672* ; *SUMO1P1* | Intergenic |
| cg24475058 | 20 | 61,008,820 | 20q13.33 | -5.44 | 5.21E-08 | 0.60 | *MIR1-1HG-AS1* | Intronic |
| cg18970070 | 20 | 61,012,386 | 20q13.33 | 5.61 | 2.02E-08 | 0.50 | *MIR1-1HG-AS1* | Intronic |
| cg01458054 | 20 | 62,200,603 | 20q13.33 | -5.69 | 1.27E-08 | 0.36 | *HELZ2* | Exonic |
| cg21520607 | 20 | 62,200,854 | 20q13.33 | -5.33 | 1.01E-07 | 0.48 | *HELZ2* | Exonic |
| cg16989086 | 20 | 62,203,971 | 20q13.33 | -5.34 | 9.12E-08 | 0.52 | *HELZ2* | 5'UTR |
| cg03568835 | 20 | 62,209,709 | 20q13.33 | 6.88 | 5.81E-12 | 0.57 | *HELZ2* ; *GMEB2* | Intergenic |
| cg24106834 | 20 | 62,216,768 | 20q13.33 | 5.63 | 1.82E-08 | 0.27 | *HELZ2* ; *GMEB2* | Intergenic |
| cg25903261 | 20 | 62,219,204 | 20q13.33 | 7.48 | 7.25E-14 | 0.47 | *GMEB2* | 3'UTR |
| cg00331541 | 20 | 62,220,778 | 20q13.33 | -6.30 | 2.96E-10 | 0.71 | *GMEB2* | 3'UTR |
| cg13591364 | 20 | 62,220,893 | 20q13.33 | -6.26 | 3.83E-10 | 0.70 | *GMEB2* | 3'UTR |
| cg04747225 | 20 | 62,221,039 | 20q13.33 | -7.33 | 2.33E-13 | 0.72 | *GMEB2* | 3'UTR |
| cg03425785 | 20 | 62,221,387 | 20q13.33 | -7.22 | 5.34E-13 | 0.71 | *GMEB2* | 3'UTR |
| cg10664469 | 20 | 62,223,314 | 20q13.33 | 7.49 | 6.82E-14 | 0.44 | *GMEB2* | Intronic |
| cg01245102 | 20 | 62,236,331 | 20q13.33 | 5.69 | 1.28E-08 | 0.38 | *GMEB2* | Intronic |
| cg04115418 | 20 | 62,273,971 | 20q13.33 | -5.80 | 6.66E-09 | 0.43 | *STMN3* | Intronic |
| cg15320416 | 20 | 62,289,024 | 20q13.33 | -6.04 | 1.50E-09 | 0.23 | *RTEL1* ; *RTEL1-TNFRSF6B* | TSS200 |
| cg27236539 | 20 | 62,289,627 | 20q13.33 | 6.59 | 4.34E-11 | 0.66 | *RTEL1-TNFRSF6B* | Exonic |
| cg06103076 | 20 | 62,296,247 | 20q13.33 | 6.70 | 2.11E-11 | 0.29 | *RTEL1-TNFRSF6B* | Intronic |
| cg13830297 | 20 | 62,314,161 | 20q13.33 | -5.25 | 1.50E-07 | 0.24 | *RTEL1-TNFRSF6B* | Intronic |
| cg00622799 | 20 | 62,318,588 | 20q13.33 | 6.38 | 1.79E-10 | 0.31 | *RTEL1-TNFRSF6B* | Intronic |
| cg08554554 | 20 | 62,330,618 | 20q13.33 | 5.17 | 2.36E-07 | 0.21 | *ARFRP1* | 3'UTR |
| cg15272956 | 20 | 62,332,704 | 20q13.33 | 8.82 | 1.10E-18 | 0.27 | *ARFRP1* | Intronic |
| cg14396214 | 20 | 62,367,998 | 20q13.33 | 5.52 | 3.31E-08 | 0.22 | *LIME1* | Intronic |
| cg17355385 | 20 | 62,368,837 | 20q13.33 | -6.33 | 2.47E-10 | 0.58 | *LIME1* | Intronic |
| cg18725573 | 20 | 62,368,956 | 20q13.33 | -8.10 | 5.62E-16 | 0.70 | *LIME1* | Exonic |
| cg01176363 | 20 | 62,369,445 | 20q13.33 | -8.72 | 2.72E-18 | 0.68 | *LIME1* | Intronic |
| cg21849932 | 20 | 62,369,462 | 20q13.33 | -8.69 | 3.58E-18 | 0.65 | *LIME1* | Intronic |
| cg05643964 | 20 | 62,369,504 | 20q13.33 | -8.61 | 7.13E-18 | 0.64 | *LIME1* | Intronic |
| cg02966332 | 20 | 62,369,605 | 20q13.33 | -8.67 | 4.19E-18 | 0.68 | *LIME1* | Exonic |
| cg14039929 | 20 | 62,369,784 | 20q13.33 | -7.88 | 3.22E-15 | 0.55 | *LIME1* | Exonic |
| cg06226150 | 20 | 62,369,956 | 20q13.33 | -8.47 | 2.51E-17 | 0.41 | *LIME1* | Exonic |
| cg22961457 | 20 | 62,370,310 | 20q13.33 | 8.50 | 1.84E-17 | 0.32 | *LIME1* | 3'UTR |
| cg07196408 | 20 | 62,373,115 | 20q13.33 | -7.40 | 1.39E-13 | 0.51 | *SLC2A4RG* | Intronic |
| cg08258867 | 20 | 62,373,333 | 20q13.33 | -6.44 | 1.19E-10 | 0.42 | *SLC2A4RG* | Exonic |
| cg07294624 | 20 | 62,374,166 | 20q13.33 | -8.19 | 2.52E-16 | 0.44 | *SLC2A4RG* | Intronic |
| cg19655510 | 20 | 62,374,329 | 20q13.33 | -9.52 | 1.69E-21 | 0.47 | *SLC2A4RG* | Exonic |

| | | | | | | | | |
|---|---|---|---|---|---|---|---|---|
| cg21160105 | 20 | 62,384,326 | 20q13.33 | -8.41 | 4.08E-17 | 0.33 | *ZBTB46* | Intronic |
| cg03431903 | 20 | 62,665,188 | 20q13.33 | 5.71 | 1.14E-08 | 0.27 | *C20orf204* | Intronic |
| cg27370028 | 20 | 62,693,943 | 20q13.33 | 5.41 | 6.44E-08 | 0.42 | *TCEA2* | Intronic |
| cg24348495 | 20 | 62,693,971 | 20q13.33 | 6.11 | 9.74E-10 | 0.39 | *TCEA2* | Intronic |
| cg07434253 | 20 | 62,809,306 | 20q13.33 | 5.76 | 8.27E-09 | 0.25 | *MYT1* | Intronic |
| cg11988872 | 21 | 45,349,847 | 21q22.3 | 5.25 | 1.55E-07 | 0.72 | *AGPAT3* | Intronic |
| cg16373820 | 21 | 45,349,849 | 21q22.3 | 5.22 | 1.81E-07 | 0.74 | *AGPAT3* | Intronic |
| cg09574757 | 22 | 19,747,024 | 22q11.21 | -6.74 | 1.59E-11 | 0.28 | *TBX1* | Intronic |
| cg05298458 | 22 | 19,749,188 | 22q11.21 | 6.00 | 2.01E-09 | 0.60 | *TBX1* | Intronic |
| cg09781936 | 22 | 19,749,961 | 22q11.21 | 6.91 | 4.73E-12 | 0.63 | *TBX1* | Intronic |
| cg00204782 | 22 | 19,753,364 | 22q11.21 | -5.66 | 1.54E-08 | 0.47 | *TBX1* | Intronic |
| cg01697719 | 22 | 19,754,125 | 22q11.21 | -6.02 | 1.76E-09 | 0.71 | *TBX1* | Exonic |
| cg08382235 | 22 | 19,754,251 | 22q11.21 | -6.09 | 1.14E-09 | 0.72 | *TBX1* | Exonic |
| cg21507095 | 22 | 19,754,448 | 22q11.21 | -6.14 | 8.29E-10 | 0.68 | *TBX1* | 3'UTR |
| cg05033341 | 22 | 40,419,857 | 22q13.1 | -5.89 | 3.90E-09 | 0.63 | *FAM83F* | Intronic |
| cg15388430 | 22 | 43,010,952 | 22q13.2 | -5.96 | 2.59E-09 | 0.22 | *POLDIP3* | 5'UTR |
| cg25664606 | 22 | 43,356,260 | 22q13.2 | -7.10 | 1.28E-12 | 0.22 | *PACSIN2* | 5'UTR |
| cg01341487 | 22 | 43,505,954 | 22q13.2 | 12.59 | 2.36E-36 | 0.37 | *BIK* | TSS1500 |
| cg26453588 | 22 | 43,506,021 | 22q13.2 | 9.53 | 1.59E-21 | 0.26 | *BIK* | TSS1500 |
| cg20662856 | 22 | 43,506,238 | 22q13.2 | 7.85 | 4.14E-15 | 0.25 | *BIK* | TSS1500 |
| cg21065704 | 22 | 43,506,641 | 22q13.2 | 11.69 | 1.49E-31 | 0.23 | *BIK* | TSS200 |
| cg13458462 | 22 | 43,521,975 | 22q13.2 | 8.77 | 1.77E-18 | 0.36 | *BIK* | Intronic |
| cg05251389 | 22 | 43,525,330 | 22q13.2 | -7.80 | 6.16E-15 | 0.40 | *BIK* | 3'UTR |
| cg13698860 | 22 | 43,525,679 | 22q13.2 | -8.90 | 5.75E-19 | 0.33 | *BIK* | 3'UTR |
| cg27443850 | 22 | 43,547,778 | 22q13.2 | 9.89 | 4.56E-23 | 0.25 | *TSPO* | Intronic |
| cg00343092 | 22 | 43,547,974 | 22q13.2 | 6.29 | 3.24E-10 | 0.48 | *TSPO* | 5'UTR |
| cg08909806 | 22 | 43,548,696 | 22q13.2 | 5.60 | 2.15E-08 | 0.57 | *TSPO* | Intronic |
| cg17456022 | 22 | 43,548,888 | 22q13.2 | 5.62 | 1.94E-08 | 0.66 | *TSPO* | Intronic |
| cg16265130 | 22 | 43,550,246 | 22q13.2 | 7.86 | 3.76E-15 | 0.52 | *TSPO* | Intronic |
| cg24710387 | 22 | 43,551,216 | 22q13.2 | -5.71 | 1.13E-08 | 0.22 | *TSPO* | Intronic |
| cg10697883 | 22 | 43,552,455 | 22q13.2 | 7.68 | 1.53E-14 | 0.37 | *TSPO* | Intronic |
| cg04740405 | 22 | 43,560,830 | 22q13.2 | -7.88 | 3.16E-15 | 0.38 | *TSPO* ; *TTLL12* | Intergenic |
| cg06266566 | 22 | 43,795,397 | 22q13.2 | 5.12 | 3.03E-07 | 0.39 | *LINC01639* | Upstream |
| cg14239351 | 22 | 43,821,115 | 22q13.2 | 5.48 | 4.26E-08 | 0.23 | *MPPED1* | Exonic |
| cg03517341 | 22 | 43,873,298 | 22q13.2 | 8.05 | 7.98E-16 | 0.21 | *MPPED1* | Intronic |

OR, odds ratio per standard deviation (SD) increase in genetically predicted DNA methylation level; CI, confidence interval; TSS, transcription start site; UTR, untranslated region.

[a] Association Z scores, ORs, 95% CIs, and *P* values were estimated using SPrediXcan. All statistical tests were two-sided.

[b] Correlation coefficients of predicted and measured DNA methylation levels.

[c] CpGs that are >1Mb away from any GWAS-identified SNPs and remain significantly associated with prostate cancer risk after conditioning on nearest GWAS signals.

[d] CpGs within 1Mb of GWAS-identified SNPs but remain significantly associated with prostate cancer risk after conditioning on nearest GWAS signals.

**Supplementary Table 7. CpGs showing a significant association with testicular germ cell cancer risk**

| CpG | Chr | Pos | Cytoband | Z score [a] | *P* [a] | R [b] | Closest gene | Classification |
|---|---|---|---|---|---|---|---|---|
| cg15896939 | 1 | 156,030,809 | 1q22 | -7.41 | 1.22E-13 | 0.33 | *RAB25* | TSS200 |
| cg11398659 | 1 | 156,124,471 | 1q22 | -5.14 | 2.77E-07 | 0.49 | *SEMA4A* | Exonic |
| cg25208724 | 1 | 156,163,844 | 1q22 | 5.58 | 2.47E-08 | 0.78 | *SLC25A44* | 5'UTR |
| cg24450063 | 1 | 156,163,899 | 1q22 | 6.91 | 4.90E-12 | 0.67 | *SLC25A44* | 5'UTR |
| cg05112850 | 1 | 156,165,865 | 1q22 | -5.40 | 6.79E-08 | 0.57 | *SLC25A44* | Intronic |
| cg15120454 | 1 | 156,339,198 | 1q22 | -6.21 | 5.43E-10 | 0.31 | *RHBG* | Exonic |
| cg01783153 | 2 | 111,644,949 | 2q13 | -5.31 | 1.10E-07 | 0.34 | *ACOXL* | 3'UTR |
| cg04780086 | 2 | 111,875,790 | 2q13 | 5.51 | 3.67E-08 | 0.38 | *ACOXL* | 3'UTR |
| cg11108060 | 2 | 111,879,843 | 2q13 | -5.27 | 1.40E-07 | 0.30 | *BCL2L11* | Intronic |
| cg18807144 | 2 | 111,906,697 | 2q13 | -5.11 | 3.14E-07 | 0.39 | *BCL2L11* | Intronic |
| cg13319804 | 2 | 111,913,750 | 2q13 | -5.28 | 1.26E-07 | 0.74 | *BCL2L11* | Intronic |
| cg19211484 | 3 | 16,650,168 | 3p24.3 | -7.07 | 1.57E-12 | 0.39 | *DAZL* ; *PLCL2* | Intergenic |
| cg08061417 | 3 | 156,322,035 | 3q25.31 | 6.35 | 2.14E-10 | 0.42 | *SSR3* ; *TIPARP-AS1* | Intergenic |
| cg04067573 | 3 | 169,899,625 | 3q26.2 | 5.59 | 2.26E-08 | 0.33 | *PHC3* | TSS200 |
| cg05967747 | 4 | 95,127,681 | 4q22.3 | -6.75 | 1.48E-11 | 0.32 | *LOC101929210* | Intronic |
| cg14368149 | 4 | 103,932,567 | 4q24 | -5.52 | 3.32E-08 | 0.71 | *SLC9B1* | Intronic |
| cg07932102 | 4 | 103,967,986 | 4q24 | -7.84 | 4.45E-15 | 0.40 | *SLC9B2* | Intronic |
| cg24988451 | 4 | 104,019,643 | 4q24 | 6.09 | 1.15E-09 | 0.38 | *BDH2* | Intronic |
| cg08656504 | 4 | 104,119,605 | 4q24 | 9.28 | 1.64E-20 | 0.41 | *CENPE* | TSS200 |
| cg00260715 | 4 | 104,120,051 | 4q24 | -7.64 | 2.24E-14 | 0.48 | *CENPE* | TSS1500 |
| cg08204402 | 5 | 851,892 | 5p15.33 | -5.87 | 4.27E-09 | 0.39 | *ZDHHC11* | TSS1500 |
| cg19764541 | 5 | 1,063,554 | 5p15.33 | -7.17 | 7.58E-13 | 0.42 | *SLC12A7* | Intronic |
| cg26330510 | 5 | 1,155,853 | 5p15.33 | -10.08 | 6.60E-24 | 0.36 | *SLC12A7* ; *CTD-3080P12.3* | Intergenic |
| cg25045012 | 5 | 1,277,563 | 5p15.33 | 5.75 | 9.07E-09 | 0.55 | *TERT* | Intronic |
| cg00624418 | 5 | 1,282,357 | 5p15.33 | 11.38 | 5.30E-30 | 0.38 | *TERT* | Intronic |
| cg13325231 | 5 | 1,342,170 | 5p15.33 | -6.57 | 4.98E-11 | 0.58 | *CLPTM1L* | Intronic |
| cg07493874 | 5 | 1,342,172 | 5p15.33 | -6.91 | 4.70E-12 | 0.67 | *CLPTM1L* | Intronic |
| cg22739207 | 5 | 1,356,382 | 5p15.33 | 6.88 | 6.01E-12 | 0.42 | *CLPTM1L* ; *LINC01511* | Intergenic |
| cg02579469 | 5 | 1,378,350 | 5p15.33 | -5.76 | 8.65E-09 | 0.32 | *LINC01511* | Intronic |
| cg13982541 | 5 | 1,466,431 | 5p15.33 | -5.18 | 2.26E-07 | 0.52 | *LPCAT1* | Intronic |
| cg05613935 | 5 | 1,667,205 | 5p15.33 | 5.15 | 2.66E-07 | 0.31 | *LOC728613; MIR4277* | Intergenic |
| cg22331671 | 5 | 141,362,962 | 5q31.3 | -9.88 | 5.07E-23 | 0.34 | *RNF14* | Exonic |
| cg21359049 | 5 | 141,385,912 | 5q31.3 | 14.66 | 1.24E-48 | 0.31 | *GNPDA1* | Intronic |
| cg00604900 | 5 | 141,659,783 | 5q31.3 | -11.18 | 5.39E-29 | 0.34 | *NDFIP1; SPRY4* | Intergenic |
| cg12425151 | 5 | 141,663,347 | 5q31.3 | 16.07 | 3.83E-58 | 0.33 | *NDFIP1; SPRY4* | Intergenic |
| cg12947757 | 6 | 33,146,725 | 6p21.32 | -8.90 | 5.35E-19 | 0.44 | *COL11A2* | Exonic |
| cg26757097 | 6 | 33,260,263 | 6p21.32 | -5.13 | 2.94E-07 | 0.39 | *RGL2* | Exonic |
| cg00701755 | 6 | 33,272,136 | 6p21.32 | 5.67 | 1.46E-08 | 0.45 | *TAPBP* | Exonic |
| cg20400383 | 6 | 33,359,642 | 6p21.32 | 5.27 | 1.34E-07 | 0.39 | *KIFC1* | 5'UTR |
| cg23888902 | 6 | 33,385,929 | 6p21.32 | -6.68 | 2.46E-11 | 0.33 | *CUTA* | Exonic |
| cg12607514 | 6 | 33,422,080 | 6p21.32 | 6.58 | 4.61E-11 | 0.37 | *ZBTB9; SYNGAP1* | TSS1500 |
| cg19326232 | 6 | 33,422,095 | 6p21.32 | 7.31 | 2.66E-13 | 0.40 | *ZBTB9; SYNGAP1* | TSS1500 |
| cg13560919 | 6 | 33,536,144 | 6p21.31 | 8.31 | 9.88E-17 | 0.47 | *ZBTB9; BAK1* | Intergenic |
| cg18037239 | 6 | 33,551,492 | 6p21.31 | 14.56 | 5.03E-48 | 0.38 | *GGNBP1* | Exonic |
| cg04201373 | 6 | 33,551,533 | 6p21.31 | 6.39 | 1.68E-10 | 0.45 | *GGNBP1* | Exonic |
| cg10728189 | 6 | 33,679,579 | 6p21.31 | 6.75 | 1.45E-11 | 0.44 | *UQCC2* | TSS200 |
| cg11050792 | 6 | 149,960,166 | 6q25.1 | 5.72 | 1.04E-08 | 0.64 | *KATNA1* | Intronic |
| cg13206674 | 6 | 150,067,644 | 6q25.1 | -5.73 | 1.01E-08 | 0.35 | *NUP43* | 5'UTR |
| cg22239180 | 6 | 150,070,039 | 6q25.1 | -5.83 | 5.47E-09 | 0.45 | *PCMT1* | TSS1500 |
| cg00933542 | 6 | 150,070,202 | 6q25.1 | -5.74 | 9.25E-09 | 0.60 | *PCMT1* | TSS1500 |
| cg12239602 | 6 | 150,087,483 | 6q25.1 | 5.75 | 9.16E-09 | 0.38 | *PCMT1* | Intronic |
| cg11878867 | 6 | 150,167,359 | 6q25.1 | 5.71 | 1.16E-08 | 0.43 | *LRP11* | Intronic |
| cg23544286 | 6 | 150,183,650 | 6q25.1 | -5.21 | 1.90E-07 | 0.38 | *LRP11* | Intronic |
| cg19626747 | 6 | 150,205,807 | 6q25.1 | -5.65 | 1.63E-08 | 0.44 | *RAET1E-AS1* | Intronic |

| | | | | | | | | |
|---|---|---|---|---|---|---|---|---|
| cg18538426 | 6 | 150,231,004 | 6q25.1 | -5.10 | 3.39E-07 | 0.46 | *RAET1E-AS1* | Intronic |
| cg18702839 | 6 | 150,238,296 | 6q25.1 | -5.59 | 2.28E-08 | 0.42 | *RAET1E-AS1* | Intronic |
| cg19673125 | 6 | 150,240,577 | 6q25.1 | -5.48 | 4.35E-08 | 0.32 | *RAET1E-AS1* | Exonic |
| cg25987200 | 6 | 150,254,830 | 6q25.1 | -5.29 | 1.22E-07 | 0.50 | *LOC105378052* | Upstream |
| cg07132326 | 6 | 150,258,058 | 6q25.1 | -5.65 | 1.62E-08 | 0.31 | *LOC105378052* | Downstream |
| cg00552369 | 7 | 1,521,973 | 7p22.3 | 7.37 | 1.65E-13 | 0.36 | *INTS1* | Intronic |
| cg17324095 | 7 | 1,783,946 | 7p22.3 | 5.17 | 2.35E-07 | 0.39 | *ELFN1* | 5'UTR |
| cg09911010 | 7 | 1,819,220 | 7p22.3 | 6.64 | 3.21E-11 | 0.36 | *ELFN1; MAD1L1* | Intergenic |
| cg12432903 | 7 | 1,882,776 | 7p22.3 | -8.53 | 1.46E-17 | 0.56 | *MAD1L1* | Intronic |
| cg22365446 | 7 | 1,885,050 | 7p22.3 | 7.56 | 3.91E-14 | 0.48 | *MAD1L1* | Intronic |
| cg25122824 | 7 | 1,886,917 | 7p22.3 | -8.04 | 9.19E-16 | 0.59 | *MAD1L1* | Intronic |
| cg26580761 | 7 | 1,894,151 | 7p22.3 | 8.55 | 1.23E-17 | 0.66 | *MAD1L1* | Intronic |
| cg05863683 | 7 | 1,912,471 | 7p22.3 | 9.02 | 1.88E-19 | 0.48 | *MAD1L1* | Intronic |
| cg08006332 | 7 | 1,928,891 | 7p22.3 | 7.80 | 6.00E-15 | 0.37 | *MAD1L1* | Intronic |
| cg12678507 | 7 | 1,934,097 | 7p22.3 | 7.27 | 3.54E-13 | 0.38 | *MAD1L1* | Intronic |
| cg10028823 | 7 | 1,952,561 | 7p22.3 | 6.10 | 1.06E-09 | 0.37 | *MAD1L1* | Intronic |
| cg02311337 | 7 | 1,955,432 | 7p22.3 | -8.11 | 5.20E-16 | 0.33 | *MAD1L1* | Intronic |
| cg04342202 | 7 | 1,961,968 | 7p22.3 | 5.79 | 7.18E-09 | 0.32 | *MAD1L1* | Intronic |
| cg24189917 | 7 | 1,970,923 | 7p22.3 | 5.11 | 3.15E-07 | 0.30 | *MAD1L1* | Intronic |
| cg13791269 | 7 | 1,971,233 | 7p22.3 | 5.10 | 3.35E-07 | 0.56 | *MAD1L1* | Intronic |
| cg03634735 | 7 | 1,992,524 | 7p22.3 | 6.71 | 1.91E-11 | 0.34 | *MAD1L1* | Intronic |
| cg00183100 | 7 | 2,006,518 | 7p22.3 | 6.05 | 1.49E-09 | 0.32 | *MAD1L1* | Intronic |
| cg22340370 | 7 | 2,019,882 | 7p22.3 | 8.22 | 2.06E-16 | 0.50 | *MAD1L1* | Intronic |
| cg23048001 | 7 | 2,026,167 | 7p22.3 | -7.54 | 4.80E-14 | 0.53 | *MAD1L1* | Intronic |
| cg21782813 | 7 | 2,030,301 | 7p22.3 | 7.52 | 5.44E-14 | 0.71 | *MAD1L1* | Intronic |
| cg08962362 | 7 | 2,033,847 | 7p22.3 | -8.42 | 3.83E-17 | 0.31 | *MAD1L1* | Intronic |
| cg18279126 | 7 | 2,041,391 | 7p22.3 | 5.54 | 3.02E-08 | 0.51 | *MAD1L1* | Intronic |
| cg08722586 | 7 | 2,046,821 | 7p22.3 | 6.19 | 5.92E-10 | 0.34 | *MAD1L1* | Intronic |
| cg02951883 | 7 | 2,050,386 | 7p22.3 | -7.78 | 7.24E-15 | 0.72 | *MAD1L1* | Intronic |
| cg17354002 | 7 | 2,071,166 | 7p22.3 | -5.53 | 3.24E-08 | 0.59 | *MAD1L1* | Intronic |
| cg08972190 | 7 | 2,138,995 | 7p22.3 | 5.70 | 1.17E-08 | 0.52 | *MAD1L1* | Intronic |
| cg19146587 | 7 | 2,165,490 | 7p22.3 | 6.54 | 6.17E-11 | 0.31 | *MAD1L1* | Intronic |
| cg02568153 | 7 | 2,184,201 | 7p22.3 | -6.93 | 4.29E-12 | 0.31 | *MAD1L1* | Intronic |
| cg20185615 | 7 | 2,316,906 | 7p22.3 | -5.40 | 6.54E-08 | 0.40 | *SNX8* | Intronic |
| cg01655824 | 8 | 71,471,544 | 8q13.3 | -5.19 | 2.10E-07 | 0.36 | *LOC101926892; TRAM1* | Intergenic |
| cg08198861 | 8 | 71,485,402 | 8q13.3 | -5.27 | 1.34E-07 | 0.32 | *TRAM1* | Downstream |
| cg02324356 | 9 | 271,757 | 9p24.3 | -5.47 | 4.62E-08 | 0.36 | *DOCK8* | Intronic |
| cg11281704 | 9 | 469,824 | 9p24.3 | -6.40 | 1.57E-10 | 0.34 | *KANK1* | TSS1500 |
| cg08677535 | 9 | 505,721 | 9p24.3 | -13.57 | 5.64E-42 | 0.39 | *KANK1* | Intronic |
| cg14021170 | 9 | 826,657 | 9p24.3 | 12.66 | 1.04E-36 | 0.51 | *KANK1; DMRT1* | Intergenic |
| cg01137471 | 9 | 841,340 | 9p24.3 | 8.84 | 9.51E-19 | 0.35 | *DMRT1* | TSS1500 |
| cg02023728 | 11 | 77,925,099 | 11q14.1 | 5.44 | 5.26E-08 | 0.43 | *USP35* | 3'UTR |
| cg18577493 | 11 | 78,001,065 | 11q14.1 | -5.51 | 3.62E-08 | 0.34 | *GAB2* | Intronic |
| cg22494377 | 11 | 78,023,011 | 11q14.1 | -5.60 | 2.15E-08 | 0.36 | *GAB2* | Intronic |
| cg00042736 | 11 | 78,251,475 | 11q14.1 | 5.41 | 6.48E-08 | 0.41 | *LOC101928896* | Intronic |
| cg21366298 | 11 | 78,278,518 | 11q14.1 | 5.27 | 1.34E-07 | 0.37 | *NARS2* | Intronic |
| cg17045582 | 11 | 125,081,966 | 11q24.2 | 5.15 | 2.67E-07 | 0.56 | *PKNOX2* | Intronic |
| cg03538663 | 12 | 88,423,137 | 12q21.32 | 7.09 | 1.35E-12 | 0.36 | *C12orf50* | 5'UTR |
| cg01309052 | 12 | 88,428,972 | 12q21.32 | -6.06 | 1.32E-09 | 0.31 | *C12orf29* | TSS1500 |
| cg23471314 | 12 | 88,429,605 | 12q21.32 | -9.97 | 1.96E-23 | 0.68 | *C12orf29* | Intronic |
| cg12161468 | 12 | 88,953,786 | 12q21.32 | -21.72 | 1.32E-104 | 0.45 | *KITLG* | Intronic |
| cg22688836 | 12 | 88,967,594 | 12q21.32 | -5.16 | 2.44E-07 | 0.41 | *KITLG* | Intronic |
| cg17396834 | 15 | 56,014,697 | 15q21.3 | -8.27 | 1.38E-16 | 0.32 | *PRTG* | Intronic |
| cg17221584 | 15 | 56,033,606 | 15q21.3 | 8.52 | 1.64E-17 | 0.57 | *PRTG* | Intronic |
| cg13982454 | 15 | 56,034,020 | 15q21.3 | 8.10 | 5.31E-16 | 0.44 | *PRTG* | Intronic |
| cg10982231 | 15 | 56,536,562 | 15q21.3 | 5.57 | 2.52E-08 | 0.33 | *RFX7* | Intronic |
| cg14635047 | 16 | 50,099,262 | 16q12.1 | -5.11 | 3.29E-07 | 0.38 | *HEATR3* | TSS1500 |

| | | | | | | | | |
|---|---|---|---|---|---|---|---|---|
| cg01561092 | 16 | 50,100,815 | 16q12.1 | -5.62 | 1.90E-08 | 0.47 | *HEATR3* | 5'UTR |
| cg21039885 | 16 | 74,498,381 | 16q23.1 | -6.00 | 1.95E-09 | 0.54 | *GLG1* | Intronic |
| cg03628122 | 16 | 74,666,669 | 16q23.1 | -6.26 | 3.82E-10 | 0.44 | *RFWD3* | Intronic |
| cg15874865 | 16 | 74,698,901 | 16q23.1 | 7.08 | 1.40E-12 | 0.48 | *RFWD3* | Intronic |
| cg03930899 | 16 | 74,701,487 | 16q23.1 | 7.03 | 2.10E-12 | 0.45 | *RFWD3* | TSS1500 |
| cg26884787 | 16 | 88,543,447 | 16q24.2 | 6.86 | 6.71E-12 | 0.54 | *ZFPM1* | Intronic |
| cg06212670 | 16 | 88,543,627 | 16q24.2 | -6.13 | 8.85E-10 | 0.39 | *ZFPM1* | Intronic |
| cg08374494 | 16 | 88,551,489 | 16q24.2 | 6.75 | 1.43E-11 | 0.57 | *ZFPM1* | Intronic |
| cg11497526 | 17 | 35,902,366 | 17q12 | -5.54 | 3.00E-08 | 0.49 | *SYNRG* | Exonic |
| cg19378036 | 17 | 36,105,364 | 17q12 | -6.39 | 1.66E-10 | 0.35 | *HNF1B* | TSS1500 |
| cg04198914 | 17 | 36,106,025 | 17q12 | -7.80 | 6.11E-15 | 0.44 | *HNF1B* | TSS1500 |
| cg04105270 | 17 | 36,106,301 | 17q12 | -9.45 | 3.30E-21 | 0.44 | *HNF1B; YWHAEP7* | Intergenic |
| cg13062819 | 17 | 56,428,420 | 17q22 | 6.23 | 4.76E-10 | 0.43 | *TSPOAP1-AS1* | Intronic |
| cg02118635 | 17 | 56,770,003 | 17q22 | 6.24 | 4.34E-10 | 0.43 | *RAD51C* | 5'UTR |
| cg10487724 | 17 | 56,770,010 | 17q22 | 5.26 | 1.44E-07 | 0.45 | *RAD51C* | Exonic |
| cg23459817 | 17 | 56,806,313 | 17q22 | 5.98 | 2.17E-09 | 0.33 | *RAD51C* | Intronic |
| cg27528727 | 17 | 56,864,423 | 17q22 | 6.30 | 2.95E-10 | 0.53 | *PPM1E* | Intronic |
| cg02765693 | 17 | 57,043,425 | 17q22 | -7.46 | 8.58E-14 | 0.35 | *PPM1E* | Intronic |
| cg19379959 | 17 | 57,232,725 | 17q22 | 6.98 | 2.93E-12 | 0.33 | *PRR11* | 5'UTR |
| cg24955191 | 17 | 57,255,866 | 17q22 | -5.37 | 7.73E-08 | 0.38 | *PRR11* | Intronic |
| cg22717650 | 17 | 57,292,547 | 17q22 | -5.09 | 3.64E-07 | 0.33 | *SMG8* | 3'UTR |
| cg15443223 | 17 | 57,401,781 | 17q22 | 5.83 | 5.40E-09 | 0.72 | *GDPD1; YPEL2* | Intergenic |
| cg18488204 | 19 | 21,658,645 | 19p12 | 5.30 | 1.19E-07 | 0.35 | *ZNF493; LINC00664* | Intergenic |
| cg02937080 | 19 | 23,183,750 | 19p12 | -6.60 | 4.00E-11 | 0.35 | *ZNF728* | Intronic |
| cg23929809 | 21 | 47,602,432 | 21q22.3 | -5.08 | 3.69E-07 | 0.51 | *SPATC1L* | Intronic |
| cg23289605 | 21 | 47,614,477 | 21q22.3 | -6.61 | 3.90E-11 | 0.62 | *LSS* | Exonic |
| cg12120016 | 21 | 47,639,967 | 21q22.3 | -5.81 | 6.42E-09 | 0.47 | *LSS* | Intronic |
| cg09831081 | 21 | 47,647,568 | 21q22.3 | -5.67 | 1.40E-08 | 0.51 | *LSS* | Exonic |
| cg11715092 | 21 | 47,705,294 | 21q22.3 | 5.96 | 2.50E-09 | 0.36 | *MCM3AP* | 5'UTR |
| cg10486505 | 21 | 47,743,117 | 21q22.3 | -6.25 | 4.19E-10 | 0.31 | *C21orf58* | 5'UTR |
| cg05196905 | 21 | 47,877,665 | 21q22.3 | 5.62 | 1.91E-08 | 0.43 | *PCNT*; *DIP2A* | Intergenic |
| cg18042004 [c] | 22 | 41,940,657 | 22q13.2 | -5.15 | 2.65E-07 | 0.38 | *POLR3H* | TSS200 |
| cg11677105 [c] | 22 | 42,078,707 | 22q13.2 | 5.17 | 2.37E-07 | 0.44 | *SNU13* | Intronic |
| cg11536612 [c] | 22 | 42,078,723 | 22q13.2 | 5.23 | 1.74E-07 | 0.43 | *SNU13* | Intronic |

OR, odds ratio per standard deviation (SD) increase in genetically predicted DNA methylation level; CI, confidence interval; TSS, transcription start site; UTR, untranslated region.

[a] Association Z scores, ORs, 95% CIs, and *P* values were estimated using SPrediXcan. All statistical tests were two-sided.

[b] Correlation coefficients of predicted and measured DNA methylation levels.

[c] CpGs that are >1Mb away from any GWAS-identified SNPs and remain significantly associated with testicular germ cell cancer risk after conditioning on nearest GWAS signals.

**Supplementary Table 8. Breast cancer susceptibility CpGs showing consistent differential DNA methylation between breast cancer tumor and adjacent normal tissues in TCGA**

| CpG | CpG-cancer associations | | Differential methylation (TCGA) | |
|---|---|---|---|---|
| | Z score | *P* | Effect [a] | *P* [a] |
| cg08353955 | -20.70 | 3.48E-95 | -0.33 | 0.03 |
| cg21032395 | 15.29 | 9.35E-53 | 0.43 | 4.37E-03 |
| cg14557534 | -14.59 | 3.47E-48 | -0.60 | 2.40E-06 |
| cg13560030 | -13.14 | 1.91E-39 | -0.50 | 1.26E-03 |
| cg18204589 | 11.15 | 7.31E-29 | 0.56 | 1.54E-04 |
| cg14861570 | -10.21 | 1.88E-24 | -0.59 | 2.04E-04 |
| cg02258246 | 9.96 | 2.22E-23 | 0.38 | 0.01 |
| cg27567761 | 9.52 | 1.74E-21 | 1.31 | 5.59E-14 |
| cg09529667 | -9.43 | 4.01E-21 | -0.28 | 0.03 |
| cg17710779 | -9.00 | 2.18E-19 | -0.78 | 3.27E-06 |
| cg14306534 | 8.12 | 4.72E-16 | 0.52 | 5.85E-04 |
| cg22633036 | -7.97 | 1.54E-15 | -0.48 | 1.25E-03 |
| cg10511902 | 7.41 | 1.30E-13 | 0.36 | 0.02 |
| cg23478547 | -7.27 | 3.55E-13 | -0.65 | 2.66E-05 |
| cg20207973 | -7.24 | 4.57E-13 | -0.33 | 0.04 |
| cg06498964 | -6.99 | 2.72E-12 | -0.54 | 3.84E-04 |
| cg25457927 | 6.98 | 3.02E-12 | 0.20 | 0.03 |
| cg22872885 | -6.92 | 4.61E-12 | -0.31 | 0.03 |
| cg16228356 | 6.91 | 4.85E-12 | 1.15 | 1.94E-12 |
| cg01882395 | -6.87 | 6.50E-12 | -0.41 | 6.99E-03 |
| cg04055107 | 6.74 | 1.59E-11 | 0.36 | 7.38E-03 |
| cg18032502 | 6.68 | 2.34E-11 | 1.23 | 1.10E-15 |
| cg08337959 | 6.61 | 3.77E-11 | 1.26 | 3.71E-15 |
| cg07241660 | -6.56 | 5.53E-11 | -0.42 | 4.76E-03 |
| cg20931134 | -6.49 | 8.68E-11 | -1.01 | 4.97E-11 |
| cg08158518 | -6.47 | 9.80E-11 | -1.29 | 1.76E-15 |
| cg01689657 | 6.41 | 1.44E-10 | 0.41 | 2.06E-04 |
| cg10224600 | -6.33 | 2.38E-10 | -0.66 | 3.09E-05 |
| cg06733794 | -6.31 | 2.72E-10 | -0.32 | 0.02 |
| cg01934064 | -6.30 | 2.99E-10 | -0.41 | 6.08E-03 |
| cg18362448 | -6.24 | 4.50E-10 | -0.31 | 0.02 |
| cg26263234 | 6.22 | 4.85E-10 | 0.49 | 9.23E-04 |
| cg05511855 | -6.10 | 1.09E-09 | -0.52 | 8.72E-04 |
| cg18604888 | 6.03 | 1.67E-09 | 0.37 | 0.01 |
| cg07152837 | -5.99 | 2.08E-09 | -0.34 | 0.03 |
| cg06046490 | 5.97 | 2.44E-09 | 1.02 | 6.78E-11 |
| cg03854071 | -5.82 | 5.97E-09 | -0.38 | 0.01 |
| cg22842085 | -5.82 | 6.00E-09 | -0.39 | 8.57E-03 |
| cg03714773 | 5.70 | 1.19E-08 | 0.36 | 1.85E-03 |
| cg05231888 | -5.70 | 1.23E-08 | -0.76 | 8.19E-08 |
| cg07104417 | -5.68 | 1.32E-08 | -0.62 | 5.09E-05 |
| cg24895155 | 5.56 | 2.68E-08 | 0.31 | 0.02 |
| cg14124917 | -5.52 | 3.43E-08 | -0.78 | 3.60E-08 |
| cg02465979 | -5.47 | 4.38E-08 | -0.30 | 0.04 |

| | | | | |
|---|---|---|---|---|
| cg19019371 | -5.44 | 5.32E-08 | -0.44 | 1.26E-03 |
| cg11688376 | -5.39 | 7.03E-08 | -0.26 | 0.05 |
| cg06782035 | -5.38 | 7.27E-08 | -0.41 | 7.19E-03 |
| cg14250997 | 5.36 | 8.28E-08 | 0.30 | 0.03 |
| cg17111895 | -5.31 | 1.09E-07 | -0.30 | 0.05 |
| cg22415969 | -5.29 | 1.24E-07 | -0.59 | 2.33E-04 |
| cg11029475 | 5.28 | 1.26E-07 | 0.61 | 6.38E-07 |
| cg20054786 | 5.27 | 1.39E-07 | 0.42 | 5.47E-03 |
| cg00609961 | -5.26 | 1.43E-07 | -0.66 | 3.15E-05 |
| cg20303441 | -5.22 | 1.83E-07 | -0.33 | 2.34E-03 |
| cg21679810 | 5.16 | 2.48E-07 | 0.42 | 2.96E-03 |
| cg07160278 | 5.15 | 2.63E-07 | 0.30 | 0.03 |
| cg02327530 | 5.13 | 2.94E-07 | 0.27 | 0.05 |
| cg16465129 | 5.13 | 2.95E-07 | 0.49 | 1.51E-03 |
| cg22652561 | 5.12 | 3.09E-07 | 0.29 | 0.04 |
| cg26218269 | 5.09 | 3.57E-07 | 1.02 | 1.96E-11 |
| cg21138405 | -5.08 | 3.69E-07 | -0.90 | 3.37E-10 |
| cg20980112 | -5.07 | 3.99E-07 | -0.38 | 0.02 |
| cg02887458 | -5.07 | 4.05E-07 | -0.67 | 2.19E-05 |

TCGA, The Cancer Genome Atlas.

[a] Differential DNA methylation analyses were performed using Illumina 450K BeadChip data of 791 breast invasive carcinoma and 97 adjacent normal tissue samples from TCGA-BRCA. Effect sizes and *P* values were estimated using linear mixed models, adjusting for age and top three DNA methylation principle components (PCs).

**Supplementary Table 9. Colorectal cancer susceptibility CpGs showing consistent differential DNA methylation between colorectal cancer tumor and adjacent normal tissues in TCGA**

| CpG | CpG-cancer associations | | Differential methylation (TCGA) | |
|---|---|---|---|---|
| | Z score | *P* | Fold change [a] | *P* [a] |
| cg20556304 | 12.53 | 5.06E-36 | 1.03 | 3.37E-05 |
| cg05820396 | -12.36 | 4.47E-35 | -0.48 | 0.01 |
| cg11909137 | -12.17 | 4.24E-34 | -1.58 | 4.59E-07 |
| cg02605258 | 11.34 | 8.16E-30 | 0.83 | 1.97E-03 |
| cg07689907 | -9.44 | 3.85E-21 | -0.83 | 2.17E-03 |
| cg03907575 | -9.02 | 1.82E-19 | -0.75 | 9.42E-04 |
| cg24519393 | -8.45 | 2.89E-17 | -0.74 | 2.45E-03 |
| cg00387115 | -8.44 | 3.22E-17 | -0.80 | 4.39E-05 |
| cg05202204 | -8.42 | 3.62E-17 | -0.68 | 3.72E-03 |
| cg01034396 | 8.37 | 5.66E-17 | 1.23 | 3.19E-06 |
| cg27176264 | 8.29 | 1.10E-16 | 0.61 | 5.42E-03 |
| cg02730678 | -8.16 | 3.34E-16 | -0.64 | 0.02 |
| cg06902929 | 7.89 | 3.11E-15 | 0.74 | 6.85E-04 |
| cg10101109 | 7.88 | 3.32E-15 | 0.63 | 0.03 |
| cg27598979 | 7.87 | 3.61E-15 | 1.61 | 2.10E-07 |
| cg02584939 | 7.76 | 8.29E-15 | 0.72 | 4.78E-03 |
| cg26528311 | -7.67 | 1.71E-14 | -0.64 | 1.60E-03 |
| cg15082992 | 7.35 | 2.02E-13 | 0.67 | 0.01 |
| cg11920449 | -7.26 | 4.00E-13 | -0.93 | 1.93E-04 |
| cg01101522 | 7.07 | 1.53E-12 | 0.69 | 1.85E-03 |
| cg09037630 | -6.98 | 3.01E-12 | -0.66 | 2.56E-05 |
| cg04105091 | -6.95 | 3.63E-12 | -0.78 | 4.58E-03 |
| cg04797846 | -6.90 | 5.14E-12 | -0.78 | 1.01E-03 |
| cg18595780 | -6.65 | 2.95E-11 | -0.69 | 8.12E-04 |
| cg07663732 | -6.60 | 4.10E-11 | -0.51 | 0.03 |
| cg26954695 | -6.57 | 5.10E-11 | -0.74 | 1.23E-04 |
| cg06879302 | 6.52 | 7.02E-11 | 1.96 | 8.88E-09 |
| cg02773045 | 6.51 | 7.47E-11 | 0.51 | 0.03 |
| cg23531748 | -6.49 | 8.72E-11 | -0.61 | 8.05E-03 |
| cg15106294 | 6.35 | 2.22E-10 | 0.62 | 0.02 |
| cg25415966 | 6.30 | 3.02E-10 | 0.85 | 2.85E-05 |
| cg25273936 | 6.28 | 3.33E-10 | 0.47 | 0.02 |
| cg13876325 | -6.19 | 5.98E-10 | -1.24 | 1.49E-07 |
| cg16518266 | -6.14 | 8.03E-10 | -0.52 | 0.02 |
| cg16615454 | -6.12 | 9.49E-10 | -0.73 | 1.84E-03 |
| cg02985381 | 6.12 | 9.57E-10 | 0.61 | 0.02 |
| cg01400685 | 6.11 | 9.77E-10 | 0.78 | 4.22E-04 |
| cg03894242 | 6.06 | 1.32E-09 | 0.76 | 7.10E-03 |
| cg06855286 | -6.05 | 1.43E-09 | -0.50 | 0.03 |
| cg23917844 | 6.05 | 1.46E-09 | 0.42 | 0.03 |
| cg16585132 | -6.05 | 1.49E-09 | -0.73 | 6.85E-04 |
| cg24875484 | -6.03 | 1.61E-09 | -1.01 | 4.68E-06 |

| | | | | |
|---|---|---|---|---|
| cg09661503 | 6.03 | 1.62E-09 | 1.07 | 1.33E-04 |
| cg26433975 | -5.99 | 2.04E-09 | -0.27 | 0.04 |
| cg19341630 | 5.97 | 2.42E-09 | 0.53 | 0.03 |
| cg10384570 | 5.94 | 2.85E-09 | 0.56 | 0.02 |
| cg12670525 | 5.82 | 5.72E-09 | 0.75 | 5.35E-03 |
| cg16398511 | -5.81 | 6.17E-09 | -0.74 | 1.34E-03 |
| cg11613427 | 5.81 | 6.27E-09 | 1.05 | 2.71E-06 |
| cg14748795 | -5.80 | 6.47E-09 | -0.57 | 1.53E-03 |
| cg20800216 | -5.79 | 7.00E-09 | -0.52 | 0.05 |
| cg00686197 | 5.78 | 7.61E-09 | 1.16 | 1.09E-05 |
| cg07786254 | -5.70 | 1.17E-08 | -0.62 | 5.72E-03 |
| cg21214455 | -5.70 | 1.23E-08 | -0.46 | 0.04 |
| cg20080983 | 5.69 | 1.25E-08 | 1.11 | 1.86E-04 |
| cg23646888 | -5.69 | 1.28E-08 | -0.49 | 8.38E-03 |
| cg24216966 | 5.66 | 1.53E-08 | 0.76 | 1.55E-03 |
| cg10591797 | -5.63 | 1.75E-08 | -0.92 | 5.42E-04 |
| cg12738718 | -5.63 | 1.84E-08 | -0.81 | 2.16E-03 |
| cg07007382 | -5.62 | 1.96E-08 | -0.79 | 5.14E-04 |
| cg17709873 | 5.58 | 2.43E-08 | 1.00 | 8.82E-05 |
| cg08610982 | -5.55 | 2.80E-08 | -0.80 | 8.70E-05 |
| cg00086577 | 5.55 | 2.94E-08 | 0.80 | 2.63E-04 |
| cg14050782 | 5.54 | 2.95E-08 | 0.59 | 0.03 |
| cg26915799 | -5.54 | 3.02E-08 | -0.54 | 0.02 |
| cg21222743 | -5.52 | 3.35E-08 | -0.67 | 0.01 |
| cg07749664 | -5.50 | 3.80E-08 | -0.76 | 2.52E-03 |
| cg00674004 | 5.50 | 3.81E-08 | 0.85 | 6.36E-04 |
| cg05133205 | -5.47 | 4.42E-08 | -0.51 | 0.03 |
| cg26818629 | -5.47 | 4.47E-08 | -0.52 | 7.26E-03 |
| cg12128696 | 5.46 | 4.80E-08 | 0.65 | 0.02 |
| cg19674797 | 5.43 | 5.53E-08 | 0.54 | 0.02 |
| cg23805594 | -5.39 | 7.06E-08 | -0.52 | 0.02 |
| cg02002586 | 5.39 | 7.12E-08 | 1.17 | 2.45E-06 |
| cg14731391 | -5.34 | 9.18E-08 | -0.62 | 0.02 |
| cg26668675 | -5.33 | 9.65E-08 | -0.41 | 5.56E-03 |
| cg09179646 | -5.31 | 1.08E-07 | -0.37 | 0.01 |
| cg12169661 | -5.31 | 1.09E-07 | -0.47 | 0.04 |
| cg16235197 | 5.30 | 1.13E-07 | 0.48 | 0.01 |
| cg03552151 | -5.29 | 1.25E-07 | -0.72 | 0.02 |
| cg21052932 | -5.28 | 1.28E-07 | -1.09 | 2.18E-05 |
| cg26101402 | -5.26 | 1.42E-07 | -0.76 | 9.27E-03 |
| cg03185022 | -5.26 | 1.43E-07 | -0.62 | 0.02 |
| cg03835151 | 5.26 | 1.44E-07 | 1.24 | 2.85E-05 |
| cg17931227 | -5.24 | 1.64E-07 | -0.40 | 4.55E-03 |
| cg02575483 | -5.21 | 1.88E-07 | -1.59 | 4.79E-08 |
| cg12585943 | -5.20 | 2.01E-07 | -0.94 | 3.33E-04 |
| cg14036627 | -5.19 | 2.06E-07 | -0.31 | 0.03 |

TCGA, The Cancer Genome Atlas.

[a] Differential DNA methylation analyses were performed using Illumina 450K BeadChip data of 404 colorectal adenocarcinoma (306 colon and 98 rectum) and 45 adjacent normal tissue samples from TCGA-COAD and TCGA-READ. Effect sizes and *P* values were estimated using linear mixed models, adjusting for age, sex, tumor subtype, and top three DNA methylation PCs.

**Supplementary Table 10. Renal cell cancer susceptibility CpGs showing consistent differential DNA methylation between renal cell cancer and adjacent normal tissues in TCGA**

| CpG | CpG-cancer associations | | Differential methylation (TCGA) | |
|---|---|---|---|---|
| | Z score | ***P*** | Fold change [a] | ***P*** [a] |
| cg10755058 | -5.20 | 1.94E-07 | -0.68 | 1.48E-04 |
| cg06917998 | -5.20 | 2.00E-07 | -0.50 | 4.74E-03 |

TCGA, The Cancer Genome Atlas.

[a] Differential DNA methylation analyses were performed using Illumina 450K BeadChip data of 597 RCC (275 papillary and 322 clear cell carcinoma) and 205 adjacent normal tissue samples from TCGA-KIRP and TCGA-KIRC. effect sizes and *P* values were estimated using linear mixed models, adjusting for age, sex, tumor subtype, and top three DNA methylation PCs.

**Supplementary Table 11. Lung cancer susceptibility CpGs showing consistent differential DNA methylation between lung cancer tumor and adjacent normal tissues in TCGA**

| CpG | CpG-cancer associations | | Differential methylation (TCGA) | |
|---|---|---|---|---|
| | Z score | *P* | Effect size [a] | *P* [a] |
| cg08092050 | 19.28 | 7.62E-83 | 0.33 | 0.03 |
| cg08426200 | 13.24 | 5.41E-40 | 0.39 | 0.02 |
| cg21202862 | -12.06 | 1.80E-33 | -0.44 | 5.23E-03 |
| cg26209169 | -11.57 | 5.76E-31 | -0.57 | 9.26E-06 |
| cg11624060 | -11.54 | 8.48E-31 | -0.53 | 1.01E-04 |
| cg10441424 | -11.38 | 5.26E-30 | -0.60 | 2.16E-04 |
| cg04140906 | -11.08 | 1.59E-28 | -0.39 | 0.02 |
| cg01786994 | -9.12 | 7.31E-20 | -0.65 | 8.77E-04 |
| cg09476067 | -8.88 | 6.67E-19 | -0.41 | 6.49E-03 |
| cg19579005 | -8.66 | 4.69E-18 | -0.89 | 1.31E-08 |
| cg09508356 | -8.10 | 5.71E-16 | -0.59 | 1.09E-04 |
| cg20802826 | -7.90 | 2.90E-15 | -0.64 | 2.03E-05 |
| cg15745284 | -7.88 | 3.38E-15 | -0.42 | 7.71E-03 |
| cg04560098 | 7.41 | 1.23E-13 | 0.56 | 6.43E-05 |
| cg00061860 | -7.20 | 5.89E-13 | -0.45 | 0.01 |
| cg11358777 | 7.17 | 7.29E-13 | 1.68 | 2.57E-25 |
| cg12463578 | -7.11 | 1.18E-12 | -0.38 | 5.57E-03 |
| cg06061002 | 6.95 | 3.59E-12 | 1.10 | 1.21E-13 |
| cg00787996 | -6.90 | 5.34E-12 | -0.68 | 3.86E-05 |
| cg16868572 | 6.88 | 5.97E-12 | 0.50 | 2.71E-03 |
| cg13537510 | 6.81 | 9.95E-12 | 0.84 | 1.26E-05 |
| cg12833048 | 6.78 | 1.18E-11 | 0.52 | 2.51E-03 |
| cg23880581 | -6.76 | 1.39E-11 | -0.44 | 8.17E-03 |
| cg16809040 | 6.70 | 2.09E-11 | 0.32 | 0.03 |
| cg12914966 | -6.64 | 3.18E-11 | -0.59 | 4.41E-04 |
| cg02014853 | -6.63 | 3.30E-11 | -0.53 | 1.50E-04 |
| cg11485823 | 6.62 | 3.69E-11 | 0.54 | 1.15E-03 |
| cg20927242 | -6.62 | 3.71E-11 | -0.49 | 1.62E-03 |
| cg05053818 | -6.58 | 4.70E-11 | -0.48 | 2.10E-03 |
| cg18940843 | -6.58 | 4.73E-11 | -0.71 | 5.29E-06 |
| cg21519219 | 6.53 | 6.38E-11 | 1.35 | 8.18E-19 |
| cg05680196 | 6.50 | 8.14E-11 | 0.77 | 1.90E-06 |
| cg00903577 | -6.40 | 1.53E-10 | -0.89 | 5.65E-08 |
| cg05133205 | -6.37 | 1.88E-10 | -0.46 | 1.16E-03 |
| cg23756442 | -6.35 | 2.17E-10 | -0.50 | 2.06E-03 |
| cg26152188 | -6.28 | 3.42E-10 | -1.31 | 2.97E-16 |
| cg22184136 | -6.27 | 3.66E-10 | -0.59 | 5.88E-05 |
| cg25729445 | -6.23 | 4.58E-10 | -0.30 | 0.04 |
| cg21100518 | -6.06 | 1.35E-09 | -0.51 | 4.46E-04 |
| cg16751781 | -6.05 | 1.44E-09 | -0.79 | 2.04E-06 |
| cg05875819 | -6.00 | 1.98E-09 | -0.45 | 9.63E-03 |
| cg13092958 | 5.99 | 2.08E-09 | 0.49 | 4.49E-03 |
| cg18964706 | 5.98 | 2.17E-09 | 0.73 | 1.55E-05 |
| cg03147503 | 5.94 | 2.87E-09 | 0.51 | 1.59E-03 |

| | | | | |
|---|---|---|---|---|
| cg14708905 | 5.90 | 3.67E-09 | 0.66 | 1.94E-05 |
| cg00433866 | 5.84 | 5.20E-09 | 1.46 | 4.92E-20 |
| cg24055029 | -5.83 | 5.69E-09 | -0.53 | 2.29E-03 |
| cg22961241 | 5.77 | 7.71E-09 | 0.45 | 0.01 |
| cg15063443 | 5.76 | 8.40E-09 | 0.28 | 0.02 |
| cg17329164 | -5.74 | 9.33E-09 | -0.58 | 3.81E-05 |
| cg02772121 | -5.74 | 9.63E-09 | -0.68 | 3.34E-07 |
| cg24444631 | 5.70 | 1.20E-08 | 0.59 | 1.40E-04 |
| cg02956255 | -5.61 | 2.00E-08 | -0.79 | 4.80E-07 |
| cg08862148 | -5.55 | 2.87E-08 | -0.31 | 0.02 |
| cg03921216 | -5.54 | 3.00E-08 | -0.41 | 3.59E-03 |
| cg24446242 | 5.54 | 3.09E-08 | 0.47 | 2.58E-03 |
| cg06051311 | -5.52 | 3.47E-08 | -0.46 | 1.59E-03 |
| cg14661811 | -5.46 | 4.77E-08 | -0.34 | 0.04 |
| cg02053920 | -5.45 | 5.08E-08 | -0.50 | 3.36E-03 |
| cg00552704 | -5.44 | 5.28E-08 | -0.38 | 8.99E-03 |
| cg18060330 | 5.43 | 5.63E-08 | 0.43 | 0.01 |
| cg03081173 | -5.37 | 7.88E-08 | -0.42 | 5.88E-03 |
| cg09085842 | -5.30 | 1.18E-07 | -0.59 | 1.34E-03 |
| cg26266427 | -5.29 | 1.19E-07 | -0.86 | 1.33E-07 |
| cg20644330 | 5.29 | 1.26E-07 | 0.63 | 3.43E-04 |
| cg01992382 | -5.22 | 1.76E-07 | -0.94 | 5.17E-08 |
| cg01084722 | 5.19 | 2.12E-07 | 0.42 | 8.66E-03 |
| cg13866093 | 5.19 | 2.15E-07 | 0.86 | 5.59E-07 |

TCGA, The Cancer Genome Atlas; FDR

[a] Differential DNA methylation analyses were performed using Illumina 450K BeadChip data of 839 lung cancer tumor (469 adenocarcinoma and 370 squamous cell carcinoma) and 74 adjacent normal tissue samples from TCGA-LUAD and TCGA-LUSC. Effect sizes and *P* values were estimated using linear mixed models, adjusting for age, sex, smoking status and pack-years, and top three DNA methylation PCs. TCGA-LUAD and TCGA-LUSC data were analyzed sepaprately and results were combined by fixed effects meta-analyses.

**Supplementary Table 12. Psostate cancer susceptibility CpGs showing consistent differential DNA methylation between prostate cancer tumor and adjacent normal tissues in TCGA**

| CpG | CpG-cancer associations | | Differential methylation (TCGA) | |
|---|---|---|---|---|
| | Z score | *P* | Effect size [a] | *P* [a] |
| cg18825076 | -19.69 | 2.40E-86 | -0.50 | 1.05E-03 |
| cg19993258 | -13.60 | 3.94E-42 | -0.31 | 0.02 |
| cg11624060 | -11.54 | 8.48E-31 | -0.31 | 0.03 |
| cg10441424 | -11.38 | 5.26E-30 | -0.36 | 0.03 |
| cg17108064 | -8.44 | 3.28E-17 | -0.62 | 4.90E-04 |
| cg06180152 | -8.10 | 5.72E-16 | -0.48 | 4.44E-03 |
| cg04176169 | 8.09 | 6.06E-16 | 0.23 | 0.03 |
| cg22498099 | -7.94 | 1.95E-15 | -0.51 | 3.07E-03 |
| cg04884612 | 7.36 | 1.89E-13 | 0.55 | 1.87E-03 |
| cg13126982 | -7.30 | 2.86E-13 | -0.53 | 5.93E-03 |
| cg25721950 | 7.13 | 1.03E-12 | 0.72 | 1.13E-04 |
| cg12463578 | -7.11 | 1.18E-12 | -0.47 | 1.03E-03 |
| cg03782043 | -7.06 | 1.70E-12 | -0.56 | 6.26E-04 |
| cg13537510 | 6.81 | 9.95E-12 | 0.46 | 8.11E-03 |
| cg08325845 | 6.75 | 1.47E-11 | 0.23 | 8.92E-03 |
| cg01554625 | -6.63 | 3.39E-11 | -0.56 | 1.37E-03 |
| cg11485823 | 6.62 | 3.69E-11 | 0.51 | 2.83E-04 |
| cg05053818 | -6.58 | 4.70E-11 | -0.79 | 7.57E-06 |
| cg21519219 | 6.53 | 6.38E-11 | 0.57 | 1.05E-04 |
| cg05680196 | 6.50 | 8.14E-11 | 0.49 | 9.17E-03 |
| cg05353884 | -6.50 | 8.28E-11 | -0.29 | 9.17E-03 |
| cg03513762 | 6.49 | 8.55E-11 | 0.38 | 0.01 |
| cg15026241 | 6.48 | 9.13E-11 | 0.56 | 4.66E-04 |
| cg00903577 | -6.40 | 1.53E-10 | -0.68 | 8.79E-06 |
| cg05868516 | -6.36 | 1.96E-10 | -0.53 | 4.82E-03 |
| cg25318809 | 6.33 | 2.52E-10 | 0.25 | 4.43E-03 |
| cg01692379 | -6.31 | 2.80E-10 | -0.50 | 3.23E-03 |
| cg03689048 | -6.26 | 3.94E-10 | -0.93 | 8.18E-08 |
| cg11811828 | -6.23 | 4.81E-10 | -0.29 | 0.04 |
| cg26747317 | -6.21 | 5.22E-10 | -0.44 | 9.64E-04 |
| cg10432093 | -6.17 | 6.95E-10 | -0.64 | 3.94E-05 |
| cg19141489 | 6.10 | 1.08E-09 | 0.39 | 7.80E-03 |
| cg13688808 | 6.09 | 1.14E-09 | 0.47 | 2.84E-03 |
| cg14432143 | 6.08 | 1.21E-09 | 0.37 | 7.49E-04 |
| cg21100518 | -6.06 | 1.35E-09 | -0.37 | 4.17E-04 |
| cg26668675 | -6.05 | 1.41E-09 | -0.36 | 5.22E-03 |
| cg16751781 | -6.05 | 1.44E-09 | -0.95 | 9.87E-07 |
| cg05875819 | -6.00 | 1.98E-09 | -0.48 | 1.69E-03 |
| cg22291762 | -5.99 | 2.06E-09 | -0.32 | 0.02 |
| cg01738650 | 5.98 | 2.30E-09 | 0.26 | 0.01 |
| cg15726260 | 5.94 | 2.80E-09 | 0.26 | 6.52E-03 |
| cg16055914 | 5.90 | 3.62E-09 | 0.64 | 1.23E-04 |
| cg14708905 | 5.90 | 3.67E-09 | 0.26 | 0.03 |

| | | | | |
|---|---|---|---|---|
| cg00599564 | -5.89 | 3.88E-09 | -0.40 | 2.89E-04 |
| cg03293507 | -5.87 | 4.25E-09 | -0.41 | 0.02 |
| cg03078486 | -5.86 | 4.52E-09 | -0.34 | 6.40E-03 |
| cg22701603 | -5.86 | 4.54E-09 | -0.31 | 0.02 |
| cg02405128 | 5.86 | 4.70E-09 | 0.41 | 0.02 |
| cg13918754 | -5.85 | 5.02E-09 | -0.34 | 0.03 |
| cg17931227 | -5.82 | 6.03E-09 | -0.32 | 0.02 |
| cg22961241 | 5.77 | 7.71E-09 | 0.39 | 0.02 |
| cg15063443 | 5.76 | 8.40E-09 | 0.54 | 2.44E-04 |
| cg20914572 | -5.75 | 9.02E-09 | -0.30 | 0.04 |
| cg02772121 | -5.74 | 9.63E-09 | -0.39 | 0.03 |
| cg09179646 | -5.72 | 1.09E-08 | -0.28 | 0.03 |
| cg23252259 | -5.65 | 1.58E-08 | -0.36 | 0.01 |
| cg24446242 | 5.54 | 3.09E-08 | 0.30 | 0.01 |
| cg12247101 | -5.53 | 3.24E-08 | -0.41 | 6.57E-03 |
| cg26531141 | -5.52 | 3.33E-08 | -0.43 | 0.01 |
| cg06051311 | -5.52 | 3.47E-08 | -0.41 | 0.01 |
| cg24813995 | -5.48 | 4.17E-08 | -0.27 | 0.02 |
| cg20406979 | -5.48 | 4.36E-08 | -0.54 | 1.68E-03 |
| cg14661811 | -5.46 | 4.77E-08 | -0.38 | 3.70E-03 |
| cg24707219 | -5.44 | 5.40E-08 | -0.29 | 5.38E-03 |
| cg19448822 | 5.41 | 6.34E-08 | 0.23 | 0.02 |
| cg06308109 | 5.39 | 6.93E-08 | 0.54 | 4.73E-04 |
| cg03081173 | -5.37 | 7.88E-08 | -0.40 | 0.02 |
| cg17038049 | 5.37 | 7.99E-08 | 0.58 | 3.93E-05 |
| cg00474080 | 5.37 | 8.07E-08 | 0.36 | 0.01 |
| cg18416871 | 5.35 | 8.63E-08 | 0.27 | 0.03 |
| cg09093485 | -5.34 | 9.05E-08 | -0.33 | 0.04 |
| cg27185298 | 5.33 | 9.62E-08 | 0.34 | 0.01 |
| cg09085842 | -5.30 | 1.18E-07 | -0.53 | 4.03E-03 |
| cg08097157 | 5.24 | 1.57E-07 | 0.40 | 0.01 |
| cg14781281 | 5.22 | 1.82E-07 | 0.26 | 7.63E-03 |
| cg23244913 | -5.19 | 2.07E-07 | -0.23 | 0.03 |
| cg01084722 | 5.19 | 2.12E-07 | 0.33 | 0.04 |
| cg08163199 | 5.17 | 2.39E-07 | 0.23 | 7.17E-03 |
| cg04903089 | 5.16 | 2.43E-07 | 0.39 | 9.69E-03 |

TCGA, The Cancer Genome Atlas.

[a] Differential DNA methylation analyses were performed using Illumina 450K BeadChip data of 502 prostate adenocarcinoma and 50 adjacent normal tissue samples from TCGA-PRAD. Effect sizes and $P$ values were estimated using linear mixed models, adjusting for age and top three DNA methylation PCs.

**Supplementary Table 13. Consistent directions of associations across DNA methylation, gene expression, and breast cancer risk**

| CpG | Chr | Position (HG19) | Gene | Distance (Kb) | Cytoband | CpG-cancer | | CpG-gene | | Gene-cancer | |
|---|---|---|---|---|---|---|---|---|---|---|---|
| | | | | | | Dir | *P* [a] | Dir | *P* [b] | Dir | *P* [c] |
| cg04416237 | 1 | 10,051,507 | *GPR157* | 862.3 | 1p36.22 | + | 1.23E-07 | + | 1.06E-03 | + | 1.32E-29 |
| cg23231268 | 3 | 46,792,462 | *CCR9* | 847.8 | 3p21.31 | + | 5.70E-08 | - | 1.15E-04 | - | 7.36E-08 |
| cg18035979 | 5 | 81,575,199 | *ATP6AP1L* | -0.1 | 5q14.2 | + | 1.22E-07 | - | 3.05E-05 | - | **2.59E-09** |
| cg16465129 | 5 | 131,408,149 | *C5orf56* | -338.2 | 5q31.1 | + | 2.95E-07 | - | 2.54E-04 | - | 2.72E-93 |
| cg20773800 | 7 | 99,934,006 | *PILRA* | -37.1 | 7q22.1 | + | 2.28E-07 | + | 5.62E-05 | + | 1.22E-03 |
| cg20773800 | 7 | 99,934,006 | *PILRB* | -15.8 | 7q22.1 | + | 2.28E-07 | + | 7.26E-04 | + | **2.87E-07** |
| cg24531009 | 7 | 99,987,853 | *PILRB* | 22.5 | 7q22.1 | + | 2.18E-07 | + | 1.24E-04 | + | **2.87E-07** |
| cg24531009 | 7 | 99,987,853 | *PILRA* | Body | 7q22.1 | + | 2.18E-07 | + | 1.32E-04 | + | 1.22E-03 |
| cg14587961 | 7 | 99,991,523 | *PILRA* | Body | 7q22.1 | + | 2.12E-07 | + | 3.70E-05 | + | 1.22E-03 |
| cg13153964 | 7 | 100,026,424 | *PILRA* | 28.7 | 7q22.1 | + | 2.83E-07 | + | 1.20E-03 | + | 1.22E-03 |
| cg13153964 | 7 | 100,026,424 | *PILRB* | 61.1 | 7q22.1 | + | 2.83E-07 | + | 1.62E-03 | + | **2.87E-07** |
| cg21679810 | 7 | 100,027,352 | *PILRA* | 29.6 | 7q22.1 | + | 2.48E-07 | + | 4.90E-04 | + | 1.22E-03 |
| cg21679810 | 7 | 100,027,352 | *PILRB* | 62.0 | 7q22.1 | + | 2.48E-07 | + | 5.68E-04 | + | **2.87E-07** |
| cg07160278 | 7 | 100,027,354 | *PILRA* | 29.6 | 7q22.1 | + | 2.63E-07 | + | 3.19E-04 | + | 1.22E-03 |
| cg02327530 | 7 | 100,091,786 | *ZKSCAN1* | 452.5 | 7q22.1 | + | 2.94E-07 | + | 3.88E-04 | + | 1.65E-10 |
| cg07546779 | 8 | 29,495,175 | *LEPROTL1* | -457.7 | 8p12 | - | 1.46E-11 | - | 6.25E-04 | + | 4.97E-45 |
| cg22872885 | 10 | 80,615,772 | *ZMIZ1* | -213.0 | 10q22.3 | - | 4.61E-12 | - | 5.81E-04 | + | 3.63E-07 |
| cg01322412 | 11 | 781,977 | *AP006621.5* | Body | 11p15.5 | + | 1.40E-10 | + | 6.50E-04 | + | **1.09E-04** |
| cg01741372 | 11 | 783,889 | *AP006621.5* | Body | 11p15.5 | + | 3.91E-11 | + | 3.94E-04 | + | **1.09E-04** |
| cg02200722 | 17 | 43,489,199 | *DND1P1* | -174.0 | 17q21.31 | + | 4.36E-10 | - | 2.96E-05 | - | 1.09E-10 |
| cg02200722 | 17 | 43,489,199 | *RP11-259G18.3* | -847.7 | 17q21.31 | + | 4.36E-10 | - | 4.71E-05 | - | 6.09E-08 |
| cg02200722 | 17 | 43,489,199 | *RP11-707O23.1* | -180.8 | 17q21.31 | + | 4.36E-10 | - | 2.26E-04 | - | 3.27E-30 |
| cg02200722 | 17 | 43,489,199 | *KANSL1-AS1* | -781.7 | 17q21.31 | + | 4.36E-10 | - | 3.14E-04 | - | 3.84E-20 |
| cg02200722 | 17 | 43,489,199 | *LINC02210* | -208.5 | 17q21.31 | + | 4.36E-10 | - | 1.79E-03 | - | 1.30E-08 |
| cg01718965 | 17 | 43,516,991 | *DND1P1* | -146.3 | 17q21.31 | + | 7.58E-10 | - | 2.28E-05 | - | 1.09E-10 |
| cg01718965 | 17 | 43,516,991 | *KANSL1-AS1* | -754.0 | 17q21.31 | + | 7.58E-10 | - | 5.97E-05 | - | 3.84E-20 |
| cg01718965 | 17 | 43,516,991 | *LINC02210* | -180.7 | 17q21.31 | + | 7.58E-10 | - | 3.30E-04 | - | 1.30E-08 |
| cg01718965 | 17 | 43,516,991 | *RP11-259G18.3* | -819.9 | 17q21.31 | + | 7.58E-10 | - | 2.13E-03 | - | 6.09E-08 |
| cg00916973 | 17 | 43,580,461 | *KANSL1-AS1* | -690.5 | 17q21.31 | + | 3.77E-11 | - | 2.63E-05 | - | 3.84E-20 |
| cg00916973 | 17 | 43,580,461 | *DND1P1* | -82.8 | 17q21.31 | + | 3.77E-11 | - | 1.28E-04 | - | 1.09E-10 |
| cg00916973 | 17 | 43,580,461 | *LINC02210* | -117.2 | 17q21.31 | + | 3.77E-11 | - | 3.10E-04 | - | 1.30E-08 |
| cg00916973 | 17 | 43,580,461 | *RP11-259G18.3* | -756.5 | 17q21.31 | + | 3.77E-11 | - | 2.06E-03 | - | 6.09E-08 |
| cg22433210 | 17 | 43,662,623 | *RP11-259G18.3* | -674.3 | 17q21.31 | - | 3.44E-12 | + | 7.50E-05 | - | 6.09E-08 |
| cg22433210 | 17 | 43,662,623 | *LRRC37A* | -707.5 | 17q21.31 | - | 3.44E-12 | + | 1.96E-04 | - | 1.31E-25 |
| cg22433210 | 17 | 43,662,623 | *DND1P1* | -0.6 | 17q21.31 | - | 3.44E-12 | + | 2.16E-04 | - | 1.09E-10 |
| cg22433210 | 17 | 43,662,623 | *KANSL1-AS1* | -608.3 | 17q21.31 | - | 3.44E-12 | + | 3.56E-04 | - | 3.84E-20 |
| cg22433210 | 17 | 43,662,623 | *LINC02210* | -35.1 | 17q21.31 | - | 3.44E-12 | + | 4.45E-04 | - | 1.30E-08 |
| cg22433210 | 17 | 43,662,623 | *LRRC37A2* | -926.3 | 17q21.31 | - | 3.44E-12 | + | 4.31E-03 | - | 3.51E-17 |
| cg22433210 | 17 | 43,662,623 | *FAM215B* | -973.6 | 17q21.31 | - | 3.44E-12 | + | 7.94E-03 | - | 6.11E-17 |
| cg01341218 | 17 | 43,662,625 | *LINC02210* | -35.1 | 17q21.31 | - | 2.79E-12 | + | 2.27E-05 | - | 1.30E-08 |
| cg01341218 | 17 | 43,662,625 | *RP11-259G18.3* | -674.3 | 17q21.31 | - | 2.79E-12 | + | 1.03E-04 | - | 6.09E-08 |
| cg01341218 | 17 | 43,662,625 | *KANSL1-AS1* | -608.3 | 17q21.31 | - | 2.79E-12 | + | 5.34E-04 | - | 3.84E-20 |
| cg01341218 | 17 | 43,662,625 | *DND1P1* | -0.6 | 17q21.31 | - | 2.79E-12 | + | 6.46E-04 | - | 1.09E-10 |
| cg01341218 | 17 | 43,662,625 | *LRRC37A* | -707.5 | 17q21.31 | - | 2.79E-12 | + | 2.21E-03 | - | 1.31E-25 |
| cg01341218 | 17 | 43,662,625 | *LRRC37A2* | -926.3 | 17q21.31 | - | 2.79E-12 | + | 6.99E-03 | - | 3.51E-17 |
| cg20393308 | 17 | 43,728,044 | *RP11-259G18.3* | -608.9 | 17q21.31 | - | 6.50E-09 | + | 1.05E-04 | - | 6.09E-08 |
| cg20393308 | 17 | 43,728,044 | *DND1P1* | 63.8 | 17q21.31 | - | 6.50E-09 | + | 1.22E-04 | - | 1.09E-10 |
| cg20393308 | 17 | 43,728,044 | *RP11-707O23.1* | 57.3 | 17q21.31 | - | 6.50E-09 | + | 3.86E-04 | - | 3.27E-30 |
| cg20393308 | 17 | 43,728,044 | *LINC02210* | Body | 17q21.31 | - | 6.50E-09 | + | 4.97E-04 | - | 1.30E-08 |
| cg20393308 | 17 | 43,728,044 | *KANSL1-AS1* | -542.9 | 17q21.31 | - | 6.50E-09 | + | 1.65E-03 | - | 3.84E-20 |
| cg03238273 | 17 | 43,825,672 | *KANSL1-AS1* | -445.3 | 17q21.31 | - | 5.58E-12 | + | 3.49E-06 | - | 3.84E-20 |
| cg03238273 | 17 | 43,825,672 | *LINC02210* | 93.2 | 17q21.31 | - | 5.58E-12 | + | 4.58E-05 | - | 1.30E-08 |
| cg03238273 | 17 | 43,825,672 | *DND1P1* | 161.4 | 17q21.31 | - | 5.58E-12 | + | 7.65E-05 | - | 1.09E-10 |
| cg03238273 | 17 | 43,825,672 | *RP11-259G18.3* | -511.3 | 17q21.31 | - | 5.58E-12 | + | 9.81E-05 | - | 6.09E-08 |
| cg03238273 | 17 | 43,825,672 | *LRRC37A2* | -763.2 | 17q21.31 | - | 5.58E-12 | + | 6.58E-03 | - | 3.51E-17 |
| cg23202277 | 17 | 43,971,911 | *RP11-259G18.3* | -365.0 | 17q21.31 | - | 4.05E-12 | + | 2.42E-06 | - | 6.09E-08 |
| cg23202277 | 17 | 43,971,911 | *LINC02210* | 239.4 | 17q21.31 | - | 4.05E-12 | + | 2.83E-05 | - | 1.30E-08 |
| cg23202277 | 17 | 43,971,911 | *LRRC37A* | -398.2 | 17q21.31 | - | 4.05E-12 | + | 5.26E-05 | - | 1.31E-25 |
| cg23202277 | 17 | 43,971,911 | *DND1P1* | 307.6 | 17q21.31 | - | 4.05E-12 | + | 5.71E-05 | - | 1.09E-10 |
| cg23202277 | 17 | 43,971,911 | *KANSL1-AS1* | -299.0 | 17q21.31 | - | 4.05E-12 | + | 2.48E-04 | - | 3.84E-20 |
| cg23202277 | 17 | 43,971,911 | *LRRC37A2* | -617.0 | 17q21.31 | - | 4.05E-12 | + | 1.62E-03 | - | 3.51E-17 |
| cg23202277 | 17 | 43,971,911 | *RP11-707O23.1* | 301.2 | 17q21.31 | - | 4.05E-12 | + | 3.36E-03 | - | 3.27E-30 |
| cg23202277 | 17 | 43,971,911 | *FAM215B* | -664.3 | 17q21.31 | - | 4.05E-12 | + | 9.39E-03 | - | 6.11E-17 |
| cg20099416 | 17 | 43,971,919 | *RP11-259G18.3* | -365.0 | 17q21.31 | - | 3.88E-12 | + | 8.61E-06 | - | 6.09E-08 |
| cg20099416 | 17 | 43,971,919 | *KANSL1-AS1* | -299.0 | 17q21.31 | - | 3.88E-12 | + | 4.27E-05 | - | 3.84E-20 |
| cg20099416 | 17 | 43,971,919 | *DND1P1* | 307.6 | 17q21.31 | - | 3.88E-12 | + | 9.43E-05 | - | 1.09E-10 |
| cg20099416 | 17 | 43,971,919 | *LINC02210* | 239.4 | 17q21.31 | - | 3.88E-12 | + | 4.16E-04 | - | 1.30E-08 |
| cg20099416 | 17 | 43,971,919 | *LRRC37A* | -398.2 | 17q21.31 | - | 3.88E-12 | + | 4.41E-04 | - | 1.31E-25 |
| cg20099416 | 17 | 43,971,919 | *LRRC37A2* | -617.0 | 17q21.31 | - | 3.88E-12 | + | 6.38E-04 | - | 3.51E-17 |
| cg20099416 | 17 | 43,971,919 | *FAM215B* | -664.3 | 17q21.31 | - | 3.88E-12 | + | 5.60E-03 | - | 6.11E-17 |
| cg20099416 | 17 | 43,971,919 | *RP11-707O23.1* | 301.2 | 17q21.31 | - | 3.88E-12 | + | 6.14E-03 | - | 3.27E-30 |

| | | | | | | | | | | | |
|---|---|---|---|---|---|---|---|---|---|---|---|
| cg11489262 | 17 | 43,973,426 | *LINC02210* | 240.9 | 17q21.31 | - | 3.02E-12 | + | 1.09E-03 | - | 1.30E-08 |
| cg11489262 | 17 | 43,973,426 | *RP11-259G18.3* | -363.5 | 17q21.31 | - | 3.02E-12 | + | 1.19E-03 | - | 6.09E-08 |
| cg11489262 | 17 | 43,973,426 | *DND1P1* | 309.1 | 17q21.31 | - | 3.02E-12 | + | 1.44E-03 | - | 1.09E-10 |
| cg11489262 | 17 | 43,973,426 | *KANSL1-AS1* | -297.5 | 17q21.31 | - | 3.02E-12 | + | 1.89E-03 | - | 3.84E-20 |
| cg10224600 | 17 | 43,975,063 | *LINC02210* | 242.5 | 17q21.31 | - | 2.38E-10 | + | 1.76E-03 | - | 1.30E-08 |
| cg10224600 | 17 | 43,975,063 | *RP11-259G18.3* | -361.9 | 17q21.31 | - | 2.38E-10 | + | 2.81E-03 | - | 6.09E-08 |
| cg10224600 | 17 | 43,975,063 | *DND1P1* | 310.8 | 17q21.31 | - | 2.38E-10 | + | 4.50E-03 | - | 1.09E-10 |
| cg21599490 | 17 | 43,998,761 | *LINC02210* | 266.2 | 17q21.31 | - | 1.13E-08 | + | 7.42E-04 | - | 1.30E-08 |
| cg21599490 | 17 | 43,998,761 | *DND1P1* | 334.5 | 17q21.31 | - | 1.13E-08 | + | 4.65E-03 | - | 1.09E-10 |
| cg23926235 | 17 | 44,024,256 | *KANSL1-AS1* | -246.7 | 17q21.31 | + | 3.72E-11 | - | 9.79E-04 | - | 3.84E-20 |
| cg23926235 | 17 | 44,024,256 | *DND1P1* | 360.0 | 17q21.31 | + | 3.72E-11 | - | 1.05E-03 | - | 1.09E-10 |
| cg23926235 | 17 | 44,024,256 | *LINC02210* | 291.7 | 17q21.31 | + | 3.72E-11 | - | 3.51E-03 | - | 1.30E-08 |
| cg23926235 | 17 | 44,024,256 | *RP11-259G18.3* | -312.7 | 17q21.31 | + | 3.72E-11 | - | 4.42E-03 | - | 6.09E-08 |
| cg18586914 | 17 | 44,040,821 | *KANSL1-AS1* | -230.1 | 17q21.31 | + | 8.77E-12 | - | 7.78E-07 | - | 3.84E-20 |
| cg18586914 | 17 | 44,040,821 | *DND1P1* | 376.5 | 17q21.31 | + | 8.77E-12 | - | 1.07E-05 | - | 1.09E-10 |
| cg18586914 | 17 | 44,040,821 | *LINC02210* | 308.3 | 17q21.31 | + | 8.77E-12 | - | 7.30E-05 | - | 1.30E-08 |
| cg18586914 | 17 | 44,040,821 | *RP11-259G18.3* | -296.1 | 17q21.31 | + | 8.77E-12 | - | 2.04E-04 | - | 6.09E-08 |
| cg18586914 | 17 | 44,040,821 | *LRRC37A2* | -548.1 | 17q21.31 | + | 8.77E-12 | - | 1.66E-03 | - | 3.51E-17 |
| cg18586914 | 17 | 44,040,821 | *RP11-259G18.1* | -303.6 | 17q21.31 | + | 8.77E-12 | - | 2.65E-03 | - | 0.05 |
| cg18586914 | 17 | 44,040,821 | *LRRC37A* | -329.3 | 17q21.31 | + | 8.77E-12 | - | 9.61E-03 | - | 1.31E-25 |
| cg01934064 | 17 | 44,064,242 | *LINC02210* | 331.7 | 17q21.31 | - | 2.99E-10 | + | 1.18E-03 | - | 1.30E-08 |
| cg05721485 | 17 | 44,071,124 | *DND1P1* | 406.8 | 17q21.31 | - | 3.25E-12 | + | 4.74E-04 | - | 1.09E-10 |
| cg05721485 | 17 | 44,071,124 | *KANSL1-AS1* | -199.8 | 17q21.31 | - | 3.25E-12 | + | 1.32E-03 | - | 3.84E-20 |
| cg05721485 | 17 | 44,071,124 | *RP11-259G18.3* | -265.8 | 17q21.31 | - | 3.25E-12 | + | 2.65E-03 | - | 6.09E-08 |
| cg05721485 | 17 | 44,071,124 | *CTB-39G8.3* | 596.3 | 17q21.31 | - | 3.25E-12 | + | 4.44E-03 | - | 2.15E-65 |
| cg08318660 | 17 | 44,122,580 | *LINC02210* | 390.1 | 17q21.31 | - | 6.37E-11 | + | 2.42E-06 | - | 1.30E-08 |
| cg08318660 | 17 | 44,122,580 | *DND1P1* | 458.3 | 17q21.31 | - | 6.37E-11 | + | 2.06E-04 | - | 1.09E-10 |
| cg08318660 | 17 | 44,122,580 | *KANSL1-AS1* | -148.4 | 17q21.31 | - | 6.37E-11 | + | 1.02E-03 | - | 3.84E-20 |
| cg08318660 | 17 | 44,122,580 | *RP11-259G18.3* | -214.3 | 17q21.31 | - | 6.37E-11 | + | 3.29E-03 | - | 6.09E-08 |
| cg08318660 | 17 | 44,122,580 | *LRRC37A2* | -466.3 | 17q21.31 | - | 6.37E-11 | + | 6.64E-03 | - | 3.51E-17 |
| cg08318660 | 17 | 44,122,580 | *LRRC37A* | -247.5 | 17q21.31 | - | 6.37E-11 | + | 7.92E-03 | - | 1.31E-25 |
| cg02478991 | 17 | 44,180,961 | *LINC02210* | 448.4 | 17q21.31 | + | 5.22E-11 | - | 2.01E-03 | - | 1.30E-08 |
| cg01906119 | 17 | 44,208,035 | *RP11-259G18.3* | -128.9 | 17q21.31 | + | 5.10E-11 | - | 3.22E-08 | - | 6.09E-08 |
| cg01906119 | 17 | 44,208,035 | *DND1P1* | 543.7 | 17q21.31 | + | 5.10E-11 | - | 1.52E-06 | - | 1.09E-10 |
| cg01906119 | 17 | 44,208,035 | *LINC02210* | 475.5 | 17q21.31 | + | 5.10E-11 | - | 1.84E-06 | - | 1.30E-08 |
| cg01906119 | 17 | 44,208,035 | *KANSL1-AS1* | -62.9 | 17q21.31 | + | 5.10E-11 | - | 2.27E-06 | - | 3.84E-20 |
| cg01906119 | 17 | 44,208,035 | *LRRC37A* | -162.1 | 17q21.31 | + | 5.10E-11 | - | 1.68E-03 | - | 1.31E-25 |
| cg01906119 | 17 | 44,208,035 | *LRRC37A2* | -380.8 | 17q21.31 | + | 5.10E-11 | - | 1.84E-03 | - | 3.51E-17 |
| cg01906119 | 17 | 44,208,035 | *RP11-707O23.1* | 537.3 | 17q21.31 | + | 5.10E-11 | - | 3.05E-03 | - | 3.27E-30 |
| cg01906119 | 17 | 44,208,035 | *RP11-259G18.1* | -136.4 | 17q21.31 | + | 5.10E-11 | - | 3.89E-03 | - | 0.05 |
| cg01906119 | 17 | 44,208,035 | *RP11-156P1.3* | -853.4 | 17q21.31 | + | 5.10E-11 | - | 8.35E-03 | - | 2.05E-07 |
| cg19064792 | 17 | 44,257,101 | *LRRC37A2* | -331.8 | 17q21.31 | + | 2.80E-10 | - | 1.16E-06 | - | 3.51E-17 |
| cg19064792 | 17 | 44,257,101 | *ARL17A* | -337.0 | 17q21.31 | + | 2.80E-10 | - | 1.04E-05 | - | 2.80E-21 |
| cg19064792 | 17 | 44,257,101 | *KANSL1-AS1* | -13.8 | 17q21.31 | + | 2.80E-10 | - | 1.14E-05 | - | 3.84E-20 |
| cg19064792 | 17 | 44,257,101 | *LRRC37A* | -113.0 | 17q21.31 | + | 2.80E-10 | - | 1.36E-04 | - | 1.31E-25 |
| cg19064792 | 17 | 44,257,101 | *RP11-259G18.3* | -79.8 | 17q21.31 | + | 2.80E-10 | - | 2.68E-04 | - | 6.09E-08 |
| cg19064792 | 17 | 44,257,101 | *DND1P1* | 592.8 | 17q21.31 | + | 2.80E-10 | - | 3.51E-04 | - | 1.09E-10 |
| cg19064792 | 17 | 44,257,101 | *LINC02210* | 524.6 | 17q21.31 | + | 2.80E-10 | - | 5.41E-04 | - | 1.30E-08 |
| cg19064792 | 17 | 44,257,101 | *CTB-39G8.3* | 782.3 | 17q21.31 | + | 2.80E-10 | - | 3.30E-03 | - | 2.15E-65 |
| cg19064792 | 17 | 44,257,101 | *RP11-259G18.1* | -87.3 | 17q21.31 | + | 2.80E-10 | - | 6.59E-03 | - | 0.05 |
| cg19064792 | 17 | 44,257,101 | *RP11-798G7.8* | 645.9 | 17q21.31 | + | 2.80E-10 | - | 8.18E-03 | - | 1.24E-10 |
| cg19064792 | 17 | 44,257,101 | *RP11-798G7.6* | 616.5 | 17q21.31 | + | 2.80E-10 | - | 0.01 | - | 6.02E-14 |
| cg02322039 | 17 | 44,269,535 | *LRRC37A2* | -319.3 | 17q21.31 | + | 1.27E-10 | - | 3.08E-06 | - | 3.51E-17 |
| cg02322039 | 17 | 44,269,535 | *ARL17A* | -324.5 | 17q21.31 | + | 1.27E-10 | - | 8.98E-06 | - | 2.80E-21 |
| cg02322039 | 17 | 44,269,535 | *KANSL1-AS1* | -1.4 | 17q21.31 | + | 1.27E-10 | - | 1.01E-04 | - | 3.84E-20 |
| cg02322039 | 17 | 44,269,535 | *RP11-798G7.6* | 628.9 | 17q21.31 | + | 1.27E-10 | - | 2.60E-04 | - | 6.02E-14 |
| cg02322039 | 17 | 44,269,535 | *RP11-798G7.8* | 658.3 | 17q21.31 | + | 1.27E-10 | - | 5.99E-04 | - | 1.24E-10 |
| cg02322039 | 17 | 44,269,535 | *KANSL1* | Body | 17q21.31 | + | 1.27E-10 | - | 1.66E-03 | - | 3.83E-19 |
| cg02322039 | 17 | 44,269,535 | *RP11-259G18.1* | -74.9 | 17q21.31 | + | 1.27E-10 | - | 1.70E-03 | - | 0.05 |
| cg02322039 | 17 | 44,269,535 | *RP11-259G18.3* | -67.4 | 17q21.31 | + | 1.27E-10 | - | 9.24E-03 | - | 6.09E-08 |
| cg02322039 | 17 | 44,269,535 | *DND1P1* | 605.2 | 17q21.31 | + | 1.27E-10 | - | 9.46E-03 | - | 1.09E-10 |
| cg02322039 | 17 | 44,269,535 | *LRRC37A* | -100.6 | 17q21.31 | + | 1.27E-10 | - | 9.56E-03 | - | 1.31E-25 |
| cg24992007 | 17 | 44,270,238 | *CTB-39G8.3* | 795.4 | 17q21.31 | + | 1.51E-09 | - | 8.76E-04 | - | 2.15E-65 |
| cg24992007 | 17 | 44,270,238 | *RP11-798G7.8* | 659.0 | 17q21.31 | + | 1.51E-09 | - | 1.98E-03 | - | 1.24E-10 |
| cg24992007 | 17 | 44,270,238 | *KANSL1-AS1* | -0.7 | 17q21.31 | + | 1.51E-09 | - | 2.61E-03 | - | 3.84E-20 |
| cg16132033 | 17 | 44,292,685 | *ARL17A* | -301.4 | 17q21.31 | + | 1.49E-11 | - | 4.57E-05 | - | 2.80E-21 |
| cg16132033 | 17 | 44,292,685 | *LRRC37A2* | -296.2 | 17q21.31 | + | 1.49E-11 | - | 9.93E-05 | - | 3.51E-17 |
| cg16132033 | 17 | 44,292,685 | *KANSL1-AS1* | 18.6 | 17q21.31 | + | 1.49E-11 | - | 5.23E-04 | - | 3.84E-20 |
| cg16132033 | 17 | 44,292,685 | *RP11-798G7.8* | 681.5 | 17q21.31 | + | 1.49E-11 | - | 9.33E-04 | - | 1.24E-10 |
| cg16132033 | 17 | 44,292,685 | *FAM215B* | -343.5 | 17q21.31 | + | 1.49E-11 | - | 3.47E-03 | - | 6.11E-17 |
| cg16132033 | 17 | 44,292,685 | *CTB-39G8.3* | 817.8 | 17q21.31 | + | 1.49E-11 | - | 3.78E-03 | - | 2.15E-65 |
| cg16132033 | 17 | 44,292,685 | *DND1P1* | 628.4 | 17q21.31 | + | 1.49E-11 | - | 5.03E-03 | - | 1.09E-10 |
| cg16132033 | 17 | 44,292,685 | *RP11-259G18.3* | -44.2 | 17q21.31 | + | 1.49E-11 | - | 5.10E-03 | - | 6.09E-08 |
| cg16132033 | 17 | 44,292,685 | *LRRC37A* | -77.4 | 17q21.31 | + | 1.49E-11 | - | 7.85E-03 | - | 1.31E-25 |

| | | | | | | | | | | | |
|---|---|---|---|---|---|---|---|---|---|---|---|
| cg16132033 | 17 | 44,292,685 | *LINC02210* | 560.2 | 17q21.31 | + | 1.49E-11 | - | 0.01 | - | 1.30E-08 |
| cg16132033 | 17 | 44,292,685 | *ARHGAP27* | 780.9 | 17q21.31 | + | 1.49E-11 | - | 0.01 | - | 5.64E-43 |
| cg16132033 | 17 | 44,292,685 | *RP11-798G7.6* | 652.1 | 17q21.31 | + | 1.49E-11 | - | 0.01 | - | 6.02E-14 |
| cg17911788 | 17 | 44,343,683 | *RP11-259G18.3* | 5.7 | 17q21.31 | + | 1.23E-09 | - | 3.25E-04 | - | 6.09E-08 |
| cg17911788 | 17 | 44,343,683 | *DND1P1* | 679.4 | 17q21.31 | + | 1.23E-09 | - | 6.78E-04 | - | 1.09E-10 |
| cg17911788 | 17 | 44,343,683 | *LINC02210* | 611.2 | 17q21.31 | + | 1.23E-09 | - | 9.73E-04 | - | 1.30E-08 |
| cg17911788 | 17 | 44,343,683 | *RP11-707O23.1* | 673.0 | 17q21.31 | + | 1.23E-09 | - | 1.72E-03 | - | 3.27E-30 |
| cg17911788 | 17 | 44,343,683 | *KANSL1-AS1* | 69.6 | 17q21.31 | + | 1.23E-09 | - | 5.81E-03 | - | 3.84E-20 |
| cg05159804 | 17 | 44,343,776 | *RP11-259G18.3* | 5.8 | 17q21.31 | + | 5.98E-12 | - | 7.14E-07 | - | 6.09E-08 |
| cg05159804 | 17 | 44,343,776 | *LINC02210* | 611.3 | 17q21.31 | + | 5.98E-12 | - | 1.62E-06 | - | 1.30E-08 |
| cg05159804 | 17 | 44,343,776 | *DND1P1* | 679.5 | 17q21.31 | + | 5.98E-12 | - | 4.57E-05 | - | 1.09E-10 |
| cg05159804 | 17 | 44,343,776 | *RP11-707O23.1* | 673.0 | 17q21.31 | + | 5.98E-12 | - | 3.33E-04 | - | 3.27E-30 |
| cg05159804 | 17 | 44,343,776 | *KANSL1-AS1* | 69.7 | 17q21.31 | + | 5.98E-12 | - | 3.37E-04 | - | 3.84E-20 |
| cg05159804 | 17 | 44,343,776 | *RP5-867C24.1* | -892.8 | 17q21.31 | + | 5.98E-12 | - | 9.42E-03 | - | 6.89E-67 |
| cg27342294 | 20 | 34,570,157 | *MMP24* | 705.9 | 20q11.23 | + | 3.55E-08 | - | 1.38E-04 | - | 3.44E-10 |
| cg02100011 | 22 | 38,635,773 | *CARD10* | 720.2 | 22q13.1 | - | 1.97E-08 | + | 2.35E-05 | - | 2.83E-40 |
| cg02100011 | 22 | 38,635,773 | *MFNG* | 753.4 | 22q13.1 | - | 1.97E-08 | + | 1.17E-03 | - | 3.22E-86 |
| cg05951419 | 22 | 41,310,349 | *TNRC6B* | 578.5 | 22q13.2 | - | 5.32E-08 | - | 1.19E-03 | + | **1.62E-03** |

Chr, chromosome; Dir, direction of association/correlation.

[a] *P* values were calculated using SPrediXcan. All statistical tests were two-sided.

[b] *P* values were calculated using linear regression. All statistical tests were two-sided.

[c] *P* values were calculated using SPrediXcan and R *limma* package for data from GTEx and GEPIA2, respectively. For genes showing in both GTEx and GEPIA2 data, *P* values from GEPIA2 data are presented. *P* values from GTEx data are highlighted in bold. All statistical tests were two-sided.

**Supplementary Table 14. Consistent directions of associations across DNA methylation, gene expression, and colorectal cancer risk**

| CpG | Chr | Position (HG19) | Gene | Distance (Kb) | Cytoband | CpG-cancer | | CpG-gene | | Gene-cancer | |
|---|---|---|---|---|---|---|---|---|---|---|---|
| | | | | | | Dir | *P* [a] | Dir | *P* [b] | Dir | *P* [c] |
| cg12658694 | 1 | 38,397,304 | *RHBDL2* | -954.2 | 1p34.3 | - | 1.44E-15 | - | 1.72E-05 | + | 4.39E-57 |
| cg23805594 | 1 | 221,933,302 | *TAF1A* | -797.9 | 1q41 | - | 7.06E-08 | - | 1.76E-03 | + | 4.26E-14 |
| cg19445579 | 1 | 222,065,665 | *C1orf140* | 556.0 | 1q41 | + | 4.04E-10 | - | 1.45E-03 | - | 6.36E-85 |
| cg00012203 | 2 | 219,082,015 | *RP11-378A13.1* | -38.0 | 2q35 | + | 1.48E-11 | + | 2.72E-04 | + | **4.27E-19** |
| cg20019365 | 2 | 219,134,978 | *RP11-378A13.1* | 12.9 | 2q35 | + | 2.97E-16 | + | 7.41E-05 | + | **4.27E-19** |
| cg01329789 | 2 | 219,157,515 | *WNT6* | -567.0 | 2q35 | + | 1.85E-09 | - | 7.56E-04 | - | 2.19E-52 |
| cg02383163 | 2 | 219,157,675 | *CYP27A1* | -488.8 | 2q35 | + | 1.64E-15 | - | 7.66E-05 | - | 0.02 |
| cg18892128 | 6 | 31,409,575 | *SKIV2L* | -517.3 | 6p21.33 | + | 6.22E-10 | - | 4.05E-04 | - | 1.92E-11 |
| cg12988118 | 6 | 31,744,524 | *XXbac-BPG299F13.17* | 578.7 | 6p21.33 | + | 5.56E-09 | - | 1.26E-04 | - | 1.16E-148 |
| cg05133205 | 6 | 32,121,249 | *HLA-DPA1* | -911.1 | 6p21.32 | - | 4.42E-08 | - | 5.21E-04 | + | 1.02E-12 |
| cg05133205 | 6 | 32,121,249 | *HLA-DRB1* | -425.3 | 6p21.32 | - | 4.42E-08 | - | 4.67E-04 | + | 1.83E-13 |
| cg05133205 | 6 | 32,121,249 | *MICA* | 738.2 | 6p21.32 | - | 4.42E-08 | + | 4.42E-04 | - | 7.77E-38 |
| cg00440797 | 6 | 32,493,873 | *HLA-DRB5* | Body | 6p21.32 | - | 5.28E-11 | + | 2.33E-07 | - | **5.68E-03** |
| cg00440797 | 6 | 32,493,873 | *HLA-DQA1* | -102.1 | 6p21.32 | - | 5.28E-11 | + | 1.65E-03 | - | **1.88E-04** |
| cg00440797 | 6 | 32,493,873 | *HLA-DQB1-AS1* | -134.3 | 6p21.32 | - | 5.28E-11 | + | 1.25E-04 | - | 8.31E-29 |
| cg00440797 | 6 | 32,493,873 | *HLA-DRB6* | -26.6 | 6p21.32 | - | 5.28E-11 | - | 4.02E-12 | + | 7.19E-32 |
| cg00440797 | 6 | 32,493,873 | *HLA-DQA2* | -215.3 | 6p21.32 | - | 5.28E-11 | - | 1.43E-04 | + | 6.16E-33 |
| cg14645244 | 6 | 32,552,205 | *HLA-DRB5* | 54.1 | 6p21.32 | + | 1.94E-11 | - | 3.08E-05 | - | **5.68E-03** |
| cg14645244 | 6 | 32,552,205 | *HLA-DRB6* | 24.4 | 6p21.32 | + | 1.94E-11 | + | 1.10E-04 | + | 7.19E-32 |
| cg07007382 | 6 | 32,578,070 | *HLA-DRB5* | 80.0 | 6p21.32 | - | 1.96E-08 | + | 1.17E-08 | - | **5.68E-03** |
| cg07007382 | 6 | 32,578,070 | *HLA-DRB1* | 20.5 | 6p21.32 | - | 1.96E-08 | + | 2.77E-03 | - | **3.84E-03** |
| cg07007382 | 6 | 32,578,070 | *HLA-DQA1* | -17.9 | 6p21.32 | - | 1.96E-08 | + | 4.37E-05 | - | **1.88E-04** |
| cg07007382 | 6 | 32,578,070 | *HLA-DQB1-AS1* | -50.1 | 6p21.32 | - | 1.96E-08 | + | 3.62E-04 | - | 8.31E-29 |
| cg07007382 | 6 | 32,578,070 | *HLA-DRB6* | 50.3 | 6p21.32 | - | 1.96E-08 | - | 3.30E-04 | + | 7.19E-32 |
| cg07007382 | 6 | 32,578,070 | *HLA-DQA2* | -131.1 | 6p21.32 | - | 1.96E-08 | - | 1.37E-03 | + | 6.16E-33 |
| cg07007382 | 6 | 32,578,070 | *HLA-DQB2* | -145.8 | 6p21.32 | - | 1.96E-08 | - | 4.09E-03 | + | 4.15E-41 |
| cg22421503 | 6 | 36,623,153 | *CPNE5* | -85.4 | 6p21.2 | + | 2.68E-13 | - | 6.87E-05 | - | 4.31E-12 |
| cg02730678 | 6 | 36,641,554 | *RP1-179N16.3* | 581.1 | 6p21.2 | - | 3.34E-16 | + | 5.45E-04 | - | 1.09E-70 |
| cg12444411 | 7 | 2,802,554 | *GNA12* | Body | 7p22.2 | + | 3.94E-08 | - | 1.83E-03 | - | 1.02E-03 |
| cg19524238 | 7 | 2,802,976 | *GNA12* | Body | 7p22.2 | + | 2.10E-07 | - | 2.03E-03 | - | 1.02E-03 |
| cg16459265 | 7 | 45,025,080 | *RP4-647J21.1* | 24.6 | 7p13 | + | 2.53E-10 | - | 1.98E-04 | - | **8.72E-10** |
| cg12169661 | 7 | 73,183,394 | *WBSCR27* | -65.5 | 7q11.23 | - | 1.09E-07 | + | 1.31E-04 | - | **5.29E-05** |
| cg09559225 | 8 | 117,636,287 | *RAD21-AS1* | -250.5 | 8q23.3 | - | 7.14E-16 | - | 3.74E-03 | + | 2.49E-21 |
| cg08956474 | 10 | 101,284,950 | *SEC31B* | -961.5 | 10q24.2 | - | 2.14E-08 | - | 6.40E-03 | + | **0.02** |
| cg08956474 | 10 | 101,284,950 | *RP11-129J12.1* | -1.8 | 10q24.2 | - | 2.14E-08 | + | 2.68E-03 | - | 1.55E-23 |
| cg08956474 | 10 | 101,284,950 | *LINC01475* | -1.2 | 10q24.2 | - | 2.14E-08 | + | 5.32E-04 | - | 5.59E-31 |
| cg08956474 | 10 | 101,284,950 | *NKX2-3* | -7.7 | 10q24.2 | - | 2.14E-08 | + | 1.48E-04 | - | 5.87E-76 |
| cg12338214 | 10 | 133,783,113 | *LRRC27* | -362.6 | 10q26.3 | - | 1.01E-08 | + | 2.33E-03 | - | 1.59E-06 |
| cg03894242 | 12 | 57,513,121 | *RDH16* | 160.0 | 12q13.3 | + | 1.32E-09 | + | 5.05E-04 | + | 2.84E-17 |
| cg01493522 | 13 | 37,497,338 | *SMAD9* | 2.4 | 13q13.3 | - | 2.12E-16 | + | 1.43E-03 | - | 9.81E-201 |
| cg24467989 | 13 | 41,560,747 | *KBTBD7* | -203.2 | 13q14.11 | - | 1.35E-07 | - | 1.16E-03 | + | 7.82E-37 |
| cg20070323 | 15 | 72,520,350 | *SENP8* | 84.3 | 15q23 | - | 1.35E-08 | + | 1.93E-03 | - | **9.49E-04** |
| cg20070323 | 15 | 72,520,350 | *THSD4* | 444.6 | 15q23 | - | 1.35E-08 | - | 1.90E-03 | + | 3.13E-20 |
| cg12934461 | 15 | 90,792,652 | *MAN2A2* | -652.8 | 15q26.1 | + | 1.52E-08 | - | 6.27E-04 | - | 5.05E-130 |
| cg21136508 | 16 | 80,058,605 | *MAF* | 424.0 | 16q23.2 | + | 4.69E-08 | - | 1.66E-03 | - | 3.32E-127 |
| cg07827796 | 19 | 33,622,959 | *RHPN2* | 67.2 | 19q13.11 | - | 7.74E-08 | - | 8.37E-04 | + | 1.56E-19 |
| cg19133199 | 19 | 41,869,409 | *B9D2* | Body | 19q13.2 | + | 5.48E-15 | - | 3.14E-05 | - | **3.08E-10** |
| cg10384570 | 19 | 41,947,118 | *TGFB1* | 87.3 | 19q13.2 | + | 2.85E-09 | - | 1.29E-03 | - | 1.76E-11 |
| cg11506090 | 20 | 6,427,265 | *RP5-1056H1.2* | 378.7 | 20p12.3 | + | 1.75E-18 | + | 1.06E-03 | + | 2.54E-43 |
| cg04969764 | 20 | 60,906,201 | *CABLES2* | -57.5 | 20q13.33 | + | 6.12E-28 | - | 9.07E-04 | - | **5.96E-14** |
| cg23531748 | 20 | 60,969,906 | *RP4-563E14.1* | -533.8 | 20q13.33 | - | 8.72E-11 | + | 1.04E-03 | - | 0.02 |
| cg14208357 | 20 | 60,977,008 | *RP11-93B14.10* | -249.3 | 20q13.33 | - | 1.28E-49 | - | 3.60E-04 | + | 4.04E-28 |

Chr, chromosome; Dir, direction of association/correlation.

[a] *P* values were calculated using SPrediXcan. All statistical tests were two-sided.

[b] *P* values were calculated using linear regression. All statistical tests were two-sided.

[c] *P* values were calculated using SPrediXcan and R *limma* package for data from GTEx and GEPIA2, respectively. For genes showing in both GTEx and GEPIA2 data, *P* values from GEPIA2 data are presented. *P* values from GTEx data are highlighted in bold. All statistical tests were two-sided.

**Supplementary Table 15. Consistent directions of associations across DNA methylation, gene expression, and renal cell cancer risk**

| CpG | Chr | Position (HG19) | Gene | Distance (Kb) | Cytoband | CpG-cancer | | CpG-gene | | Gene-cancer | |
|---|---|---|---|---|---|---|---|---|---|---|---|
| | | | | | | Dir | *P* [a] | Dir | *P* [b] | Dir | *P* [c] |
| cg27476253 | 11 | 69,066,655 | *TPCN2* | 162.9 | 11q13.3 | - | 1.46E-14 | - | 4.19E-03 | + | 2.40E-10 |
| cg13524857 | 11 | 69,240,192 | *CCND1* | -215.7 | 11q13.3 | + | 5.88E-09 | + | 3.61E-03 | + | 6.11E-62 |
| cg06511653 | 12 | 26,472,706 | *SSPN* | 20.5 | 12p12.1 | - | 8.84E-08 | - | 1.76E-04 | + | 1.08E-30 |

Chr, chromosome; Dir, direction of association/correlation.

[a] *P* values were calculated using SPrediXcan. All statistical tests were two-sided.

[b] *P* values were obtained from Liu H et al. *Nat Genet* 2022. All statistical tests were two-sided.

[c] *P* values were calculated using R *limma* package for GEPIA2 data.

**Supplementary Table 16. Consistent directions of associations across DNA methylation, gene expression, and lung cancer risk**

| CpG | Chr | Position (HG19) | Gene | Distance (Kb) | Cytoband | CpG-cancer | | CpG-gene | | Gene-cancer | |
|---|---|---|---|---|---|---|---|---|---|---|---|
| | | | | | | Dir | ***P*** [a] | Dir | ***P*** [b] | Dir | ***P*** [c] |
| cg25606641 | 5 | 1,729,927 | *SLC12A7* | 617.8 | 5p15.33 | + | 1.50E-11 | - | 1.94E-03 | - | 3.10E-14 |
| cg16898833 | 6 | 26,189,333 | *BTN3A2* | -176.1 | 6p22.2 | + | 7.51E-09 | - | 1.90E-05 | - | 9.99E-16 |
| cg12315302 | 6 | 26,189,340 | *BTN3A2* | -176.1 | 6p22.2 | + | 1.44E-08 | - | 1.53E-05 | - | 9.99E-16 |
| cg01168740 | 6 | 26,229,103 | *RP1-221C16.8* | 56.1 | 6p22.2 | + | 5.96E-08 | + | 1.36E-04 | + | **2.80E-06** |
| cg19918208 | 6 | 28,828,364 | *PGBD1* | 558.0 | 6p22.1 | - | 6.07E-13 | + | 7.19E-04 | - | 7.02E-31 |
| cg00903577 | 6 | 28,831,109 | *ZFP57* | -809.1 | 6p22.1 | - | 1.53E-10 | - | 2.02E-04 | + | 2.03E-03 |
| cg10420223 | 6 | 29,526,561 | *ZFP57* | -113.6 | 6p22.1 | - | 7.65E-16 | - | 1.89E-04 | + | 2.03E-03 |
| cg10420223 | 6 | 29,526,561 | *ZBED9* | 942.6 | 6p22.1 | - | 7.65E-16 | - | 3.65E-04 | + | 1.09E-20 |
| cg10420223 | 6 | 29,526,561 | *HLA-K* | -368.4 | 6p22.1 | - | 7.65E-16 | + | 1.45E-03 | - | 7.54E-09 |
| cg16118803 | 6 | 29,629,716 | *HLA-V* | -129.0 | 6p22.1 | + | 2.28E-12 | - | 2.71E-04 | - | 3.55E-83 |
| cg16118803 | 6 | 29,629,716 | *ZFP57* | -10.5 | 6p22.1 | + | 2.28E-12 | + | 5.25E-04 | + | 2.03E-03 |
| cg16118803 | 6 | 29,629,716 | *HCG4* | -129.8 | 6p22.1 | + | 2.28E-12 | - | 1.42E-03 | - | 2.46E-03 |
| cg16118803 | 6 | 29,629,716 | *HCG4B* | -264.0 | 6p22.1 | + | 2.28E-12 | - | 2.40E-03 | - | 2.92E-11 |
| cg16118803 | 6 | 29,629,716 | *HLA-J* | -344.0 | 6p22.1 | + | 2.28E-12 | - | 4.31E-03 | - | 3.82E-25 |
| cg05279622 | 6 | 29,629,758 | *ZFP57* | -10.4 | 6p22.1 | + | 3.70E-13 | + | 1.75E-05 | + | 2.03E-03 |
| cg03147503 | 6 | 29,633,969 | *ZFP57* | -6.2 | 6p22.1 | + | 2.87E-09 | + | 1.10E-04 | + | 2.03E-03 |
| cg11358777 | 6 | 29,638,498 | *RANP1* | -815.2 | 6p22.1 | + | 7.29E-13 | + | 5.58E-04 | + | 0.04 |
| cg12463578 | 6 | 29,644,756 | *TRIM39* | -650.3 | 6p22.1 | - | 1.18E-12 | + | 5.90E-04 | - | 3.29E-17 |
| cg12463578 | 6 | 29,644,756 | *ZFP57* | Body | 6p22.1 | - | 1.18E-12 | - | 1.25E-03 | + | 2.03E-03 |
| cg25978138 | 6 | 29,648,161 | *ZFP57* | Body | 6p22.1 | - | 4.56E-13 | - | 1.62E-19 | + | 2.03E-03 |
| cg25978138 | 6 | 29,648,161 | *HCG4* | -111.4 | 6p22.1 | - | 4.56E-13 | + | 1.21E-06 | - | 2.46E-03 |
| cg25978138 | 6 | 29,648,161 | *HLA-K* | -246.8 | 6p22.1 | - | 4.56E-13 | + | 3.05E-05 | - | 7.54E-09 |
| cg25978138 | 6 | 29,648,161 | *HCP5B* | -191.5 | 6p22.1 | - | 4.56E-13 | - | 1.27E-04 | + | **5.97E-10** |
| cg25978138 | 6 | 29,648,161 | *HLA-H* | -208.1 | 6p22.1 | - | 4.56E-13 | - | 6.57E-04 | + | 6.78E-22 |
| cg25978138 | 6 | 29,648,161 | *HCG4B* | -245.6 | 6p22.1 | - | 4.56E-13 | + | 1.66E-03 | - | 2.92E-11 |
| cg11747594 | 6 | 29,648,225 | *ZFP57* | Body | 6p22.1 | - | 3.81E-12 | - | 4.44E-26 | + | 2.03E-03 |
| cg11747594 | 6 | 29,648,225 | *HLA-K* | -246.7 | 6p22.1 | - | 3.81E-12 | + | 1.13E-07 | - | 7.54E-09 |
| cg11747594 | 6 | 29,648,225 | *HCG4* | -111.3 | 6p22.1 | - | 3.81E-12 | + | 1.72E-05 | - | 2.46E-03 |
| cg11747594 | 6 | 29,648,225 | *HCP5B* | -191.5 | 6p22.1 | - | 3.81E-12 | - | 2.29E-04 | + | **5.97E-10** |
| cg11747594 | 6 | 29,648,225 | *HCG4B* | -245.5 | 6p22.1 | - | 3.81E-12 | + | 2.92E-04 | - | 2.92E-11 |
| cg11747594 | 6 | 29,648,225 | *HLA-H* | -208.0 | 6p22.1 | - | 3.81E-12 | - | 4.42E-04 | + | 6.78E-22 |
| cg11747594 | 6 | 29,648,225 | *HCG4P5* | -261.7 | 6p22.1 | - | 3.81E-12 | + | 1.83E-03 | - | 2.20E-09 |
| cg11747594 | 6 | 29,648,225 | *HLA-J* | -325.5 | 6p22.1 | - | 3.81E-12 | + | 2.13E-03 | - | 3.82E-25 |
| cg11747594 | 6 | 29,648,225 | *MRPS18B* | -937.3 | 6p22.1 | - | 3.81E-12 | - | 5.31E-03 | + | 7.65E-49 |
| cg11747594 | 6 | 29,648,225 | *Xbac-BPG170G13.3* | -71.5 | 6p22.1 | - | 3.81E-12 | + | 6.54E-03 | - | 2.21E-38 |
| cg15708526 | 6 | 29,648,271 | *ZFP57* | Body | 6p22.1 | - | 6.88E-12 | - | 2.61E-27 | + | 2.03E-03 |
| cg15708526 | 6 | 29,648,271 | *HLA-K* | -246.7 | 6p22.1 | - | 6.88E-12 | + | 8.61E-07 | - | 7.54E-09 |
| cg15708526 | 6 | 29,648,271 | *HCG4* | -111.3 | 6p22.1 | - | 6.88E-12 | + | 3.45E-06 | - | 2.46E-03 |
| cg15708526 | 6 | 29,648,271 | *HLA-H* | -208.0 | 6p22.1 | - | 6.88E-12 | - | 9.40E-05 | + | 6.78E-22 |
| cg15708526 | 6 | 29,648,271 | *MOG* | 8.1 | 6p22.1 | - | 6.88E-12 | - | 1.02E-04 | + | **6.22E-10** |
| cg15708526 | 6 | 29,648,271 | *HCG4P5* | -261.6 | 6p22.1 | - | 6.88E-12 | + | 1.47E-04 | - | 2.20E-09 |
| cg15708526 | 6 | 29,648,271 | *HCG4B* | -245.5 | 6p22.1 | - | 6.88E-12 | + | 1.54E-04 | - | 2.92E-11 |
| cg15708526 | 6 | 29,648,271 | *HCP5B* | -191.4 | 6p22.1 | - | 6.88E-12 | - | 2.55E-04 | + | **5.97E-10** |
| cg04071440 | 6 | 29,648,275 | *ZFP57* | Body | 6p22.1 | - | 1.38E-12 | - | 1.70E-24 | + | 2.03E-03 |
| cg04071440 | 6 | 29,648,275 | *HLA-K* | -246.7 | 6p22.1 | - | 1.38E-12 | + | 1.26E-06 | - | 7.54E-09 |
| cg04071440 | 6 | 29,648,275 | *HCG4* | -111.3 | 6p22.1 | - | 1.38E-12 | + | 3.22E-06 | - | 2.46E-03 |
| cg04071440 | 6 | 29,648,275 | *HLA-H* | -208.0 | 6p22.1 | - | 1.38E-12 | - | 5.81E-05 | + | 6.78E-22 |
| cg04071440 | 6 | 29,648,275 | *HCP5B* | -191.4 | 6p22.1 | - | 1.38E-12 | - | 1.11E-04 | + | **5.97E-10** |
| cg04071440 | 6 | 29,648,275 | *IFITM4P* | -70.2 | 6p22.1 | - | 1.38E-12 | + | 2.56E-03 | - | 3.64E-60 |
| cg04071440 | 6 | 29,648,275 | *HCG4B* | -245.5 | 6p22.1 | - | 1.38E-12 | + | 3.18E-03 | - | 2.92E-11 |
| cg04071440 | 6 | 29,648,275 | *HCG4P5* | -261.6 | 6p22.1 | - | 1.38E-12 | + | 3.52E-03 | - | 2.20E-09 |
| cg04071440 | 6 | 29,648,275 | *RNF39* | -389.8 | 6p22.1 | - | 1.38E-12 | + | 3.73E-03 | - | **1.57E-13** |
| cg08022281 | 6 | 29,648,345 | *ZFP57* | Body | 6p22.1 | - | 1.46E-13 | - | 6.40E-24 | + | 2.03E-03 |
| cg08022281 | 6 | 29,648,345 | *HLA-K* | -246.6 | 6p22.1 | - | 1.46E-13 | + | 4.67E-06 | - | 7.54E-09 |
| cg08022281 | 6 | 29,648,345 | *HLA-H* | -207.9 | 6p22.1 | - | 1.46E-13 | - | 2.45E-05 | + | 6.78E-22 |
| cg08022281 | 6 | 29,648,345 | *HCG4* | -111.2 | 6p22.1 | - | 1.46E-13 | + | 3.45E-05 | - | 2.46E-03 |
| cg08022281 | 6 | 29,648,345 | *HCP5B* | -191.3 | 6p22.1 | - | 1.46E-13 | - | 5.63E-04 | + | **5.97E-10** |
| cg08022281 | 6 | 29,648,345 | *HCG4B* | -245.4 | 6p22.1 | - | 1.46E-13 | + | 2.37E-03 | - | 2.92E-11 |
| cg10648573 | 6 | 29,648,348 | *ZFP57* | Body | 6p22.1 | - | 1.53E-12 | - | 8.19E-25 | + | 2.03E-03 |
| cg10648573 | 6 | 29,648,348 | *HLA-K* | -246.6 | 6p22.1 | - | 1.53E-12 | + | 4.05E-07 | - | 7.54E-09 |
| cg10648573 | 6 | 29,648,348 | *HCG4* | -111.2 | 6p22.1 | - | 1.53E-12 | + | 5.38E-06 | - | 2.46E-03 |
| cg10648573 | 6 | 29,648,348 | *HLA-H* | -207.9 | 6p22.1 | - | 1.53E-12 | - | 8.50E-05 | + | 6.78E-22 |
| cg10648573 | 6 | 29,648,348 | *HCG4B* | -245.4 | 6p22.1 | - | 1.53E-12 | + | 4.94E-04 | - | 2.92E-11 |
| cg10648573 | 6 | 29,648,348 | *HCP5B* | -191.3 | 6p22.1 | - | 1.53E-12 | - | 1.80E-03 | + | **5.97E-10** |
| cg10648573 | 6 | 29,648,348 | *HCG4P5* | -261.5 | 6p22.1 | - | 1.53E-12 | + | 2.70E-03 | - | 2.20E-09 |
| cg12644888 | 6 | 29,648,360 | *ZFP57* | Body | 6p22.1 | - | 4.87E-12 | - | 1.01E-22 | + | 2.03E-03 |
| cg12644888 | 6 | 29,648,360 | *HLA-K* | -246.6 | 6p22.1 | - | 4.87E-12 | + | 1.20E-06 | - | 7.54E-09 |
| cg12644888 | 6 | 29,648,360 | *HLA-H* | -207.9 | 6p22.1 | - | 4.87E-12 | - | 6.32E-06 | + | 6.78E-22 |

| | | | | | | | | | | | |
|---|---|---|---|---|---|---|---|---|---|---|---|
| cg12644888 | 6 | 29,648,360 | *HCG4* | -111.2 | 6p22.1 | - | 4.87E-12 | + | 5.26E-05 | - | 2.46E-03 |
| cg12644888 | 6 | 29,648,360 | *HCP5B* | -191.3 | 6p22.1 | - | 4.87E-12 | - | 1.18E-04 | + | **5.97E-10** |
| cg12644888 | 6 | 29,648,360 | *HCG4B* | -245.4 | 6p22.1 | - | 4.87E-12 | + | 7.54E-04 | - | 2.92E-11 |
| cg12644888 | 6 | 29,648,360 | *IFITM4P* | -70.2 | 6p22.1 | - | 4.87E-12 | + | 3.34E-03 | - | 3.64E-60 |
| cg12644888 | 6 | 29,648,360 | *HCG4P5* | -261.5 | 6p22.1 | - | 4.87E-12 | + | 5.13E-03 | - | 2.20E-09 |
| cg22494932 | 6 | 29,648,379 | *ZFP57* | Body | 6p22.1 | - | 5.78E-12 | - | 6.06E-25 | + | 2.03E-03 |
| cg22494932 | 6 | 29,648,379 | *HCG4* | -111.2 | 6p22.1 | - | 5.78E-12 | + | 3.03E-07 | - | 2.46E-03 |
| cg22494932 | 6 | 29,648,379 | *HLA-K* | -246.6 | 6p22.1 | - | 5.78E-12 | + | 1.30E-06 | - | 7.54E-09 |
| cg22494932 | 6 | 29,648,379 | *HCG4B* | -245.4 | 6p22.1 | - | 5.78E-12 | + | 2.61E-04 | - | 2.92E-11 |
| cg22494932 | 6 | 29,648,379 | *HLA-H* | -207.9 | 6p22.1 | - | 5.78E-12 | - | 5.25E-04 | + | 6.78E-22 |
| cg22494932 | 6 | 29,648,379 | *HCP5B* | -191.3 | 6p22.1 | - | 5.78E-12 | - | 1.88E-03 | + | **5.97E-10** |
| cg22494932 | 6 | 29,648,379 | *HCG4P5* | -261.5 | 6p22.1 | - | 5.78E-12 | + | 1.94E-03 | - | 2.20E-09 |
| cg22494932 | 6 | 29,648,379 | *HLA-J* | -325.4 | 6p22.1 | - | 5.78E-12 | + | 5.01E-03 | - | 3.82E-25 |
| cg22494932 | 6 | 29,648,379 | *HCG16* | 692.1 | 6p22.1 | - | 5.78E-12 | - | 8.11E-03 | + | 2.61E-33 |
| cg22494932 | 6 | 29,648,379 | *TRIM31* | -422.3 | 6p22.1 | - | 5.78E-12 | + | 8.97E-03 | - | **4.64E-14** |
| cg25699073 | 6 | 29,648,381 | *ZFP57* | Body | 6p22.1 | - | 4.35E-13 | - | 7.16E-20 | + | 2.03E-03 |
| cg25699073 | 6 | 29,648,381 | *HLA-K* | -246.6 | 6p22.1 | - | 4.35E-13 | + | 2.86E-06 | - | 7.54E-09 |
| cg25699073 | 6 | 29,648,381 | *HCG4* | -111.2 | 6p22.1 | - | 4.35E-13 | + | 4.42E-06 | - | 2.46E-03 |
| cg25699073 | 6 | 29,648,381 | *HLA-H* | -207.9 | 6p22.1 | - | 4.35E-13 | - | 8.35E-04 | + | 6.78E-22 |
| cg25699073 | 6 | 29,648,381 | *HCG4P5* | -261.5 | 6p22.1 | - | 4.35E-13 | + | 9.12E-04 | - | 2.20E-09 |
| cg25699073 | 6 | 29,648,381 | *HCG4B* | -245.4 | 6p22.1 | - | 4.35E-13 | + | 2.58E-03 | - | 2.92E-11 |
| cg25699073 | 6 | 29,648,381 | *HCP5B* | -191.3 | 6p22.1 | - | 4.35E-13 | - | 2.84E-03 | + | **5.97E-10** |
| cg25699073 | 6 | 29,648,381 | *HLA-J* | -325.4 | 6p22.1 | - | 4.35E-13 | + | 7.12E-03 | - | 3.82E-25 |
| cg07134666 | 6 | 29,648,400 | *ZFP57* | Body | 6p22.1 | - | 2.69E-12 | - | 8.38E-28 | + | 2.03E-03 |
| cg07134666 | 6 | 29,648,400 | *HCG4* | -111.2 | 6p22.1 | - | 2.69E-12 | + | 7.66E-07 | - | 2.46E-03 |
| cg07134666 | 6 | 29,648,400 | *HLA-K* | -246.6 | 6p22.1 | - | 2.69E-12 | + | 1.07E-06 | - | 7.54E-09 |
| cg07134666 | 6 | 29,648,400 | *HCG4B* | -245.4 | 6p22.1 | - | 2.69E-12 | + | 3.80E-04 | - | 2.92E-11 |
| cg07134666 | 6 | 29,648,400 | *HLA-H* | -207.8 | 6p22.1 | - | 2.69E-12 | - | 6.51E-04 | + | 6.78E-22 |
| cg07134666 | 6 | 29,648,400 | *HCP5B* | -191.3 | 6p22.1 | - | 2.69E-12 | - | 1.36E-03 | + | **5.97E-10** |
| cg07134666 | 6 | 29,648,400 | *HLA-J* | -325.4 | 6p22.1 | - | 2.69E-12 | + | 2.10E-03 | - | 3.82E-25 |
| cg07134666 | 6 | 29,648,400 | *HCG4P5* | -261.5 | 6p22.1 | - | 2.69E-12 | + | 7.11E-03 | - | 2.20E-09 |
| cg00588198 | 6 | 29,648,452 | *ZFP57* | Body | 6p22.1 | - | 4.60E-10 | - | 1.12E-22 | + | 2.03E-03 |
| cg00588198 | 6 | 29,648,452 | *HLA-K* | -246.5 | 6p22.1 | - | 4.60E-10 | + | 1.50E-06 | - | 7.54E-09 |
| cg00588198 | 6 | 29,648,452 | *HCG4* | -111.1 | 6p22.1 | - | 4.60E-10 | + | 3.37E-05 | - | 2.46E-03 |
| cg00588198 | 6 | 29,648,452 | *HLA-H* | -207.8 | 6p22.1 | - | 4.60E-10 | - | 4.99E-05 | + | 6.78E-22 |
| cg00588198 | 6 | 29,648,452 | *HCG4P5* | -261.4 | 6p22.1 | - | 4.60E-10 | + | 3.89E-04 | - | 2.20E-09 |
| cg00588198 | 6 | 29,648,452 | *HCG4B* | -245.3 | 6p22.1 | - | 4.60E-10 | + | 1.03E-03 | - | 2.92E-11 |
| cg00588198 | 6 | 29,648,452 | *HCP5B* | -191.2 | 6p22.1 | - | 4.60E-10 | - | 1.78E-03 | + | **5.97E-10** |
| cg00588198 | 6 | 29,648,452 | *RNF39* | -389.7 | 6p22.1 | - | 4.60E-10 | + | 2.77E-03 | - | **1.57E-13** |
| cg00588198 | 6 | 29,648,452 | *TRIM31* | -422.2 | 6p22.1 | - | 4.60E-10 | + | 3.07E-03 | - | **4.64E-14** |
| cg00588198 | 6 | 29,648,452 | *IFITM4P* | -70.1 | 6p22.1 | - | 4.60E-10 | + | 4.96E-03 | - | 3.64E-60 |
| cg16885113 | 6 | 29,648,507 | *ZFP57* | Body | 6p22.1 | - | 9.19E-12 | - | 1.25E-29 | + | 2.03E-03 |
| cg16885113 | 6 | 29,648,507 | *HLA-K* | -246.5 | 6p22.1 | - | 9.19E-12 | + | 1.24E-06 | - | 7.54E-09 |
| cg16885113 | 6 | 29,648,507 | *HLA-H* | -207.7 | 6p22.1 | - | 9.19E-12 | - | 1.65E-06 | + | 6.78E-22 |
| cg16885113 | 6 | 29,648,507 | *HCG4* | -111.1 | 6p22.1 | - | 9.19E-12 | + | 4.15E-05 | - | 2.46E-03 |
| cg16885113 | 6 | 29,648,507 | *HCG4B* | -245.3 | 6p22.1 | - | 9.19E-12 | + | 9.14E-05 | - | 2.92E-11 |
| cg16885113 | 6 | 29,648,507 | *HCP5B* | -191.2 | 6p22.1 | - | 9.19E-12 | - | 2.61E-04 | + | **5.97E-10** |
| cg16885113 | 6 | 29,648,507 | *HCG4P5* | -261.4 | 6p22.1 | - | 9.19E-12 | + | 2.21E-03 | - | 2.20E-09 |
| cg16885113 | 6 | 29,648,507 | *TRIM31* | -422.2 | 6p22.1 | - | 9.19E-12 | + | 3.43E-03 | - | **4.64E-14** |
| cg20228636 | 6 | 29,648,525 | *ZFP57* | Body | 6p22.1 | - | 6.60E-12 | - | 3.32E-30 | + | 2.03E-03 |
| cg20228636 | 6 | 29,648,525 | *HLA-K* | -246.4 | 6p22.1 | - | 6.60E-12 | + | 5.15E-08 | - | 7.54E-09 |
| cg20228636 | 6 | 29,648,525 | *HLA-H* | -207.7 | 6p22.1 | - | 6.60E-12 | - | 4.37E-06 | + | 6.78E-22 |
| cg20228636 | 6 | 29,648,525 | *HCG4* | -111.0 | 6p22.1 | - | 6.60E-12 | + | 6.72E-06 | - | 2.46E-03 |
| cg20228636 | 6 | 29,648,525 | *HCG4B* | -245.2 | 6p22.1 | - | 6.60E-12 | + | 9.05E-05 | - | 2.92E-11 |
| cg20228636 | 6 | 29,648,525 | *HCP5B* | -191.2 | 6p22.1 | - | 6.60E-12 | - | 1.46E-04 | + | **5.97E-10** |
| cg20228636 | 6 | 29,648,525 | *IFITM4P* | -70.0 | 6p22.1 | - | 6.60E-12 | + | 2.58E-03 | - | 3.64E-60 |
| cg20228636 | 6 | 29,648,525 | *HCG4P5* | -261.4 | 6p22.1 | - | 6.60E-12 | + | 3.67E-03 | - | 2.20E-09 |
| cg20228636 | 6 | 29,648,525 | *TRIM31* | -422.2 | 6p22.1 | - | 6.60E-12 | + | 4.51E-03 | - | **4.64E-14** |
| cg11383134 | 6 | 29,648,590 | *ZFP57* | Body | 6p22.1 | - | 1.36E-11 | - | 7.01E-33 | + | 2.03E-03 |
| cg11383134 | 6 | 29,648,590 | *HCG4* | -111.0 | 6p22.1 | - | 1.36E-11 | + | 9.88E-08 | - | 2.46E-03 |
| cg11383134 | 6 | 29,648,590 | *HLA-K* | -246.4 | 6p22.1 | - | 1.36E-11 | + | 3.90E-06 | - | 7.54E-09 |
| cg11383134 | 6 | 29,648,590 | *HLA-H* | -207.7 | 6p22.1 | - | 1.36E-11 | - | 1.44E-05 | + | 6.78E-22 |
| cg11383134 | 6 | 29,648,590 | *HCP5B* | -191.1 | 6p22.1 | - | 1.36E-11 | - | 8.29E-05 | + | **5.97E-10** |
| cg11383134 | 6 | 29,648,590 | *HCG4B* | -245.2 | 6p22.1 | - | 1.36E-11 | + | 1.20E-03 | - | 2.92E-11 |
| cg11383134 | 6 | 29,648,590 | *HCG4P5* | -261.3 | 6p22.1 | - | 1.36E-11 | + | 1.27E-03 | - | 2.20E-09 |
| cg11383134 | 6 | 29,648,590 | *TRIM31* | -422.1 | 6p22.1 | - | 1.36E-11 | + | 2.76E-03 | - | **4.64E-14** |
| cg11383134 | 6 | 29,648,590 | *HLA-J* | -325.2 | 6p22.1 | - | 1.36E-11 | + | 3.73E-03 | - | 3.82E-25 |
| cg03198009 | 6 | 29,648,604 | *ZFP57* | Body | 6p22.1 | - | 9.74E-11 | - | 2.19E-28 | + | 2.03E-03 |
| cg03198009 | 6 | 29,648,604 | *HLA-K* | -246.4 | 6p22.1 | - | 9.74E-11 | + | 3.87E-07 | - | 7.54E-09 |
| cg03198009 | 6 | 29,648,604 | *HLA-H* | -207.6 | 6p22.1 | - | 9.74E-11 | - | 9.73E-06 | + | 6.78E-22 |
| cg03198009 | 6 | 29,648,604 | *HCG4* | -111.0 | 6p22.1 | - | 9.74E-11 | + | 2.72E-05 | - | 2.46E-03 |
| cg03198009 | 6 | 29,648,604 | *HCG4P5* | -261.3 | 6p22.1 | - | 9.74E-11 | + | 7.67E-05 | - | 2.20E-09 |

| | | | | | | | | | | | |
|---|---|---|---|---|---|---|---|---|---|---|---|
| cg03198009 | 6 | 29,648,604 | *HCG4B* | -245.2 | 6p22.1 | - | 9.74E-11 | + | 2.33E-04 | - | 2.92E-11 |
| cg03198009 | 6 | 29,648,604 | *RNF39* | -389.5 | 6p22.1 | - | 9.74E-11 | + | 3.00E-03 | - | **1.57E-13** |
| cg03198009 | 6 | 29,648,604 | *HCP5B* | -191.1 | 6p22.1 | - | 9.74E-11 | - | 4.46E-03 | + | **5.97E-10** |
| cg03198009 | 6 | 29,648,604 | *TRIM31* | -422.1 | 6p22.1 | - | 9.74E-11 | + | 7.14E-03 | - | **4.64E-14** |
| cg03198009 | 6 | 29,648,604 | *IFITM4P* | -69.9 | 6p22.1 | - | 9.74E-11 | + | 9.04E-03 | - | 3.64E-60 |
| cg03449857 | 6 | 29,648,623 | *ZFP57* | Body | 6p22.1 | - | 3.39E-11 | - | 2.87E-30 | + | 2.03E-03 |
| cg03449857 | 6 | 29,648,623 | *HLA-H* | -207.6 | 6p22.1 | - | 3.39E-11 | - | 6.01E-07 | + | 6.78E-22 |
| cg03449857 | 6 | 29,648,623 | *HLA-K* | -246.3 | 6p22.1 | - | 3.39E-11 | + | 2.51E-06 | - | 7.54E-09 |
| cg03449857 | 6 | 29,648,623 | *HCG4* | -110.9 | 6p22.1 | - | 3.39E-11 | + | 4.31E-06 | - | 2.46E-03 |
| cg03449857 | 6 | 29,648,623 | *HCP5B* | -191.1 | 6p22.1 | - | 3.39E-11 | - | 6.81E-05 | + | **5.97E-10** |
| cg03449857 | 6 | 29,648,623 | *HCG4B* | -245.1 | 6p22.1 | - | 3.39E-11 | + | 3.33E-04 | - | 2.92E-11 |
| cg03449857 | 6 | 29,648,623 | *IFITM4P* | -69.9 | 6p22.1 | - | 3.39E-11 | + | 1.42E-03 | - | 3.64E-60 |
| cg03449857 | 6 | 29,648,623 | *HCG4P5* | -261.3 | 6p22.1 | - | 3.39E-11 | + | 4.02E-03 | - | 2.20E-09 |
| cg03449857 | 6 | 29,648,623 | *TRIM31* | -422.1 | 6p22.1 | - | 3.39E-11 | + | 4.87E-03 | - | **4.64E-14** |
| cg03449857 | 6 | 29,648,623 | *RNF39* | -389.5 | 6p22.1 | - | 3.39E-11 | + | 8.81E-03 | - | **1.57E-13** |
| cg15570656 | 6 | 29,648,628 | *ZFP57* | Body | 6p22.1 | - | 4.27E-12 | - | 5.64E-28 | + | 2.03E-03 |
| cg15570656 | 6 | 29,648,628 | *HLA-K* | -246.3 | 6p22.1 | - | 4.27E-12 | + | 5.04E-07 | - | 7.54E-09 |
| cg15570656 | 6 | 29,648,628 | *HLA-H* | -207.6 | 6p22.1 | - | 4.27E-12 | - | 1.12E-06 | + | 6.78E-22 |
| cg15570656 | 6 | 29,648,628 | *HCG4* | -110.9 | 6p22.1 | - | 4.27E-12 | + | 2.13E-06 | - | 2.46E-03 |
| cg15570656 | 6 | 29,648,628 | *HCG4P5* | -261.3 | 6p22.1 | - | 4.27E-12 | + | 1.30E-04 | - | 2.20E-09 |
| cg15570656 | 6 | 29,648,628 | *HCP5B* | -191.0 | 6p22.1 | - | 4.27E-12 | - | 5.26E-04 | + | **5.97E-10** |
| cg15570656 | 6 | 29,648,628 | *HCG4B* | -245.1 | 6p22.1 | - | 4.27E-12 | + | 5.68E-04 | - | 2.92E-11 |
| cg15570656 | 6 | 29,648,628 | *TRIM31* | -422.1 | 6p22.1 | - | 4.27E-12 | + | 8.68E-04 | - | **4.64E-14** |
| cg15570656 | 6 | 29,648,628 | *RNF39* | -389.5 | 6p22.1 | - | 4.27E-12 | + | 1.32E-03 | - | **1.57E-13** |
| cg13835168 | 6 | 29,648,756 | *ZFP57* | Body | 6p22.1 | - | 7.00E-11 | - | 1.23E-25 | + | 2.03E-03 |
| cg13835168 | 6 | 29,648,756 | *HCG4* | -110.8 | 6p22.1 | - | 7.00E-11 | + | 8.26E-07 | - | 2.46E-03 |
| cg13835168 | 6 | 29,648,756 | *HLA-K* | -246.2 | 6p22.1 | - | 7.00E-11 | + | 8.88E-07 | - | 7.54E-09 |
| cg13835168 | 6 | 29,648,756 | *HLA-H* | -207.5 | 6p22.1 | - | 7.00E-11 | - | 1.89E-04 | + | 6.78E-22 |
| cg13835168 | 6 | 29,648,756 | *HCG4P5* | -261.1 | 6p22.1 | - | 7.00E-11 | + | 4.11E-04 | - | 2.20E-09 |
| cg13835168 | 6 | 29,648,756 | *IFITM4P* | -69.8 | 6p22.1 | - | 7.00E-11 | + | 1.54E-03 | - | 3.64E-60 |
| cg13835168 | 6 | 29,648,756 | *HCG4B* | -245.0 | 6p22.1 | - | 7.00E-11 | + | 1.73E-03 | - | 2.92E-11 |
| cg13835168 | 6 | 29,648,756 | *TRIM31* | -421.9 | 6p22.1 | - | 7.00E-11 | + | 2.30E-03 | - | **4.64E-14** |
| cg13835168 | 6 | 29,648,756 | *HCP5B* | -190.9 | 6p22.1 | - | 7.00E-11 | - | 3.19E-03 | + | **5.97E-10** |
| cg13835168 | 6 | 29,648,756 | *HLA-J* | -325.0 | 6p22.1 | - | 7.00E-11 | + | 4.87E-03 | - | 3.82E-25 |
| cg08041448 | 6 | 29,648,901 | *ZFP57* | 0.0 | 6p22.1 | - | 7.18E-08 | - | 2.25E-22 | + | 2.03E-03 |
| cg08041448 | 6 | 29,648,901 | *HCG4* | -110.7 | 6p22.1 | - | 7.18E-08 | + | 1.36E-06 | - | 2.46E-03 |
| cg08041448 | 6 | 29,648,901 | *HLA-H* | -207.3 | 6p22.1 | - | 7.18E-08 | - | 1.78E-05 | + | 6.78E-22 |
| cg08041448 | 6 | 29,648,901 | *HLA-K* | -246.1 | 6p22.1 | - | 7.18E-08 | + | 7.52E-05 | - | 7.54E-09 |
| cg08041448 | 6 | 29,648,901 | *IFITM4P* | -69.6 | 6p22.1 | - | 7.18E-08 | + | 9.06E-04 | - | 3.64E-60 |
| cg08041448 | 6 | 29,648,901 | *HCP5B* | -190.8 | 6p22.1 | - | 7.18E-08 | - | 1.21E-03 | + | **5.97E-10** |
| cg08041448 | 6 | 29,648,901 | *HCG4B* | -244.9 | 6p22.1 | - | 7.18E-08 | + | 1.71E-03 | - | 2.92E-11 |
| cg08041448 | 6 | 29,648,901 | *TRIM31* | -421.8 | 6p22.1 | - | 7.18E-08 | + | 1.73E-03 | - | **4.64E-14** |
| cg08041448 | 6 | 29,648,901 | *HCG4P5* | -261.0 | 6p22.1 | - | 7.18E-08 | + | 6.08E-03 | - | 2.20E-09 |
| cg08041448 | 6 | 29,648,901 | *RNF39* | -389.2 | 6p22.1 | - | 7.18E-08 | + | 7.00E-03 | - | **1.57E-13** |
| cg03278703 | 6 | 29,691,107 | *IFITM4P* | -27.4 | 6p22.1 | + | 7.25E-11 | - | 3.87E-04 | - | 3.64E-60 |
| cg25252977 | 6 | 29,696,650 | *ZFP57* | 47.8 | 6p22.1 | - | 2.10E-14 | - | 1.42E-05 | + | 2.03E-03 |
| cg25252977 | 6 | 29,696,650 | *HCG4* | -62.9 | 6p22.1 | - | 2.10E-14 | + | 5.37E-04 | - | 2.46E-03 |
| cg25252977 | 6 | 29,696,650 | *HCG4P5* | -213.2 | 6p22.1 | - | 2.10E-14 | + | 7.20E-04 | - | 2.20E-09 |
| cg08451469 | 6 | 29,711,588 | *ZFP57* | 62.7 | 6p22.1 | - | 4.29E-10 | - | 9.43E-12 | + | 2.03E-03 |
| cg08451469 | 6 | 29,711,588 | *HLA-H* | -144.7 | 6p22.1 | - | 4.29E-10 | - | 1.95E-05 | + | 6.78E-22 |
| cg08451469 | 6 | 29,711,588 | *HCG4* | -48.0 | 6p22.1 | - | 4.29E-10 | + | 2.40E-05 | - | 2.46E-03 |
| cg08451469 | 6 | 29,711,588 | *HCP5B* | -128.1 | 6p22.1 | - | 4.29E-10 | - | 3.27E-05 | + | **5.97E-10** |
| cg08451469 | 6 | 29,711,588 | *HLA-K* | -183.4 | 6p22.1 | - | 4.29E-10 | + | 1.19E-04 | - | 7.54E-09 |
| cg08696107 | 6 | 29,759,949 | *TRIM39* | -535.1 | 6p22.1 | + | 1.08E-08 | - | 6.04E-04 | - | 3.29E-17 |
| cg03432955 | 6 | 29,795,501 | *HLA-K* | -99.5 | 6p22.1 | - | 4.09E-09 | + | 2.50E-06 | - | 7.54E-09 |
| cg03432955 | 6 | 29,795,501 | *ZFP57* | 146.6 | 6p22.1 | - | 4.09E-09 | - | 1.37E-04 | + | 2.03E-03 |
| cg03432955 | 6 | 29,795,501 | *HCG4* | 35.3 | 6p22.1 | - | 4.09E-09 | + | 8.46E-04 | - | 2.46E-03 |
| cg03432955 | 6 | 29,795,501 | *HLA-H* | -60.7 | 6p22.1 | - | 4.09E-09 | - | 1.56E-03 | + | 6.78E-22 |
| cg00126638 | 6 | 29,795,583 | *HLA-K* | -99.4 | 6p22.1 | - | 3.83E-08 | + | 1.68E-04 | - | 7.54E-09 |
| cg00126638 | 6 | 29,795,583 | *HLA-H* | -60.7 | 6p22.1 | - | 3.83E-08 | - | 1.35E-03 | + | 6.78E-22 |
| cg00126638 | 6 | 29,795,583 | *ZFP57* | 146.7 | 6p22.1 | - | 3.83E-08 | - | 2.32E-03 | + | 2.03E-03 |
| cg03521696 | 6 | 29,795,595 | *HLA-K* | -99.4 | 6p22.1 | - | 2.44E-08 | + | 2.74E-04 | - | 7.54E-09 |
| cg11862551 | 6 | 29,817,963 | *IFITM4P* | 99.0 | 6p22.1 | - | 1.56E-07 | + | 1.03E-05 | - | 3.64E-60 |
| cg11862551 | 6 | 29,817,963 | *Xbac-BPG170G13.3* | 98.0 | 6p22.1 | - | 1.56E-07 | + | 2.60E-04 | - | 2.21E-38 |
| cg03863342 | 6 | 29,831,661 | *HCG4* | 71.5 | 6p22.1 | - | 1.13E-15 | + | 1.92E-05 | - | 2.46E-03 |
| cg03863342 | 6 | 29,831,661 | *ZFP57* | 182.8 | 6p22.1 | - | 1.13E-15 | - | 7.33E-05 | + | 2.03E-03 |
| cg03863342 | 6 | 29,831,661 | *HLA-K* | -63.3 | 6p22.1 | - | 1.13E-15 | + | 4.30E-04 | - | 7.54E-09 |
| cg26121931 | 6 | 29,895,204 | *ZFP57* | 246.3 | 6p22.1 | + | 5.70E-10 | + | 3.44E-12 | + | 2.03E-03 |
| cg26121931 | 6 | 29,895,204 | *HLA-K* | Body | 6p22.1 | + | 5.70E-10 | - | 1.65E-05 | - | 7.54E-09 |
| cg26121931 | 6 | 29,895,204 | *HCG4* | 135.0 | 6p22.1 | + | 5.70E-10 | - | 1.02E-04 | - | 2.46E-03 |
| cg26121931 | 6 | 29,895,204 | *HCP5B* | 53.6 | 6p22.1 | + | 5.70E-10 | + | 1.44E-04 | + | **5.97E-10** |
| cg26121931 | 6 | 29,895,204 | *HCG4B* | 0.5 | 6p22.1 | + | 5.70E-10 | - | 1.68E-04 | - | 2.92E-11 |

| | | | | | | | | | | | |
|---|---|---|---|---|---|---|---|---|---|---|---|
| cg26121931 | 6 | 29,895,204 | *HLA-J* | -78.5 | 6p22.1 | + | 5.70E-10 | - | 1.27E-03 | - | 3.82E-25 |
| cg24838316 | 6 | 29,895,260 | *ZFP57* | 246.4 | 6p22.1 | + | 3.09E-11 | + | 2.56E-11 | + | 2.03E-03 |
| cg24838316 | 6 | 29,895,260 | *HLA-K* | Body | 6p22.1 | + | 3.09E-11 | - | 4.67E-07 | - | 7.54E-09 |
| cg24838316 | 6 | 29,895,260 | *HCG4* | 135.1 | 6p22.1 | + | 3.09E-11 | - | 4.72E-07 | - | 2.46E-03 |
| cg24838316 | 6 | 29,895,260 | *HCG4B* | 0.5 | 6p22.1 | + | 3.09E-11 | - | 3.54E-05 | - | 2.92E-11 |
| cg24838316 | 6 | 29,895,260 | *HLA-J* | -78.5 | 6p22.1 | + | 3.09E-11 | - | 7.40E-05 | - | 3.82E-25 |
| cg24838316 | 6 | 29,895,260 | *HCG4P5* | -14.6 | 6p22.1 | + | 3.09E-11 | - | 3.89E-04 | - | 2.20E-09 |
| cg24838316 | 6 | 29,895,260 | *IFITM4P* | 176.3 | 6p22.1 | + | 3.09E-11 | - | 1.28E-03 | - | 3.64E-60 |
| cg05523662 | 6 | 29,910,101 | *HCG4P5* | Body | 6p22.1 | + | 1.51E-08 | - | 4.54E-04 | - | 2.20E-09 |
| cg05523662 | 6 | 29,910,101 | *ZFP57* | 261.2 | 6p22.1 | + | 1.51E-08 | + | 7.78E-04 | + | 2.03E-03 |
| cg05523662 | 6 | 29,910,101 | *IFITM4P* | 191.2 | 6p22.1 | + | 1.51E-08 | - | 1.09E-03 | - | 3.64E-60 |
| cg05523662 | 6 | 29,910,101 | *HCG4* | 149.9 | 6p22.1 | + | 1.51E-08 | - | 1.76E-03 | - | 2.46E-03 |
| cg15319255 | 6 | 29,910,269 | *HCG4P5* | Body | 6p22.1 | - | 3.27E-10 | + | 8.65E-05 | - | 2.20E-09 |
| cg15319255 | 6 | 29,910,269 | *HLA-K* | 13.3 | 6p22.1 | - | 3.27E-10 | + | 1.44E-04 | - | 7.54E-09 |
| cg15319255 | 6 | 29,910,269 | *HLA-J* | -63.5 | 6p22.1 | - | 3.27E-10 | + | 3.22E-04 | - | 3.82E-25 |
| cg19448822 | 6 | 29,974,773 | *IFITM4P* | 255.9 | 6p22.1 | + | 6.34E-08 | - | 4.87E-04 | - | 3.64E-60 |
| cg08163199 | 6 | 29,974,858 | *TRIM39* | -320.2 | 6p22.1 | + | 2.39E-07 | - | 4.30E-04 | - | 3.29E-17 |
| cg08325845 | 6 | 29,974,886 | *TRIM39* | -320.2 | 6p22.1 | + | 1.47E-11 | - | 3.03E-04 | - | 3.29E-17 |
| cg08325845 | 6 | 29,974,886 | *DHX16* | -646.1 | 6p22.1 | + | 1.47E-11 | - | 7.08E-04 | - | 2.25E-14 |
| cg15448721 | 6 | 30,045,041 | *HLA-K* | 148.0 | 6p22.1 | - | 1.25E-11 | + | 2.91E-05 | - | 7.54E-09 |
| cg15448721 | 6 | 30,045,041 | *ZFP57* | 396.2 | 6p22.1 | - | 1.25E-11 | - | 7.50E-04 | + | 2.03E-03 |
| cg15448721 | 6 | 30,045,041 | *TRIM39* | -250.0 | 6p22.1 | - | 1.25E-11 | + | 2.08E-03 | - | 3.29E-17 |
| cg15448721 | 6 | 30,045,041 | *HCG4B* | 150.3 | 6p22.1 | - | 1.25E-11 | + | 3.03E-03 | - | 2.92E-11 |
| cg16219552 | 6 | 30,057,025 | *HLA-V* | 291.4 | 6p22.1 | - | 8.43E-13 | + | 3.25E-04 | - | 3.55E-83 |
| cg16219552 | 6 | 30,057,025 | *HLA-K* | 160.0 | 6p22.1 | - | 8.43E-13 | + | 6.94E-04 | - | 7.54E-09 |
| cg04906656 | 6 | 30,069,681 | *HCG4B* | 174.9 | 6p22.1 | + | 1.84E-11 | - | 2.57E-04 | - | 2.92E-11 |
| cg23642250 | 6 | 30,071,394 | *ZFP57* | 422.5 | 6p22.1 | + | 3.61E-08 | + | 1.36E-04 | + | 2.03E-03 |
| cg22377040 | 6 | 30,071,412 | *ZFP57* | 422.5 | 6p22.1 | + | 9.56E-09 | + | 2.37E-05 | + | 2.03E-03 |
| cg25721950 | 6 | 30,074,078 | *ZFP57* | 425.2 | 6p22.1 | + | 1.03E-12 | + | 3.15E-04 | + | 2.03E-03 |
| cg25721950 | 6 | 30,074,078 | *HCG4P5* | 163.2 | 6p22.1 | + | 1.03E-12 | - | 1.11E-03 | - | 2.20E-09 |
| cg25721950 | 6 | 30,074,078 | *TRIM31* | Body | 6p22.1 | + | 1.03E-12 | - | 1.70E-03 | - | **4.64E-14** |
| cg11229497 | 6 | 30,078,954 | *HCG4* | 318.7 | 6p22.1 | + | 1.16E-08 | - | 1.17E-04 | - | 2.46E-03 |
| cg11229497 | 6 | 30,078,954 | *IFITM4P* | 360.0 | 6p22.1 | + | 1.16E-08 | - | 4.29E-04 | - | 3.64E-60 |
| cg11229497 | 6 | 30,078,954 | *HCG4B* | 184.2 | 6p22.1 | + | 1.16E-08 | - | 7.39E-04 | - | 2.92E-11 |
| cg21232488 | 6 | 30,079,203 | *HLA-K* | 182.2 | 6p22.1 | + | 7.78E-08 | - | 1.07E-04 | - | 7.54E-09 |
| cg21232488 | 6 | 30,079,203 | *HCG4B* | 184.5 | 6p22.1 | + | 7.78E-08 | - | 1.37E-03 | - | 2.92E-11 |
| cg21232488 | 6 | 30,079,203 | *TRIM31* | Body | 6p22.1 | + | 7.78E-08 | - | 1.79E-03 | - | **4.64E-14** |
| cg21232488 | 6 | 30,079,203 | *ZFP57* | 430.3 | 6p22.1 | + | 7.78E-08 | + | 2.72E-03 | + | 2.03E-03 |
| cg09476067 | 6 | 30,418,581 | *TRIM39* | 107.1 | 6p21.33 | - | 6.67E-19 | + | 1.51E-04 | - | 3.29E-17 |
| cg27185298 | 6 | 30,885,205 | *HCG4B* | 990.5 | 6p21.33 | + | 9.62E-08 | - | 9.06E-05 | - | 2.92E-11 |
| cg03059420 | 6 | 30,923,241 | *VARS* | -822.1 | 6p21.33 | + | 9.09E-09 | + | 3.97E-04 | + | 3.18E-68 |
| cg15878568 | 6 | 30,923,865 | *LY6G5B* | -714.1 | 6p21.33 | + | 1.62E-07 | + | 2.33E-04 | + | **3.67E-03** |
| cg15878568 | 6 | 30,923,865 | *ABCF1* | 358.9 | 6p21.33 | + | 1.62E-07 | + | 2.44E-03 | + | 3.34E-03 |
| cg17849733 | 6 | 31,082,200 | *HCG22* | 54.5 | 6p21.33 | - | 6.10E-09 | + | 1.74E-04 | - | 6.62E-08 |
| cg27536700 | 6 | 31,166,759 | *PSORS1C3* | 21.1 | 6p21.33 | - | 1.70E-08 | + | 2.45E-04 | - | 3.30E-06 |
| cg10473907 | 6 | 31,167,664 | *PSORS1C3* | 22.0 | 6p21.33 | + | 1.93E-07 | - | 6.66E-06 | - | 3.30E-06 |
| cg21091985 | 6 | 31,706,222 | *VARS* | -39.1 | 6p21.33 | + | 6.87E-11 | + | 1.24E-04 | + | 3.18E-68 |
| cg16143804 | 6 | 31,706,344 | *VARS* | -39.0 | 6p21.33 | + | 2.68E-14 | + | 2.24E-04 | + | 3.18E-68 |
| cg16505946 | 6 | 31,870,305 | *CYP21A1P* | -103.2 | 6p21.33 | + | 1.18E-16 | - | 1.80E-04 | - | 2.34E-07 |
| cg01347674 | 11 | 118,134,073 | *HYOU1* | -781.4 | 11q23.3 | + | 5.14E-08 | + | 4.96E-04 | + | 1.01E-53 |
| cg18825076 | 15 | 78,729,989 | *CRABP1* | 89.4 | 15q25.1 | - | 2.40E-86 | - | 7.52E-04 | + | 2.06E-23 |
| cg08426200 | 15 | 78,802,280 | *CHRNA3* | -83.1 | 15q25.1 | + | 5.41E-40 | + | 6.56E-04 | + | 1.01E-09 |
| cg22563815 | 15 | 78,856,949 | *CHRNA3* | -28.5 | 15q25.1 | - | 2.28E-14 | + | 4.08E-05 | - | **1.33E-28** |
| cg17108064 | 15 | 78,857,060 | *CHRNA3* | -28.3 | 15q25.1 | - | 3.28E-17 | + | 2.26E-04 | - | **1.33E-28** |
| cg19696491 | 15 | 78,857,124 | *CHRNA3* | -28.3 | 15q25.1 | - | 2.76E-14 | + | 1.07E-04 | - | **1.33E-28** |
| cg21242079 | 15 | 79,101,063 | *KIAA1024* | -623.8 | 15q25.1 | + | 9.26E-10 | + | 1.09E-03 | + | 7.29E-28 |
| cg13835168 | 6 | 29,648,756 | *RNF39* | -389.4 | 6p22.1 | - | 7.00E-11 | + | 9.69E-03 | - | **1.57E-13** |

Chr, chromosome; Dir, direction of association/correlation.

[a] *P* values were calculated using SPrediXcan. All statistical tests were two-sided.

[b] *P* values were calculated using linear regression. All statistical tests were two-sided.

[c] *P* values were calculated using SPrediXcan and R *limma* package for data from GTEx and GEPIA2, respectively. For genes showing in both GTEx and GEPIA2 data, *P* values from GEPIA2 data are presented. *P* values from GTEx data are highlighted in bold. All statistical tests were two-sided.

**Supplementary Table 17. Consistent directions of associations across DNA methylation, gene expression, and ovarian cancer risk**

| CpG | Chr | Position (HG19) | Gene | Distance (Kb) | Cytoband | CpG-cancer | | CpG-gene | | Gene-cancer | |
|---|---|---|---|---|---|---|---|---|---|---|---|
| | | | | | | Dir | ***P*** [a] | Dir | ***P*** [b] | Dir | ***P*** [c] |
| cg03321133 | 2 | 176,993,115 | *HAGLROS* | -50.0 | 2q31.1 | - | 3.64E-11 | - | 1.67E-03 | + | 7.41E-52 |
| cg01293179 | 2 | 176,996,285 | *LINC01117* | -506.4 | 2q31.1 | - | 4.91E-11 | - | 3.37E-03 | + | 3.27E-16 |
| cg21932416 | 2 | 176,997,707 | *LINC01117* | -505.0 | 2q31.1 | - | 6.12E-10 | - | 4.74E-03 | + | 3.27E-16 |
| cg21932416 | 2 | 176,997,707 | *HOXD9* | 8.0 | 2q31.1 | - | 6.12E-10 | - | 6.11E-03 | + | **7.53E-06** |
| cg07011913 | 2 | 177,014,626 | *HAGLROS* | -28.5 | 2q31.1 | - | 4.00E-13 | - | 1.24E-03 | + | 7.41E-52 |
| cg12127282 | 2 | 177,014,685 | *HAGLROS* | -28.5 | 2q31.1 | - | 3.56E-08 | - | 3.16E-03 | + | 7.41E-52 |
| cg12127282 | 2 | 177,014,685 | *ATP5G3* | 965.7 | 2q31.1 | - | 3.56E-08 | - | 3.29E-03 | + | 6.05E-37 |
| cg00014998 | 2 | 177,014,962 | *ATP5G3* | 966.0 | 2q31.1 | - | 1.65E-13 | - | 3.06E-04 | + | 6.05E-37 |
| cg01152019 | 2 | 177,015,044 | *ATP5G3* | 966.1 | 2q31.1 | - | 1.98E-14 | - | 1.40E-03 | + | 6.05E-37 |
| cg01152019 | 2 | 177,015,044 | *HAGLROS* | -28.1 | 2q31.1 | - | 1.98E-14 | - | 3.37E-03 | + | 7.41E-52 |
| cg00767581 | 2 | 177,015,125 | *HAGLROS* | -28.0 | 2q31.1 | - | 5.69E-14 | - | 1.59E-03 | + | 7.41E-52 |
| cg19478682 | 2 | 177,015,812 | *HAGLROS* | -27.3 | 2q31.1 | - | 4.66E-13 | - | 1.27E-03 | + | 7.41E-52 |
| cg19478682 | 2 | 177,015,812 | *ATP5G3* | 966.9 | 2q31.1 | - | 4.66E-13 | - | 2.02E-03 | + | 6.05E-37 |
| cg21011616 | 2 | 177,015,992 | *ATP5G3* | 967.1 | 2q31.1 | - | 6.41E-14 | - | 7.39E-04 | + | 6.05E-37 |
| cg21011616 | 2 | 177,015,992 | *HAGLROS* | -27.2 | 2q31.1 | - | 6.41E-14 | - | 7.87E-04 | + | 7.41E-52 |
| cg12539415 | 2 | 177,016,167 | *HAGLROS* | -27.0 | 2q31.1 | - | 5.69E-14 | - | 9.53E-05 | + | 7.41E-52 |
| cg12539415 | 2 | 177,016,167 | *ATP5G3* | 967.2 | 2q31.1 | - | 5.69E-14 | - | 3.96E-03 | + | 6.05E-37 |
| cg18750960 | 2 | 177,016,417 | *HAGLROS* | -26.7 | 2q31.1 | - | 4.46E-14 | - | 7.29E-04 | + | 7.41E-52 |
| cg18750960 | 2 | 177,016,417 | *HOXD4* | Body | 2q31.1 | - | 4.46E-14 | - | 3.74E-03 | + | **1.74E-08** |
| cg19509393 | 2 | 177,016,490 | *HAGLROS* | -26.7 | 2q31.1 | - | 3.70E-14 | - | 2.23E-05 | + | 7.41E-52 |
| cg19509393 | 2 | 177,016,490 | *ATP5G3* | 967.6 | 2q31.1 | - | 3.70E-14 | - | 2.78E-03 | + | 6.05E-37 |
| cg19509393 | 2 | 177,016,490 | *HOXD4* | Body | 2q31.1 | - | 3.70E-14 | - | 3.90E-03 | + | **1.74E-08** |
| cg25951210 | 2 | 177,016,619 | *HAGLROS* | -26.5 | 2q31.1 | - | 4.30E-14 | - | 2.19E-04 | + | 7.41E-52 |
| cg10803577 | 2 | 177,016,837 | *HAGLROS* | -26.3 | 2q31.1 | - | 2.53E-14 | - | 5.26E-04 | + | 7.41E-52 |
| cg09981233 | 2 | 177,016,899 | *HAGLROS* | -26.3 | 2q31.1 | - | 3.42E-14 | - | 1.01E-03 | + | 7.41E-52 |
| cg26654148 | 2 | 177,016,978 | *HAGLROS* | -26.2 | 2q31.1 | - | 2.44E-14 | - | 1.13E-03 | + | 7.41E-52 |
| cg25304107 | 2 | 177,017,331 | *HAGLROS* | -25.8 | 2q31.1 | - | 1.15E-13 | - | 1.18E-04 | + | 7.41E-52 |
| cg16218721 | 2 | 177,017,352 | *HAGLROS* | -25.8 | 2q31.1 | - | 3.05E-14 | - | 6.39E-04 | + | 7.41E-52 |
| cg26590744 | 2 | 177,017,389 | *HAGLROS* | -25.8 | 2q31.1 | - | 1.32E-12 | - | 3.52E-04 | + | 7.41E-52 |
| cg01414185 | 2 | 177,017,449 | *HAGLROS* | -25.7 | 2q31.1 | - | 3.62E-14 | - | 3.84E-04 | + | 7.41E-52 |
| cg14900124 | 2 | 177,017,630 | *HAGLROS* | -25.5 | 2q31.1 | - | 2.41E-13 | - | 1.13E-03 | + | 7.41E-52 |
| cg09701582 | 2 | 177,017,797 | *HAGLROS* | -25.4 | 2q31.1 | - | 2.90E-14 | - | 3.61E-03 | + | 7.41E-52 |
| cg09701582 | 2 | 177,017,797 | *ATP5G3* | 968.9 | 2q31.1 | - | 2.90E-14 | - | 3.96E-03 | + | 6.05E-37 |
| cg26091021 | 2 | 177,021,826 | *ATP5G3* | 972.9 | 2q31.1 | - | 2.36E-13 | - | 2.12E-03 | + | 6.05E-37 |
| cg05342515 | 2 | 177,024,172 | *ATP5G3* | 975.2 | 2q31.1 | - | 3.53E-14 | - | 2.00E-03 | + | 6.05E-37 |
| cg24541426 | 2 | 177,029,459 | *HAGLROS* | -13.7 | 2q31.1 | - | 5.94E-13 | - | 3.75E-03 | + | 7.41E-52 |
| cg24541426 | 2 | 177,029,459 | *ATP5G3* | 980.5 | 2q31.1 | - | 5.94E-13 | - | 4.77E-03 | + | 6.05E-37 |
| cg24541426 | 2 | 177,029,459 | *HOXD4* | 11.5 | 2q31.1 | - | 5.94E-13 | - | 5.19E-03 | + | **1.74E-08** |
| cg04316624 | 2 | 177,036,809 | *HAGLROS* | -6.3 | 2q31.1 | - | 8.25E-08 | - | 3.69E-05 | + | 7.41E-52 |
| cg04316624 | 2 | 177,036,809 | *ATP5G3* | 987.9 | 2q31.1 | - | 8.25E-08 | - | 2.57E-03 | + | 6.05E-37 |
| cg00278547 | 2 | 177,037,212 | *HAGLROS* | -5.9 | 2q31.1 | - | 7.98E-13 | - | 3.76E-06 | + | 7.41E-52 |
| cg10159630 | 2 | 177,037,364 | *HAGLROS* | -5.8 | 2q31.1 | - | 8.03E-14 | - | 1.45E-05 | + | 7.41E-52 |
| cg10159630 | 2 | 177,037,364 | *ATP5G3* | 988.4 | 2q31.1 | - | 8.03E-14 | - | 3.17E-03 | + | 6.05E-37 |
| cg13053653 | 2 | 177,037,631 | *HAGLROS* | -5.5 | 2q31.1 | - | 3.03E-14 | - | 2.06E-04 | + | 7.41E-52 |
| cg21902187 | 2 | 177,037,710 | *HAGLROS* | -5.4 | 2q31.1 | - | 7.28E-11 | - | 4.48E-05 | + | 7.41E-52 |
| cg22934308 | 2 | 177,038,617 | *HAGLROS* | -4.5 | 2q31.1 | - | 4.61E-14 | - | 1.60E-04 | + | 7.41E-52 |
| cg10477905 | 2 | 177,039,295 | *HAGLROS* | -3.9 | 2q31.1 | - | 1.19E-13 | - | 7.75E-05 | + | 7.41E-52 |
| cg26754761 | 2 | 177,040,938 | *HAGLROS* | -2.2 | 2q31.1 | - | 3.97E-10 | - | 3.03E-03 | + | 7.41E-52 |
| cg26754761 | 2 | 177,040,938 | *LINC01117* | -461.7 | 2q31.1 | - | 3.97E-10 | - | 5.47E-03 | + | 3.27E-16 |
| cg23139698 | 2 | 177,041,515 | *HAGLROS* | -1.6 | 2q31.1 | - | 2.40E-13 | - | 3.26E-03 | + | 7.41E-52 |
| cg07797397 | 2 | 177,052,527 | *HAGLROS* | 8.8 | 2q31.1 | - | 7.20E-14 | - | 2.81E-04 | + | 7.41E-52 |
| cg00916973 | 17 | 43,580,461 | *LRRC37A4P* | -3.7 | 17q21.31 | - | 1.52E-11 | + | 3.08E-06 | - | 1.13E-13 |
| cg00916973 | 17 | 43,580,461 | *KANSL1-AS1* | -690.5 | 17q21.31 | - | 1.52E-11 | - | 3.32E-04 | + | **7.68E-12** |
| cg00916973 | 17 | 43,580,461 | *MAPT* | -391.3 | 17q21.31 | - | 1.52E-11 | + | 4.96E-03 | - | **5.20E-04** |
| cg00916973 | 17 | 43,580,461 | *LRRC37A* | -789.6 | 17q21.31 | - | 1.52E-11 | - | 5.73E-03 | + | **1.71E-07** |
| cg22433210 | 17 | 43,662,623 | *KANSL1-AS1* | -608.3 | 17q21.31 | + | 1.28E-12 | + | 7.99E-16 | + | **7.68E-12** |
| cg22433210 | 17 | 43,662,623 | *DND1P1* | -0.6 | 17q21.31 | + | 1.28E-12 | + | 1.30E-15 | + | 1.56E-07 |
| cg22433210 | 17 | 43,662,623 | *LRRC37A4P* | 34.9 | 17q21.31 | + | 1.28E-12 | - | 4.55E-10 | - | 1.13E-13 |
| cg22433210 | 17 | 43,662,623 | *LRRC37A2* | -926.3 | 17q21.31 | + | 1.28E-12 | + | 3.98E-09 | + | **2.36E-13** |
| cg22433210 | 17 | 43,662,623 | *LRRC37A* | -707.5 | 17q21.31 | + | 1.28E-12 | + | 7.24E-05 | + | **1.71E-07** |

| | | | | | | | | | | | |
|---|---|---|---|---|---|---|---|---|---|---|---|
| cg22433210 | 17 | 43,662,623 | *ARL17A* | -931.5 | 17q21.31 | + | 1.28E-12 | + | 4.30E-04 | + | **4.32E-07** |
| cg22433210 | 17 | 43,662,623 | *RP11-259G18.1* | -681.8 | 17q21.31 | + | 1.28E-12 | + | 4.63E-04 | + | 1.80E-06 |
| cg22433210 | 17 | 43,662,623 | *FAM215B* | -973.6 | 17q21.31 | + | 1.28E-12 | + | 7.45E-04 | + | **1.90E-07** |
| cg01341218 | 17 | 43,662,625 | *DND1P1* | -0.6 | 17q21.31 | + | 8.75E-13 | + | 1.51E-13 | + | 1.56E-07 |
| cg01341218 | 17 | 43,662,625 | *KANSL1-AS1* | -608.3 | 17q21.31 | + | 8.75E-13 | + | 1.42E-12 | + | **7.68E-12** |
| cg01341218 | 17 | 43,662,625 | *LRRC37A4P* | 34.9 | 17q21.31 | + | 8.75E-13 | - | 7.96E-10 | - | 1.13E-13 |
| cg01341218 | 17 | 43,662,625 | *LRRC37A2* | -926.3 | 17q21.31 | + | 8.75E-13 | + | 1.01E-07 | + | **2.36E-13** |
| cg01341218 | 17 | 43,662,625 | *LRRC37A* | -707.5 | 17q21.31 | + | 8.75E-13 | + | 2.81E-04 | + | **1.71E-07** |
| cg01341218 | 17 | 43,662,625 | *ARL17A* | -931.4 | 17q21.31 | + | 8.75E-13 | + | 3.62E-04 | + | **4.32E-07** |
| cg01341218 | 17 | 43,662,625 | *RP11-259G18.1* | -681.8 | 17q21.31 | + | 8.75E-13 | + | 5.36E-04 | + | 1.80E-06 |
| cg01341218 | 17 | 43,662,625 | *FAM215B* | -973.6 | 17q21.31 | + | 8.75E-13 | + | 1.52E-03 | + | **1.90E-07** |
| cg01341218 | 17 | 43,662,625 | *C1QL1* | 617.2 | 17q21.31 | + | 8.75E-13 | + | 6.12E-03 | + | 5.74E-23 |
| cg12297030 | 17 | 43,662,878 | *DND1P1* | -0.4 | 17q21.31 | + | 4.79E-12 | + | 1.05E-07 | + | 1.56E-07 |
| cg12297030 | 17 | 43,662,878 | *LRRC37A2* | -926.0 | 17q21.31 | + | 4.79E-12 | + | 6.43E-07 | + | **2.36E-13** |
| cg12297030 | 17 | 43,662,878 | *LRRC37A4P* | 35.2 | 17q21.31 | + | 4.79E-12 | - | 9.72E-07 | - | 1.13E-13 |
| cg12297030 | 17 | 43,662,878 | *KANSL1-AS1* | -608.1 | 17q21.31 | + | 4.79E-12 | + | 2.47E-06 | + | **7.68E-12** |
| cg12297030 | 17 | 43,662,878 | *ARL17A* | -931.2 | 17q21.31 | + | 4.79E-12 | + | 1.28E-04 | + | **4.32E-07** |
| cg12297030 | 17 | 43,662,878 | *LRRC37A* | -707.2 | 17q21.31 | + | 4.79E-12 | + | 7.01E-04 | + | **1.71E-07** |
| cg17117718 | 17 | 43,663,208 | *DND1P1* | 0.0 | 17q21.31 | + | 9.40E-13 | + | 6.51E-16 | + | 1.56E-07 |
| cg17117718 | 17 | 43,663,208 | *LRRC37A4P* | 35.5 | 17q21.31 | + | 9.40E-13 | - | 1.28E-12 | - | 1.13E-13 |
| cg17117718 | 17 | 43,663,208 | *LRRC37A2* | -925.7 | 17q21.31 | + | 9.40E-13 | + | 1.55E-11 | + | **2.36E-13** |
| cg17117718 | 17 | 43,663,208 | *KANSL1-AS1* | -607.7 | 17q21.31 | + | 9.40E-13 | + | 8.18E-11 | + | **7.68E-12** |
| cg17117718 | 17 | 43,663,208 | *ARL17A* | -930.9 | 17q21.31 | + | 9.40E-13 | + | 3.03E-07 | + | **4.32E-07** |
| cg17117718 | 17 | 43,663,208 | *LRRC37A* | -706.9 | 17q21.31 | + | 9.40E-13 | + | 6.96E-06 | + | **1.71E-07** |
| cg17117718 | 17 | 43,663,208 | *RP11-259G18.1* | -681.2 | 17q21.31 | + | 9.40E-13 | + | 1.03E-04 | + | 1.80E-06 |
| cg17117718 | 17 | 43,663,208 | *FAM215B* | -973.0 | 17q21.31 | + | 9.40E-13 | + | 9.24E-04 | + | **1.90E-07** |
| cg17117718 | 17 | 43,663,208 | *RPS26P8* | -22.7 | 17q21.31 | + | 9.40E-13 | + | 8.62E-03 | + | 3.65E-08 |
| cg05727186 | 17 | 43,697,356 | *RPS26P8* | 11.1 | 17q21.31 | - | 7.60E-10 | - | 9.95E-04 | + | 3.65E-08 |
| cg05727186 | 17 | 43,697,356 | *KANSL1-AS1* | -573.6 | 17q21.31 | - | 7.60E-10 | - | 1.39E-03 | + | **7.68E-12** |
| cg05727186 | 17 | 43,697,356 | *LRRC37A2* | -891.5 | 17q21.31 | - | 7.60E-10 | - | 3.09E-03 | + | **2.36E-13** |
| cg05727186 | 17 | 43,697,356 | *ARL17A* | -896.7 | 17q21.31 | - | 7.60E-10 | - | 4.25E-03 | + | **4.32E-07** |
| cg05727186 | 17 | 43,697,356 | *DND1P1* | 33.1 | 17q21.31 | - | 7.60E-10 | - | 5.57E-03 | + | 1.56E-07 |
| cg18815117 | 17 | 43,698,604 | *DND1P1* | 34.3 | 17q21.31 | - | 1.13E-12 | - | 1.97E-05 | + | 1.56E-07 |
| cg18815117 | 17 | 43,698,604 | *KANSL1-AS1* | -572.3 | 17q21.31 | - | 1.13E-12 | - | 1.14E-04 | + | **7.68E-12** |
| cg18815117 | 17 | 43,698,604 | *LRRC37A4P* | 70.9 | 17q21.31 | - | 1.13E-12 | + | 2.88E-04 | - | 1.13E-13 |
| cg18815117 | 17 | 43,698,604 | *LRRC37A2* | -890.3 | 17q21.31 | - | 1.13E-12 | - | 5.83E-04 | + | **2.36E-13** |
| cg01882395 | 17 | 43,717,810 | *LRRC37A4P* | 90.1 | 17q21.31 | + | 1.93E-13 | - | 2.53E-07 | - | 1.13E-13 |
| cg01882395 | 17 | 43,717,810 | *KANSL1-AS1* | -553.1 | 17q21.31 | + | 1.93E-13 | + | 5.20E-07 | + | **7.68E-12** |
| cg01882395 | 17 | 43,717,810 | *LRRC37A2* | -871.1 | 17q21.31 | + | 1.93E-13 | + | 2.04E-06 | + | **2.36E-13** |
| cg01882395 | 17 | 43,717,810 | *DND1P1* | 53.5 | 17q21.31 | + | 1.93E-13 | + | 9.15E-05 | + | 1.56E-07 |
| cg01882395 | 17 | 43,717,810 | *ARL17A* | -876.3 | 17q21.31 | + | 1.93E-13 | + | 8.04E-04 | + | **4.32E-07** |
| cg01882395 | 17 | 43,717,810 | *LRRC37A* | -652.3 | 17q21.31 | + | 1.93E-13 | + | 2.76E-03 | + | **1.71E-07** |
| cg18753072 | 17 | 43,719,194 | *KANSL1-AS1* | -551.8 | 17q21.31 | + | 2.37E-13 | + | 5.01E-05 | + | **7.68E-12** |
| cg18753072 | 17 | 43,719,194 | *LRRC37A2* | -869.7 | 17q21.31 | + | 2.37E-13 | + | 1.10E-04 | + | **2.36E-13** |
| cg18753072 | 17 | 43,719,194 | *DND1P1* | 54.9 | 17q21.31 | + | 2.37E-13 | + | 1.53E-04 | + | 1.56E-07 |
| cg18753072 | 17 | 43,719,194 | *ARL17A* | -874.9 | 17q21.31 | + | 2.37E-13 | + | 5.75E-04 | + | **4.32E-07** |
| cg18753072 | 17 | 43,719,194 | *LRRC37A* | -650.9 | 17q21.31 | + | 2.37E-13 | + | 1.63E-03 | + | **1.71E-07** |
| cg18753072 | 17 | 43,719,194 | *LRRC37A4P* | 91.5 | 17q21.31 | + | 2.37E-13 | - | 4.27E-03 | - | 1.13E-13 |
| cg20059597 | 17 | 43,726,219 | *DND1P1* | 61.9 | 17q21.31 | + | 2.99E-12 | + | 5.31E-08 | + | 1.56E-07 |
| cg20059597 | 17 | 43,726,219 | *KANSL1-AS1* | -544.7 | 17q21.31 | + | 2.99E-12 | + | 1.01E-05 | + | **7.68E-12** |
| cg20059597 | 17 | 43,726,219 | *LRRC37A2* | -862.7 | 17q21.31 | + | 2.99E-12 | + | 7.00E-05 | + | **2.36E-13** |
| cg20059597 | 17 | 43,726,219 | *LRRC37A4P* | 98.5 | 17q21.31 | + | 2.99E-12 | - | 4.56E-04 | - | 1.13E-13 |
| cg20059597 | 17 | 43,726,219 | *FAM215B* | -910.0 | 17q21.31 | + | 2.99E-12 | + | 3.15E-03 | + | **1.90E-07** |
| cg20059597 | 17 | 43,726,219 | *ARL17A* | -867.9 | 17q21.31 | + | 2.99E-12 | + | 5.07E-03 | + | **4.32E-07** |
| cg20393308 | 17 | 43,728,044 | *DND1P1* | 63.8 | 17q21.31 | + | 1.12E-12 | + | 4.34E-07 | + | 1.56E-07 |
| cg20393308 | 17 | 43,728,044 | *KANSL1-AS1* | -542.9 | 17q21.31 | + | 1.12E-12 | + | 2.30E-05 | + | **7.68E-12** |
| cg20393308 | 17 | 43,728,044 | *LRRC37A4P* | 100.3 | 17q21.31 | + | 1.12E-12 | - | 3.15E-05 | - | 1.13E-13 |
| cg20393308 | 17 | 43,728,044 | *LRRC37A2* | -860.8 | 17q21.31 | + | 1.12E-12 | + | 3.01E-04 | + | **2.36E-13** |
| cg20393308 | 17 | 43,728,044 | *ARL17A* | -866.0 | 17q21.31 | + | 1.12E-12 | + | 4.07E-03 | + | **4.32E-07** |
| cg20393308 | 17 | 43,728,044 | *LRRC37A* | -642.1 | 17q21.31 | + | 1.12E-12 | + | 6.51E-03 | + | **1.71E-07** |
| cg08276280 | 17 | 43,743,868 | *DND1P1* | 79.6 | 17q21.31 | + | 3.37E-11 | + | 1.84E-04 | + | 1.56E-07 |
| cg19447040 | 17 | 43,750,272 | *DND1P1* | 86.0 | 17q21.31 | + | 6.42E-09 | + | 4.12E-04 | + | 1.56E-07 |
| cg19447040 | 17 | 43,750,272 | *KANSL1-AS1* | -520.7 | 17q21.31 | + | 6.42E-09 | + | 8.07E-04 | + | **7.68E-12** |

| | | | | | | | | | | | |
|---|---|---|---|---|---|---|---|---|---|---|---|
| cg19447040 | 17 | 43,750,272 | *LRRC37A* | -619.8 | 17q21.31 | + | 6.42E-09 | + | 1.77E-03 | + | **1.71E-07** |
| cg06475362 | 17 | 43,763,639 | *KANSL1-AS1* | -507.3 | 17q21.31 | + | 4.39E-12 | + | 8.20E-04 | + | **7.68E-12** |
| cg07817266 | 17 | 43,765,890 | *DND1P1* | 101.6 | 17q21.31 | + | 2.46E-12 | + | 3.85E-09 | + | 1.56E-07 |
| cg07817266 | 17 | 43,765,890 | *LRRC37A4P* | 138.2 | 17q21.31 | + | 2.46E-12 | - | 9.39E-09 | - | 1.13E-13 |
| cg07817266 | 17 | 43,765,890 | *KANSL1-AS1* | -505.1 | 17q21.31 | + | 2.46E-12 | + | 1.86E-08 | + | **7.68E-12** |
| cg07817266 | 17 | 43,765,890 | *LRRC37A2* | -823.0 | 17q21.31 | + | 2.46E-12 | + | 1.43E-07 | + | **2.36E-13** |
| cg07817266 | 17 | 43,765,890 | *LRRC37A* | -604.2 | 17q21.31 | + | 2.46E-12 | + | 3.06E-05 | + | **1.71E-07** |
| cg07817266 | 17 | 43,765,890 | *ARL17A* | -828.2 | 17q21.31 | + | 2.46E-12 | + | 3.01E-04 | + | **4.32E-07** |
| cg07817266 | 17 | 43,765,890 | *RPS26P8* | 79.6 | 17q21.31 | + | 2.46E-12 | + | 6.51E-04 | + | 3.65E-08 |
| cg07817266 | 17 | 43,765,890 | *FAM215B* | -870.3 | 17q21.31 | + | 2.46E-12 | + | 2.97E-03 | + | **1.90E-07** |
| cg06537391 | 17 | 43,799,898 | *DND1P1* | 135.6 | 17q21.31 | + | 3.68E-13 | + | 4.73E-07 | + | 1.56E-07 |
| cg06537391 | 17 | 43,799,898 | *KANSL1-AS1* | -471.0 | 17q21.31 | + | 3.68E-13 | + | 8.64E-06 | + | **7.68E-12** |
| cg06537391 | 17 | 43,799,898 | *LRRC37A2* | -789.0 | 17q21.31 | + | 3.68E-13 | + | 3.22E-05 | + | **2.36E-13** |
| cg06537391 | 17 | 43,799,898 | *ARL17A* | -794.2 | 17q21.31 | + | 3.68E-13 | + | 1.30E-04 | + | **4.32E-07** |
| cg06537391 | 17 | 43,799,898 | *LRRC37A4P* | 172.2 | 17q21.31 | + | 3.68E-13 | - | 1.92E-03 | - | 1.13E-13 |
| cg06537391 | 17 | 43,799,898 | *FAM215B* | -836.3 | 17q21.31 | + | 3.68E-13 | + | 0.01 | + | **1.90E-07** |
| cg06537391 | 17 | 43,799,898 | *RP11-259G18.1* | -544.5 | 17q21.31 | + | 3.68E-13 | + | 0.01 | + | 1.80E-06 |
| cg06537391 | 17 | 43,799,898 | *LRRC37A* | -570.2 | 17q21.31 | + | 3.68E-13 | + | 0.01 | + | **1.71E-07** |
| cg11760414 | 17 | 43,863,398 | *KANSL1-AS1* | -407.5 | 17q21.31 | + | 3.44E-11 | + | 3.35E-03 | + | **7.68E-12** |
| cg15295732 | 17 | 43,942,128 | *DND1P1* | 277.8 | 17q21.31 | - | 4.53E-11 | - | 2.86E-08 | + | 1.56E-07 |
| cg15295732 | 17 | 43,942,128 | *LRRC37A2* | -646.8 | 17q21.31 | - | 4.53E-11 | - | 4.75E-07 | + | **2.36E-13** |
| cg15295732 | 17 | 43,942,128 | *KANSL1-AS1* | -328.8 | 17q21.31 | - | 4.53E-11 | - | 4.93E-06 | + | **7.68E-12** |
| cg15295732 | 17 | 43,942,128 | *LRRC37A4P* | 314.4 | 17q21.31 | - | 4.53E-11 | + | 1.03E-05 | - | 1.13E-13 |
| cg15295732 | 17 | 43,942,128 | *ARL17A* | -651.9 | 17q21.31 | - | 4.53E-11 | - | 1.31E-05 | + | **4.32E-07** |
| cg15295732 | 17 | 43,942,128 | *MAPT* | -29.6 | 17q21.31 | - | 4.53E-11 | + | 3.30E-03 | - | **5.20E-04** |
| cg15295732 | 17 | 43,942,128 | *RP11-259G18.1* | -402.3 | 17q21.31 | - | 4.53E-11 | - | 8.50E-03 | + | 1.80E-06 |
| cg05301556 | 17 | 43,971,177 | *DND1P1* | 306.9 | 17q21.31 | - | 4.84E-13 | - | 1.13E-08 | + | 1.56E-07 |
| cg05301556 | 17 | 43,971,177 | *KANSL1-AS1* | -299.8 | 17q21.31 | - | 4.84E-13 | - | 7.84E-05 | + | **7.68E-12** |
| cg05301556 | 17 | 43,971,177 | *LRRC37A4P* | 343.5 | 17q21.31 | - | 4.84E-13 | + | 2.46E-04 | - | 1.13E-13 |
| cg05301556 | 17 | 43,971,177 | *LRRC37A2* | -617.7 | 17q21.31 | - | 4.84E-13 | - | 5.11E-03 | + | **2.36E-13** |
| cg05301556 | 17 | 43,971,177 | *RP11-259G18.1* | -373.2 | 17q21.31 | - | 4.84E-13 | - | 5.41E-03 | + | 1.80E-06 |
| cg16520312 | 17 | 43,971,471 | *DND1P1* | 307.2 | 17q21.31 | - | 9.16E-13 | - | 4.36E-06 | + | 1.56E-07 |
| cg16520312 | 17 | 43,971,471 | *KANSL1-AS1* | -299.5 | 17q21.31 | - | 9.16E-13 | - | 1.14E-03 | + | **7.68E-12** |
| cg16520312 | 17 | 43,971,471 | *LRRC37A4P* | 343.8 | 17q21.31 | - | 9.16E-13 | + | 2.88E-03 | - | 1.13E-13 |
| cg16520312 | 17 | 43,971,471 | *LRRC37A2* | -617.4 | 17q21.31 | - | 9.16E-13 | - | 6.20E-03 | + | **2.36E-13** |
| cg13773723 | 17 | 43,971,894 | *KANSL1-AS1* | -299.1 | 17q21.31 | + | 3.59E-08 | + | 2.79E-04 | + | **7.68E-12** |
| cg13773723 | 17 | 43,971,894 | *LRRC37A2* | -617.0 | 17q21.31 | + | 3.59E-08 | + | 9.95E-04 | + | **2.36E-13** |
| cg13773723 | 17 | 43,971,894 | *DND1P1* | 307.6 | 17q21.31 | + | 3.59E-08 | + | 1.25E-03 | + | 1.56E-07 |
| cg13773723 | 17 | 43,971,894 | *FAM215B* | -664.3 | 17q21.31 | + | 3.59E-08 | + | 2.04E-03 | + | **1.90E-07** |
| cg13773723 | 17 | 43,971,894 | *LRRC37A4P* | 344.2 | 17q21.31 | + | 3.59E-08 | - | 6.47E-03 | - | 1.13E-13 |
| cg07163735 | 17 | 43,971,906 | *DND1P1* | 307.6 | 17q21.31 | + | 1.89E-11 | + | 8.16E-04 | + | 1.56E-07 |
| cg07163735 | 17 | 43,971,906 | *LRRC37A2* | -617.0 | 17q21.31 | + | 1.89E-11 | + | 1.56E-03 | + | **2.36E-13** |
| cg07163735 | 17 | 43,971,906 | *KANSL1-AS1* | -299.0 | 17q21.31 | + | 1.89E-11 | + | 3.66E-03 | + | **7.68E-12** |
| cg23202277 | 17 | 43,971,911 | *DND1P1* | 307.6 | 17q21.31 | + | 4.10E-12 | + | 9.67E-18 | + | 1.56E-07 |
| cg23202277 | 17 | 43,971,911 | *KANSL1-AS1* | -299.0 | 17q21.31 | + | 4.10E-12 | + | 4.31E-15 | + | **7.68E-12** |
| cg23202277 | 17 | 43,971,911 | *LRRC37A4P* | 344.2 | 17q21.31 | + | 4.10E-12 | - | 2.10E-10 | - | 1.13E-13 |
| cg23202277 | 17 | 43,971,911 | *LRRC37A2* | -617.0 | 17q21.31 | + | 4.10E-12 | + | 2.34E-09 | + | **2.36E-13** |
| cg23202277 | 17 | 43,971,911 | *RP11-259G18.1* | -372.5 | 17q21.31 | + | 4.10E-12 | + | 1.65E-06 | + | 1.80E-06 |
| cg23202277 | 17 | 43,971,911 | *ARL17A* | -622.2 | 17q21.31 | + | 4.10E-12 | + | 9.48E-06 | + | **4.32E-07** |
| cg23202277 | 17 | 43,971,911 | *LRRC37A* | -398.2 | 17q21.31 | + | 4.10E-12 | + | 1.67E-05 | + | **1.71E-07** |
| cg23202277 | 17 | 43,971,911 | *RPS26P8* | 285.6 | 17q21.31 | + | 4.10E-12 | + | 2.46E-04 | + | 3.65E-08 |
| cg23202277 | 17 | 43,971,911 | *FAM215B* | -664.3 | 17q21.31 | + | 4.10E-12 | + | 3.02E-03 | + | **1.90E-07** |
| cg20099416 | 17 | 43,971,919 | *DND1P1* | 307.6 | 17q21.31 | + | 1.26E-11 | + | 2.81E-16 | + | 1.56E-07 |
| cg20099416 | 17 | 43,971,919 | *KANSL1-AS1* | -299.0 | 17q21.31 | + | 1.26E-11 | + | 2.59E-14 | + | **7.68E-12** |
| cg20099416 | 17 | 43,971,919 | *LRRC37A2* | -617.0 | 17q21.31 | + | 1.26E-11 | + | 3.65E-10 | + | **2.36E-13** |
| cg20099416 | 17 | 43,971,919 | *LRRC37A4P* | 344.2 | 17q21.31 | + | 1.26E-11 | - | 9.32E-10 | - | 1.13E-13 |
| cg20099416 | 17 | 43,971,919 | *ARL17A* | -622.2 | 17q21.31 | + | 1.26E-11 | + | 3.13E-05 | + | **4.32E-07** |
| cg20099416 | 17 | 43,971,919 | *LRRC37A* | -398.2 | 17q21.31 | + | 1.26E-11 | + | 4.49E-05 | + | **1.71E-07** |
| cg20099416 | 17 | 43,971,919 | *RP11-259G18.1* | -372.5 | 17q21.31 | + | 1.26E-11 | + | 2.02E-03 | + | 1.80E-06 |
| cg20099416 | 17 | 43,971,919 | *RPS26P8* | 285.6 | 17q21.31 | + | 1.26E-11 | + | 2.16E-03 | + | 3.65E-08 |
| cg20099416 | 17 | 43,971,919 | *FAM215B* | -664.3 | 17q21.31 | + | 1.26E-11 | + | 4.13E-03 | + | **1.90E-07** |
| cg00891649 | 17 | 43,972,573 | *LRRC37A4P* | 344.9 | 17q21.31 | - | 2.90E-13 | + | 1.71E-03 | - | 1.13E-13 |
| cg00891649 | 17 | 43,972,573 | *DND1P1* | 308.3 | 17q21.31 | - | 2.90E-13 | - | 2.33E-03 | + | 1.56E-07 |

| | | | | | | | | | | | |
|---|---|---|---|---|---|---|---|---|---|---|---|
| cg11489262 | 17 | 43,973,426 | *LRRC37A4P* | 345.7 | 17q21.31 | + | 1.04E-12 | - | 7.62E-08 | - | 1.13E-13 |
| cg11489262 | 17 | 43,973,426 | *KANSL1-AS1* | -297.5 | 17q21.31 | + | 1.04E-12 | + | 5.21E-07 | + | **7.68E-12** |
| cg11489262 | 17 | 43,973,426 | *DND1P1* | 309.1 | 17q21.31 | + | 1.04E-12 | + | 9.19E-07 | + | 1.56E-07 |
| cg11489262 | 17 | 43,973,426 | *LRRC37A2* | -615.5 | 17q21.31 | + | 1.04E-12 | + | 5.69E-04 | + | **2.36E-13** |
| cg11489262 | 17 | 43,973,426 | *LRRC37A* | -396.7 | 17q21.31 | + | 1.04E-12 | + | 8.37E-03 | + | **1.71E-07** |
| cg10780632 | 17 | 43,973,522 | *DND1P1* | 309.2 | 17q21.31 | + | 1.40E-08 | + | 5.46E-11 | + | 1.56E-07 |
| cg10780632 | 17 | 43,973,522 | *LRRC37A4P* | 345.8 | 17q21.31 | + | 1.40E-08 | - | 1.14E-07 | - | 1.13E-13 |
| cg10780632 | 17 | 43,973,522 | *KANSL1-AS1* | -297.4 | 17q21.31 | + | 1.40E-08 | + | 5.89E-07 | + | **7.68E-12** |
| cg10780632 | 17 | 43,973,522 | *LRRC37A2* | -615.4 | 17q21.31 | + | 1.40E-08 | + | 2.52E-05 | + | **2.36E-13** |
| cg10780632 | 17 | 43,973,522 | *LRRC37A* | -396.6 | 17q21.31 | + | 1.40E-08 | + | 5.24E-04 | + | **1.71E-07** |
| cg10780632 | 17 | 43,973,522 | *ARL17A* | -620.6 | 17q21.31 | + | 1.40E-08 | + | 2.94E-03 | + | **4.32E-07** |
| cg10780632 | 17 | 43,973,522 | *RP11-259G18.1* | -370.9 | 17q21.31 | + | 1.40E-08 | + | 3.17E-03 | + | 1.80E-06 |
| cg10780632 | 17 | 43,973,522 | *MAPT* | Body | 17q21.31 | + | 1.40E-08 | - | 8.12E-03 | - | **5.20E-04** |
| cg01630519 | 17 | 43,992,416 | *DND1P1* | 328.1 | 17q21.31 | + | 5.11E-13 | + | 3.47E-05 | + | 1.56E-07 |
| cg01630519 | 17 | 43,992,416 | *KANSL1-AS1* | -278.5 | 17q21.31 | + | 5.11E-13 | + | 1.35E-03 | + | **7.68E-12** |
| cg01630519 | 17 | 43,992,416 | *RP11-259G18.1* | -352.0 | 17q21.31 | + | 5.11E-13 | + | 5.54E-03 | + | 1.80E-06 |
| cg01630519 | 17 | 43,992,416 | *LRRC37A2* | -596.5 | 17q21.31 | + | 5.11E-13 | + | 6.34E-03 | + | **2.36E-13** |
| cg23926235 | 17 | 44,024,256 | *DND1P1* | 360.0 | 17q21.31 | - | 8.65E-13 | - | 6.36E-04 | + | 1.56E-07 |
| cg23926235 | 17 | 44,024,256 | *KANSL1-AS1* | -246.7 | 17q21.31 | - | 8.65E-13 | - | 3.16E-03 | + | **7.68E-12** |
| cg23926235 | 17 | 44,024,256 | *LRRC37A2* | -564.6 | 17q21.31 | - | 8.65E-13 | - | 4.35E-03 | + | **2.36E-13** |
| cg05772917 | 17 | 44,027,251 | *DND1P1* | 363.0 | 17q21.31 | + | 4.14E-12 | + | 1.91E-07 | + | 1.56E-07 |
| cg05772917 | 17 | 44,027,251 | *KANSL1-AS1* | -243.7 | 17q21.31 | + | 4.14E-12 | + | 1.40E-04 | + | **7.68E-12** |
| cg05772917 | 17 | 44,027,251 | *LRRC37A2* | -561.6 | 17q21.31 | + | 4.14E-12 | + | 2.74E-03 | + | **2.36E-13** |
| cg05772917 | 17 | 44,027,251 | *LRRC37A4P* | 399.6 | 17q21.31 | + | 4.14E-12 | - | 9.25E-03 | - | 1.13E-13 |
| cg00112026 | 17 | 44,033,451 | *DND1P1* | 369.2 | 17q21.31 | + | 4.90E-11 | + | 1.49E-06 | + | 1.56E-07 |
| cg00112026 | 17 | 44,033,451 | *KANSL1-AS1* | -237.5 | 17q21.31 | + | 4.90E-11 | + | 4.85E-06 | + | **7.68E-12** |
| cg00112026 | 17 | 44,033,451 | *LRRC37A2* | -555.4 | 17q21.31 | + | 4.90E-11 | + | 2.87E-05 | + | **2.36E-13** |
| cg00112026 | 17 | 44,033,451 | *LRRC37A4P* | 405.8 | 17q21.31 | + | 4.90E-11 | - | 8.45E-05 | - | 1.13E-13 |
| cg00112026 | 17 | 44,033,451 | *ARL17A* | -560.6 | 17q21.31 | + | 4.90E-11 | + | 1.23E-03 | + | **4.32E-07** |
| cg22635938 | 17 | 44,039,549 | *LRRC37A2* | -549.3 | 17q21.31 | - | 1.36E-14 | - | 3.75E-05 | + | **2.36E-13** |
| cg22635938 | 17 | 44,039,549 | *ARL17A* | -554.5 | 17q21.31 | - | 1.36E-14 | - | 4.57E-05 | + | **4.32E-07** |
| cg22635938 | 17 | 44,039,549 | *KANSL1-AS1* | -231.4 | 17q21.31 | - | 1.36E-14 | - | 7.44E-04 | + | **7.68E-12** |
| cg18586914 | 17 | 44,040,821 | *LRRC37A2* | -548.1 | 17q21.31 | - | 5.10E-12 | - | 1.40E-09 | + | **2.36E-13** |
| cg18586914 | 17 | 44,040,821 | *KANSL1-AS1* | -230.1 | 17q21.31 | - | 5.10E-12 | - | 2.49E-09 | + | **7.68E-12** |
| cg18586914 | 17 | 44,040,821 | *LRRC37A4P* | 413.1 | 17q21.31 | - | 5.10E-12 | + | 5.25E-08 | - | 1.13E-13 |
| cg18586914 | 17 | 44,040,821 | *DND1P1* | 376.5 | 17q21.31 | - | 5.10E-12 | - | 3.84E-06 | + | 1.56E-07 |
| cg18586914 | 17 | 44,040,821 | *ARL17A* | -553.3 | 17q21.31 | - | 5.10E-12 | - | 2.60E-05 | + | **4.32E-07** |
| cg18586914 | 17 | 44,040,821 | *FAM215B* | -595.4 | 17q21.31 | - | 5.10E-12 | - | 1.38E-03 | + | **1.90E-07** |
| cg18586914 | 17 | 44,040,821 | *RP11-259G18.1* | -303.6 | 17q21.31 | - | 5.10E-12 | - | 2.12E-03 | + | 1.80E-06 |
| cg18586914 | 17 | 44,040,821 | *C1QL1* | 995.4 | 17q21.31 | - | 5.10E-12 | - | 4.31E-03 | + | 5.74E-23 |
| cg18586914 | 17 | 44,040,821 | *NSF* | -627.2 | 17q21.31 | - | 5.10E-12 | + | 6.97E-03 | - | **7.08E-11** |
| cg18586914 | 17 | 44,040,821 | *ARHGAP27* | 529.0 | 17q21.31 | - | 5.10E-12 | - | 0.01 | + | 2.23E-13 |
| cg01934064 | 17 | 44,064,242 | *DND1P1* | 400.0 | 17q21.31 | + | 4.92E-12 | + | 3.34E-04 | + | 1.56E-07 |
| cg01934064 | 17 | 44,064,242 | *KANSL1-AS1* | -206.7 | 17q21.31 | + | 4.92E-12 | + | 4.85E-04 | + | **7.68E-12** |
| cg01934064 | 17 | 44,064,242 | *LRRC37A2* | -524.6 | 17q21.31 | + | 4.92E-12 | + | 7.28E-04 | + | **2.36E-13** |
| cg01934064 | 17 | 44,064,242 | *ARL17A* | -529.8 | 17q21.31 | + | 4.92E-12 | + | 2.29E-03 | + | **4.32E-07** |
| cg05721485 | 17 | 44,071,124 | *DND1P1* | 406.8 | 17q21.31 | + | 5.94E-13 | + | 2.74E-04 | + | 1.56E-07 |
| cg05721485 | 17 | 44,071,124 | *KANSL1-AS1* | -199.8 | 17q21.31 | + | 5.94E-13 | + | 5.19E-04 | + | **7.68E-12** |
| cg05721485 | 17 | 44,071,124 | *LRRC37A4P* | 443.4 | 17q21.31 | + | 5.94E-13 | - | 3.24E-03 | - | 1.13E-13 |
| cg00792758 | 17 | 44,087,219 | *DND1P1* | 422.9 | 17q21.31 | - | 2.36E-13 | - | 5.33E-09 | + | 1.56E-07 |
| cg00792758 | 17 | 44,087,219 | *KANSL1-AS1* | -183.7 | 17q21.31 | - | 2.36E-13 | - | 1.84E-08 | + | **7.68E-12** |
| cg00792758 | 17 | 44,087,219 | *LRRC37A2* | -501.7 | 17q21.31 | - | 2.36E-13 | - | 7.07E-07 | + | **2.36E-13** |
| cg00792758 | 17 | 44,087,219 | *LRRC37A4P* | 459.5 | 17q21.31 | - | 2.36E-13 | + | 1.50E-06 | - | 1.13E-13 |
| cg00792758 | 17 | 44,087,219 | *ARL17A* | -506.9 | 17q21.31 | - | 2.36E-13 | - | 4.90E-05 | + | **4.32E-07** |
| cg00792758 | 17 | 44,087,219 | *WNT3* | -752.7 | 17q21.31 | - | 2.36E-13 | - | 5.89E-03 | + | **2.96E-11** |
| cg07694853 | 17 | 44,116,674 | *RP11-156P1.3* | -944.7 | 17q21.31 | - | 8.46E-09 | + | 4.43E-06 | - | 0.02 |
| cg08318660 | 17 | 44,122,580 | *DND1P1* | 458.3 | 17q21.31 | + | 2.06E-10 | + | 3.71E-04 | + | 1.56E-07 |
| cg08318660 | 17 | 44,122,580 | *LRRC37A* | -247.5 | 17q21.31 | + | 2.06E-10 | + | 9.47E-04 | + | **1.71E-07** |
| cg08318660 | 17 | 44,122,580 | *RP11-259G18.1* | -221.8 | 17q21.31 | + | 2.06E-10 | + | 2.79E-03 | + | 1.80E-06 |
| cg08318660 | 17 | 44,122,580 | *KANSL1-AS1* | -148.4 | 17q21.31 | + | 2.06E-10 | + | 3.28E-03 | + | **7.68E-12** |
| cg22192454 | 17 | 44,135,007 | *KANSL1-AS1* | -135.9 | 17q21.31 | - | 6.96E-10 | - | 2.94E-03 | + | **7.68E-12** |
| cg22192454 | 17 | 44,135,007 | *LRRC37A4P* | 507.3 | 17q21.31 | - | 6.96E-10 | + | 3.87E-03 | - | 1.13E-13 |
| cg21214508 | 17 | 44,248,233 | *LRRC37A4P* | 620.5 | 17q21.31 | - | 4.29E-14 | + | 8.91E-06 | - | 1.13E-13 |

| | | | | | | | | | | | |
|---|---|---|---|---|---|---|---|---|---|---|---|
| cg21214508 | 17 | 44,248,233 | *DND1P1* | 583.9 | 17q21.31 | - | 4.29E-14 | - | 1.31E-05 | + | 1.56E-07 |
| cg21214508 | 17 | 44,248,233 | *KANSL1-AS1* | -22.7 | 17q21.31 | - | 4.29E-14 | - | 1.89E-05 | + | **7.68E-12** |
| cg21214508 | 17 | 44,248,233 | *LRRC37A2* | -340.6 | 17q21.31 | - | 4.29E-14 | - | 8.80E-05 | + | **2.36E-13** |
| cg21214508 | 17 | 44,248,233 | *ARL17A* | -345.8 | 17q21.31 | - | 4.29E-14 | - | 3.79E-04 | + | **4.32E-07** |
| cg21214508 | 17 | 44,248,233 | *LRRC37A* | -121.9 | 17q21.31 | - | 4.29E-14 | - | 6.14E-03 | + | **1.71E-07** |
| cg21214508 | 17 | 44,248,233 | *FAM215B* | -388.0 | 17q21.31 | - | 4.29E-14 | - | 0.01 | + | **1.90E-07** |
| cg10191506 | 17 | 44,260,227 | *LRRC37A4P* | 632.5 | 17q21.31 | - | 9.99E-12 | + | 4.73E-04 | - | 1.13E-13 |
| cg04739613 | 17 | 44,270,960 | *DND1P1* | 606.7 | 17q21.31 | + | 7.64E-11 | + | 1.15E-03 | + | 1.56E-07 |
| cg09250749 | 17 | 44,271,316 | *KANSL1-AS1* | Body | 17q21.31 | + | 2.45E-13 | + | 1.66E-03 | + | **7.68E-12** |
| cg09250749 | 17 | 44,271,316 | *LRRC37A2* | -317.6 | 17q21.31 | + | 2.45E-13 | + | 1.78E-03 | + | **2.36E-13** |
| cg17642057 | 17 | 44,274,734 | *LRRC37A2* | -314.1 | 17q21.31 | - | 2.24E-08 | - | 2.11E-04 | + | **2.36E-13** |
| cg17642057 | 17 | 44,274,734 | *ARL17A* | -319.3 | 17q21.31 | - | 2.24E-08 | - | 2.30E-04 | + | **4.32E-07** |
| cg17642057 | 17 | 44,274,734 | *KANSL1-AS1* | 0.7 | 17q21.31 | - | 2.24E-08 | - | 1.36E-03 | + | **7.68E-12** |
| cg15790431 | 17 | 44,292,828 | *KANSL1-AS1* | 18.7 | 17q21.31 | - | 5.23E-12 | - | 7.67E-06 | + | **7.68E-12** |
| cg15790431 | 17 | 44,292,828 | *ARL17A* | -301.2 | 17q21.31 | - | 5.23E-12 | - | 2.92E-04 | + | **4.32E-07** |
| cg15790431 | 17 | 44,292,828 | *LRRC37A2* | -296.1 | 17q21.31 | - | 5.23E-12 | - | 5.80E-04 | + | **2.36E-13** |
| cg15790431 | 17 | 44,292,828 | *LRRC37A4P* | 665.1 | 17q21.31 | - | 5.23E-12 | + | 3.10E-03 | - | 1.13E-13 |
| cg15790431 | 17 | 44,292,828 | *DND1P1* | 628.5 | 17q21.31 | - | 5.23E-12 | - | 3.46E-03 | + | 1.56E-07 |
| cg15790431 | 17 | 44,292,828 | *ARHGAP27* | 781.0 | 17q21.31 | - | 5.23E-12 | - | 9.29E-03 | + | 2.23E-13 |
| cg15790431 | 17 | 44,292,828 | *RP11-259G18.1* | -51.6 | 17q21.31 | - | 5.23E-12 | - | 0.02 | + | 1.80E-06 |
| cg17911788 | 17 | 44,343,683 | *KANSL1-AS1* | 69.6 | 17q21.31 | - | 4.82E-09 | - | 2.75E-04 | + | **7.68E-12** |
| cg17911788 | 17 | 44,343,683 | *DND1P1* | 679.4 | 17q21.31 | - | 4.82E-09 | - | 4.41E-04 | + | 1.56E-07 |
| cg17911788 | 17 | 44,343,683 | *LRRC37A2* | -245.2 | 17q21.31 | - | 4.82E-09 | - | 2.19E-03 | + | **2.36E-13** |
| cg17911788 | 17 | 44,343,683 | *LRRC37A4P* | 716.0 | 17q21.31 | - | 4.82E-09 | + | 3.58E-03 | - | 1.13E-13 |
| cg05159804 | 17 | 44,343,776 | *DND1P1* | 679.5 | 17q21.31 | - | 1.04E-12 | - | 3.97E-03 | + | 1.56E-07 |

Chr, chromosome; Dir, direction of association/correlation.

[a] *P* values were calculated using SPrediXcan. All statistical tests were two-sided.

[b] *P* values were calculated using linear regression. All statistical tests were two-sided.

[c] *P* values were calculated using SPrediXcan and R limma package for data from GTEx and GEPIA2, respectively. For genes showing in both GTEx and GEPIA2 data, *P* values from GEPIA2 data are presented. *P* values from GTEx data are highlighted in bold. All statistical tests were two-sided.

**Supplementary Table 18. Consistent directions of associations across DNA methylation, gene expression, and prostate cancer risk**

| CpG | Chr | Position (HG19) | Gene | Distance (Kb) | Cytoband | CpG-cancer | | CpG-gene | | Gene-cancer | |
|---|---|---|---|---|---|---|---|---|---|---|---|
| | | | | | | Dir | *P* [a] | Dir | *P* [b] | Dir | *P* [c] |
| cg09590377 | 2 | 8,597,389 | *CPSF3* | -966.3 | 2p25.1 | - | 5.13E-09 | - | 1.22E-03 | + | 1.91E-31 |
| cg27658811 | 2 | 62,494,078 | *CCT4* | 378.2 | 2p15 | - | 5.10E-10 | - | 2.54E-03 | + | 2.68E-45 |
| cg19825503 | 2 | 63,155,443 | *WDPCP* | -193.1 | 2p15 | + | 1.58E-08 | + | 1.48E-03 | + | **1.51E-24** |
| cg17397364 | 2 | 63,273,436 | *WDPCP* | -75.1 | 2p15 | + | 4.80E-23 | + | 2.41E-05 | + | **1.51E-24** |
| cg17397364 | 2 | 63,273,436 | *TMEM17* | 540.0 | 2p15 | + | 4.80E-23 | + | 6.17E-03 | + | 2.18E-23 |
| cg15853117 | 2 | 63,276,966 | *WDPCP* | -71.6 | 2p15 | - | 4.72E-10 | - | 2.74E-03 | + | **1.51E-24** |
| cg19179185 | 6 | 30,731,397 | *BAG6* | -875.4 | 6p21.33 | + | 4.13E-10 | + | 5.33E-05 | + | 0.02 |
| cg14166520 | 6 | 31,532,351 | *XXbac-BPG248L24.10* | 256.0 | 6p21.33 | - | 2.02E-07 | - | 1.81E-04 | + | 3.79E-10 |
| cg17360552 | 6 | 32,725,332 | *HLA-DQA2* | 10.3 | 6p21.32 | + | 1.22E-09 | + | 8.52E-06 | + | 2.88E-06 |
| cg17360552 | 6 | 32,725,332 | *HLA-DRB6* | 197.5 | 6p21.32 | + | 1.22E-09 | + | 1.72E-03 | + | 7.14E-16 |
| cg27176262 | 6 | 32,797,830 | *HLA-DRB5* | 299.8 | 6p21.32 | - | 3.56E-08 | - | 8.91E-05 | + | 0.03 |
| cg12209329 | 7 | 97,929,748 | *RP11-307C18.1* | -22.4 | 7q21.3 | + | 4.41E-20 | - | 9.17E-04 | - | 2.75E-32 |
| cg06500932 | 8 | 127,882,188 | *CASC8* | -419.9 | 8q24.21 | - | 2.03E-13 | - | 5.49E-03 | + | 6.41E-24 |
| cg01725759 | 8 | 127,916,305 | *RP11-89K10.1* | 190.7 | 8q24.21 | - | 1.96E-13 | - | 2.72E-03 | + | 3.50E-29 |
| cg04657445 | 8 | 128,390,627 | *RP11-89K10.1* | 665.0 | 8q24.21 | - | 5.97E-67 | - | 4.68E-03 | + | 3.50E-29 |
| cg09800781 | 10 | 104,253,015 | *RP11-724N1.1* | -421.3 | 10q24.32 | - | 2.78E-09 | - | 1.68E-04 | + | **8.62E-04** |
| cg09800781 | 10 | 104,253,015 | *MFSD13A* | 16.2 | 10q24.32 | - | 2.78E-09 | - | 7.91E-04 | + | **1.96E-10** |
| cg09800781 | 10 | 104,253,015 | *C10orf95* | 41.9 | 10q24.32 | - | 2.78E-09 | - | 2.21E-03 | + | 2.83E-22 |
| cg02742593 | 11 | 68,481,342 | *ALDH3B1* | 684.6 | 11q13.3 | - | 1.56E-09 | + | 1.44E-03 | - | 1.08E-34 |
| cg00448220 | 17 | 624,386 | *MIR22HG* | -990.4 | 17p13.3 | + | 2.49E-19 | - | 9.45E-04 | - | 1.78E-38 |
| cg07420359 | 18 | 76,768,732 | *CTDP1* | -671.1 | 18q23 | - | 4.35E-16 | + | 1.02E-03 | - | 1.18E-18 |
| cg15272956 | 20 | 62,332,704 | *RTEL1* | 5.1 | 20q13.33 | + | 1.10E-18 | + | 4.23E-04 | + | **3.40E-22** |
| cg00343092 | 22 | 43,547,974 | *NFAM1* | 719.6 | 22q13.2 | + | 3.24E-10 | - | 2.01E-03 | - | 2.83E-03 |

Chr, chromosome; Dir, direction of association/correlation.

[a] *P* values were calculated using SPrediXcan. All statistical tests were two-sided.

[b] *P* values were calculated using linear regression. All statistical tests were two-sided.

[c] *P* values were calculated using SPrediXcan and R *limma* package for data from GTEx and GEPIA2, respectively. For genes showing in both GTEx and GEPIA2 data, *P* values from GEPIA2 data are presented. *P* values from GTEx data are highlighted in bold. All statistical tests were two-sided.

**Supplementary Table 19. Consistent directions of associations across DNA methylation, gene expression, and testicular germ cell cancer risk**

| CpG | Chr | Position (HG19) | Gene | Distance (Kb) | Cytoband | CpG-cancer | | CpG-gene | | Gene-cancer | |
|---|---|---|---|---|---|---|---|---|---|---|---|
| | | | | | | Dir | *P* [a] | Dir | *P* [b] | Dir | *P* [c] |
| cg15120454 | 1 | 156,339,198 | *BGLAP* | 126.1 | 1q22 | - | 5.43E-10 | + | 2.66E-04 | - | **8.14E-04** |
| cg19626747 | 6 | 150,205,807 | *RP1-111D6.3* | 639.1 | 6q25.1 | - | 1.63E-08 | + | 3.70E-04 | - | 1.74E-19 |
| cg13206674 | 6 | 150,067,644 | *RP1-111D6.4* | 489.0 | 6q25.1 | - | 1.01E-08 | + | 5.71E-04 | - | 2.11E-69 |
| cg18279126 | 7 | 2,041,391 | *AC073957.15* | 957.9 | 7p22.3 | + | 3.02E-08 | - | 9.33E-04 | - | 1.18E-46 |
| cg25122824 | 7 | 1,886,917 | *AC102953.4* | 383.3 | 7p22.3 | - | 9.19E-16 | + | 1.03E-03 | - | 1.39E-109 |
| cg22340370 | 7 | 2,019,882 | *MRM2* | -254.0 | 7p22.3 | + | 2.06E-16 | + | 1.15E-03 | + | 3.02E-99 |
| cg20185615 | 7 | 2,316,906 | *GRIFIN* | -197.6 | 7p22.3 | - | 6.54E-08 | + | 1.41E-03 | - | 1.83E-18 |

Chr, chromosome; Dir, direction of association/correlation; DNAm, DNA methylation of CpG; GEx, gene expression.

[a] *P* values were calculated using SPrediXcan. All statistical tests were two-sided.

[b] *P* values were calculated using linear regression. All statistical tests were two-sided.

[c] *P* values were calculated using SPrediXcan and R *limma* package for data from GTEx and GEPIA2, respectively. For genes showing in both GTEx and GEPIA2 data, *P* values from GEPIA2 data are presented. *P* values from GTEx data are highlighted in bold. All statistical tests were two-sided.
